# Discovery of 95 PTSD loci provides insight into genetic architecture and neurobiology of trauma and stress-related disorders

**DOI:** 10.1101/2023.08.31.23294915

**Authors:** Caroline M Nievergelt, Adam X Maihofer, Elizabeth G Atkinson, Chia-Yen Chen, Karmel W Choi, Jonathan RI Coleman, Nikolaos P Daskalakis, Laramie E Duncan, Renato Polimanti, Cindy Aaronson, Ananda B Amstadter, Soren B Andersen, Ole A Andreassen, Paul A Arbisi, Allison E Ashley-Koch, S Bryn Austin, Esmina Avdibegoviç, Dragan Babic, Silviu-Alin Bacanu, Dewleen G Baker, Anthony Batzler, Jean C Beckham, Sintia Belangero, Corina Benjet, Carisa Bergner, Linda M Bierer, Joanna M Biernacka, Laura J Bierut, Jonathan I Bisson, Marco P Boks, Elizabeth A Bolger, Amber Brandolino, Gerome Breen, Rodrigo Affonseca Bressan, Richard A Bryant, Angela C Bustamante, Jonas Bybjerg-Grauholm, Marie Bækvad-Hansen, Anders D Børglum, Sigrid Børte, Leah Cahn, Joseph R Calabrese, Jose Miguel Caldas-de-Almeida, Chris Chatzinakos, Sheraz Cheema, Sean A P Clouston, LucÍa Colodro-Conde, Brandon J Coombes, Carlos S Cruz-Fuentes, Anders M Dale, Shareefa Dalvie, Lea K Davis, Jürgen Deckert, Douglas L Delahanty, Michelle F Dennis, Terri deRoon-Cassini, Frank Desarnaud, Christopher P DiPietro, Seth G Disner, Anna R Docherty, Katharina Domschke, Grete Dyb, Alma Dzubur Kulenovic, Howard J Edenberg, Alexandra Evans, Chiara Fabbri, Negar Fani, Lindsay A Farrer, Adriana Feder, Norah C Feeny, Janine D Flory, David Forbes, Carol E Franz, Sandro Galea, Melanie E Garrett, Bizu Gelaye, Joel Gelernter, Elbert Geuze, Charles F Gillespie, Aferdita Goci, Slavina B Goleva, Scott D Gordon, Lana Ruvolo Grasser, Camila Guindalini, Magali Haas, Saskia Hagenaars, Michael A Hauser, Andrew C Heath, Sian MJ Hemmings, Victor Hesselbrock, Ian B Hickie, Kelleigh Hogan, David Michael Hougaard, Hailiang Huang, Laura M Huckins, Kristian Hveem, Miro Jakovljevic, Arash Javanbakht, Gregory D Jenkins, Jessica Johnson, Ian Jones, Tanja Jovanovic, Karen-Inge Karstoft, Milissa L Kaufman, James L Kennedy, Ronald C Kessler, Alaptagin Khan, Nathan A Kimbrel, Anthony P King, Nastassja Koen, Roman Kotov, Henry R Kranzler, Kristi Krebs, William S Kremen, Pei-Fen Kuan, Bruce R Lawford, Lauren A M Lebois, Kelli Lehto, Daniel F Levey, Catrin Lewis, Israel Liberzon, Sarah D Linnstaedt, Mark W Logue, Adriana Lori, Yi Lu, Benjamin J Luft, Michelle K Lupton, Jurjen J Luykx, Iouri Makotkine, Jessica L Maples-Keller, Shelby Marchese, Charles Marmar, Nicholas G Martin, Gabriela A MartÍnez-Levy, Kerrie McAloney, Alexander McFarlane, Katie A McLaughlin, Samuel A McLean, Sarah E Medland, Divya Mehta, Jacquelyn Meyers, Vasiliki Michopoulos, Elizabeth A Mikita, Lili Milani, William Milberg, Mark W Miller, Rajendra A Morey, Charles Phillip Morris, Ole Mors, Preben Bo Mortensen, Mary S Mufford, Elliot C Nelson, Merete Nordentoft, Sonya B Norman, Nicole R Nugent, Meaghan O’Donnell, Holly K Orcutt, Pedro M Pan, Matthew S Panizzon, Gita A Pathak, Edward S Peters, Alan L Peterson, Matthew Peverill, Robert H Pietrzak, Melissa A Polusny, Bernice Porjesz, Abigail Powers, Xue-Jun Qin, Andrew Ratanatharathorn, Victoria B Risbrough, Andrea L Roberts, Barbara O Rothbaum, Alex O Rothbaum, Peter Roy-Byrne, Kenneth J Ruggiero, Ariane Rung, Heiko Runz, Bart P F Rutten, Stacey Saenz de Viteri, Giovanni Abrahão Salum, Laura Sampson, Sixto E Sanchez, Marcos Santoro, Carina Seah, Soraya Seedat, Julia S Seng, Andrey Shabalin, Christina M Sheerin, Derrick Silove, Alicia K Smith, Jordan W Smoller, Scott R Sponheim, Dan J Stein, Synne Stensland, Jennifer S Stevens, Jennifer A Sumner, Martin H Teicher, Wesley K Thompson, Arun K Tiwari, Edward Trapido, Monica Uddin, Robert J Ursano, Unnur Valdimarsdóttir, Leigh Luella van den Heuvel, Miranda Van Hooff, Sanne JH van Rooij, Eric Vermetten, Christiaan H Vinkers, Joanne Voisey, Zhewu Wang, Yunpeng Wang, Monika Waszczuk, Heike Weber, Frank R Wendt, Thomas Werge, Michelle A Williams, Douglas E Williamson, Bendik S Winsvold, Sherry Winternitz, Erika J Wolf, Christiane Wolf, Yan Xia, Ying Xiong, Rachel Yehuda, Ross McD Young, Keith A Young, Clement C Zai, Gwyneth C Zai, Mark Zervas, Hongyu Zhao, Lori A Zoellner, John-Anker Zwart, Murray B Stein, Kerry J Ressler, Karestan C Koenen

## Abstract

Posttraumatic stress disorder (PTSD) genetics are characterized by lower discoverability than most other psychiatric disorders. The contribution to biological understanding from previous genetic studies has thus been limited. We performed a multi-ancestry meta-analysis of genome-wide association studies across 1,222,882 individuals of European ancestry (137,136 cases) and 58,051 admixed individuals with African and Native American ancestry (13,624 cases). We identified 95 genome-wide significant loci (80 novel). Convergent multi-omic approaches identified 43 potential causal genes, broadly classified as neurotransmitter and ion channel synaptic modulators (e.g., *GRIA1, GRM8, CACNA1E*), developmental, axon guidance, and transcription factors (e.g., *FOXP2, EFNA5, DCC*), synaptic structure and function genes (e.g., *PCLO, NCAM1, PDE4B*), and endocrine or immune regulators (e.g., *ESR1, TRAF3, TANK*). Additional top genes influence stress, immune, fear, and threat-related processes, previously hypothesized to underlie PTSD neurobiology. These findings strengthen our understanding of neurobiological systems relevant to PTSD pathophysiology, while also opening new areas for investigation.

## Introduction

Posttraumatic stress disorder (PTSD) is characterized by intrusive thoughts, hyperarousal, avoidance, and negative alterations in cognitions and mood that can become persistent for some individuals after traumatic event exposure. Approximately 5.6% of trauma-exposed adults world-wide have PTSD during their lifetimes, and rates are higher in those with high levels and certain types of trauma exposure such as combat survivors and assault victims.^1^ PTSD is a chronic condition for many, posing a substantial quality-of-life and economic burden to individuals and society.^2^

Substantial advances are being made in the understanding of PTSD biology through preclinical studies,3 many of which are focused on fear systems in the brain, and some of which are being translated to human studies of PTSD.4 Human neuroimaging studies highlight probable dysfunction in brain fear circuitry that includes deficits in top-down modulation of the amygdala by regulatory regions such as the anterior cingulate and ventromedial prefrontal cortex.^5,6^ Neuroendocrine studies have identified abnormalities in the HPA axis and glucocorticoid-induced gene expression in the development and maintenance of PTSD.^7,8^ However, many questions remain about the pathophysiology of PTSD and new targets are needed for prevention and treatment.

While twin and genetic studies demonstrated that risk of developing PTSD conditional on trauma exposure is partly driven by genetic factors,^9,10^ the specific characterization of the genetic architecture of PTSD is just emerging as very large meta-analyses of genome-wide association studies (GWAS) become available. Recent research by our workgroup – the Psychiatric Genomic Consortium for PTSD (PGC-PTSD),^11,12^ and the VA Million Veterans Program (MVP)^13^, contributed to an increased appreciation for the genetic complexity of PTSD as a highly polygenic disorder. Despite sample sizes of over 200,000 individuals, these studies identified up to 15 PTSD risk loci, which were not consistent across datasets, indicating the necessity of still larger sample sizes. In addition, these studies did not examine the X chromosome, which comprises 5% of the human genome, and may be particularly important given sex differences in PTSD prevalence.

Furthermore, GWAS to date have had limited power to identify credible treatment candidates. PTSD is also known frequently to be comorbid and genetically correlated with other mental (e.g., major depressive disorder [MDD]; attention deficit hyperactivity disorder)14 and physical health conditions (e.g., cardiovascular disease; obesity),^15-17^ but studies to date are limited in their ability to parse shared and disorder-specific loci and link them to underlying biological systems. Importantly, prior GWAS are severely limited in generalizing their findings to non-European ancestries. Recent work on polygenic risk scores (PRS) in PTSD shows potential utility of these measures in research,^16-18^ but also, vexingly, limited cross-population transferability. Without expansion to other ancestries, there is a risk that recent advances in PTSD genetics will result in the widening of research and treatment disparities. This inequity is particularly troubling in the US given the disproportionately high burden of trauma and PTSD faced by populations of African, Native, and Latin American origin.^19,20^

In the present analysis, we synthesize data from 88 studies to perform a multi-ancestry meta-analysis of GWAS data from European ancestry (EA) (N = 137,136 cases and 1,085,746 controls), African ancestry (AA) (N=11,560 cases and 39,474 controls), and Native American ancestry (LAT) (N=2,064 cases and 4,953 controls) samples, including analyses of the X chromosome. We follow-up on GWAS findings to examine global and local heritability, infer involvement of brain regions and neuronal systems using transcriptomic data, describe shared genetic effects with comorbid conditions, and use multi-omic data to prioritize a set of 43 putatively causal genes (Fig. 1). Lastly, we use this information to identify potential candidate pathways for future PTSD treatment studies. Together, these novel findings mark significant progress towards discovering the pathophysiology of trauma and stress-related disorders and inform future intervention approaches for PTSD and related conditions.

**Figure 1:**
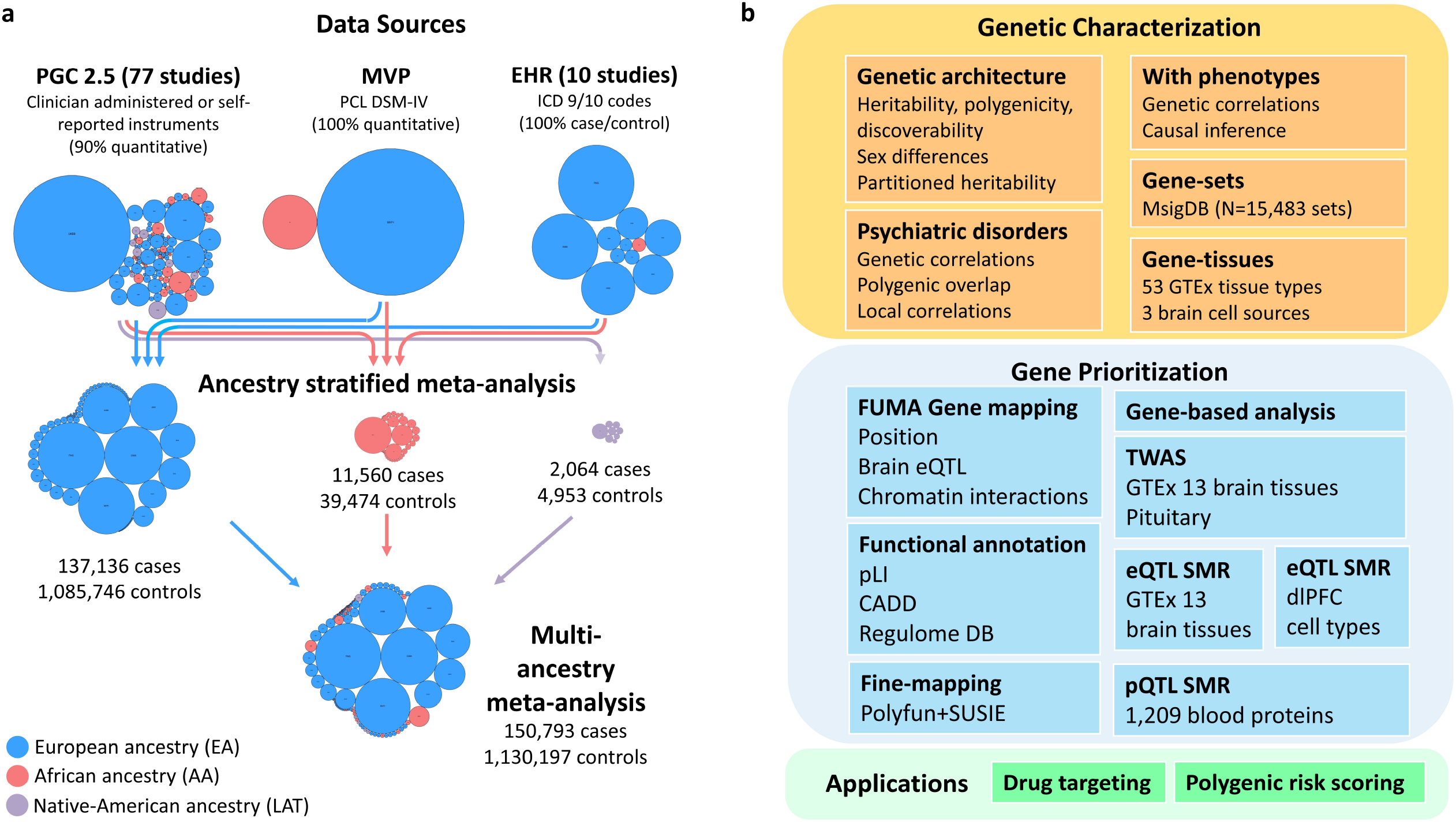
Data sources and analyses in PTSD Freeze 3. **a**, Data sources of genome-wide association studies (GWAS) included in PGC-PTSD Freeze 3. Collections of contributing studies are pictured as bubble plots where each circle represents a contributing study. Circle areas are proportional to sample size and colors indicate the ancestry classification of participants (blue, EA; red, AA; purple, LAT). Arrowed lines indicate data sources being pooled together to perform GWAS meta-analyses stratified by ancestry. **b**, Methods applied for genetic characterization of PTSD, gene prioritization analyses, and translational applications. Abbreviations: EA, European ancestry, AA, African ancestry, LAT, Native-American ancestry (Latinx); EHR, electronic health record

## Results

### Data collection and GWAS

The PGC-PTSD^21^ Freeze 3 data collection includes 1,307,247 individuals from 88 studies (Supplementary Table 1). Data in this freeze were assembled from three primary sources (Fig. 1A): PTSD studies based on clinician administered or self-reported instruments (Freeze 2.5^11,12^ plus subsequently collected studies), MVP release 3 GWASs utilizing the Posttraumatic Stress Disorder Checklist (PCL for DSM-IV),^13^ and 10 biobank studies with electronic health record (EHR)-derived PTSD status. We included 95 GWASs, including EA (N=1,222,882; effective sample size (N_eff_)=641,533), AA (N=51,034; N_eff_=42,804) and LAT (N=7,017; N_eff_=6,530) participants (Supplementary Table 2).

**Table 1.**
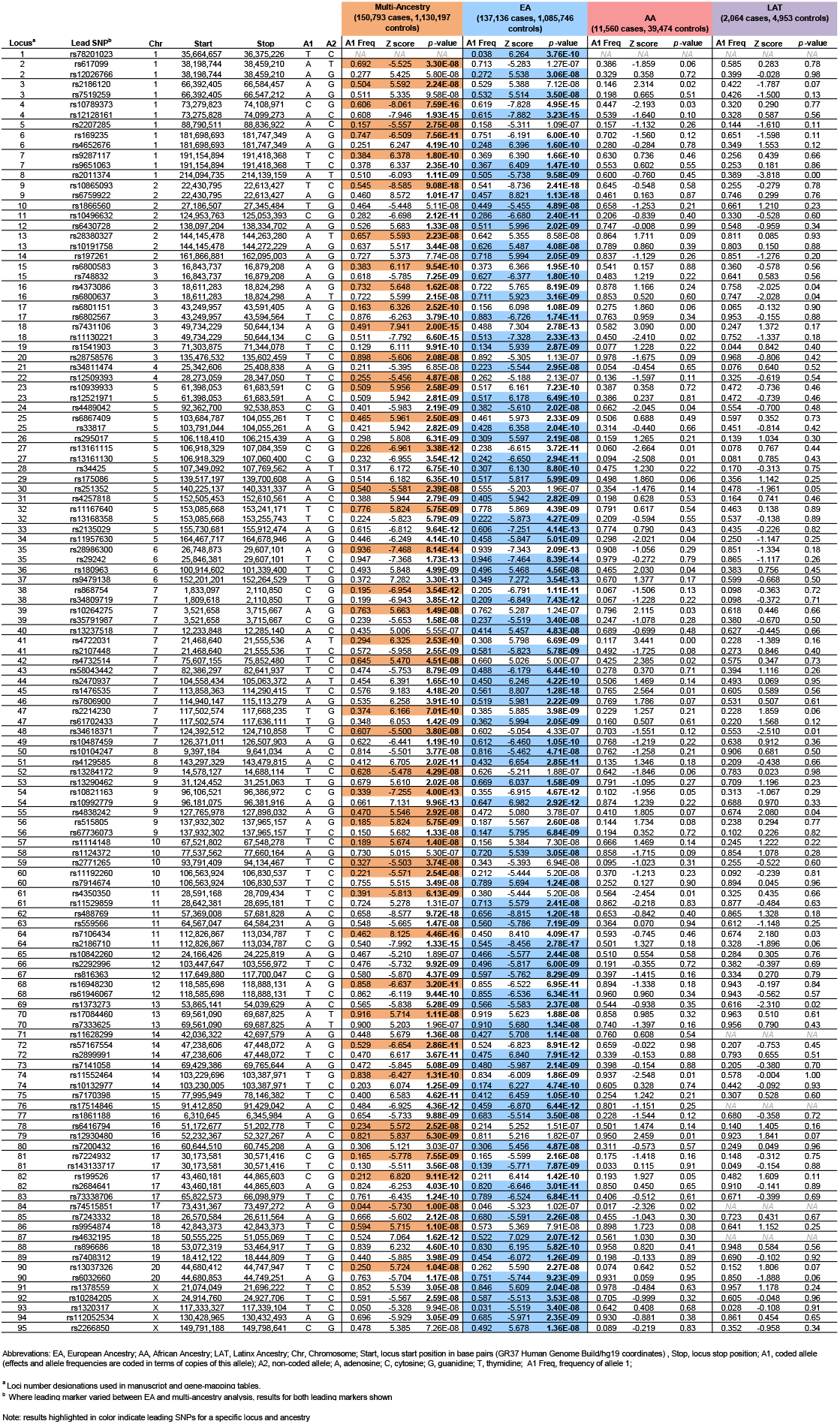
Genome-Wide Significant Loci Associated With PTSD in the Multi-Ancestry and European PGC-PTSD Freeze 3 Data.

### Comparisons of PTSD data subsets

Population, screening, and case ascertainment differences between datasets led to the assumption that there would be substantial cross-dataset variation in PTSD genetic signal. We investigated this possibility using the software MiXeR, which incorporates an LD score regression (LDSC) genetic correlation model plus an extension to estimate sets of causal variants and model their shared and unshared polygenic overlap.^22,23^ In univariate analyses, we identified comparable genetic architecture between PGC2.5 and EHR datasets in regards to h^2^_SNP_, polygenicity, and discoverability. However, the uniformly assessed MVP had a higher h^2^ SNP, was more discoverable and less polygenic than the other datasets (p < 0.05; Supplementary Table 3). Bivariate analyses identified high genetic correlations between the three subsets (r_g_ range = 0.79 - 0.87) (Extended Data Fig. 1, Supplementary Table 4). The MiXeR model did not provide a substantially better fit than the r_g_ model (all AIC < 0), indicating that there is no evidence for subset-specific genetic causal variation.

### European ancestry PTSD GWAS

Given the similarities of the PTSD subsets, we performed a sample-size weighted fixed-effects meta-analysis of GWAS. For the EA meta-analysis (137,136 cases and 1,085,746 controls), the GC lambda was 1.55, the LDSC^24^ intercept was 1.0524 (SE = 0.0097) (Supplementary Table 5), and the attenuation ratio was 0.0729 (SE=0.0134), indicating that 92.7% of the observed inflation in test-statistics was due to polygenic signal; thus artifacts produced only minimal inflation.

The EA meta-analysis identified 81 independent genome-wide significant (GWS) loci, including 5 GWS loci on the X chromosome (Extended data Fig. 2, Supplementary Figs. 1 and 2, Supplementary Table 6, regional association plots in Supplementary Data 1, forest plots in Supplementary Data 2, Supplementary Text). Relative to recent prior PTSD GWAS, 67 loci are novel^11-13^(Supplementary Table 7). No region exhibited significant effect size heterogeneity (Supplementary Fig. 3).

We next sought to gain insights into whether loci harbor multiple independent variants. While FUMA^25^ annotations reported independent lead SNPs within risk loci based on pair-wise LD (Supplementary Table 8), COJO^26^ analysis of each locus conditional on the leading variants suggested that only one locus carried a conditionally independent GWS SNP (rs3132388 on chromosome 6, p=2.86 ×10^-9^). This locus however, is in the MHC region, whose complicated linkage disequilibrium (LD) structure^27^ may not be accurately captured by reference panels.

### African and Native American ancestry PTSD GWAS meta-analyses

The AA meta-analysis included 51,034 predominantly admixed subjects (N=11,560 cases and 39,474 controls). There was minimal inflation of test statistics, with GC lambda = 1.031. No GWS loci were identified (Supplementary Fig. 4). The LAT meta-analysis was performed in 7,017 subjects (N=2,064 cases and 4,953 controls). There was minimal inflation of test statistics (GC Lambda=0.993) and no GWS loci were identified (Supplementary Fig. 5).

### Multi-ancestry GWAS meta-analysis

A multi-ancestry fixed-effects meta-analysis of EA, AA, and LAT GWAS (150,793 cases, 1,130,197 controls) identified 85 GWS loci. Compared to the EA meta-analysis, 10 loci lost GWS, while 14 previously suggestive loci (p < 5 x 10^-7^) became GWS (Fig. 2). In total, the present study identified 95 unique GWS PTSD loci between the EA and multi-ancestry meta-analyses (Table 1). Due to the complex local ancestry structure in individuals with African and Native American ancestry, which complicates LD modeling, we focused subsequent fine-mapping analyses (Fig. 1B) on data from the EA GWAS.

**Figure 2:**
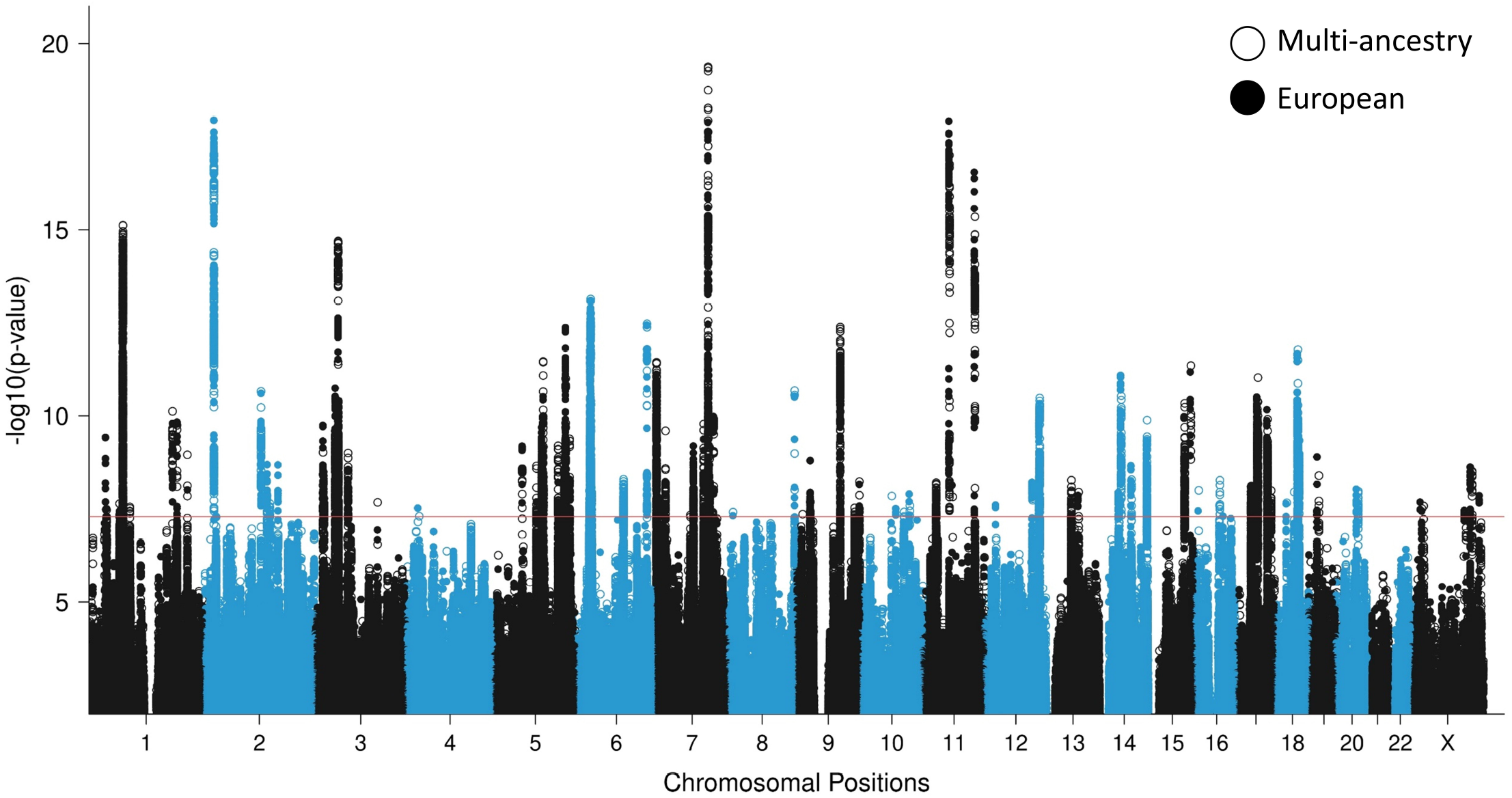
GWAS meta-analyses in European and multi-ancestry individuals identify a total of 95 PTSD risk loci. Overlaid Manhattan plots of European ancestry (EA; 137,136 cases and 1,085,746 controls) and multi-ancestry meta-analyses (150,760 cases and 1,130,173 controls), showing 81 genome-wide significant (GWS) loci for the EA (full circles) and 85 GWS loci for the multi-ancestry (hollow circles) analyses. Circle colors alternate between chromosomes, with even chromosomes colored blue and odd chromosomes colored black. The *y* axis refers to −log_10_ p-values. The horizontal red bar indicates the threshold for GWS associations (p < 5×10-8).

### Gene-mapping

To link GWS SNPs to relevant protein coding genes, we applied three gene mapping approaches implemented in FUMA: positional mapping, expression quantitative trait loci (eQTL), and chromatin interaction mapping (Supplementary Table 9). GWS SNPs within the 81 EA loci mapped to 415 protein coding genes under at least one mapping strategy. A total of 230 genes (55%) were mapped by two or more strategies, and 85 (20%) genes were mapped by all three strategies (Supplementary Fig. 6). Notably, some genes were implicated across independent risk loci by chromatin interactions/eQTL mapping, including *EFNA5, GRIA1, FOXP2, MDFIC, WSB2, VSIG10, PEBP1*, and *C17orf58*. Chromatin interaction plots are shown in Supplementary Data 3.

### Functional annotation and fine-mapping of risk loci

Functional annotations were used to gain insights into the functional role of SNPs within the 81 risk loci (Supplementary Table 10): 72 loci contained at least one SNP with Combined Annotation Dependent Depletion (CADD)^28^ scores suggestive of deleteriousness to gene function (≥12.37), 43 loci contained GWS SNPs with Regulome DB^29^ scores likely to affect binding, and 23 loci contained at least one SNP in the exon region of a gene.

To narrow the credible window of risk loci and identify potentially causal SNPs, we fine-mapped loci using Polyfun+SUSIE^30^, which identified a credible set for 67 loci. Credible set window lengths were on average 62% of the original set lengths (Supplementary Table 11) and contained a median of 23 credible SNPs (range 1-252). Only one contained a SNP with posterior inclusion probability > 0.95, a missense SNP in the exon of *ANAPC4* (rs34811474, R[CGA]>Q[CAA]; Supplementary Table 12).

### Gene-based, gene-set, and gene-tissue analyses

As an alternative approach to SNP-based association analysis, we tested the joint association of markers within genes using a gene-based association analysis in MAGMA,^31^ which is a 2-stage method that first maps SNPs to genes and then tests whether a gene is significantly associated with PTSD. The gene-based analysis identified 175 GWS genes (Supplementary Table 13, Supplementary Fig. 7). Of these, 52 were distinct from the genes implicated by the gene-mapping of individual SNPs within GWS loci. These notably include *DRD2*, which has been thoroughly investigated in the context of psychiatric disorders and is a significant GWAS locus for multiple psychiatric disorders including schizophrenia.^32^ Refer to the Supplementary Text and Supplementary Table 14 for further investigation of conditionally independent SNPs within these 52 genes.

MAGMA gene-set analysis of 15,483 pathways and gene ontology (GO) terms from MSigDB^33^ identified 12 significant GO terms. Significant terms were related to the development and differentiation of neurons (e.g. go_central_nervous_system_development, p=2.0×10^-7^), the synaptic membrane (e.g. go_postsynaptic_membrane, p=6.9×10^-7^), regulation (go_positive_regulation_of_gene_expression 1.0×10^-6^), and nucleic acid binding (p=1.52×10^-6^) (Extended Data Fig. 3, Supplementary Table 15).

MAGMA gene-tissue analysis of 54 tissue types showed PTSD gene enrichment in the brain (most notably in cerebellum, but also cortex, hypothalamus, hippocampus and amygdala) and in the pituitary, with enrichment found across all 13 examined brain regions (Extended Data Fig 4). Cell type analysis conducted in midbrain tissue data^34^ identified GABAergic neurons, GABA neuroblasts, and mediolateral neuroblast type 5 cell types as having enriched associations above other brain cell types tested (p<0.05/268) (Extended Data Fig 5). GABAergic neurons remained significant (p=4.4×10^-5^) after stepwise conditional analysis of other significant cell types.

### Multi-omic investigation of PTSD

To gain insights into which particular genes in enriched brain tissues were contributing to PTSD, we conducted a combination of a transcriptome-wide association study (TWAS)^35^ and summary based mendelian randomization (SMR) analyses^36^ using GTEx brain tissue data based on the EA GWAS summary data. TWAS identified 25 genes within 9 loci with Bonferroni-significantly different genetically regulated expression levels between PTSD cases and controls (p<0.05/14,935 unique genes tested) (Fig. 3A, Supplementary Fig. 8, Supplementary Table 16). SMR identified 26 genes within 4 loci whose expression levels were putatively causally associated with PTSD (p<0.05/9,003 unique genes tested) (Fig. 3B, Supplementary Table 17). Many of these genes have been previously implicated in PTSD37 and other psychiatric disorders (e.g., *CACNA1E, CRHR1, FOXP2, MAPT, WNT3*). Notably, the 3p21.31 (*incl*., *RBM6, RNF123, MST1R, GMPPB, INKA1*), 6p22.1 (incl., *ZCAN9* and *HCG17)* and 17q21.31 (incl., *ARHGAP27, ARL17A, CRHR1, MAPT, FAM215B, LRRC37A2, PLEKHM1*, and *SPPL2C)* regions contained >10 putative causal genes each.

**Figure 3:**
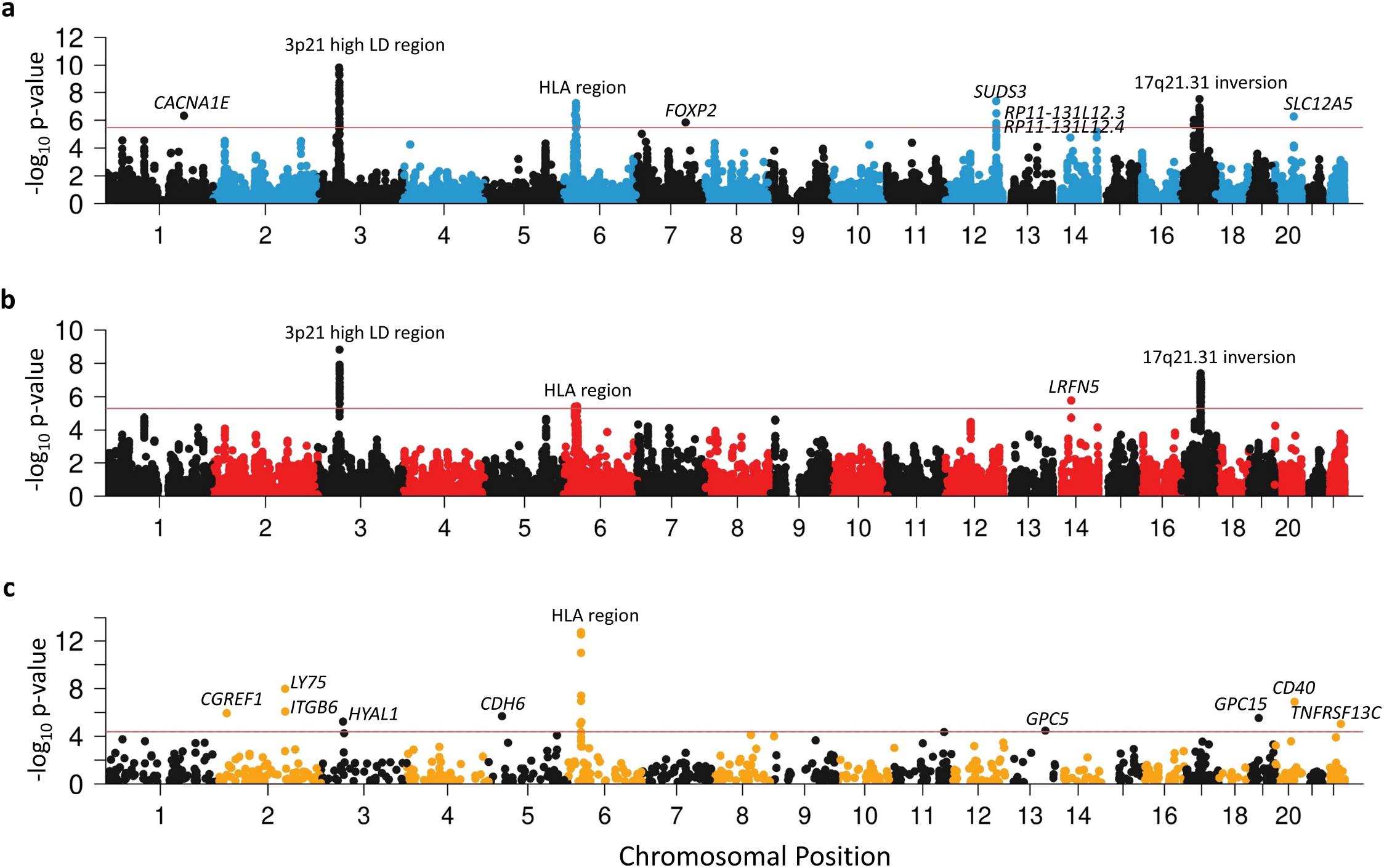
Manhattan plots of PTSD associations in multi-omic analyses. Gene expression data from 13 brain tissue types and the pituitary were used to conduct **a**, Transcriptome-wide association study (TWAS) identifying 9 loci with differential expression between PTSD cases and controls and **b**, expression quantitative trait locus summary based mendelian randomization (eQTL SMR) identifying 4 loci where gene expression has putative causal effects on PTSD. **c**, Blood protein quantitative trait locus (pQTL) SMR identify 16 blood proteins whose abundance has a putative causal effect on PTSD. The *y* axis refers to -log_10_ p-values of each respective analysis. The horizontal red bars indicate gene-wide significance (p < 0.05/14,935 for TWAS, p < 0.05/9,903 for eQTL SMR, and p < 0.05/1,209 for pQTL SMR). Significant findings are labeled.

Among the GTEx tissues with the most TWAS and SMR signals was the dorsolateral prefrontal cortex (dlPFC). To gain insight into cell type resolution, we conducted MAGMA for cell-type-specific markers of dlPFC and cell-type-specific SMR. MAGMA showed a significant enrichment of dlPFC inhibitory and excitatory neurons, but also of oligodendrocytes and oligodendrocyte precursor cells (Supplementary Table 18), while the SMR analyses identified cell-type-specific SMR signals for 8 genes (*KANSL1, ARL17B, LINC02210-CRHR1, LRRC37A2, ENSG00000262633, MAPT, ENSG00000273919, PLEKHM1*) over 3 loci (6 out of 8 from 17q21.31) and all cell types (p<0.05/1,885 unique genes tested) whose expression levels were potentially causally associated with PTSD (Supplementary Table 19). The top-gene, *KANSL1*, was significant in all cell types.

Given previously reported associations between blood-based protein levels and PTSD,^38,39^ we performed protein quantitative trait loci (pQTL) SMR^36^ analysis for PTSD using data from the UK Biobank Pharma Proteomics Project^40^ (N=54,306 samples and N=1,209 proteins). We identified 16 genes within 9 loci whose protein levels were significantly associated with PTSD (p<0.05/1,209 and p HEIDI > 0.05) (Fig. 3C, Supplementary Table 20), including members of the TNF superfamily (e.g., *CD40, TNFRSF13C*) implicating TNF-related immune activation in PTSD.

### Gene prioritization

One research objective was to identify the genes with the greatest evidence of being responsible for the associations observed at each identified PTSD locus. Following recent research methods,41 we prioritized genes based on weighted sum of evidence scores taken across the functional annotation and post-GWAS analyses (Fig. 1B). Based on the absolute and relative scores of genes within risk loci, we ranked genes into Tier 1 (greater likelihood of being the causal risk gene) and Tier 2 (prioritized over other GWAS-implicated genes, but lower likelihood than Tier 1 of being the causal gene). 75% of loci contained prioritized genes (Tier 1 or Tier 2), the remaining loci did not contain any genes over the minimum threshold of evidence (score ≥ 4) to suggest prioritization. The prioritized genes for the top 20% of loci (ranked by locus p-value) are shown in Fig 4. A complete list of scores and rankings for all 415 protein coding genes mapped to risk loci is available in Supplementary Data 4.

**Figure 4:**
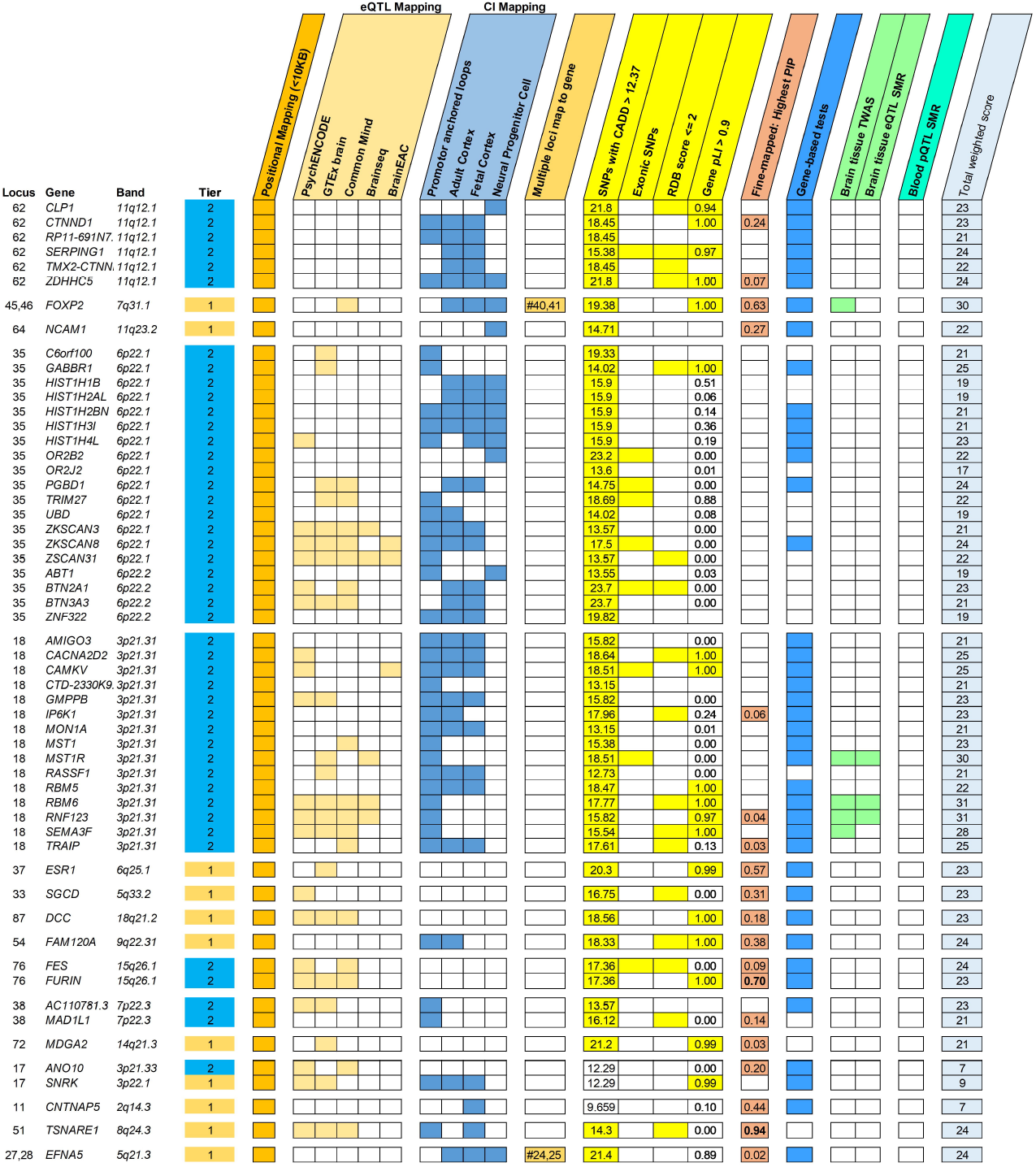
Gene prioritization in PTSD loci. Summary of evidence categories of prioritized genes (Tier 1 or 2) for the top 20% of PTSD loci (as ranked by leading SNP p-value). Locus number, prioritized genes within locus, gene locations (in terms of cytogenic band), and gene tier ranks (Tier 1, orange; Tier 2, blue) are indicated on the left. Categories of evidence are grouped and colored according to the domain they belong to. CADD scores, pLI scores and fine-mapping PIPs are written within their respective squares. The total weighted scores taken across all 9 evidence categories are shown on the rightmost squares. Abbreviations: eQTL, expression QTL; CI, chromatin interaction; CADD, combined annotation dependent depletion; RDB, regulome DB; pLI, predicted loss of impact; PIP, posterior importance probability; TWAS, transcriptome-wide association study; SMR, summary Mendelian randomization; pQTL, protein QTL;

We performed pathway enrichment analysis of the Tier 1 genes in SynGO. From Tier 1, 11 genes mapped to the set of SynGO annotated genes (*CACNA1E, DCC, EFNA5, GRIA1, GRM8, LRFN5, MDGA2, NCAM1, OLFM1, PCLO*, and *SORCS3*). Relative to other brain-expressed genes, Tier 1 genes were significantly overrepresented in the synapse (p=0.0009, qFDR=0.003), pre- and post-synapse (p=0.0086, qFDR=0.0086 and p=0.003, qFDR=0.004, respectively), and four subcategories (Extended Data Fig. 6). By contrast, there was no significant overrepresentation of genes when we applied this test to the entire set of 415 protein coding genes. Other notable Tier 1 genes included *PDE4B* related to synaptic function and TNF-related immune-regulatory genes, including *TANK* and *TRAF3*.

### Genetic architecture of PTSD

SNP-based heritability (h^2^_SNP_) estimated by LDSC was 0.053 (SE=0.002, p=6.8×10^-156^). Whereas previous reports suggested sex-specific differences in PTSD,^11^ no significant differences were found (p=0.13), and r_g_ between male and female subsets was high (r_g_=0.98,SE=0.05, p=1.2×10^-98^; Supplementary Table 5). MiXeR estimated 10,863 (SE=377) influential variants and a discoverability of 7.4×10^-6^ (SE=2.2×10-7) (Supplementary Table 3), indicating a genetic architecture comparable to other psychiatric disorders.^42^

Partitioned heritability across 28 functional categories identified enrichment in histone markers (H3K9ac peaks: 6.3 fold enrichment, SE = 1.12, p=3.11×10^-6^; H3K4me1: 1.5 fold enrichment, SE=0.14, p=3.3×10^-4^; Supplementary Table 21), and in evolutionary constrained regions across 29 Eutherians (18.37 fold enrichment, SE = 1.18, p=1.29×10-17). This is consistent with findings for multiple other psychiatric disorders, but has not been previously identified in PTSD.^42^

### Contextualization of PTSD among psychiatric disorders

We measured the genetic overlap between PTSD and other psychiatric disorders using the most recent available datasets.^32,43-52^ We observed moderate to high positive r_g_ between PTSD and other psychiatric disorders (Extended Data Fig. 7A). To gain further insights into this overlap, we used MiXeR to quantify the genetic overlap in causal variation between PTSD and bipolar disorder (BPD), MDD, and schizophrenia (SCZ) (Extended Data Fig. 7B). The strong majority (79-99%) of the variation influencing PTSD risk also influenced these disorders (Extended Data Fig. 7B, Supplementary Tables 22 and 23). Similar to r_g_, PTSD had the highest fraction of concordant effect directions with MDD (among the shared variation) (87% concordant, SE=2%), significantly higher than the directional concordance with BPD (67%, SE=1%) and SCZ (65%, SE=0.5%).

While our results indicate an overall strong r_g_ between PTSD and MDD (r_g_=0.85, SE = 0.008, p< 2×10^-16^), the correlation between PTSD and MDD varied significantly across PTSD subsets, with the most homogeneously assessed subset, MVP, showing the lowest correlation, and the biobank subset being most strongly associated (Supplementary Table 24). Further, to evaluate if specific genetic regions differ substantially from genome-wide estimates we used LAVA^53^ to estimate the local h^2^ SNP and r _g_ of PTSD and MDD across the genome, as partitioned into 2,495 approximately independent regions (Supplementary Table 25). Local h^2^ SNP was significant (P<0.05/2,495) for both PTSD and MDD in 141 regions. Of these, local r_g_ was significant (p < 0.05/141) in 40 regions, all in the positive effect direction, where the mean local rg^2^ was 0.57 (SD=0.24). In addition, we assessed the local r_g_ between PTSD and MDD specifically for the 76 autosomal GWS EA loci (Supplementary Table 26). While LAVA identified 20 significantly correlated loci (r_g_<6.58×10^-4^), there was also evidence for PTSD loci lacking evidence for correlation with MDD (Supplementary Figures 9 and 10 showcase 6 selected loci with low and high r_g_).

### Contextualization of PTSD across other phenotype domains

Considering all 1,114 traits with SNP-based heritability z>6 available from the Pan-UKB^54^ analysis, we observed Bonferroni-significant r_g_ of PTSD with 73% of them (Supplementary Table 27). Examining the extremes of estimates observed, the top positive r_g_ was with sertraline prescription (r_g_=0.88, p=3.25 x 10^-20^), a medication frequently prescribed for PTSD and other internalizing disorders^55^. Other leading associations included medication poisonings (e.g. “Poisoning by psychotropic agents” r_g_=0.88, p=3.92×10^-20^), which could support a link with accidental poisonings or self-harm behaviors.^56,57^ Converging with epidemiologic studies, there were correlations with gastrointestinal symptoms^58^ (e.g., “Nausea and vomiting” r_g_=0.80, p=2.39×10-16), mental health comorbidities^59^ (e.g., Probable Recurrent major depression (severe)” r_g_=0.87, p=1.18×10^-18^ ; “Recent restlessness” r_g_=0.86, p=4.21×10^-54^;), chronic pain^60^ (multi-site chronic pain r_g_=0.63, p=7.5×10^-301^) and reduced longevity^61-63^ (“Mother’s age at death” (r_g_=-0.51, p=7.6×10^-27^).

### Drug target and class analysis

We extended MAGMA gene-set analysis to investigate 1530 gene sets comprising known drug targets (Supplementary Table 28). We identified one drug (stanozolol, an anabolic steroid) significantly enriched for targets associated with PTSD (p=1.62×10^−5^). However, stanozolol has only two target genes in our analyses (*ESR1, JUN*), and likely reflects the strong association of *ESR1* with PTSD in gene-level analyses (p=8.94×10^-12^).

We further examined whether high-ranking drug targets were enriched for 159 drug classes defined by Anatomical Therapeutic Chemical (ATC) codes. We identified two broad classes where drugs were significantly enriched for association in drug target analyses (Supplementary Table 29). These were opioid drugs (ATC code N02A, p=2.75×10^-4^), and psycholeptics (ATC code N05, p=3.62×10^-5^), particularly antipsychotics (ATC code N05A, p=3.55×10^-7^). However, sensitivity analyses limited to drugs with 10 or more targets identified no significant drug target sets nor drug classes.

### Polygenic predictive scoring

We evaluated the predictive accuracy of PRS based on PTSD Freeze 3 in a set of MVP holdout samples (Fig. 5). In EA holdouts, risk was significantly different across the range of PTSD PRS: For example, individuals in the highest quintile of PTSD PRS had 2.4 times the relative risk of PTSD (log relative risk SE=0.032; 95%CI = [2.25, 2.56]; p=1.16×10^-167^) than individuals in the lowest quintile. PRS explained 6.6% of the phenotypic variation in PTSD (Nagelkerke’s R^2^ transformed to the liability scale at 15% population and sample prevalence), representing a major improvement over PRS based on Freeze 2. In contrast, among AA holdout samples, PRS explained only 0.9% (liability scale) of the variation in PTSD, consistent with previous work suggesting that AA PRS based on EA data lag behind in prediction.^65^

**Figure 5:**
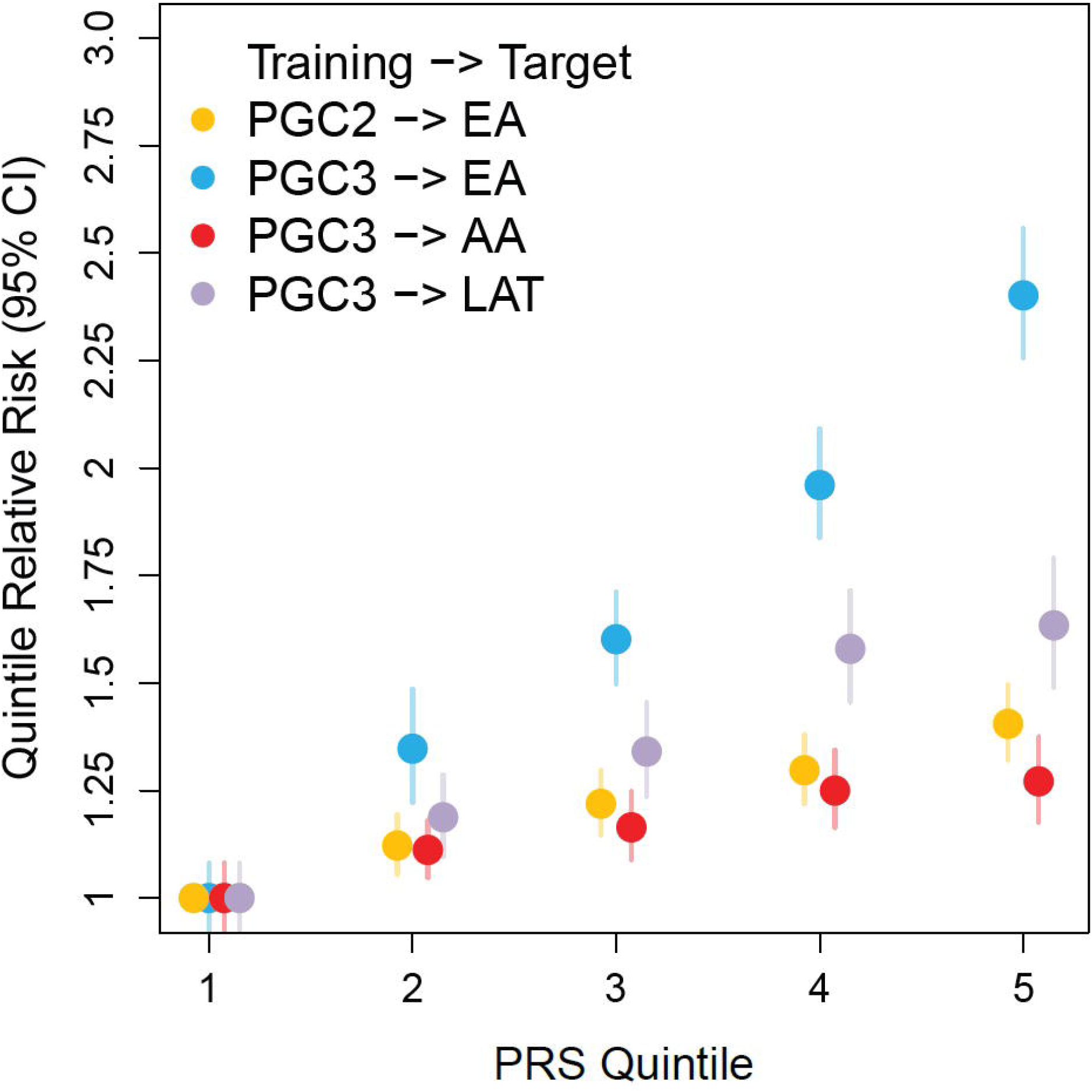
Polygenic risk score analysis for PTSD across different data sets and ancestries. PGC-PTSD Freeze 2 and Freeze 3 European ancestry (EA) based genetic risk score (PRS) predictions into independent samples of different ancestries. The *y* axis represents PTSD odds ratios relative to the lowest quintile of PRS. For EA, predictions based on Freeze 3 training data (10,334 cases and 55,504 controls; blue circles) demonstrate a significant performance increase compared to predictions based on the previous Freeze 2 training GWAS (Nievergelt et al. 2019; yellow circles). Based on Freeze 3 EA training data, EA individuals in the highest quintile of PRS have 2.8 fold the odds of PTSD relative to individuals in the lowest quintile PRS (blue circles). Lower prediction accuracies are found for individuals of African (AA; 10,151 cases and 22,420 controls; red circles)) and Native American (Latinx; LAT; 5,346 cases and 10,821 controls; purple circles) ancestries, indicating poor PRS transferability across ancestries.

## Discussion

In the largest PTSD GWAS to date we analyzed data from over one million subjects and identified a total of 95 independent risk loci across analyses, a five-fold increase over the most recent PTSD GWAS.^11-13^ Compared to previous PTSD GWAS, we confirmed 14 out of 24 loci, and identified 80 novel PTSD loci. Variant discovery in psychiatric GWAS follows a sigmoid curve, rapidly increasing once sample size passes a given threshold. This analysis passes that inflection point in PTSD,^66^ thus representing a major milestone in PTSD genetics. Moreover, by leveraging complementary research methodologies, our findings provide new functional insights and a deeper characterization of the genetic architecture of PTSD.

Tissue and cell-type enrichments revealed involvement of cerebellum, in addition to other traditionally PTSD-associated brain regions, and interneurons in PTSD risk. Structural alterations in the cerebellum are associated with PTSD67 and large postmortem transcriptomic studies of PTSD consistently reveal differential expression of interneuron markers in prefrontal cortical tissue and amygdala nuclei.^68-70^ We used a combination of TWAS and SMR to probe the causal genes operating within the enriched tissues and cell types with brain transcriptomic data. The identified signals were concentrated in some GWAS loci like 17q21.31 whose inversion region is associated with a range of psychiatric phenotypes and linked to changes in brain structure and function. *KANSL1, ARL17B, LINC02210-CRHR1* (encoding a fusion protein with CRHR1) *and LRRC37A2* were the top causal genes in both neuronal and non-neuronal cell-types. *KANSL1* plays a critical role in brain development. Furthermore, the first single cell transcriptomic study of PTSD confirmed neuronal, excitatory and inhibitory, alterations in 17q21.31 with top alterations in *ARL17B, LINC02210-CRHR1 and LRRC37A2*, while also emphasizing the involvement of immune and glucocorticoid response in neurons (Chatzinakos et al. 2023, *in press*).

Notably, although PTSD risk in epidemiological studies is higher in women than men,^71^ here we found no sex differences in heritability. Five loci on the X chromosome associated with the disorder. Our finding that the estrogen receptor (*ESR1*) gene was identified in GWAS, as well as observations of differential effects of estrogen levels on a variety PTSD symptoms,^72,73^ suggests the importance of further analyses of *ESR1* as a potential mediator of observed sex differences.

Our analyses prioritized 43 genes as Tier 1 (likely causal) based on weighted sum of evidence scores taken across the functional annotation and post-GWAS analyses. These genes can broadly be classified as neurotransmitter and ion channel synaptic plasticity modulators (e.g., *GRIA1, GRM8, CACNA1E*), developmental, axon guidance and transcription factors (e.g., *FOXP2, EFNA5, DCC*), synaptic structure and function genes (e.g., *PCLO, NCAM1, PDE4B*), and endocrine and immune regulators (e.g., *ESR1, TRAF3, TANK*). Furthermore, many additional genes with known function in related pathways were genome-wide significant and met Tier 2 prioritization criteria (e.g., *GABBR1, CACNA2D2, SLC12A5, CAMKV, SEMA3F, CTNND1, and CD40)*. Together, these top genes show a remarkable convergence with neural network, synaptic plasticity and immune processes implicated in psychiatric disease. Furthermore, *CRHR1*,^*74,75*^ *WNT3*,^*76,77*^ and *FOXP2*,^*78,79*^ among other genes, are implicated in preclinical and clinical work related to stress, fear and threat-processing brain regions thought to underlie the neurobiology of PTSD. These findings largely support existing mechanistic hypotheses, and it will be important to examine how these genes and pathways function in already identified stress-related neural circuits and biological systems. Furthermore, while some of the prioritized genes are largely within pathways currently indicated in PTSD, many of the specific genes and encoded proteins were not previously established and warrant further investigation. Additionally, many genes and noncoding RNAs were not previously identified in any psychiatric or stress-related disorder, and offer an important road map for determining next steps in understanding new mechanisms of vulnerability for posttraumatic psychopathology. Future mechanistic research in preclinical models should examine whether targeting combinations of these genes, for example via polygenic targeting, epigenetic, or knockdown approaches, would have increased power in regulating stress, fear, cognitive dysfunction or other symptoms and behaviors seen in PTSD.

We observed highly shared polygenicity between PTSD and other psychiatric disorders, albeit with effect discordance across the shared variation. In particular, in some cases we found that the genetic correlation of PTSD with MDD is as high or higher than genetic correlations between different cohorts, with different measures, of PTSD. Thus, our findings corroborate the hypothesis that psychiatric disorders share a substantial amount of risk variation but are differentiated by disorder-specific effect sizes.^43^ Across the disorders we assessed, the correlation between PTSD and MDD was highest, in agreement with existing genetic multi-factor models of psychopathology that consistently cluster these disorders together^42,80^ and concordant with their epidemiologic co-morbidity.^81^ Evaluation of local patterns of heritability and genetic correlation however indicates disorder-specific risk variation, which will serve as targets for follow-up in cross-disorder investigations. We note that as GWAS of psychiatric traits grow in size and power, the field is seeing relatively strong genetic correlations among these traits, as well as with other behavioral and medical traits. This likely reflects, in part, the reality that there is substantial shared genetic variance among these traits, while not excluding the consistent observations that: (1) these traits do vary considerably in the magnitude of their genetic correlations, and (2) local genetic correlations reveal even greater genetic heterogeneity among these traits than global genetic correlations alone would lead us to believe. Finally, while PTSD is the most well-understood psychiatric outcome of trauma exposure, it is well documented that trauma is a risk factor for many different psychiatric disorders, with perhaps depression as the highest risk. Thus these shared areas of overlap may represent general trauma vulnerability as well.

Despite the high level of overall correlation between PTSD and depression, we also note certain areas of clear distinction. When we examined local genetic correlations between PTSD and depression within all significant loci from the EA PTSD GWAS, we found that there were some regions with significant local heritability for PTSD but not depression, suggestive of PTSD-specific signals. In contrast, we also find other regions with clear shared signals showing local correlation across depression and PTSD, indicating that we have the power to detect shared and distinct local heritability. Together these findings suggest several PTSD-specific loci worthy of further investigation.

Further identification of PTSD genetic loci will provide therapeutic insights.^89^ We explored whether genes targeted by specific drugs (and drug classes) were enriched for GWAS signal. These analyses provided tentative support for antipsychotics and opioid drugs – known psychiatric drug classes – and were driven by gene-wise associations with *DRD2* (antipsychotics) and *CYP2D6* (opioids). Atypical antipsychotics may have efficacy in treating severe PTSD, but otherwise their use is not supported.^90^ Similarly, whereas some observational studies find that chronic opioid use worsens PTSD outcomes,^91^ there is preclinical work motivating the further study of opioid subtype-specific targeting (e.g., partial MOR1 agonism, κ-type opioid receptor [KOR1] antagonism) in the treatment of comorbid PTSD and opioid use disorders.^92^ Analyses in better-powered datasets may identify drug repositioning opportunities and could use the predicted effect of associated variants on gene expression to indicate whether drug candidates would be beneficial or contraindicated in people with PTSD.

In summary, we reported 81 loci associated with PTSD in a EA meta-analysis, and 85 loci when expanding to trans-ancestry analyses. While these results represent a milestone in PTSD genetics and point to exciting potential target genes, further investment into data collection from underrepresented populations of diverse ancestries is needed for identification of additional risk variants and to generate equitable and more robust PRS.

## Methods

### Participants and studies

PTSD assessment and DNA collection for GWAS analysis were performed by each study following their protocols. A description of the studies included and the phenotypic and genotyping methods for each study Supplementary Methods and Supplementary Table 1. We complied with relevant ethical regulations for human research. All subjects provided written informed consent and studies were approved by the relevant institutional review boards and the UCSD IRB (protocol #16097×).

### EHR Studies

A total of 10 EHR-based cohorts (not including the MVP, which also contributed data) provided GWAS summary statistics. These cohorts consisted of four US-based sites (Vanderbilt University Medical Center’s BioVu, the Mass General Brigham Biobank, Mount Sinai’s BioMe, and Mayo Clinic’s MayoGC) and six non-US sites (iPSYCH from Denmark, FinnGen, HUNT Study from Norway, STR-STAGE from Sweden, UK Biobank, and Estonia Biobank). More details on procedures at each site are provided in the Supplementary Text. At each site, a broad definition of PTSD cases was defined based on patients having at least 1 PTSD or other stress disorder code (see Supplementary Text for the list of corresponding ICD-9 and 10 codes). All other patients without such a code were defined as controls. From a total of 817,181 participants across all cohorts, this case definition resulted in 78,687 cases based on the broad definition (9.6%).

### Data assimilation

Subjects were genotyped on Illumina (N=84 studies) or Affymetrix genotyping arrays (N=5 studies) (Supplementary Table 1). Studies which provided direct access to pre-quality control genotype data (N=64 studies) were deposited on the LISA server for central processing and analysis by the PGC-PTSD analyst. Studies with data sharing restrictions (N=24 studies) were processed and analyzed following their own site-specific protocols (Supplementary Table 28), and shared GWAS summary statistics for inclusion in meta-analysis.

### Genotype quality control and imputation

Genotype data was processed separately by study. For genotype data processed by the PGC-PTSD analyst, quality control was performed using a uniform set of criteria, as implemented in the RICOPILI^93^ pipeline version 2019_Oct_15.001. Modifications were made to the pipeline to allow for ancestrally diverse data and are noted where applicable. Quality control: using SNPs with call rates >95%, samples were excluded with call rates <98%, deviation from expected inbreeding coefficient (fhet < −0.2 or >0.2), or a sex discrepancy between reported and estimated sex based on inbreeding coefficients calculated from SNPs on X chromosomes. SNPs were excluded for call rates <98%, a > 2% difference in missing genotypes between cases and controls, or being monomorphic. Hardy-Weinberg equilibrium was calculated within only in the largest homogenous ancestry group found in the data. SNPs with a Hardy-Weinberg equilibrium P-value < 1 × 10−6 in controls were excluded.

After quality control, datasets were lifted over to the GRCh37/hg19 human genome reference build. SNP name inconsistencies were corrected, and genotypes were aligned to the strand of the imputation reference panel. Markers with non-matching allele codes or with excessive MAF difference (> 0.15) with the selected corresponding population in the reference data were removed. The pipeline was modified so that only the largest homogenous ancestry group in the data was used for the calculation of MAF. For ambiguous markers, strand was matched by comparing allele frequencies: if a strand flip resulted in a lower MAF difference between the study and the reference data, the strand was flipped. Ambiguous markers with high MAF (> 0.4) were removed. The genome was broken into 132 approximately equally sized chunks. For each chunk, genotypes were phased using Eagle v2.3.5 and phased genotypes were imputed into the Haplotype Reference Consortium panel^94^ using minimac3. Imputed datasets were deposited with the PGC DAC and are available for approved requests.

Studies with data sharing restrictions followed similar criteria for quality control, as detailed in Supplementary Table 28 and in the references in the supplemental material. Studies were imputed to either the 1000G phase 3, HRC, SISu panel, or a composite panel. GWAS summary data were lifted to the GRCh37 reference build where required. As differences in the imputation panels and genome reference build can result in SNP-level discrepancies between datasets, each set of summary data was examined for correspondence to the centrally imputed data. Multi-allelic SNPs and SNPs with non-matching allele codes were excluded. Stand ambiguous SNPs with high MAF difference (>20%) from the average frequency calculated the PGC-PTSD data were flagged and examined for strand correspondence.

### Ancestry determination

For studies where the PGC analyst had genotype data access, ancestry was determined using a global reference panel11 using SNPweights^95^. The ancestry pipeline was shared with external sites to be utilized where possible. Subjects were placed into three large groupings: European and European Americans (EA; subjects with ≥90% European ancestry), African and African-Americans (AA; subjects with ≥5% African ancestry, <90% European ancestry, <5% East Asian, Native American, Oceanian, and Central-South Asian ancestry; and subjects with ≥50% African ancestry, <5% Native American, Oceanian, and <1% Asian ancestry), and Latinos (LAT; subjects with ≥5% Native American ancestry, <90% European, <5% African, East Asian, Oceanian, and Central-South Asian ancestry). Native Americans (subjects with ≥60% Native American ancestry, <20% East Asian, <15% Central-South Asian, and <5% African and Oceanian ancestry) were grouped together with LAT. All other subjects were excluded from the current analyses. For the MVP cohort, ancestry was determined using standard principal components analysis approach where MVP samples were projected onto a PC space made from 1000 Genomes Phase 3 (KGP3) samples with known population origins (EUR, AFR, EAS, SAS, and AMR populations). EHR cohorts followed their own site-specific ancestry classification protocols.

### GWAS

GWAS was performed with stratification by ancestry group and study. Strata were only analyzed if they had a minimum of 50 cases and 50 controls, or alternatively 200 subjects total. Where noted (Supplementary Table 2), small studies of similar composition were jointly genotyped so that they could be analyzed together as a single unit. For GWAS, the association between each SNP and PTSD was tested under an additive genetic model, using a regression model appropriate to the data structure. The statistical model, covariates, and analysis software used to analyze each study is detailed in Supplementary Table 30. In brief, studies of unrelated subjects with continuous (case/control) measures of PTSD were analyzed using PLINK 1.9,^96^ using a linear (logistic) regression model which included 5 PCs as covariates. For studies that retained related subjects, analyses were performed using methods that account for relatedness. QIMR was analyzed using GEMMA^97^ v0.96, including the first five PCs as covariates. RCOG was analyzed using the generalized disequilibrium test.^98^ UKBB was analyzed using Bolt-LMM^99^ including 6 PCs, and batch and center indicator variables as covariates. VETS was analyzed using BOLT-LMM including 5 PCs as covariates. EHR based studies that included related subjects were analyzed using saddle point approximation methods to account for case/control imbalances. AGDS and QIM2 were analyzed using SAIGE^100^ including 4 PCs and study specific covariates. BIOV was analyzed using SAIGE including 10 PCs and age of record. ESBB, FING, HUNT, and SWED were analyzed using SAIGE including 5 PCs. UKB2 was analyzed using REGENIE^101^ including 6 PCs, assessment center, and genotyping batch covariates. GWAS was additionally performed stratified by sex. For the X chromosome analysis, sex was added as a covariate.

### Meta-analysis

Sample-size weighted fixed-effects meta-analysis was performed with METAL.^102^ Within each dataset and ancestry group, summary statistics were filtered to MAF ≥1% and imputation information score ≥0.6. Meta-analyses were performed within the EA, AA, and LAT ancestry groups. A multi-ancestry meta-analysis was performed as the meta-analysis of the three meta-analyses. Genome-wide significance was declared at P < 5 × 10^−8^. Heterogeneity between datasets was tested with the Cochran test. Markers with summary statistics in less than 80% of the total effective sample size were removed from meta-analyses. LDSC^24^ intercept was used to estimate inflation of test statistics related to artifacts rather than genetic signal. The proportion of inflation of test statistics due to the actual polygenic signal (rather than other causes such as population stratification) was estimated as 1-(LDSC intercept-1)/(mean observed Chi-square-1).

### Regional Association Plots

Regional association plots were generated using LocusZoom^103^ with 1.5MB windows around the index variant (unless the locus region was wider than 1.5MB, in which case it was the locus region plotted plus an additional buffer to include data up to the recombination region). The LD patterns plotted were based on the 1000 Genomes Phase 3 reference data,^104^ where a sample ancestry appropriate subpopulation (EUR, AFR, or AMR) was used.

### Conditional analysis of significant loci

To determine if there were independent significant SNPs within risk loci, GCTA Conditional and Joint Analysis^26^ was performed. Stepwise selection was performed using the --cojo-slct option and default parameters, where UKBB European genotype data was used to model LD structure.

### SNP heritability

h^2^ SNPof PTSD was estimated using LDSC. LD scores calculated within KGP3 European populations (https://data.broadinstitute.org/alkesgroup/LDSCORE/) were used for the input. Analyses were limited to HapMap 3 SNPs, with the MHC region excluded (chr6: 26–34 million base pairs). SNP-based heritability was also calculated as partitioned across 28 functional annotation categories (https://data.broadinstitute.org/alkesgroup/LDSCORE/) using stratified LDSC.^105^

### Comparisons of Genetic Architecture

We used univariate MiXeR (version 1.3)^22,23^ to contrast the genetic architecture of phenotypes. MiXeR estimates SNP-based heritability and two components that are proportional to heritability: the proportion of non-null SNPs (polygenicity), and the variance of effect sizes of non-null SNPs (discoverability). MiXeR was applied to GWAS summary statistics under the default settings with the supplied European ancestry LD reference panel. The results reported for the number of influential variants reflects the number of SNPs necessary to explain 90% of SNP-based heritability. Bivariate MiXeR was used to estimate phenotype-specific polygenicity and the shared polygenicity between phenotypes. Goodness of fit of the MiXeR model relative to simpler models of polygenic overlap was assessed using AIC values. Heritability, polygenicity and discoverability estimates were contrasted between datasets using the z-test.

### Local genetic correlation analyses

Local h^2^ SNP and r _g_ between PTSD and MDD^50^ were estimated using LAVA.^53^ KGP3 European data was used as the LD reference. Local h^2^_SNP_ and r_g_ were evaluated across the genome, as partitioned into 2,495 approximately equally sized LD blocks. Local rg was only evaluated for loci where local heritability was significant (P < 0.05/2,495) in both phenotypes. Significance of local rg was based on Bonferroni adjustment for the number of r_g_ evaluated.

### Polygenic risk scores (PRS)

PRS were calculated in ancestry-stratified MVP holdout samples, based on the EA Freeze 3 PTSD GWAS. GWAS summary statistics were filtered to common (MAF >1%), well-imputed variants (INFO > 0.8). Indels and ambiguous SNPs were removed. PRS-CS^106^ was used to infer posterior effect sizes of SNPs, using the KGP3 EUR based LD reference panel supplied with the program, with the global shrinkage parameter set to 0.01, 1,000 MCMC iterations with 500 burn-in iterations, and the Markov chain thinning factor set to 5. PRS were calculated using the --score option in PLINK 1.9, using the best-guess genotype data of target samples, where for each SNP the risk score was estimated as the posterior effect size multiplied by the number of copies of the risk allele. PRS was estimated as the sum of risk scores over all SNPs. PRS were used to predict PTSD status under logistic regression, adjusting for 5 PCs. The proportion of variance explained by PRS for each study was estimated as the difference in Nagelkerke’s R*2* between a model containing PRS plus covariates and a model with only covariates.

### Functional Mapping and Annotation

We used the SNP2GENE module in FUMA^25^ v1.4.1 (https://fuma.ctglab.nl) to annotate and visualize GWAS results. The complete set of parameters used for FUMA analysis are shown in the Supplementary Text. Independent genomic risk loci were identified (r^2^ < 0.6, calculated using ancestry-appropriate KGP3 reference genotypes). SNPs within risk loci were mapped to protein coding genes using positional mapping (10KB window), eQTL mapping (GTEx v8 brain tissue,^107^ BRAINEAC,^108^ and CommonMind^109^ data sources), and chromatin interaction mapping (PsychENCODE^110^ and HiC^111,112^ of brain tissue types) methods. Chromatin interactions and eQTLs were plotted in circos plots. SNPs were annotated to functional annotation databases including ANNOVAR,^113^ CADD,28 and RegulomeDB.^29^

### Novelty of risk loci

The start and stop positions of independent risk loci were assessed for positional overlap with existing PTSD loci^11-13^. Loci were declared novel if their boundaries did not overlap with a variant reported significant in prior GWAS.

### MAGMA gene-based and gene-set analyses

Gene-based association analyses were conducted using MAGMA^31^ v1.08. SNPs were positionally mapped (0KB window) to 19,106 protein-coding genes. The SNP-wide mean model was used to derive gene-level p-values, with an ancestry appropriate KGP3 reference panel was used to model LD. Significance was declared based on Bonferroni adjustment for the number of genes tested. Gene-based association statistics were used in MAGMA for gene-set and gene-property analyses. Gene-set analysis used the MsigDB^33^ version 7.0 including 15,483 curated gene-sets and gene-ontology (GO) terms. Gene-property analysis of tissues and tissue subtypes was performed using GTEx v8 expression data, with adjustment for the average expression of all tissues in the dataset. To evaluate cell type specific enrichment, the FUMA cell type module was used, selecting 12 datasets related to the brain (full list in Supplementary Text). Finally, MAGMA was used to estimate the enrichment of dlPFC cell types in PTSD risk based on the DER21 marker gene list from PsychEncode Consortium Phase 1 resource release.^110^

### GWAS Fine-mapping

Polygenic functionally informed fine-mapping (Polyfun)^30^ software was used to annotate our results data with per-SNP heritabilities, as derived from a meta-analysis of 15 UK Biobank traits. PTSD risk loci were fine-mapped using SUSIE,^114^ with these per SNP heritabilities used as priors, pre-computed UKB based summary LD information used as the LD reference, and locus start and end positions as determined by FUMA. The SUSIE model assumed a maximum of two causal variants.

### Expression quantitative trait loci (eQTL) and blood protein quantitative trait loci (pQTL) analyses

To test for a joint association between GWAS summary statistics SNPs and eQTL, the SMR method,^36^ a Mendelian randomization approach, was used. SMR software (version 1.03) was run using the default settings. The European samples of the 1000G were used as a reference panel. Bonferroni multiple-testing correction was applied on SMR *P-value (P*_SMR_). Moreover, a post-filtering step was applied by conducting heterogeneity in dependent instruments (HEIDI) test. The HEIDI test distinguishes the causality and pleiotropy models from the linkage model by considering the pattern of associations using all SNPs significantly associated with gene expression in the cis-eQTL region. The null hypothesis is that a single variant is associated with both trait and gene expression, while the alternative hypothesis is that trait and gene expression are associated with two distinct variants. Finally, gene-trait associations based on SMR-HEIDI were defined as the ones for which *P*_SMR_ met the Bonferroni significance threshold and had *P*_HEIDI_>0.05. We conducted a combination of SMR and HEIDI based on GTEx project latest (version 8) multi-tissue cis-eQTL databases^107^ from 13 brain regions and pituitary tissue that showed significant enrichment in MAGMA/FUMA analyses (see above). We also used cell-type-specific eQTLs in dlPFC for SMR analyses.^115^ Finally, we used a blood UK Biobank pQTLs database of 1,463 plasma proteins^40^ relying on a very large population (54,306) for SMR/HEIDI analysis to evaluate biomarker potential.

### Brain focused TWAS

JEPEGMIX2-P^116^ software with default settings was used to conduct TWAS on 13 brain regions and pituitary tissue that showed significant enrichment in MAGMA/FUMA analyses using our PEC-DLPFC GReX model. JEPEGMIX2-P was applied on GWAS summary statistics to estimate gene-trait associations. This method was preferable since it relied on a covariance matrix based on 33K samples compared to other TWAS methods which use less than 3k samples.^117^ To determine significance, a Bonferroni correction threshold for the unique number of genes tested was applied) P < 0.05/14,935). As a less conservative approach, we also applied FDR at a q value threshold of 0.05.

### Gene prioritization

Genes within risk loci were prioritized following the general approach previously described.^41^ Genes were given prioritization scores based on the weighted sum of evidence across all evidence categories: FUMA positional, eQTL, and CI mapping, variant and gene annotation scores (CADD, predicted loss of impact [pLI], and RDB scores), positional overlap in fine-mapping, significance in gene-based analyses, brain tissue TWAS, eQTL SMR, and pQTL SMR. Weights for each evidence category are provided in Supplementary Table 31. Within a given locus, the evidence scores were compared across genes to identify the most likely causal gene. Genes with scores >=4 were ranked as either Tier 1 (greater likelihood of being the causal risk gene) or Tier 2 (lower likelihood of being the causal risk gene) and genes with scores < 4 were left unranked. The ranking algorithm is as follows: For a given locus, if there was a gene whose evidence score >= 4 and this gene’s score was > 20% higher than all other genes in the locus, it was ranked as a Tier 1 gene (greater likelihood of being the causal risk gene). Within a locus with a Tier 1 gene, other genes with scores between 20% and 50% lower than the Tier 1 gene were labeled as Tier 2. For loci without a Tier 1 gene, all genes with scores >= 4 that were within 50% of the leading gene were ranked as Tier 2.

### SynGO

PTSD related genes were tested for overrepresentation among genes related to synaptic terms in the SynGO^118^ web interface (https://www.syngoportal.org/). Brain expressed genes were selected as the background list for the overrepresentation tests. SynGO terms with FDR q < 0.05 were considered as being overrepresented.

### Drug Targeting Analyses

Following a previously described approach,^119^ we analyzed the enrichment of gene-level associations with PTSD in genes targeted by individual drugs. We then examined the enrichment of specific drug classes among these drug target associations. We obtained gene-level associations using MAGMA^31^ v1.08. Variant-level associations were converted to gene-level associations using the “multi=snp-wise” model, which aggregates Z scores derived from the lowest and the mean variant-level P value within the gene boundary. We set gene boundaries 35 kilobases upstream and 10 kilobases downstream of the transcribed regions from build 37 reference data (National Center for Biotechnology Information, available at https://ctg.cncr.nl/software/magma).

We performed drug target analysis using competitive gene-set tests implemented in MAGMA. Drug target sets were defined as the targets of each drug from: the Drug–Gene Interaction database DGIdb v.4.2.0,^120^ the Psychoactive Drug Screening Database Ki DB,^121^ ChEMBL v27,^122^ the Target Central Resource Database v6.7.0,^123^ and DSigDB v1.0,^124^ all downloaded in October 2020. We additionally used the drug target sets to identify targets of drugs of interest from gene-based analyses.

We grouped drugs according to the Anatomical Therapeutic Chemical class of the drug.^119^ Results from the drug target analysis were ranked, and the enrichment of each class in the drug target analysis was assessed with enrichment curves. We calculated the area under the enrichment curve and compared the ranks of drugs within the class to those outside the class using the Wilcoxon Mann-Whitney test. Multiple testing was controlled using a Bonferroni-corrected significance threshold of P < 3.27 × 10^−5^ for drug target analysis and P < 4.42 × 10^−4^ for drug class analysis, accounting for 1530 drug sets and 113 drug classes tested.

We initially limited drug target analyses to drugs with two or more targets. However, results suggested this low limit may lead to false positive findings. As a sensitivity analysis, we further limited these analyses to drugs with 10 or more targets. Multiple testing was controlled using a Bonferroni-corrected significance threshold of P < 5.42 × 10^−5^ for drug target analysis and P < 7.94 × 10^−4^ for drug class analysis, accounting for 923 drug sets and 63 drug classes tested.

### Genetic correlations and causal associations with other phenotypes

Using LDSC, we assessed the r_g_ of PTSD derived from the PGC meta-analysis conducted in EUR cohorts with traits available from the Pan-UKB analysis conducted in EUR samples. Details regarding the Pan-UKB analysis are available at https://pan.ukbb.broadinstitute.org/. Briefly, Pan-UKB genome-wide association statistics were generated using the SAIGE and including a kinship matrix as a random effect and covariates as fixed effects. The covariates included age, sex, age x sex, age^2^, age^2^ x sex, and the top-10 within-ancestry principal components. We limited our analysis to data derived from UKB participants of European descent (N=420,531) because of the limited sample size available in the other ancestry groups. Initially, we calculated SNP-based heritability of phenotypes available from Pan-UKB, retaining only those with SNP-based heritability z>6 (Supplemental Table 25) as recommended by the developers of LDSC.^125^ To define traits genetically correlated with PTSD, we applied a Bonferroni correction accounting for the number of tests performed.

## Data availability

Summary statistics for PGC2.5 will be made available upon publication via the PGC (https://pgc.unc.edu/for-researchers/download-results/). Access to study level summary statistics and genotype data can be applied for by using the PGC data access portal (https://pgc.unc.edu/for-researchers/data-access-committee/data-access-portal/). Summary statistics for MVP are available from dbGAP (accession id phs001672.v3.p1) to qualified researchers. EHR dataset summary statistics availability follows the policies of the individual contributing cohorts.

## Code availability

Analysis code is made available in a public repository (https://github.com/nievergeltlab/freeze3_gwas).

## Acknowledgements

Major financial support for the PTSD-PGC was provided by the Cohen Veterans Bioscience, Stanley Center for Psychiatric Research at the Broad Institute, and the National Institute of Mental Health (NIMH; R01MH106595, R01MH124847, R01MH124851).

Statistical analyses were carried out on the NL Genetic Cluster computer (URL) hosted by SURFsara. Genotyping of samples was supported in part through the Stanley Center for Psychiatric Genetics at the Broad Institute of MIT and Harvard. This research has been conducted using the UK Biobank resource under application number 41209. This work would not have been possible without the contributions of the investigators who comprise the PGC-PTSD working group, and especially the more than 1,307,247 research participants worldwide who shared their life experiences and biological samples with PGC-PTSD investigators.

We would like to thank Allison E. Aiello, Bekh Bradley, Aarti Gautam, Rasha Hammamieh, Marti Jett, Michael J. Lyons, Douglas Maurer, Matig R. Mavissakalian, and the late Christopher R. Erbes and Regina E. McGlinchey for their contributions to this study.

For the purposes of open access, the author has applied a Creative Commons Attribution (CC BY) license to any Accepted Author Manuscript version arising from this submission.

## Author Contributions

PGC-PTSD writing group: E.G.A., S.-A.B., C.-Y.C., K.W.C., J.R.I.C., N.P.D., L.E.D., K.C.K., A.X.M., R.A.M., C.M.N., R.P., K.J.R., and M.B.S.

Study PI or co-PI: A.B.A., S.B. Andersen, P.A.A., A.E.A.-K., S.B. Austin, E.A., D.B., D.G.B., J.C.B., S. Belangero, C. Benjet, J.M.B., L.J.B., J.I.B., G.B., R.B., A.D.B., J.R.C., C.S.C., L.K.B., J.D., D.L.D., T.d-C, K.D., G.D., A.D.-K., N.F., L.A.F., A.F., N.C.F., B.G., J.G., E.G., C.F.G., A.G.U., M.A.H., A.C.H., V.H., I.B.H., D.M.H., K. Hveem, M. Jakovljevic, A.J., I.J., T.J., K.-I.K., M.L.K., R.C.K., N.A.K., K.C.K., R.K., H.R.K., W.S.K., B.R.L., K.L., I.L., B.L., C.M., N.G.M., K.A.M., S.A.M., S.E.M., D.M., W.P.M., M.W.M., C.P.M., O.M., P.B.M, E.C.N., C.M.N., M.N., S.B.N., N.R.N., P.M.P., A.L.P., R.H.P., M.A.P., B.P., A.P., K.J.R., V.R., P.R.B., K.R., H.R., G.S., S. Seedat, J.S. Seng, A.K.S., S.R.S., D.J.S., M.B.S., R.J.U., U.V., S.J.H.V.R., E.V., J.V., Z.W., M.W., H.W., T.W., M.A.W., D.E.W., C.W., R.M.Y., H.Z., L.A.Z., and J.Z.

Obtained funding for studies: A.B.A., P.A.A., A.E.A.-K., S.B. Austin, J.C.B., S. Belangero, C. Benjet, J.M.B., L.J.B., G.B., A.D.B., C.S.C., J.D., T.d-C, A.F., N.C.F., J.D.F., C.E.F., E.G., C.F.G., M.H., M.A.H., A.C.H., V.H., I.B.H., D.M.H., K. Hveem, T.J., N.A.K., K.C.K., R.K., W.S.K., B.R.L., B.L., C.M., N.G.M., K.A.M., S.A.M., S.E.M., J.M., W.P.M., M.W.M., C.P.M., O.M., P.B.M, E.C.N., C.M.N., M.N., N.R.N., H.K.O., M.A.P., B.P., K.J.R., B.O.R., G.S., M.S., A.K.S., S.R.S., M.H.T., R.J.U., U.V., E.V., J.V., Z.W., M.W., T.W., M.A.W., D.E.W., R.Y., R.M.Y., and L.A.Z.

Clinical: C.A., P.A.A., E.A., D.B., D.G.B., J.C.B., L.B., L.J.B., E.A.B., R.B., A.C.B., A.D.B., S. Børte, L.C., J.R.C., K.W.C., L.K.B., M.F.D., T.d-C, S.G.D., G.D., A.D.-K., N.F., N.C.F., J.D.F., C.E.F., S.G., E.G., A.G.U., S.B.G., L.G., C.G., V.H., D.M.H., M. Jakovljevic, A.J., G.D.J., M.L.K., A.K., N.A.K., N.K., R.K., W.S.K., B.R.L., L.A.M.L., K.L., C.E.L., B.L., J.L.M.-K., S.A.M., P.B.M, H.K.O., P.M.P., M.S.P., E.S.P., A.L.P., M.P., R.H.P., M.A.P., B.P., A.P., B.O.R., A.O.R., G.S., L.S., J.S. Seng, C.M.S., S. Stensland, M.H.T., W.K.T., E.T., M.U., U.V., L.L.V.D.H., E.V., Z.W., Y.W., T.W., D.E.W., B.S.W., S.W., E.J.W., R.Y., K.A.Y., and L.A.Z.

Contributed data: O.A.A., P.A.A., S.B. Austin, D.G.B., S. Belangero, L.J.B., R.B., R.A.B., A.D.B., J.R.C., J.M.C.-D.-A., S.Y.C., S.A.P.C., A.M.D., L.K.B., D.L.D., A.E., N.C.F., D.F., C.E.F., S.G., B.G., S.M.J.H., D.M.H., L.M.H., K. Hveem, A.J., I.J., M.L.K., J.L.K., R.C.K., A.P.K., R.K., W.S.K., .A.M.L., K.L., D.F.L., C.E.L., I.L., B.L., M.K.L., S.M., G.A.M., K.M., A.M., K.A.M., S.E.M., J.M., L.M., O.M., P.B.M, M.N., S.B.N., N.R.N., M.O., P.M.P., M.S.P., E.S.P., A.L.P., M.P., R.H.P., M.A.P., K.J.R., V.R., P.R.B., A. Rung, G.S., L.S., S.E.S., M.S., C.S., S. Seedat, J.S. Seng, D. Silove, J.W.S., S.R.S., M.B.S., A.K.T., E.T., U.V., L.L.V.D.H., M.V.H., M.W., T.W., D.E.W., S.W., K.A.Y., C.C.Z., G.C.Z., L.A.Z., and J.Z.

Statistical analysis: A.E.A.-K., A. Batzler, C. Bergner, A. Brandolino, S. Børte, C.C., C.-Y.C., S.A.P.C., J.R.I.C., L.C.-C., B.J.C., S.D., S.G.D., A.D., L.E.D., C.F., M.E.G., B.G., S.B.G., S.D.G., C.G., S.H., E.M.H., K. Hogan, H.H., G.D.J., K.K., P.-F.K., D.F.L., M.W.L., A.L, Y.L., A.X.M., S.M., C.M., D.M., J.M., V.M., E.A.M., M.S.M., C.M.N., G.A.P., M.P., X-J.Q., A.R., A.L.R., S.S.V., C.S., A.S., C.M.S., S. Stensland, J.S.S., J.A.S., F.R.W., B.S.W., Y. Xia, Y. Xiong, and C.C.Z.

Bioinformatics: A.E.A.-K., A. Batzler, M.P.B., S. Børte, C.C., C.-Y.C., J.R.I.C., N.P.D., C.D.P., S.G.D., A.D., H.E., M.E.G., K. Hogan, H.H., K.K., P.-F.K., D.F.L., S.D.L., A.L, A.X.M., G.A.M., D.M., J.M., V.M., E.A.M., G.A.P., A.R., A.S., J.S.S., F.R.W., B.S.W., C.W., Y. Xia, Y. Xiong, and C.C.Z.

Genomics: M.P.B., J.B.-G., M.B.-H., N.P.D., T.d-C, F.D., A.D., K.D., H.E., L.G., M.A.H., J.J., P.-F.K., S.D.L., J.J.L., I.K., J.M., L.M., K.J.R., B.P.F.R., S.S.V., A.S., C.H.V., and D.E.W.

PGC-PTSD management group: M.H. and M.Z.

## Ethics Declarations

L.J.B. is listed as an inventor on Issued U.S. Patent 8,080,371, “Markers for Addiction” covering the use of certain SNPs in determining the diagnosis, prognosis, and treatment of addiction. C.-Y.C. and H.R. are employees of Biogen. A.M.D. holds equity in CorTechs Labs, Inc., and serves on the Scientific Advisory Board of Human Longevity, Inc., and the Mohn Medical Imaging and Visualization Centre; A.M.D. receives funding through research grants with General Electric Healthcare. C.F. was a speaker for Janssen in 2021. I.B.H. is the Co-Director, Health and Policy at the Brain and Mind Centre (BMC) University of Sydney; the BMC operates an early-intervention youth services at Camperdown under contract to headspace. I.B.H. is the Chief Scientific Advisor to, and a 3.2% equity shareholder in, InnoWell Pty Ltd; InnoWell was formed by the University of Sydney (45% equity) and PwC (Australia; 45% equity) to deliver the $30 M Australian Government-funded Project Synergy. H.H. received consultancy fees from Ono Pharmaceutical and honorarium from Xian Janssen Pharmaceutical. In the past 3 years, R.C.K. was a consultant for Cambridge Health Alliance, Canandaigua VA Medical Center, Holmusk, Partners Healthcare, Inc., RallyPoint Networks, Inc., and Sage Therapeutics. He has stock options in Cerebral Inc., Mirah, PYM, Roga Sciences and Verisense Health. L.A.M.L. reports spousal IP payments from Vanderbilt University for technology licensed to Acadia Pharmaceuticals unrelated to the present work. C.M. has served on advisory boards of Receptor Life Sciences, Otsuka Pharmaceuticals and Roche Products Limited and has received support from National Institute on Alcohol Abuse and Alcoholism, National Institute of Mental Health, Department of Defense-CDMRP * US Army Research Office * DARPA, Bank of America Foundation, Brockman Foundation, Cohen Veterans Bioscience, Cohen Veterans Network, McCormick Foundation, Home Depot Foundation, New York City Council, New York State Health, Mother Cabrini Foundation, Tilray Pharmaceuticals, and Ananda Scientific. P.M.P. received payment or honoraria for lectures and presentations in educational events for Sandoz, Daiichi Sankyo, Eurofarma, Abbot, Libbs, Instituto Israelita de Pesquisa e Ensino Albert Einstein, Instituto D’Or de Pesquisa e Ensino. R.P. paid for his editorial work on the journal Complex Psychiatry and received a research grant outside the scope of this study from Alkermes. J.W.S. is a member of the Scientific Advisory Board of Sensorium Therapeutics (with equity), and has received grant support from Biogen, Inc.; J.W.S. is PI of a collaborative study of the genetics of depression and bipolar disorder sponsored by 23andMe for which 23andMe provides analysis time as in-kind support but no payments. M.B.S. has in the past 3 years received consulting income from Acadia Pharmaceuticals, Aptinyx, atai Life Sciences, BigHealth, Biogen, Bionomics, BioXcel Therapeutics, Boehringer Ingelheim, Clexio, Eisai, EmpowerPharm, Engrail Therapeutics, Janssen, Jazz Pharmaceuticals, NeuroTrauma Sciences, PureTech Health, Sage Therapeutics, Sumitomo Pharma, and Roche/Genentech. M.B.S. has stock options in Oxeia Biopharmaceuticals and EpiVario. M.B.S. has been paid for his editorial work on Depression and Anxiety (Editor-in-Chief), Biological Psychiatry (Deputy Editor), and UpToDate (Co-Editor-in-Chief for Psychiatry). M.B.S. has also received research support from NIH, Department of Veterans Affairs, and the Department of Defense. M.B.S. is on the scientific advisory board for the Brain and Behavior Research Foundation and the Anxiety and Depression Association of America. In the past 3 years, D.J.S. has received consultancy honoraria from Discovery Vitality, Johnson & Johnson, Kanna, L’Oreal, Lundbeck, Orion, Sanofi, Servier, Takeda and Vistagen. MLK reports unpaid membership on the Scientific Committee for the ISSTD.

## Figure Legends

### Extended data figure legends

***Extended data Figure 1: Comparison of the genetic architecture of PTSD in the three main data sources***.

Quantification of polygenicity and polygenic overlap in the three main data subsets based on (1) symptom scores in clinical studies and cohorts assessed on a variety of instruments in Freeze 2.5 (yellow; 26,080 cases and 192,966 controls), (2) PCL (for DSM-IV) based symptom scores in the MVP (red; 32,372 cases and 154,317 controls), and (3) ICD9/10 codes in EHR studies (blue; 78,684 cases and 738,463 controls) indicate a similar genetic architecture. The circles on the top half of the plot depict univariate MiXeR estimates of the total polygenicity for each data subset. Numbers within circles indicate polygenicity values, expressed as the number of variants (in thousands, with SE in parenthesis) necessary to explain 90% of SNP based heritability (h^2^_SNP_). h^2^_SNP_ estimates are written in the boxes at the bottom of the circles. The Euler diagrams on the bottom half of the plot depict bivariate MiXeR estimates of the polygenic overlap between data subsets. Values in the overlapping part of the Euler diagrams denote shared polygenicity and values on the non-overlapping parts note dataset-specific polygenicity. Genetic correlations (r_g_) between dataset pairs are noted in the boxes below the Euler diagrams. Arrowed lines are drawn between univariate and bivariate results to indicate which dataset pairs are being evaluated. Abbreviations: Neff, effective sample size.

***Extended data Figure 2: Manhattan plot of the PTSD GWAS meta-analysis in individuals of European ancestry (EA)***.

Results of the EA GWAS meta-analysis (137,136 PTSD cases, 1,085,746 controls) identifying 81 genome-wide significant PTSD loci. The *y axis* refers to the −log10 p-value from a meta-analysis using a sample size weighted fixed-effects model. Circle colors alternate between chromosomes: even chromosomes are colored blue and odd chromosomes are colored black. The horizontal red bar indicates genome-wide significant associations (p < 5×10^-8^).

***Extended Data Figure 3: Significant PTSD gene-sets***.

MAGMA gene-set analysis using the Molecular Signatures database (MSigDB) identifies 11 significant gene-sets. The dotted line indicates significance adjusted for the number of comparisons (p < 0.05/15,483 gene-sets). Bars depict -log_10_ p-values. Corresponding gene-set names are indicated to the left of bars. Terms are clustered and colored according to their Gene Ontology term category (biological processes, yellow; molecular function, blue; cellular component, red).

***Extended Data Figure 4: MAGMA tissue enrichment analysis***.

MAGMA gene-property analysis in 53 specific tissue types from GTEx v8 shows enrichment of PTSD-related genes in 13 brain tissue types and in the pituitary. Bars depict -log_10_ p-values. Corresponding tissue names are indicated below bars. The dotted horizontal line indicates statistical significance adjusted for the number of comparisons (p < 0.05/53). Significant tissues are colored red.

***Extended Data Figure 5: MAGMA cell-type enrichment analysis in midbrain***.

MAGMA gene-property analysis of 25 midbrain cell types (GSE76381) indicate enrichment of GABAergic neurons, GABAergic neuroblasts, and mediolateral neuroblasts. Vertical bars depict -log_10_ p-values. Significant cell types are colored blue and grey if not. The dotted horizontal line indicates statistical significance adjusted for the number of comparisons (p < 0.05/25). The asterisk (*) indicates that GABAergic neurons remained significant in stepwise conditional analysis of the other significant cell types. Abbreviations: Gaba - GABAergic neurons; NbGaba - neuroblast gabaergic; NbML1-5, mediolateral neuroblasts; DA0-2 - dopaminergicneurons; Sert, serotonergic; RN, red nucleus; Rgl 1-3, radial glia-like cells; NbM, medial neuroblast; OPC, oligodendrocyte precursor cells. ProgFPL - progenitor lateral floorplate ; OMTN - oculomotor and trochlear nucleus; Endo, endothelial cells; ProgM, progenitor midline;NProg, neuronal progenitor; ProgBP, progenitorbasal plate; Mgl, microglia; ProgFPM, progenitor medial floorplate; Peric – pericytes.

***Extended Data Figure 6: PTSD genes in SynGO***.

Sunburst plots show enrichment of PTSD-related genes in SynGO cellular components. The synapse is at the center ring, pre- and post-synaptic locations are at the first rings, and child terms are in subsequent outer rings. **a**, enrichment test results for all 415 genes mapped to PTSD GWAS loci by FUMA from one of three gene-mapping strategies (positional, expression quantitative trait loci, and chromatin interaction mapping). **b**, enrichment test results for 43 genes prioritized into Tier 1 using a gene prioritization strategy. Plots are colored by -log10 Q-value (see color code in the bar at left) from enrichment of PTSD genes relative to a brain expressed background set.

***Extended Data Figure 7: Genetic correlations and polygenic overlap between PTSD and other psychiatric disorders***.

**a**, Genetic correlations (r_g_) between PTSD and other psychiatric disorders are indicated by circles that are drawn along the *x* axis. Red dots indicate SNP based heritability (h^2^_SNP_) z-score > 6 in the psychiatric disorder GWAS and colored grey to indicate z-score < 6 (r_g_ estimates may be unreliable). The first author and publication year of source summary data is noted in parenthesis following the disorder name. **b**, Quantification of the polygenic overlap between PTSD and other psychiatric disorders. Euler diagrams depict Bivariate MiXeR analysis of PTSD (blue circles) and bipolar disorder (BIP), major depression (MDD), and schizophrenia (SCZ) (red circles). Values in the overlapping part of the Euler diagrams denote shared polygenicity (expressed as the number of influential variants, in thousands, with SE in parenthesis), and values in the non-overlapping part indicate dataset-specific variation. r_g_ between dataset pairs are noted in the boxes below the Euler plots. Abbreviations: ADHD, attention deficit hyperactive disorder; alc. dep, alcohol dependence; BIP, bipolar disorder; MDD, major depression; OCD, obsessive compulsive disorder; SCZ, schizophrenia.

***Extended Data Figure 8: Genetic correlation and causal relationships with non-psychiatric traits***.

A total of 1,114 traits from the Pan-UKB database were analyzed. The 12 traits with a significant shared genetic causality proportion (GCP) with PTSD are depicted. **a**, Genetic correlation between PTSD with each trait. Red circles indicate genetic correlation estimates. **b**, GCP estimates between PTSD and each trait. Blue circles indicate the GCP estimates. The vertical dotted line indicates zero shared causality. GCP estimates to the right of the dotted line indicate the causal influence of PTSD on the trait, whereas values to the left the line indicate a causal influence of the trait on PTSD.

***Extended Data Figure 9: Mendelian randomization analysis identifies causal effects of PTSD on lipid traits***.

Two-sample Mendelian randomization (MR) of PTSD and lipid traits, including disorders of lipoid metabolism (phecode 272); hyperlipidemia (phecode 272.1); hypercholesterolemia (phecode 272.11); disorders of lipoprotein metabolism and other lipidemias (ICD-10 E78), and “Non-cancer illness code, self-reported: High cholesterol”. Results are shown for MR analyses as corrected for sample overlap between datasets (orange) and uncorrected inverse variance weighted MR (blue).

## Notes

### Author Declarations

IRB of University of California San Diego gave ethical approval for this work

