## Supplementary figures and images for "Discovery of 95 PTSD loci provides insight into genetic architecture and neurobiology of trauma and stress-related disorders"

### Extended Data Figure 1

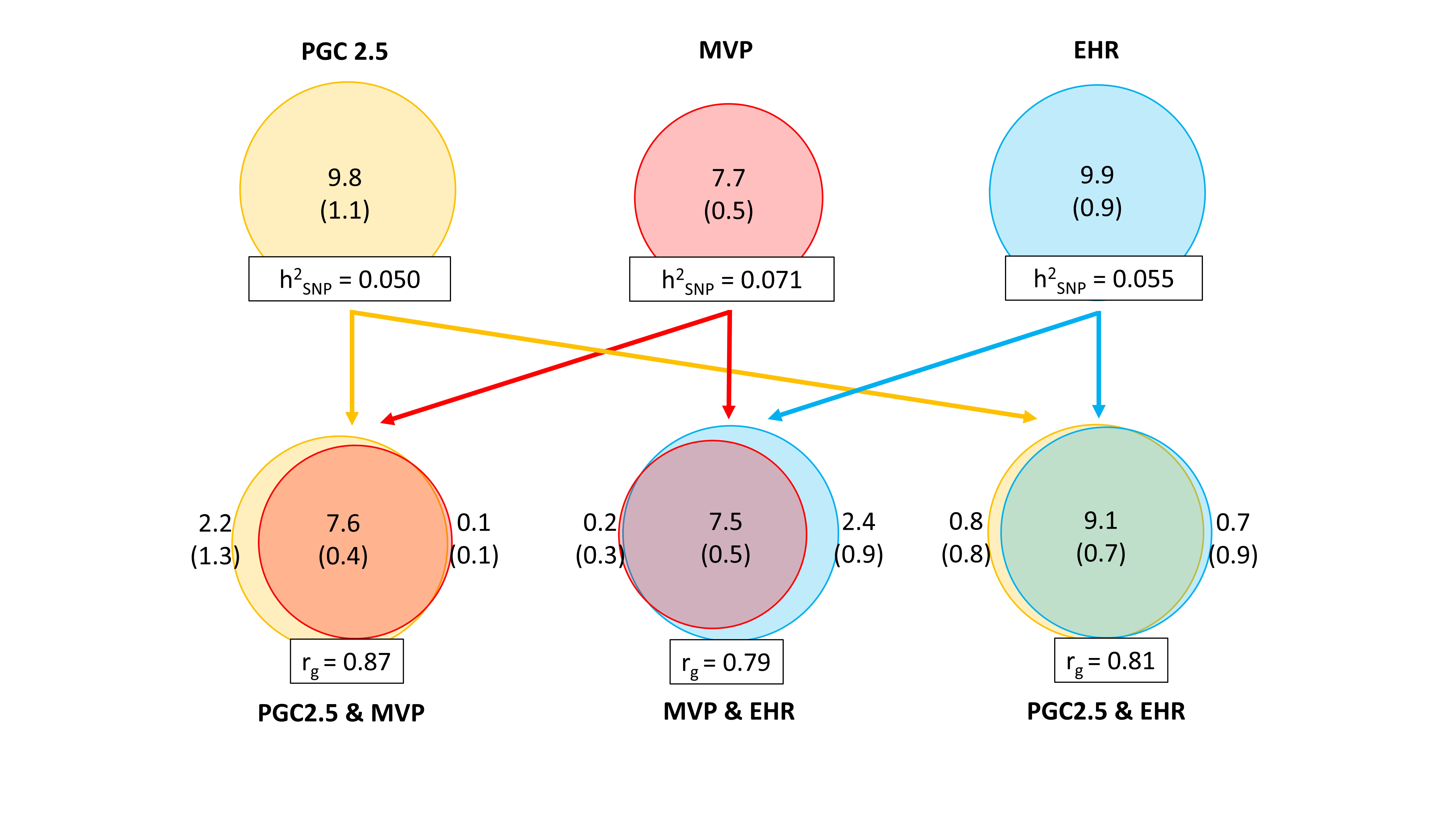

### Extended Data Figure 2

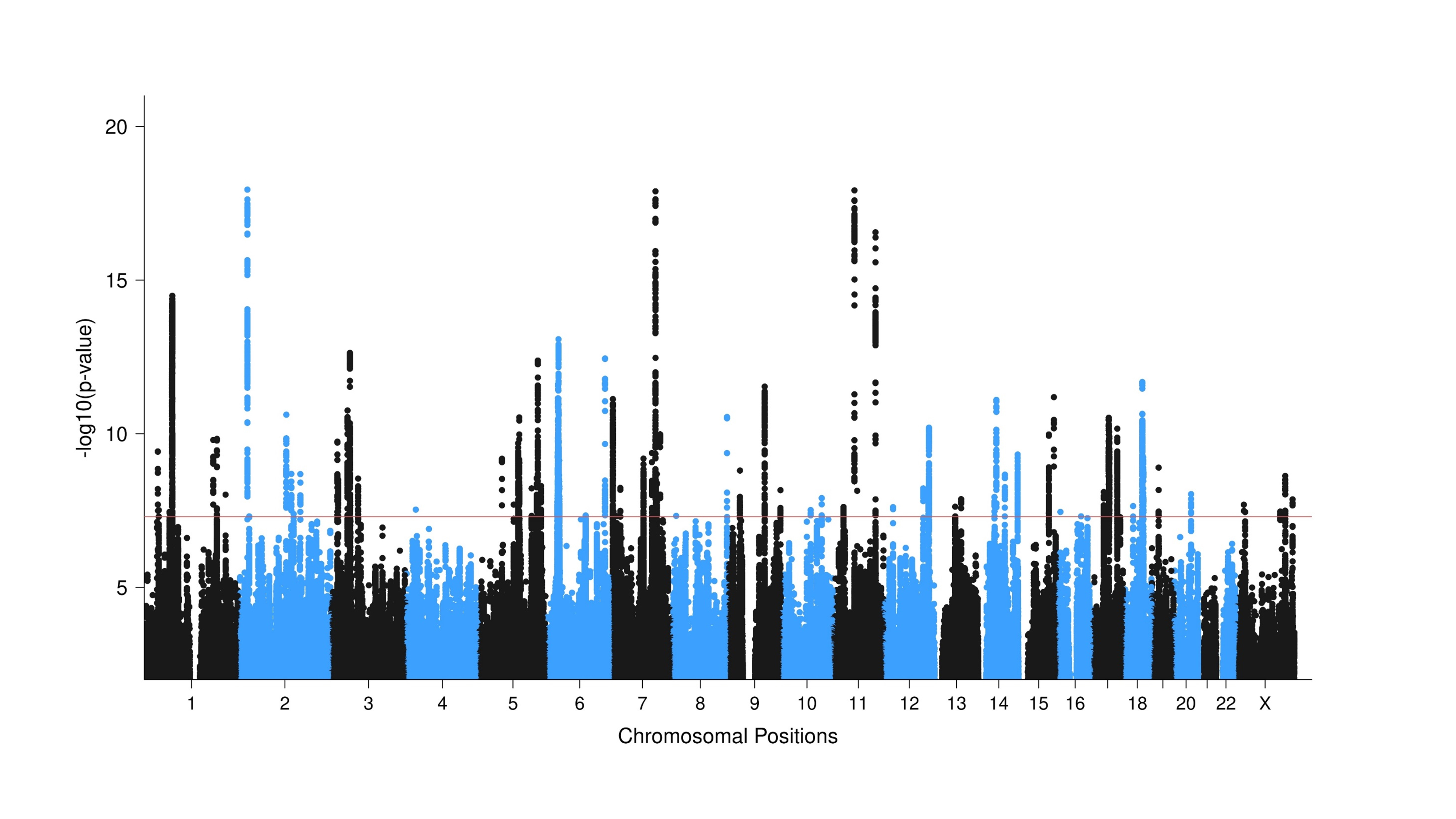

### Extended Data Figure 3

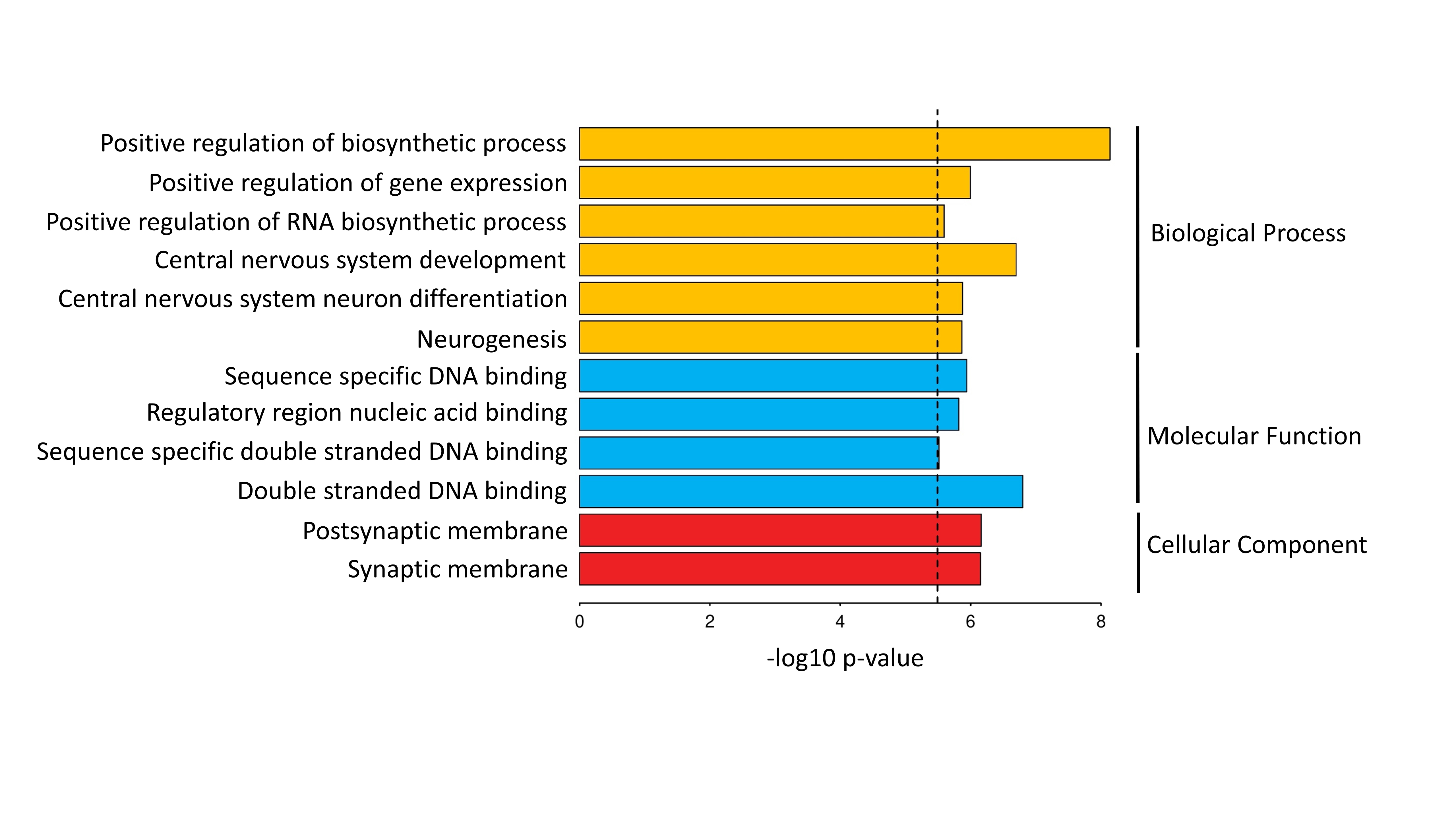

### Extended Data Figure 4

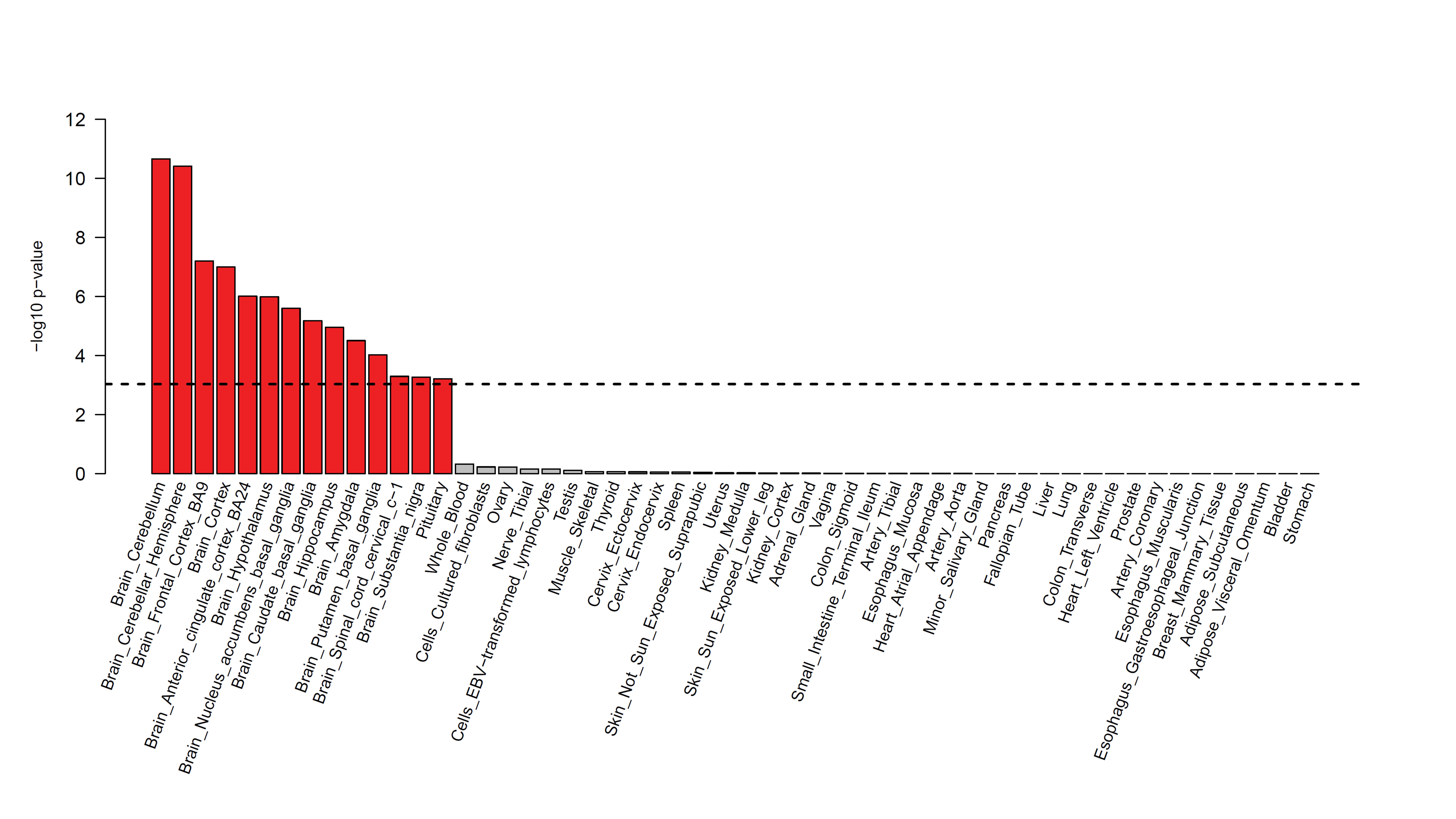

### Extended Data Figure 5

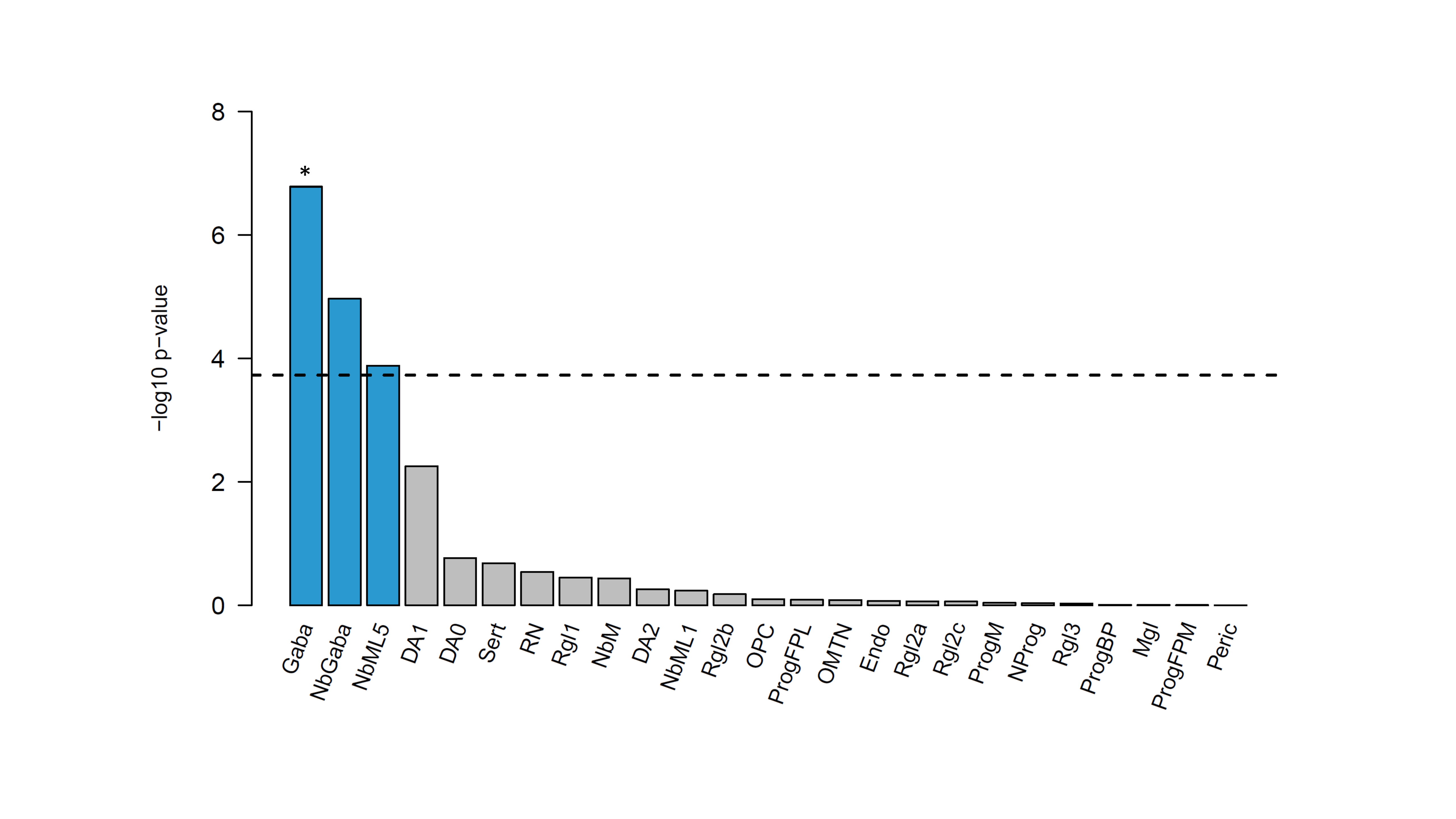

### Extended Data Figure 6

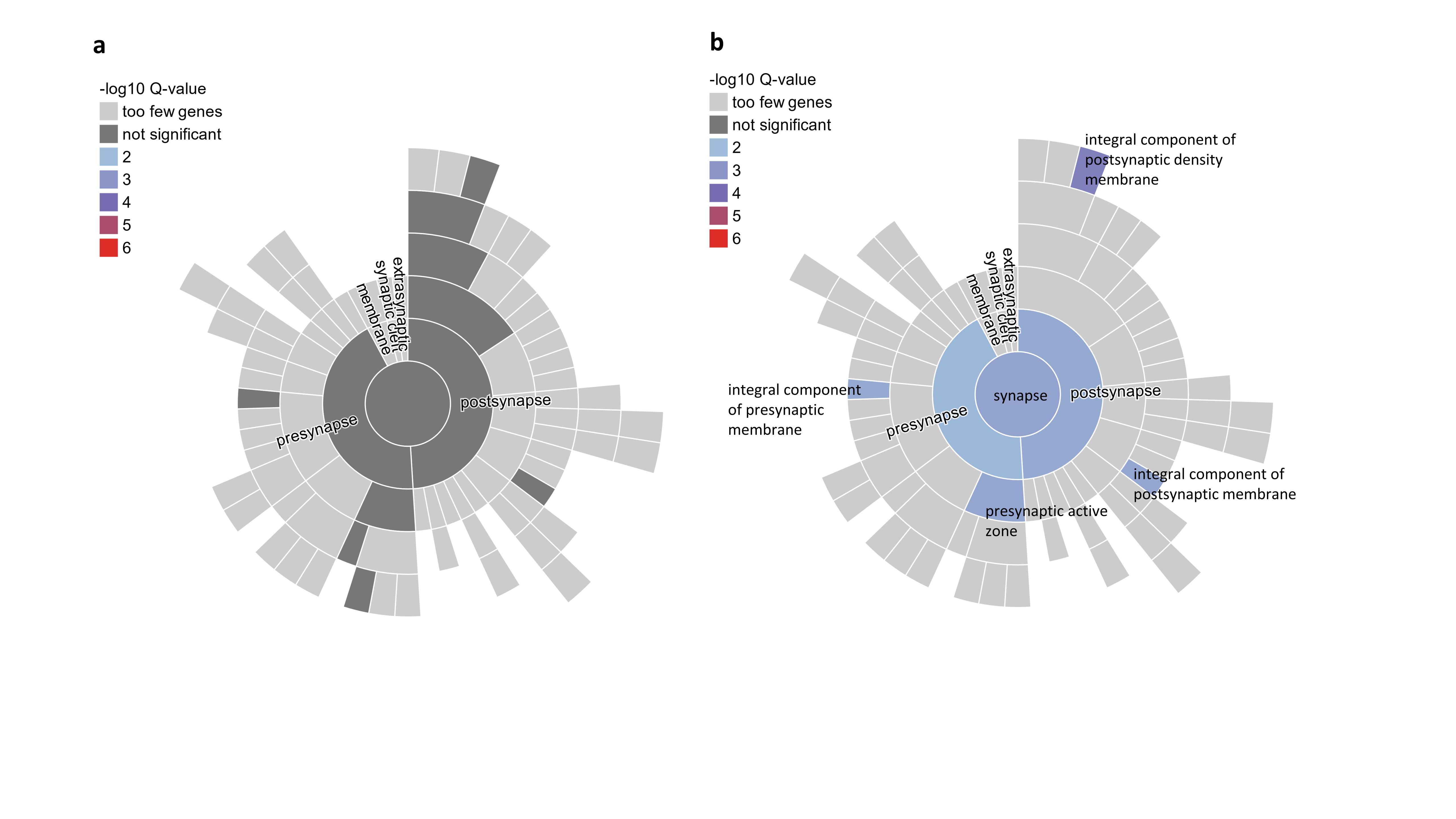

### Extended Data Figure 7

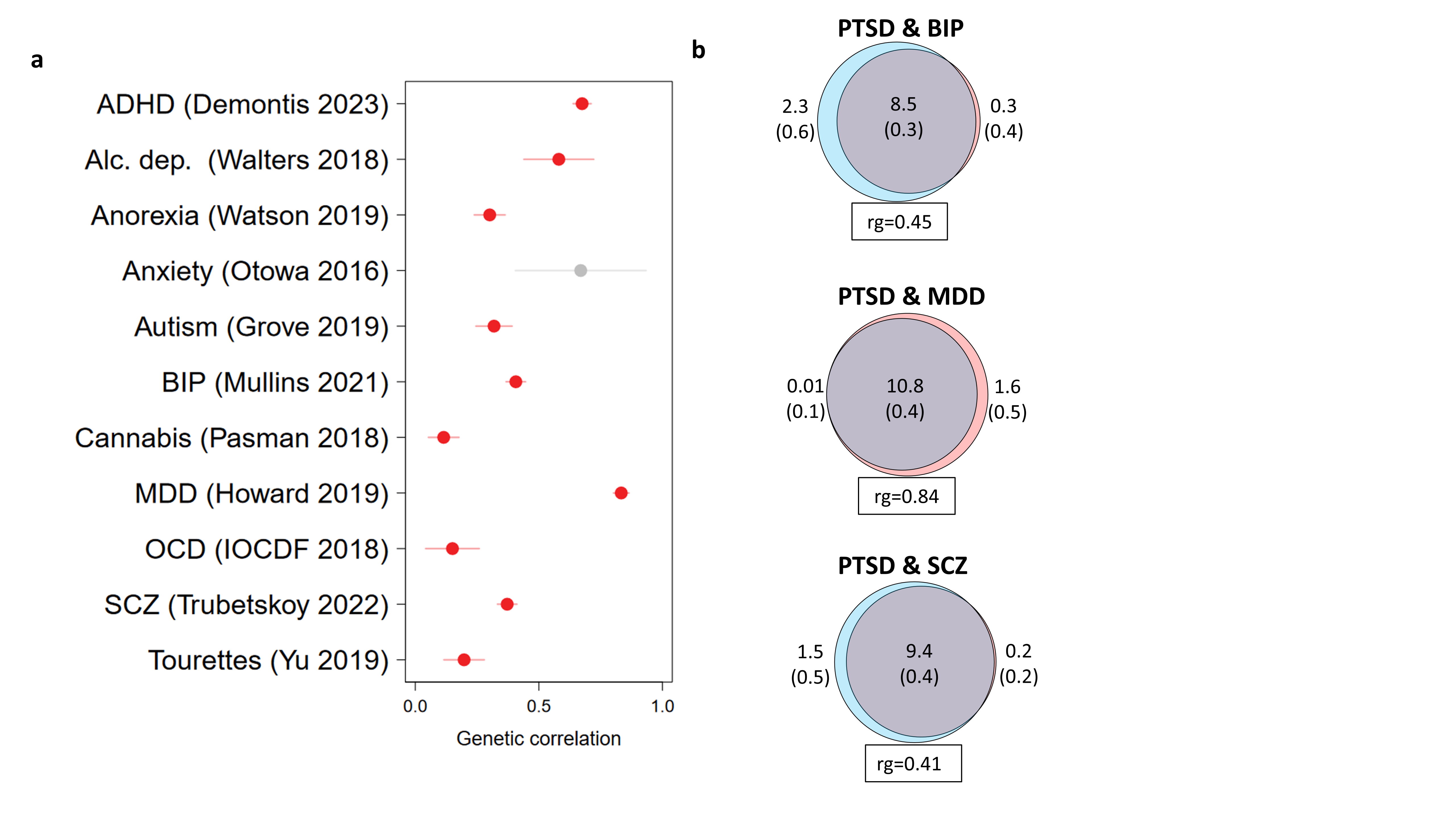
