## Supplemental Data 1 for "Discovery of 95 PTSD loci provides insight into genetic architecture and neurobiology of trauma and stress-related disorders"

### Supplementary Data 1

Regional association plots of each significant locus from the EA GWAS meta-analysis of 137,136 PTSD cases and 1,085,746 controls. The x-axis represents the base pair (hg19) genetic position of SNPs. The y-axis represents –log p-values of SNP association with PTSD. LD estimates of surrounding SNPs with the labeled index SNP (LD $r^2$ values estimated based on 1KGP3 Europeans) is indicated by color (color bar on side of plot indicates color coding of $r^2$ values). Local estimates of recombination rate are indicated in light blue (legend on vertical axis at right). Gene names, strands, and boundaries are shown in the box below the regional plot.

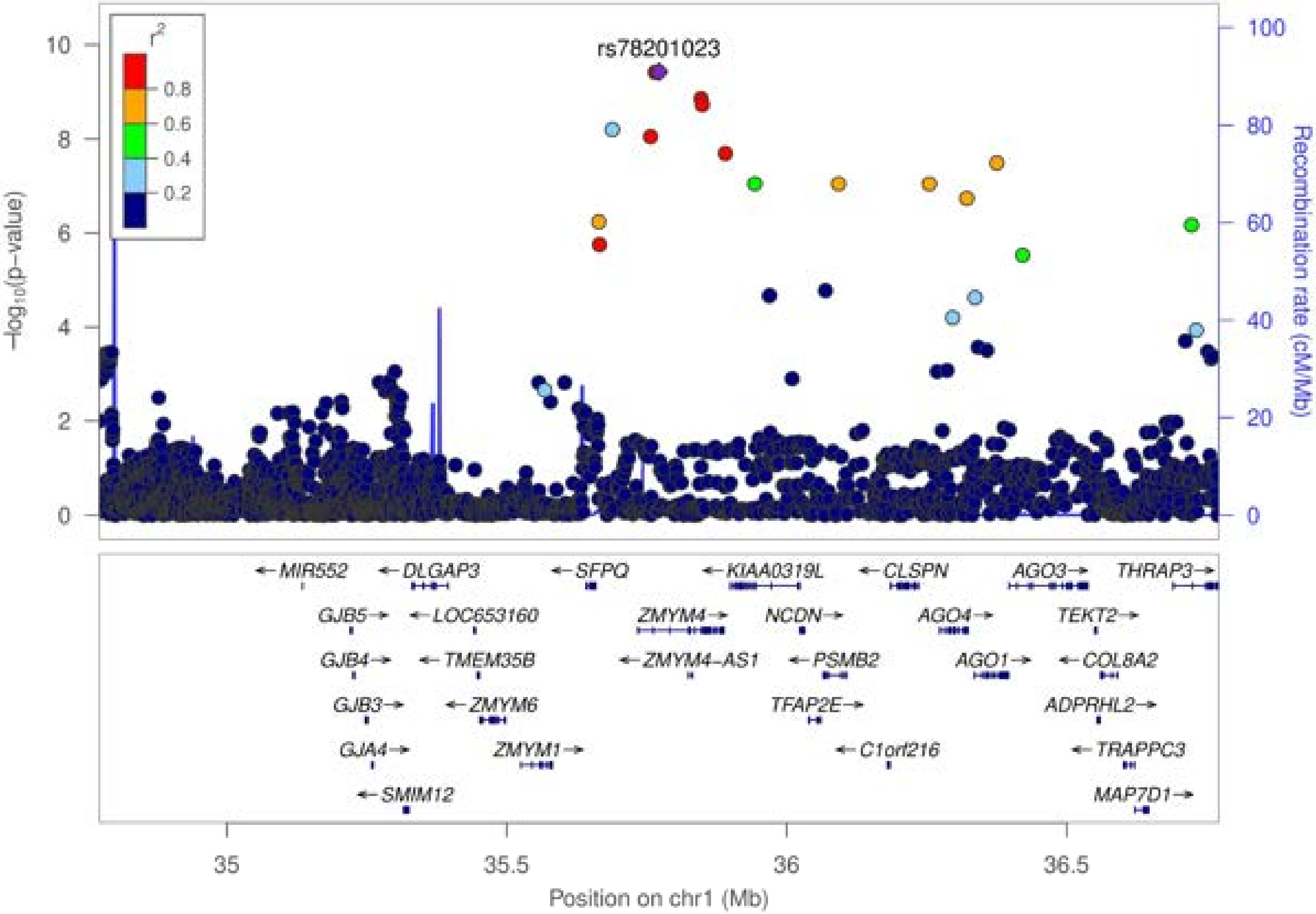

rs78201023
r²
0.8
0.6
0.4
0.2
−log₁₀(p−value)
Recombination rate (cM/Mb)
MIR552
DLGAP3
SFPQ
KIAA0319L
CLSPN
AGO3
THRAP3
GJB5
LOC653160
ZMYM4
NCDN
AGO4
TEKT2
GJB4
TMEM35B
ZMYM4−AS1
PSMB2
AGO1
COL8A2
GJB3
ZMYM6
TFAP2E
ADPRHL2
GJA4
ZMYM1
C1orf216
TRAPPC3
SMIM12
MAP7D1
Position on chr1 (Mb)

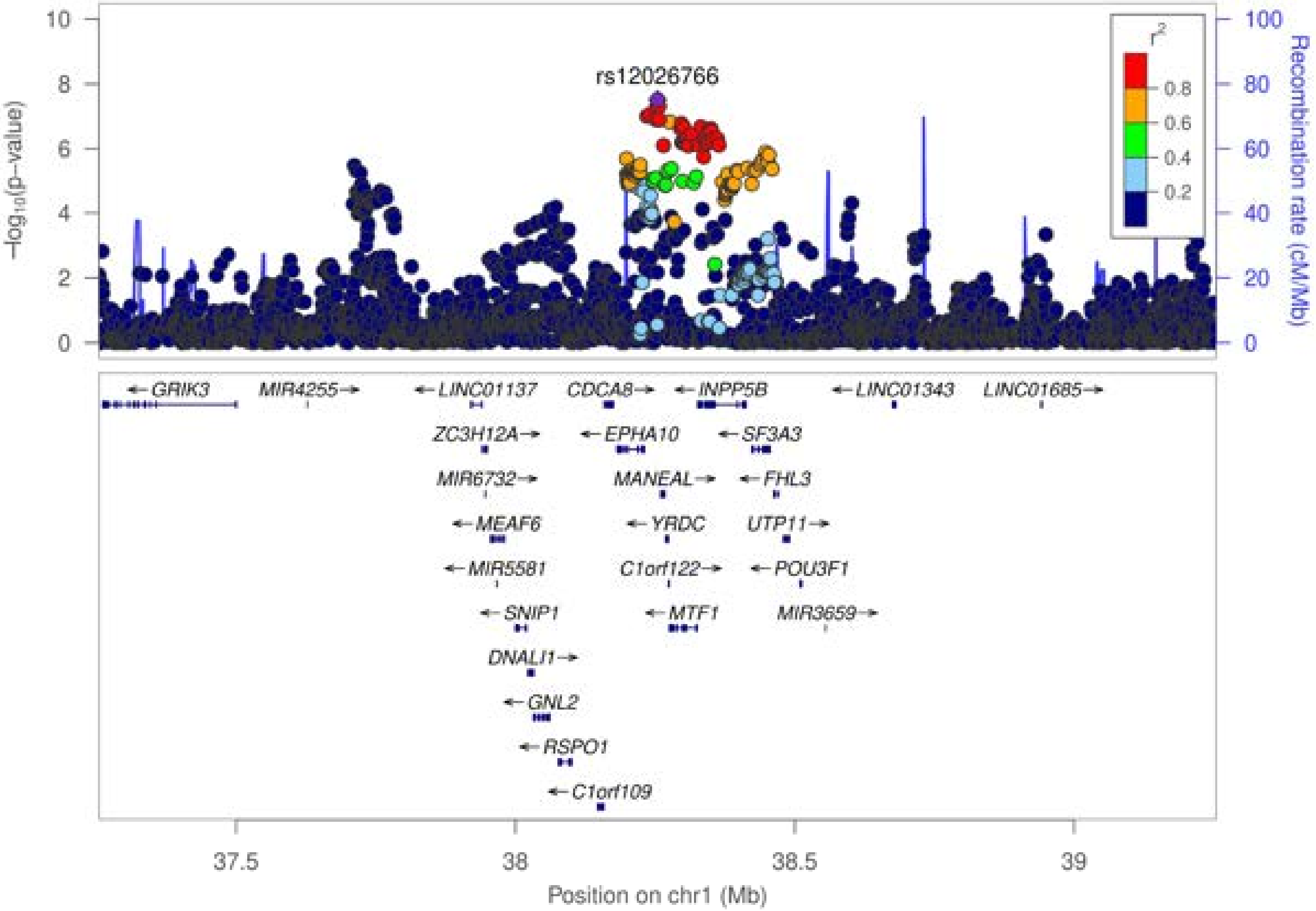

rs12026766
−log10(p−value)
Recombination rate (cM/Mb)
r2
0.8
0.6
0.4
0.2
GRIK3
MIR4255
LINC01137
CDCA8
INPP5B
LINC01343
LINC01685
ZC3H12A
EPHA10
SF3A3
MIR6732
MANEAL
FHL3
MEAF6
YRDC
UTP11
MIR5581
C1orf122
POU3F1
SNIP1
MTF1
MIR3659
DNALI1
GNL2
RSPO1
C1orf109
37.5
38
38.5
39
Position on chr1 (Mb)

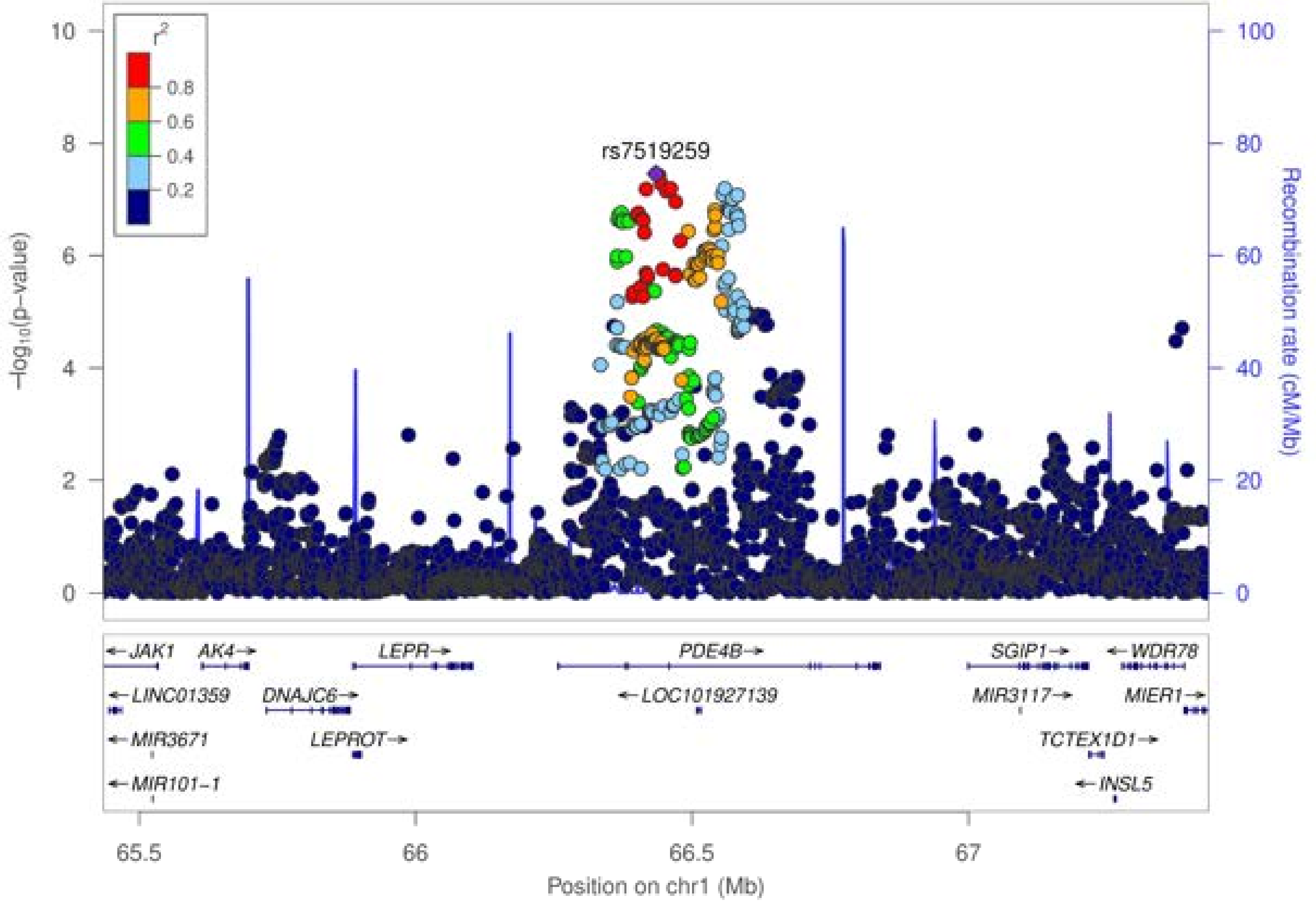

r²
0.8
0.6
0.4
0.2
rs7519259
−log₁₀(p−value)
Recombination rate (cM/Mb)
10
8
6
4
2
0
100
80
60
40
20
0
JAK1
AK4
LEPR
PDE4B
SGIP1
WDR78
LINC01359
DNAJC6
LOC101927139
MIR3117
MIER1
MIR3671
LEPROT
TCTEX1D1
MIR101-1
INSL5
65.5
66
66.5
67
Position on chr1 (Mb)

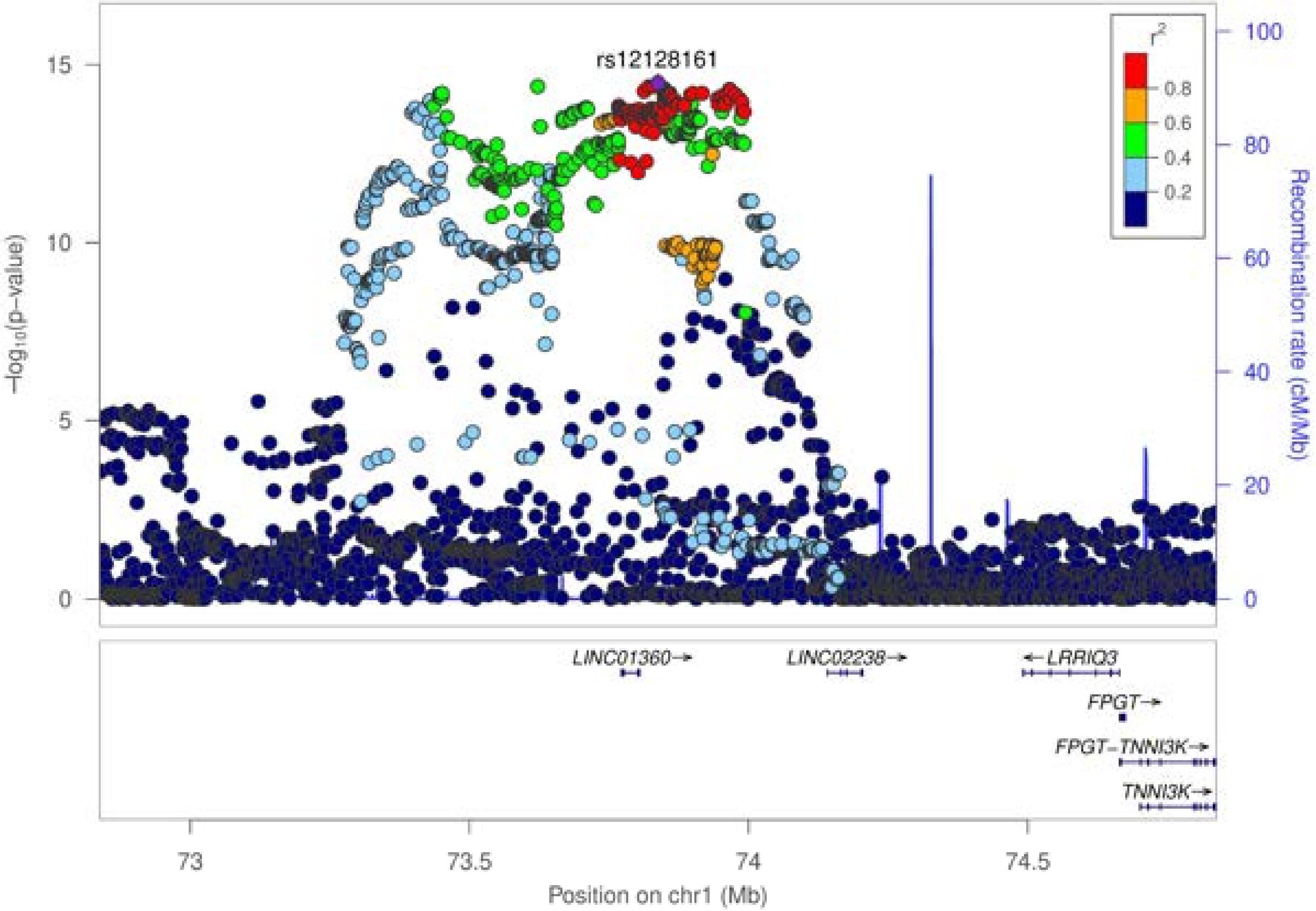
rs12128161
r²
0.8
0.6
0.4
0.2
−log₁₀(p−value)
Recombination rate (cM/Mb)
0
5
10
15
0
20
40
60
80
100
LINC01360→
LINC02238→
←LRRIQ3
FPGT→
FPGT−TNNI3K→
TNNI3K→
73
73.5
74
74.5
Position on chr1 (Mb)

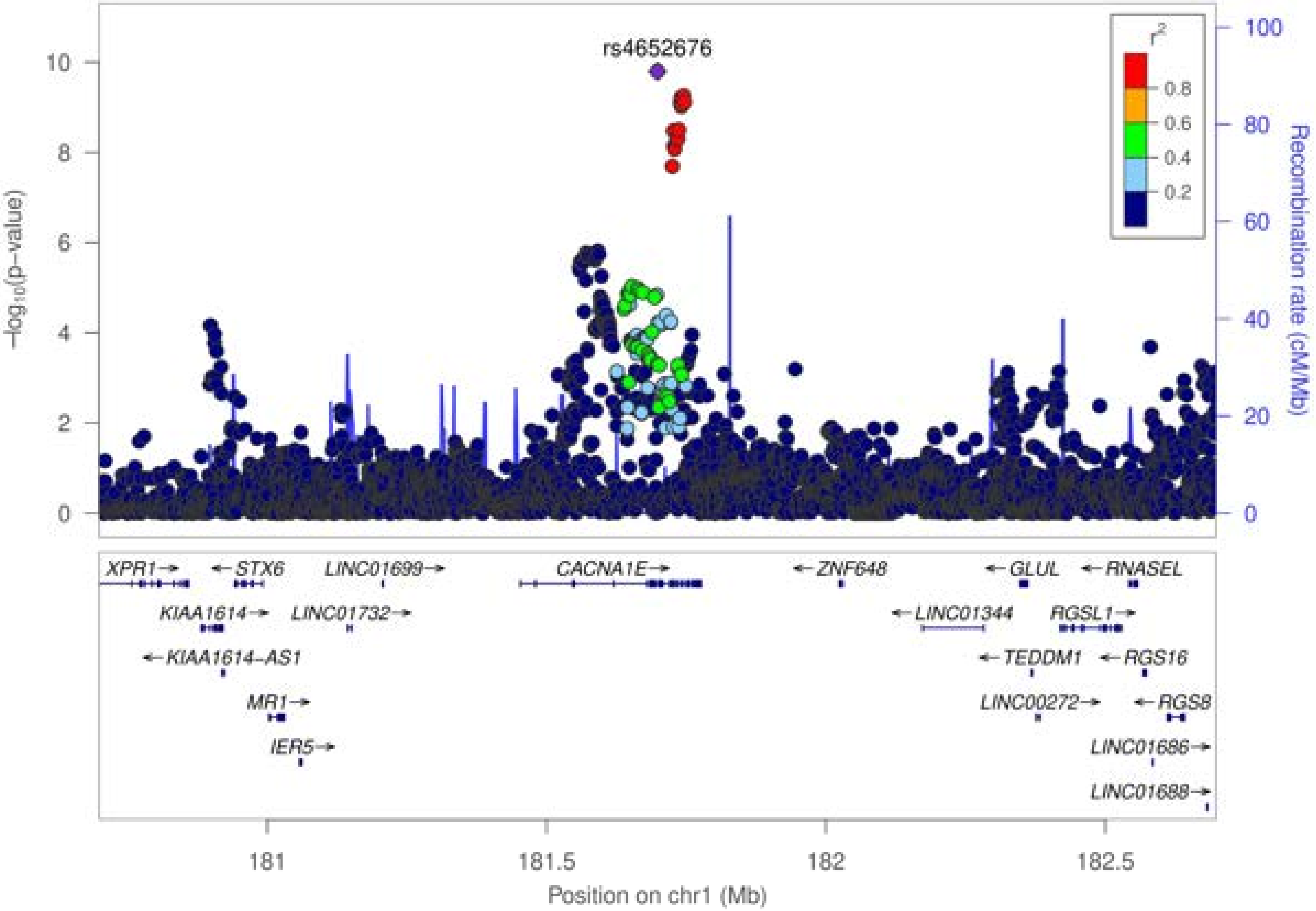
rs4652676
−log10(p−value)
Recombination rate (cM/Mb)
r²
0.8
0.6
0.4
0.2
0
2
4
6
8
10
0
20
40
60
80
100
XPR1
STX6
LINC01699
CACNA1E
ZNF648
GLUL
RNASEL
KIAA1614
LINC01732
LINC01344
RGSL1
KIAA1614-AS1
TEDDM1
RGS16
MR1
LINC00272
RGS8
IER5
LINC01686
LINC01688
181
181.5
182
182.5
Position on chr1 (Mb)

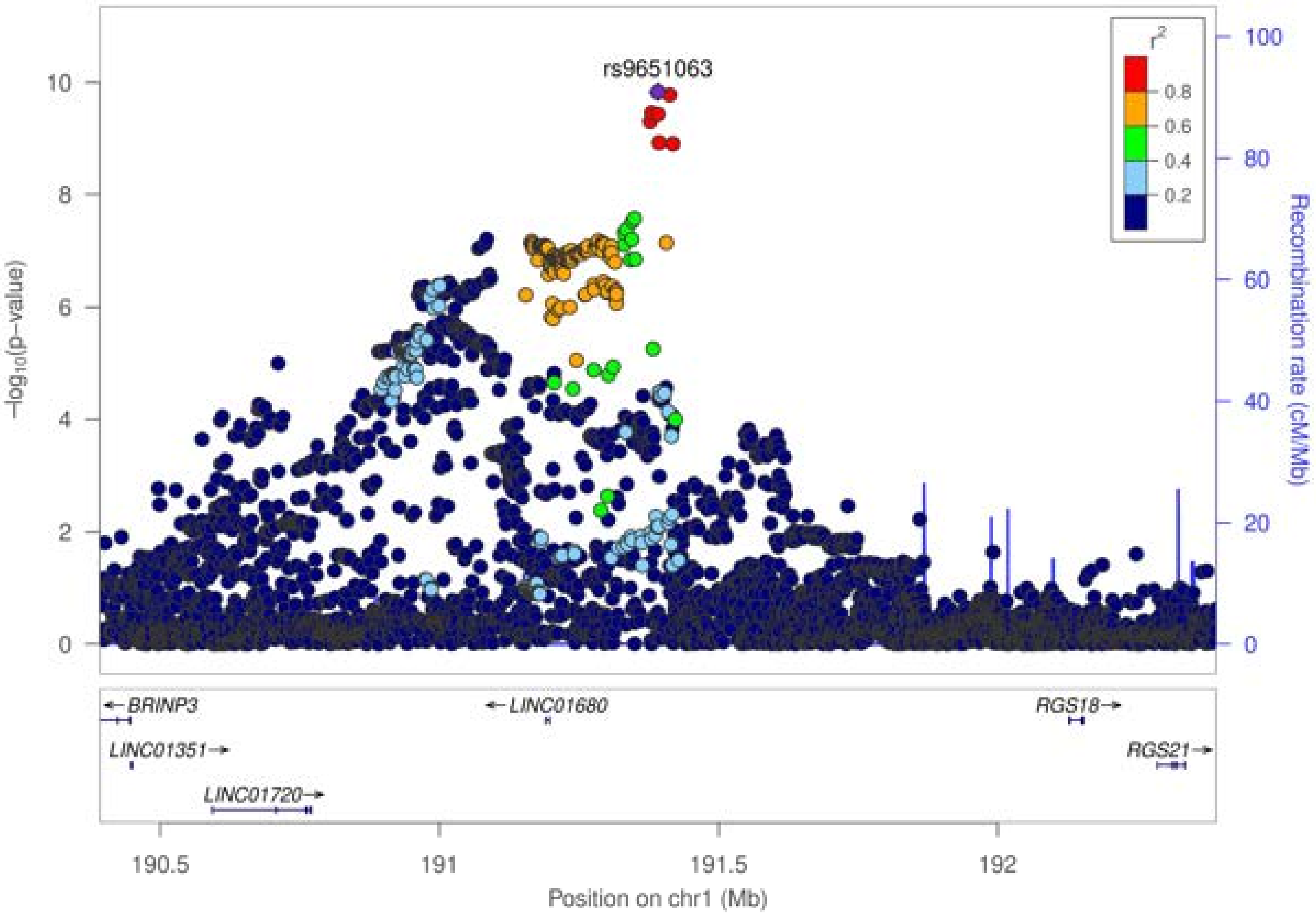

rs9651063
r²
0.8
0.6
0.4
0.2
10
8
6
4
2
0
100
80
60
40
20
0
−log10(p−value)
Recombination rate (cM/Mb)
BRINP3
LINC01680
RGS18
LINC01351
RGS21
LINC01720
190.5
191
191.5
192
Position on chr1 (Mb)

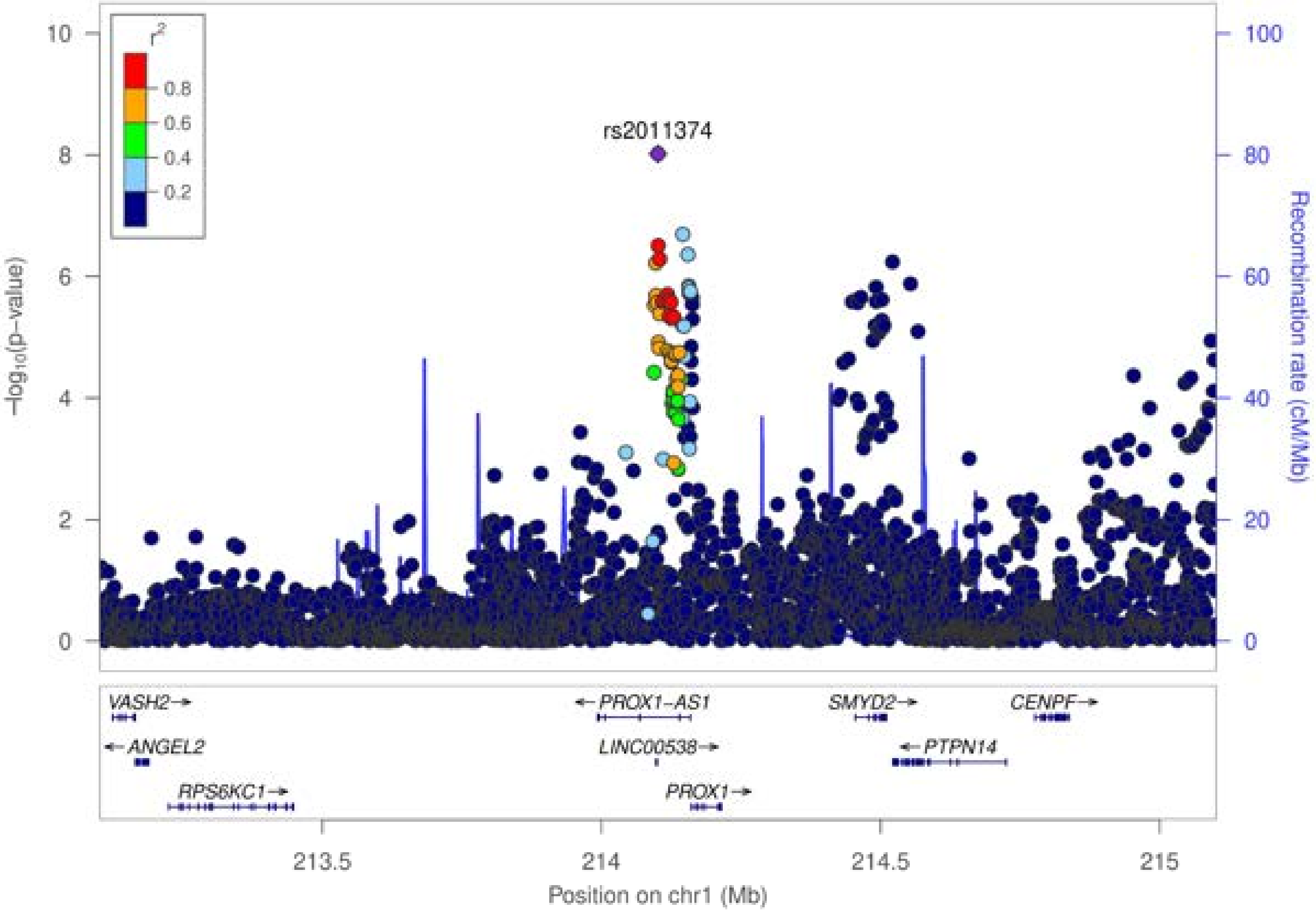

r²
0.8
0.6
0.4
0.2
rs2011374
-log₁₀(p-value)
Recombination rate (cM/Mb)
0
2
4
6
8
10
0
20
40
60
80
100
VASH2
ANGEL2
RPS6KC1
PROX1-AS1
LINC00538
PROX1
SMYD2
PTPN14
CENPF
213.5
214
214.5
215
Position on chr1 (Mb)

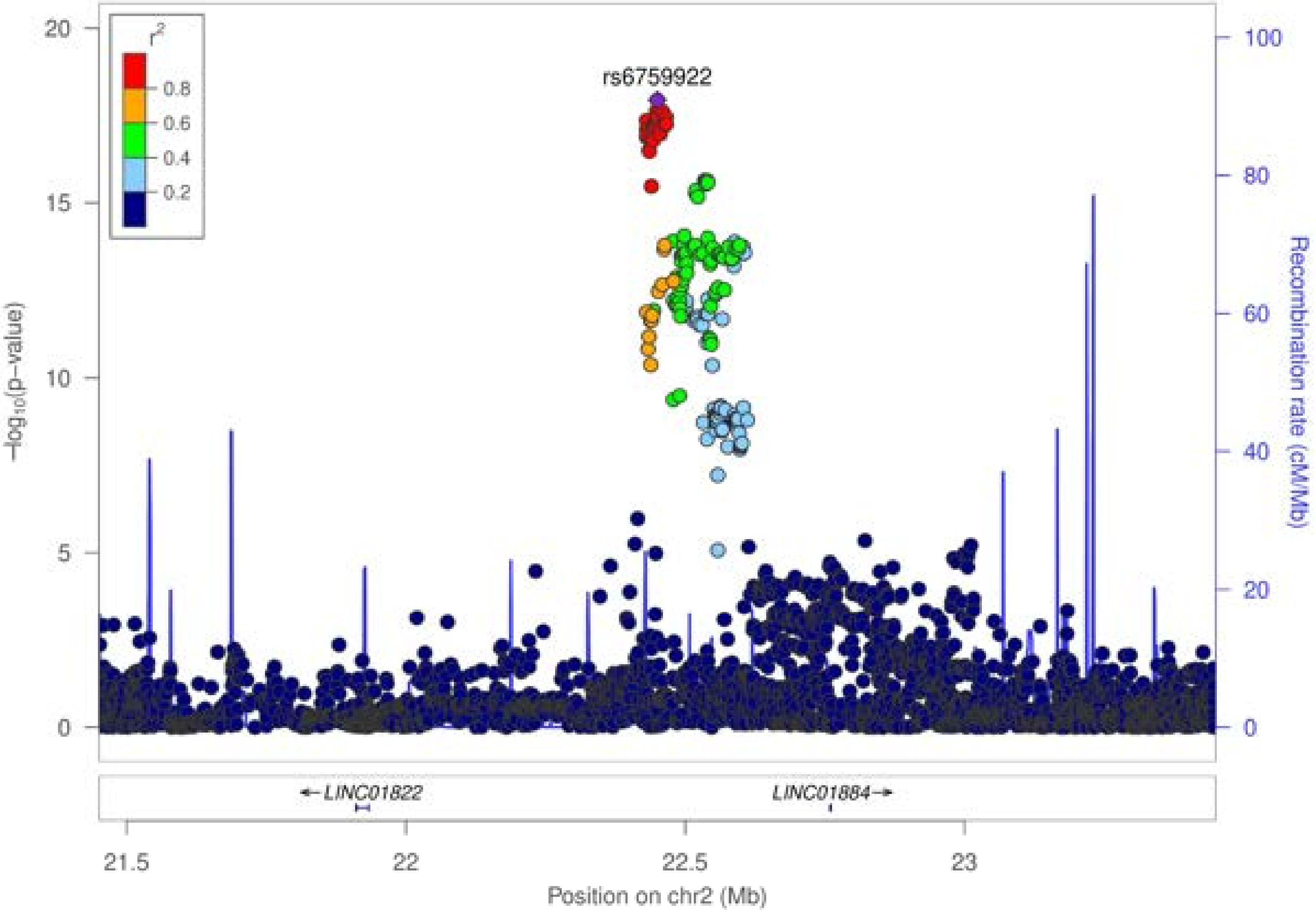

rs6759922
r²
0.8
0.6
0.4
0.2
−log10(p−value)
Recombination rate (cM/Mb)
0
5
10
15
20
0
20
40
60
80
100
LINC01822
LINC01884
21.5
22
22.5
23
Position on chr2 (Mb)

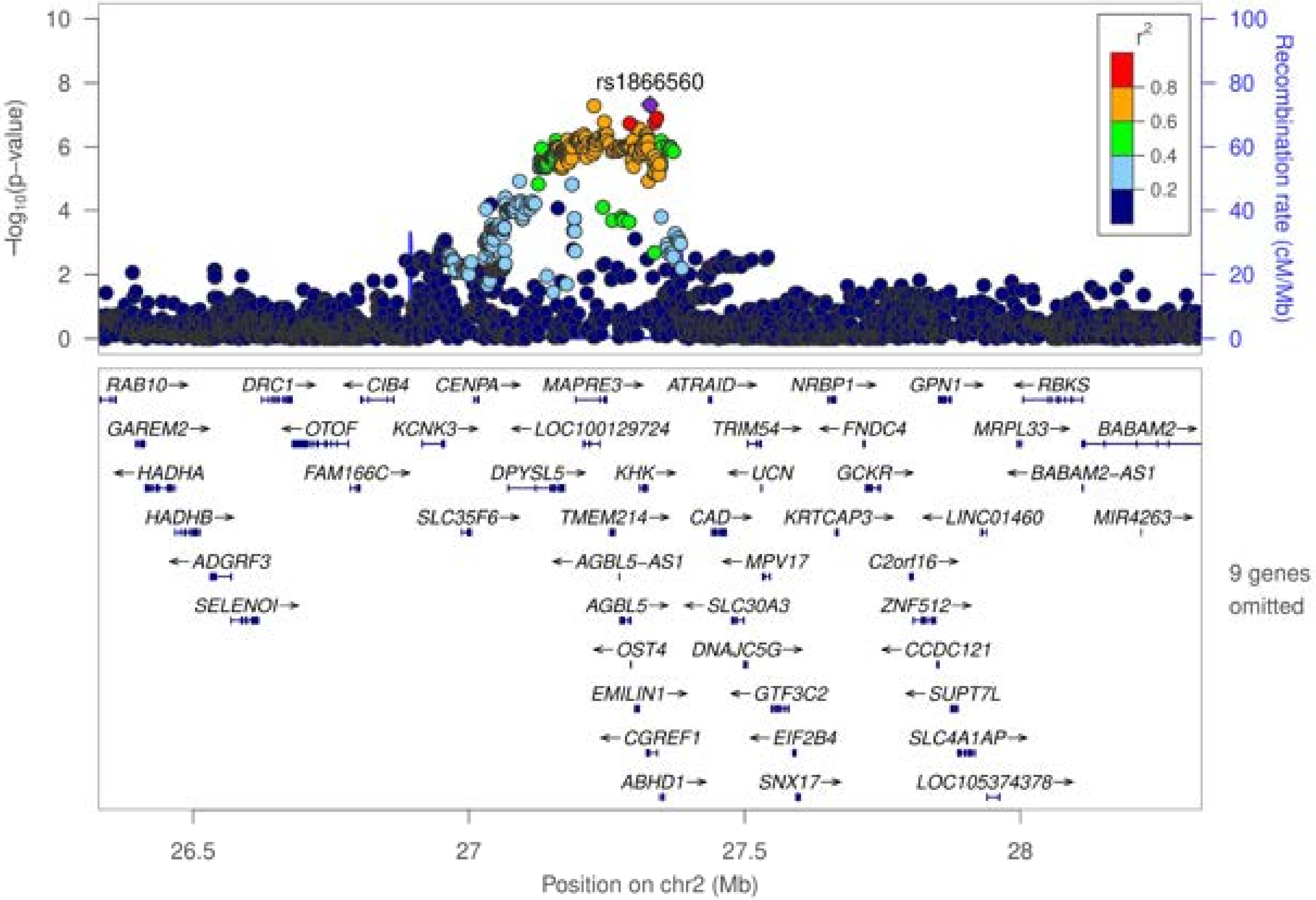
rs1866560
−log10(p−value)
Recombination rate (cM/Mb)
r2
0.8
0.6
0.4
0.2
0
2
4
6
8
10
20
40
60
80
100
RAB10 DRC1 CIB4 CENPA MAPRE3 ATRAID NRBP1 GPN1 RBKS
GAREM2 OTOF KCNK3 LOC100129724 TRIM54 FNDC4 MRPL33 BABAM2
HADHA FAM166C DPYSL5 KHK UCN GCKR BABAM2-AS1
HADHB SLC35F6 TMEM214 CAD KRTCAP3 LINC01460 MIR4263
ADGRF3 AGBL5-AS1 MPV17 C2orf16
SELENOI AGBL5 SLC30A3 ZNF512
OST4 DNAJC5G CCDC121
EMILIN1 GTF3C2 SUPT7L
CGREF1 EIF2B4 SLC4A1AP
ABHD1 SNX17 LOC105374378
9 genes omitted
26.5
27
27.5
28
Position on chr2 (Mb)

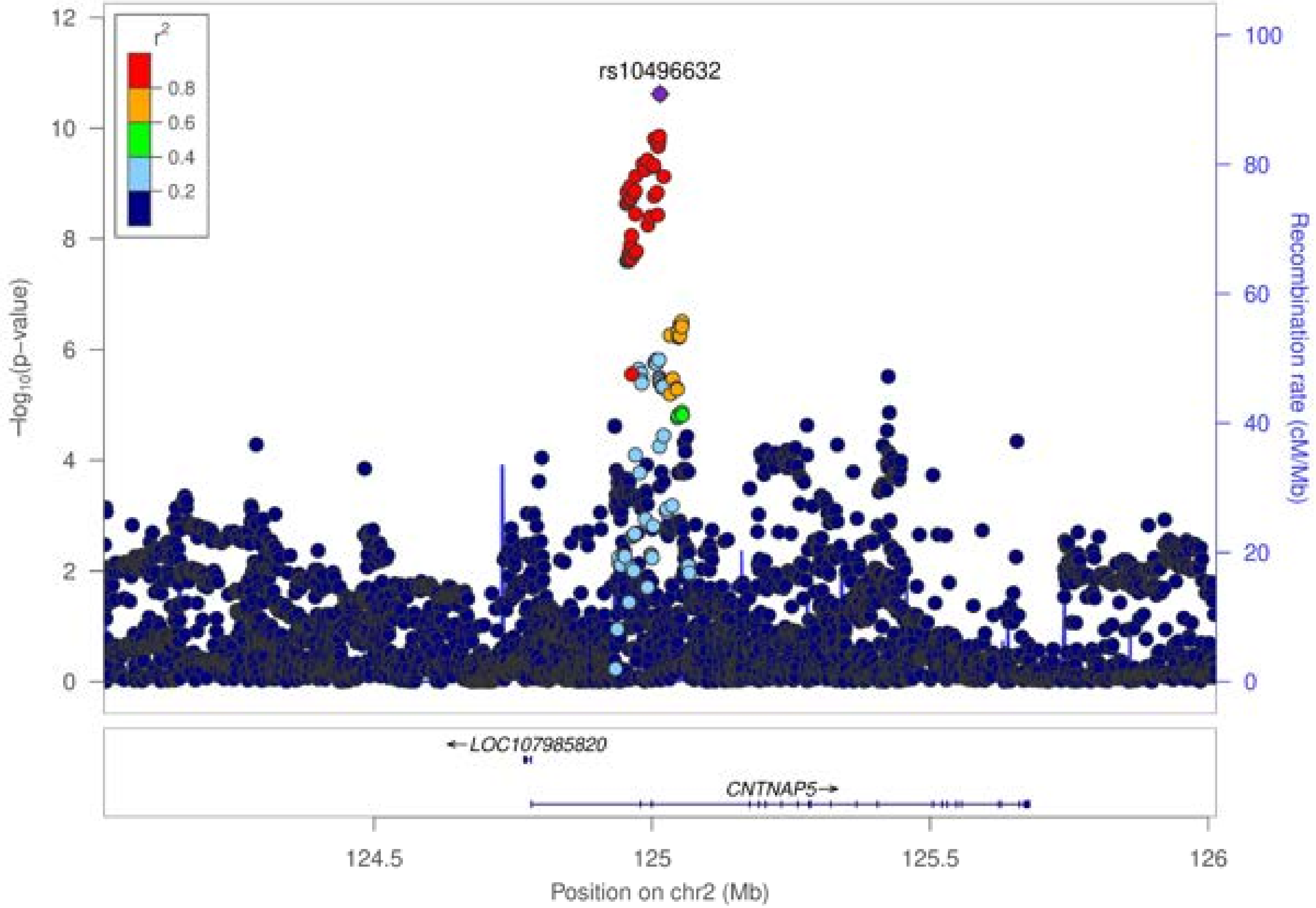
rs10496632
$r^2$
0.8
0.6
0.4
0.2
$-\log_{10}$(p-value)
Recombination rate (cM/Mb)
LOC107985820
CNTNAP5
124.5
125
125.5
126
Position on chr2 (Mb)

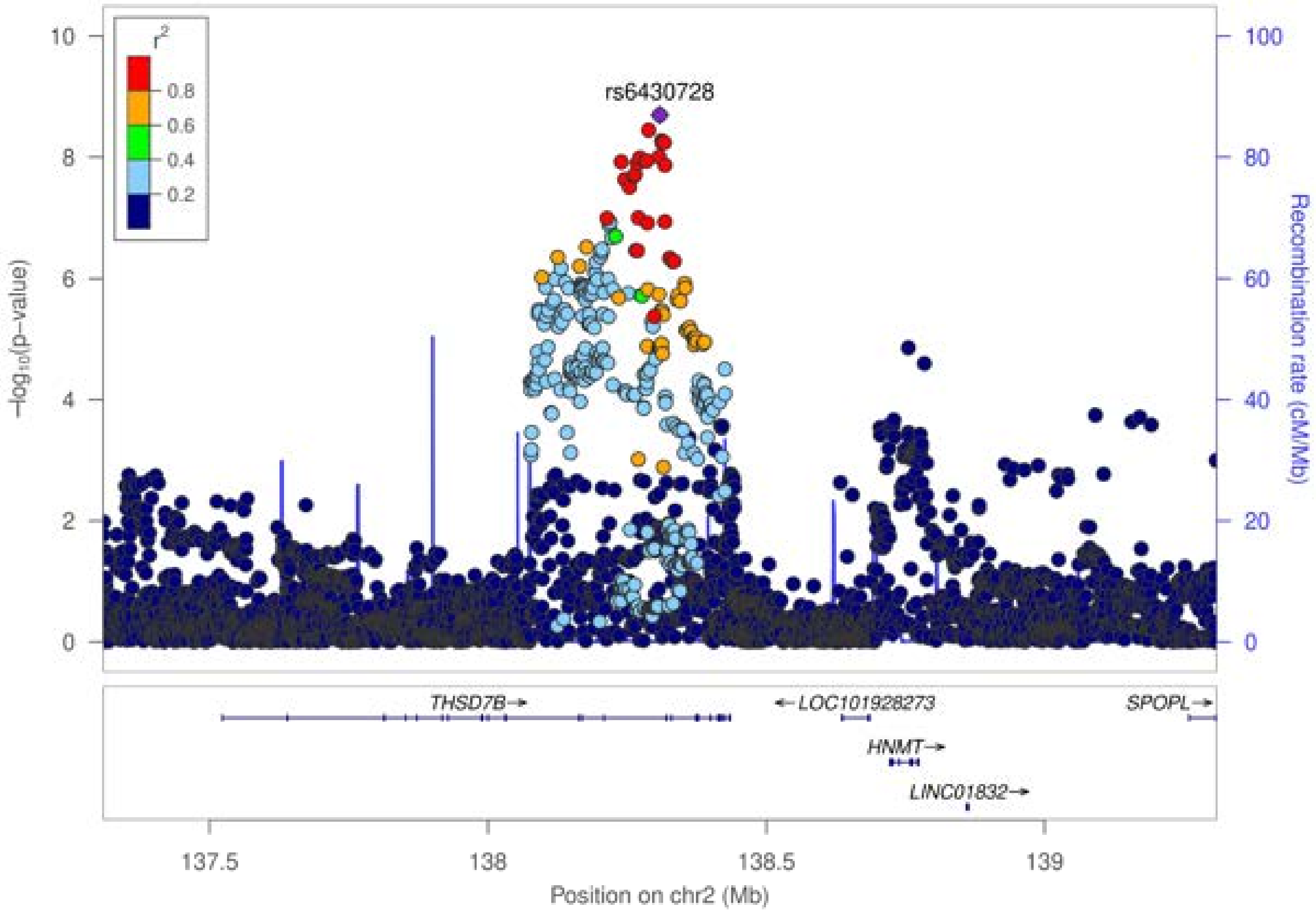

r²
0.8
0.6
0.4
0.2
rs6430728
−log10(p−value)
Recombination rate (cM/Mb)
THSD7B→
← LOC101928273
SPOPL→
HNMT→
LINC01832→
137.5
138
138.5
139
Position on chr2 (Mb)

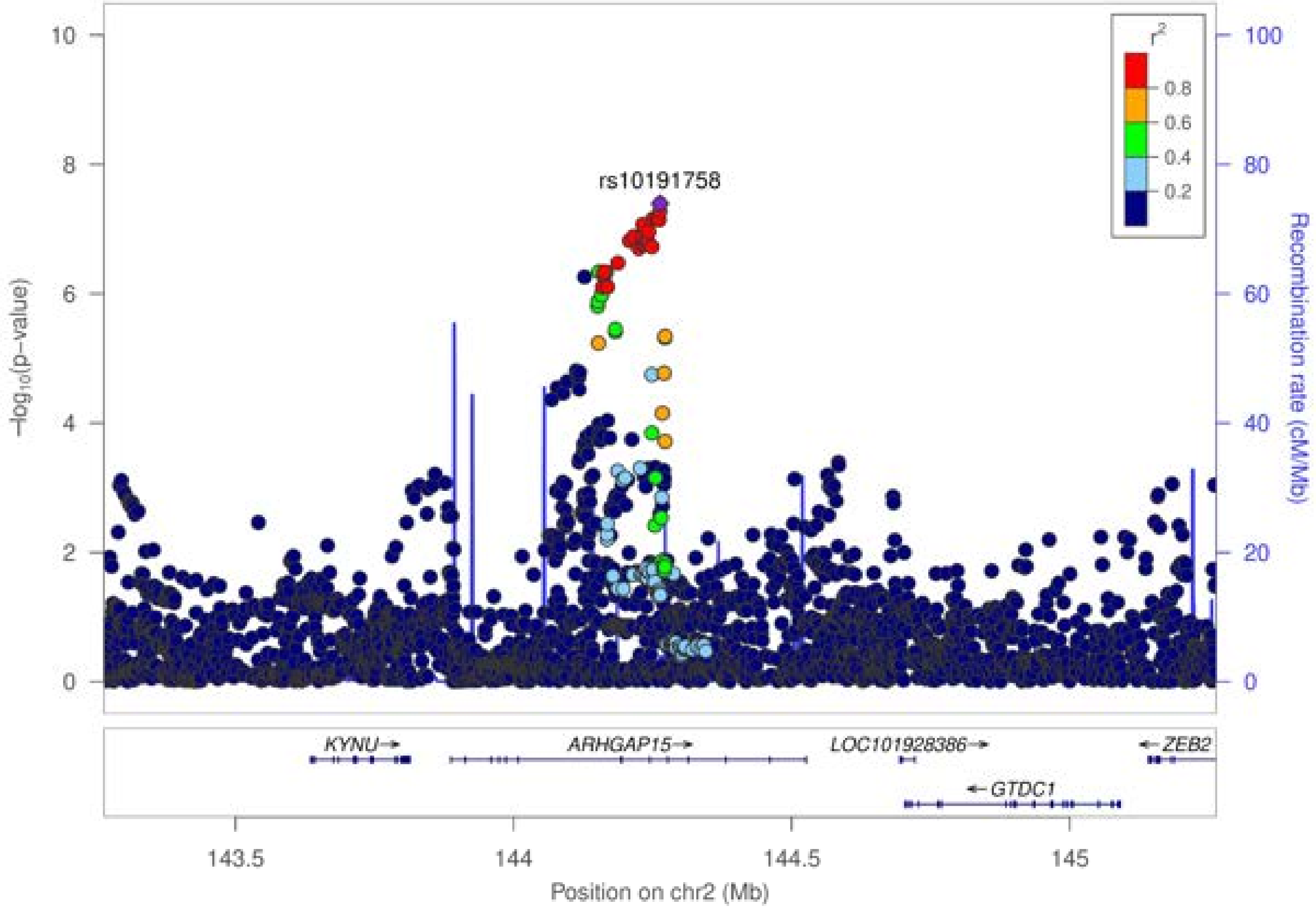

rs10191758
r2
0.8
0.6
0.4
0.2
-log10(p-value)
Recombination rate (cM/Mb)
KYNU→
ARHGAP15→
LOC101928386→
←ZEB2
←GTDC1
143.5
144
144.5
145
Position on chr2 (Mb)

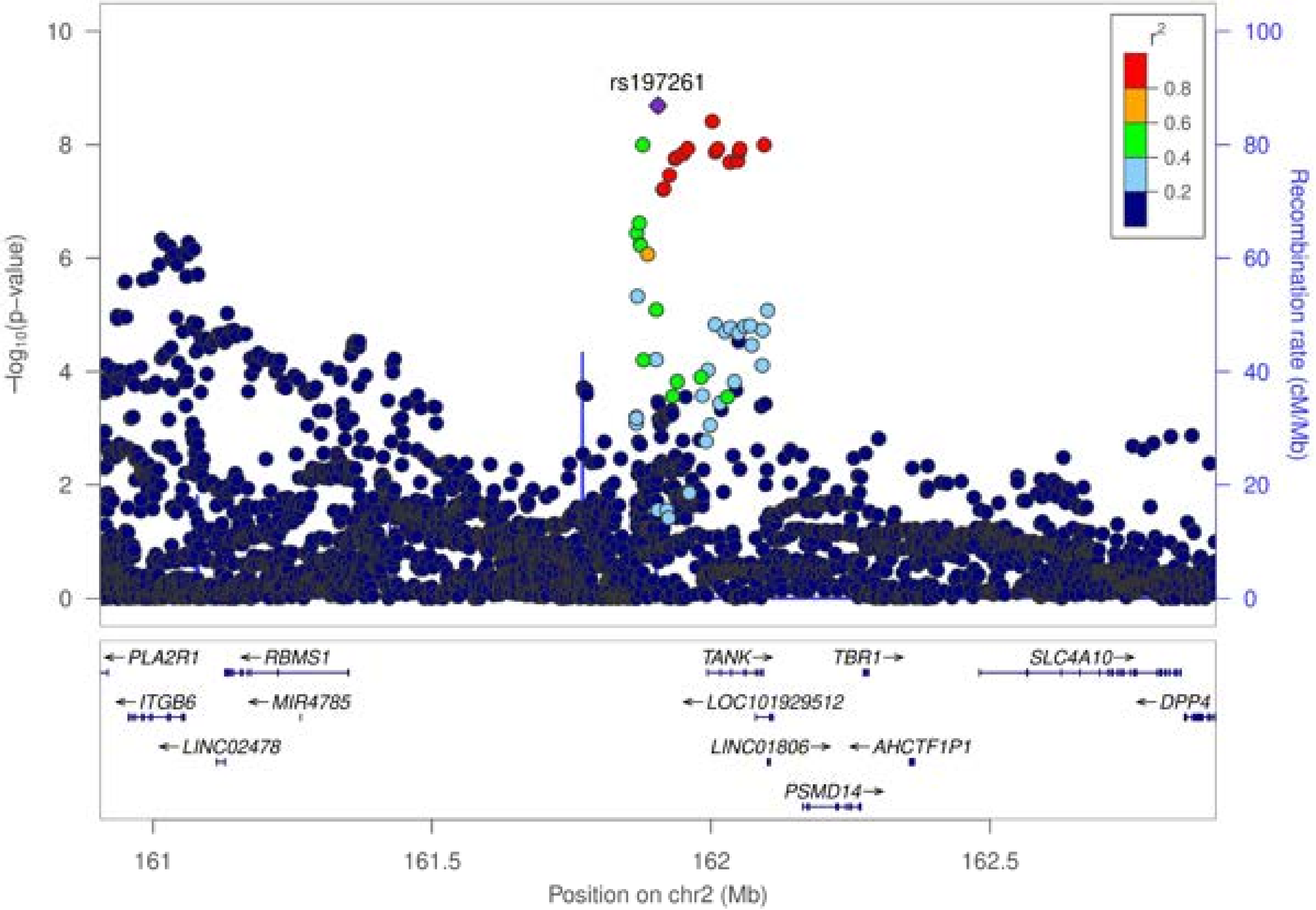

rs197261
r²
0.8
0.6
0.4
0.2
−log₁₀(p−value)
Recombination rate (cM/Mb)
PLA2R1
RBMS1
ITGB6
MIR4785
LINC02478
TANK
TBR1
SLC4A10
LOC101929512
DPP4
LINC01806
AHCTF1P1
PSMD14
161
161.5
162
162.5
Position on chr2 (Mb)

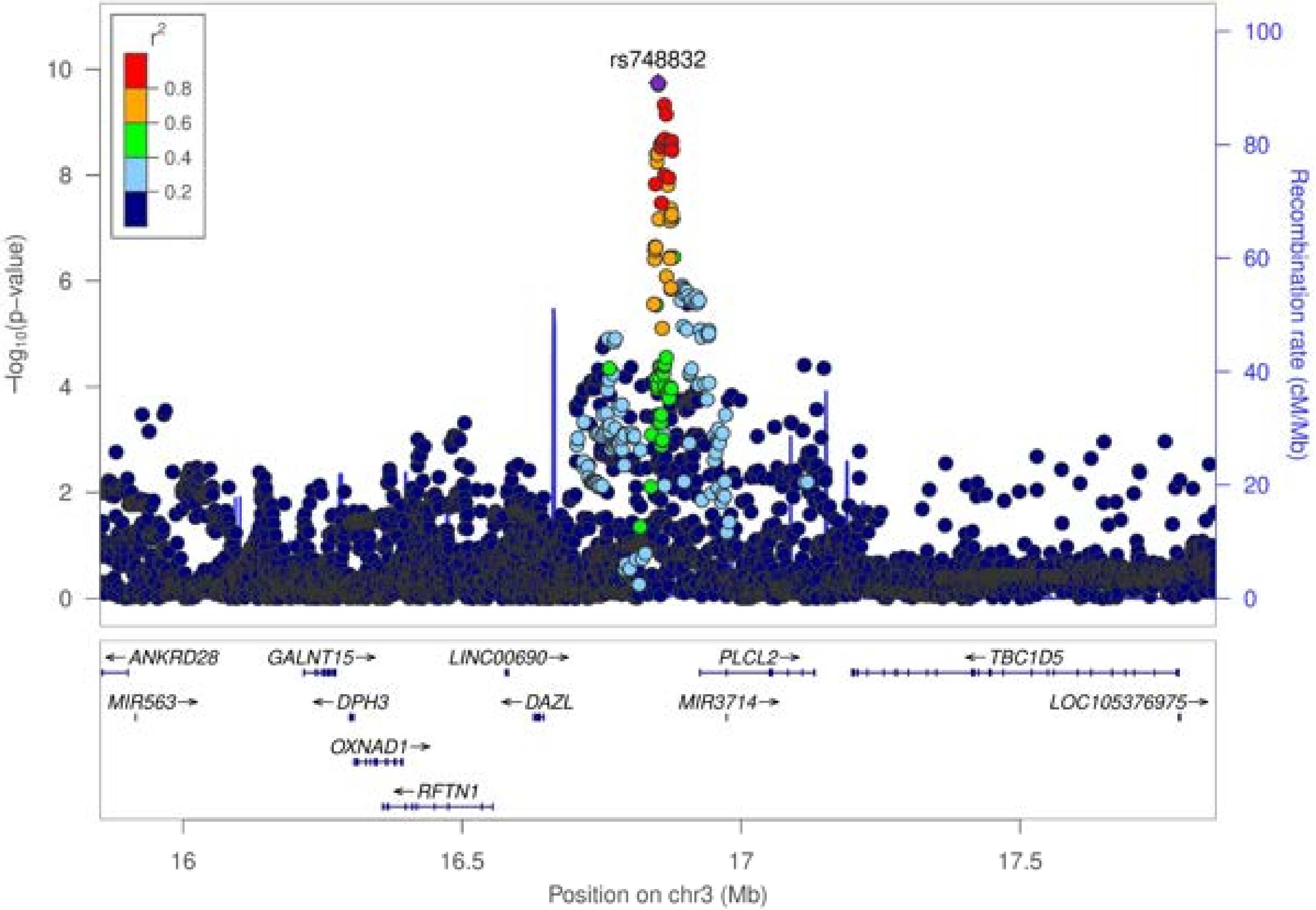

rs748832
$r^2$
0.8
0.6
0.4
0.2
−log10(p−value)
Recombination rate (cM/Mb)
10
8
6
4
2
0
100
80
60
40
20
0
← ANKRD28
GALNT15 →
LINC00690 →
PLCL2 →
← TBC1D5
MIR563 →
← DPH3
← DAZL
MIR3714 →
LOC105376975 →
OXNAD1 →
← RFTN1
16
16.5
17
17.5
Position on chr3 (Mb)

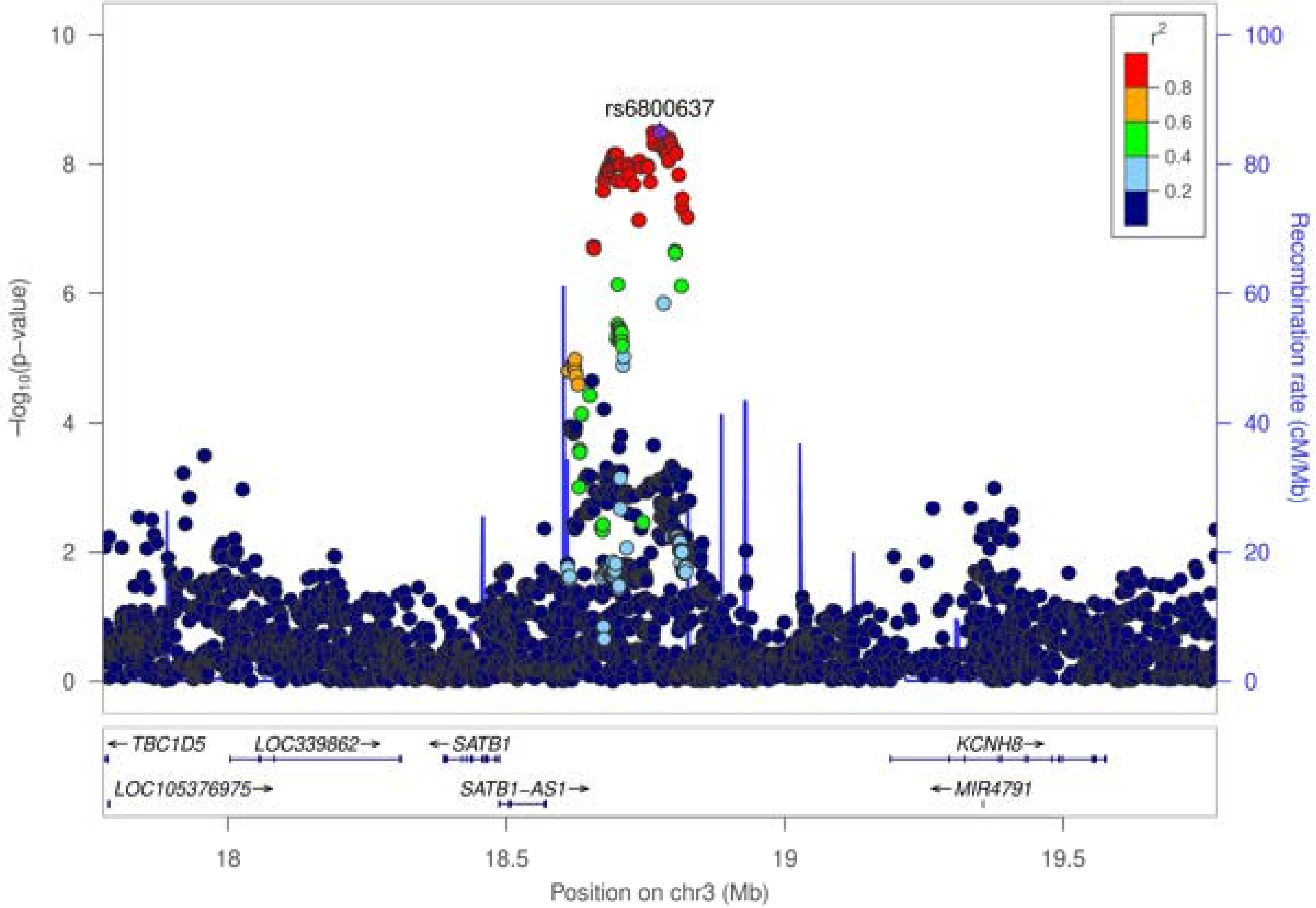

rs6800637
r2
0.8
0.6
0.4
0.2
−log10(p−value)
Recombination rate (cM/Mb)
10
8
6
4
2
0
100
80
60
40
20
0
← TBC1D5
LOC339862→
← SATB1
KCNH8→
LOC105376975→
SATB1−AS1→
← MIR4791
18
18.5
19
19.5
Position on chr3 (Mb)

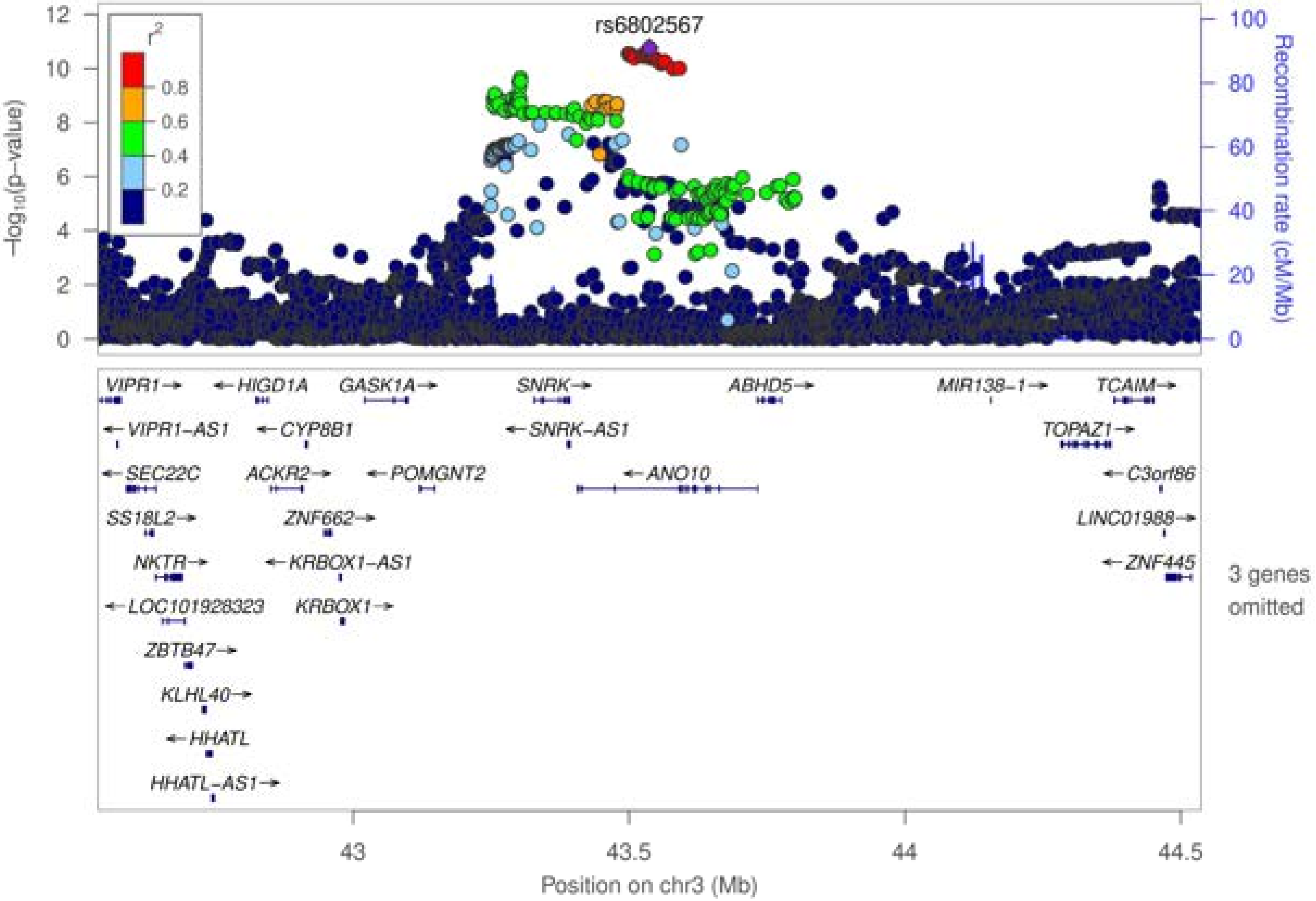
rs6802567
−log10(p−value)
Recombination rate (cM/Mb)
r2
0.8
0.6
0.4
0.2
VIPR1→
←HIGD1A
GASK1A→
SNRK→
ABHD5→
MIR138−1→
TCAIM→
←VIPR1−AS1
←CYP8B1
←SNRK−AS1
TOPAZ1→
←SEC22C
ACKR2→
←POMGNT2
←ANO10
←C3orf86
SS18L2→
ZNF662→
LINC01988→
NKTR→
←KRBOX1−AS1
←ZNF445
3 genes omitted
←LOC101928323
KRBOX1→
ZBTB47→
KLHL40→
←HHATL
HHATL−AS1→
Position on chr3 (Mb)

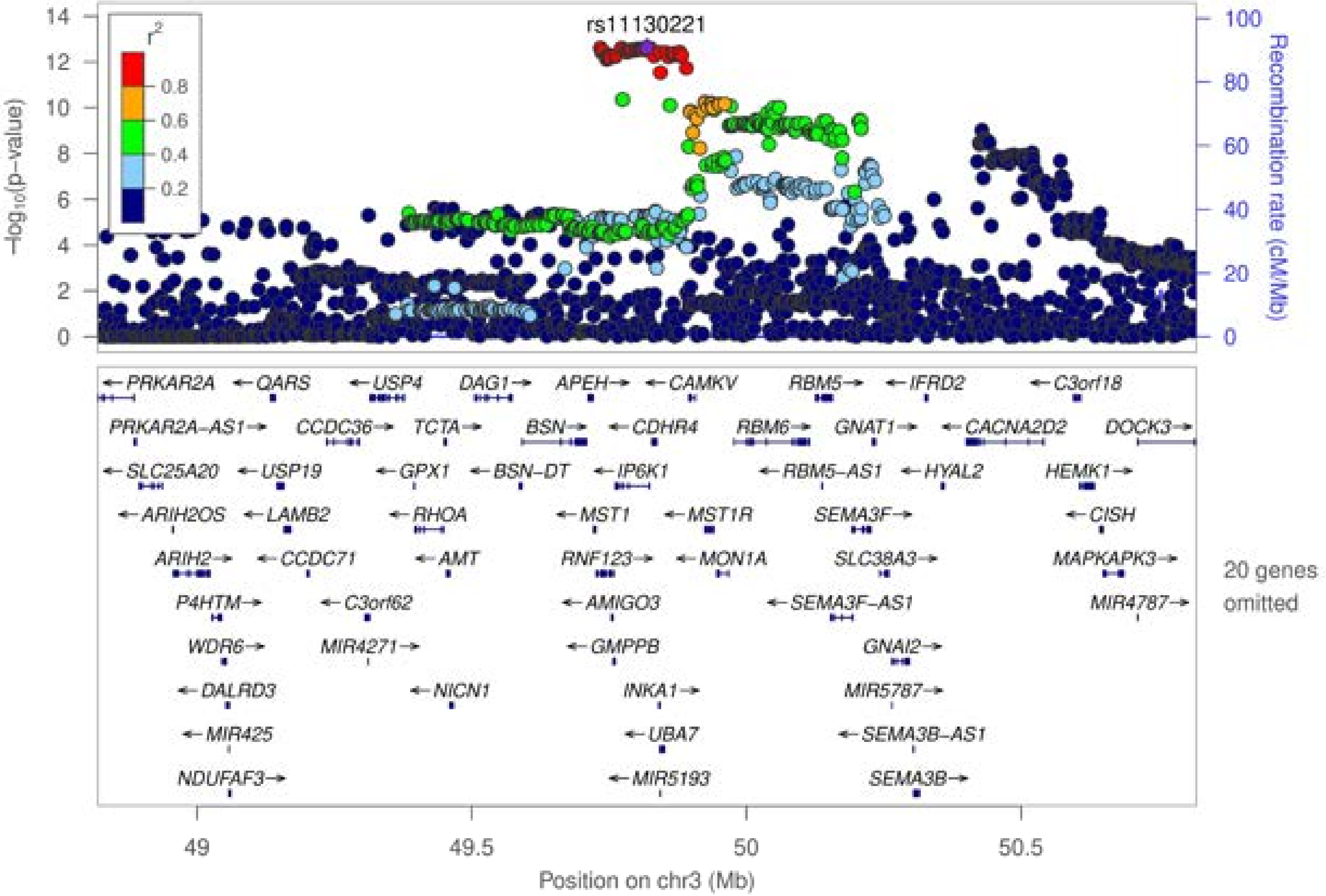
rs11130221
r²
0.8
0.6
0.4
0.2
−log10(p−value)
Recombination rate (cM/Mb)
PRKAR2A
QARS
USP4
DAG1
APEH
CAMKV
RBM5
IFRD2
C3orf18
PRKAR2A−AS1
CCDC36
TCTA
BSN
CDHR4
RBM6
GNAT1
CACNA2D2
DOCK3
SLC25A20
USP19
GPX1
BSN−DT
IP6K1
RBM5−AS1
HYAL2
HEMK1
ARIH2OS
LAMB2
RHOA
MST1
MST1R
SEMA3F
CISH
ARIH2
CCDC71
AMT
RNF123
MON1A
SLC38A3
MAPKAPK3
20 genes omitted
P4HTM
C3orf62
AMIGO3
SEMA3F−AS1
MIR4787
WDR6
MIR4271
GMPPB
GNAI2
DALRD3
NICN1
INKA1
MIR5787
MIR425
UBA7
SEMA3B−AS1
NDUFAF3
MIR5193
SEMA3B
Position on chr3 (Mb)

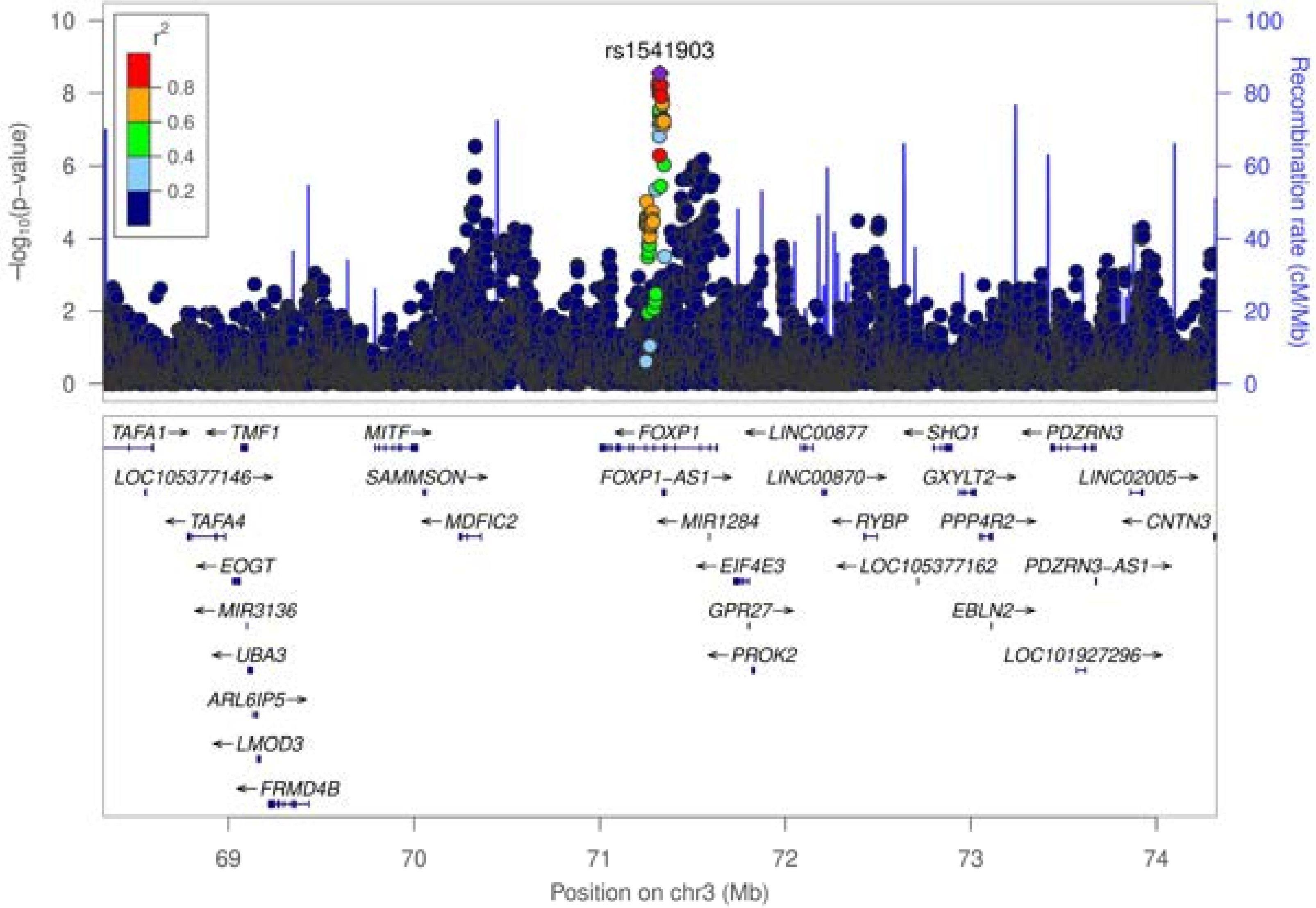
rs1541903
r²
0.8
0.6
0.4
0.2
−log10(p−value)
Recombination rate (cM/Mb)
TAFA1→
←TMF1
MITF→
←FOXP1
←LINC00877
←SHQ1
←PDZRN3
LOC105377146→
SAMMSON→
FOXP1−AS1→
LINC00870→
GXYLT2→
LINC02005→
←TAFA4
←MDFIC2
←MIR1284
←RYBP
PPP4R2→
←CNTN3
←EOGT
←EIF4E3
←LOC105377162
PDZRN3−AS1→
←MIR3136
GPR27→
EBLN2→
←UBA3
←PROK2
LOC101927296→
ARL6IP5→
←LMOD3
←FRMD4B
Position on chr3 (Mb)

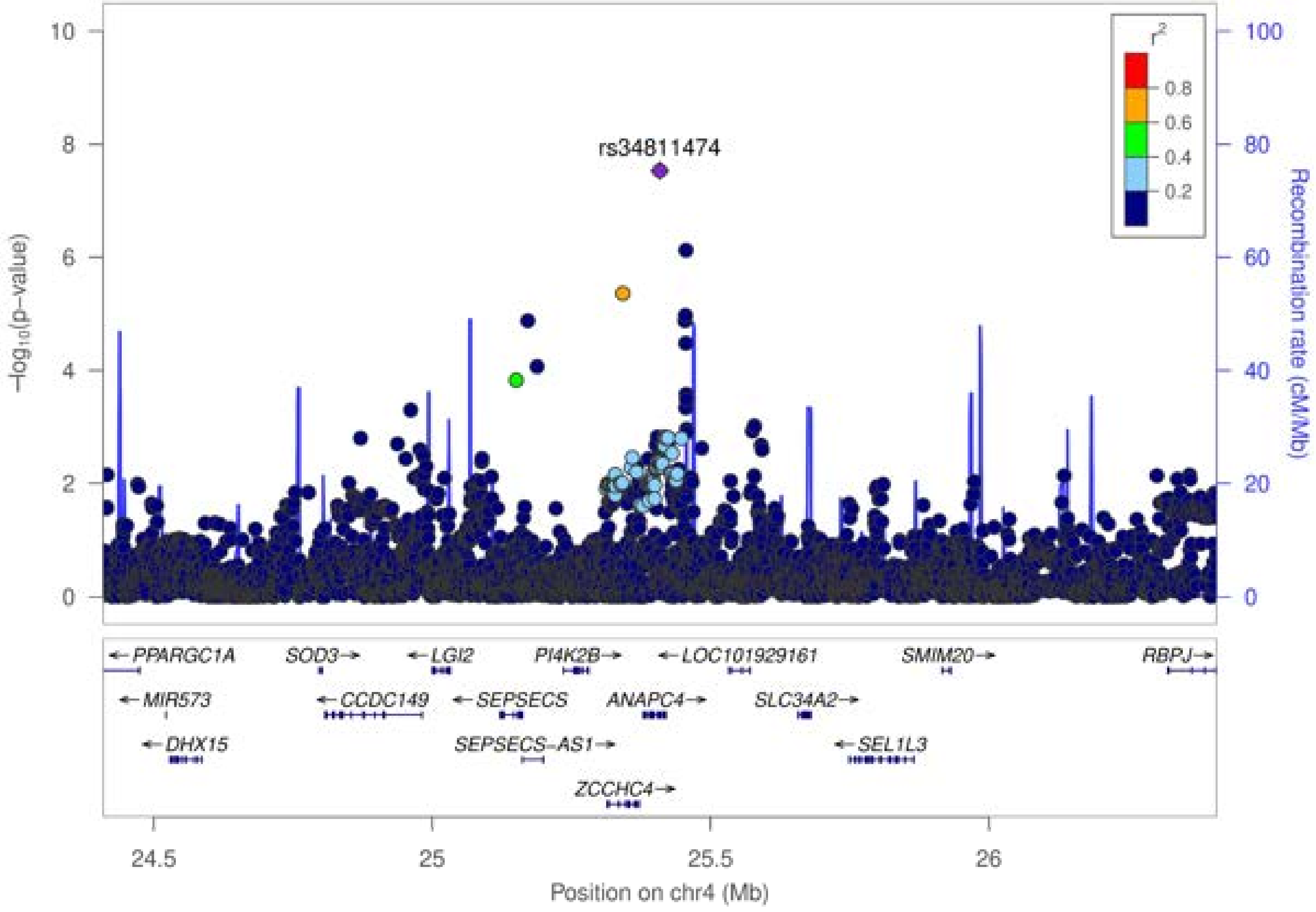

rs34811474
−log10(p−value)
Recombination rate (cM/Mb)
r2
0.8
0.6
0.4
0.2
10
8
6
4
2
0
100
80
60
40
20
0
PPARGC1A
MIR573
DHX15
SOD3
CCDC149
LGI2
SEPSECS
SEPSECS−AS1
PI4K2B
ANAPC4
ZCCHC4
LOC101929161
SLC34A2
SEL1L3
SMIM20
RBPJ
24.5
25
25.5
26
Position on chr4 (Mb)

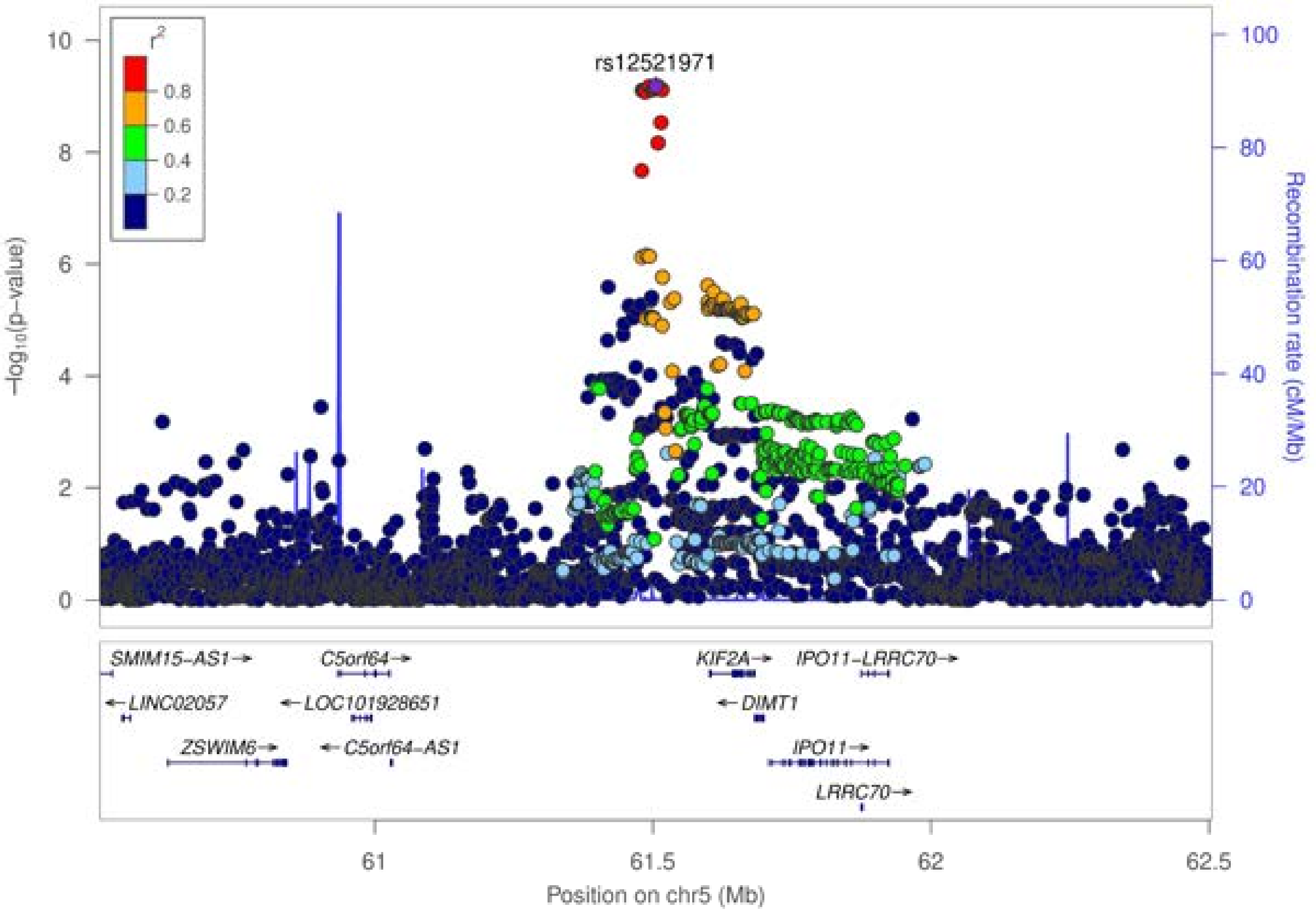
rs12521971
r²
0.8
0.6
0.4
0.2
10
8
6
4
2
0
−log₁₀(p−value)
100
80
60
40
20
0
Recombination rate (cM/Mb)
SMIM15−AS1→
C5orf64→
KIF2A→
IPO11−LRRC70→
←LINC02057
←LOC101928651
←DIMT1
ZSWIM6→
←C5orf64−AS1
IPO11→
LRRC70→
61
61.5
62
62.5
Position on chr5 (Mb)

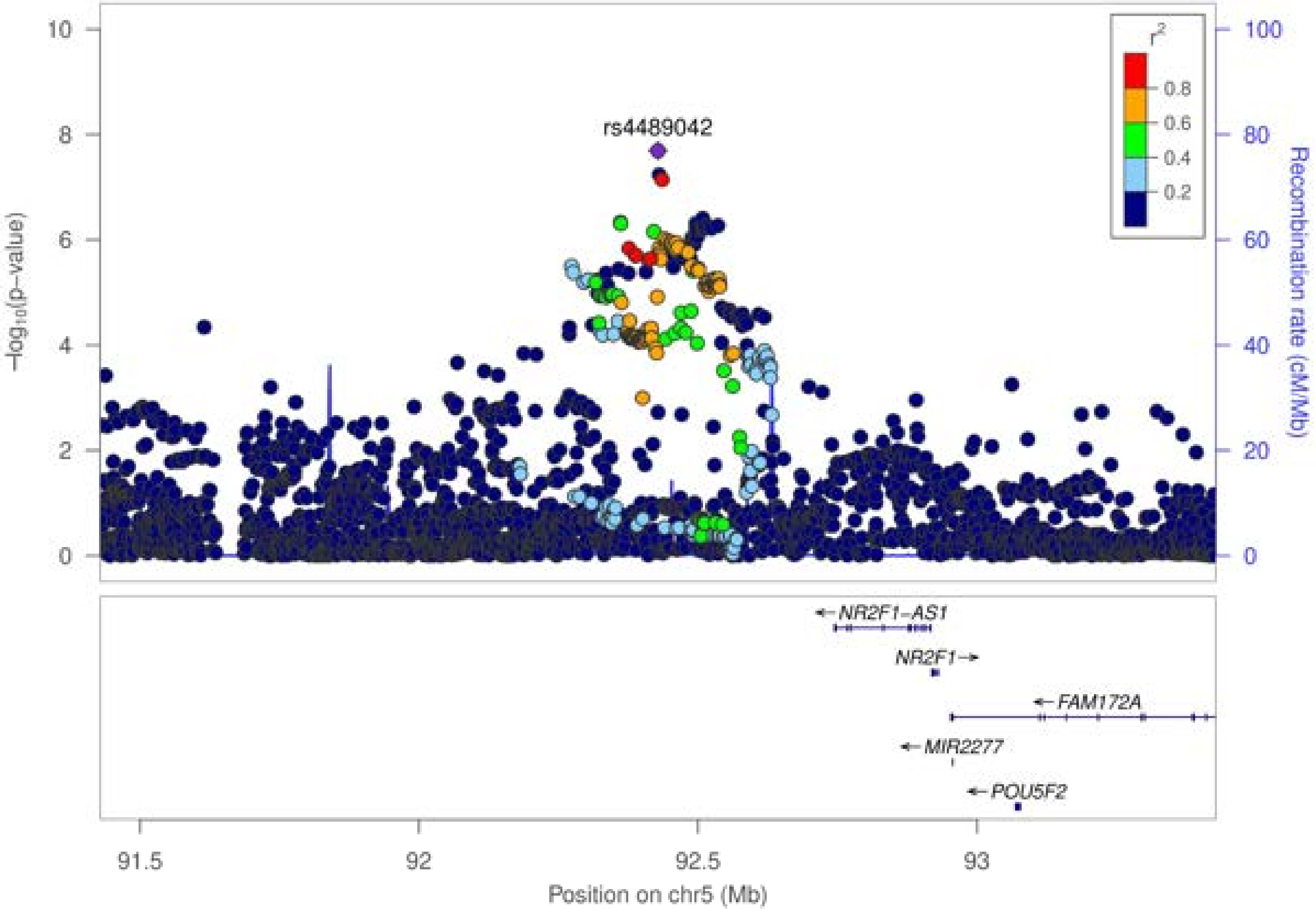

rs4489042
r2
0.8
0.6
0.4
0.2
−log10(p−value)
Recombination rate (cM/Mb)
0
2
4
6
8
10
0
20
40
60
80
100
NR2F1−AS1
NR2F1
FAM172A
MIR2277
POU5F2
91.5
92
92.5
93
Position on chr5 (Mb)

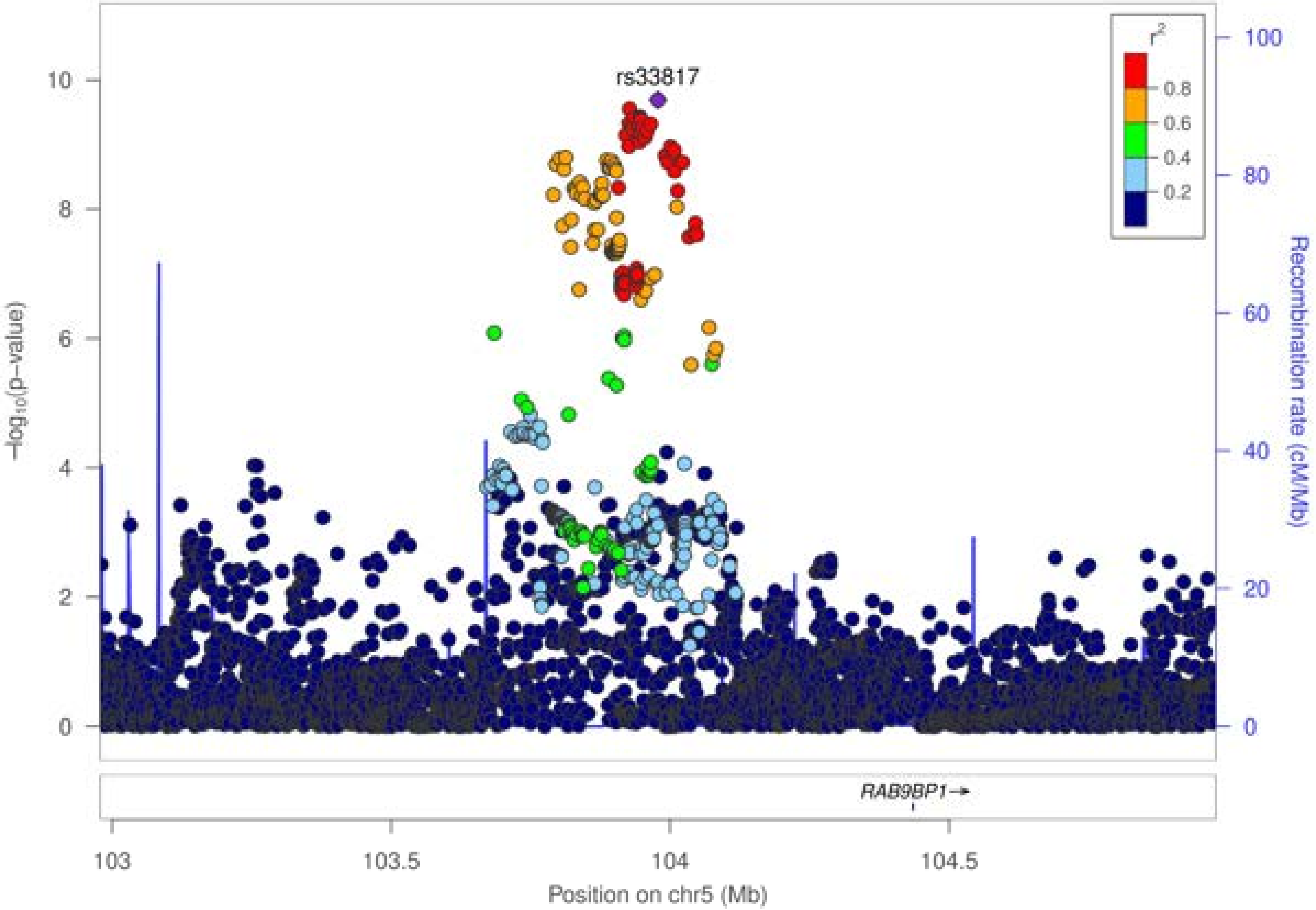

rs33817
r²
0.8
0.6
0.4
0.2
−log₁₀(p−value)
Recombination rate (cM/Mb)
RAB9BP1→
103
103.5
104
104.5
Position on chr5 (Mb)

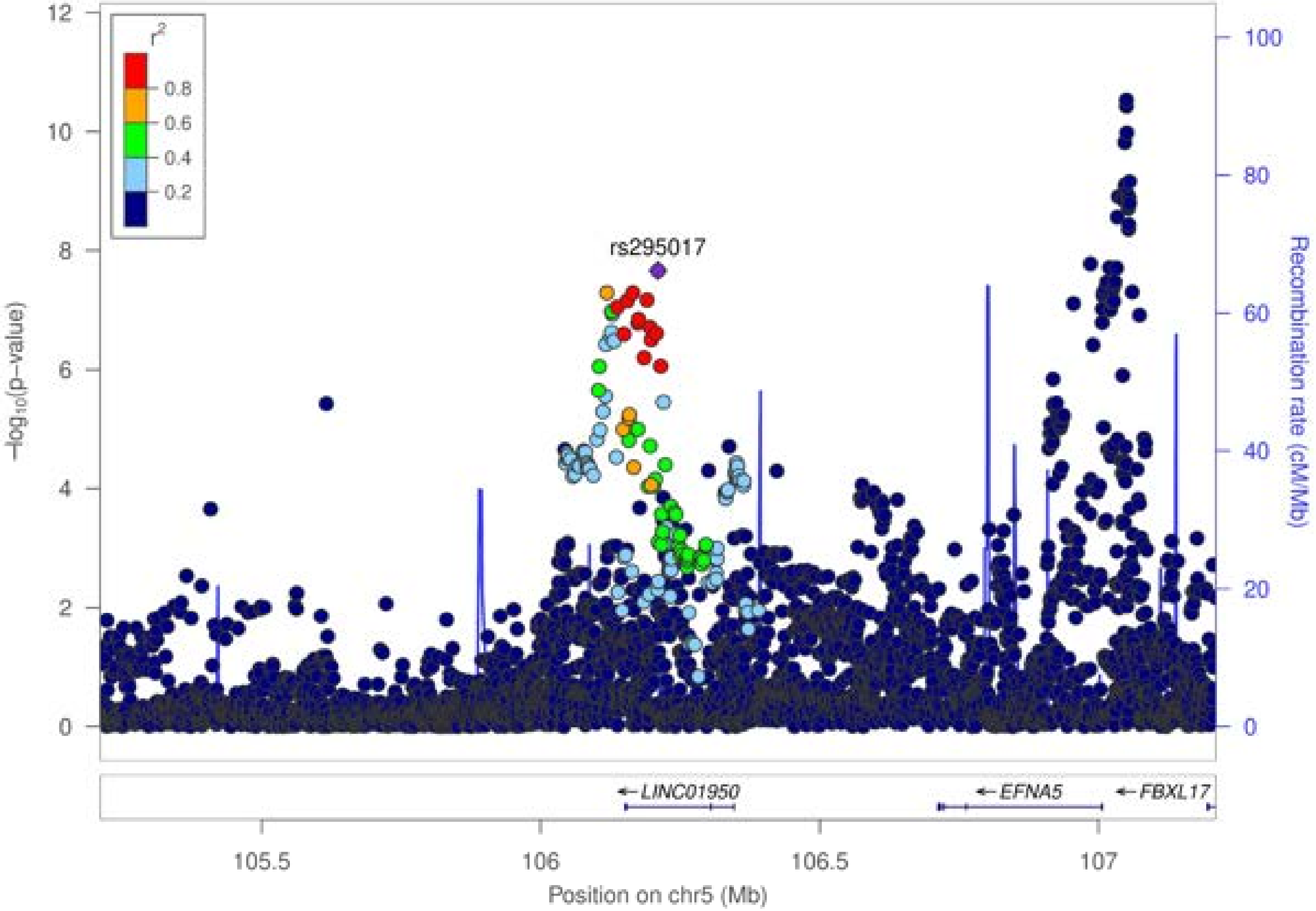

rs295017
r²
0.8
0.6
0.4
0.2
−log10(p−value)
Recombination rate (cM/Mb)
LINC01950
EFNA5
FBXL17
Position on chr5 (Mb)

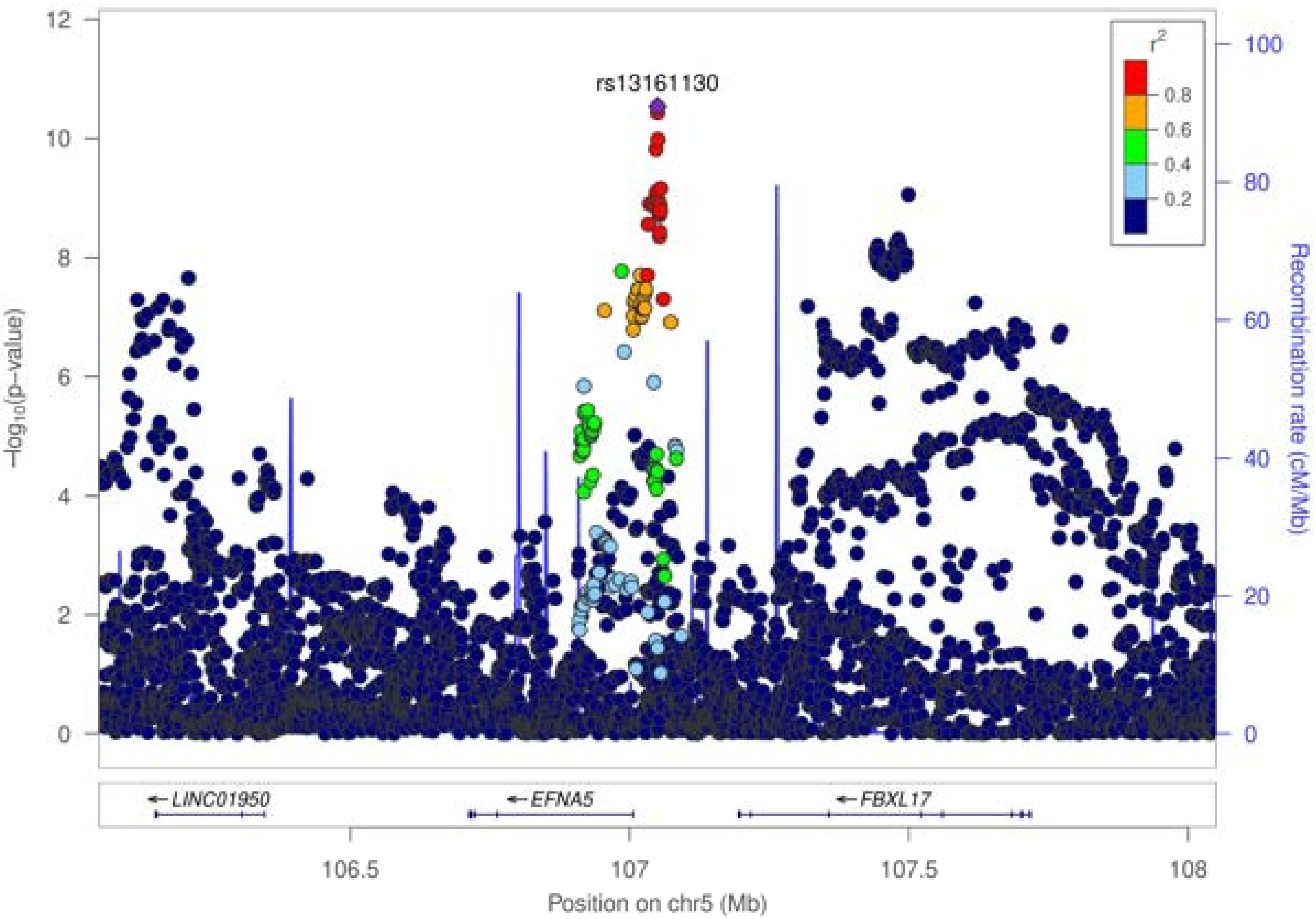

rs13161130
r²
0.8
0.6
0.4
0.2
−log₁₀(p−value)
Recombination rate (cM/Mb)
← LINC01950
← EFNA5
← FBXL17
106.5
107
107.5
108
Position on chr5 (Mb)

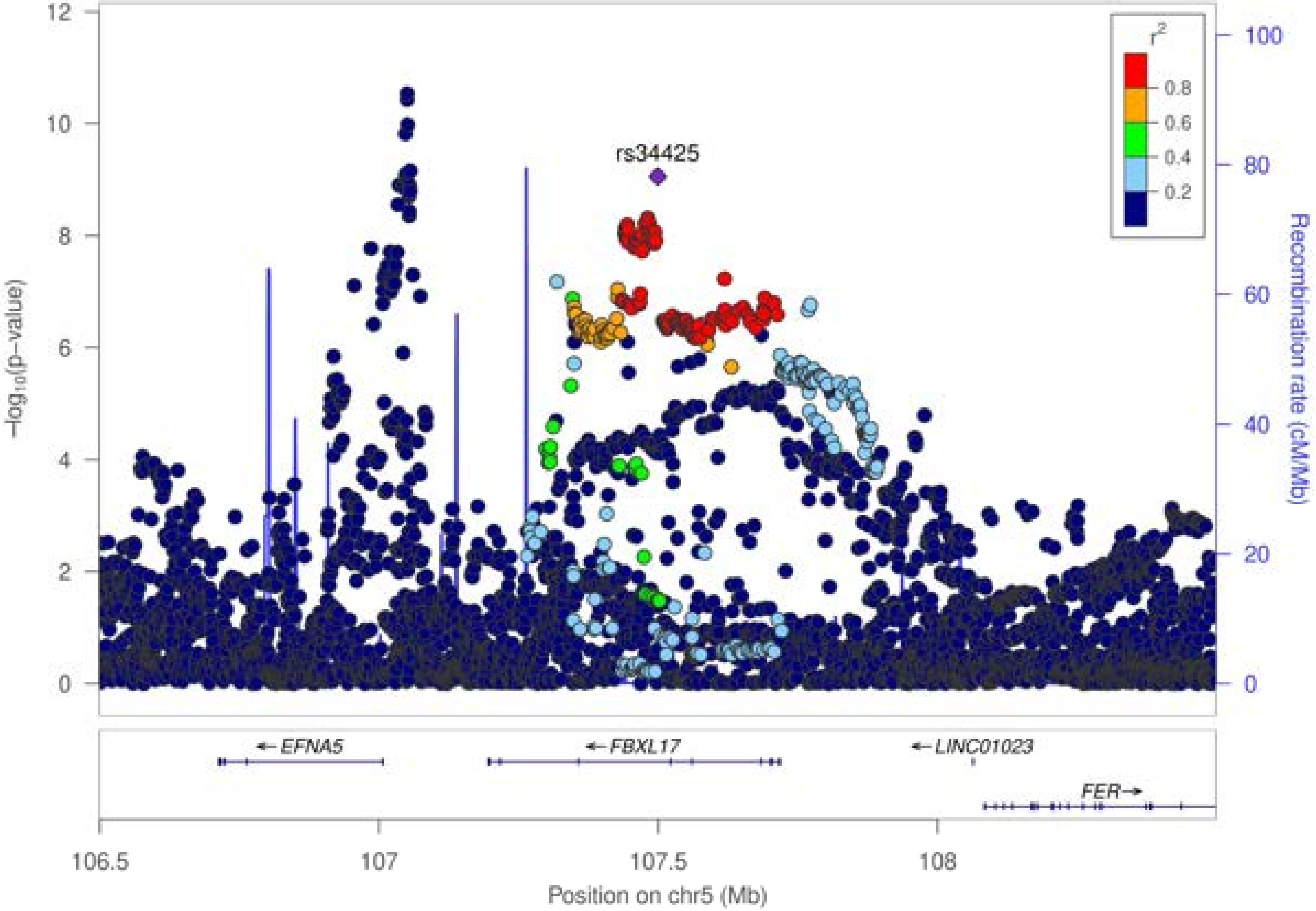

rs34425
r2
0.8
0.6
0.4
0.2
−log10(p−value)
Recombination rate (cM/Mb)
12
10
8
6
4
2
0
100
80
60
40
20
0
EFNA5
FBXL17
LINC01023
FER
106.5
107
107.5
108
Position on chr5 (Mb)

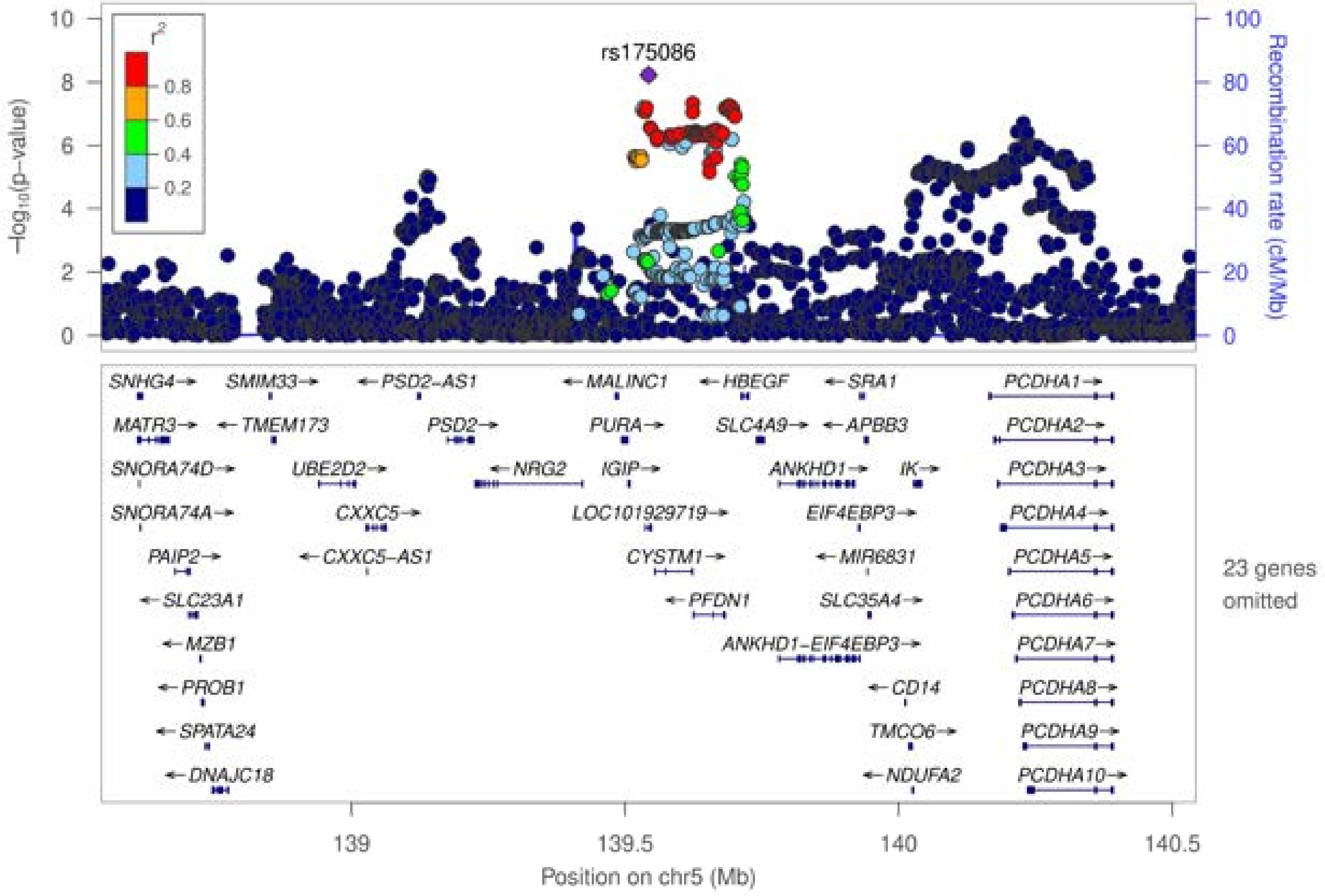
rs175086
r2
0.8
0.6
0.4
0.2
−log10(p−value)
Recombination rate (cM/Mb)
SNHG4→
SMIM33→
←PSD2-AS1
←MALINC1
←HBEGF
←SRA1
PCDHA1→
MATR3→
←TMEM173
PSD2→
PURA→
SLC4A9→
←APBB3
PCDHA2→
SNORA74D→
UBE2D2→
←NRG2
IGIP→
ANKHD1→
IK→
PCDHA3→
SNORA74A→
CXXC5→
LOC101929719→
EIF4EBP3→
PCDHA4→
PAIP2→
←CXXC5-AS1
CYSTM1→
←MIR6831
PCDHA5→
←SLC23A1
←PFDN1
SLC35A4→
PCDHA6→
←MZB1
ANKHD1-EIF4EBP3→
PCDHA7→
←PROB1
←CD14
PCDHA8→
←SPATA24
TMCO6→
PCDHA9→
←DNAJC18
←NDUFA2
PCDHA10→
23 genes omitted
139
139.5
140
140.5
Position on chr5 (Mb)

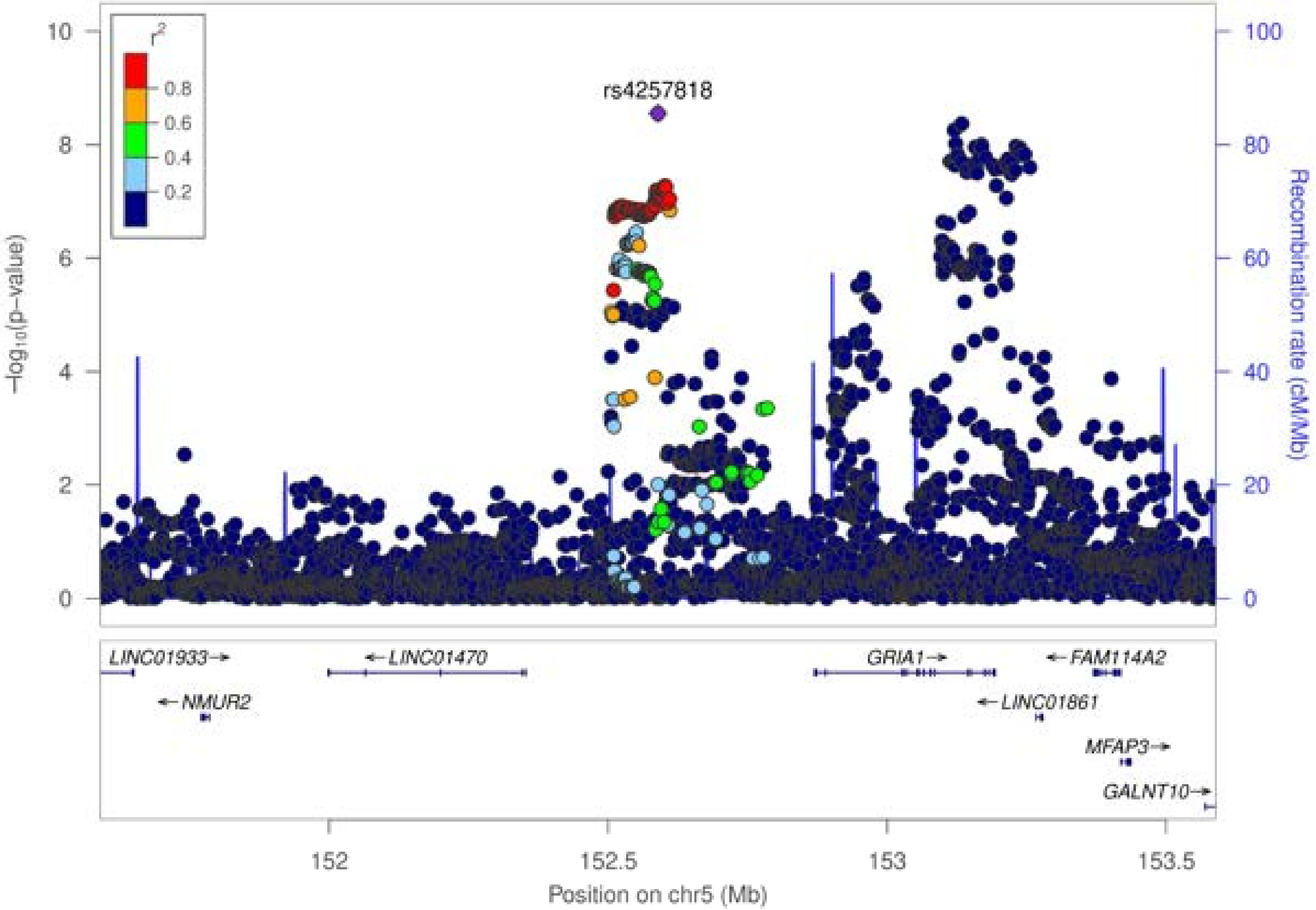

rs4257818
r²
0.8
0.6
0.4
0.2
−log10(p−value)
Recombination rate (cM/Mb)
LINC01933→
←LINC01470
GRIA1→
←FAM114A2
←NMUR2
←LINC01861
MFAP3→
GALNT10→
Position on chr5 (Mb)

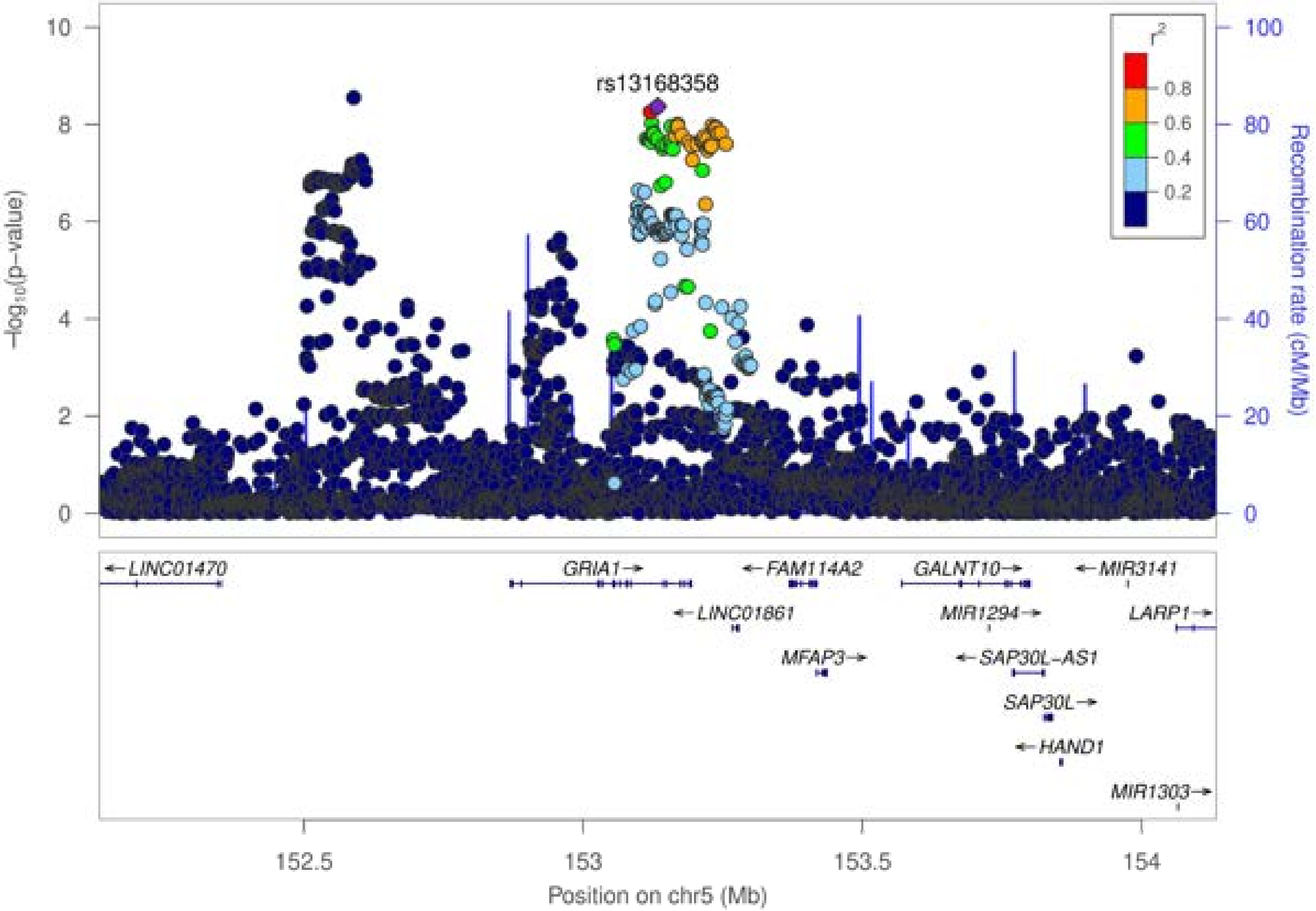

rs13168358
r2
0.8
0.6
0.4
0.2
−log10(p−value)
Recombination rate (cM/Mb)
10
8
6
4
2
0
100
80
60
40
20
0
← LINC01470
GRIA1 →
← FAM114A2
GALNT10 →
← MIR3141
← LINC01861
MIR1294 →
LARP1 →
MFAP3 →
← SAP30L-AS1
SAP30L →
← HAND1
MIR1303 →
152.5
153
153.5
154
Position on chr5 (Mb)

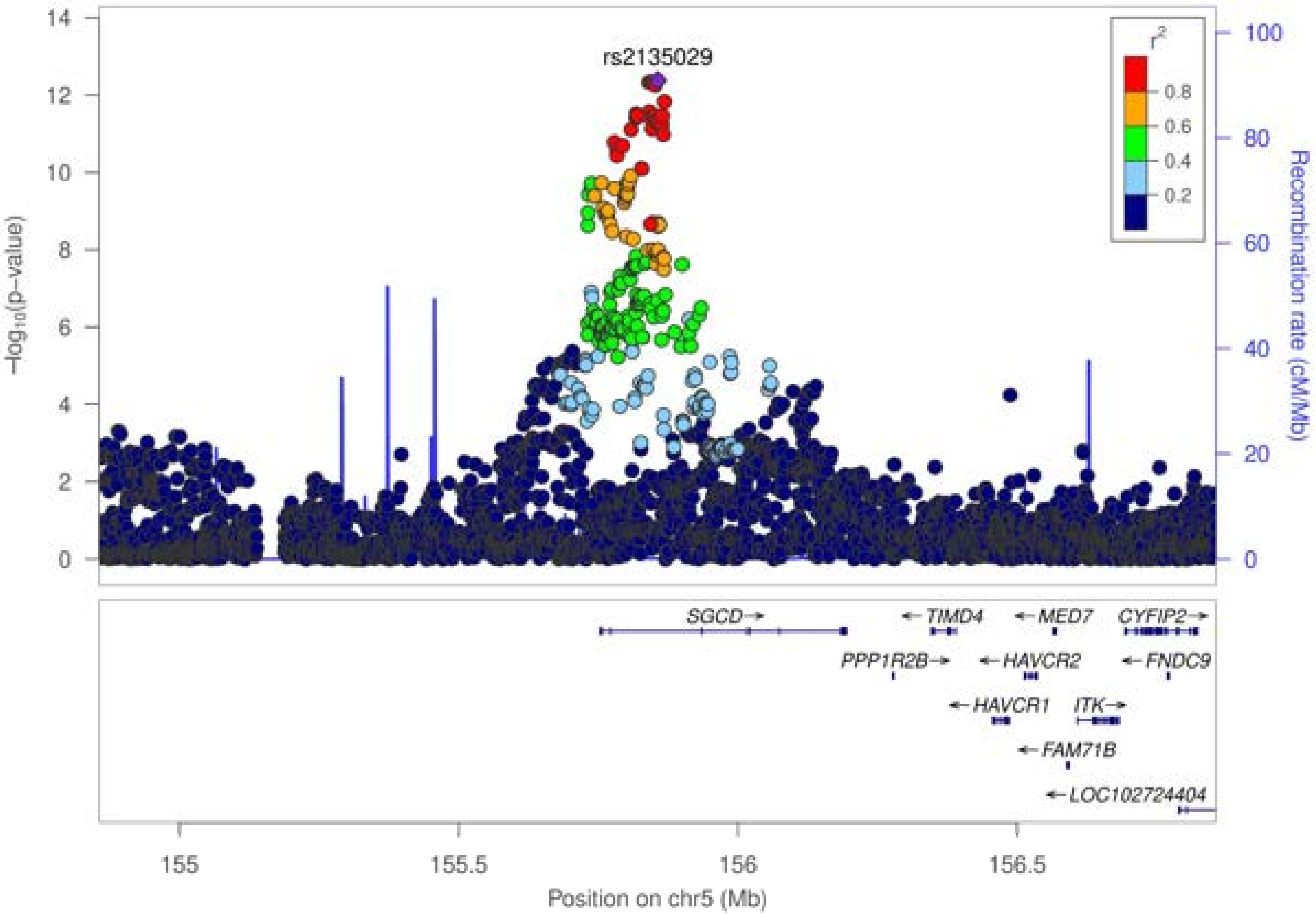

rs2135029
r²
0.8
0.6
0.4
0.2
−log10(p−value)
Recombination rate (cM/Mb)
SGCD
TIMD4
MED7
CYFIP2
PPP1R2B
HAVCR2
FNDC9
HAVCR1
ITK
FAM71B
LOC102724404
155
155.5
156
156.5
Position on chr5 (Mb)

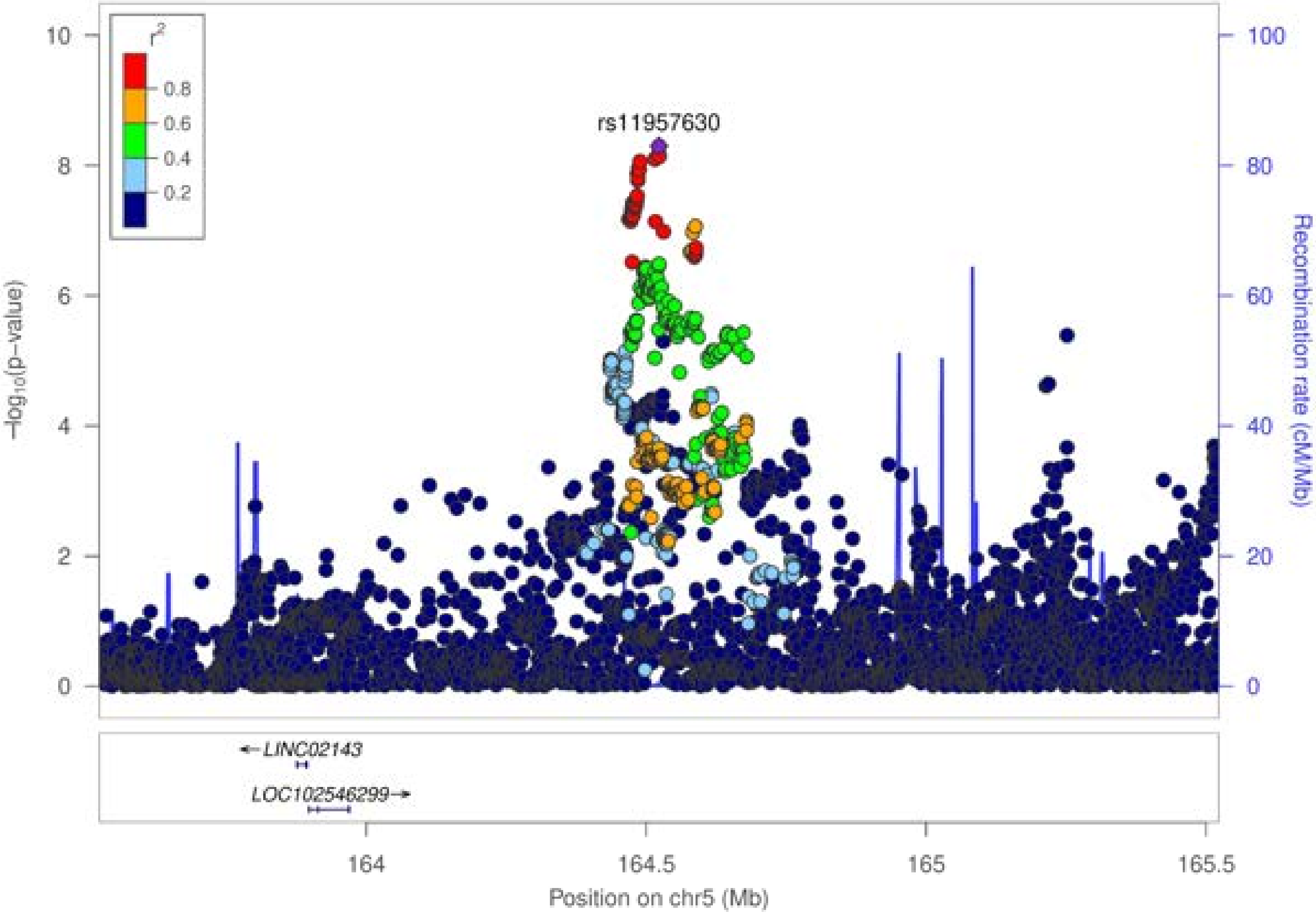
r²
0.8
0.6
0.4
0.2
rs11957630
−log10(p−value)
Recombination rate (cM/Mb)
LINC02143
LOC102546299
Position on chr5 (Mb)
164
164.5
165
165.5
0
2
4
6
8
10
20
40
60
80
100

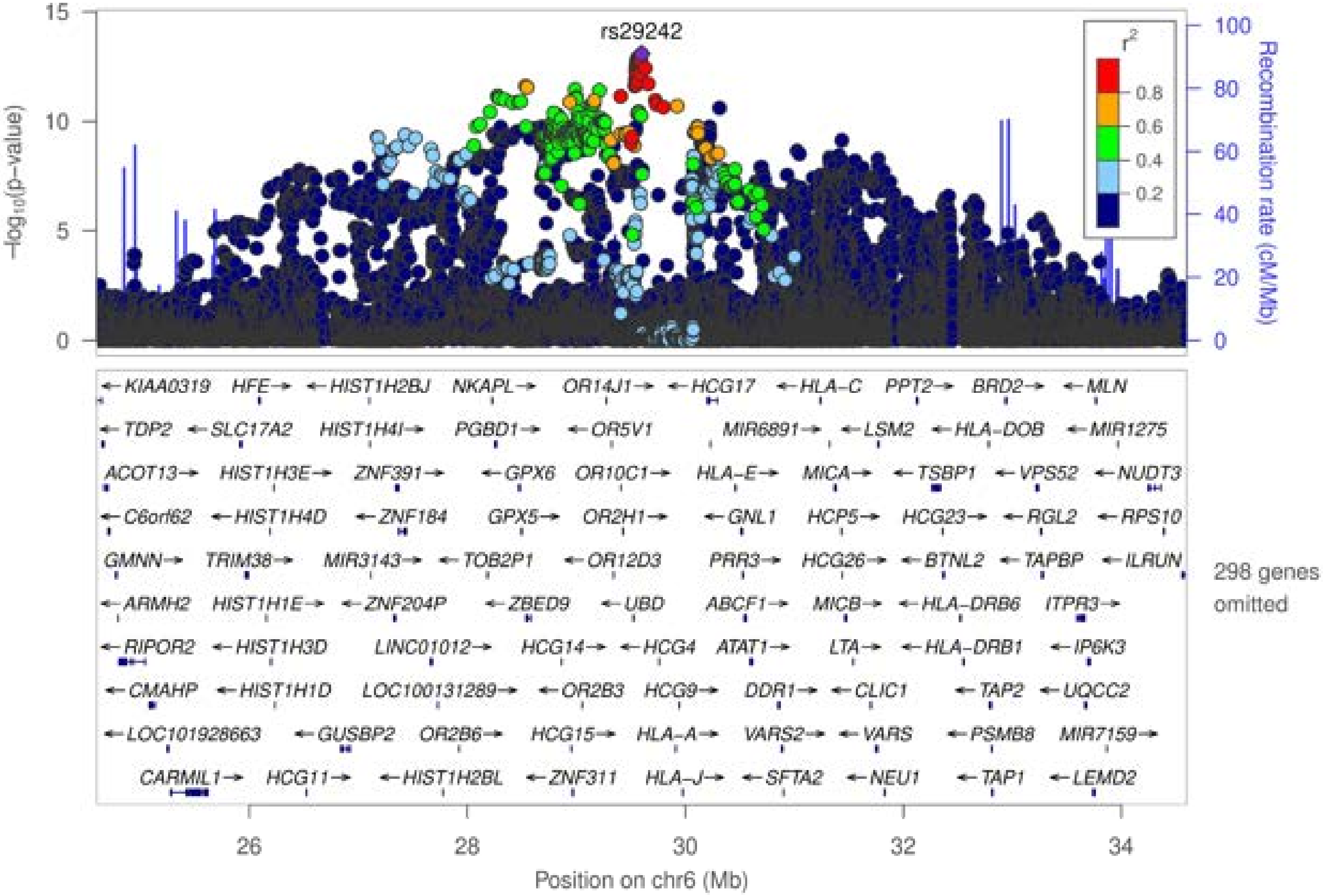

rs29242
−log10(p−value)
Recombination rate (cM/Mb)
r²
0.8
0.6
0.4
0.2
298 genes omitted
Position on chr6 (Mb)

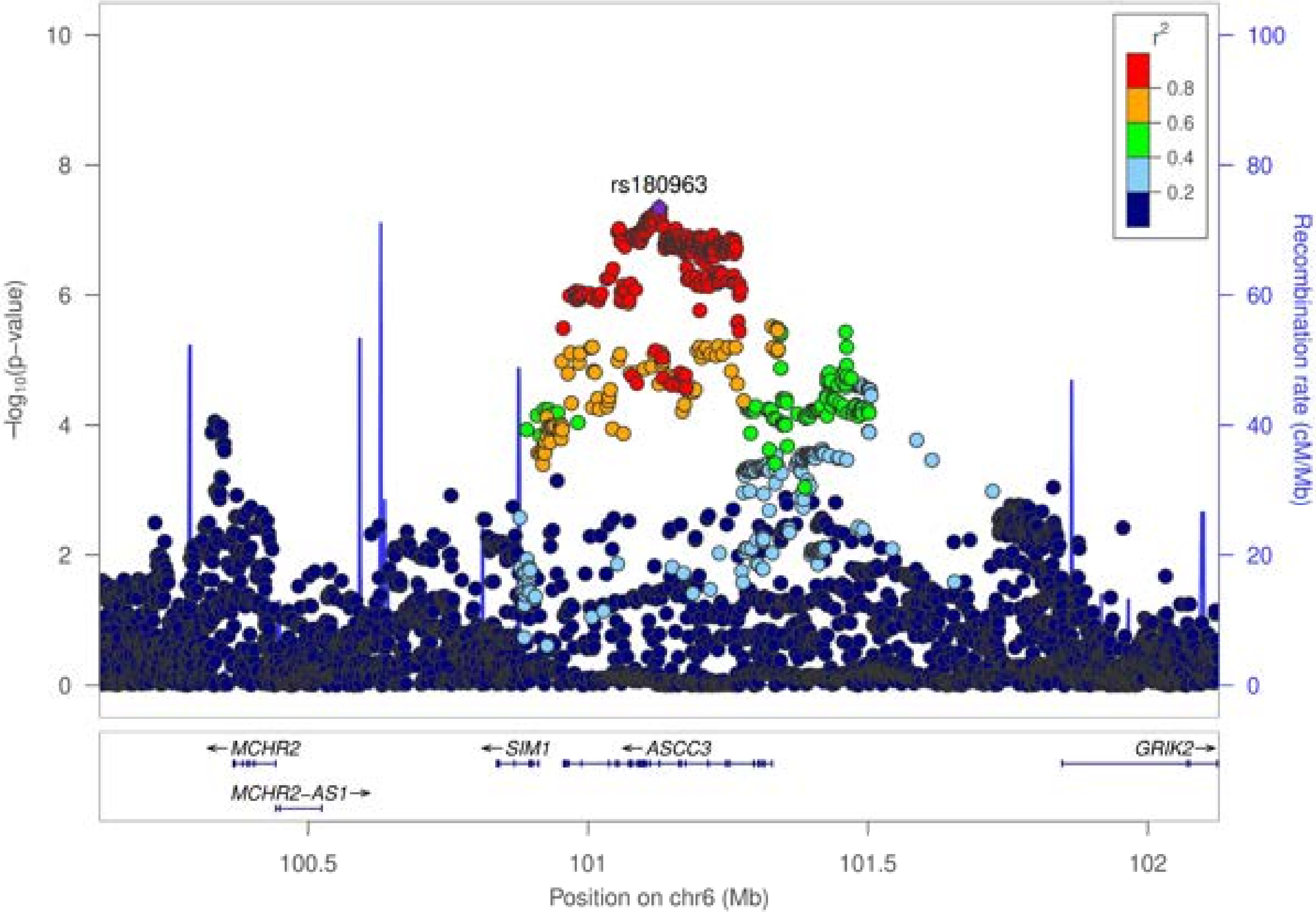

rs180963
r²
0.8
0.6
0.4
0.2
−log₁₀(p−value)
Recombination rate (cM/Mb)
0
2
4
6
8
10
0
20
40
60
80
100
←MCHR2
←SIM1
←ASCC3
GRIK2→
MCHR2−AS1→
100.5
101
101.5
102
Position on chr6 (Mb)

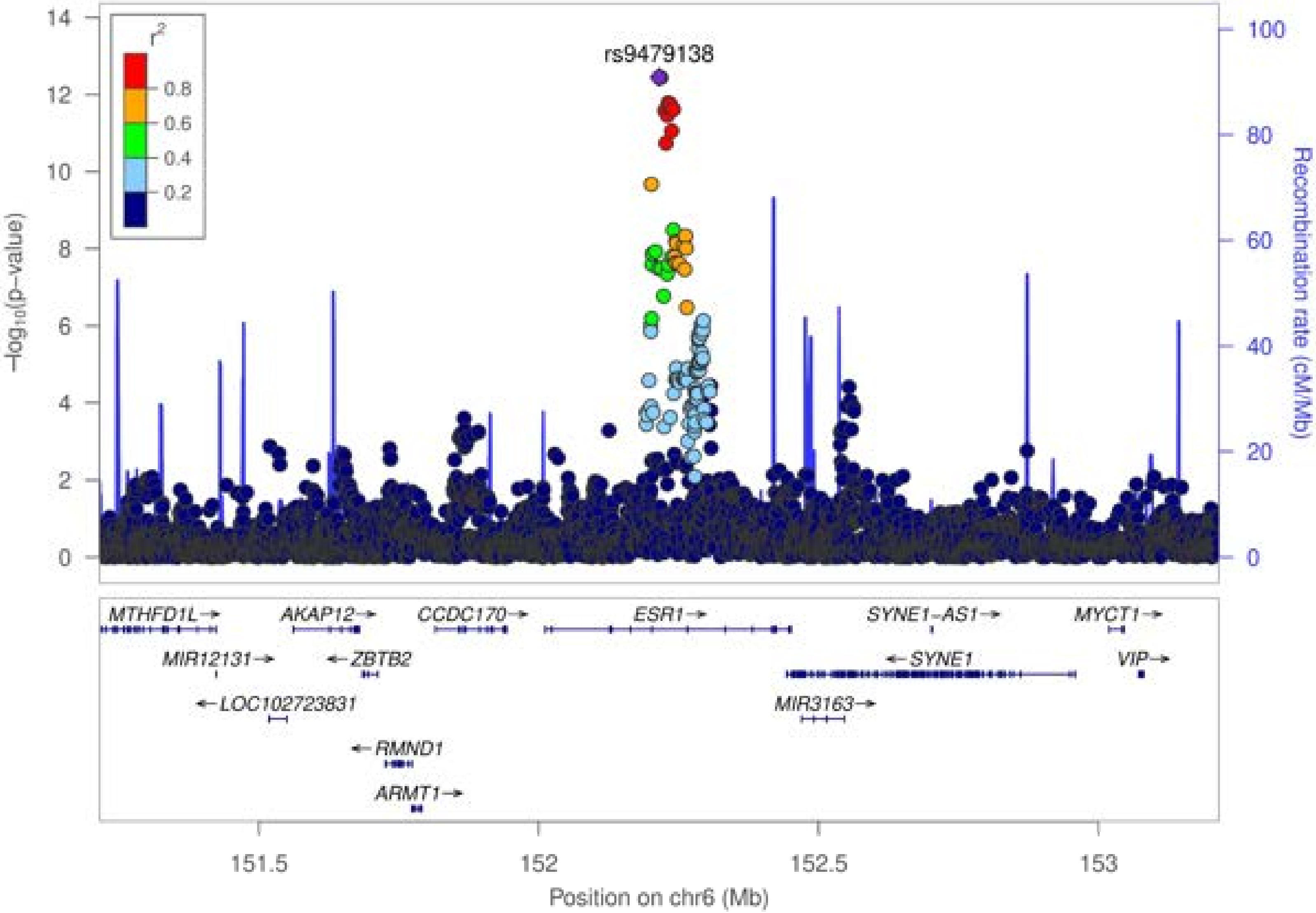

rs9479138
r²
0.8
0.6
0.4
0.2
−log₁₀(p−value)
Recombination rate (cM/Mb)
MTHFD1L→
AKAP12→
CCDC170→
ESR1→
SYNE1−AS1→
MYCT1→
MIR12131→
←ZBTB2
←SYNE1
VIP→
←LOC102723831
MIR3163→
←RMND1
ARMT1→
151.5
152
152.5
153
Position on chr6 (Mb)

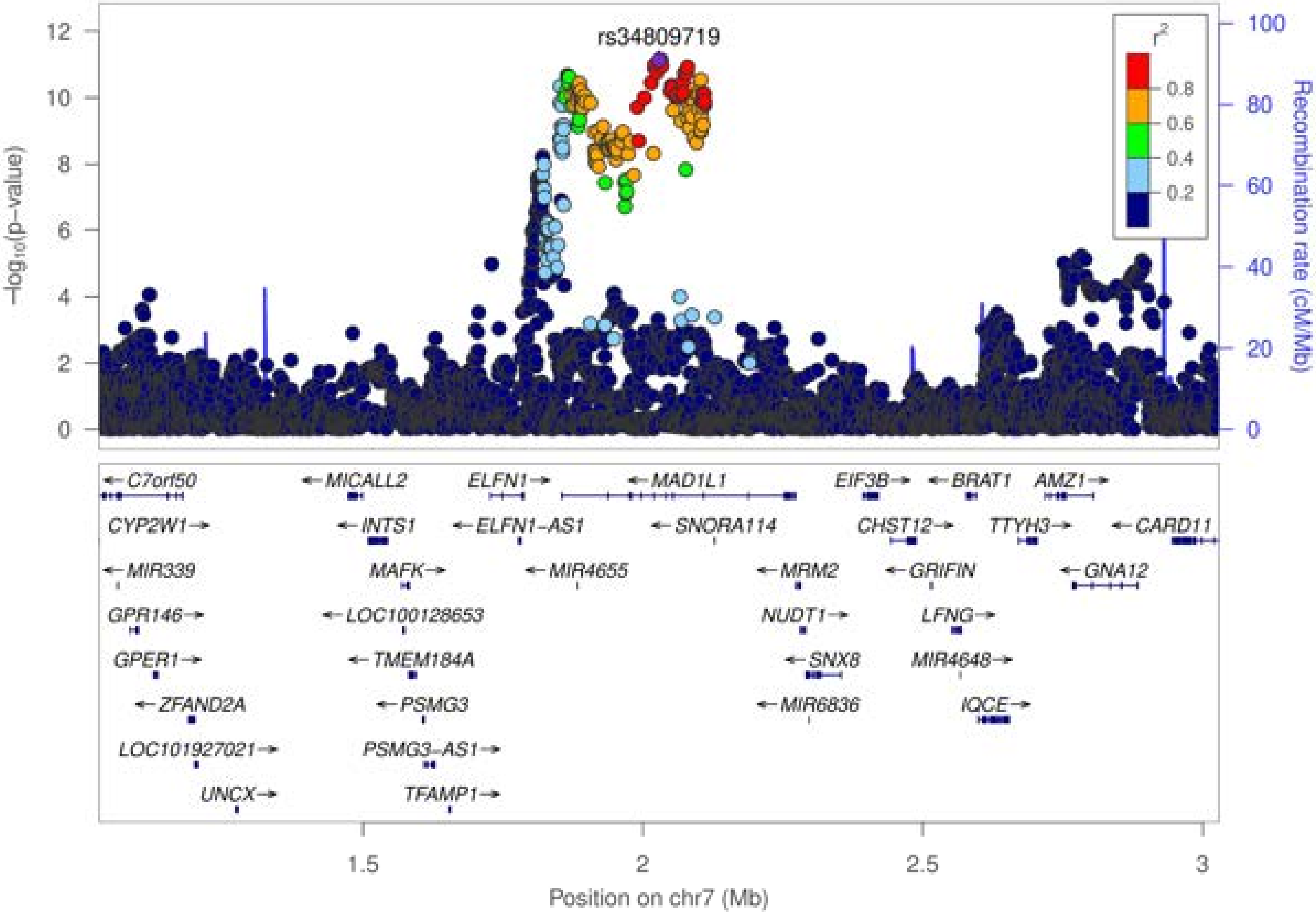
rs34809719
−log10(p−value)
Recombination rate (cM/Mb)
r2
0.8
0.6
0.4
0.2
C7orf50
MICALL2
ELFN1
MAD1L1
EIF3B
BRAT1
AMZ1
CYP2W1
INTS1
ELFN1-AS1
SNORA114
CHST12
TTYH3
CARD11
MIR339
MAFK
MIR4655
MRM2
GRIFIN
GNA12
GPR146
LOC100128653
NUDT1
LFNG
GPER1
TMEM184A
SNX8
MIR4648
ZFAND2A
PSMG3
MIR6836
IQCE
LOC101927021
PSMG3-AS1
UNCX
TFAMP1
Position on chr7 (Mb)

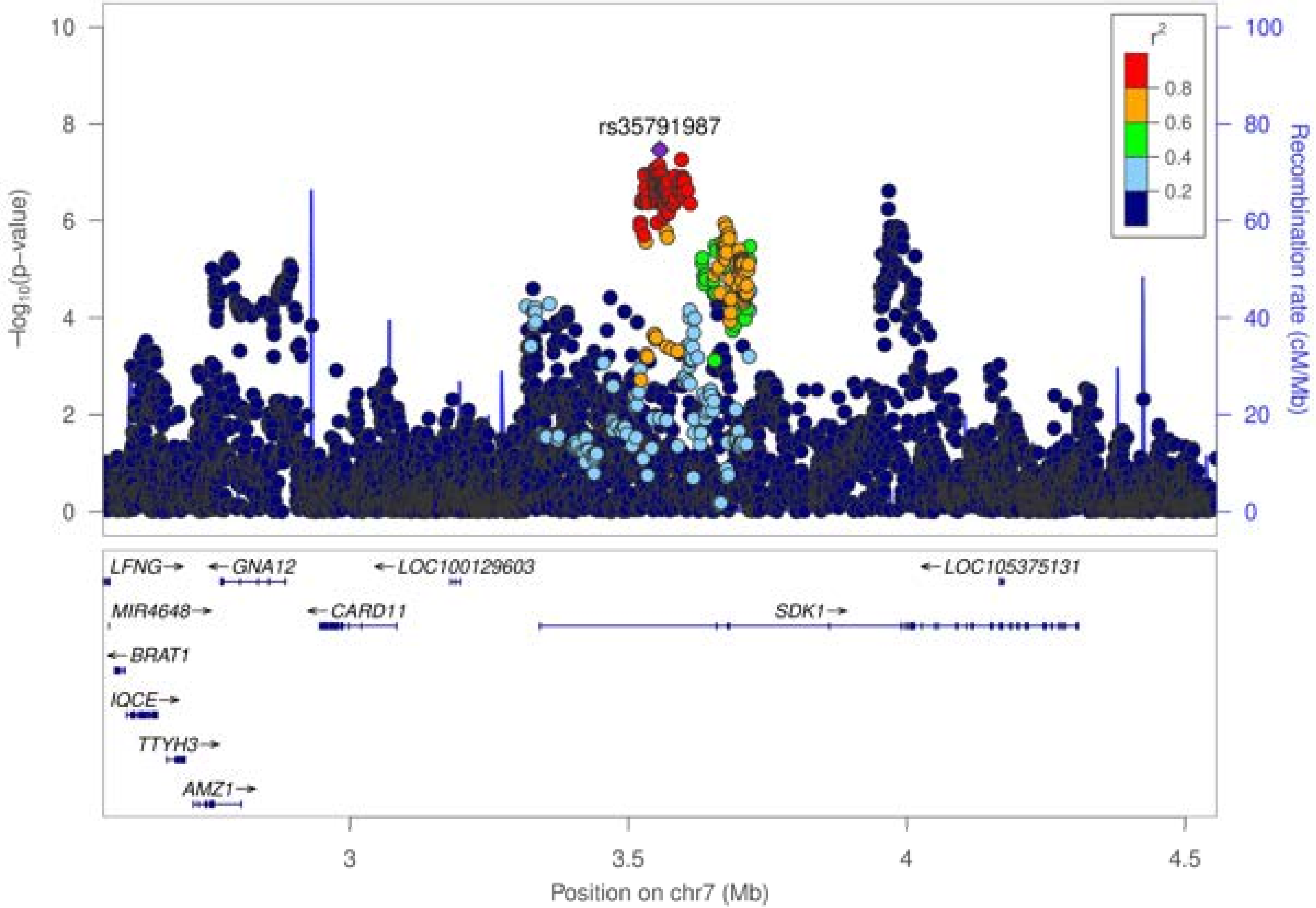

rs35791987
r2
0.8
0.6
0.4
0.2
-log10(p-value)
Recombination rate (cM/Mb)
LFNG
GNA12
LOC100129603
LOC105375131
MIR4648
CARD11
SDK1
BRAT1
IQCE
TTYH3
AMZ1
Position on chr7 (Mb)

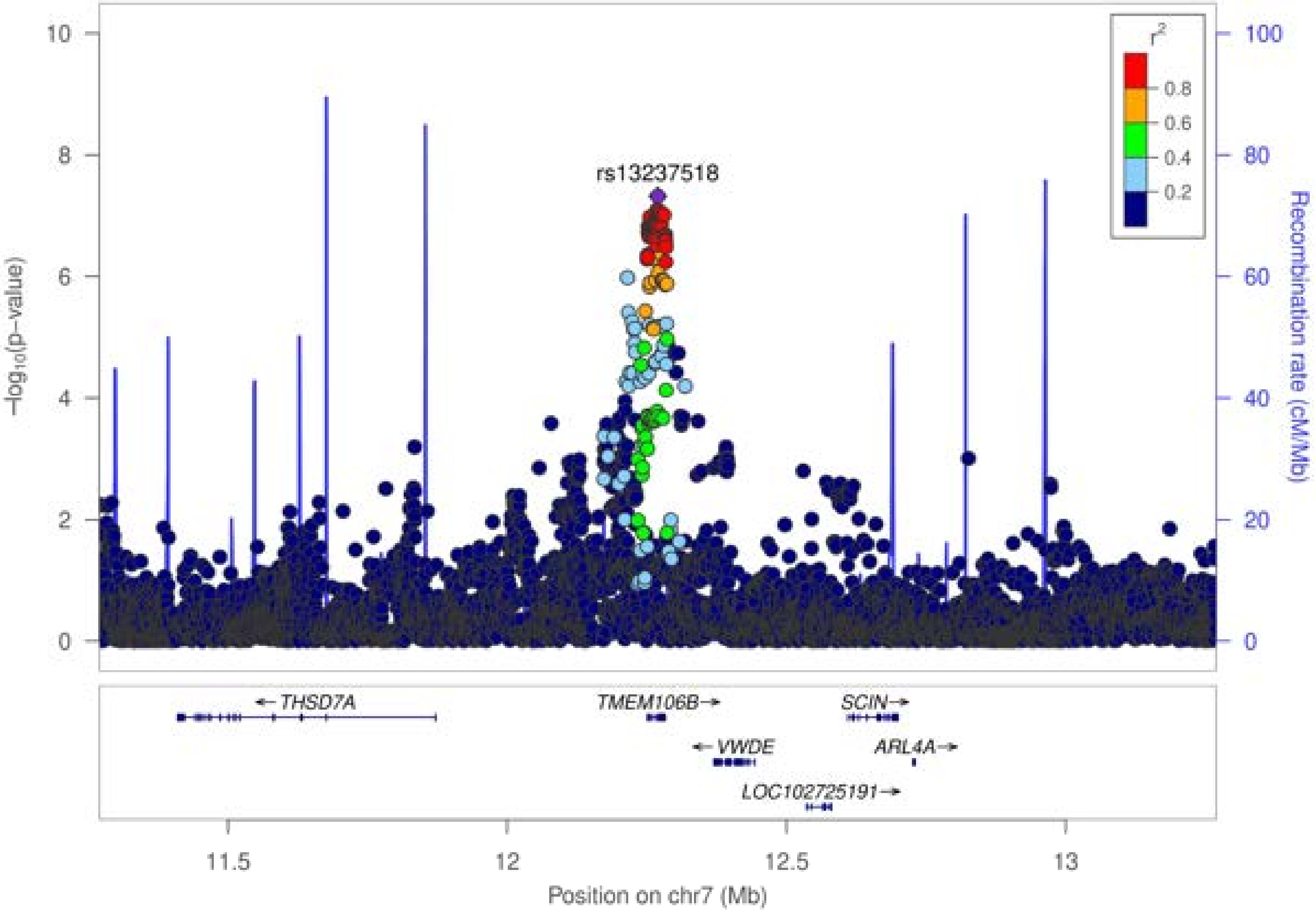

10
8
6
4
2
0
−log10(p−value)
100
80
60
40
20
0
Recombination rate (cM/Mb)
r2
0.8
0.6
0.4
0.2
rs13237518
← THSD7A
TMEM106B→
SCIN→
← VWDE
ARL4A→
LOC102725191→
11.5
12
12.5
13
Position on chr7 (Mb)

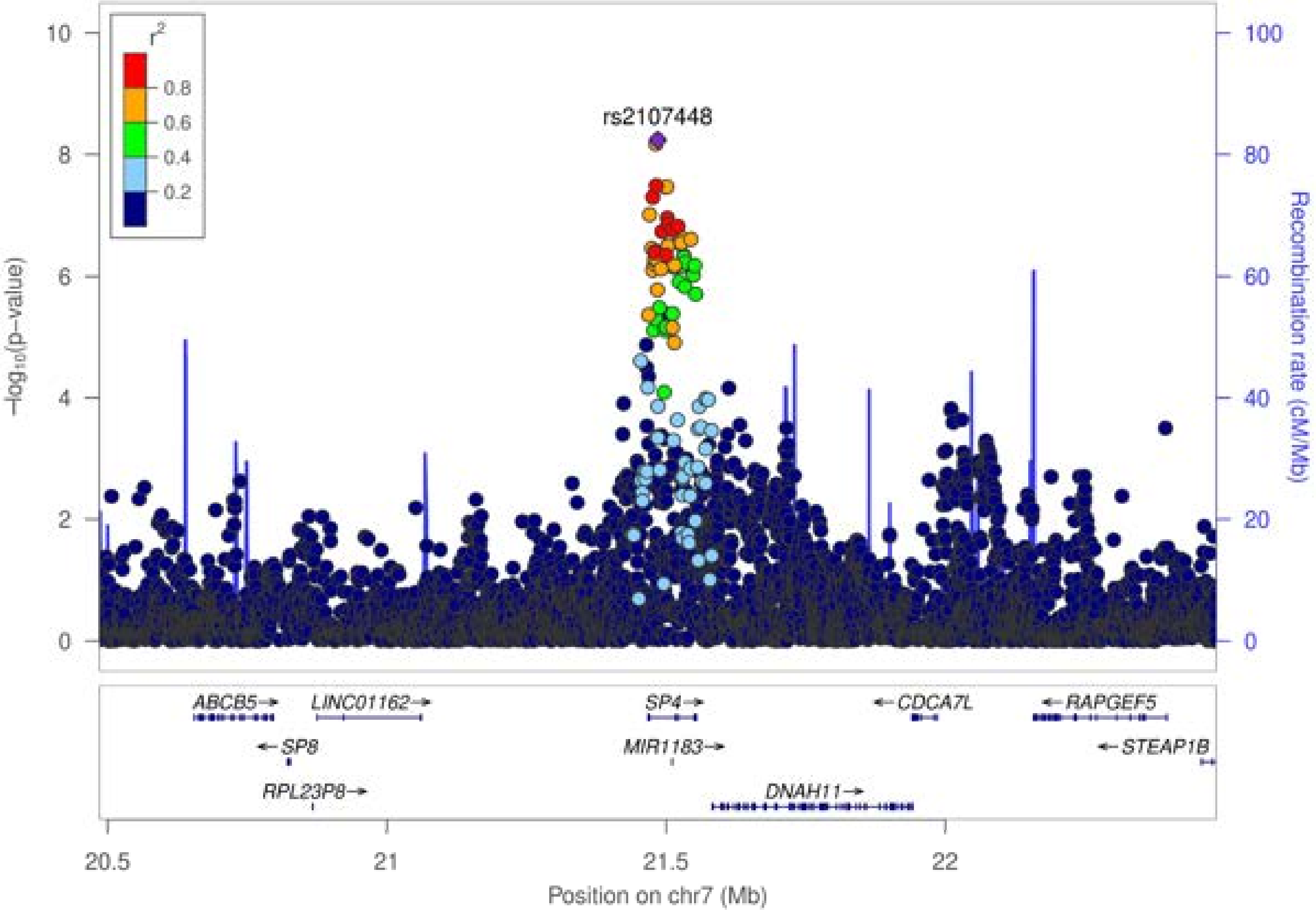

r2
0.8
0.6
0.4
0.2
rs2107448
−log10(p−value)
Recombination rate (cM/Mb)
ABCB5
LINC01162
SP4
CDCA7L
RAPGEF5
SP8
MIR1183
STEAP1B
RPL23P8
DNAH11
20.5
21
21.5
22
Position on chr7 (Mb)

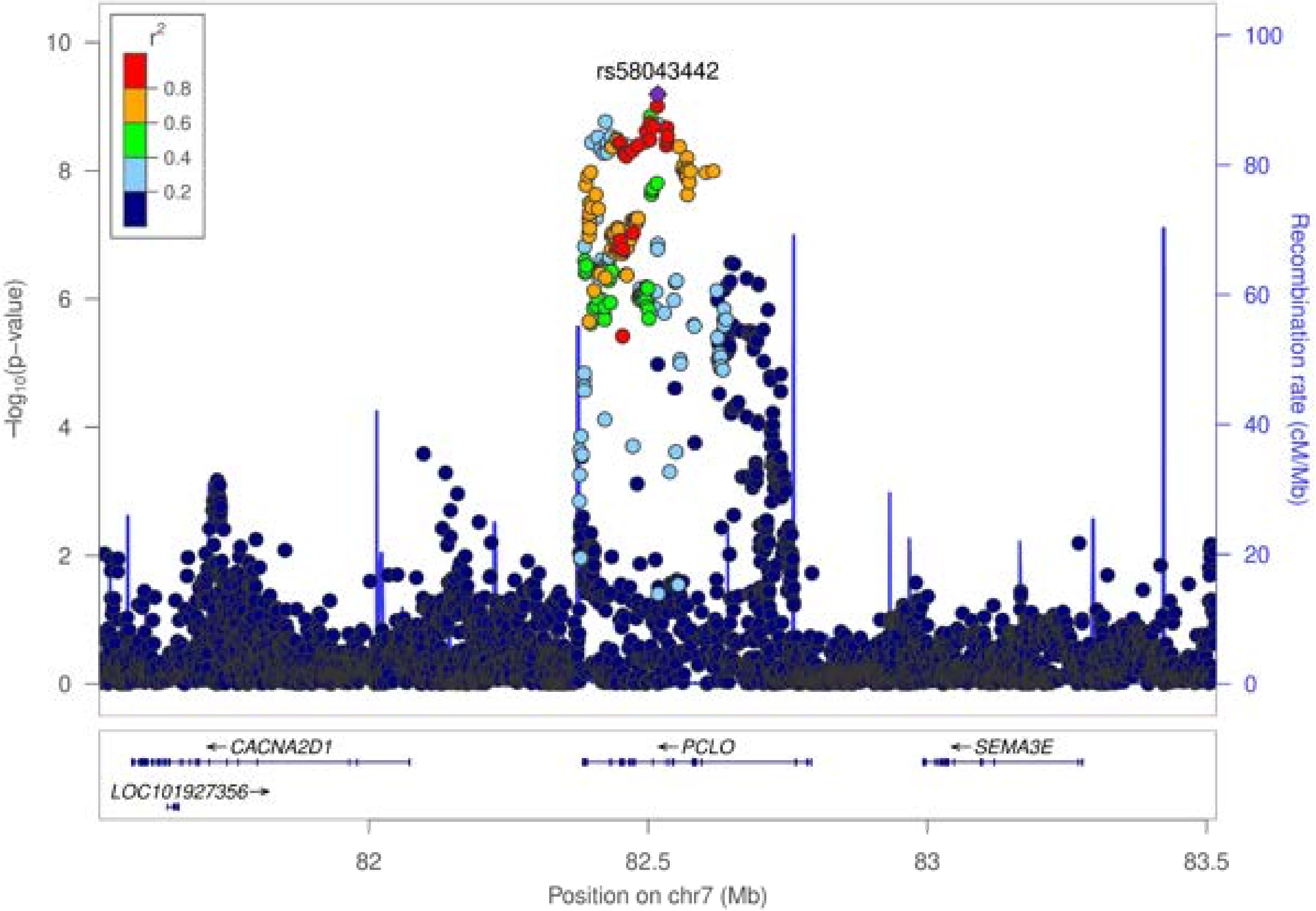

r²
0.8
0.6
0.4
0.2
rs58043442
−log₁₀(p−value)
Recombination rate (cM/Mb)
CACNA2D1
PCLO
SEMA3E
LOC101927356
82
82.5
83
83.5
Position on chr7 (Mb)

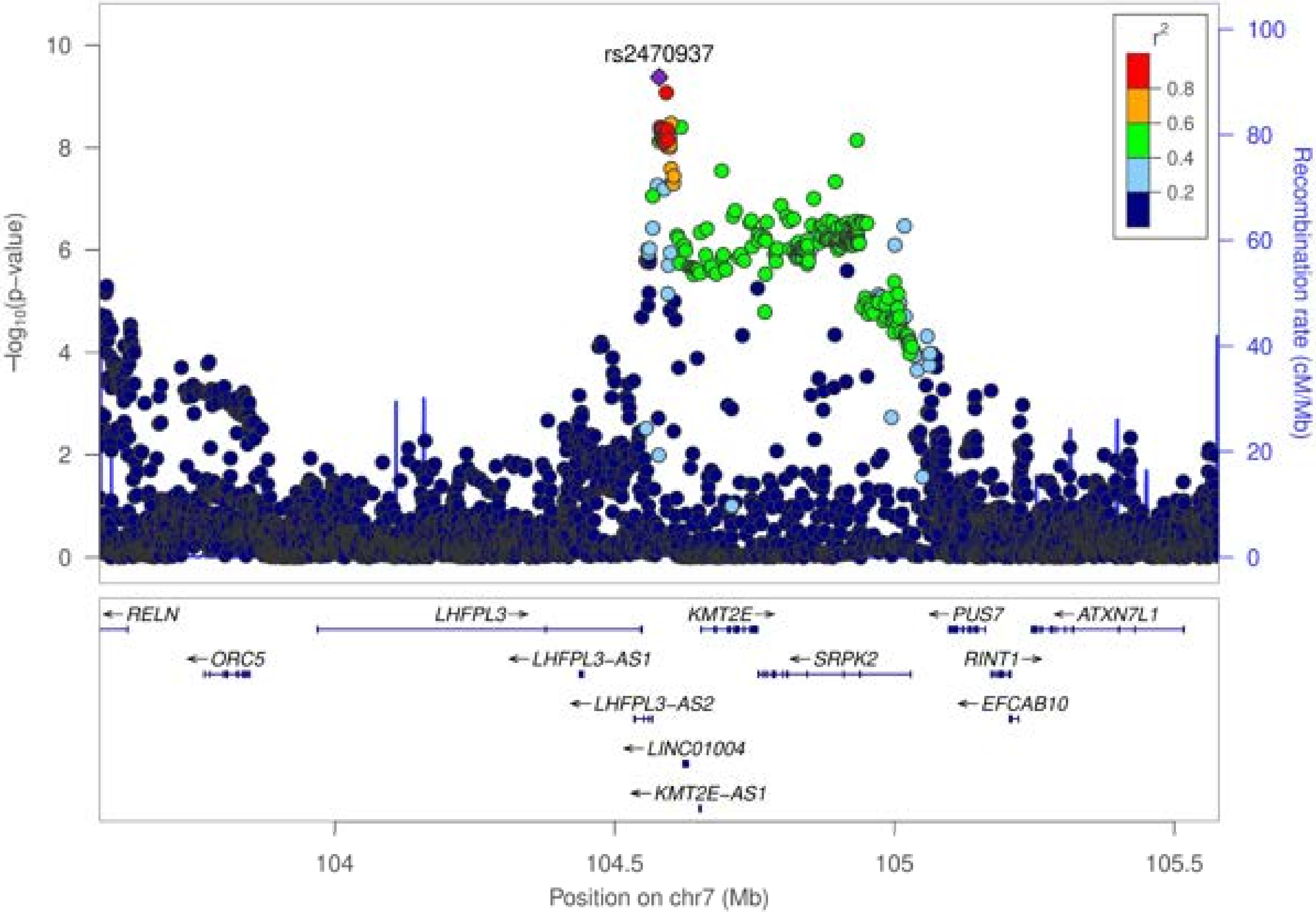
rs2470937
r2
0.8
0.6
0.4
0.2
−log10(p−value)
Recombination rate (cM/Mb)
RELN
ORC5
LHFPL3
LHFPL3-AS1
LHFPL3-AS2
LINC01004
KMT2E-AS1
KMT2E
SRPK2
PUS7
RINT1
EFCAB10
ATXN7L1
104
104.5
105
105.5
Position on chr7 (Mb)

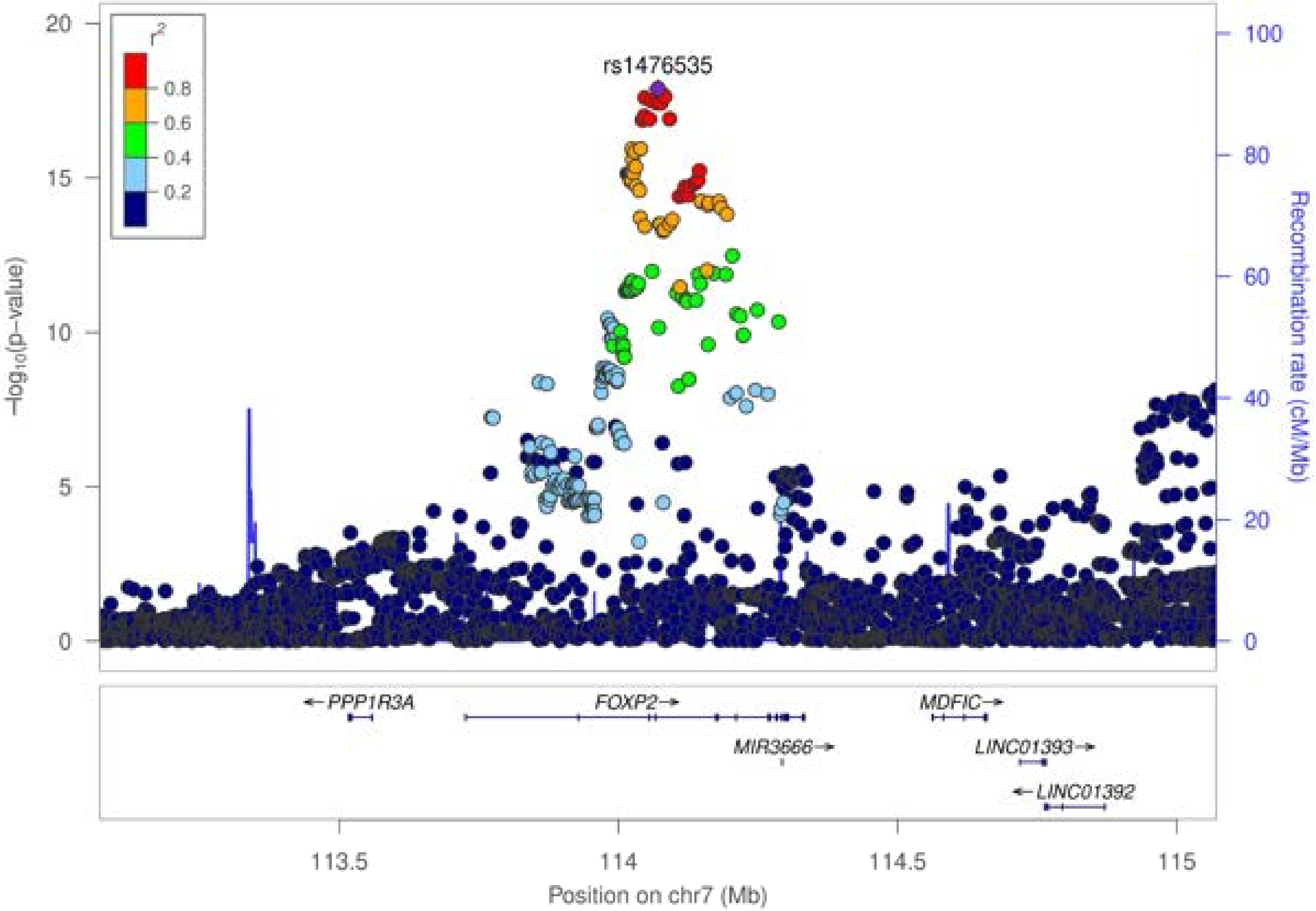

r^2
0.8
0.6
0.4
0.2
rs1476535
−log10(p−value)
Recombination rate (cM/Mb)
20
15
10
5
0
100
80
60
40
20
0
← PPP1R3A
FOXP2→
MDFIC→
MIR3666→
LINC01393→
← LINC01392
113.5
114
114.5
115
Position on chr7 (Mb)

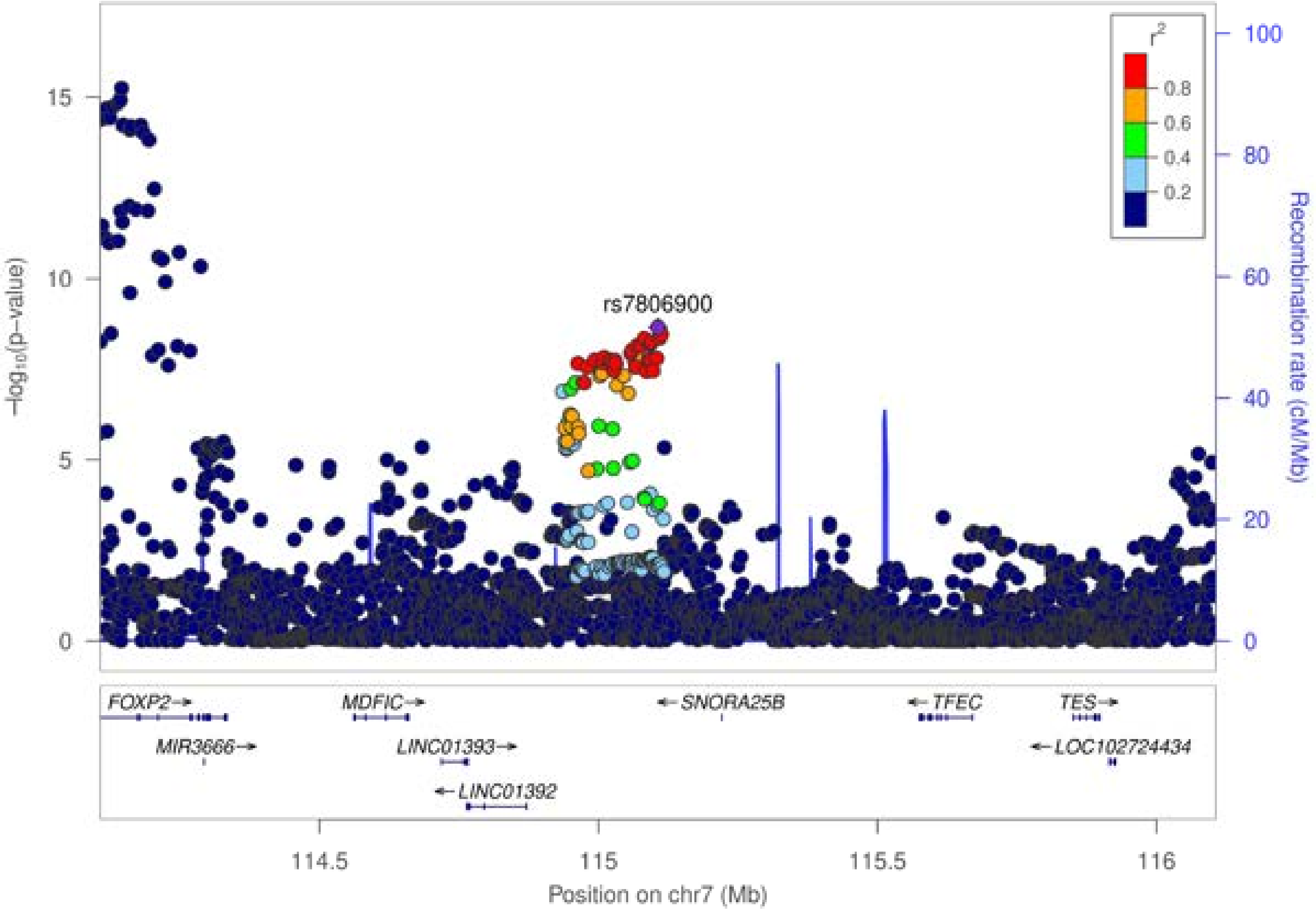

rs7806900
r2
0.8
0.6
0.4
0.2
15
10
5
0
−log10(p−value)
100
80
60
40
20
0
Recombination rate (cM/Mb)
FOXP2
MIR3666
MDFIC
LINC01393
LINC01392
SNORA25B
TFEC
TES
LOC102724434
114.5
115
115.5
116
Position on chr7 (Mb)

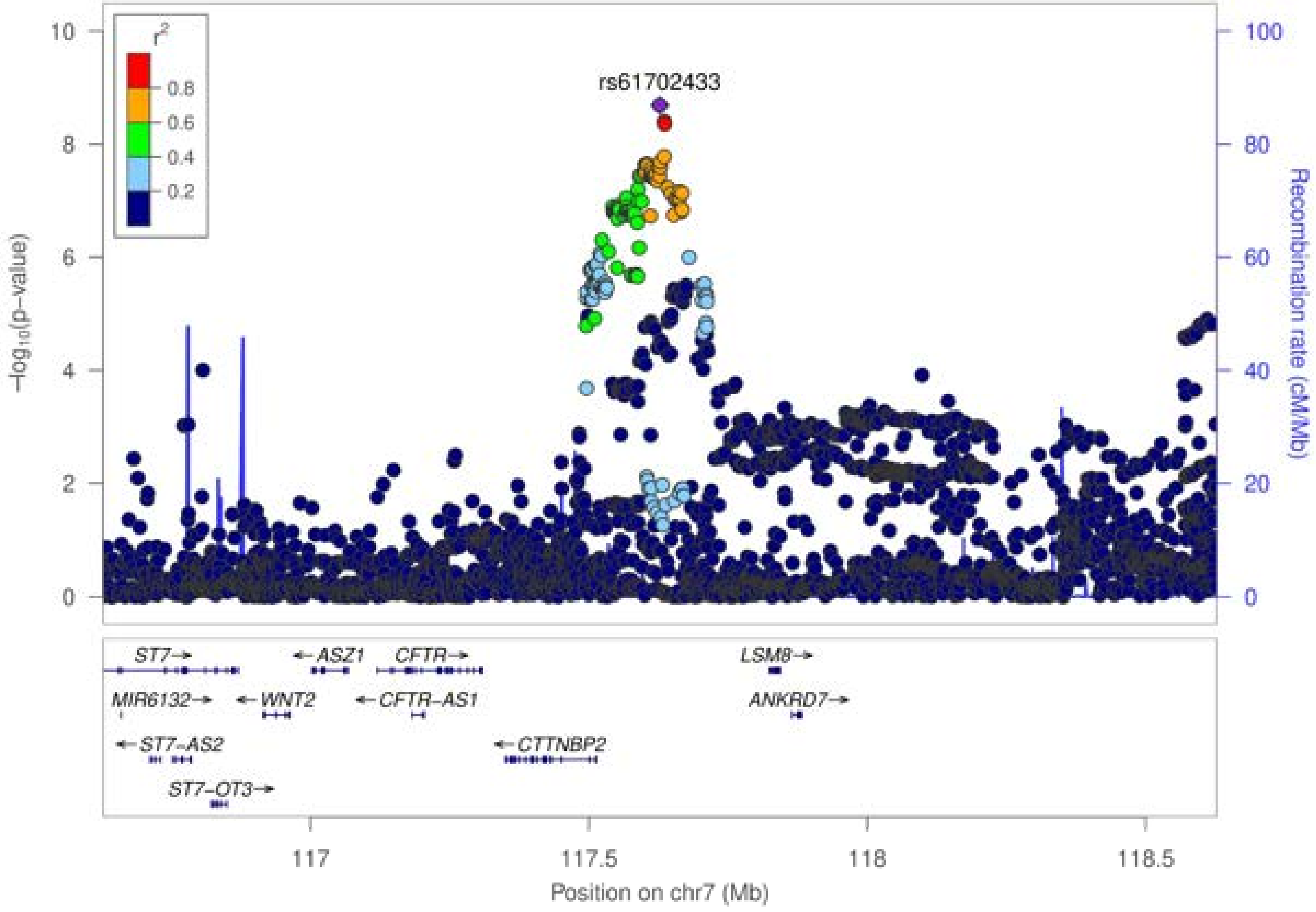

rs61702433
r2
0.8
0.6
0.4
0.2
−log10(p−value)
Recombination rate (cM/Mb)
0
2
4
6
8
10
0
20
40
60
80
100
ST7
ASZ1
CFTR
LSM8
MIR6132
WNT2
CFTR−AS1
ANKRD7
ST7−AS2
CTTNBP2
ST7−OT3
117
117.5
118
118.5
Position on chr7 (Mb)

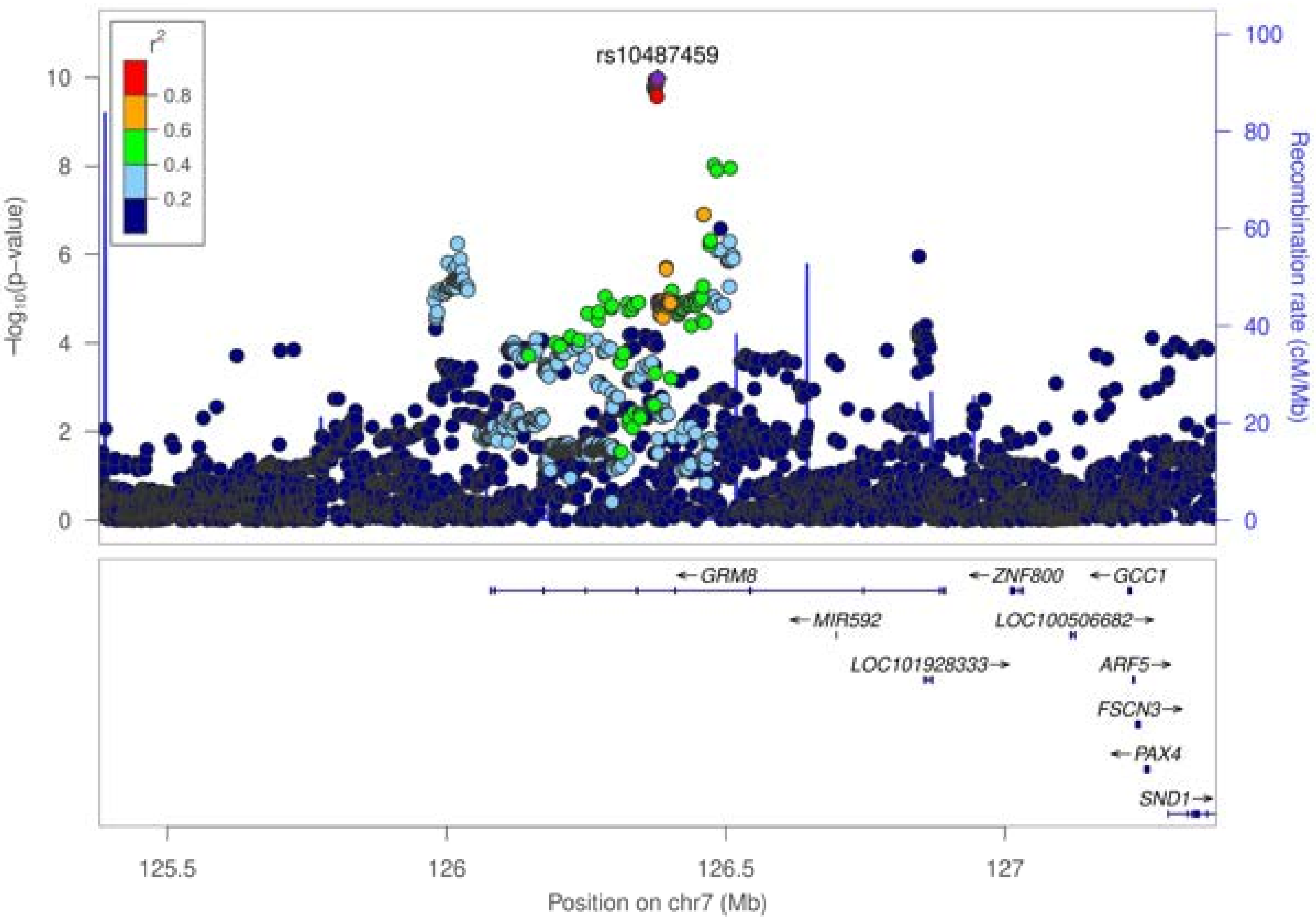

rs10487459
r²
0.8
0.6
0.4
0.2
−log₁₀(p−value)
Recombination rate (cM/Mb)
0
2
4
6
8
10
0
20
40
60
80
100
GRM8
ZNF800
GCC1
MIR592
LOC100506682
LOC101928333
ARF5
FSCN3
PAX4
SND1
125.5
126
126.5
127
Position on chr7 (Mb)

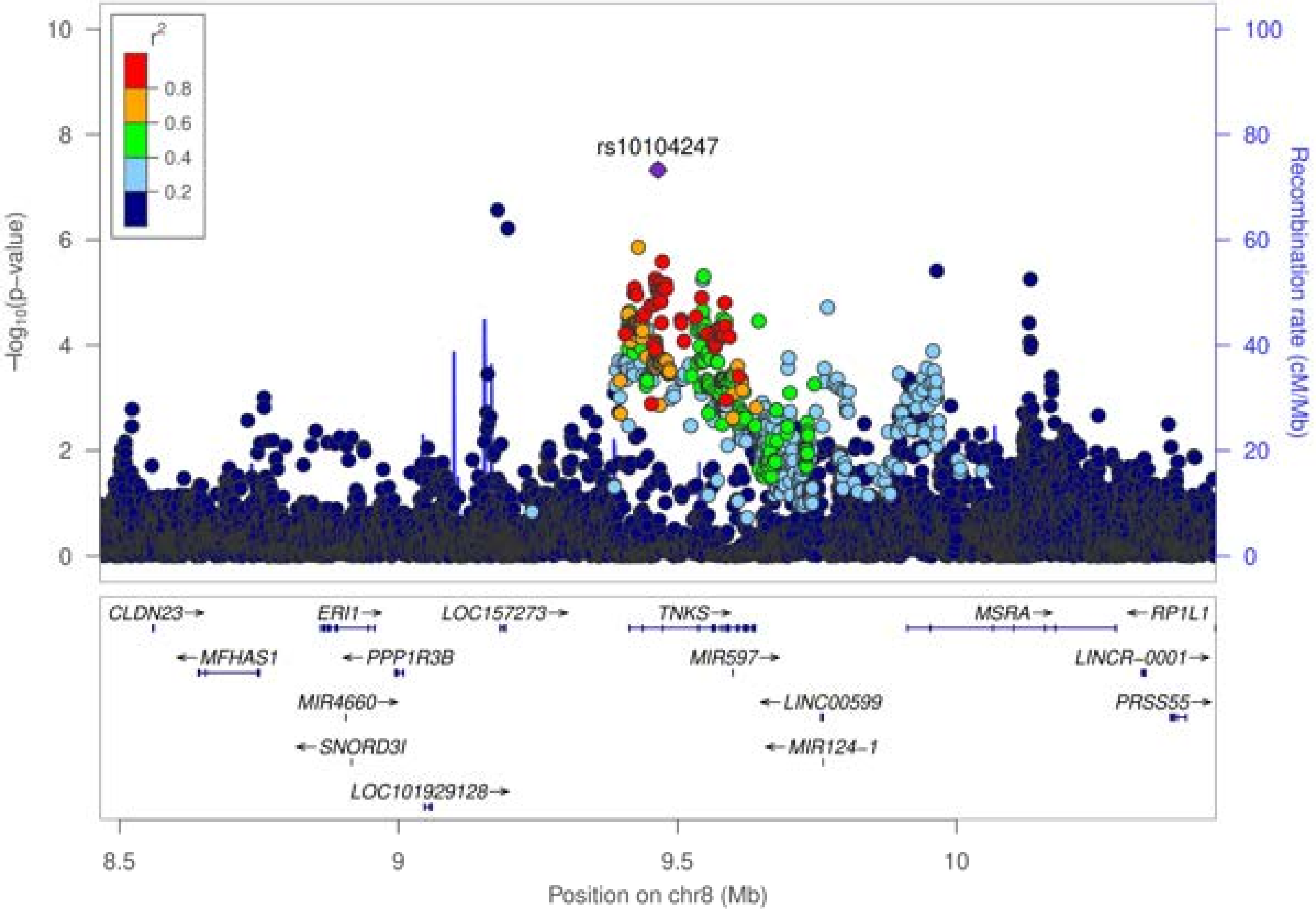

r²
0.8
0.6
0.4
0.2
rs10104247
−log10(p−value)
Recombination rate (cM/Mb)
10
8
6
4
2
0
100
80
60
40
20
0
CLDN23
MFHAS1
ERI1
PPP1R3B
MIR4660
SNORD3I
LOC101929128
LOC157273
TNKS
MIR597
LINC00599
MIR124−1
MSRA
RP1L1
LINCR−0001
PRSS55
8.5
9
9.5
10
Position on chr8 (Mb)

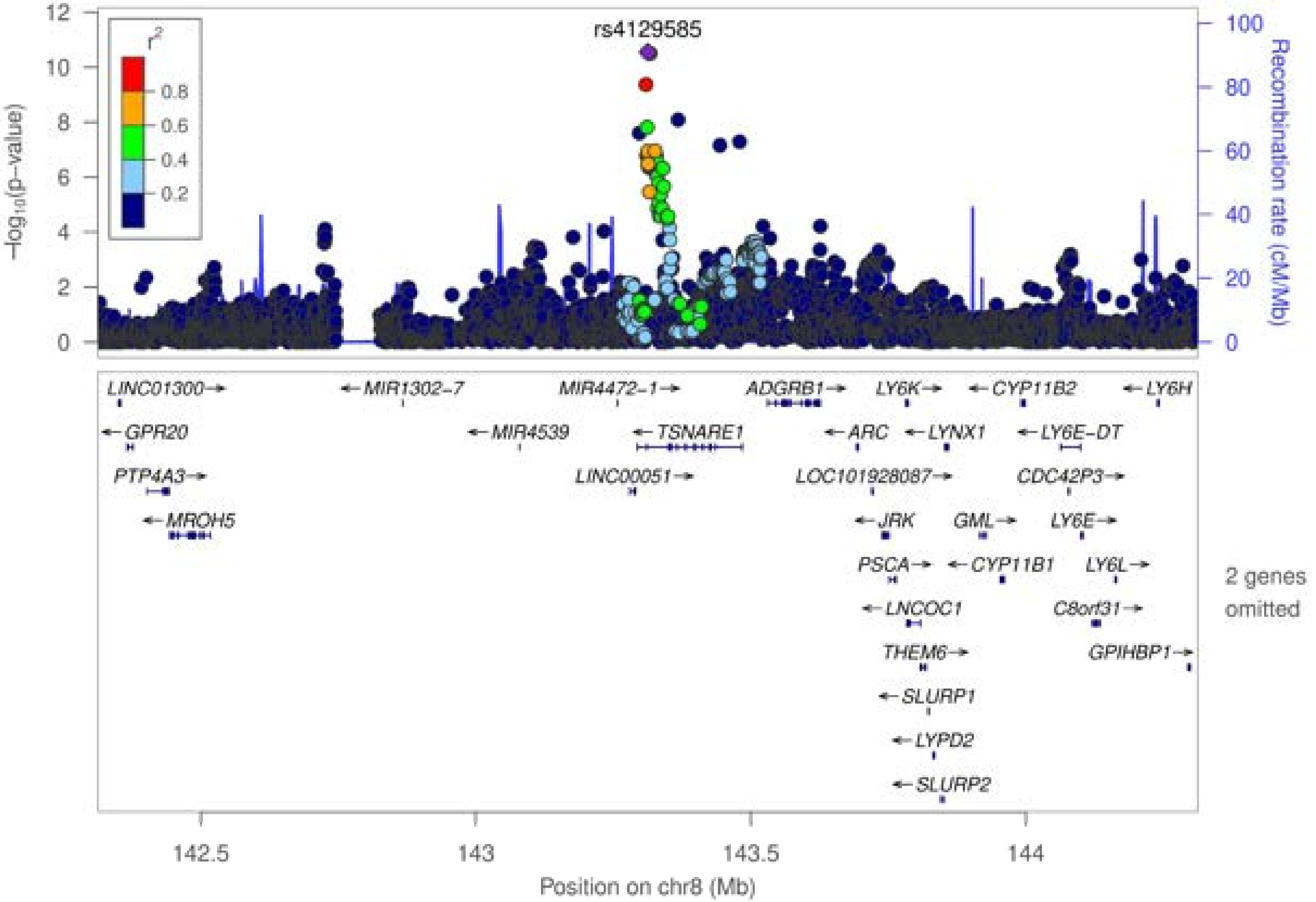
rs4129585
r²
0.8
0.6
0.4
0.2
−log10(p−value)
Recombination rate (cM/Mb)
LINC01300→
←MIR1302-7
MIR4472-1→
ADGRB1→
LY6K→
←CYP11B2
←LY6H
←GPR20
←MIR4539
←TSNARE1
←ARC
←LYNX1
←LY6E-DT
PTP4A3→
LINC00051→
LOC101928087→
CDC42P3→
←MROH5
←JRK
GML→
LY6E→
PSCA→
←CYP11B1
LY6L→
←LNCOC1
C8orf31→
THEM6→
GPIHBP1→
←SLURP1
←LYPD2
←SLURP2
2 genes omitted
142.5
143
143.5
144
Position on chr8 (Mb)

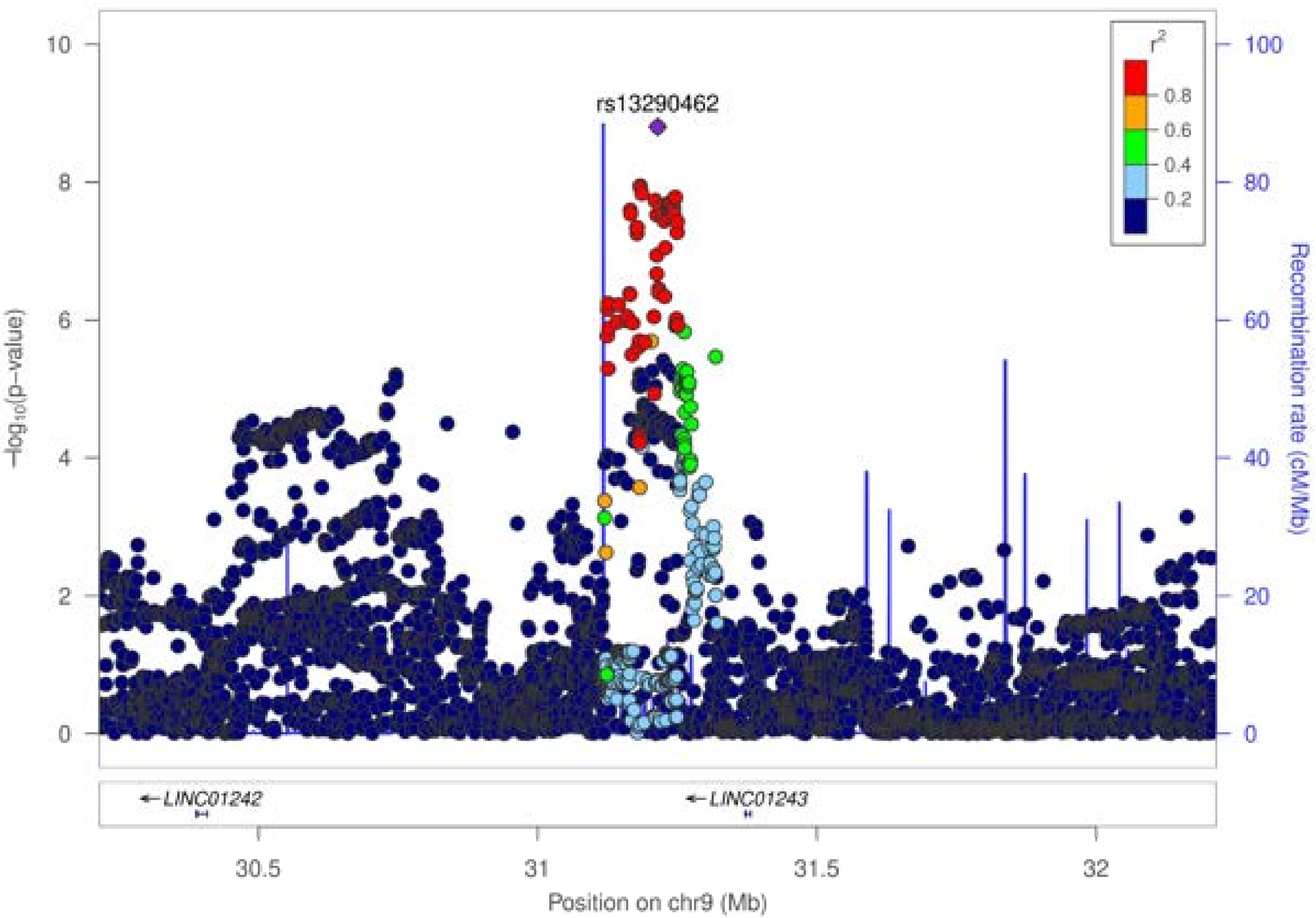

rs13290462
r²
0.8
0.6
0.4
0.2
−log10(p−value)
Recombination rate (cM/Mb)
LINC01242
LINC01243
Position on chr9 (Mb)
30.5
31
31.5
32
0
2
4
6
8
10
20
40
60
80
100

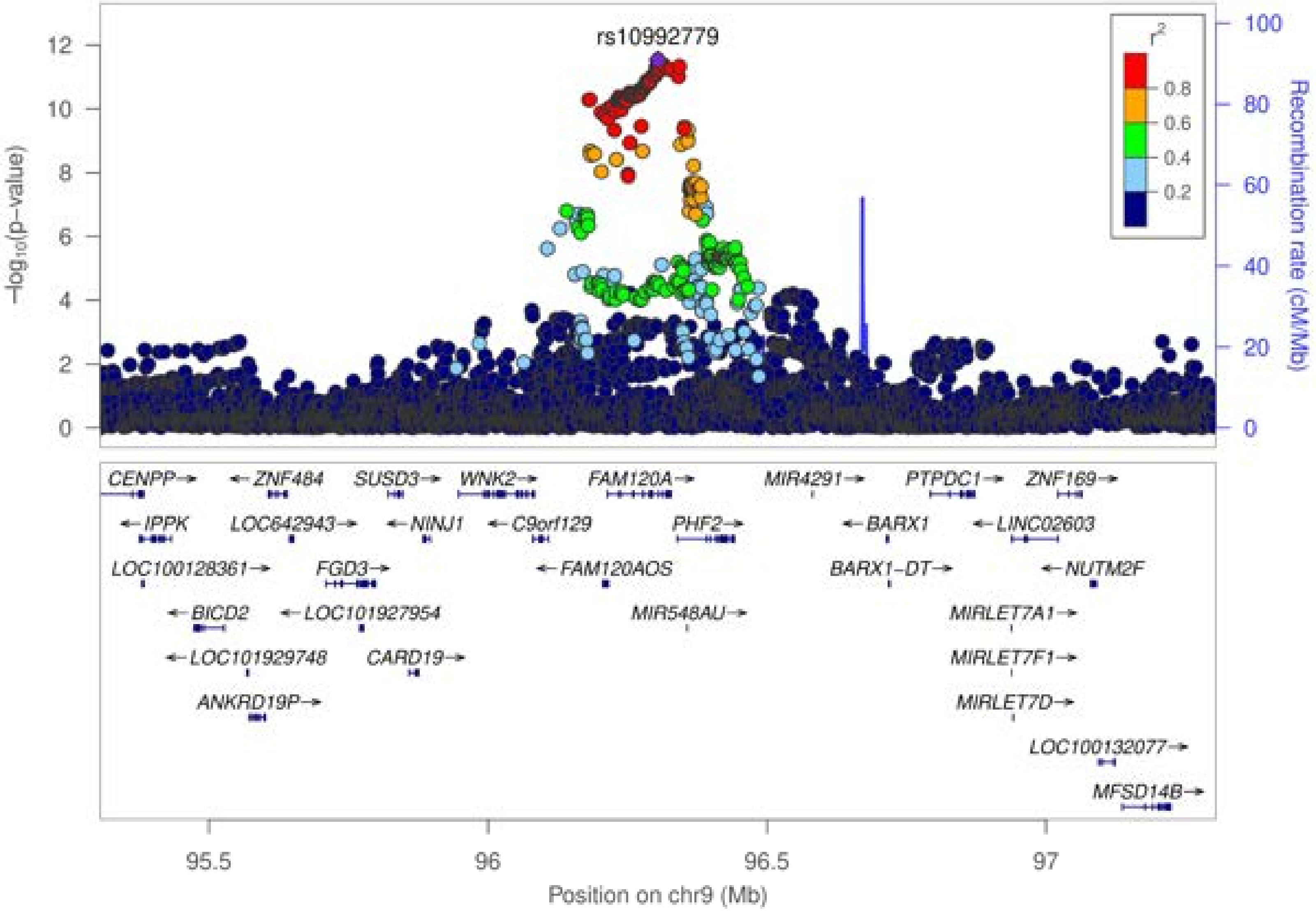

rs10992779
r²
0.8
0.6
0.4
0.2
−log10(p−value)
Recombination rate (cM/Mb)
CENPP→
←ZNF484
SUSD3→
WNK2→
FAM120A→
MIR4291→
PTPDC1→
ZNF169→
←IPPK
LOC642943→
←NINJ1
←C9orf129
PHF2→
←BARX1
←LINC02603
LOC100128361→
FGD3→
←FAM120AOS
BARX1−DT→
←NUTM2F
←BICD2
←LOC101927954
MIR548AU→
MIRLET7A1→
←LOC101929748
CARD19→
MIRLET7F1→
ANKRD19P→
MIRLET7D→
LOC100132077→
MFSD14B→
95.5
96
96.5
97
Position on chr9 (Mb)

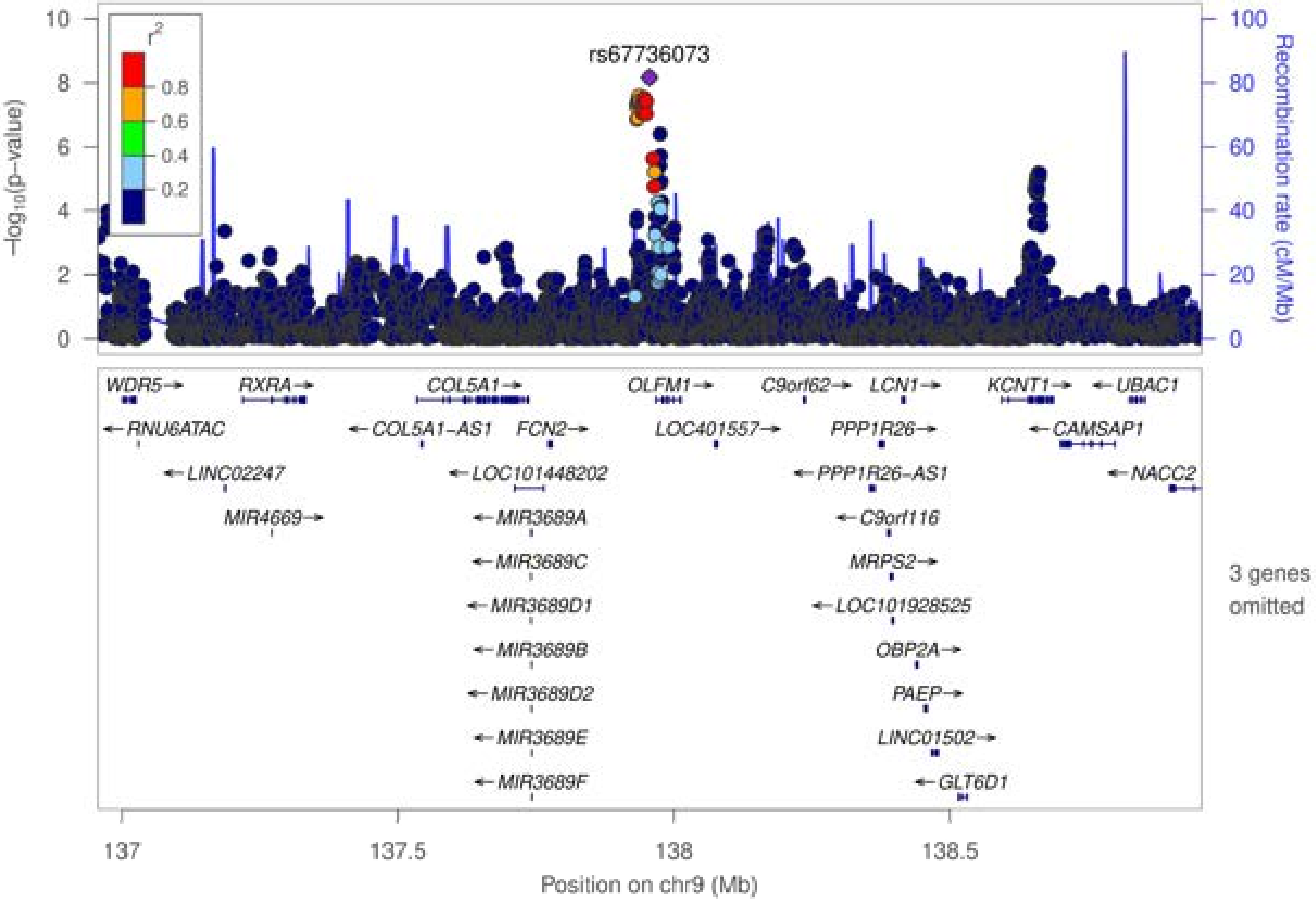

rs67736073
r²
0.8
0.6
0.4
0.2
−log10(p−value)
Recombination rate (cM/Mb)
WDR5
RXRA
COL5A1
OLFM1
C9orf62
LCN1
KCNT1
UBAC1
RNU6ATAC
COL5A1-AS1
FCN2
LOC401557
PPP1R26
CAMSAP1
LINC02247
LOC101448202
PPP1R26-AS1
NACC2
MIR4669
MIR3689A
C9orf116
MIR3689C
MRPS2
MIR3689D1
LOC101928525
MIR3689B
OBP2A
MIR3689D2
PAEP
MIR3689E
LINC01502
MIR3689F
GLT6D1
3 genes omitted
137
137.5
138
138.5
Position on chr9 (Mb)

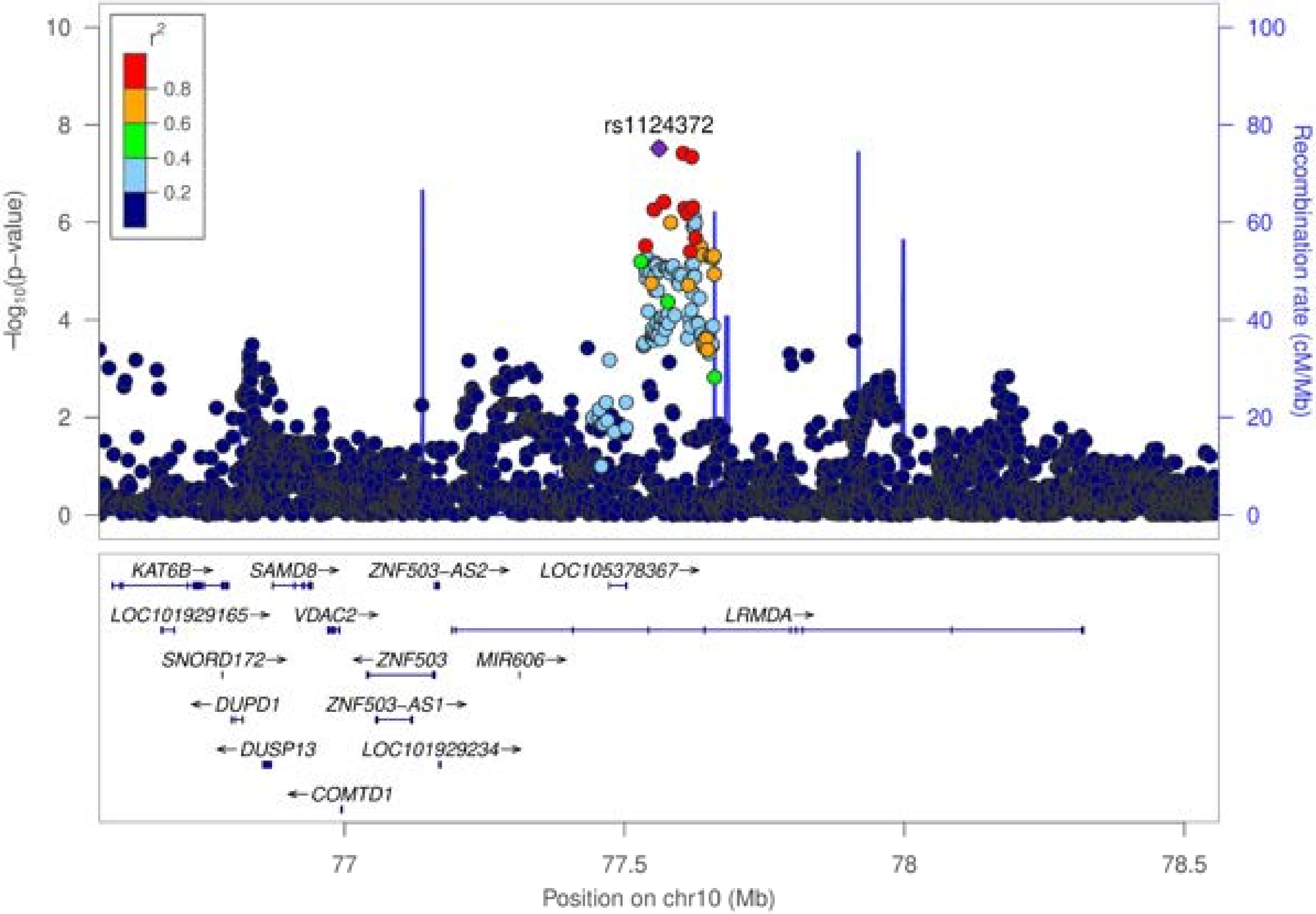

rs1124372
r^2
0.8
0.6
0.4
0.2
10
8
6
4
2
0
-log10(p-value)
100
80
60
40
20
0
Recombination rate (cM/Mb)
KAT6B→
SAMD8→
ZNF503-AS2→
LOC105378367→
LOC101929165→
VDAC2→
LRMDA→
SNORD172→
←ZNF503
MIR606→
←DUPD1
ZNF503-AS1→
←DUSP13
LOC101929234→
←COMTD1
77
77.5
78
78.5
Position on chr10 (Mb)

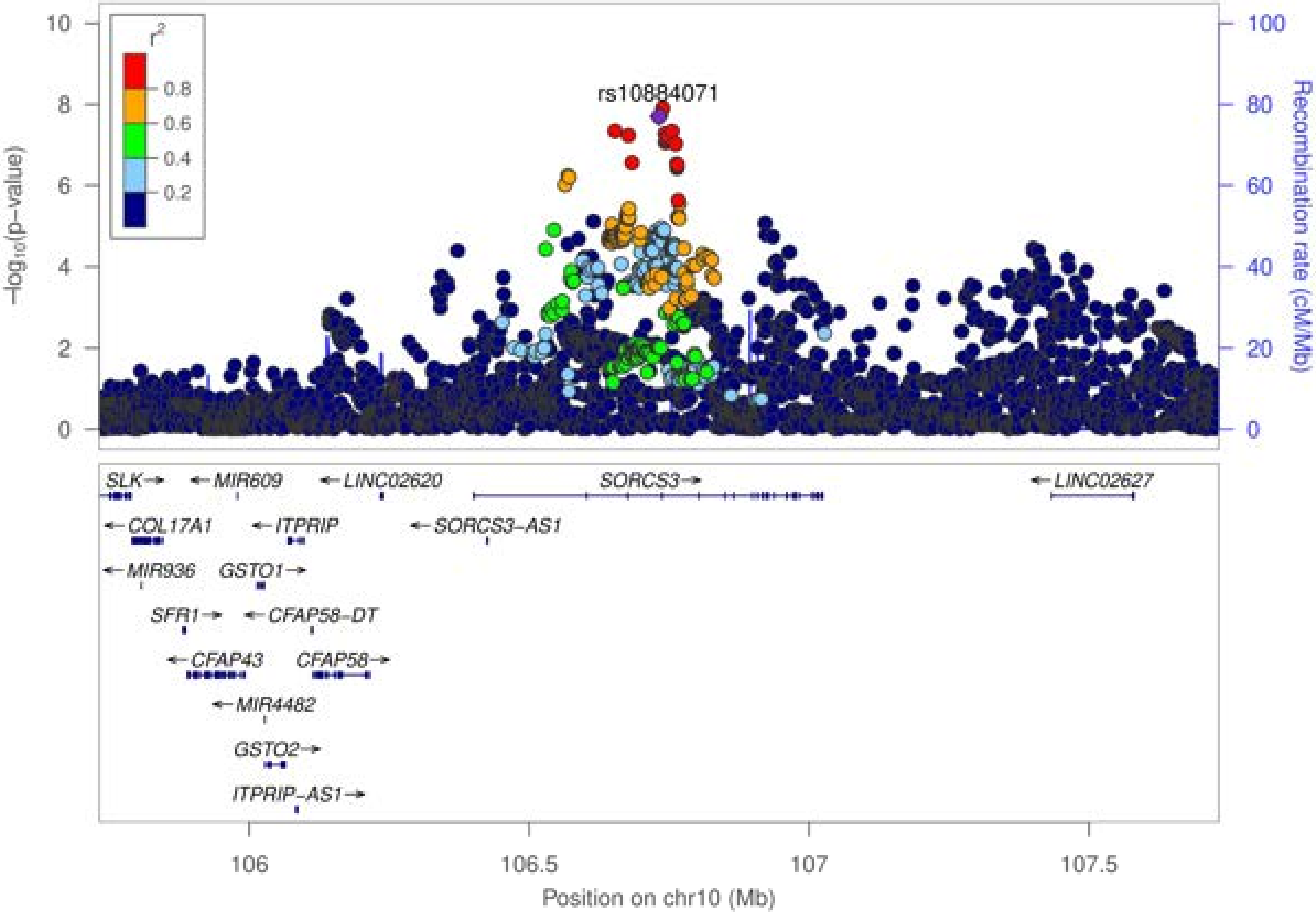

rs10884071
r2
0.8
0.6
0.4
0.2
−log10(p−value)
Recombination rate (cM/Mb)
SLK
MIR609
LINC02620
SORCS3
LINC02627
COL17A1
ITPRIP
SORCS3−AS1
MIR936
GSTO1
SFR1
CFAP58−DT
CFAP43
CFAP58
MIR4482
GSTO2
ITPRIP−AS1
106
106.5
107
107.5
Position on chr10 (Mb)

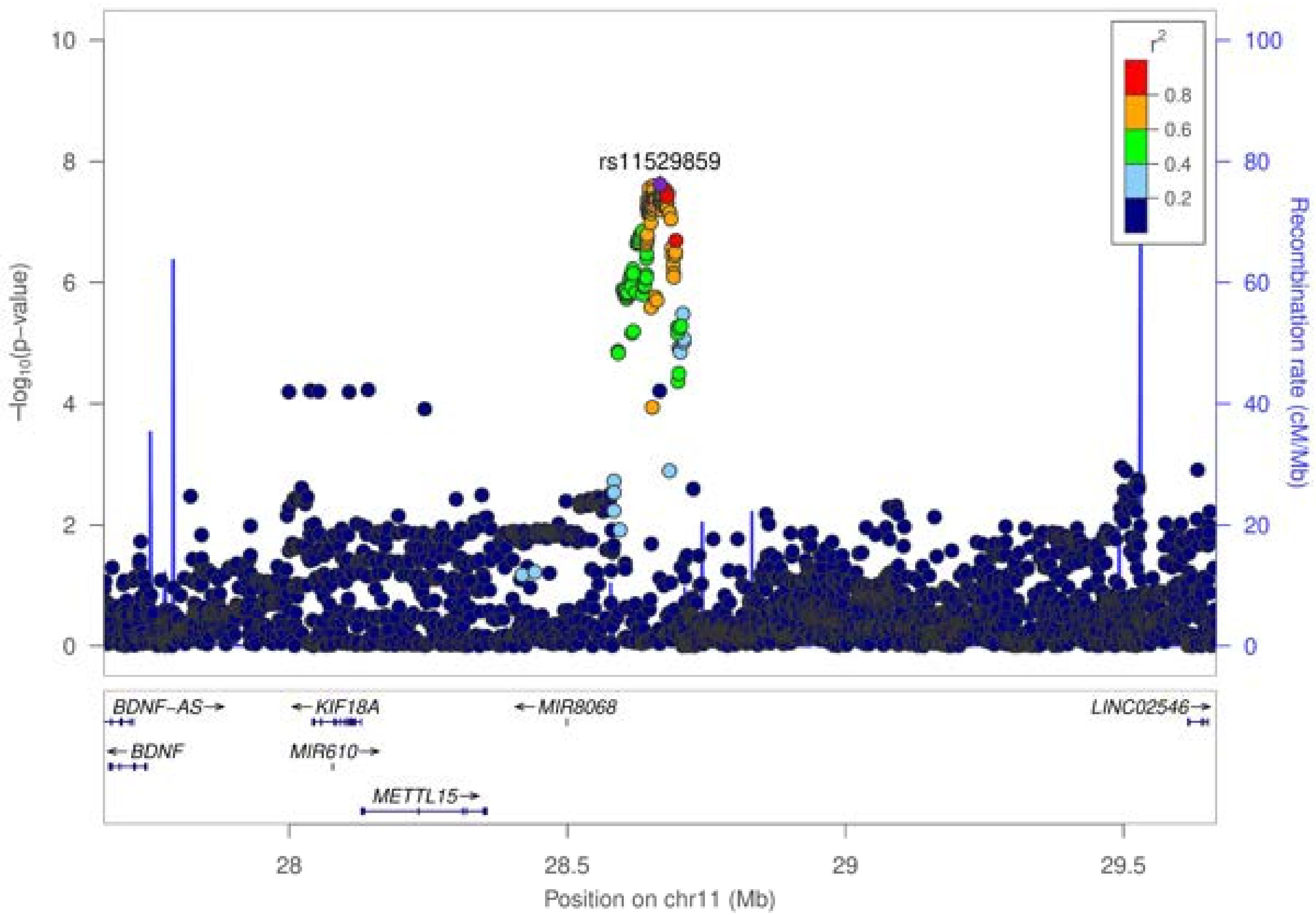

rs11529859
r²
0.8
0.6
0.4
0.2
−log₁₀(p−value)
10
8
6
4
2
0
100
80
60
40
20
0
Recombination rate (cM/Mb)
BDNF-AS→
←BDNF
←KIF18A
MIR610→
METTL15→
←MIR8068
LINC02546→
28
28.5
29
29.5
Position on chr11 (Mb)

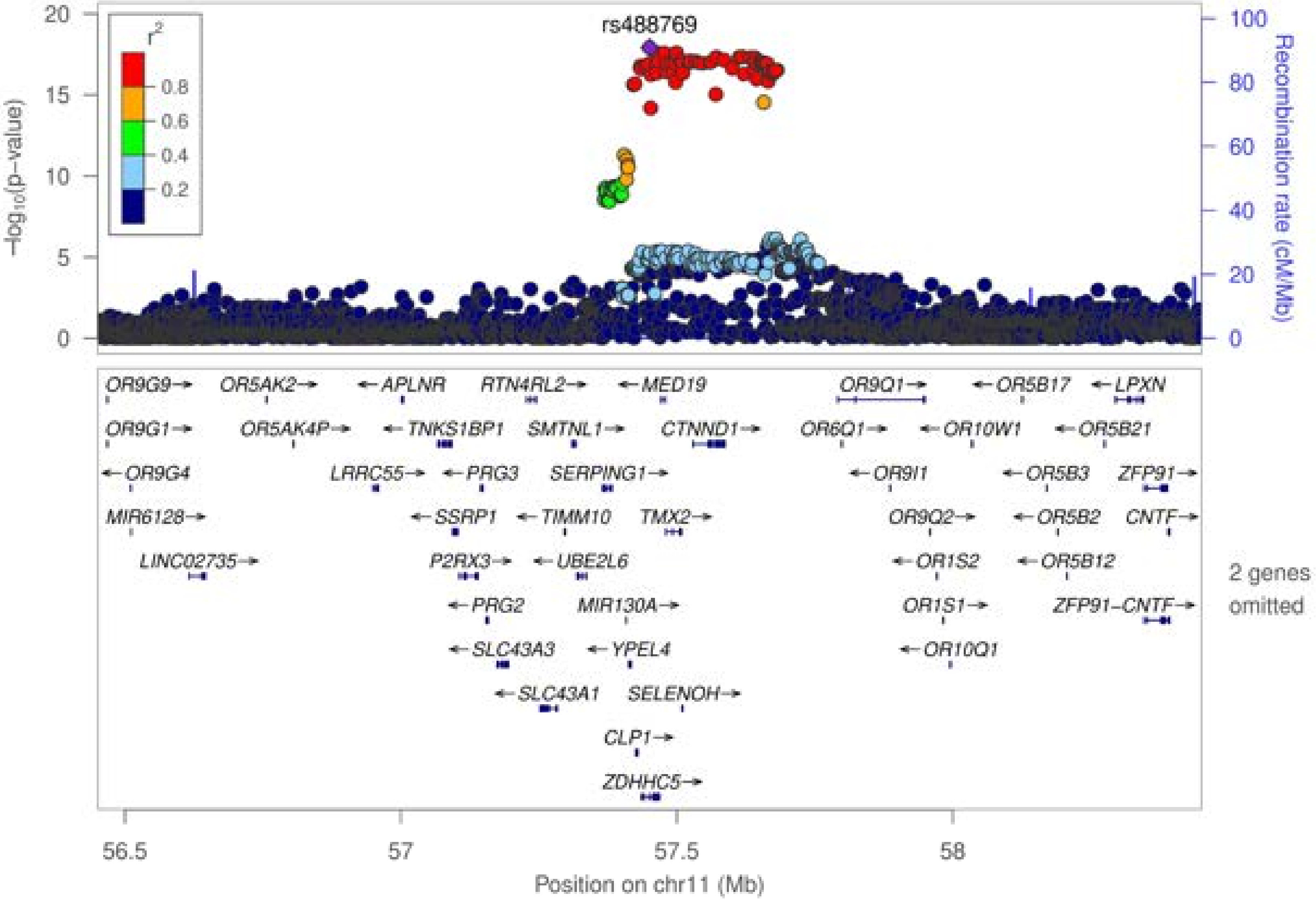

rs488769
r²
0.8
0.6
0.4
0.2
−log10(p−value)
Recombination rate (cM/Mb)
OR9G9 OR5AK2 APLNR RTN4RL2 MED19 OR9Q1 OR5B17 LPXN
OR9G1 OR5AK4P TNKS1BP1 SMTNL1 CTNND1 OR6Q1 OR10W1 OR5B21
OR9G4 LRRC55 PRG3 SERPING1 OR9I1 OR5B3 ZFP91
MIR6128 SSRP1 TIMM10 TMX2 OR9Q2 OR5B2 CNTF
LINC02735 P2RX3 UBE2L6 OR1S2 OR5B12
PRG2 MIR130A OR1S1 ZFP91−CNTF
SLC43A3 YPEL4 OR10Q1
SLC43A1 SELENOH
CLP1
ZDHHC5
2 genes omitted
56.5
57
57.5
58
Position on chr11 (Mb)

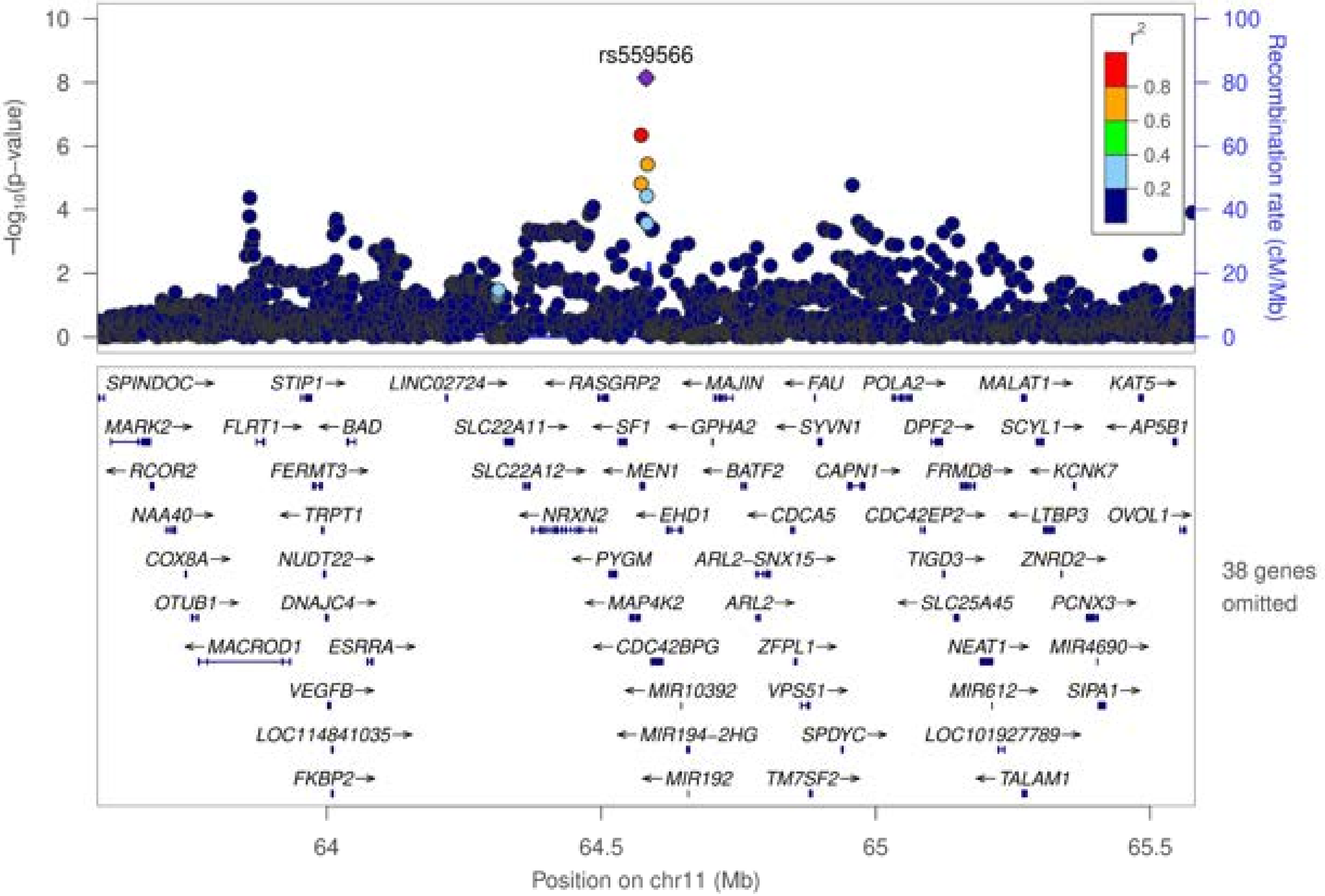
rs559566
$-\log_{10}$(p−value)
Recombination rate (cM/Mb)
$r^2$
0.8
0.6
0.4
0.2
0
2
4
6
8
10
20
40
60
80
100
SPINDOC→ STIP1→ LINC02724→ ←RASGRP2 ←MAJIN ←FAU POLA2→ MALAT1→ KAT5→
MARK2→ FLRT1→ ←BAD SLC22A11→ ←SF1 ←GPHA2 ←SYVN1 DPF2→ SCYL1→ ←AP5B1
←RCOR2 FERMT3→ SLC22A12→ ←MEN1 ←BATF2 CAPN1→ FRMD8→ ←KCNK7
NAA40→ ←TRPT1 ←NRXN2 ←EHD1 ←CDCA5 CDC42EP2→ ←LTBP3 OVOL1→
COX8A→ NUDT22→ ←PYGM ARL2−SNX15→ TIGD3→ ZNRD2→
OTUB1→ DNAJC4→ ←MAP4K2 ARL2→ ←SLC25A45 PCNX3→
←MACROD1 ESRRA→ ←CDC42BPG ZFPL1→ NEAT1→ MIR4690→
VEGFB→ ←MIR10392 VPS51→ MIR612→ SIPA1→
LOC114841035→ ←MIR194−2HG SPDYC→ LOC101927789→
FKBP2→ ←MIR192 TM7SF2→ ←TALAM1
38 genes omitted
64
64.5
65
65.5
Position on chr11 (Mb)

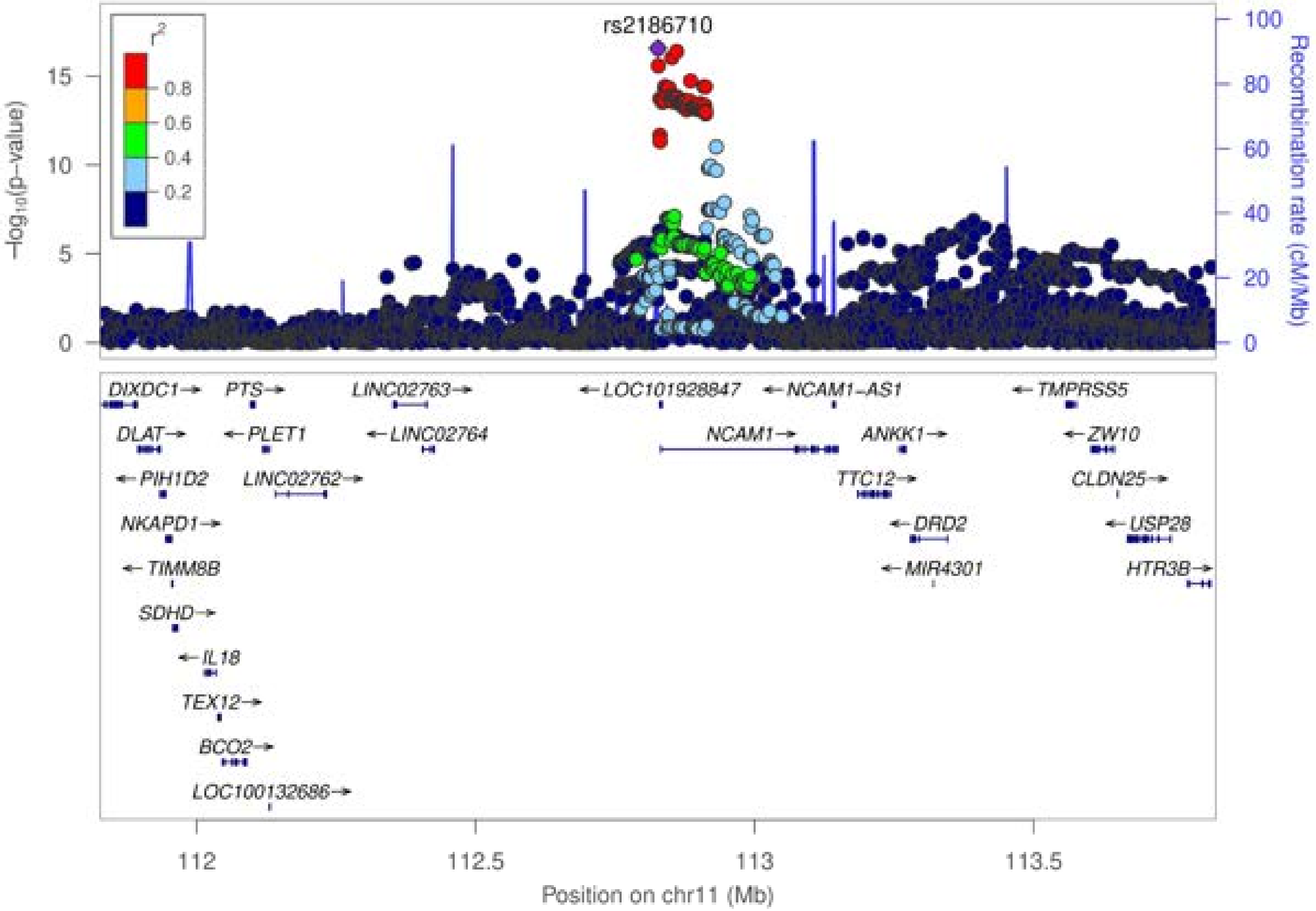

rs2186710
$r^2$
0.8
0.6
0.4
0.2
−log10(p−value)
Recombination rate (cM/Mb)
DIXDC1→
PTS→
LINC02763→
← LOC101928847
← NCAM1−AS1
← TMPRSS5
DLAT→
← PLET1
← LINC02764
NCAM1→
ANKK1→
← ZW10
← PIH1D2
LINC02762→
TTC12→
CLDN25→
NKAPD1→
← DRD2
← USP28
← TIMM8B
← MIR4301
HTR3B→
SDHD→
← IL18
TEX12→
BCO2→
LOC100132686→
Position on chr11 (Mb)

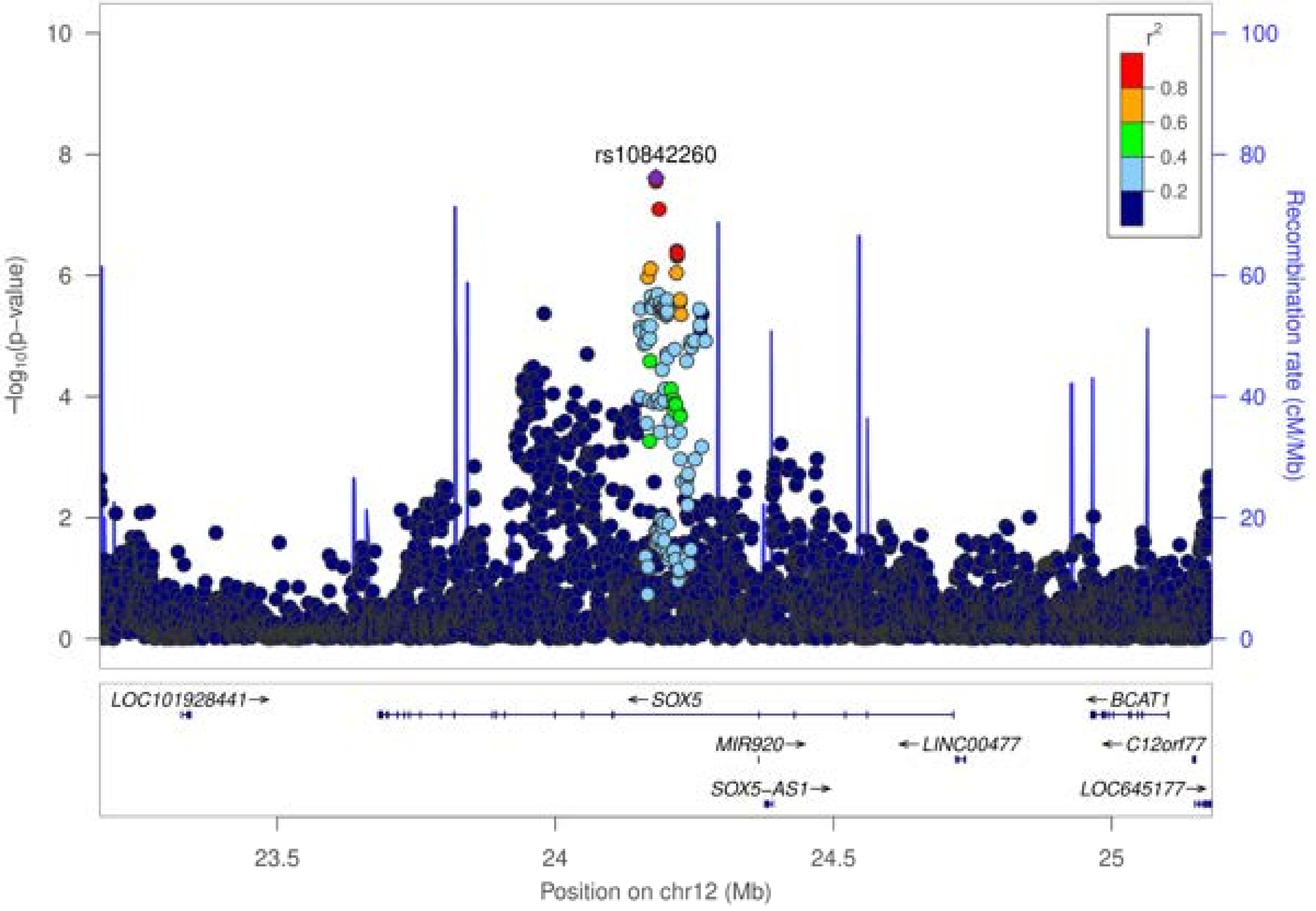

rs10842260
r2
0.8
0.6
0.4
0.2
−log10(p−value)
Recombination rate (cM/Mb)
10
8
6
4
2
0
100
80
60
40
20
0
LOC101928441→
← SOX5
← BCAT1
MIR920→
← LINC00477
← C12orf77
SOX5−AS1→
LOC645177→
23.5
24
24.5
25
Position on chr12 (Mb)

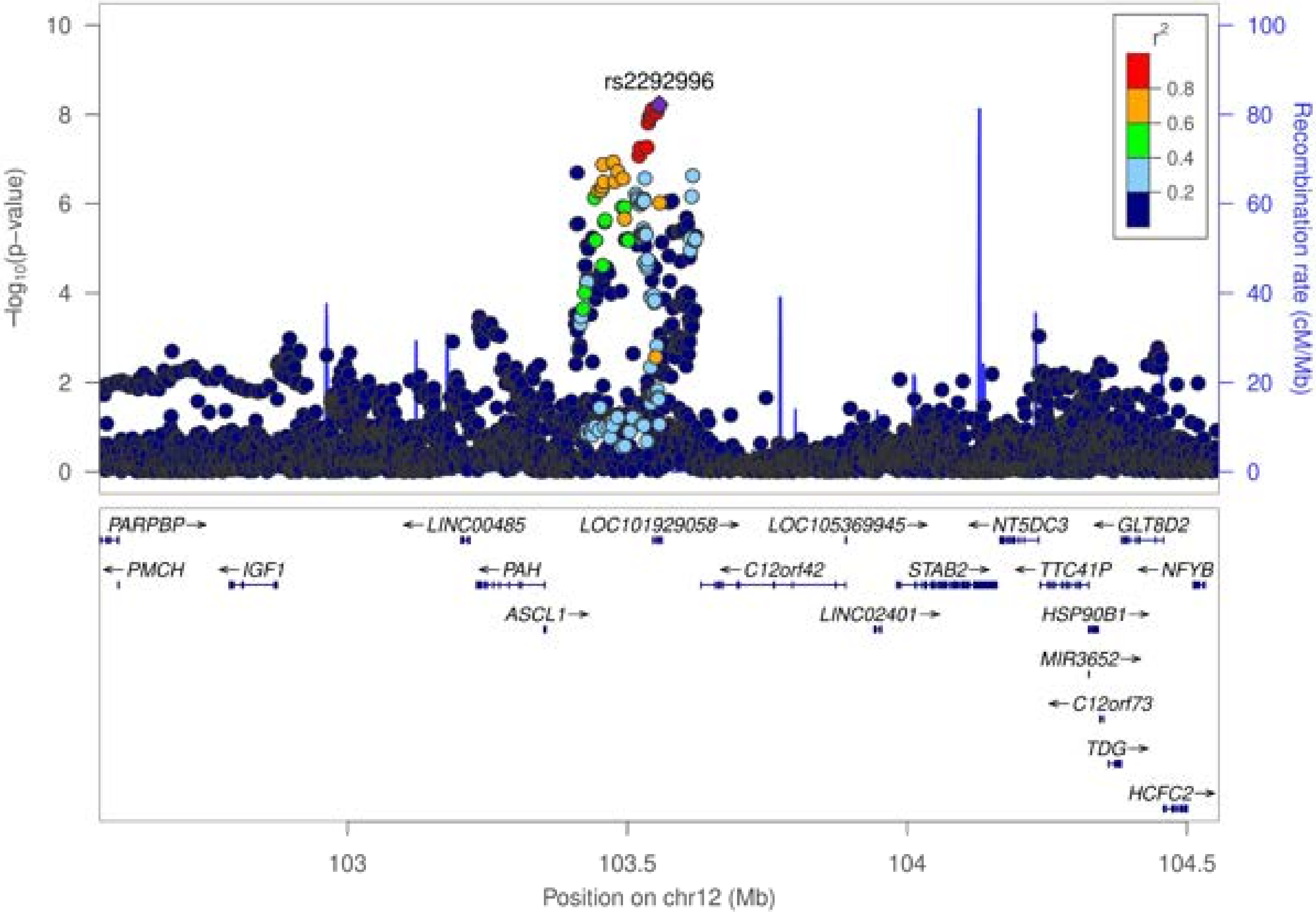

rs2292996
r2
0.8
0.6
0.4
0.2
−log10(p−value)
Recombination rate (cM/Mb)
10
8
6
4
2
0
100
80
60
40
20
0
PARPBP→
←PMCH
←IGF1
←LINC00485
←PAH
ASCL1→
LOC101929058→
←C12orf42
LOC105369945→
LINC02401→
STAB2→
←NT5DC3
←TTC41P
HSP90B1→
MIR3652→
←C12orf73
TDG→
←GLT8D2
←NFYB
HCFC2→
103
103.5
104
104.5
Position on chr12 (Mb)

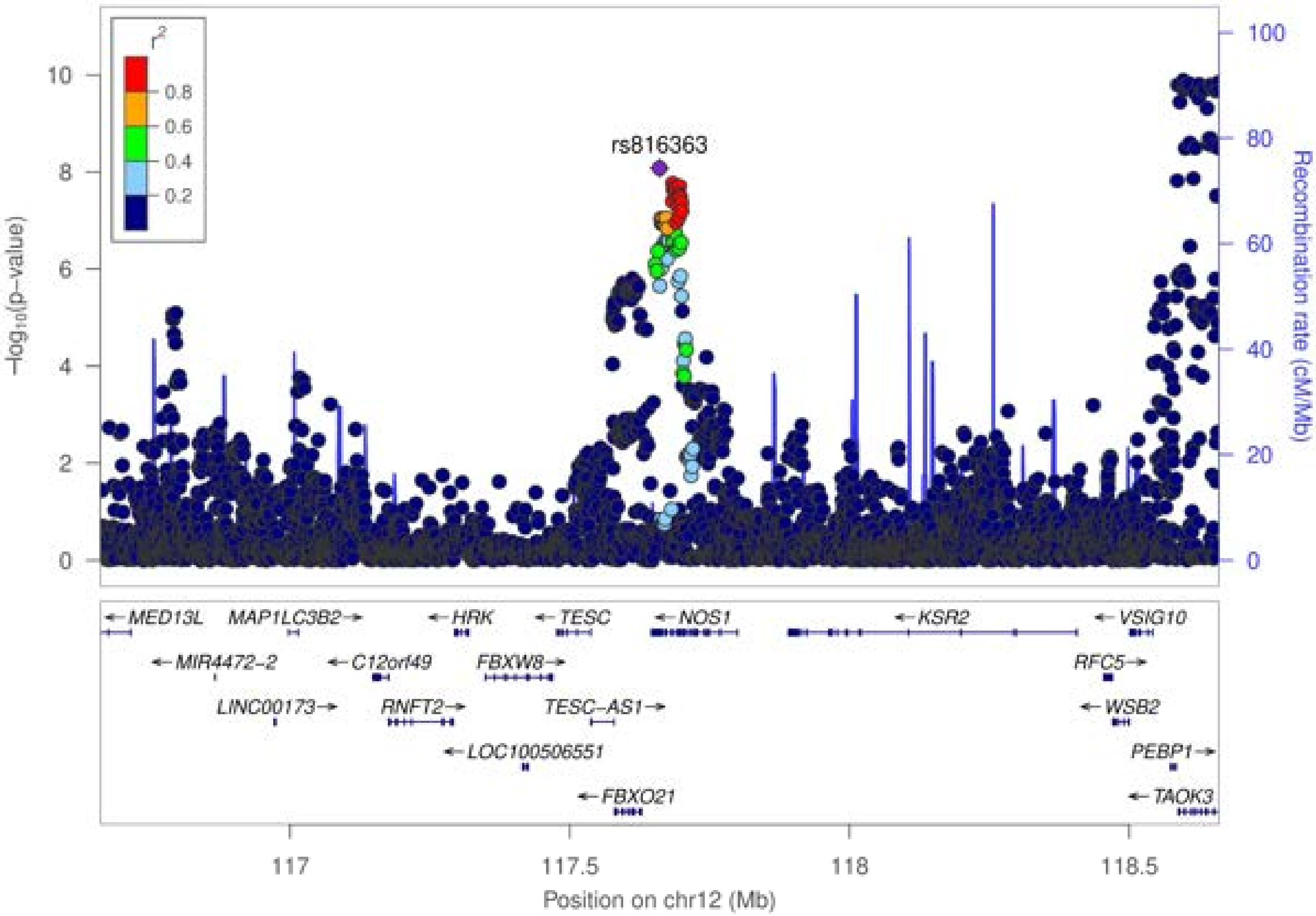

rs816363
r²
0.8
0.6
0.4
0.2
−log10(p−value)
Recombination rate (cM/Mb)
MED13L
MAP1LC3B2
HRK
TESC
NOS1
KSR2
VSIG10
MIR4472-2
C12orf49
FBXW8
RFC5
LINC00173
RNFT2
TESC-AS1
WSB2
LOC100506551
PEBP1
FBXO21
TAOK3
117
117.5
118
118.5
Position on chr12 (Mb)

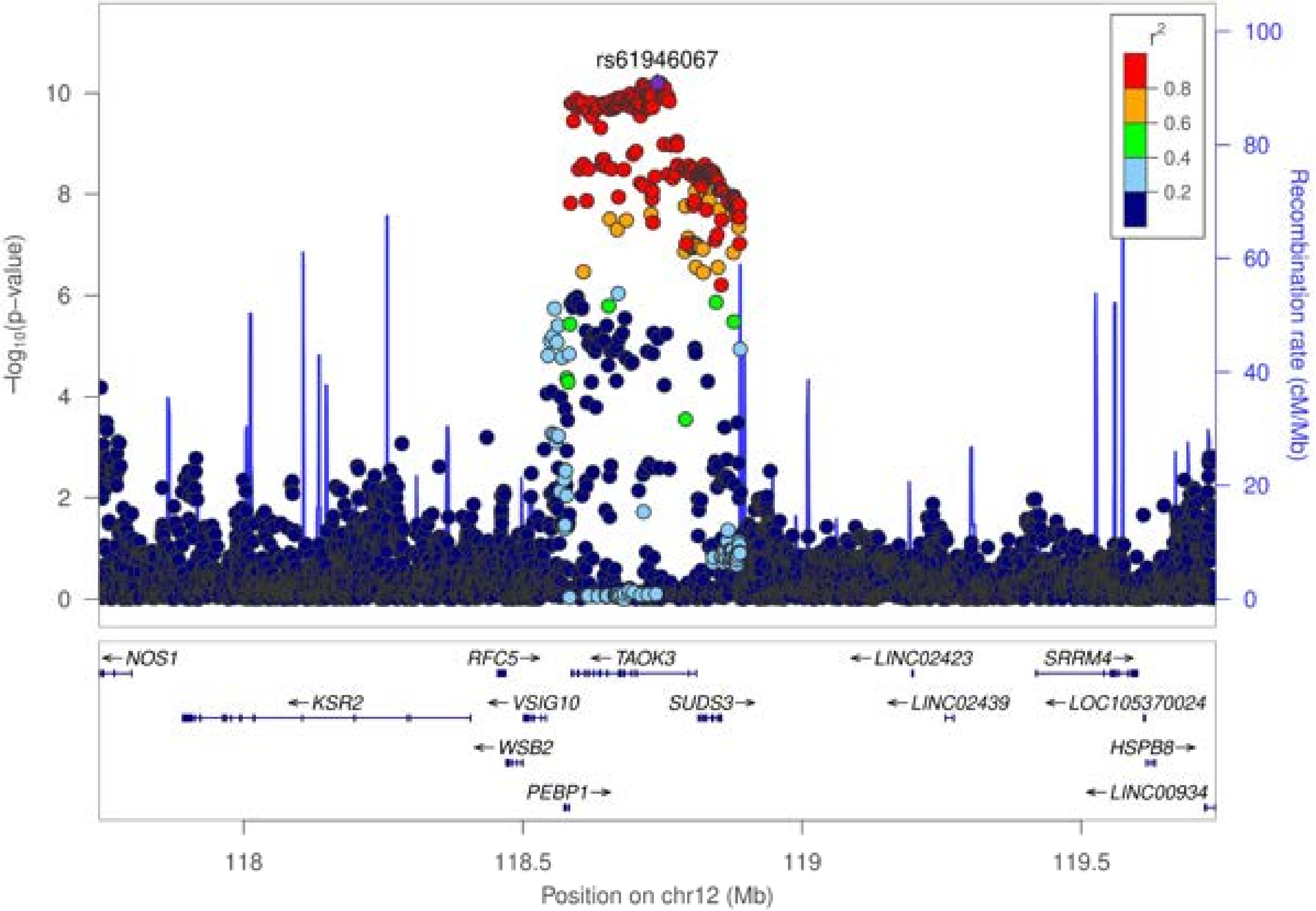

rs61946067
r²
0.8
0.6
0.4
0.2
−log₁₀(p−value)
Recombination rate (cM/Mb)
0
2
4
6
8
10
0
20
40
60
80
100
NOS1
KSR2
RFC5
VSIG10
WSB2
PEBP1
TAOK3
SUDS3
LINC02423
LINC02439
SRRM4
LOC105370024
HSPB8
LINC00934
118
118.5
119
119.5
Position on chr12 (Mb)

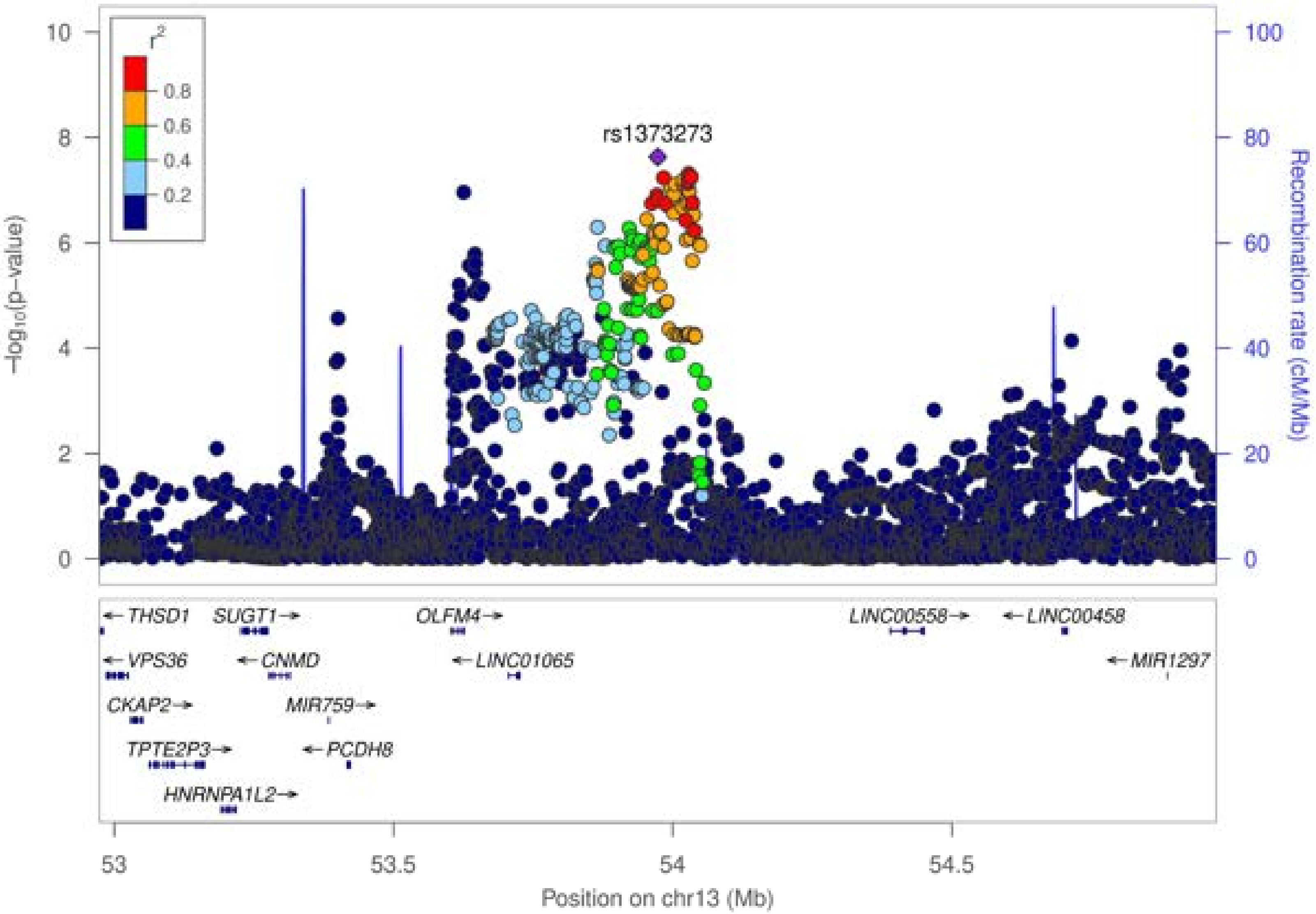

rs1373273
r²
0.8
0.6
0.4
0.2
−log10(p−value)
Recombination rate (cM/Mb)
10
8
6
4
2
0
100
80
60
40
20
0
THSD1
SUGT1
OLFM4
LINC00558
LINC00458
VPS36
CNMD
LINC01065
MIR1297
CKAP2
MIR759
TPTE2P3
PCDH8
HNRNPA1L2
53
53.5
54
54.5
Position on chr13 (Mb)

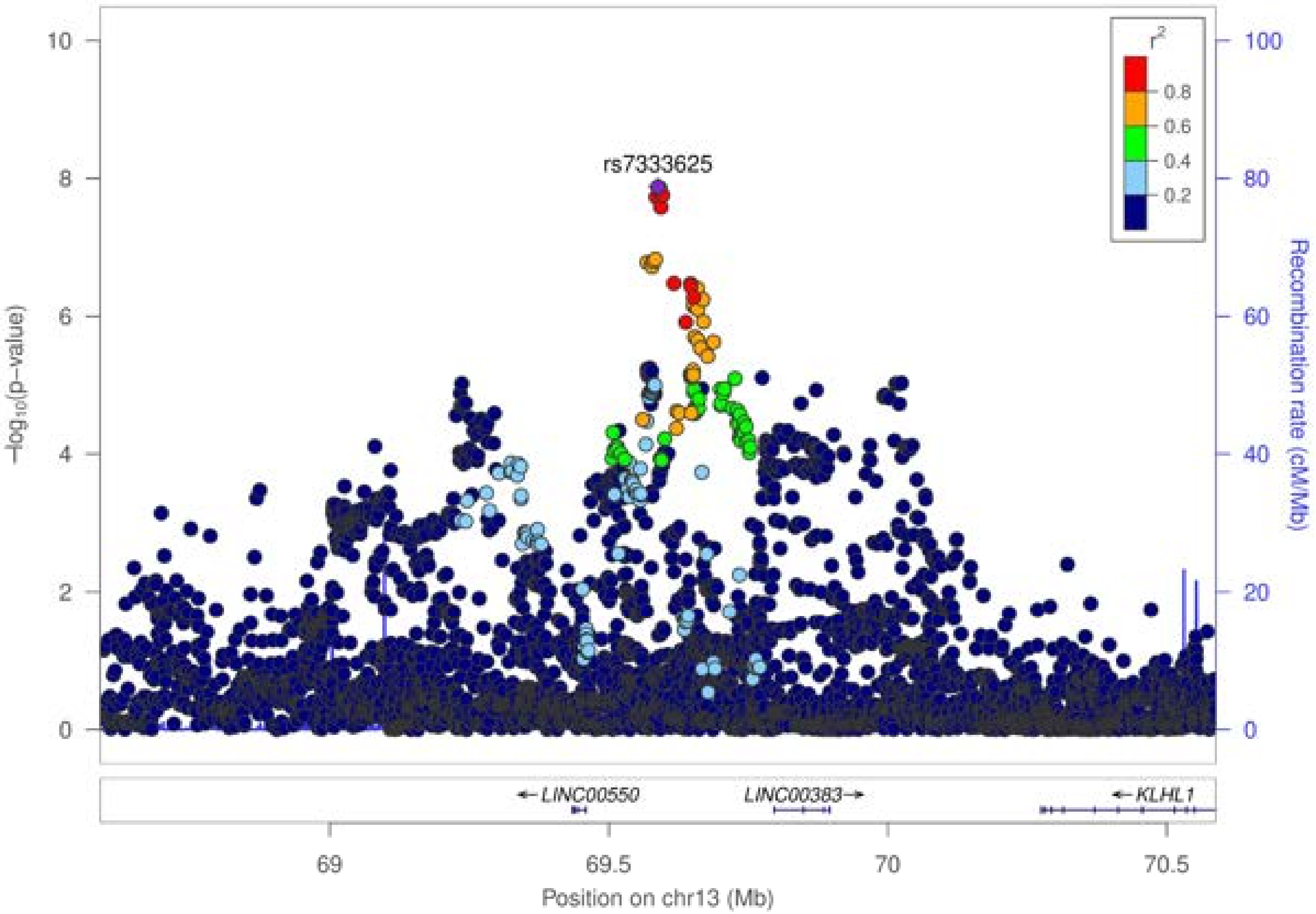

rs7333625
r2
0.8
0.6
0.4
0.2
−log10(p−value)
Recombination rate (cM/Mb)
LINC00550
LINC00383
KLHL1
69
69.5
70
70.5
Position on chr13 (Mb)

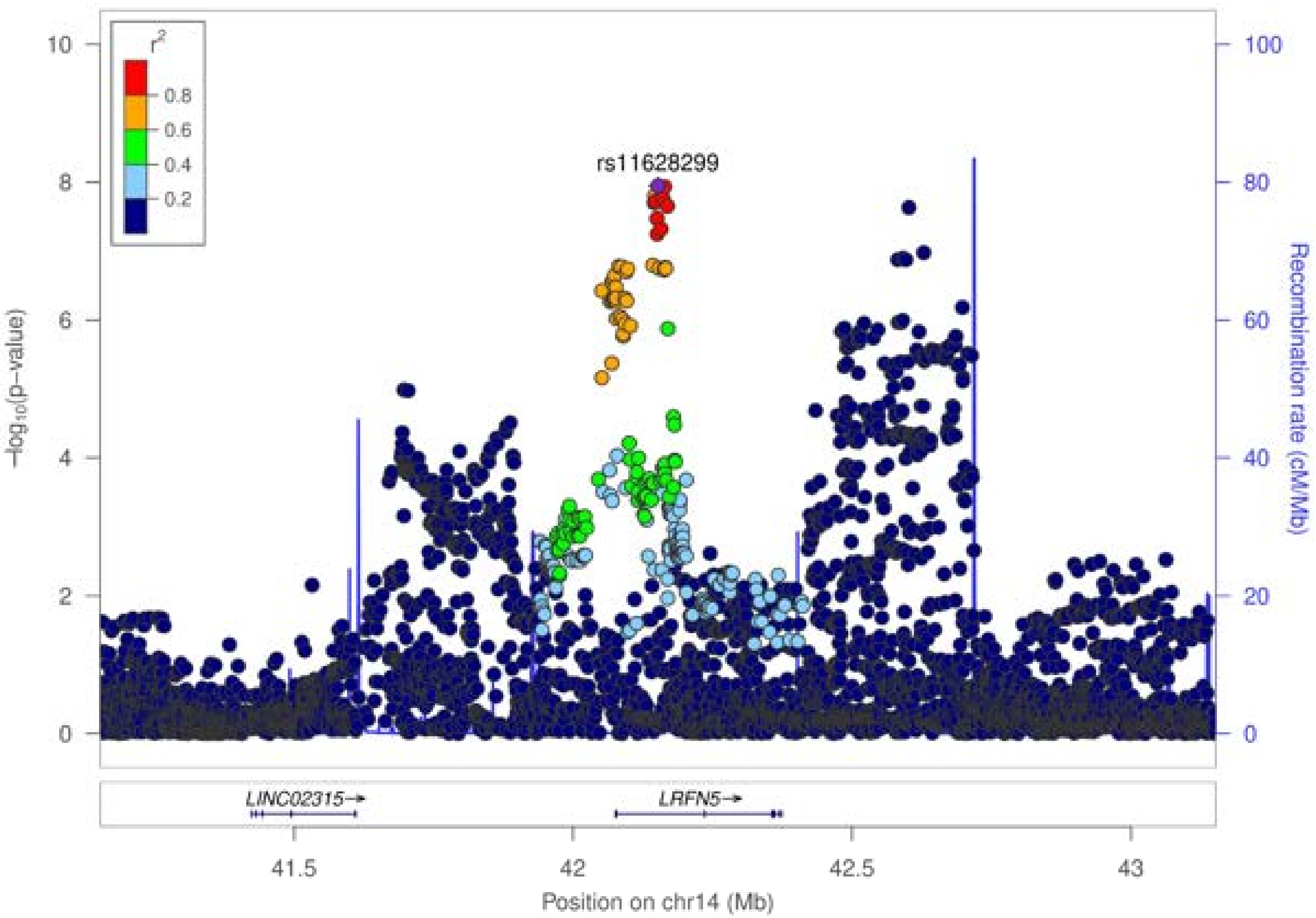

rs11628299
r²
0.8
0.6
0.4
0.2
−log10(p−value)
Recombination rate (cM/Mb)
LINC02315→
LRFN5→
41.5
42
42.5
43
Position on chr14 (Mb)

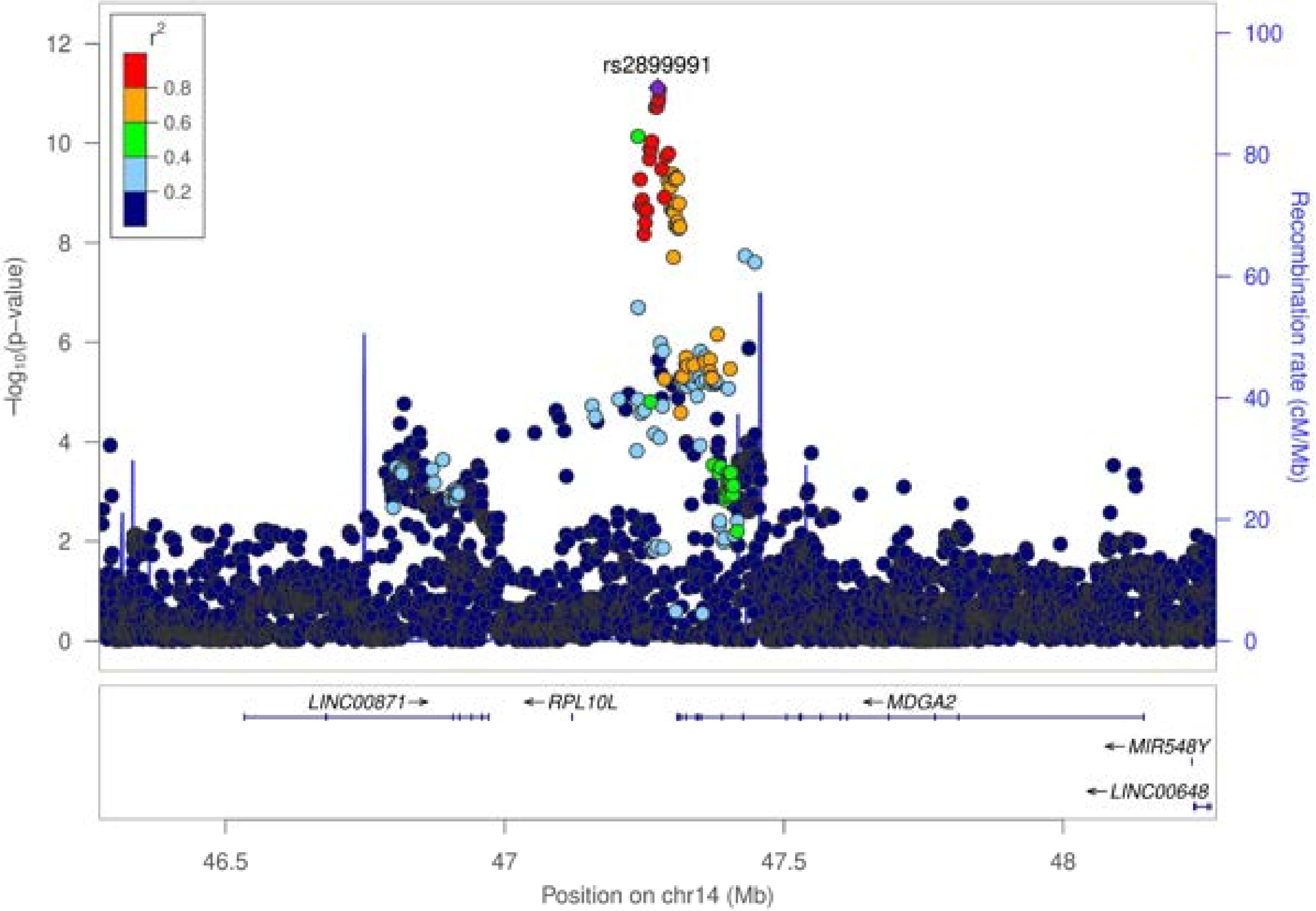
rs2899991
r²
0.8
0.6
0.4
0.2
−log₁₀(p−value)
Recombination rate (cM/Mb)
LINC00871→
←RPL10L
←MDGA2
←MIR548Y
←LINC00648
46.5
47
47.5
48
Position on chr14 (Mb)

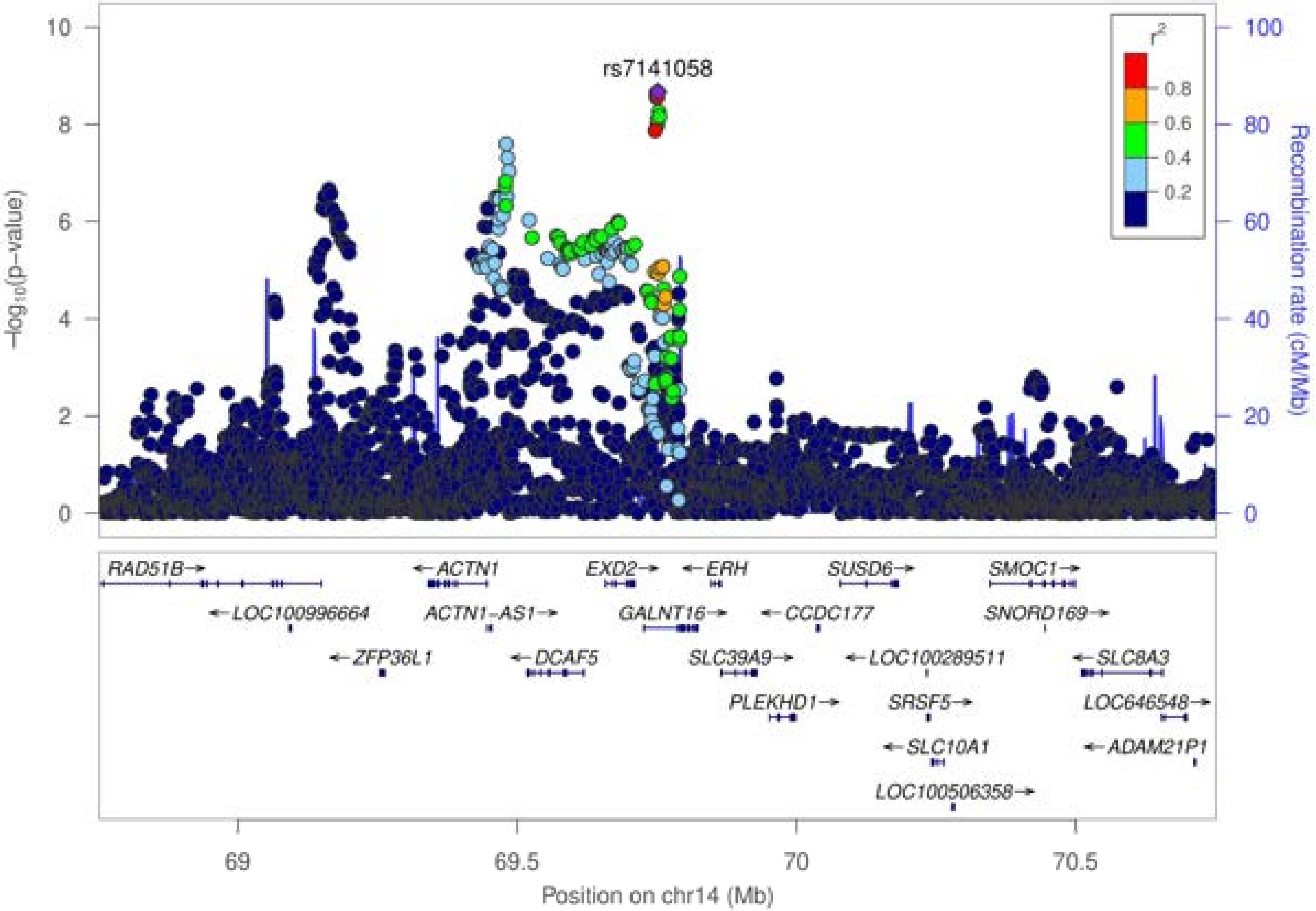

rs7141058
r²
0.8
0.6
0.4
0.2
10
8
6
4
2
0
100
80
60
40
20
0
−log10(p−value)
Recombination rate (cM/Mb)
RAD51B→
←ACTN1
EXD2→
←ERH
SUSD6→
SMOC1→
←LOC100996664
ACTN1−AS1→
GALNT16→
←CCDC177
SNORD169→
←ZFP36L1
←DCAF5
SLC39A9→
←LOC100289511
←SLC8A3
PLEKHD1→
SRSF5→
LOC646548→
←SLC10A1
←ADAM21P1
LOC100506358→
69
69.5
70
70.5
Position on chr14 (Mb)

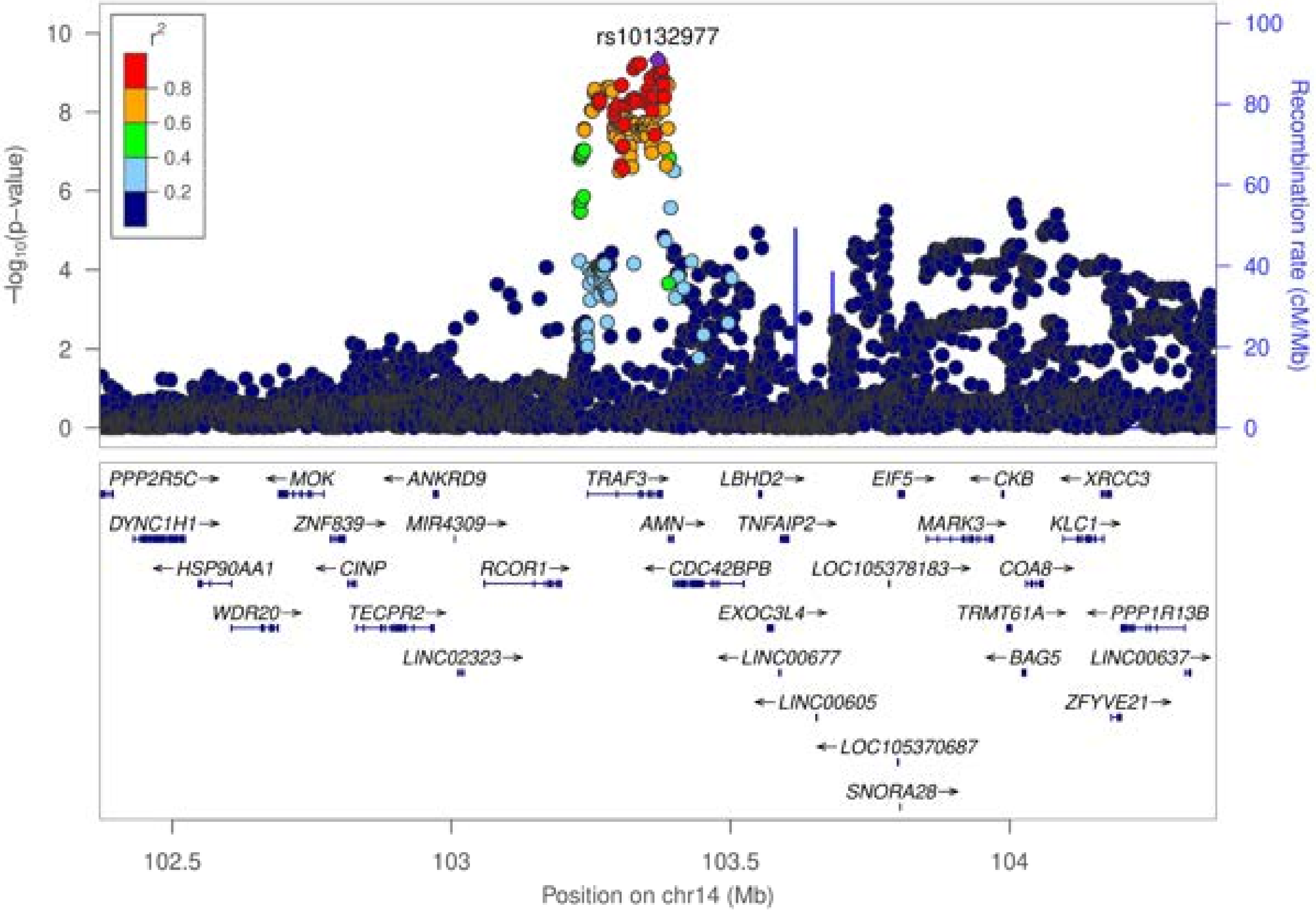

rs10132977
r²
0.8
0.6
0.4
0.2
−log10(p−value)
Recombination rate (cM/Mb)
PPP2R5C→
←MOK
←ANKRD9
TRAF3→
LBHD2→
EIF5→
←CKB
←XRCC3
DYNC1H1→
ZNF839→
MIR4309→
AMN→
TNFAIP2→
MARK3→
KLC1→
←HSP90AA1
←CINP
RCOR1→
←CDC42BPB
LOC105378183→
COA8→
WDR20→
TECPR2→
EXOC3L4→
TRMT61A→
←PPP1R13B
LINC02323→
←LINC00677
←BAG5
LINC00637→
←LINC00605
ZFYVE21→
←LOC105370687
SNORA28→
102.5
103
103.5
104
Position on chr14 (Mb)

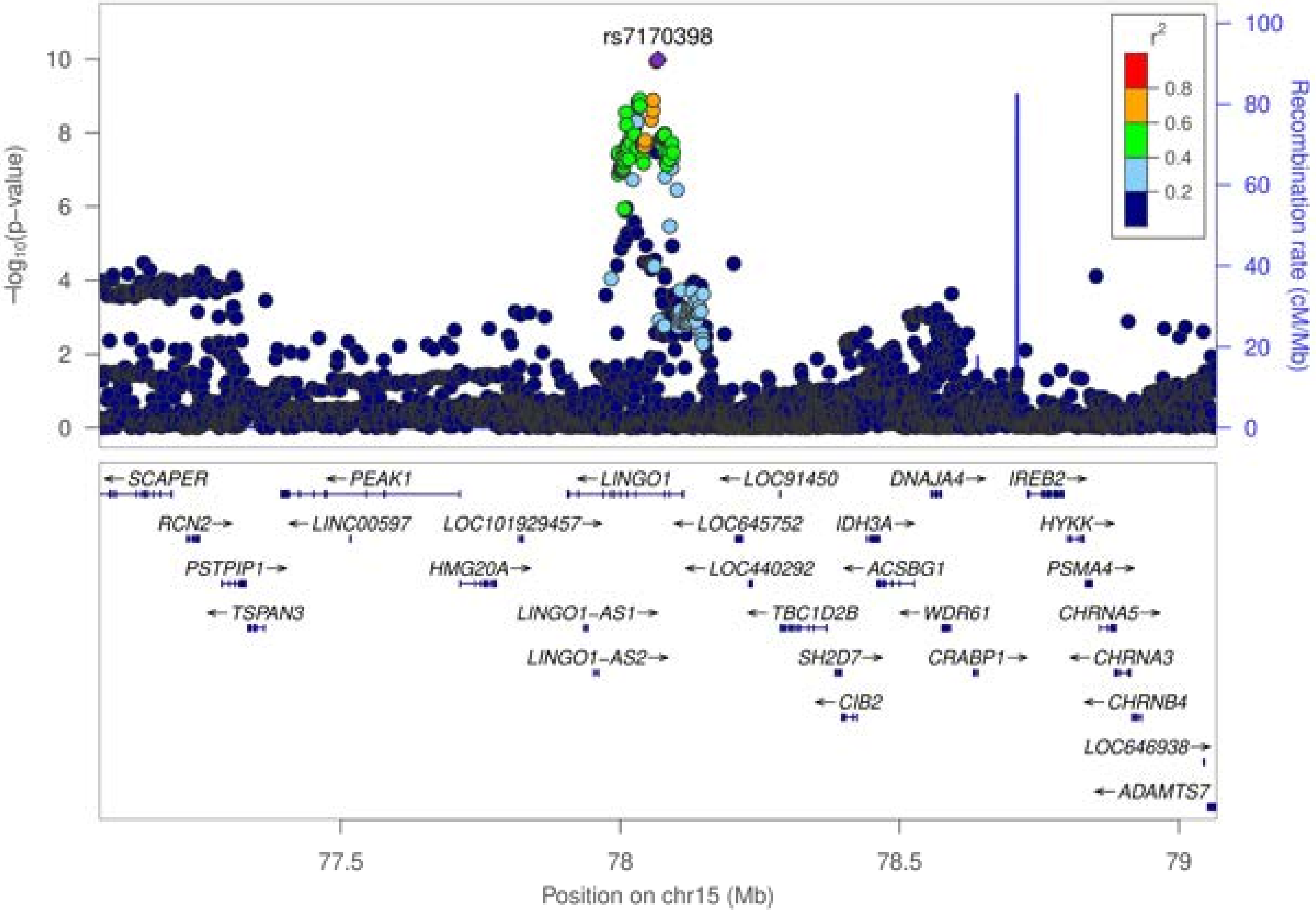

rs7170398
−log10(p−value)
Recombination rate (cM/Mb)
r2
0.8
0.6
0.4
0.2
0
2
4
6
8
10
20
40
60
80
100
SCAPER
PEAK1
LINGO1
LOC91450
DNAJA4
IREB2
RCN2
LINC00597
LOC101929457
LOC645752
IDH3A
HYKK
PSTPIP1
HMG20A
LOC440292
ACSBG1
PSMA4
TSPAN3
LINGO1-AS1
TBC1D2B
WDR61
CHRNA5
LINGO1-AS2
SH2D7
CRABP1
CHRNA3
CIB2
CHRNB4
LOC646938
ADAMTS7
77.5
78
78.5
79
Position on chr15 (Mb)

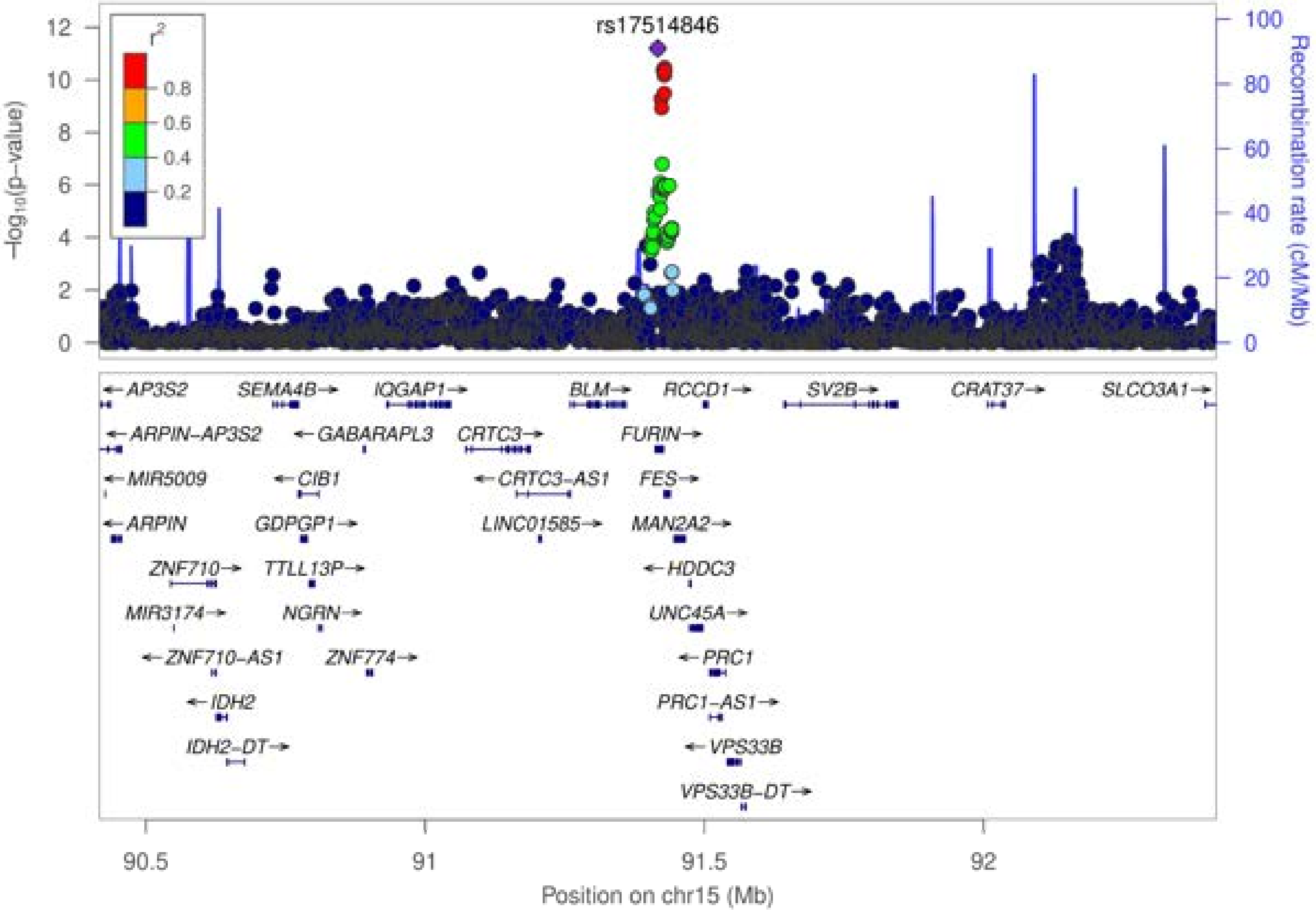

rs17514846
r²
0.8
0.6
0.4
0.2
−log10(p−value)
Recombination rate (cM/Mb)
AP3S2
SEMA4B
IQGAP1
BLM
RCCD1
SV2B
CRAT37
SLCO3A1
ARPIN−AP3S2
GABARAPL3
CRTC3
FURIN
MIR5009
CIB1
CRTC3−AS1
FES
ARPIN
GDPGP1
LINC01585
MAN2A2
ZNF710
TTLL13P
HDDC3
MIR3174
NGRN
UNC45A
ZNF710−AS1
ZNF774
PRC1
IDH2
PRC1−AS1
IDH2−DT
VPS33B
VPS33B−DT
Position on chr15 (Mb)

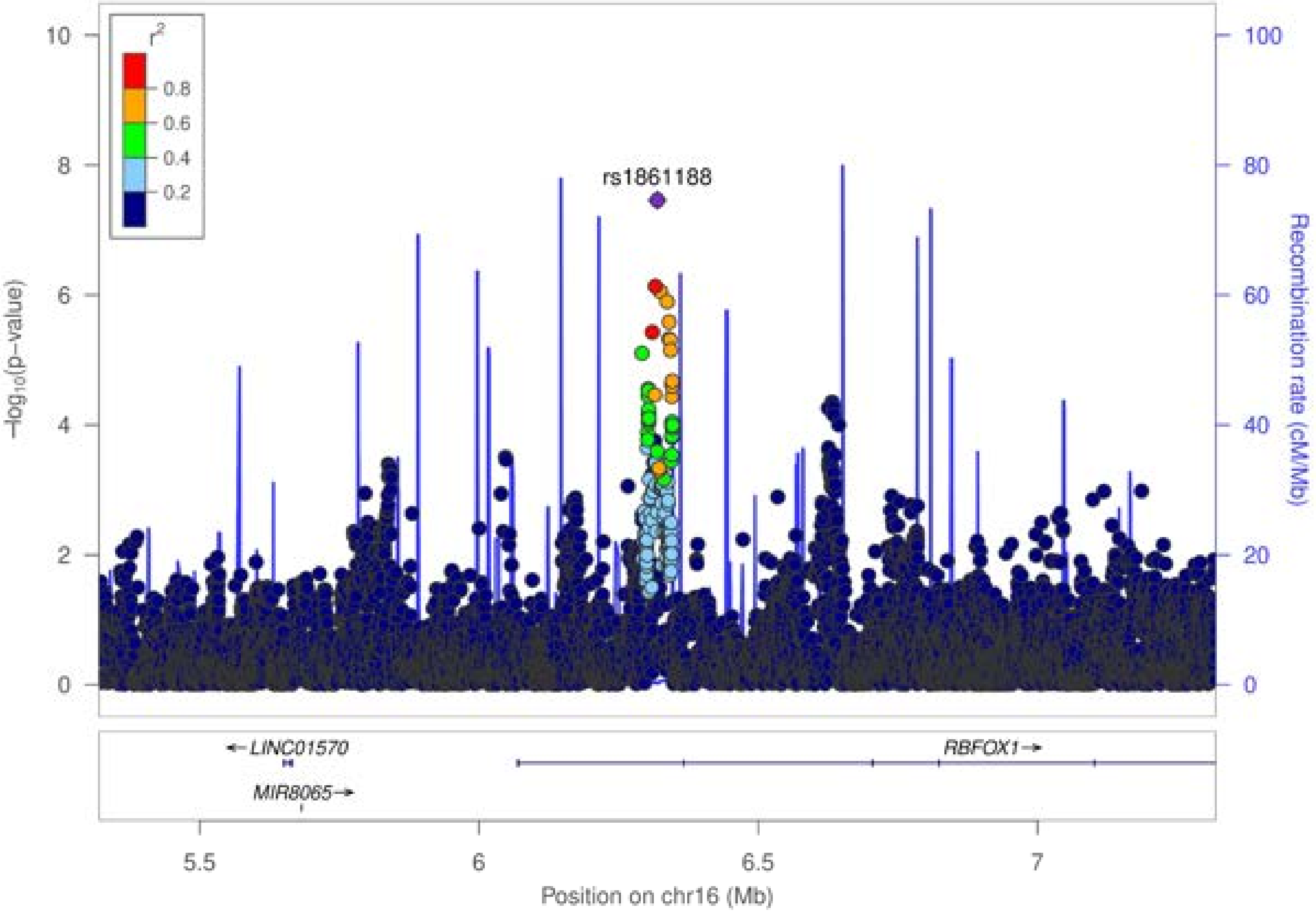

r²
0.8
0.6
0.4
0.2
rs1861188
−log₁₀(p−value)
Recombination rate (cM/Mb)
0
2
4
6
8
10
0
20
40
60
80
100
← LINC01570
MIR8065 →
RBFOX1 →
5.5
6
6.5
7
Position on chr16 (Mb)

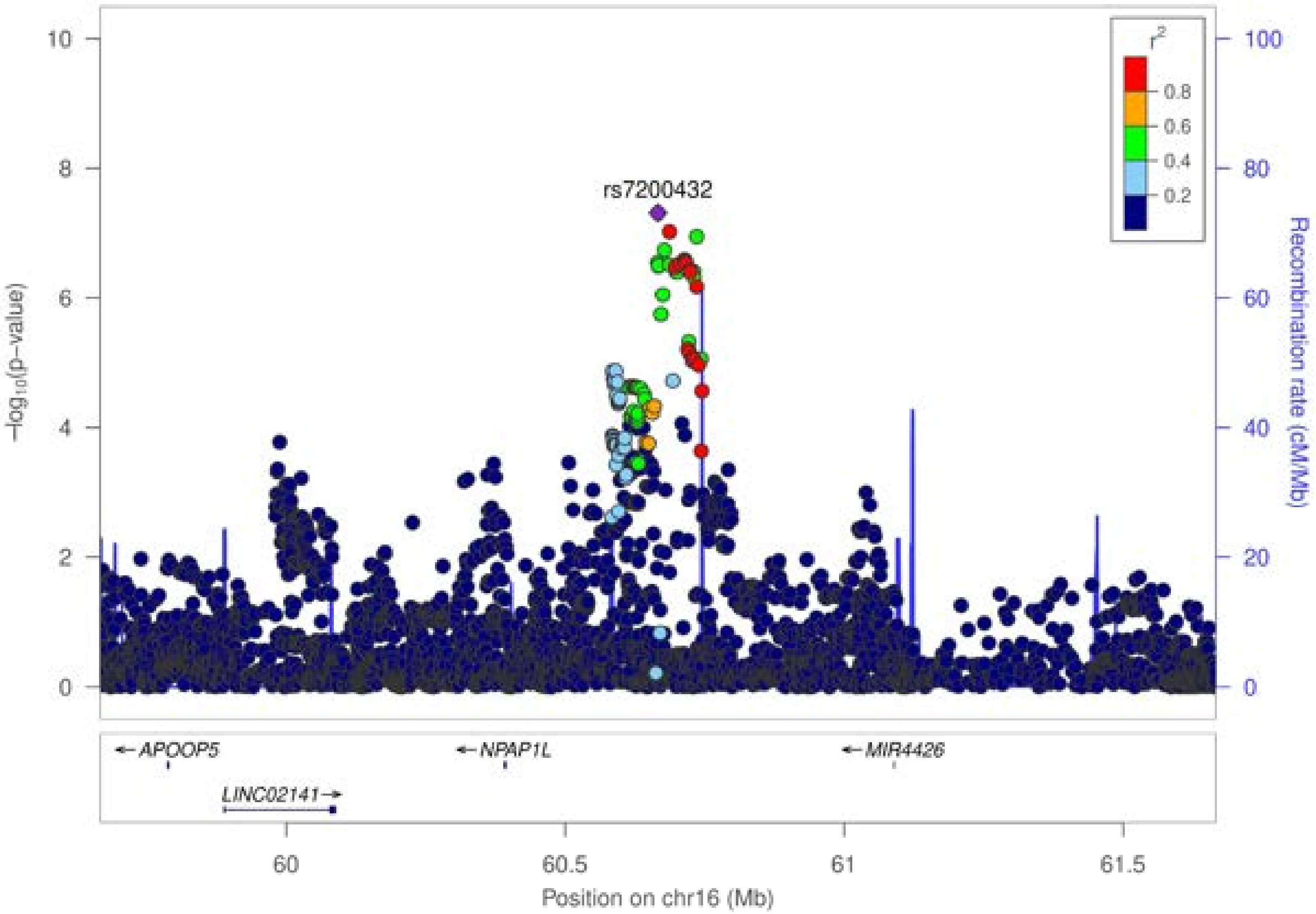

rs7200432
r²
0.8
0.6
0.4
0.2
−log10(p−value)
Recombination rate (cM/Mb)
0
2
4
6
8
10
0
20
40
60
80
100
APOOP5
LINC02141
NPAP1L
MIR4426
60
60.5
61
61.5
Position on chr16 (Mb)

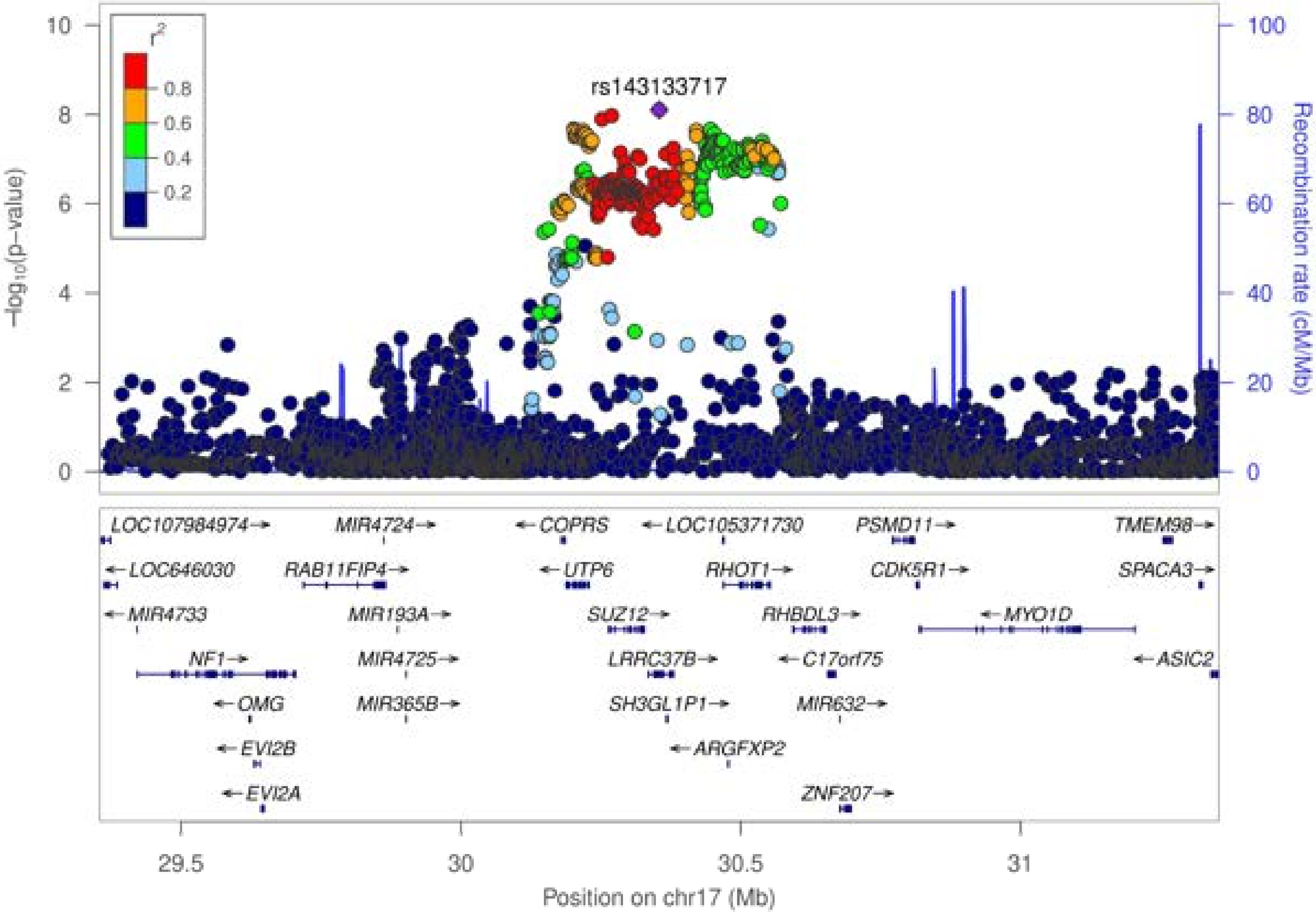

rs143133717
r²
0.8
0.6
0.4
0.2
−log10(p−value)
Recombination rate (cM/Mb)
LOC107984974→
MIR4724→
←COPRS
←LOC105371730
PSMD11→
TMEM98→
←LOC646030
RAB11FIP4→
←UTP6
RHOT1→
CDK5R1→
SPACA3→
←MIR4733
MIR193A→
SUZ12→
RHBDL3→
←MYO1D
NF1→
MIR4725→
LRRC37B→
←C17orf75
←ASIC2
←OMG
MIR365B→
SH3GL1P1→
MIR632→
←EVI2B
←ARGFXP2
←EVI2A
ZNF207→
29.5
30
30.5
31
Position on chr17 (Mb)

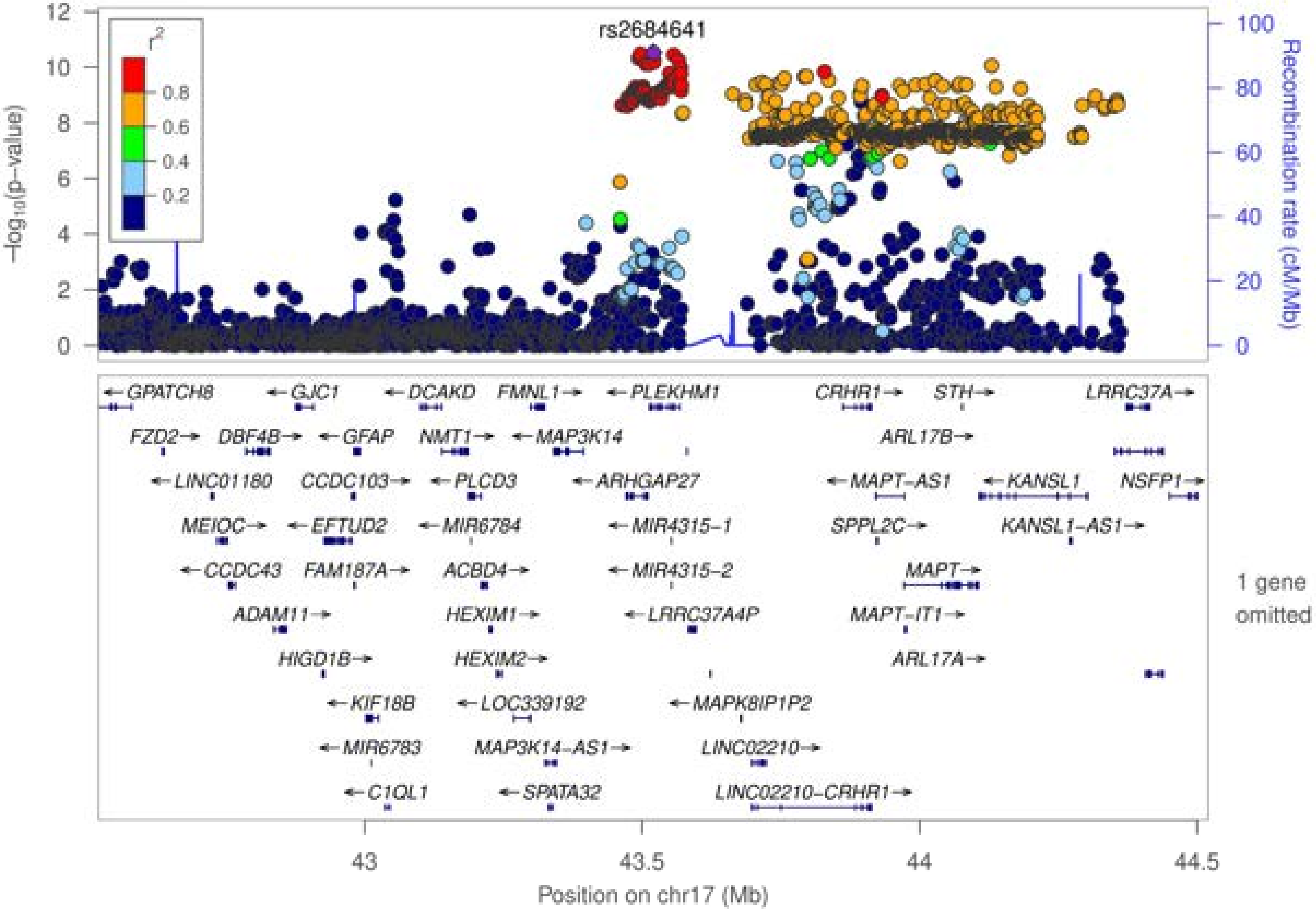

rs2684641
r²
0.8
0.6
0.4
0.2
−log10(p−value)
Recombination rate (cM/Mb)
GPATCH8
GJC1
DCAKD
FMNL1
PLEKHM1
CRHR1
STH
LRRC37A
FZD2
DBF4B
GFAP
NMT1
MAP3K14
ARL17B
LINC01180
CCDC103
PLCD3
ARHGAP27
MAPT−AS1
KANSL1
NSFP1
MEIOC
EFTUD2
MIR6784
MIR4315−1
SPPL2C
KANSL1−AS1
CCDC43
FAM187A
ACBD4
MIR4315−2
MAPT
ADAM11
HEXIM1
LRRC37A4P
MAPT−IT1
HIGD1B
HEXIM2
ARL17A
KIF18B
LOC339192
MAPK8IP1P2
MIR6783
MAP3K14−AS1
LINC02210
C1QL1
SPATA32
LINC02210−CRHR1
1 gene omitted
43
43.5
44
44.5
Position on chr17 (Mb)

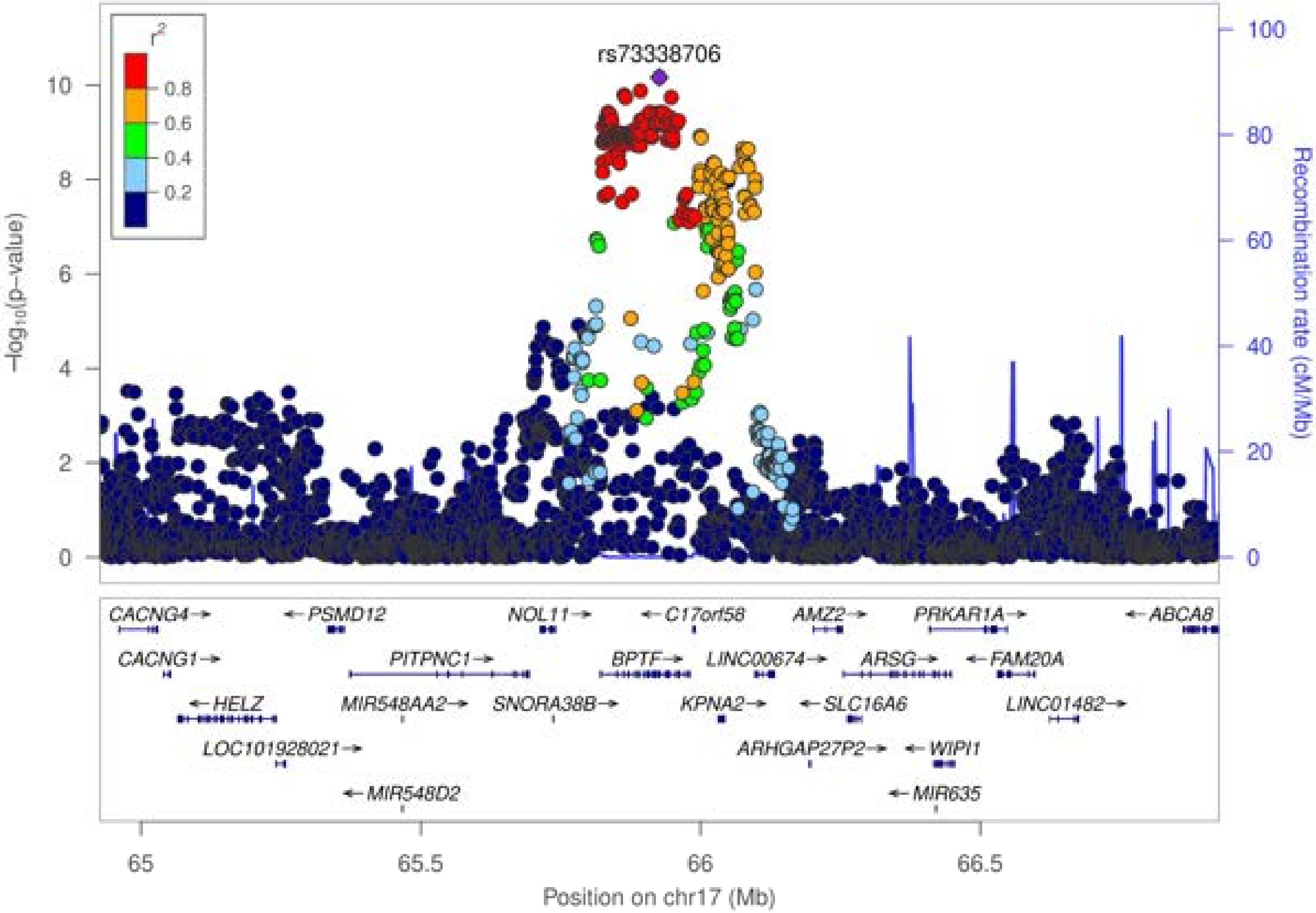

rs73338706
r²
0.8
0.6
0.4
0.2
−log₁₀(p−value)
Recombination rate (cM/Mb)
Position on chr17 (Mb)
CACNG4
PSMD12
NOL11
C17orf58
AMZ2
PRKAR1A
ABCA8
CACNG1
PITPNC1
BPTF
LINC00674
ARSG
FAM20A
HELZ
MIR548AA2
SNORA38B
KPNA2
SLC16A6
LINC01482
LOC101928021
ARHGAP27P2
WIPI1
MIR548D2
MIR635

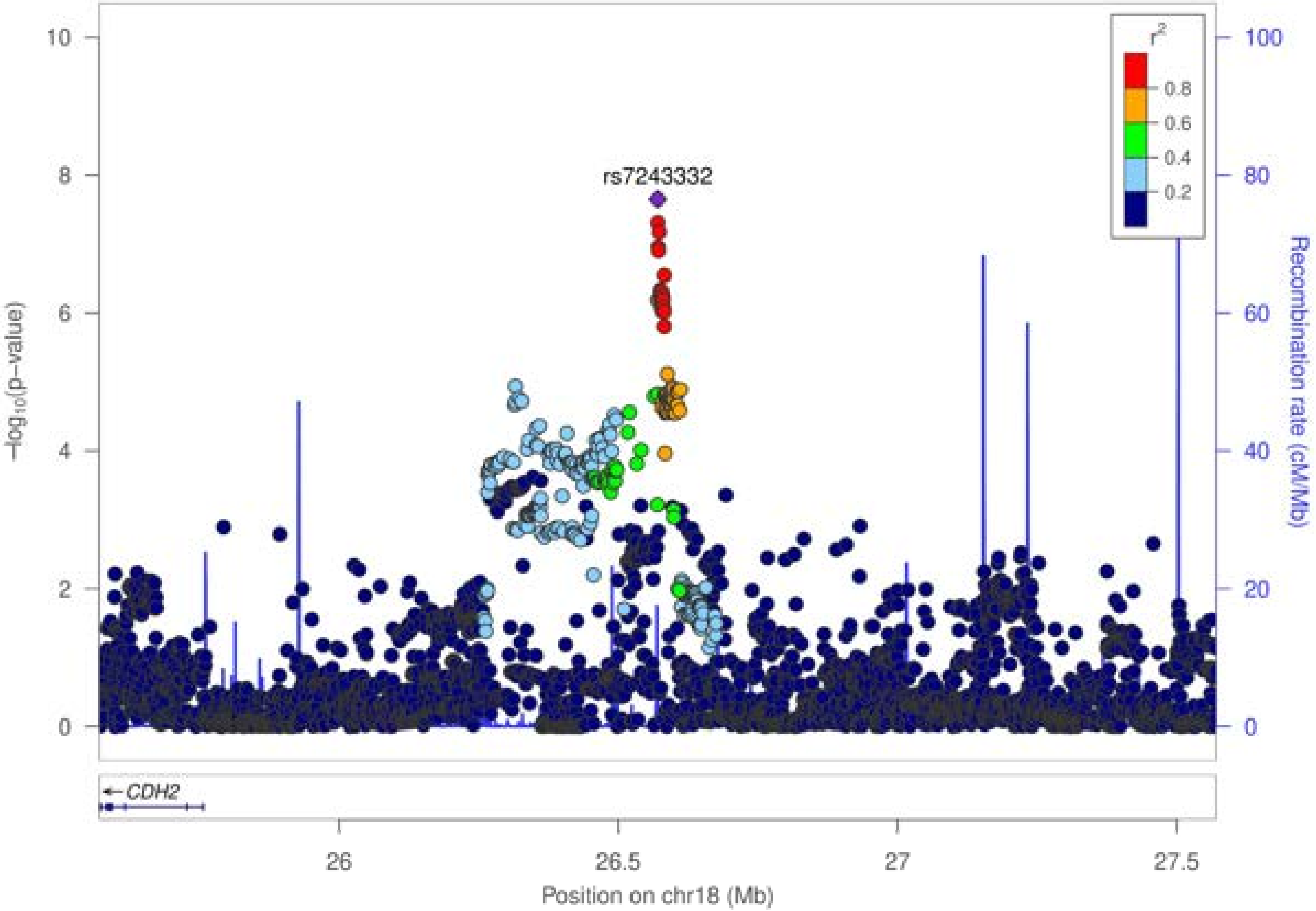

rs7243332
r²
0.8
0.6
0.4
0.2
−log10(p−value)
Recombination rate (cM/Mb)
CDH2
Position on chr18 (Mb)
26
26.5
27
27.5
0
2
4
6
8
10
20
40
60
80
100

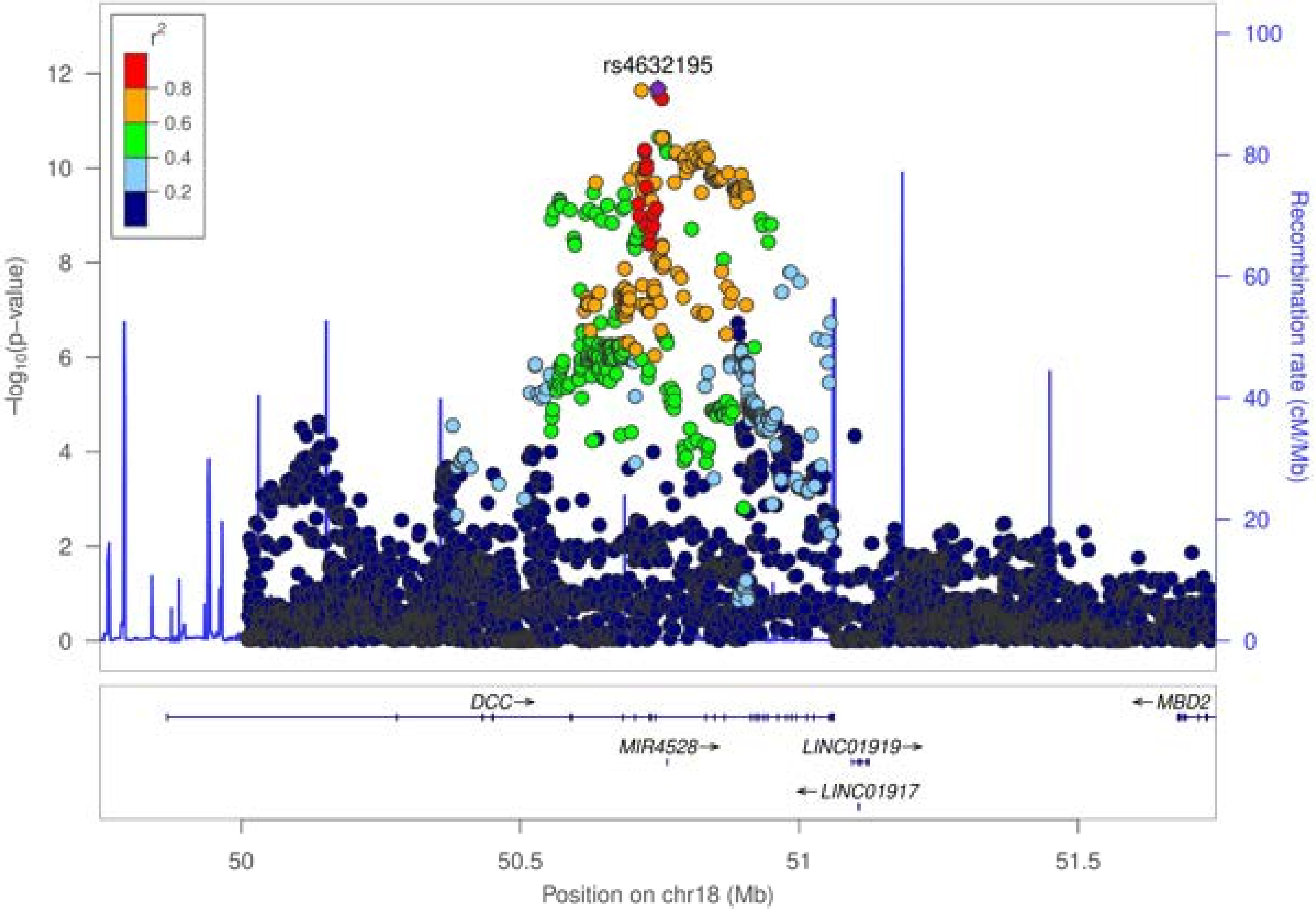

r²
0.8
0.6
0.4
0.2
rs4632195
−log10(p−value)
Recombination rate (cM/Mb)
DCC→
←MBD2
MIR4528→
LINC01919→
←LINC01917
Position on chr18 (Mb)

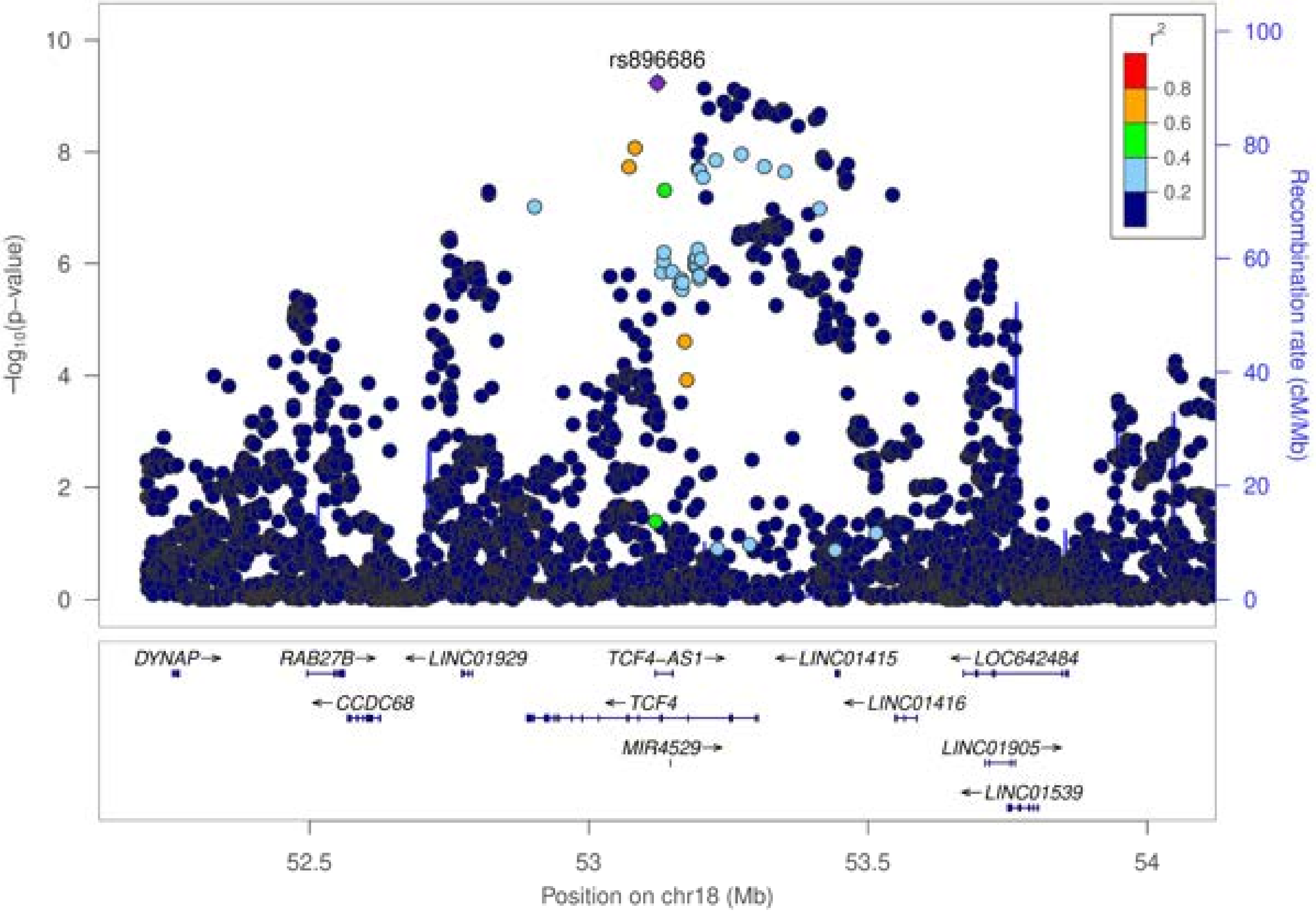

rs896686
r2
0.8
0.6
0.4
0.2
−log10(p−value)
Recombination rate (cM/Mb)
10
8
6
4
2
0
100
80
60
40
20
0
DYNAP→
RAB27B→
←LINC01929
TCF4−AS1→
←LINC01415
←LOC642484
←CCDC68
←TCF4
←LINC01416
MIR4529→
LINC01905→
←LINC01539
52.5
53
53.5
54
Position on chr18 (Mb)

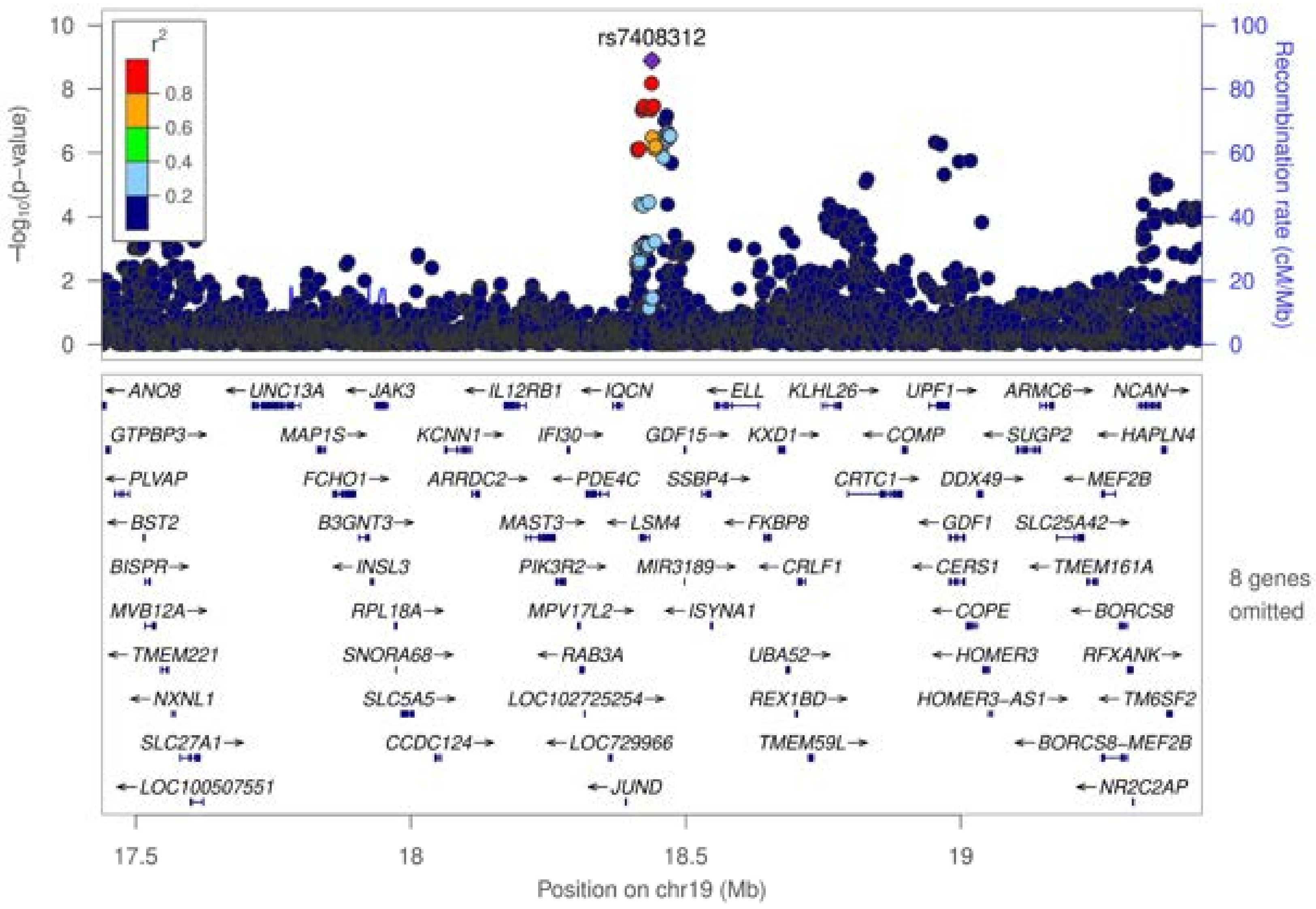

rs7408312
r²
0.8
0.6
0.4
0.2
−log10(p−value)
Recombination rate (cM/Mb)
ANO8
UNC13A
JAK3
IL12RB1
IQCN
ELL
KLHL26
UPF1
ARMC6
NCAN
GTPBP3
MAP1S
KCNN1
IFI30
GDF15
KXD1
COMP
SUGP2
HAPLN4
PLVAP
FCHO1
ARRDC2
PDE4C
SSBP4
CRTC1
DDX49
MEF2B
BST2
B3GNT3
MAST3
LSM4
FKBP8
GDF1
SLC25A42
BISPR
INSL3
PIK3R2
MIR3189
CRLF1
CERS1
TMEM161A
MVB12A
RPL18A
MPV17L2
ISYNA1
COPE
BORCS8
TMEM221
SNORA68
RAB3A
UBA52
HOMER3
RFXANK
NXNL1
SLC5A5
LOC102725254
REX1BD
HOMER3−AS1
TM6SF2
SLC27A1
CCDC124
LOC729966
TMEM59L
BORCS8−MEF2B
LOC100507551
JUND
NR2C2AP
8 genes omitted
17.5
18
18.5
19
Position on chr19 (Mb)

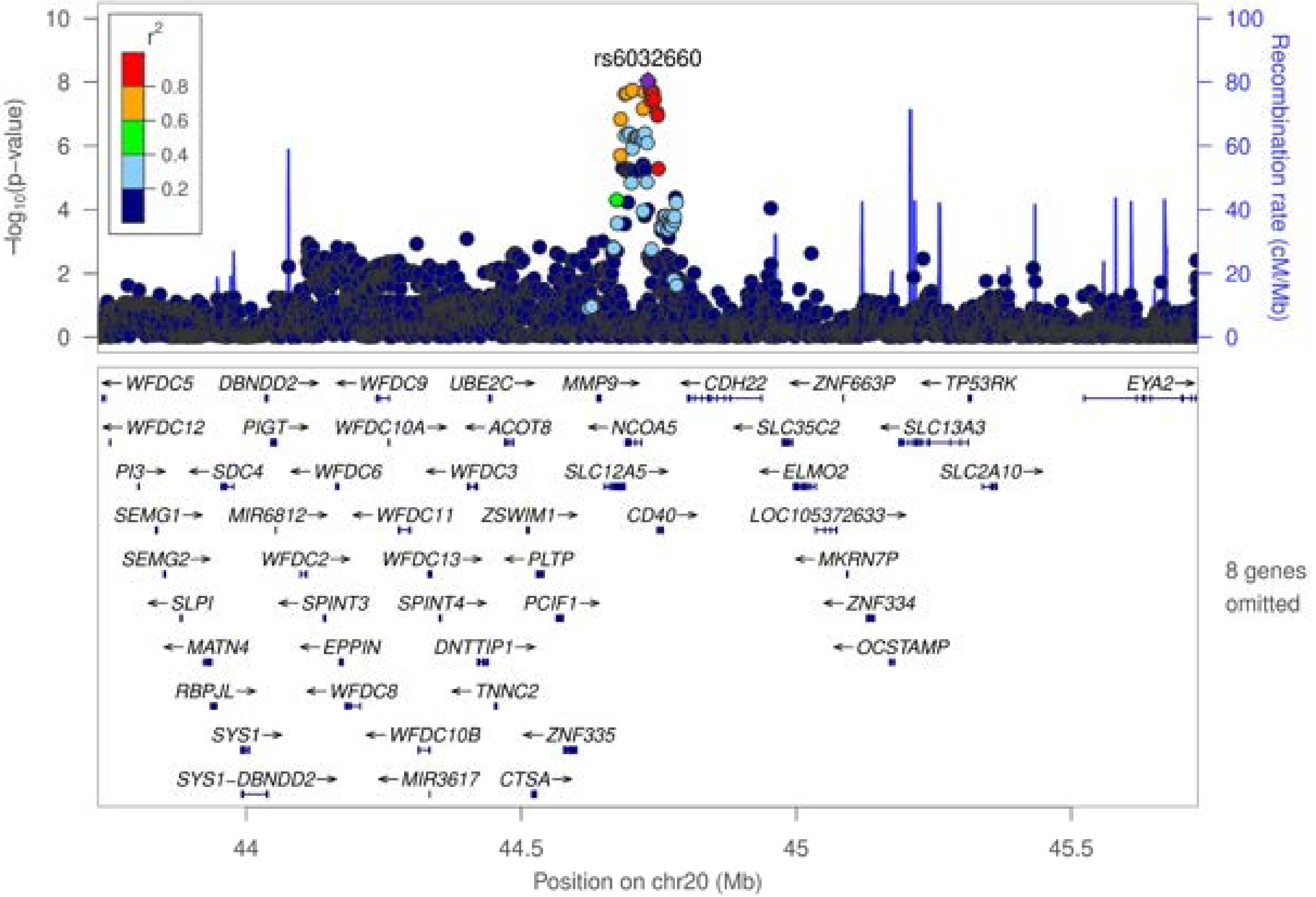
rs6032660
r²
0.8
0.6
0.4
0.2
−log₁₀(p−value)
Recombination rate (cM/Mb)
0
2
4
6
8
10
20
40
60
80
100
WFDC5
DBNDD2
WFDC9
UBE2C
MMP9
CDH22
ZNF663P
TP53RK
EYA2
WFDC12
PIGT
WFDC10A
ACOT8
NCOA5
SLC35C2
SLC13A3
PI3
SDC4
WFDC6
WFDC3
SLC12A5
ELMO2
SLC2A10
SEMG1
MIR6812
WFDC11
ZSWIM1
CD40
LOC105372633
SEMG2
WFDC2
WFDC13
PLTP
MKRN7P
SLPI
SPINT3
SPINT4
PCIF1
ZNF334
MATN4
EPPIN
DNTTIP1
OCSTAMP
RBPJL
WFDC8
TNNC2
SYS1
WFDC10B
ZNF335
SYS1-DBNDD2
MIR3617
CTSA
8 genes omitted
44
44.5
45
45.5
Position on chr20 (Mb)

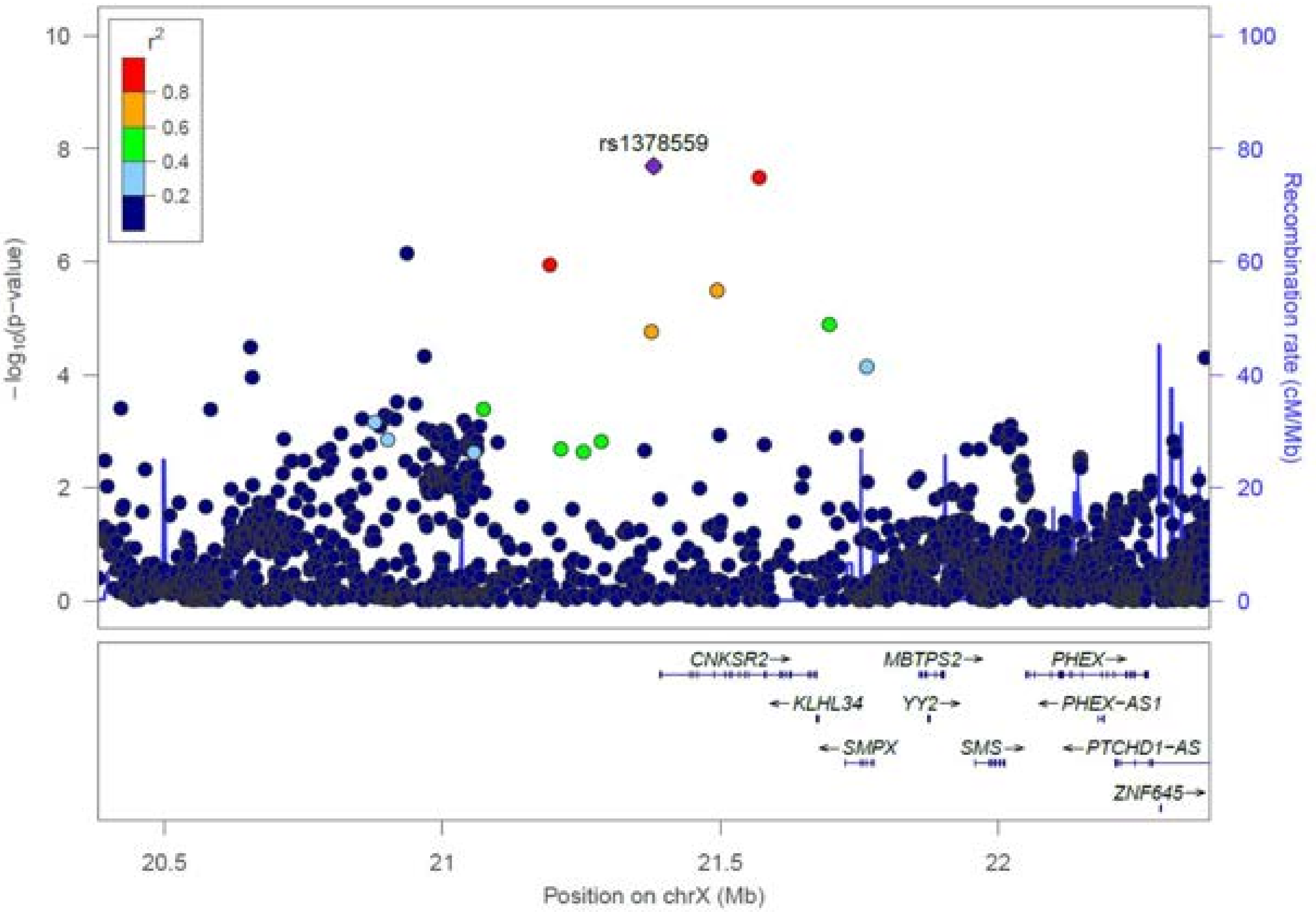

r2
0.8
0.6
0.4
0.2
rs1378559
−log10(p−value)
Recombination rate (cM/Mb)
10
8
6
4
2
0
100
80
60
40
20
0
CNKSR2→
MBTPS2→
PHEX→
←KLHL34
YY2→
←PHEX−AS1
←SMPX
SMS→
←PTCHD1−AS
ZNF645→
20.5
21
21.5
22
Position on chrX (Mb)

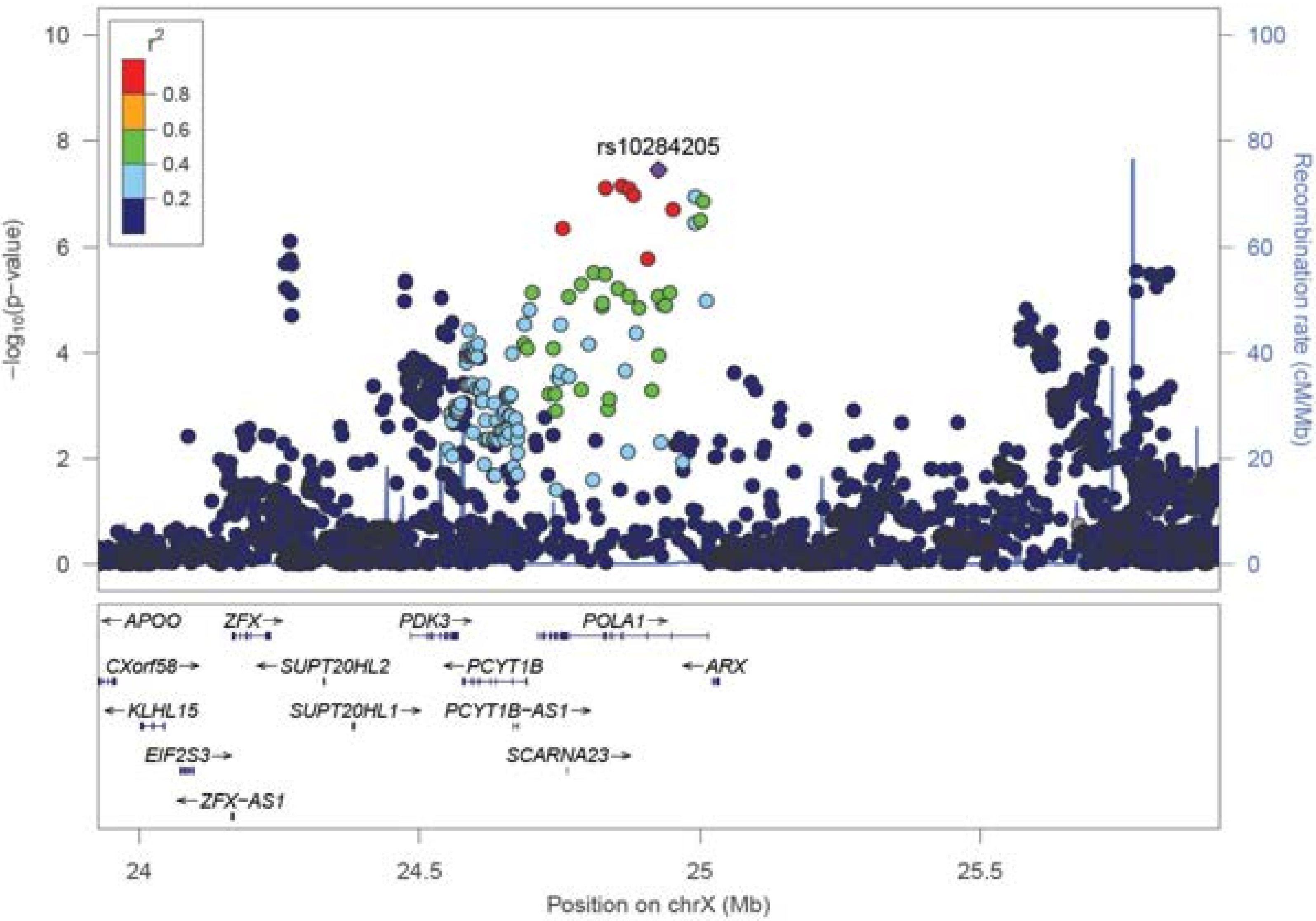

r2
0.8
0.6
0.4
0.2
rs10284205
−log10(p−value)
Recombination rate (cM/Mb)
APOO
ZFX
PDK3
POLA1
CXorf58
SUPT20HL2
PCYT1B
ARX
KLHL15
SUPT20HL1
PCYT1B−AS1
EIF2S3
SCARNA23
ZFX−AS1
Position on chrX (Mb)

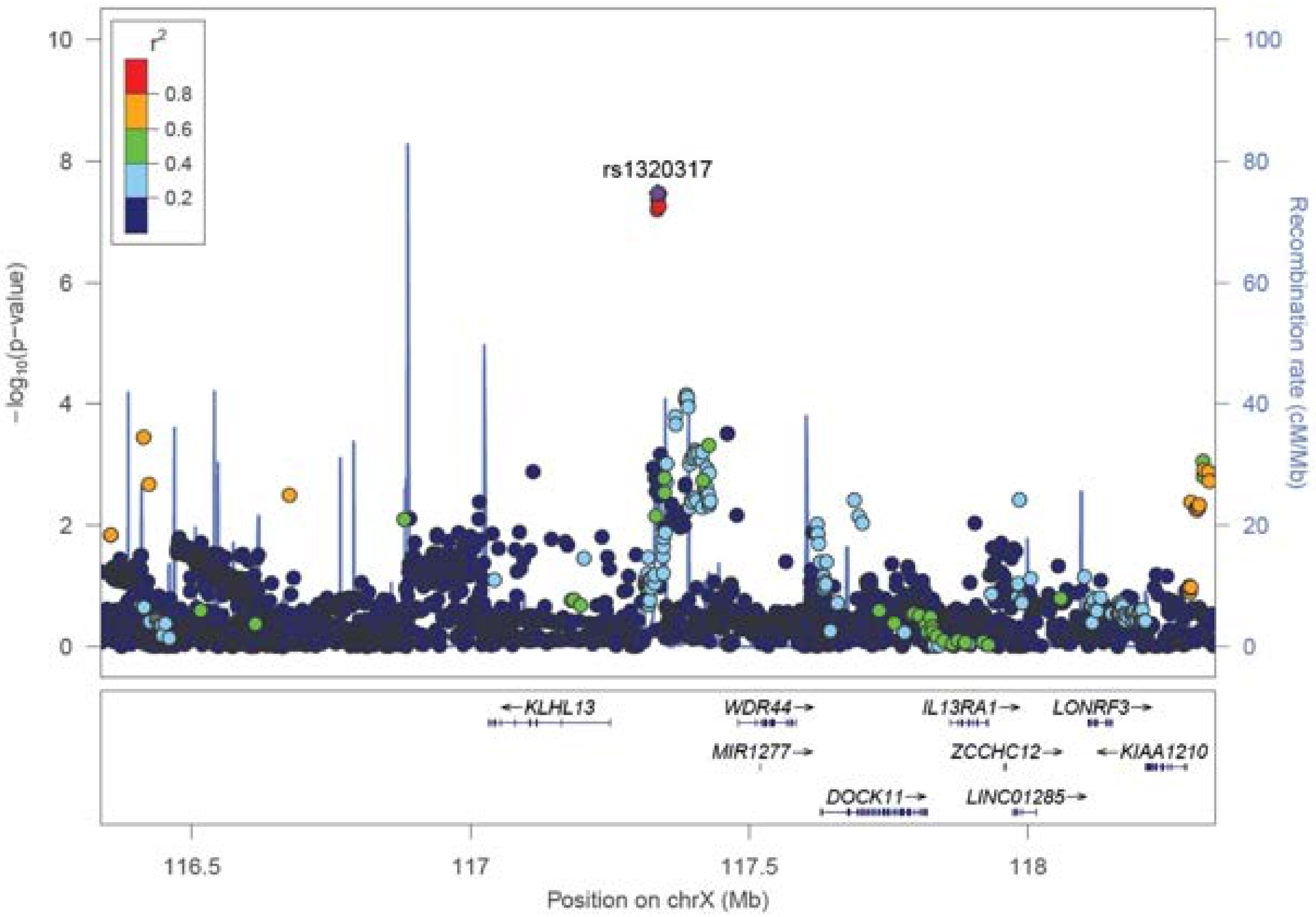

rs1320317
r2
0.8
0.6
0.4
0.2
-log10(p-value)
Recombination rate (cM/Mb)
0
2
4
6
8
10
20
40
60
80
100
KLHL13
WDR44
IL13RA1
LONRF3
MIR1277
ZCCHC12
KIAA1210
DOCK11
LINC01285
116.5
117
117.5
118
Position on chrX (Mb)

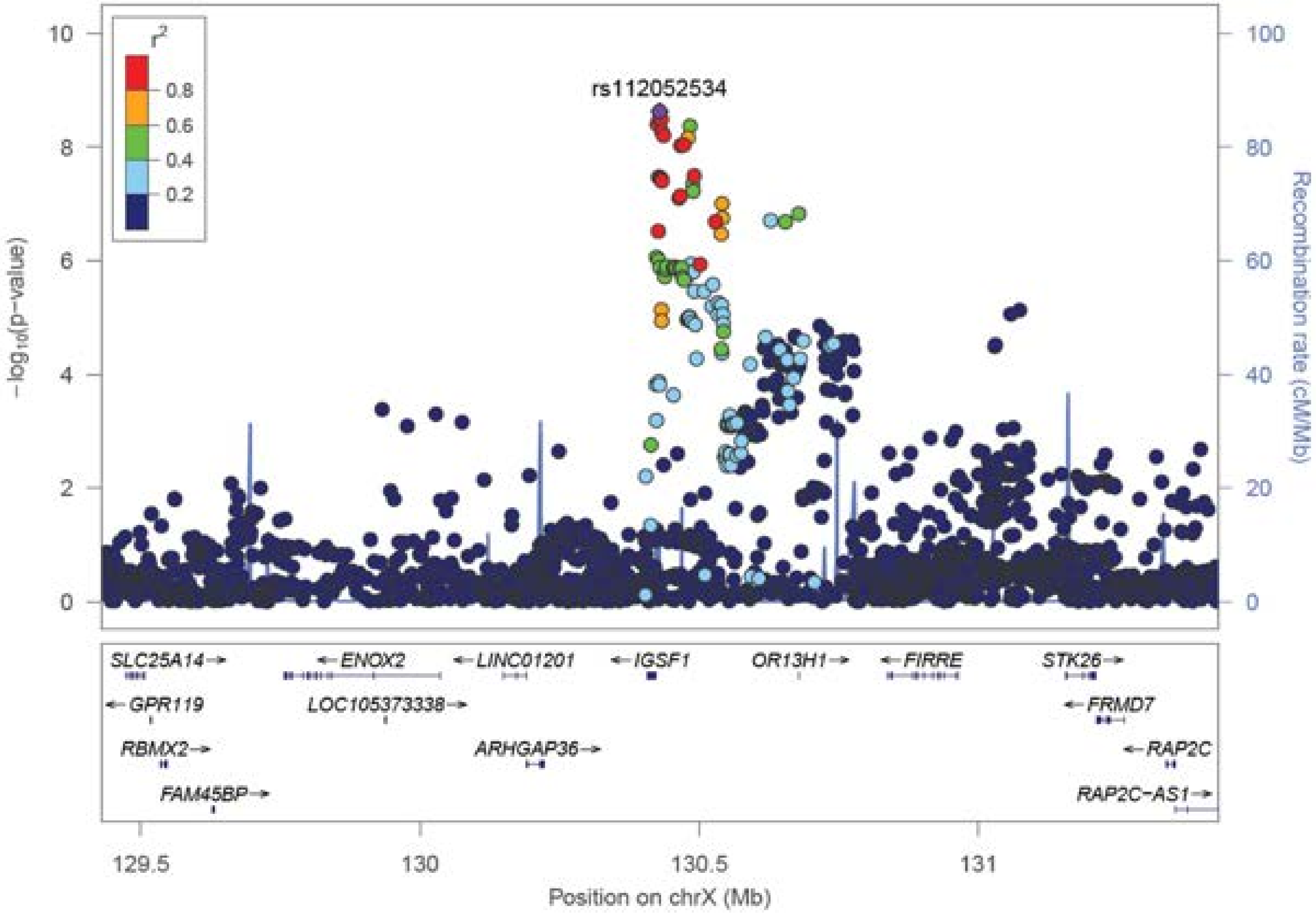

rs112052534
r²
0.8
0.6
0.4
0.2
−log₁₀(p−value)
Recombination rate (cM/Mb)
SLC25A14
ENOX2
LINC01201
IGSF1
OR13H1
FIRRE
STK26
GPR119
LOC105373338
FRMD7
RBMX2
ARHGAP36
RAP2C
FAM45BP
RAP2C−AS1
129.5
130
130.5
131
Position on chrX (Mb)

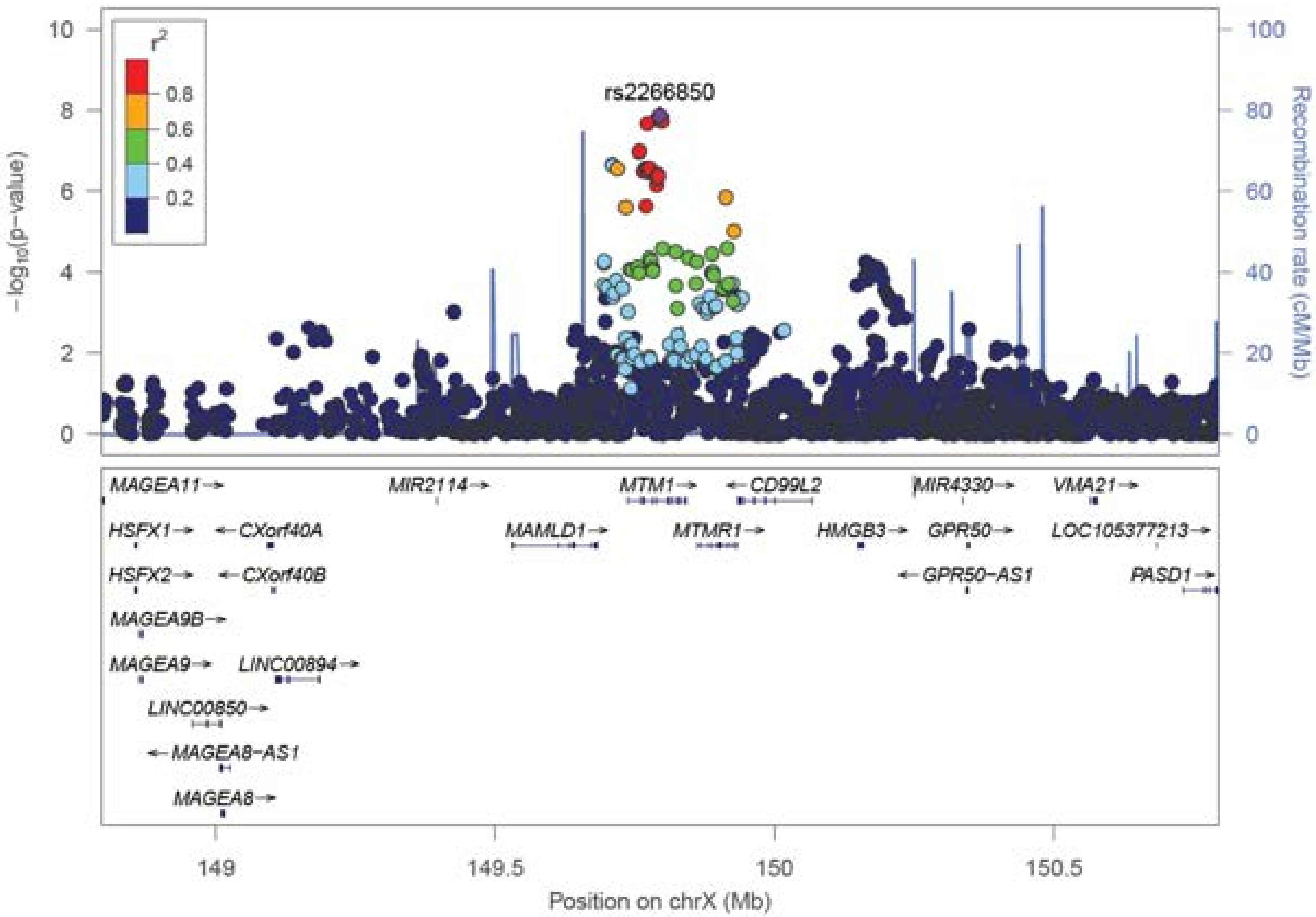

rs2266850
r²
0.8
0.6
0.4
0.2
-log10(p-value)
Recombination rate (cM/Mb)
MAGEA11
MIR2114
MTM1
CD99L2
MIR4330
VMA21
HSFX1
CXorf40A
MAMLD1
MTMR1
HMGB3
GPR50
LOC105377213
HSFX2
CXorf40B
GPR50-AS1
PASD1
MAGEA9B
MAGEA9
LINC00894
LINC00850
MAGEA8-AS1
MAGEA8
Position on chrX (Mb)
