## Supplemental Data 2 for "Discovery of 95 PTSD loci provides insight into genetic architecture and neurobiology of trauma and stress-related disorders"

**Supplementary Data 2.**

Forest plots for each of the 81 index variants of the genome-wide significant loci identified in the GWAS meta-analysis of 137,136 PTSD cases and 1,085,746 controls of European ancestry. Each plot provides a visualization of the standardized effect size estimates for each cohort, represented as black squares that are proportional to sample size weight. Horizontal bars indicate 95% confidence intervals. Sample size weighted meta-analysis results for the data subsets (Freeze 2.5 [F2.5], Electronic Health Record [EHR] based data) and the complete Freeze 3 are represented by blue diamonds.

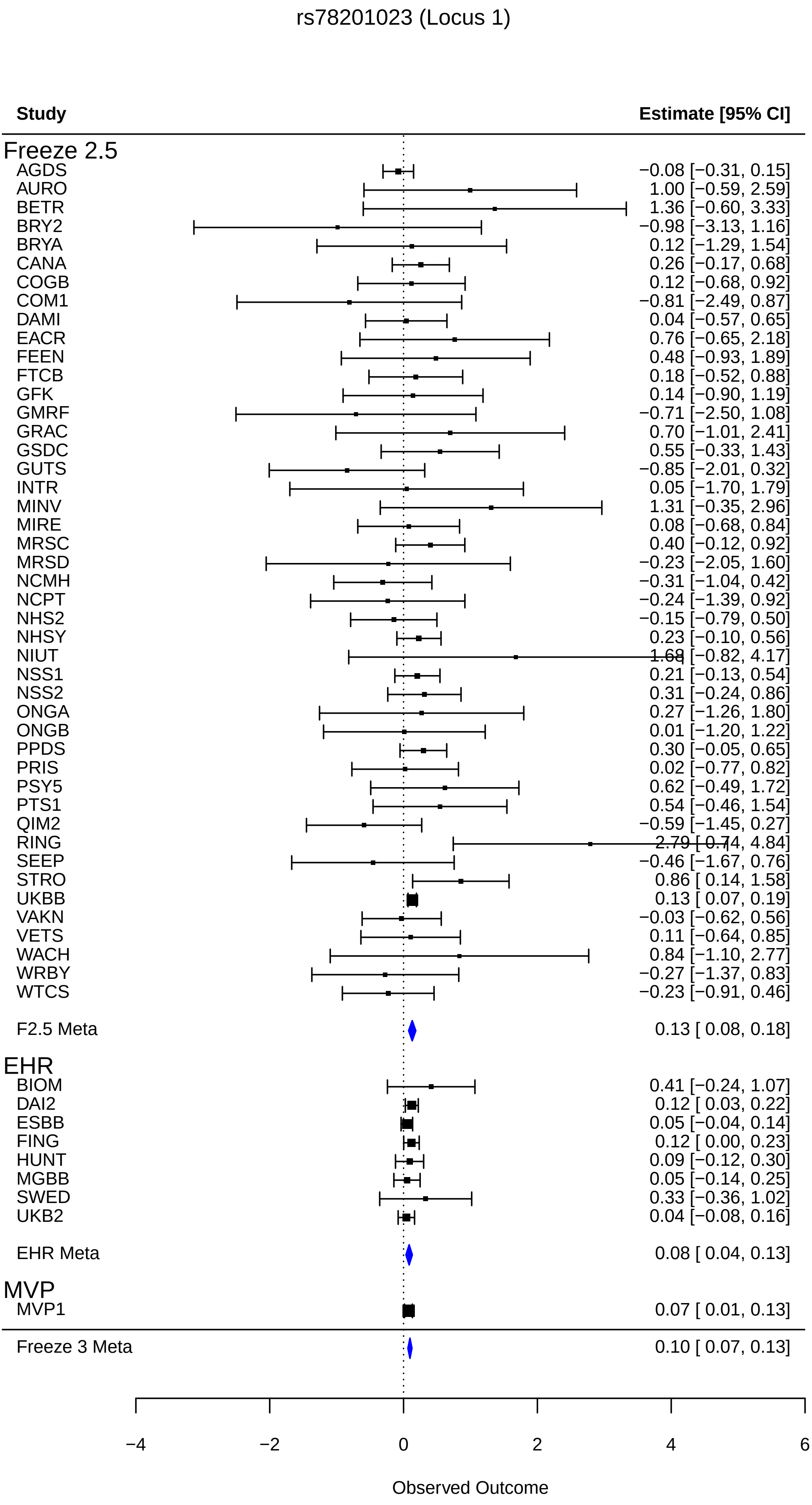
rs78201023 (Locus 1)
Study
Estimate [95% CI]
Freeze 2.5
AGDS −0.08 [−0.31, 0.15]
AURO 1.00 [−0.59, 2.59]
BETR 1.36 [−0.60, 3.33]
BRY2 −0.98 [−3.13, 1.16]
BRYA 0.12 [−1.29, 1.54]
CANA 0.26 [−0.17, 0.68]
COGB 0.12 [−0.68, 0.92]
COM1 −0.81 [−2.49, 0.87]
DAMI 0.04 [−0.57, 0.65]
EACR 0.76 [−0.65, 2.18]
FEEN 0.48 [−0.93, 1.89]
FTCB 0.18 [−0.52, 0.88]
GFK 0.14 [−0.90, 1.19]
GMRF −0.71 [−2.50, 1.08]
GRAC 0.70 [−1.01, 2.41]
GSDC 0.55 [−0.33, 1.43]
GUTS −0.85 [−2.01, 0.32]
INTR 0.05 [−1.70, 1.79]
MINV 1.31 [−0.35, 2.96]
MIRE 0.08 [−0.68, 0.84]
MRSC 0.40 [−0.12, 0.92]
MRSD −0.23 [−2.05, 1.60]
NCMH −0.31 [−1.04, 0.42]
NCPT −0.24 [−1.39, 0.92]
NHS2 −0.15 [−0.79, 0.50]
NHSY 0.23 [−0.10, 0.56]
NIUT 1.68 [−0.82, 4.17]
NSS1 0.21 [−0.13, 0.54]
NSS2 0.31 [−0.24, 0.86]
ONGA 0.27 [−1.26, 1.80]
ONGB 0.01 [−1.20, 1.22]
PPDS 0.30 [−0.05, 0.65]
PRIS 0.02 [−0.77, 0.82]
PSY5 0.62 [−0.49, 1.72]
PTS1 0.54 [−0.46, 1.54]
QIM2 −0.59 [−1.45, 0.27]
RING 2.79 [0.74, 4.84]
SEEP −0.46 [−1.67, 0.76]
STRO 0.86 [ 0.14, 1.58]
UKBB 0.13 [ 0.07, 0.19]
VAKN −0.03 [−0.62, 0.56]
VETS 0.11 [−0.64, 0.85]
WACH 0.84 [−1.10, 2.77]
WRBY −0.27 [−1.37, 0.83]
WTCS −0.23 [−0.91, 0.46]
F2.5 Meta 0.13 [ 0.08, 0.18]
EHR
BIOM 0.41 [−0.24, 1.07]
DAI2 0.12 [ 0.03, 0.22]
ESBB 0.05 [−0.04, 0.14]
FING 0.12 [ 0.00, 0.23]
HUNT 0.09 [−0.12, 0.30]
MGBB 0.05 [−0.14, 0.25]
SWED 0.33 [−0.36, 1.02]
UKB2 0.04 [−0.08, 0.16]
EHR Meta 0.08 [ 0.04, 0.13]
MVP
MVP1 0.07 [ 0.01, 0.13]
Freeze 3 Meta 0.10 [ 0.07, 0.13]
−4
−2
0
2
4
6
Observed Outcome

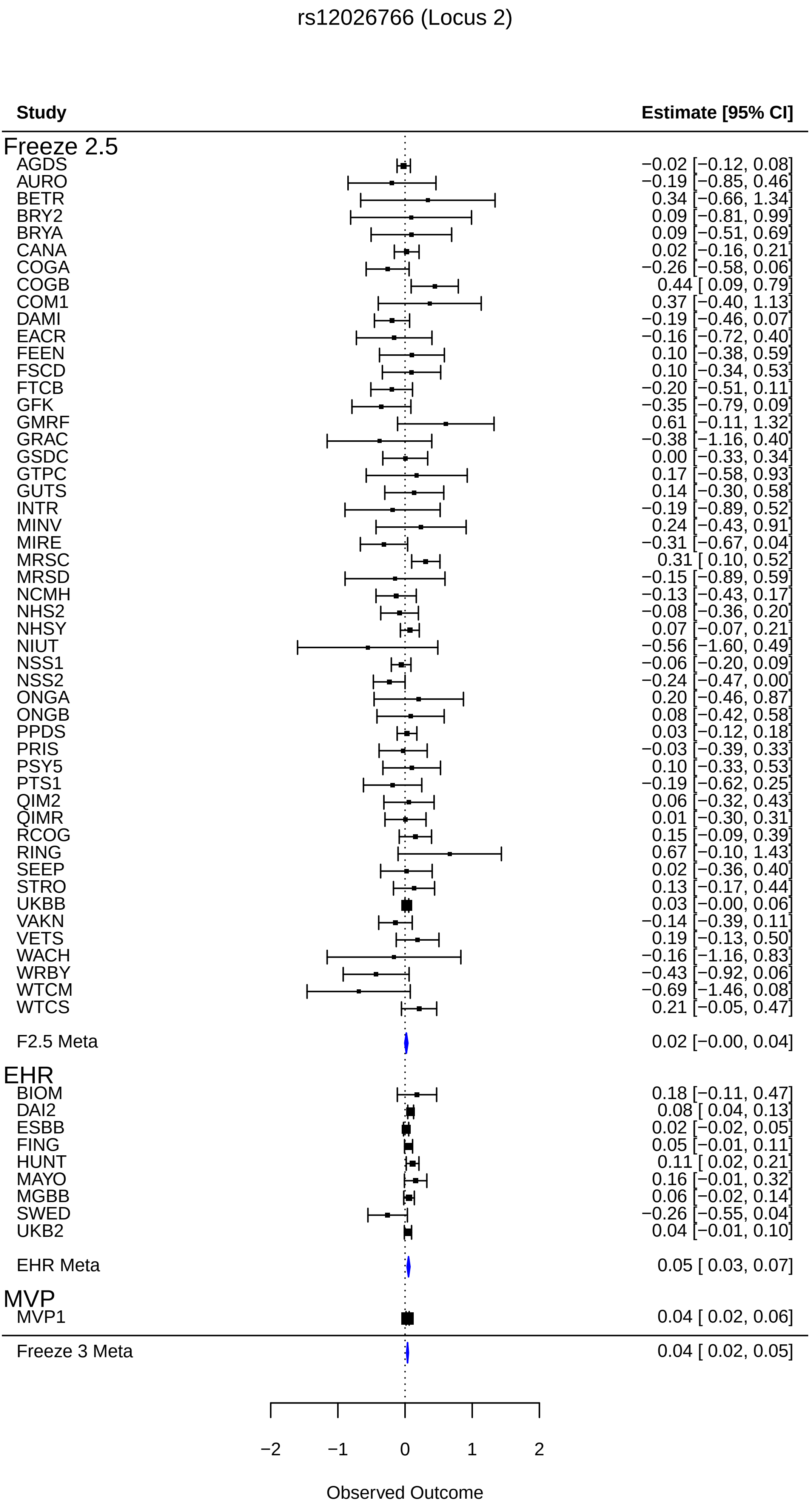
rs12026766 (Locus 2)
Study
Estimate [95% CI]
Freeze 2.5
AGDS −0.02 [−0.12, 0.08]
AURO −0.19 [−0.85, 0.46]
BETR 0.34 [−0.66, 1.34]
BRY2 0.09 [−0.81, 0.99]
BRYA 0.09 [−0.51, 0.69]
CANA 0.02 [−0.16, 0.21]
COGA −0.26 [−0.58, 0.06]
COGB 0.44 [ 0.09, 0.79]
COM1 0.37 [−0.40, 1.13]
DAMI −0.19 [−0.46, 0.07]
EACR −0.16 [−0.72, 0.40]
FEEN 0.10 [−0.38, 0.59]
FSCD 0.10 [−0.34, 0.53]
FTCB −0.20 [−0.51, 0.11]
GFK −0.35 [−0.79, 0.09]
GMRF 0.61 [−0.11, 1.32]
GRAC −0.38 [−1.16, 0.40]
GSDC 0.00 [−0.33, 0.34]
GTPC 0.17 [−0.58, 0.93]
GUTS 0.14 [−0.30, 0.58]
INTR −0.19 [−0.89, 0.52]
MINV 0.24 [−0.43, 0.91]
MIRE −0.31 [−0.67, 0.04]
MRSC 0.31 [ 0.10, 0.52]
MRSD −0.15 [−0.89, 0.59]
NCMH −0.13 [−0.43, 0.17]
NHS2 −0.08 [−0.36, 0.20]
NHSY 0.07 [−0.07, 0.21]
NIUT −0.56 [−1.60, 0.49]
NSS1 −0.06 [−0.20, 0.09]
NSS2 −0.24 [−0.47, 0.00]
ONGA 0.20 [−0.46, 0.87]
ONGB 0.08 [−0.42, 0.58]
PPDS 0.03 [−0.12, 0.18]
PRIS −0.03 [−0.39, 0.33]
PSY5 0.10 [−0.33, 0.53]
PTS1 −0.19 [−0.62, 0.25]
QIM2 0.06 [−0.32, 0.43]
QIMR 0.01 [−0.30, 0.31]
RCOG 0.15 [−0.09, 0.39]
RING 0.67 [−0.10, 1.43]
SEEP 0.02 [−0.36, 0.40]
STRO 0.13 [−0.17, 0.44]
UKBB 0.03 [−0.00, 0.06]
VAKN −0.14 [−0.39, 0.11]
VETS 0.19 [−0.13, 0.50]
WACH −0.16 [−1.16, 0.83]
WRBY −0.43 [−0.92, 0.06]
WTCM −0.69 [−1.46, 0.08]
WTCS 0.21 [−0.05, 0.47]
F2.5 Meta 0.02 [−0.00, 0.04]
EHR
BIOM 0.18 [−0.11, 0.47]
DAI2 0.08 [ 0.04, 0.13]
ESBB 0.02 [−0.02, 0.05]
FING 0.05 [−0.01, 0.11]
HUNT 0.11 [ 0.02, 0.21]
MAYO 0.16 [−0.01, 0.32]
MGBB 0.06 [−0.02, 0.14]
SWED −0.26 [−0.55, 0.04]
UKB2 0.04 [−0.01, 0.10]
EHR Meta 0.05 [ 0.03, 0.07]
MVP
MVP1 0.04 [ 0.02, 0.06]
Freeze 3 Meta 0.04 [ 0.02, 0.05]
−2
−1
0
1
2
Observed Outcome

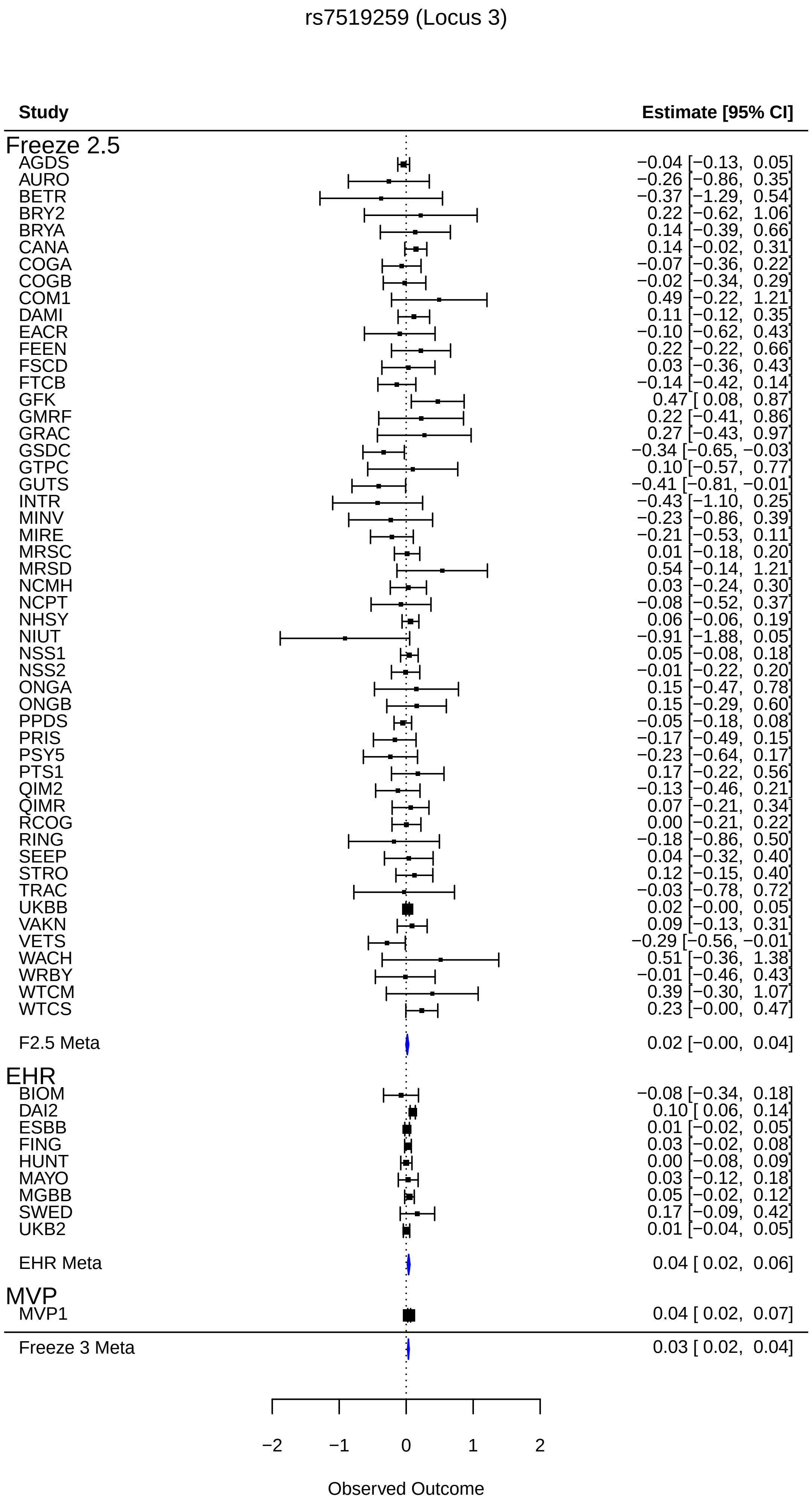
rs7519259 (Locus 3)
Study
Estimate [95% CI]
Freeze 2.5
AGDS −0.04 [−0.13, 0.05]
AURO −0.26 [−0.86, 0.35]
BETR −0.37 [−1.29, 0.54]
BRY2 0.22 [−0.62, 1.06]
BRYA 0.14 [−0.39, 0.66]
CANA 0.14 [−0.02, 0.31]
COGA −0.07 [−0.36, 0.22]
COGB −0.02 [−0.34, 0.29]
COM1 0.49 [−0.22, 1.21]
DAMI 0.11 [−0.12, 0.35]
EACR −0.10 [−0.62, 0.43]
FEEN 0.22 [−0.22, 0.66]
FSCD 0.03 [−0.36, 0.43]
FTCB −0.14 [−0.42, 0.14]
GFK 0.47 [ 0.08, 0.87]
GMRF 0.22 [−0.41, 0.86]
GRAC 0.27 [−0.43, 0.97]
GSDC −0.34 [−0.65, −0.03]
GTPC 0.10 [−0.57, 0.77]
GUTS −0.41 [−0.81, −0.01]
INTR −0.43 [−1.10, 0.25]
MINV −0.23 [−0.86, 0.39]
MIRE −0.21 [−0.53, 0.11]
MRSC 0.01 [−0.18, 0.20]
MRSD 0.54 [−0.14, 1.21]
NCMH 0.03 [−0.24, 0.30]
NCPT −0.08 [−0.52, 0.37]
NHSY 0.06 [−0.06, 0.19]
NIUT −0.91 [−1.88, 0.05]
NSS1 0.05 [−0.08, 0.18]
NSS2 −0.01 [−0.22, 0.20]
ONGA 0.15 [−0.47, 0.78]
ONGB 0.15 [−0.29, 0.60]
PPDS −0.05 [−0.18, 0.08]
PRIS −0.17 [−0.49, 0.15]
PSY5 −0.23 [−0.64, 0.17]
PTS1 0.17 [−0.22, 0.56]
QIM2 −0.13 [−0.46, 0.21]
QIMR 0.07 [−0.21, 0.34]
RCOG 0.00 [−0.21, 0.22]
RING −0.18 [−0.86, 0.50]
SEEP 0.04 [−0.32, 0.40]
STRO 0.12 [−0.15, 0.40]
TRAC −0.03 [−0.78, 0.72]
UKBB 0.02 [−0.00, 0.05]
VAKN 0.09 [−0.13, 0.31]
VETS −0.29 [−0.56, −0.01]
WACH 0.51 [−0.36, 1.38]
WRBY −0.01 [−0.46, 0.43]
WTCM 0.39 [−0.30, 1.07]
WTCS 0.23 [−0.00, 0.47]
F2.5 Meta 0.02 [−0.00, 0.04]
EHR
BIOM −0.08 [−0.34, 0.18]
DAI2 0.10 [ 0.06, 0.14]
ESBB 0.01 [−0.02, 0.05]
FING 0.03 [−0.02, 0.08]
HUNT 0.00 [−0.08, 0.09]
MAYO 0.03 [−0.12, 0.18]
MGBB 0.05 [−0.02, 0.12]
SWED 0.17 [−0.09, 0.42]
UKB2 0.01 [−0.04, 0.05]
EHR Meta 0.04 [ 0.02, 0.06]
MVP
MVP1 0.04 [ 0.02, 0.07]
Freeze 3 Meta 0.03 [ 0.02, 0.04]
−2
−1
0
1
2
Observed Outcome

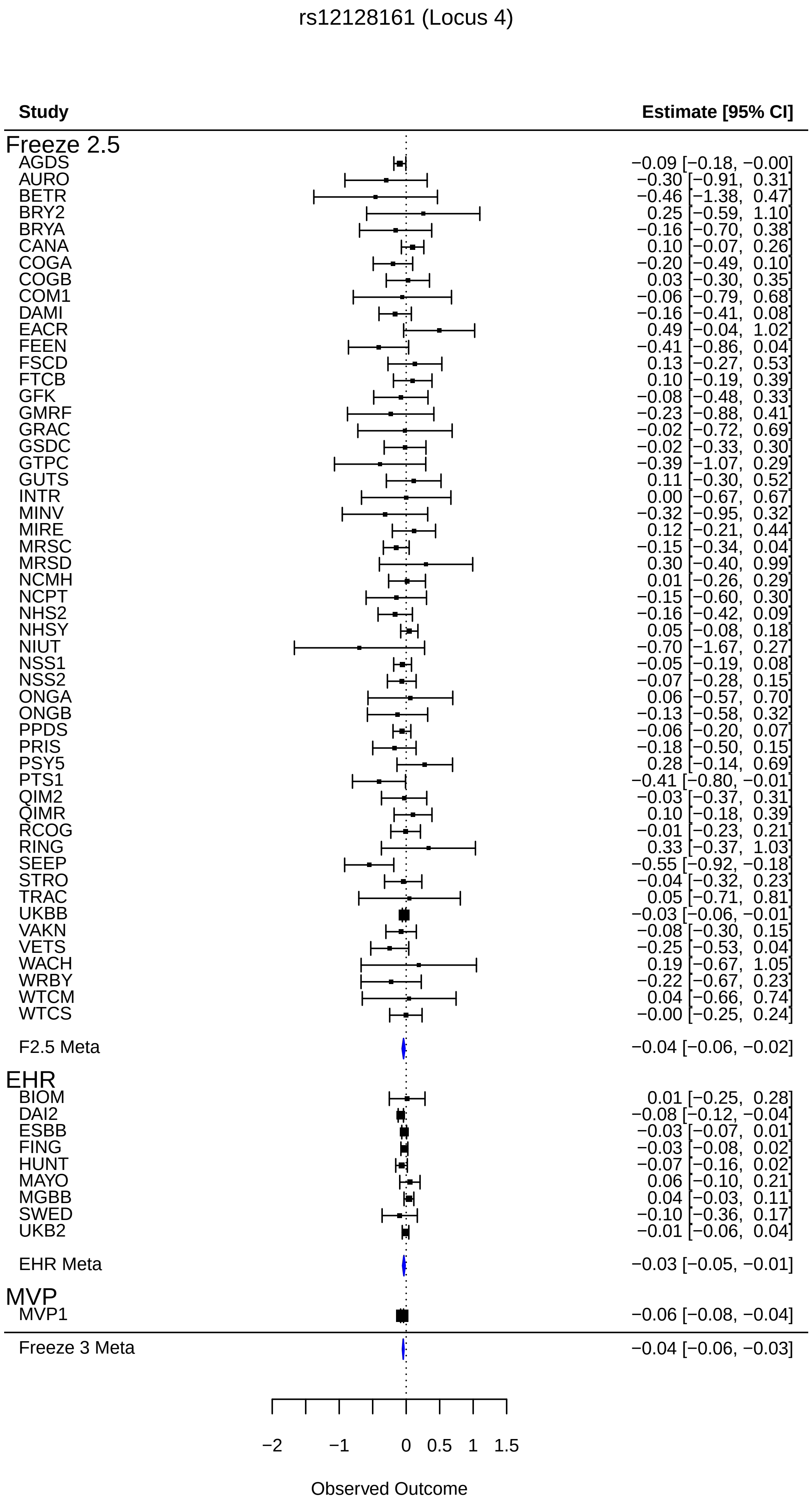
rs12128161 (Locus 4)
Study
Estimate [95% CI]
Freeze 2.5
AGDS −0.09 [−0.18, −0.00]
AURO −0.30 [−0.91, 0.31]
BETR −0.46 [−1.38, 0.47]
BRY2 0.25 [−0.59, 1.10]
BRYA −0.16 [−0.70, 0.38]
CANA 0.10 [−0.07, 0.26]
COGA −0.20 [−0.49, 0.10]
COGB 0.03 [−0.30, 0.35]
COM1 −0.06 [−0.79, 0.68]
DAMI −0.16 [−0.41, 0.08]
EACR 0.49 [−0.04, 1.02]
FEEN −0.41 [−0.86, 0.04]
FSCD 0.13 [−0.27, 0.53]
FTCB 0.10 [−0.19, 0.39]
GFK −0.08 [−0.48, 0.33]
GMRF −0.23 [−0.88, 0.41]
GRAC −0.02 [−0.72, 0.69]
GSDC −0.02 [−0.33, 0.30]
GTPC −0.39 [−1.07, 0.29]
GUTS 0.11 [−0.30, 0.52]
INTR 0.00 [−0.67, 0.67]
MINV −0.32 [−0.95, 0.32]
MIRE 0.12 [−0.21, 0.44]
MRSC −0.15 [−0.34, 0.04]
MRSD 0.30 [−0.40, 0.99]
NCMH 0.01 [−0.26, 0.29]
NCPT −0.15 [−0.60, 0.30]
NHS2 −0.16 [−0.42, 0.09]
NHSY 0.05 [−0.08, 0.18]
NIUT −0.70 [−1.67, 0.27]
NSS1 −0.05 [−0.19, 0.08]
NSS2 −0.07 [−0.28, 0.15]
ONGA 0.06 [−0.57, 0.70]
ONGB −0.13 [−0.58, 0.32]
PPDS −0.06 [−0.20, 0.07]
PRIS −0.18 [−0.50, 0.15]
PSY5 0.28 [−0.14, 0.69]
PTS1 −0.41 [−0.80, −0.01]
QIM2 −0.03 [−0.37, 0.31]
QIMR 0.10 [−0.18, 0.39]
RCOG −0.01 [−0.23, 0.21]
RING 0.33 [−0.37, 1.03]
SEEP −0.55 [−0.92, −0.18]
STRO −0.04 [−0.32, 0.23]
TRAC 0.05 [−0.71, 0.81]
UKBB −0.03 [−0.06, −0.01]
VAKN −0.08 [−0.30, 0.15]
VETS −0.25 [−0.53, 0.04]
WACH 0.19 [−0.67, 1.05]
WRBY −0.22 [−0.67, 0.23]
WTCM 0.04 [−0.66, 0.74]
WTCS −0.00 [−0.25, 0.24]
F2.5 Meta −0.04 [−0.06, −0.02]
EHR
BIOM 0.01 [−0.25, 0.28]
DAI2 −0.08 [−0.12, −0.04]
ESBB −0.03 [−0.07, 0.01]
FING −0.03 [−0.08, 0.02]
HUNT −0.07 [−0.16, 0.02]
MAYO 0.06 [−0.10, 0.21]
MGBB 0.04 [−0.03, 0.11]
SWED −0.10 [−0.36, 0.17]
UKB2 −0.01 [−0.06, 0.04]
EHR Meta −0.03 [−0.05, −0.01]
MVP
MVP1 −0.06 [−0.08, −0.04]
Freeze 3 Meta −0.04 [−0.06, −0.03]
−2
−1
0
0.5
1
1.5
Observed Outcome

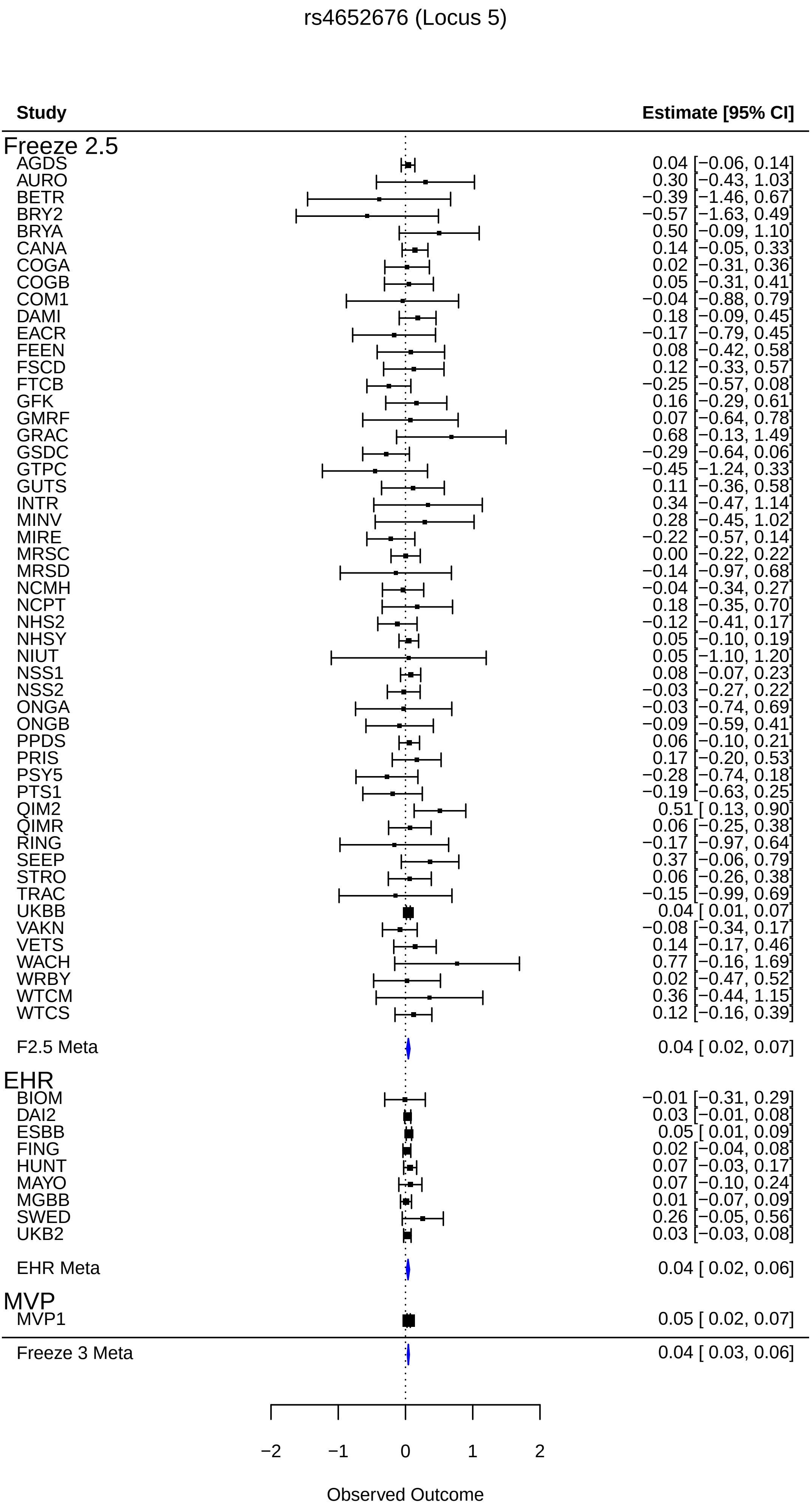
rs4652676 (Locus 5)
Study
Estimate [95% CI]
Freeze 2.5
AGDS 0.04 [−0.06, 0.14]
AURO 0.30 [−0.43, 1.03]
BETR −0.39 [−1.46, 0.67]
BRY2 −0.57 [−1.63, 0.49]
BRYA 0.50 [−0.09, 1.10]
CANA 0.14 [−0.05, 0.33]
COGA 0.02 [−0.31, 0.36]
COGB 0.05 [−0.31, 0.41]
COM1 −0.04 [−0.88, 0.79]
DAMI 0.18 [−0.09, 0.45]
EACR −0.17 [−0.79, 0.45]
FEEN 0.08 [−0.42, 0.58]
FSCD 0.12 [−0.33, 0.57]
FTCB −0.25 [−0.57, 0.08]
GFK 0.16 [−0.29, 0.61]
GMRF 0.07 [−0.64, 0.78]
GRAC 0.68 [−0.13, 1.49]
GSDC −0.29 [−0.64, 0.06]
GTPC −0.45 [−1.24, 0.33]
GUTS 0.11 [−0.36, 0.58]
INTR 0.34 [−0.47, 1.14]
MINV 0.28 [−0.45, 1.02]
MIRE −0.22 [−0.57, 0.14]
MRSC 0.00 [−0.22, 0.22]
MRSD −0.14 [−0.97, 0.68]
NCMH −0.04 [−0.34, 0.27]
NCPT 0.18 [−0.35, 0.70]
NHS2 −0.12 [−0.41, 0.17]
NHSY 0.05 [−0.10, 0.19]
NIUT 0.05 [−1.10, 1.20]
NSS1 0.08 [−0.07, 0.23]
NSS2 −0.03 [−0.27, 0.22]
ONGA −0.03 [−0.74, 0.69]
ONGB −0.09 [−0.59, 0.41]
PPDS 0.06 [−0.10, 0.21]
PRIS 0.17 [−0.20, 0.53]
PSY5 −0.28 [−0.74, 0.18]
PTS1 −0.19 [−0.63, 0.25]
QIM2 0.51 [ 0.13, 0.90]
QIMR 0.06 [−0.25, 0.38]
RING −0.17 [−0.97, 0.64]
SEEP 0.37 [−0.06, 0.79]
STRO 0.06 [−0.26, 0.38]
TRAC −0.15 [−0.99, 0.69]
UKBB 0.04 [ 0.01, 0.07]
VAKN −0.08 [−0.34, 0.17]
VETS 0.14 [−0.17, 0.46]
WACH 0.77 [−0.16, 1.69]
WRBY 0.02 [−0.47, 0.52]
WTCM 0.36 [−0.44, 1.15]
WTCS 0.12 [−0.16, 0.39]
F2.5 Meta 0.04 [ 0.02, 0.07]
EHR
BIOM −0.01 [−0.31, 0.29]
DAI2 0.03 [−0.01, 0.08]
ESBB 0.05 [ 0.01, 0.09]
FING 0.02 [−0.04, 0.08]
HUNT 0.07 [−0.03, 0.17]
MAYO 0.07 [−0.10, 0.24]
MGBB 0.01 [−0.07, 0.09]
SWED 0.26 [−0.05, 0.56]
UKB2 0.03 [−0.03, 0.08]
EHR Meta 0.04 [ 0.02, 0.06]
MVP
MVP1 0.05 [ 0.02, 0.07]
Freeze 3 Meta 0.04 [ 0.03, 0.06]
−2
−1
0
1
2
Observed Outcome

rs9651063 (Locus 6)

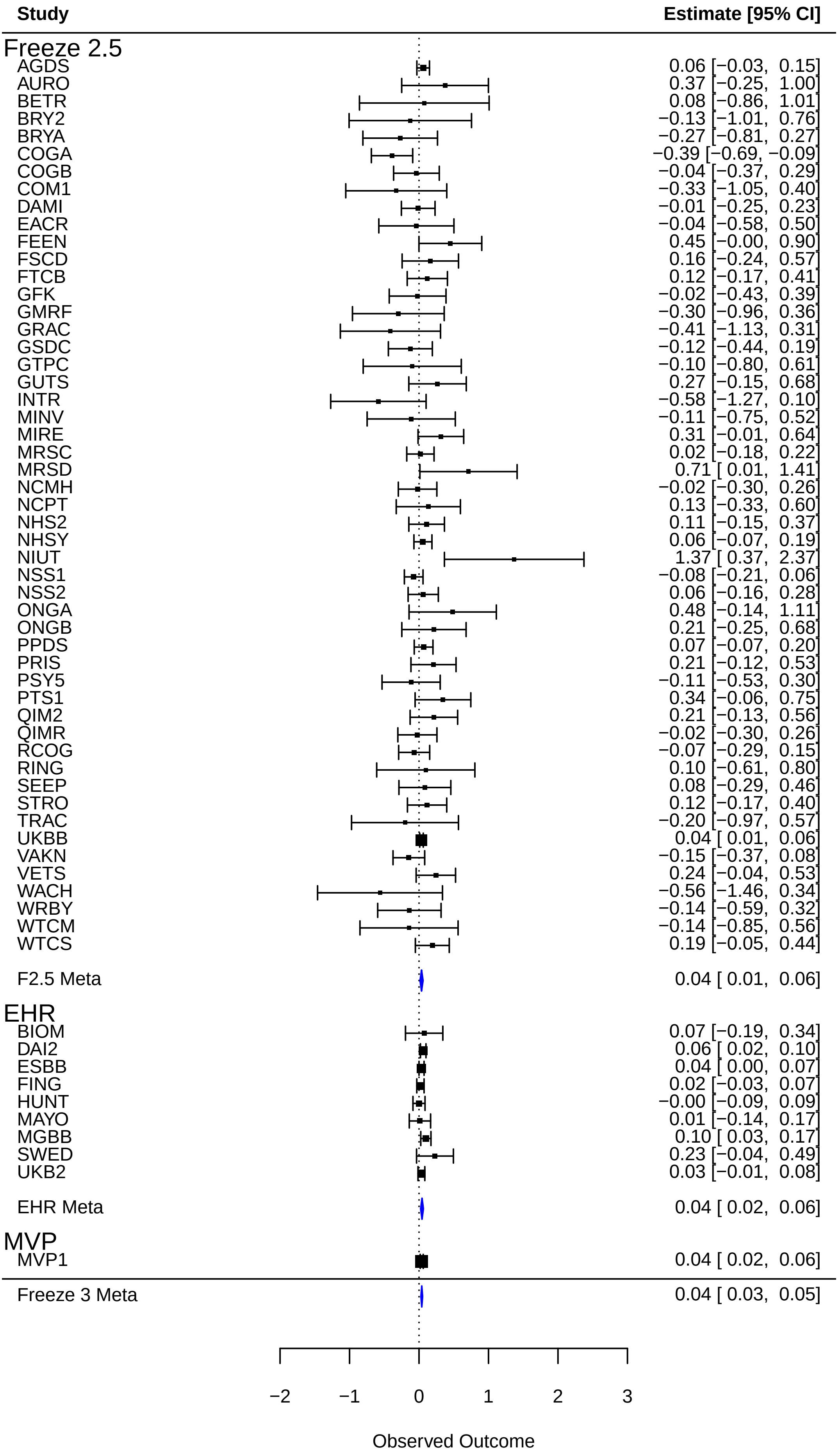

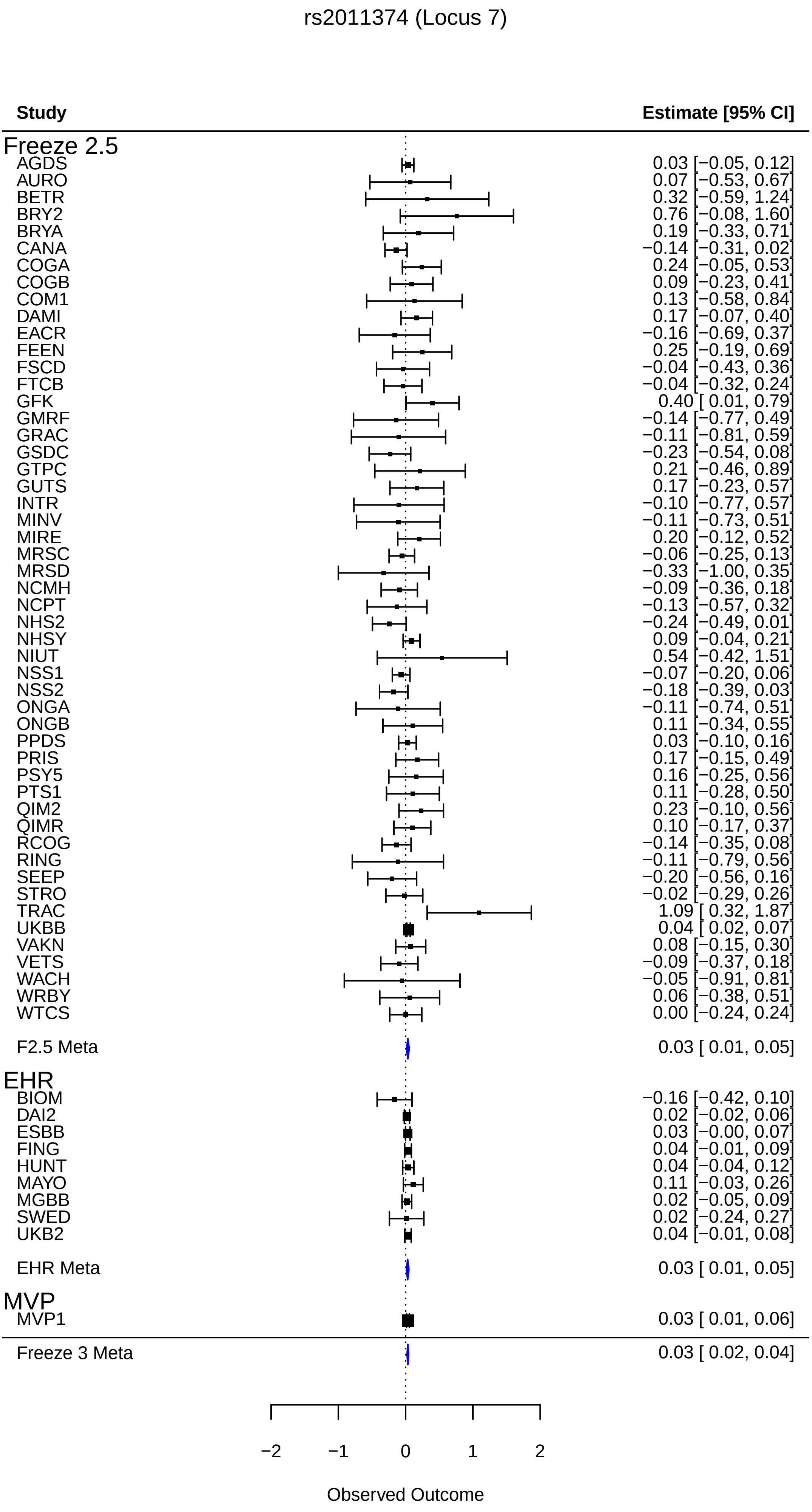
rs2011374 (Locus 7)
Study
Estimate [95% CI]
Freeze 2.5
AGDS 0.03 [−0.05, 0.12]
AURO 0.07 [−0.53, 0.67]
BETR 0.32 [−0.59, 1.24]
BRY2 0.76 [−0.08, 1.60]
BRYA 0.19 [−0.33, 0.71]
CANA −0.14 [−0.31, 0.02]
COGA 0.24 [−0.05, 0.53]
COGB 0.09 [−0.23, 0.41]
COM1 0.13 [−0.58, 0.84]
DAMI 0.17 [−0.07, 0.40]
EACR −0.16 [−0.69, 0.37]
FEEN 0.25 [−0.19, 0.69]
FSCD −0.04 [−0.43, 0.36]
FTCB −0.04 [−0.32, 0.24]
GFK 0.40 [ 0.01, 0.79]
GMRF −0.14 [−0.77, 0.49]
GRAC −0.11 [−0.81, 0.59]
GSDC −0.23 [−0.54, 0.08]
GTPC 0.21 [−0.46, 0.89]
GUTS 0.17 [−0.23, 0.57]
INTR −0.10 [−0.77, 0.57]
MINV −0.11 [−0.73, 0.51]
MIRE 0.20 [−0.12, 0.52]
MRSC −0.06 [−0.25, 0.13]
MRSD −0.33 [−1.00, 0.35]
NCMH −0.09 [−0.36, 0.18]
NCPT −0.13 [−0.57, 0.32]
NHS2 −0.24 [−0.49, 0.01]
NHSY 0.09 [−0.04, 0.21]
NIUT 0.54 [−0.42, 1.51]
NSS1 −0.07 [−0.20, 0.06]
NSS2 −0.18 [−0.39, 0.03]
ONGA −0.11 [−0.74, 0.51]
ONGB 0.11 [−0.34, 0.55]
PPDS 0.03 [−0.10, 0.16]
PRIS 0.17 [−0.15, 0.49]
PSY5 0.16 [−0.25, 0.56]
PTS1 0.11 [−0.28, 0.50]
QIM2 0.23 [−0.10, 0.56]
QIMR 0.10 [−0.17, 0.37]
RCOG −0.14 [−0.35, 0.08]
RING −0.11 [−0.79, 0.56]
SEEP −0.20 [−0.56, 0.16]
STRO −0.02 [−0.29, 0.26]
TRAC 1.09 [ 0.32, 1.87]
UKBB 0.04 [ 0.02, 0.07]
VAKN 0.08 [−0.15, 0.30]
VETS −0.09 [−0.37, 0.18]
WACH −0.05 [−0.91, 0.81]
WRBY 0.06 [−0.38, 0.51]
WTCS 0.00 [−0.24, 0.24]
F2.5 Meta 0.03 [ 0.01, 0.05]
EHR
BIOM −0.16 [−0.42, 0.10]
DAI2 0.02 [−0.02, 0.06]
ESBB 0.03 [−0.00, 0.07]
FING 0.04 [−0.01, 0.09]
HUNT 0.04 [−0.04, 0.12]
MAYO 0.11 [−0.03, 0.26]
MGBB 0.02 [−0.05, 0.09]
SWED 0.02 [−0.24, 0.27]
UKB2 0.04 [−0.01, 0.08]
EHR Meta 0.03 [ 0.01, 0.05]
MVP
MVP1 0.03 [ 0.01, 0.06]
Freeze 3 Meta 0.03 [ 0.02, 0.04]
−2
−1
0
1
2
Observed Outcome

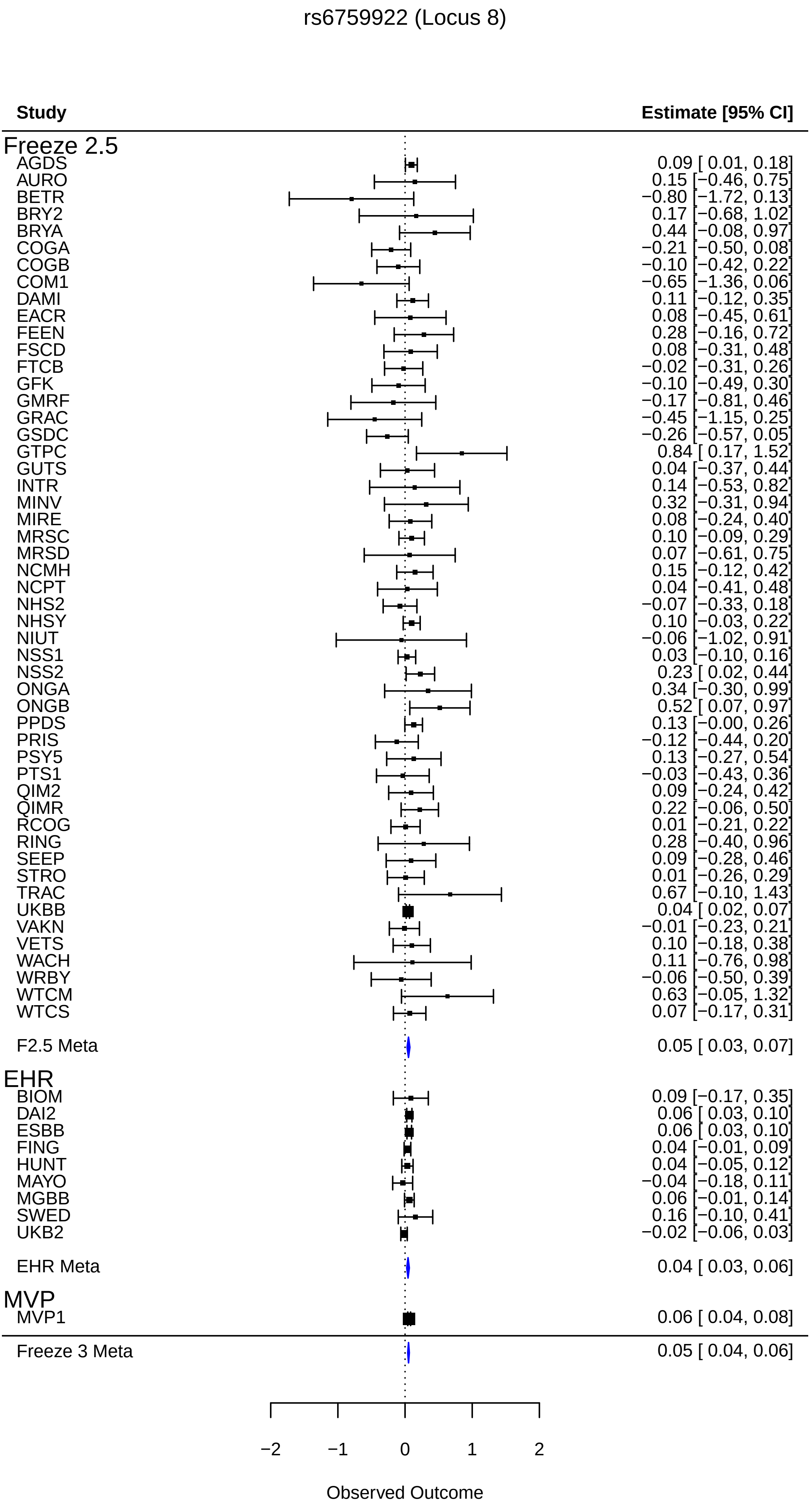
rs6759922 (Locus 8)
Study
Estimate [95% CI]
Freeze 2.5
AGDS 0.09 [ 0.01, 0.18]
AURO 0.15 [−0.46, 0.75]
BETR −0.80 [−1.72, 0.13]
BRY2 0.17 [−0.68, 1.02]
BRYA 0.44 [−0.08, 0.97]
COGA −0.21 [−0.50, 0.08]
COGB −0.10 [−0.42, 0.22]
COM1 −0.65 [−1.36, 0.06]
DAMI 0.11 [−0.12, 0.35]
EACR 0.08 [−0.45, 0.61]
FEEN 0.28 [−0.16, 0.72]
FSCD 0.08 [−0.31, 0.48]
FTCB −0.02 [−0.31, 0.26]
GFK −0.10 [−0.49, 0.30]
GMRF −0.17 [−0.81, 0.46]
GRAC −0.45 [−1.15, 0.25]
GSDC −0.26 [−0.57, 0.05]
GTPC 0.84 [ 0.17, 1.52]
GUTS 0.04 [−0.37, 0.44]
INTR 0.14 [−0.53, 0.82]
MINV 0.32 [−0.31, 0.94]
MIRE 0.08 [−0.24, 0.40]
MRSC 0.10 [−0.09, 0.29]
MRSD 0.07 [−0.61, 0.75]
NCMH 0.15 [−0.12, 0.42]
NCPT 0.04 [−0.41, 0.48]
NHS2 −0.07 [−0.33, 0.18]
NHSY 0.10 [−0.03, 0.22]
NIUT −0.06 [−1.02, 0.91]
NSS1 0.03 [−0.10, 0.16]
NSS2 0.23 [ 0.02, 0.44]
ONGA 0.34 [−0.30, 0.99]
ONGB 0.52 [ 0.07, 0.97]
PPDS 0.13 [−0.00, 0.26]
PRIS −0.12 [−0.44, 0.20]
PSY5 0.13 [−0.27, 0.54]
PTS1 −0.03 [−0.43, 0.36]
QIM2 0.09 [−0.24, 0.42]
QIMR 0.22 [−0.06, 0.50]
RCOG 0.01 [−0.21, 0.22]
RING 0.28 [−0.40, 0.96]
SEEP 0.09 [−0.28, 0.46]
STRO 0.01 [−0.26, 0.29]
TRAC 0.67 [−0.10, 1.43]
UKBB 0.04 [ 0.02, 0.07]
VAKN −0.01 [−0.23, 0.21]
VETS 0.10 [−0.18, 0.38]
WACH 0.11 [−0.76, 0.98]
WRBY −0.06 [−0.50, 0.39]
WTCM 0.63 [−0.05, 1.32]
WTCS 0.07 [−0.17, 0.31]
F2.5 Meta 0.05 [ 0.03, 0.07]
EHR
BIOM 0.09 [−0.17, 0.35]
DAI2 0.06 [ 0.03, 0.10]
ESBB 0.06 [ 0.03, 0.10]
FING 0.04 [−0.01, 0.09]
HUNT 0.04 [−0.05, 0.12]
MAYO −0.04 [−0.18, 0.11]
MGBB 0.06 [−0.01, 0.14]
SWED 0.16 [−0.10, 0.41]
UKB2 −0.02 [−0.06, 0.03]
EHR Meta 0.04 [ 0.03, 0.06]
MVP
MVP1 0.06 [ 0.04, 0.08]
Freeze 3 Meta 0.05 [ 0.04, 0.06]
−2
−1
0
1
2
Observed Outcome

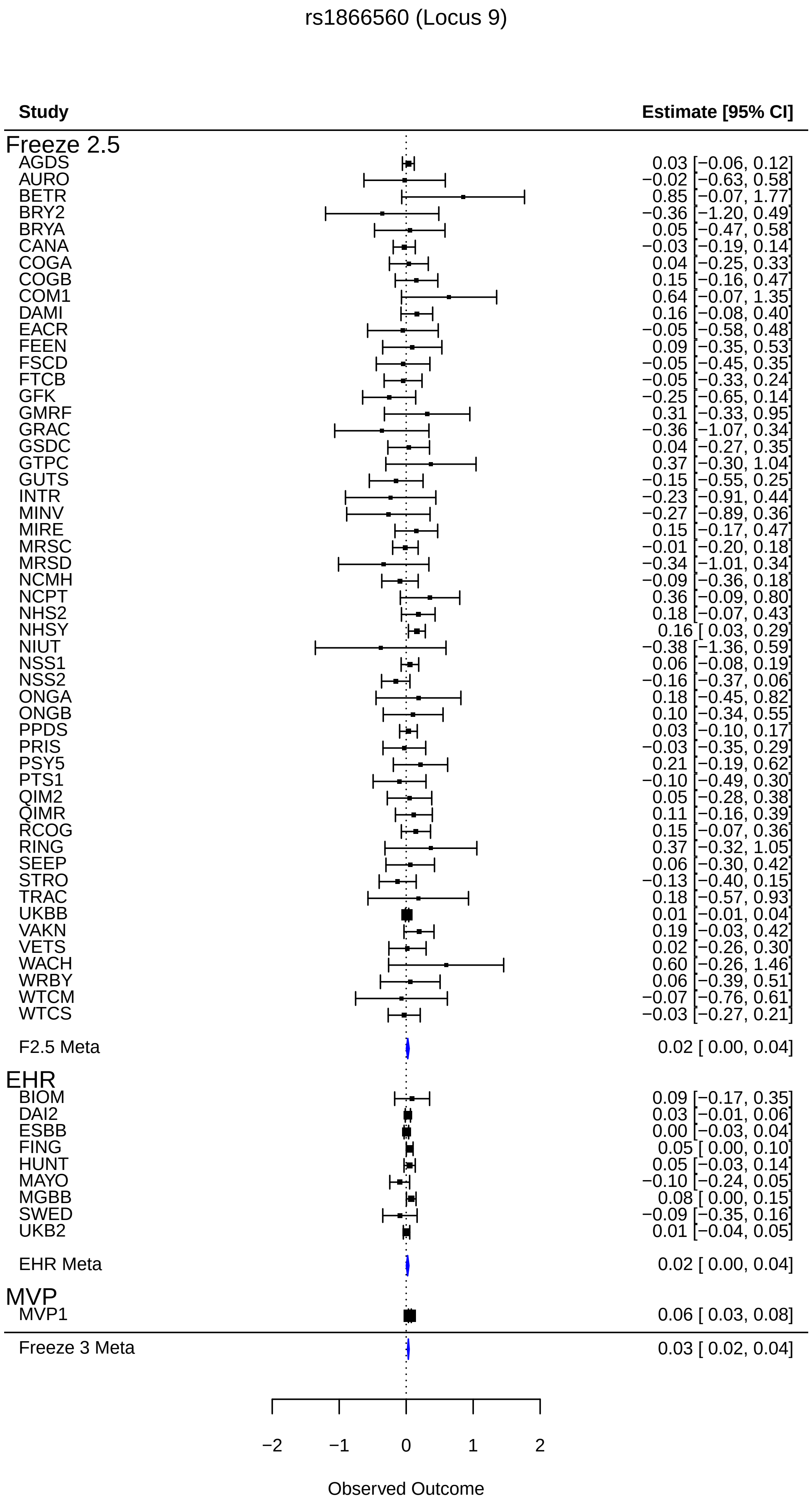
rs1866560 (Locus 9)
Study
Estimate [95% CI]
Freeze 2.5
AGDS 0.03 [−0.06, 0.12]
AURO −0.02 [−0.63, 0.58]
BETR 0.85 [−0.07, 1.77]
BRY2 −0.36 [−1.20, 0.49]
BRYA 0.05 [−0.47, 0.58]
CANA −0.03 [−0.19, 0.14]
COGA 0.04 [−0.25, 0.33]
COGB 0.15 [−0.16, 0.47]
COM1 0.64 [−0.07, 1.35]
DAMI 0.16 [−0.08, 0.40]
EACR −0.05 [−0.58, 0.48]
FEEN 0.09 [−0.35, 0.53]
FSCD −0.05 [−0.45, 0.35]
FTCB −0.05 [−0.33, 0.24]
GFK −0.25 [−0.65, 0.14]
GMRF 0.31 [−0.33, 0.95]
GRAC −0.36 [−1.07, 0.34]
GSDC 0.04 [−0.27, 0.35]
GTPC 0.37 [−0.30, 1.04]
GUTS −0.15 [−0.55, 0.25]
INTR −0.23 [−0.91, 0.44]
MINV −0.27 [−0.89, 0.36]
MIRE 0.15 [−0.17, 0.47]
MRSC −0.01 [−0.20, 0.18]
MRSD −0.34 [−1.01, 0.34]
NCMH −0.09 [−0.36, 0.18]
NCPT 0.36 [−0.09, 0.80]
NHS2 0.18 [−0.07, 0.43]
NHSY 0.16 [ 0.03, 0.29]
NIUT −0.38 [−1.36, 0.59]
NSS1 0.06 [−0.08, 0.19]
NSS2 −0.16 [−0.37, 0.06]
ONGA 0.18 [−0.45, 0.82]
ONGB 0.10 [−0.34, 0.55]
PPDS 0.03 [−0.10, 0.17]
PRIS −0.03 [−0.35, 0.29]
PSY5 0.21 [−0.19, 0.62]
PTS1 −0.10 [−0.49, 0.30]
QIM2 0.05 [−0.28, 0.38]
QIMR 0.11 [−0.16, 0.39]
RCOG 0.15 [−0.07, 0.36]
RING 0.37 [−0.32, 1.05]
SEEP 0.06 [−0.30, 0.42]
STRO −0.13 [−0.40, 0.15]
TRAC 0.18 [−0.57, 0.93]
UKBB 0.01 [−0.01, 0.04]
VAKN 0.19 [−0.03, 0.42]
VETS 0.02 [−0.26, 0.30]
WACH 0.60 [−0.26, 1.46]
WRBY 0.06 [−0.39, 0.51]
WTCM −0.07 [−0.76, 0.61]
WTCS −0.03 [−0.27, 0.21]
F2.5 Meta 0.02 [ 0.00, 0.04]
EHR
BIOM 0.09 [−0.17, 0.35]
DAI2 0.03 [−0.01, 0.06]
ESBB 0.00 [−0.03, 0.04]
FING 0.05 [ 0.00, 0.10]
HUNT 0.05 [−0.03, 0.14]
MAYO −0.10 [−0.24, 0.05]
MGBB 0.08 [ 0.00, 0.15]
SWED −0.09 [−0.35, 0.16]
UKB2 0.01 [−0.04, 0.05]
EHR Meta 0.02 [ 0.00, 0.04]
MVP
MVP1 0.06 [ 0.03, 0.08]
Freeze 3 Meta 0.03 [ 0.02, 0.04]
−2
−1
0
1
2
Observed Outcome

rs10496632 (Locus 10)

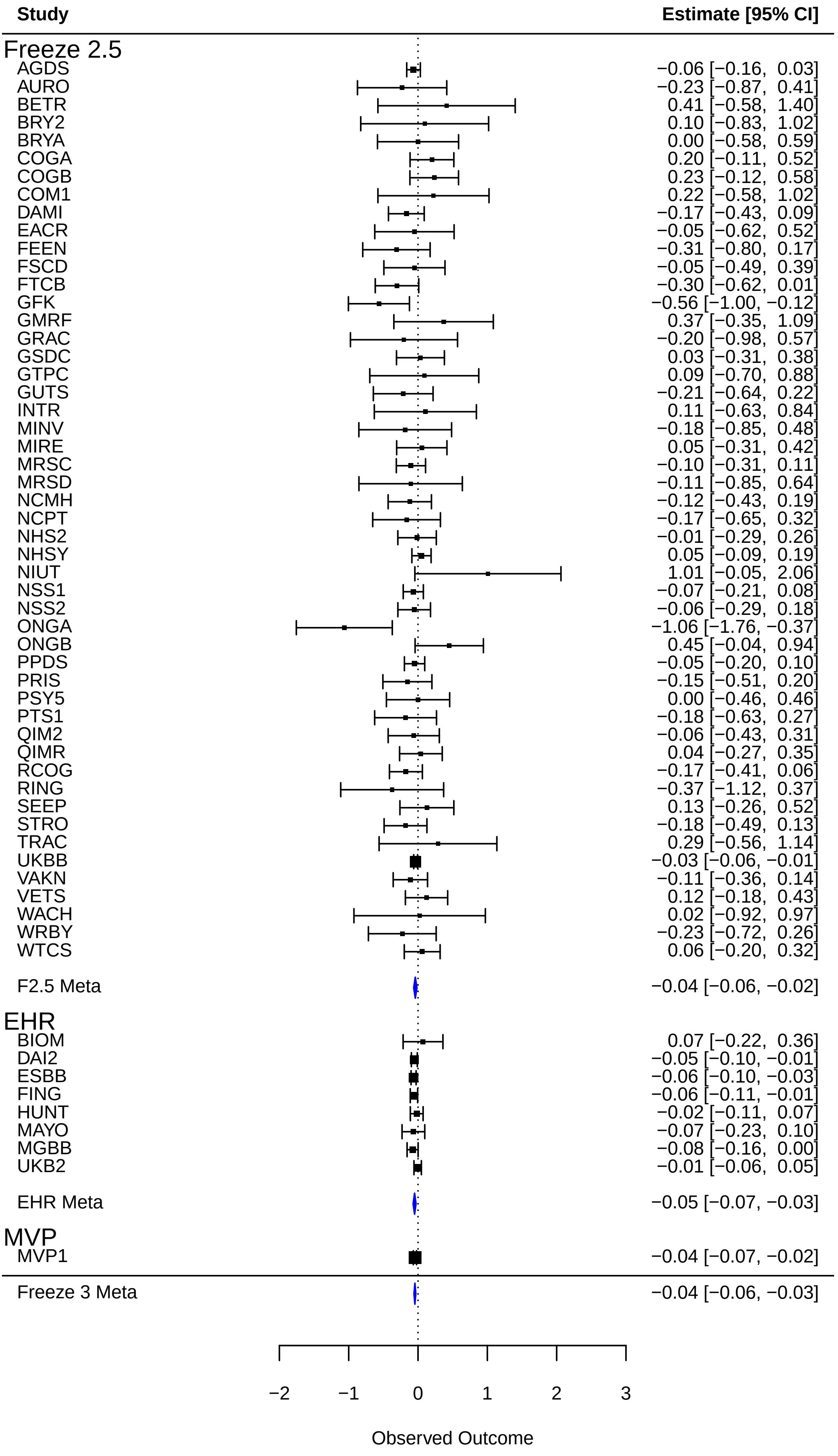

rs6430728 (Locus11)

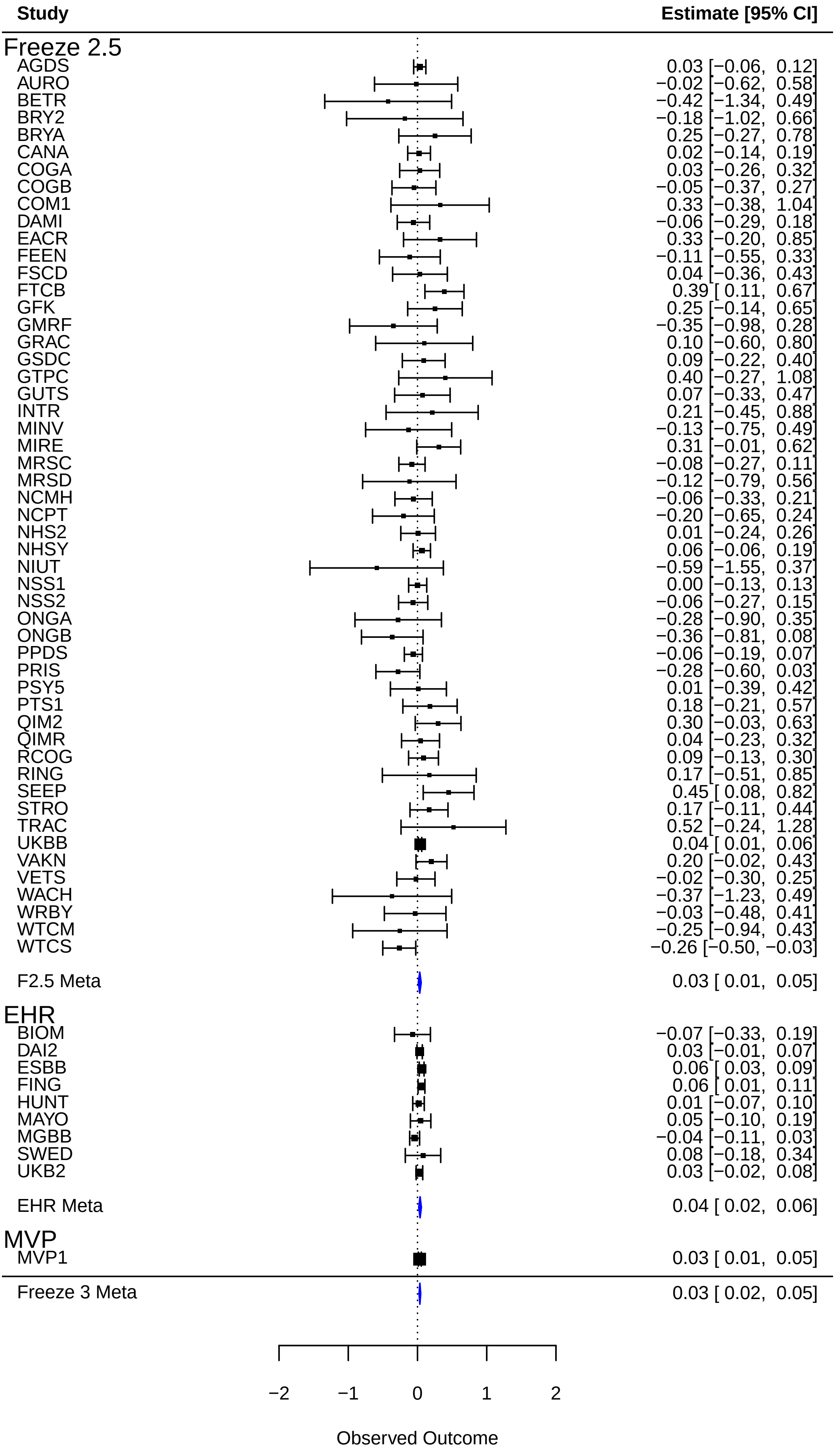

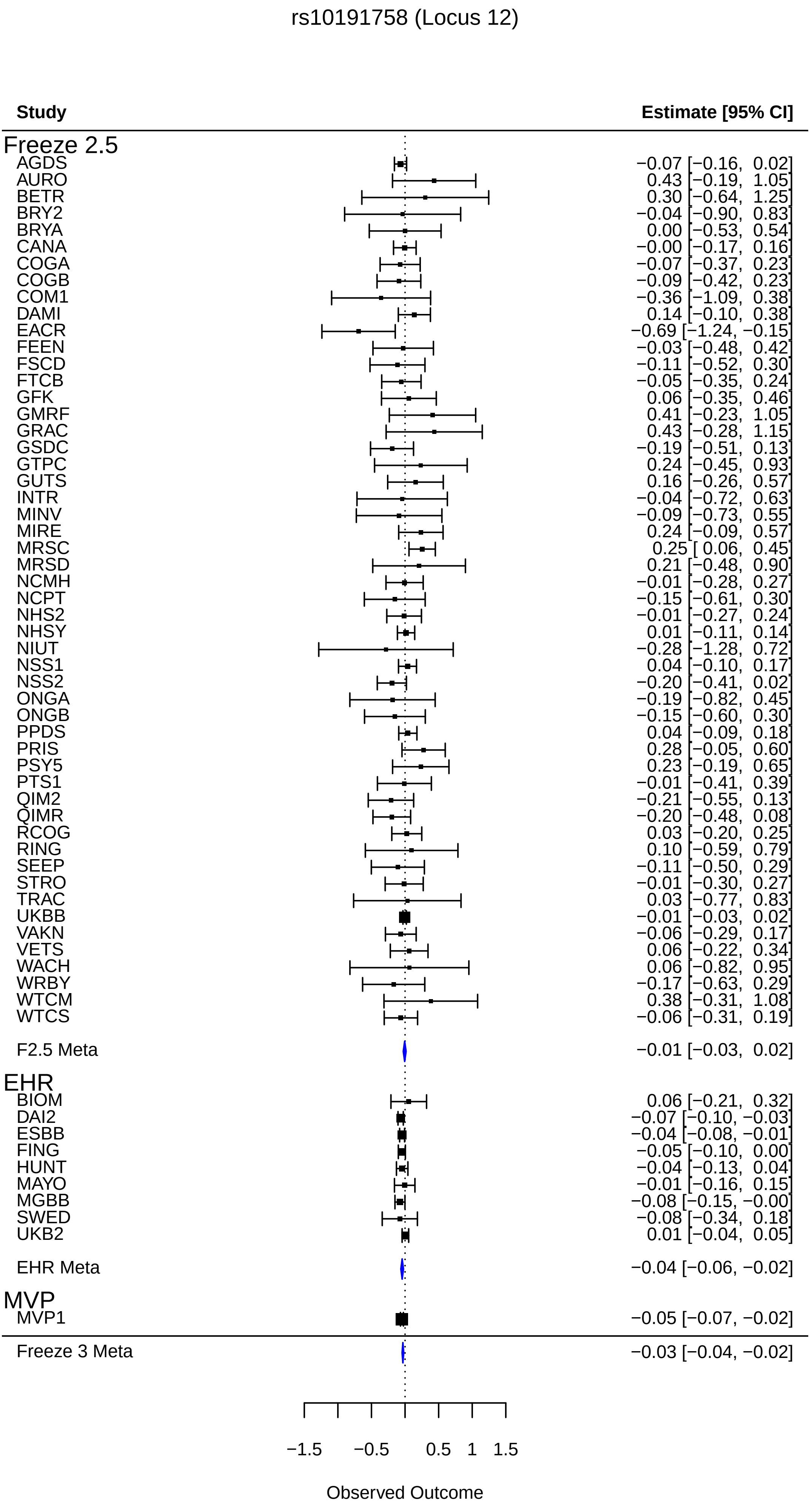
rs10191758 (Locus 12)
Study
Estimate [95% CI]
Freeze 2.5
AGDS −0.07 [−0.16, 0.02]
AURO 0.43 [−0.19, 1.05]
BETR 0.30 [−0.64, 1.25]
BRY2 −0.04 [−0.90, 0.83]
BRYA 0.00 [−0.53, 0.54]
CANA −0.00 [−0.17, 0.16]
COGA −0.07 [−0.37, 0.23]
COGB −0.09 [−0.42, 0.23]
COM1 −0.36 [−1.09, 0.38]
DAMI 0.14 [−0.10, 0.38]
EACR −0.69 [−1.24, −0.15]
FEEN −0.03 [−0.48, 0.42]
FSCD −0.11 [−0.52, 0.30]
FTCB −0.05 [−0.35, 0.24]
GFK 0.06 [−0.35, 0.46]
GMRF 0.41 [−0.23, 1.05]
GRAC 0.43 [−0.28, 1.15]
GSDC −0.19 [−0.51, 0.13]
GTPC 0.24 [−0.45, 0.93]
GUTS 0.16 [−0.26, 0.57]
INTR −0.04 [−0.72, 0.63]
MINV −0.09 [−0.73, 0.55]
MIRE 0.24 [−0.09, 0.57]
MRSC 0.25 [ 0.06, 0.45]
MRSD 0.21 [−0.48, 0.90]
NCMH −0.01 [−0.28, 0.27]
NCPT −0.15 [−0.61, 0.30]
NHS2 −0.01 [−0.27, 0.24]
NHSY 0.01 [−0.11, 0.14]
NIUT −0.28 [−1.28, 0.72]
NSS1 0.04 [−0.10, 0.17]
NSS2 −0.20 [−0.41, 0.02]
ONGA −0.19 [−0.82, 0.45]
ONGB −0.15 [−0.60, 0.30]
PPDS 0.04 [−0.09, 0.18]
PRIS 0.28 [−0.05, 0.60]
PSY5 0.23 [−0.19, 0.65]
PTS1 −0.01 [−0.41, 0.39]
QIM2 −0.21 [−0.55, 0.13]
QIMR −0.20 [−0.48, 0.08]
RCOG 0.03 [−0.20, 0.25]
RING 0.10 [−0.59, 0.79]
SEEP −0.11 [−0.50, 0.29]
STRO −0.01 [−0.30, 0.27]
TRAC 0.03 [−0.77, 0.83]
UKBB −0.01 [−0.03, 0.02]
VAKN −0.06 [−0.29, 0.17]
VETS 0.06 [−0.22, 0.34]
WACH 0.06 [−0.82, 0.95]
WRBY −0.17 [−0.63, 0.29]
WTCM 0.38 [−0.31, 1.08]
WTCS −0.06 [−0.31, 0.19]
F2.5 Meta −0.01 [−0.03, 0.02]
EHR
BIOM 0.06 [−0.21, 0.32]
DAI2 −0.07 [−0.10, −0.03]
ESBB −0.04 [−0.08, −0.01]
FING −0.05 [−0.10, 0.00]
HUNT −0.04 [−0.13, 0.04]
MAYO −0.01 [−0.16, 0.15]
MGBB −0.08 [−0.15, −0.00]
SWED −0.08 [−0.34, 0.18]
UKB2 0.01 [−0.04, 0.05]
EHR Meta −0.04 [−0.06, −0.02]
MVP
MVP1 −0.05 [−0.07, −0.02]
Freeze 3 Meta −0.03 [−0.04, −0.02]
−1.5
−0.5
0.5
1
1.5
Observed Outcome

rs197261 (Locus 13)

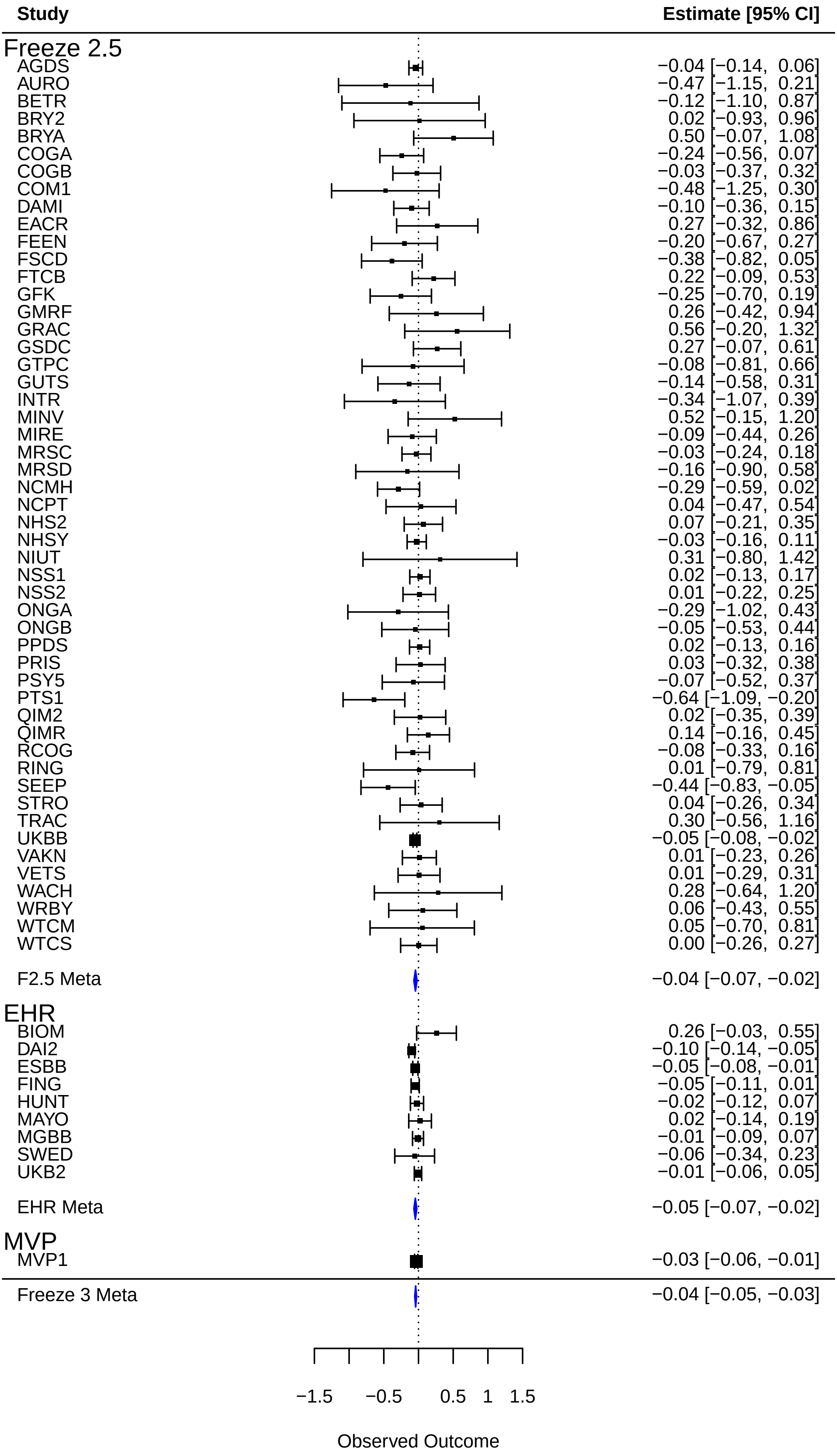

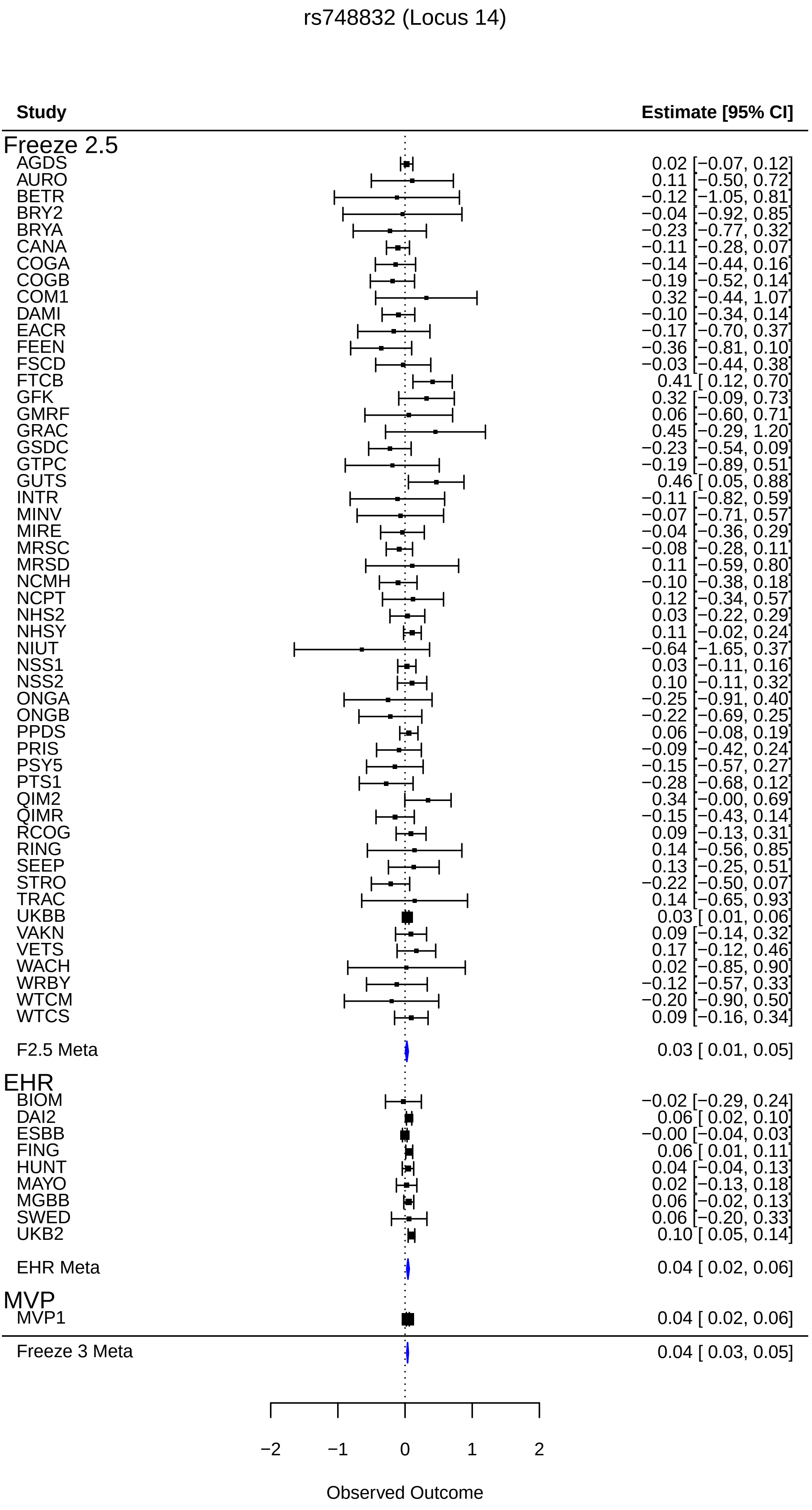

rs748832 (Locus 14)
Study
Estimate [95% CI]
Freeze 2.5
AGDS 0.02 [−0.07, 0.12]
AURO 0.11 [−0.50, 0.72]
BETR −0.12 [−1.05, 0.81]
BRY2 −0.04 [−0.92, 0.85]
BRYA −0.23 [−0.77, 0.32]
CANA −0.11 [−0.28, 0.07]
COGA −0.14 [−0.44, 0.16]
COGB −0.19 [−0.52, 0.14]
COM1 0.32 [−0.44, 1.07]
DAMI −0.10 [−0.34, 0.14]
EACR −0.17 [−0.70, 0.37]
FEEN −0.36 [−0.81, 0.10]
FSCD −0.03 [−0.44, 0.38]
FTCB 0.41 [ 0.12, 0.70]
GFK 0.32 [−0.09, 0.73]
GMRF 0.06 [−0.60, 0.71]
GRAC 0.45 [−0.29, 1.20]
GSDC −0.23 [−0.54, 0.09]
GTPC −0.19 [−0.89, 0.51]
GUTS 0.46 [ 0.05, 0.88]
INTR −0.11 [−0.82, 0.59]
MINV −0.07 [−0.71, 0.57]
MIRE −0.04 [−0.36, 0.29]
MRSC −0.08 [−0.28, 0.11]
MRSD 0.11 [−0.59, 0.80]
NCMH −0.10 [−0.38, 0.18]
NCPT 0.12 [−0.34, 0.57]
NHS2 0.03 [−0.22, 0.29]
NHSY 0.11 [−0.02, 0.24]
NIUT −0.64 [−1.65, 0.37]
NSS1 0.03 [−0.11, 0.16]
NSS2 0.10 [−0.11, 0.32]
ONGA −0.25 [−0.91, 0.40]
ONGB −0.22 [−0.69, 0.25]
PPDS 0.06 [−0.08, 0.19]
PRIS −0.09 [−0.42, 0.24]
PSY5 −0.15 [−0.57, 0.27]
PTS1 −0.28 [−0.68, 0.12]
QIM2 0.34 [−0.00, 0.69]
QIMR −0.15 [−0.43, 0.14]
RCOG 0.09 [−0.13, 0.31]
RING 0.14 [−0.56, 0.85]
SEEP 0.13 [−0.25, 0.51]
STRO −0.22 [−0.50, 0.07]
TRAC 0.14 [−0.65, 0.93]
UKBB 0.03 [ 0.01, 0.06]
VAKN 0.09 [−0.14, 0.32]
VETS 0.17 [−0.12, 0.46]
WACH 0.02 [−0.85, 0.90]
WRBY −0.12 [−0.57, 0.33]
WTCM −0.20 [−0.90, 0.50]
WTCS 0.09 [−0.16, 0.34]
F2.5 Meta 0.03 [ 0.01, 0.05]
EHR
BIOM −0.02 [−0.29, 0.24]
DAI2 0.06 [ 0.02, 0.10]
ESBB −0.00 [−0.04, 0.03]
FING 0.06 [ 0.01, 0.11]
HUNT 0.04 [−0.04, 0.13]
MAYO 0.02 [−0.13, 0.18]
MGBB 0.06 [−0.02, 0.13]
SWED 0.06 [−0.20, 0.33]
UKB2 0.10 [ 0.05, 0.14]
EHR Meta 0.04 [ 0.02, 0.06]
MVP
MVP1 0.04 [ 0.02, 0.06]
Freeze 3 Meta 0.04 [ 0.03, 0.05]
−2
−1
0
1
2
Observed Outcome

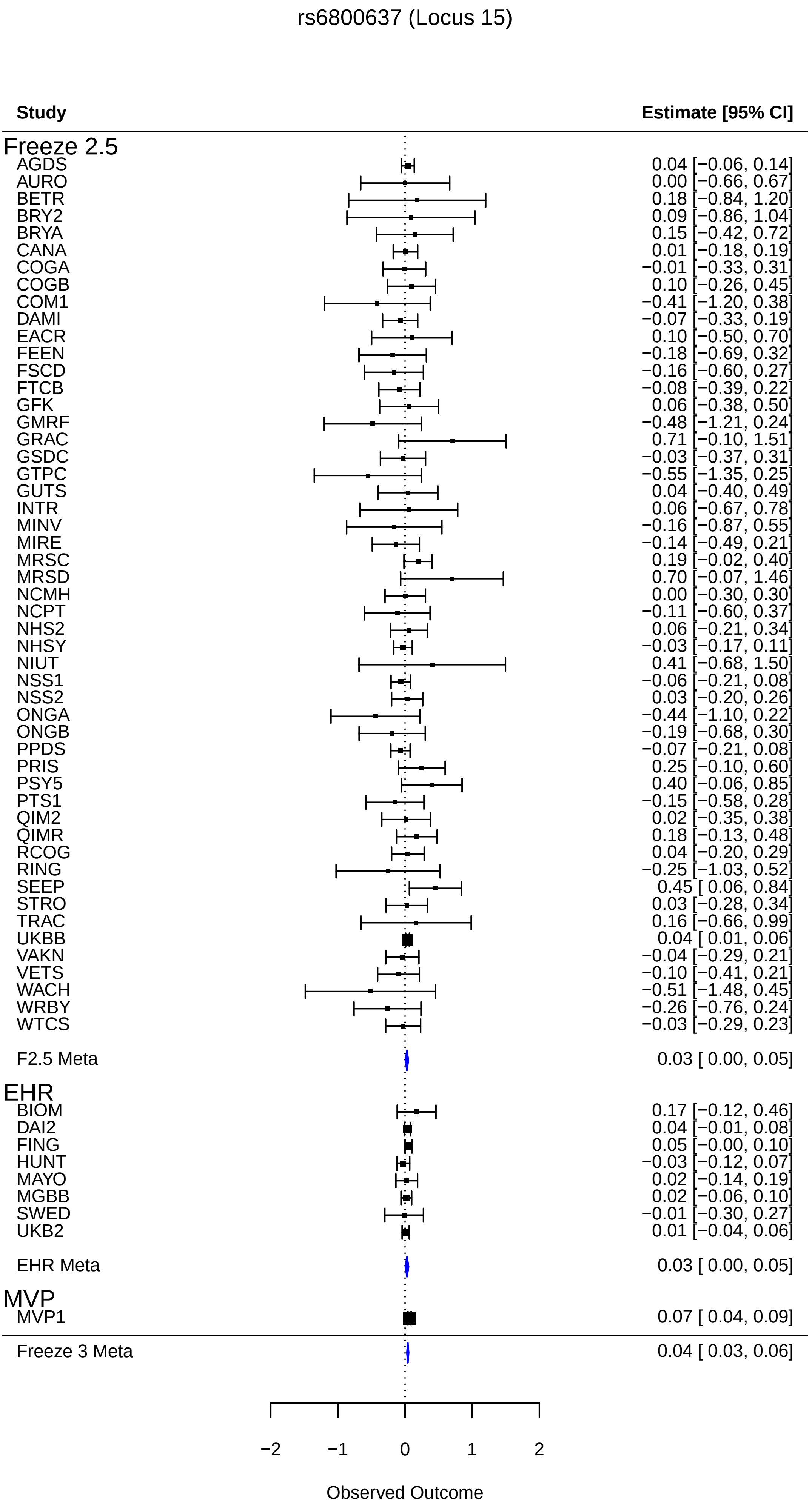
rs6800637 (Locus 15)
Study
Estimate [95% CI]
Freeze 2.5
AGDS 0.04 [−0.06, 0.14]
AURO 0.00 [−0.66, 0.67]
BETR 0.18 [−0.84, 1.20]
BRY2 0.09 [−0.86, 1.04]
BRYA 0.15 [−0.42, 0.72]
CANA 0.01 [−0.18, 0.19]
COGA −0.01 [−0.33, 0.31]
COGB 0.10 [−0.26, 0.45]
COM1 −0.41 [−1.20, 0.38]
DAMI −0.07 [−0.33, 0.19]
EACR 0.10 [−0.50, 0.70]
FEEN −0.18 [−0.69, 0.32]
FSCD −0.16 [−0.60, 0.27]
FTCB −0.08 [−0.39, 0.22]
GFK 0.06 [−0.38, 0.50]
GMRF −0.48 [−1.21, 0.24]
GRAC 0.71 [−0.10, 1.51]
GSDC −0.03 [−0.37, 0.31]
GTPC −0.55 [−1.35, 0.25]
GUTS 0.04 [−0.40, 0.49]
INTR 0.06 [−0.67, 0.78]
MINV −0.16 [−0.87, 0.55]
MIRE −0.14 [−0.49, 0.21]
MRSC 0.19 [−0.02, 0.40]
MRSD 0.70 [−0.07, 1.46]
NCMH 0.00 [−0.30, 0.30]
NCPT −0.11 [−0.60, 0.37]
NHS2 0.06 [−0.21, 0.34]
NHSY −0.03 [−0.17, 0.11]
NIUT 0.41 [−0.68, 1.50]
NSS1 −0.06 [−0.21, 0.08]
NSS2 0.03 [−0.20, 0.26]
ONGA −0.44 [−1.10, 0.22]
ONGB −0.19 [−0.68, 0.30]
PPDS −0.07 [−0.21, 0.08]
PRIS 0.25 [−0.10, 0.60]
PSY5 0.40 [−0.06, 0.85]
PTS1 −0.15 [−0.58, 0.28]
QIM2 0.02 [−0.35, 0.38]
QIMR 0.18 [−0.13, 0.48]
RCOG 0.04 [−0.20, 0.29]
RING −0.25 [−1.03, 0.52]
SEEP 0.45 [ 0.06, 0.84]
STRO 0.03 [−0.28, 0.34]
TRAC 0.16 [−0.66, 0.99]
UKBB 0.04 [ 0.01, 0.06]
VAKN −0.04 [−0.29, 0.21]
VETS −0.10 [−0.41, 0.21]
WACH −0.51 [−1.48, 0.45]
WRBY −0.26 [−0.76, 0.24]
WTCS −0.03 [−0.29, 0.23]
F2.5 Meta 0.03 [ 0.00, 0.05]
EHR
BIOM 0.17 [−0.12, 0.46]
DAI2 0.04 [−0.01, 0.08]
FING 0.05 [−0.00, 0.10]
HUNT −0.03 [−0.12, 0.07]
MAYO 0.02 [−0.14, 0.19]
MGBB 0.02 [−0.06, 0.10]
SWED −0.01 [−0.30, 0.27]
UKB2 0.01 [−0.04, 0.06]
EHR Meta 0.03 [ 0.00, 0.05]
MVP
MVP1 0.07 [ 0.04, 0.09]
Freeze 3 Meta 0.04 [ 0.03, 0.06]
−2
−1
0
1
2
Observed Outcome

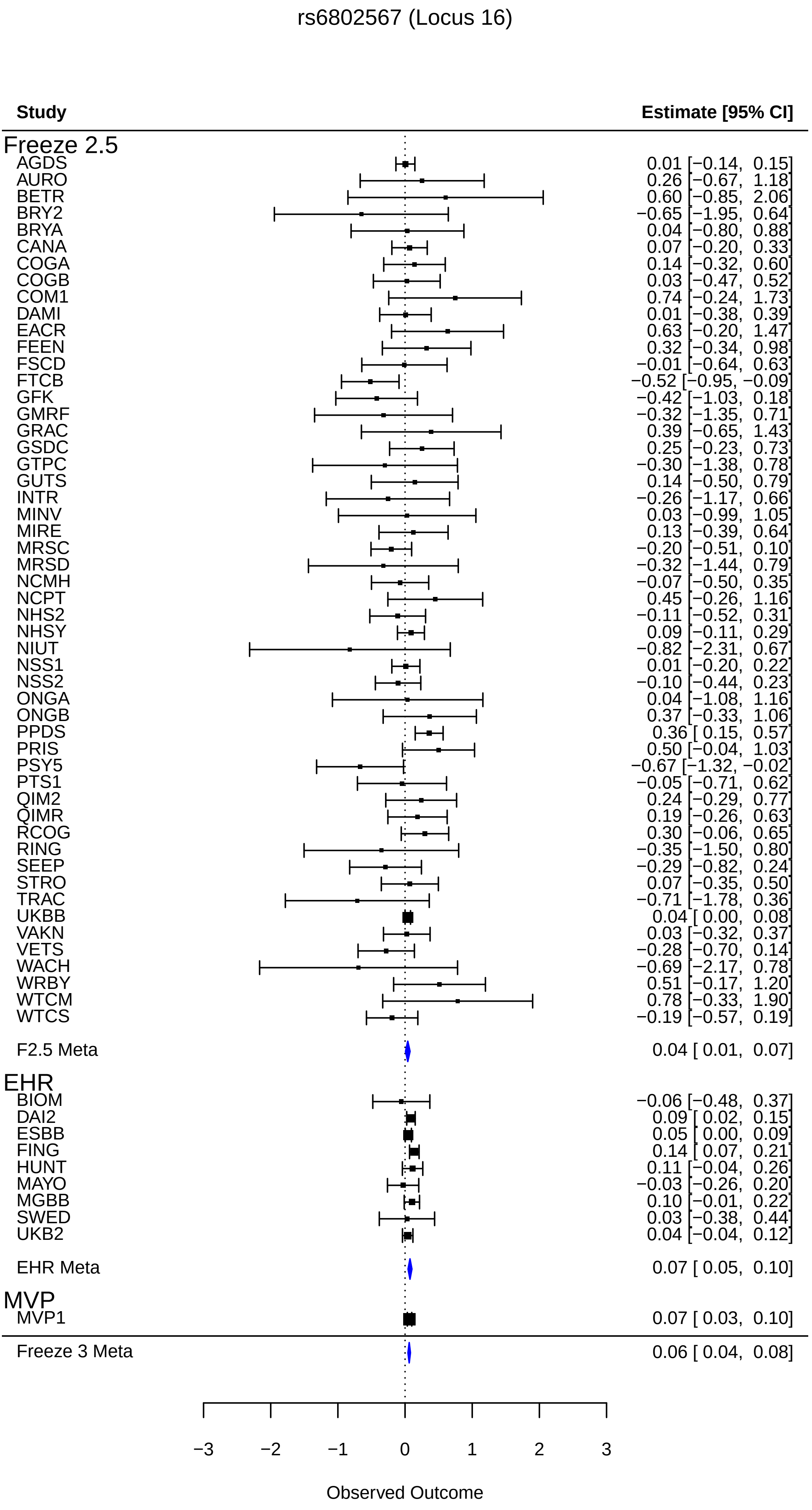

rs6802567 (Locus 16)
Study
Estimate [95% CI]
Freeze 2.5
AGDS 0.01 [−0.14, 0.15]
AURO 0.26 [−0.67, 1.18]
BETR 0.60 [−0.85, 2.06]
BRY2 −0.65 [−1.95, 0.64]
BRYA 0.04 [−0.80, 0.88]
CANA 0.07 [−0.20, 0.33]
COGA 0.14 [−0.32, 0.60]
COGB 0.03 [−0.47, 0.52]
COM1 0.74 [−0.24, 1.73]
DAMI 0.01 [−0.38, 0.39]
EACR 0.63 [−0.20, 1.47]
FEEN 0.32 [−0.34, 0.98]
FSCD −0.01 [−0.64, 0.63]
FTCB −0.52 [−0.95, −0.09]
GFK −0.42 [−1.03, 0.18]
GMRF −0.32 [−1.35, 0.71]
GRAC 0.39 [−0.65, 1.43]
GSDC 0.25 [−0.23, 0.73]
GTPC −0.30 [−1.38, 0.78]
GUTS 0.14 [−0.50, 0.79]
INTR −0.26 [−1.17, 0.66]
MINV 0.03 [−0.99, 1.05]
MIRE 0.13 [−0.39, 0.64]
MRSC −0.20 [−0.51, 0.10]
MRSD −0.32 [−1.44, 0.79]
NCMH −0.07 [−0.50, 0.35]
NCPT 0.45 [−0.26, 1.16]
NHS2 −0.11 [−0.52, 0.31]
NHSY 0.09 [−0.11, 0.29]
NIUT −0.82 [−2.31, 0.67]
NSS1 0.01 [−0.20, 0.22]
NSS2 −0.10 [−0.44, 0.23]
ONGA 0.04 [−1.08, 1.16]
ONGB 0.37 [−0.33, 1.06]
PPDS 0.36 [ 0.15, 0.57]
PRIS 0.50 [−0.04, 1.03]
PSY5 −0.67 [−1.32, −0.02]
PTS1 −0.05 [−0.71, 0.62]
QIM2 0.24 [−0.29, 0.77]
QIMR 0.19 [−0.26, 0.63]
RCOG 0.30 [−0.06, 0.65]
RING −0.35 [−1.50, 0.80]
SEEP −0.29 [−0.82, 0.24]
STRO 0.07 [−0.35, 0.50]
TRAC −0.71 [−1.78, 0.36]
UKBB 0.04 [ 0.00, 0.08]
VAKN 0.03 [−0.32, 0.37]
VETS −0.28 [−0.70, 0.14]
WACH −0.69 [−2.17, 0.78]
WRBY 0.51 [−0.17, 1.20]
WTCM 0.78 [−0.33, 1.90]
WTCS −0.19 [−0.57, 0.19]
F2.5 Meta 0.04 [ 0.01, 0.07]
EHR
BIOM −0.06 [−0.48, 0.37]
DAI2 0.09 [ 0.02, 0.15]
ESBB 0.05 [ 0.00, 0.09]
FING 0.14 [ 0.07, 0.21]
HUNT 0.11 [−0.04, 0.26]
MAYO −0.03 [−0.26, 0.20]
MGBB 0.10 [−0.01, 0.22]
SWED 0.03 [−0.38, 0.44]
UKB2 0.04 [−0.04, 0.12]
EHR Meta 0.07 [ 0.05, 0.10]
MVP
MVP1 0.07 [ 0.03, 0.10]
Freeze 3 Meta 0.06 [ 0.04, 0.08]
−3
−2
−1
0
1
2
3
Observed Outcome

rs11130221 (Locus 17)

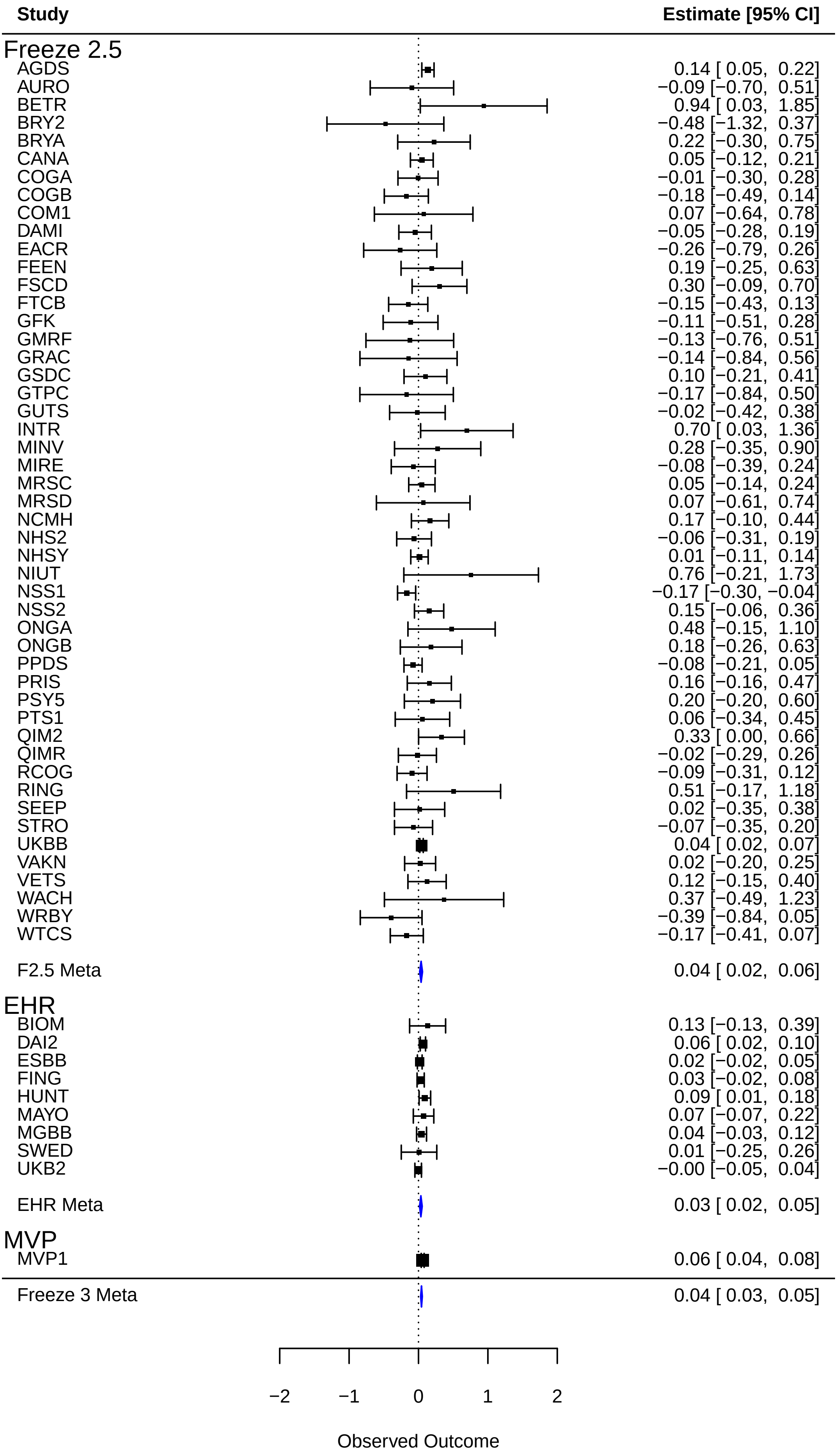

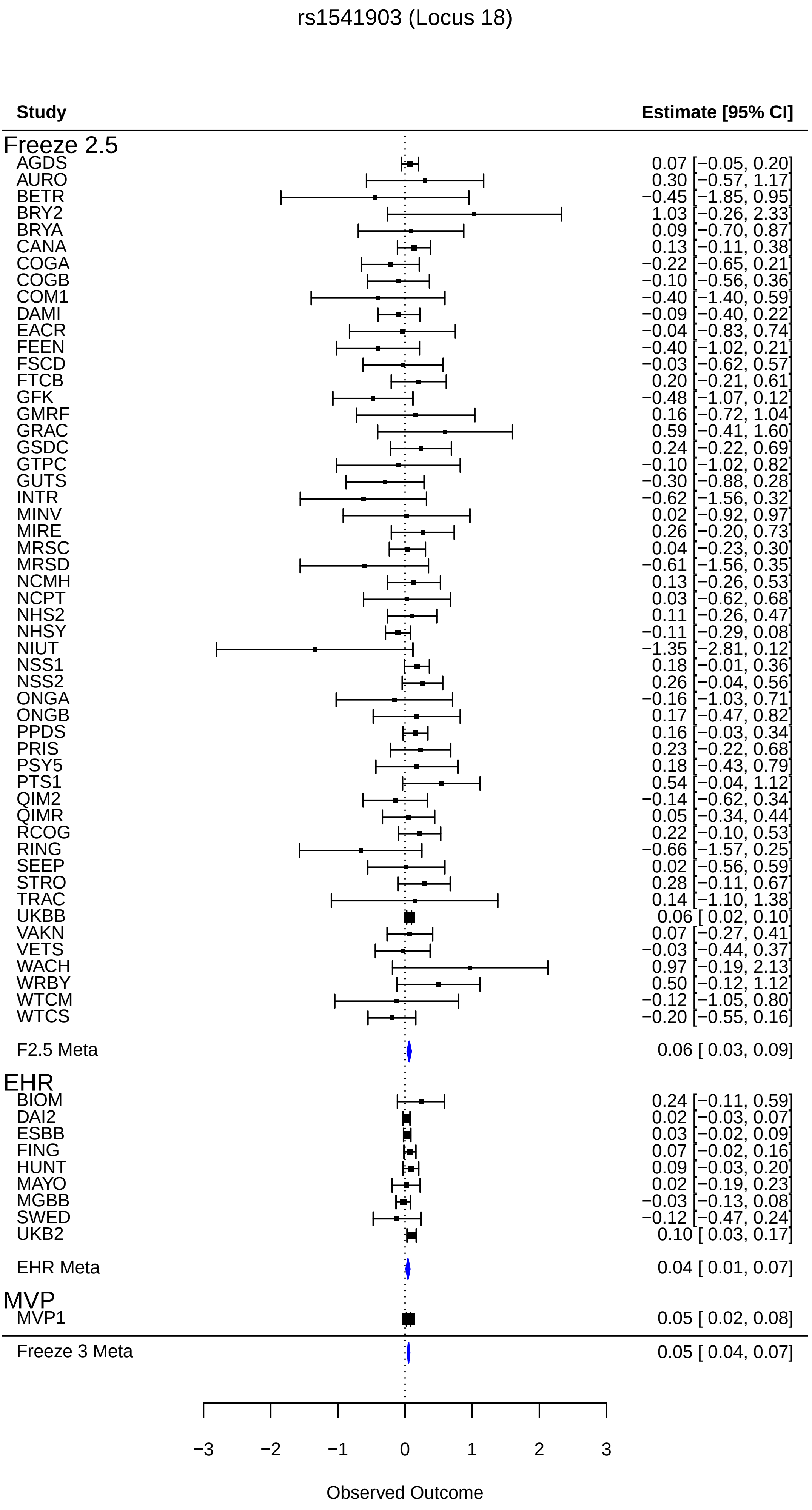
rs1541903 (Locus 18)
Study
Estimate [95% CI]
Freeze 2.5
AGDS 0.07 [−0.05, 0.20]
AURO 0.30 [−0.57, 1.17]
BETR −0.45 [−1.85, 0.95]
BRY2 1.03 [−0.26, 2.33]
BRYA 0.09 [−0.70, 0.87]
CANA 0.13 [−0.11, 0.38]
COGA −0.22 [−0.65, 0.21]
COGB −0.10 [−0.56, 0.36]
COM1 −0.40 [−1.40, 0.59]
DAMI −0.09 [−0.40, 0.22]
EACR −0.04 [−0.83, 0.74]
FEEN −0.40 [−1.02, 0.21]
FSCD −0.03 [−0.62, 0.57]
FTCB 0.20 [−0.21, 0.61]
GFK −0.48 [−1.07, 0.12]
GMRF 0.16 [−0.72, 1.04]
GRAC 0.59 [−0.41, 1.60]
GSDC 0.24 [−0.22, 0.69]
GTPC −0.10 [−1.02, 0.82]
GUTS −0.30 [−0.88, 0.28]
INTR −0.62 [−1.56, 0.32]
MINV 0.02 [−0.92, 0.97]
MIRE 0.26 [−0.20, 0.73]
MRSC 0.04 [−0.23, 0.30]
MRSD −0.61 [−1.56, 0.35]
NCMH 0.13 [−0.26, 0.53]
NCPT 0.03 [−0.62, 0.68]
NHS2 0.11 [−0.26, 0.47]
NHSY −0.11 [−0.29, 0.08]
NIUT −1.35 [−2.81, 0.12]
NSS1 0.18 [−0.01, 0.36]
NSS2 0.26 [−0.04, 0.56]
ONGA −0.16 [−1.03, 0.71]
ONGB 0.17 [−0.47, 0.82]
PPDS 0.16 [−0.03, 0.34]
PRIS 0.23 [−0.22, 0.68]
PSY5 0.18 [−0.43, 0.79]
PTS1 0.54 [−0.04, 1.12]
QIM2 −0.14 [−0.62, 0.34]
QIMR 0.05 [−0.34, 0.44]
RCOG 0.22 [−0.10, 0.53]
RING −0.66 [−1.57, 0.25]
SEEP 0.02 [−0.56, 0.59]
STRO 0.28 [−0.11, 0.67]
TRAC 0.14 [−1.10, 1.38]
UKBB 0.06 [ 0.02, 0.10]
VAKN 0.07 [−0.27, 0.41]
VETS −0.03 [−0.44, 0.37]
WACH 0.97 [−0.19, 2.13]
WRBY 0.50 [−0.12, 1.12]
WTCM −0.12 [−1.05, 0.80]
WTCS −0.20 [−0.55, 0.16]
F2.5 Meta 0.06 [ 0.03, 0.09]
EHR
BIOM 0.24 [−0.11, 0.59]
DAI2 0.02 [−0.03, 0.07]
ESBB 0.03 [−0.02, 0.09]
FING 0.07 [−0.02, 0.16]
HUNT 0.09 [−0.03, 0.20]
MAYO 0.02 [−0.19, 0.23]
MGBB −0.03 [−0.13, 0.08]
SWED −0.12 [−0.47, 0.24]
UKB2 0.10 [ 0.03, 0.17]
EHR Meta 0.04 [ 0.01, 0.07]
MVP
MVP1 0.05 [ 0.02, 0.08]
Freeze 3 Meta 0.05 [ 0.04, 0.07]
−3
−2
−1
0
1
2
3
Observed Outcome

rs34811474 (Locus 19)

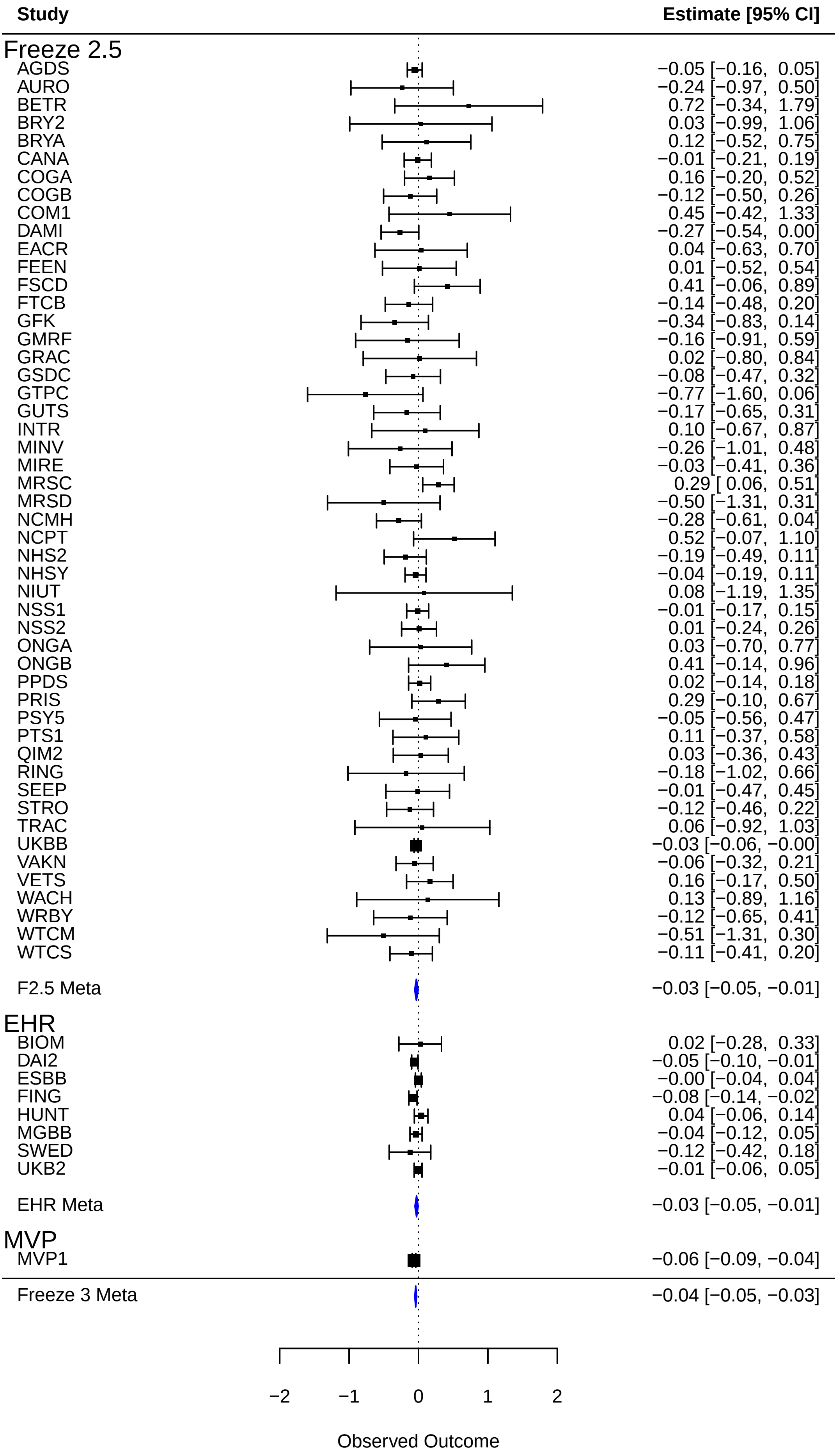

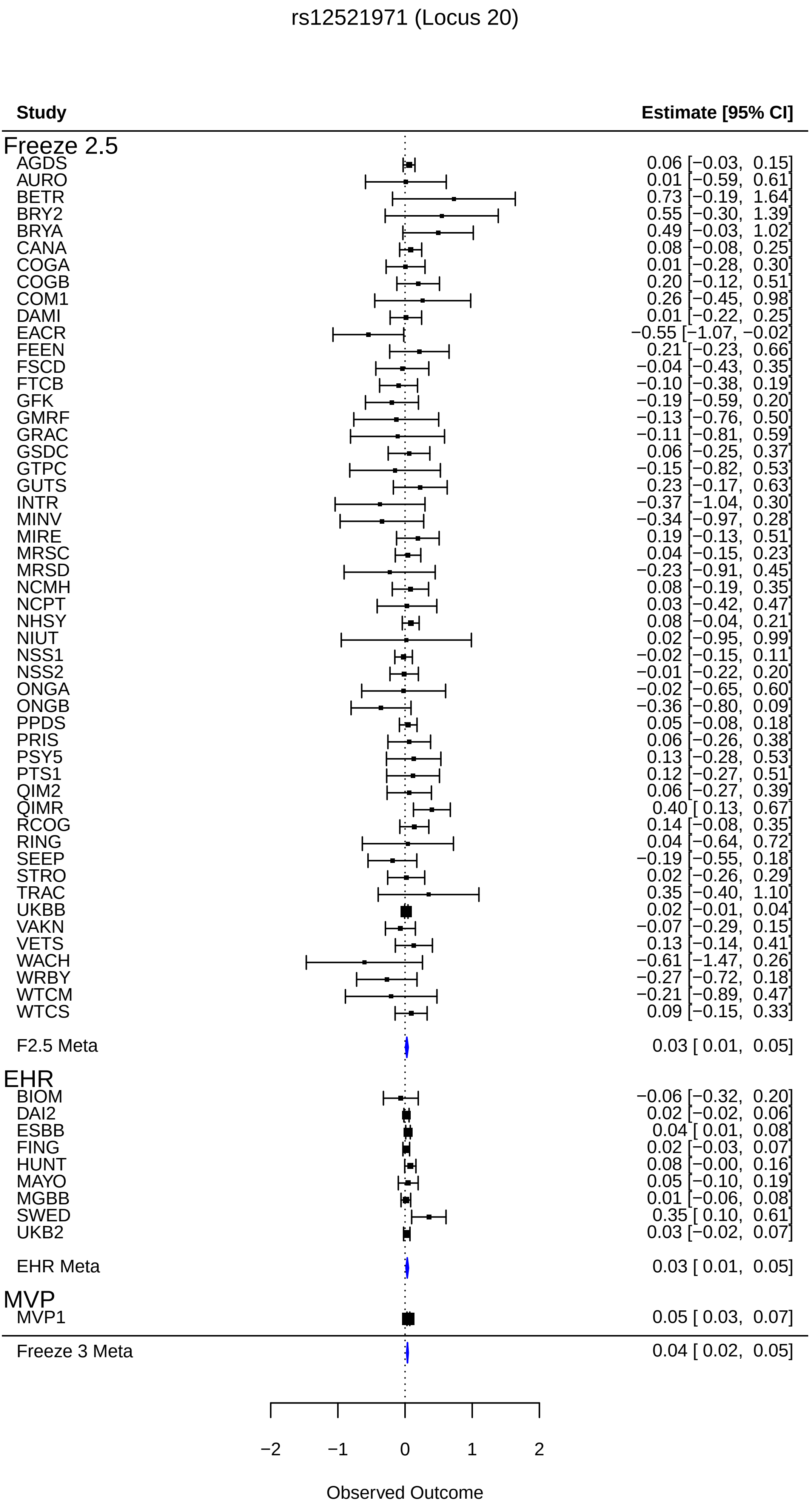
rs12521971 (Locus 20)
Study
Estimate [95% CI]
Freeze 2.5
AGDS 0.06 [−0.03, 0.15]
AURO 0.01 [−0.59, 0.61]
BETR 0.73 [−0.19, 1.64]
BRY2 0.55 [−0.30, 1.39]
BRYA 0.49 [−0.03, 1.02]
CANA 0.08 [−0.08, 0.25]
COGA 0.01 [−0.28, 0.30]
COGB 0.20 [−0.12, 0.51]
COM1 0.26 [−0.45, 0.98]
DAMI 0.01 [−0.22, 0.25]
EACR −0.55 [−1.07, −0.02]
FEEN 0.21 [−0.23, 0.66]
FSCD −0.04 [−0.43, 0.35]
FTCB −0.10 [−0.38, 0.19]
GFK −0.19 [−0.59, 0.20]
GMRF −0.13 [−0.76, 0.50]
GRAC −0.11 [−0.81, 0.59]
GSDC 0.06 [−0.25, 0.37]
GTPC −0.15 [−0.82, 0.53]
GUTS 0.23 [−0.17, 0.63]
INTR −0.37 [−1.04, 0.30]
MINV −0.34 [−0.97, 0.28]
MIRE 0.19 [−0.13, 0.51]
MRSC 0.04 [−0.15, 0.23]
MRSD −0.23 [−0.91, 0.45]
NCMH 0.08 [−0.19, 0.35]
NCPT 0.03 [−0.42, 0.47]
NHSY 0.08 [−0.04, 0.21]
NIUT 0.02 [−0.95, 0.99]
NSS1 −0.02 [−0.15, 0.11]
NSS2 −0.01 [−0.22, 0.20]
ONGA −0.02 [−0.65, 0.60]
ONGB −0.36 [−0.80, 0.09]
PPDS 0.05 [−0.08, 0.18]
PRIS 0.06 [−0.26, 0.38]
PSY5 0.13 [−0.28, 0.53]
PTS1 0.12 [−0.27, 0.51]
QIM2 0.06 [−0.27, 0.39]
QIMR 0.40 [ 0.13, 0.67]
RCOG 0.14 [−0.08, 0.35]
RING 0.04 [−0.64, 0.72]
SEEP −0.19 [−0.55, 0.18]
STRO 0.02 [−0.26, 0.29]
TRAC 0.35 [−0.40, 1.10]
UKBB 0.02 [−0.01, 0.04]
VAKN −0.07 [−0.29, 0.15]
VETS 0.13 [−0.14, 0.41]
WACH −0.61 [−1.47, 0.26]
WRBY −0.27 [−0.72, 0.18]
WTCM −0.21 [−0.89, 0.47]
WTCS 0.09 [−0.15, 0.33]
F2.5 Meta 0.03 [ 0.01, 0.05]
EHR
BIOM −0.06 [−0.32, 0.20]
DAI2 0.02 [−0.02, 0.06]
ESBB 0.04 [ 0.01, 0.08]
FING 0.02 [−0.03, 0.07]
HUNT 0.08 [−0.00, 0.16]
MAYO 0.05 [−0.10, 0.19]
MGBB 0.01 [−0.06, 0.08]
SWED 0.35 [ 0.10, 0.61]
UKB2 0.03 [−0.02, 0.07]
EHR Meta 0.03 [ 0.01, 0.05]
MVP
MVP1 0.05 [ 0.03, 0.07]
Freeze 3 Meta 0.04 [ 0.02, 0.05]
−2
−1
0
1
2
Observed Outcome

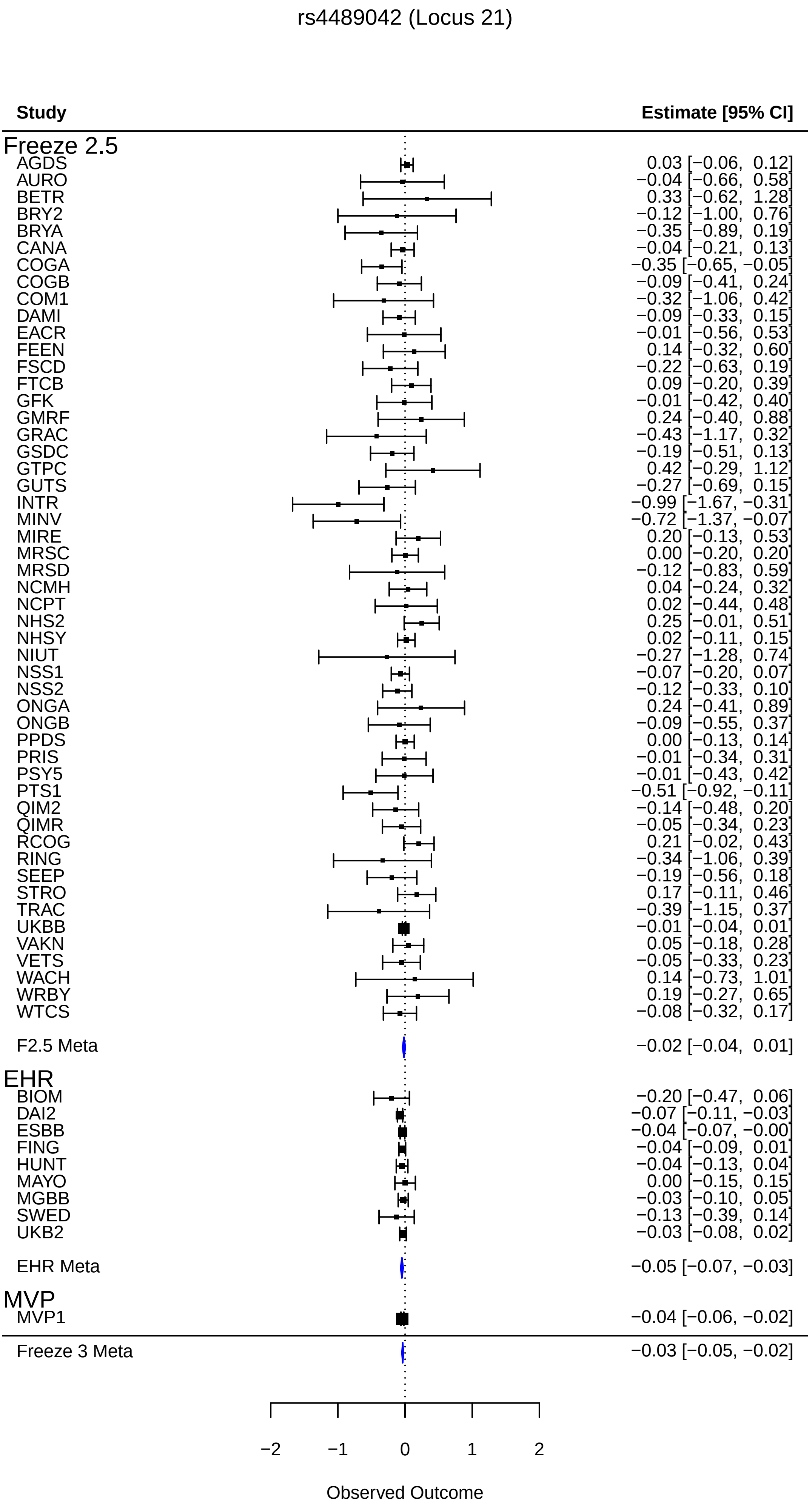
rs4489042 (Locus 21)
Study
Estimate [95% CI]
Freeze 2.5
AGDS 0.03 [−0.06, 0.12]
AURO −0.04 [−0.66, 0.58]
BETR 0.33 [−0.62, 1.28]
BRY2 −0.12 [−1.00, 0.76]
BRYA −0.35 [−0.89, 0.19]
CANA −0.04 [−0.21, 0.13]
COGA −0.35 [−0.65, −0.05]
COGB −0.09 [−0.41, 0.24]
COM1 −0.32 [−1.06, 0.42]
DAMI −0.09 [−0.33, 0.15]
EACR −0.01 [−0.56, 0.53]
FEEN 0.14 [−0.32, 0.60]
FSCD −0.22 [−0.63, 0.19]
FTCB 0.09 [−0.20, 0.39]
GFK −0.01 [−0.42, 0.40]
GMRF 0.24 [−0.40, 0.88]
GRAC −0.43 [−1.17, 0.32]
GSDC −0.19 [−0.51, 0.13]
GTPC 0.42 [−0.29, 1.12]
GUTS −0.27 [−0.69, 0.15]
INTR −0.99 [−1.67, −0.31]
MINV −0.72 [−1.37, −0.07]
MIRE 0.20 [−0.13, 0.53]
MRSC 0.00 [−0.20, 0.20]
MRSD −0.12 [−0.83, 0.59]
NCMH 0.04 [−0.24, 0.32]
NCPT 0.02 [−0.44, 0.48]
NHS2 0.25 [−0.01, 0.51]
NHSY 0.02 [−0.11, 0.15]
NIUT −0.27 [−1.28, 0.74]
NSS1 −0.07 [−0.20, 0.07]
NSS2 −0.12 [−0.33, 0.10]
ONGA 0.24 [−0.41, 0.89]
ONGB −0.09 [−0.55, 0.37]
PPDS 0.00 [−0.13, 0.14]
PRIS −0.01 [−0.34, 0.31]
PSY5 −0.01 [−0.43, 0.42]
PTS1 −0.51 [−0.92, −0.11]
QIM2 −0.14 [−0.48, 0.20]
QIMR −0.05 [−0.34, 0.23]
RCOG 0.21 [−0.02, 0.43]
RING −0.34 [−1.06, 0.39]
SEEP −0.19 [−0.56, 0.18]
STRO 0.17 [−0.11, 0.46]
TRAC −0.39 [−1.15, 0.37]
UKBB −0.01 [−0.04, 0.01]
VAKN 0.05 [−0.18, 0.28]
VETS −0.05 [−0.33, 0.23]
WACH 0.14 [−0.73, 1.01]
WRBY 0.19 [−0.27, 0.65]
WTCS −0.08 [−0.32, 0.17]
F2.5 Meta −0.02 [−0.04, 0.01]
EHR
BIOM −0.20 [−0.47, 0.06]
DAI2 −0.07 [−0.11, −0.03]
ESBB −0.04 [−0.07, −0.00]
FING −0.04 [−0.09, 0.01]
HUNT −0.04 [−0.13, 0.04]
MAYO 0.00 [−0.15, 0.15]
MGBB −0.03 [−0.10, 0.05]
SWED −0.13 [−0.39, 0.14]
UKB2 −0.03 [−0.08, 0.02]
EHR Meta −0.05 [−0.07, −0.03]
MVP
MVP1 −0.04 [−0.06, −0.02]
Freeze 3 Meta −0.03 [−0.05, −0.02]
−2
−1
0
1
2
Observed Outcome

rs33817 (Locus 22)

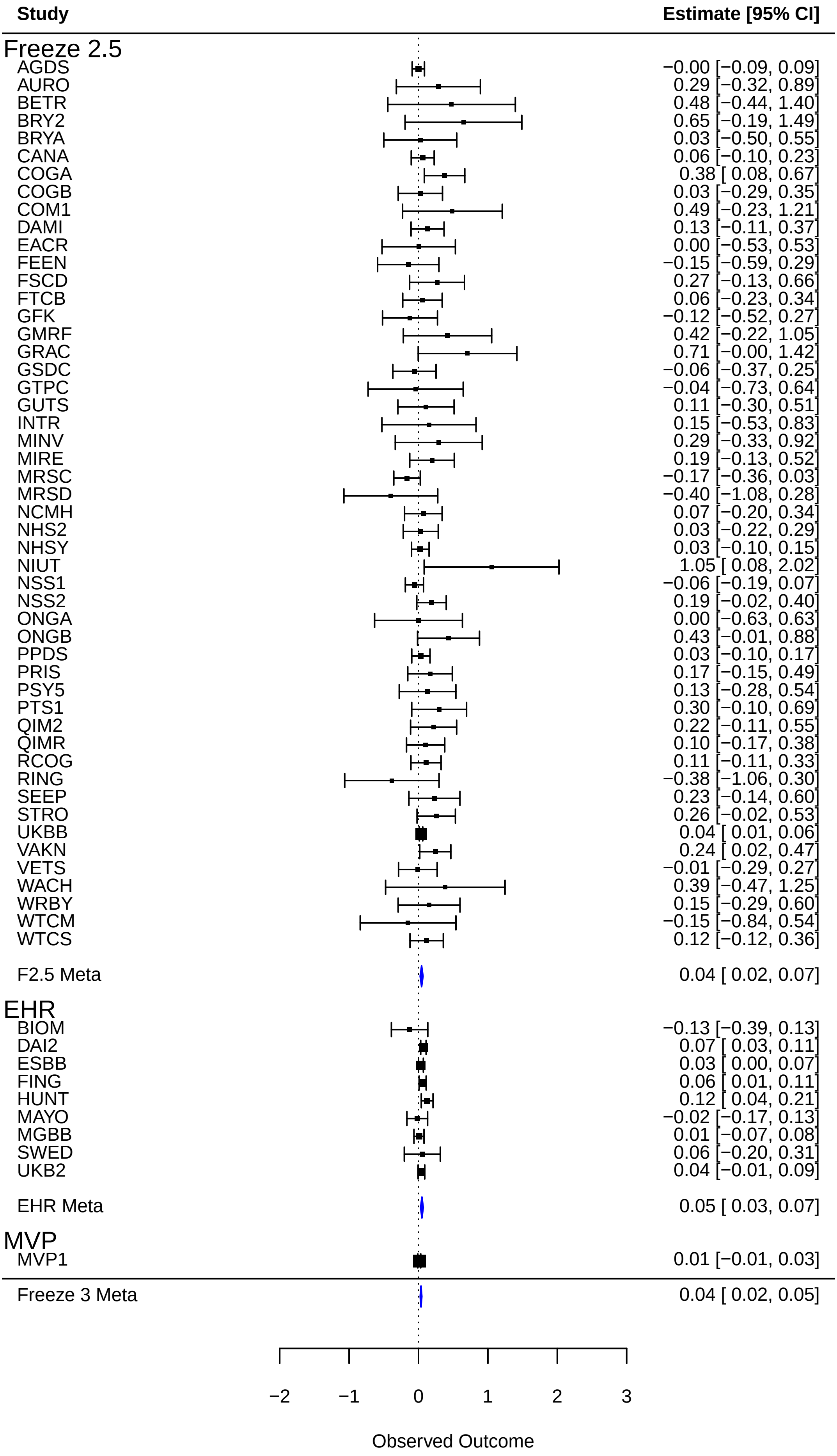

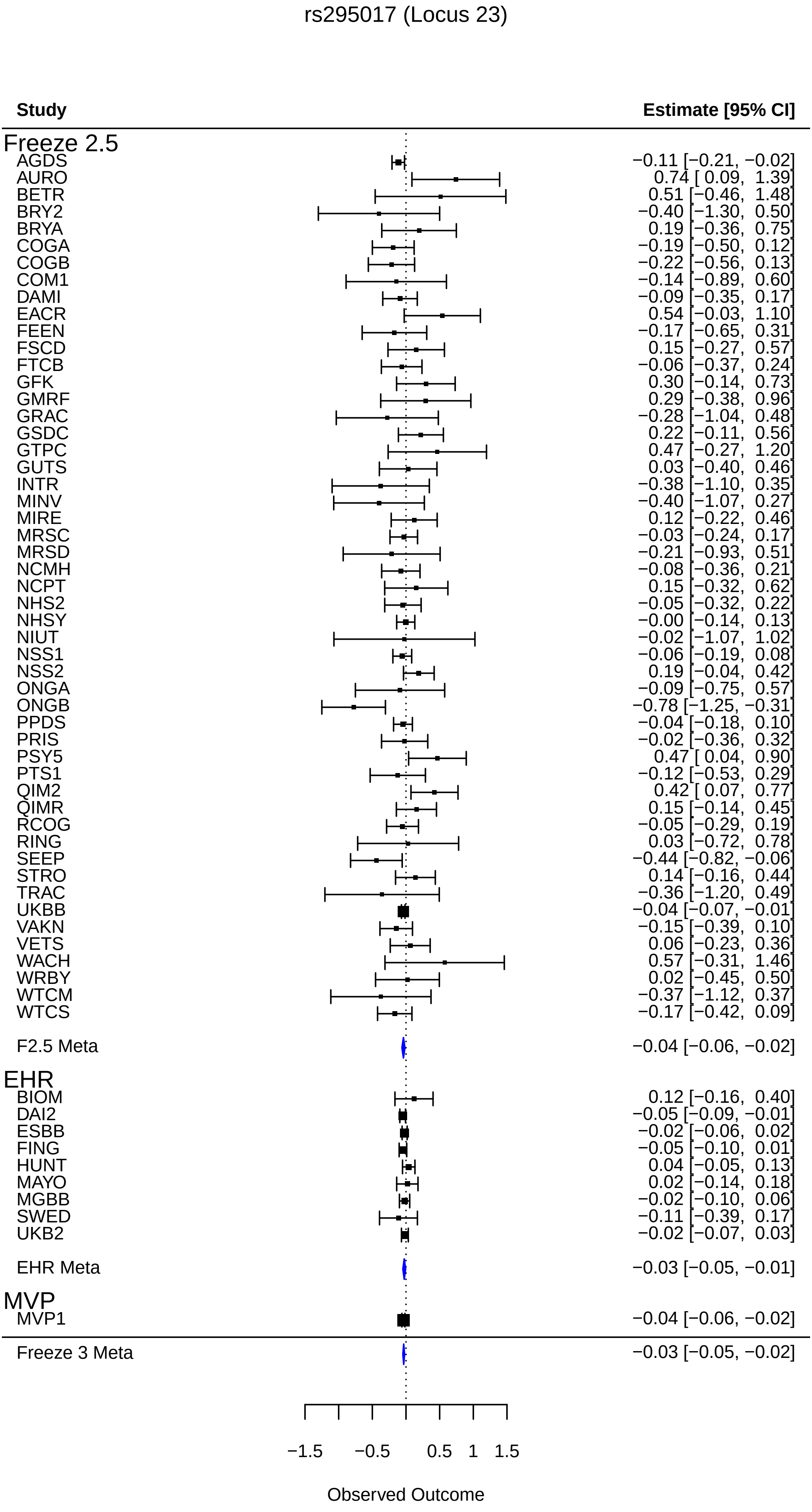
rs295017 (Locus 23)
Study
Estimate [95% CI]
Freeze 2.5
AGDS −0.11 [−0.21, −0.02]
AURO 0.74 [ 0.09, 1.39]
BETR 0.51 [−0.46, 1.48]
BRY2 −0.40 [−1.30, 0.50]
BRYA 0.19 [−0.36, 0.75]
COGA −0.19 [−0.50, 0.12]
COGB −0.22 [−0.56, 0.13]
COM1 −0.14 [−0.89, 0.60]
DAMI −0.09 [−0.35, 0.17]
EACR 0.54 [−0.03, 1.10]
FEEN −0.17 [−0.65, 0.31]
FSCD 0.15 [−0.27, 0.57]
FTCB −0.06 [−0.37, 0.24]
GFK 0.30 [−0.14, 0.73]
GMRF 0.29 [−0.38, 0.96]
GRAC −0.28 [−1.04, 0.48]
GSDC 0.22 [−0.11, 0.56]
GTPC 0.47 [−0.27, 1.20]
GUTS 0.03 [−0.40, 0.46]
INTR −0.38 [−1.10, 0.35]
MINV −0.40 [−1.07, 0.27]
MIRE 0.12 [−0.22, 0.46]
MRSC −0.03 [−0.24, 0.17]
MRSD −0.21 [−0.93, 0.51]
NCMH −0.08 [−0.36, 0.21]
NCPT 0.15 [−0.32, 0.62]
NHS2 −0.05 [−0.32, 0.22]
NHSY −0.00 [−0.14, 0.13]
NIUT −0.02 [−1.07, 1.02]
NSS1 −0.06 [−0.19, 0.08]
NSS2 0.19 [−0.04, 0.42]
ONGA −0.09 [−0.75, 0.57]
ONGB −0.78 [−1.25, −0.31]
PPDS −0.04 [−0.18, 0.10]
PRIS −0.02 [−0.36, 0.32]
PSY5 0.47 [ 0.04, 0.90]
PTS1 −0.12 [−0.53, 0.29]
QIM2 0.42 [ 0.07, 0.77]
QIMR 0.15 [−0.14, 0.45]
RCOG −0.05 [−0.29, 0.19]
RING 0.03 [−0.72, 0.78]
SEEP −0.44 [−0.82, −0.06]
STRO 0.14 [−0.16, 0.44]
TRAC −0.36 [−1.20, 0.49]
UKBB −0.04 [−0.07, −0.01]
VAKN −0.15 [−0.39, 0.10]
VETS 0.06 [−0.23, 0.36]
WACH 0.57 [−0.31, 1.46]
WRBY 0.02 [−0.45, 0.50]
WTCM −0.37 [−1.12, 0.37]
WTCS −0.17 [−0.42, 0.09]
F2.5 Meta −0.04 [−0.06, −0.02]
EHR
BIOM 0.12 [−0.16, 0.40]
DAI2 −0.05 [−0.09, −0.01]
ESBB −0.02 [−0.06, 0.02]
FING −0.05 [−0.10, 0.01]
HUNT 0.04 [−0.05, 0.13]
MAYO 0.02 [−0.14, 0.18]
MGBB −0.02 [−0.10, 0.06]
SWED −0.11 [−0.39, 0.17]
UKB2 −0.02 [−0.07, 0.03]
EHR Meta −0.03 [−0.05, −0.01]
MVP
MVP1 −0.04 [−0.06, −0.02]
Freeze 3 Meta −0.03 [−0.05, −0.02]
−1.5
−0.5
0.5
1
1.5
Observed Outcome

rs13161130 (Locus 24)

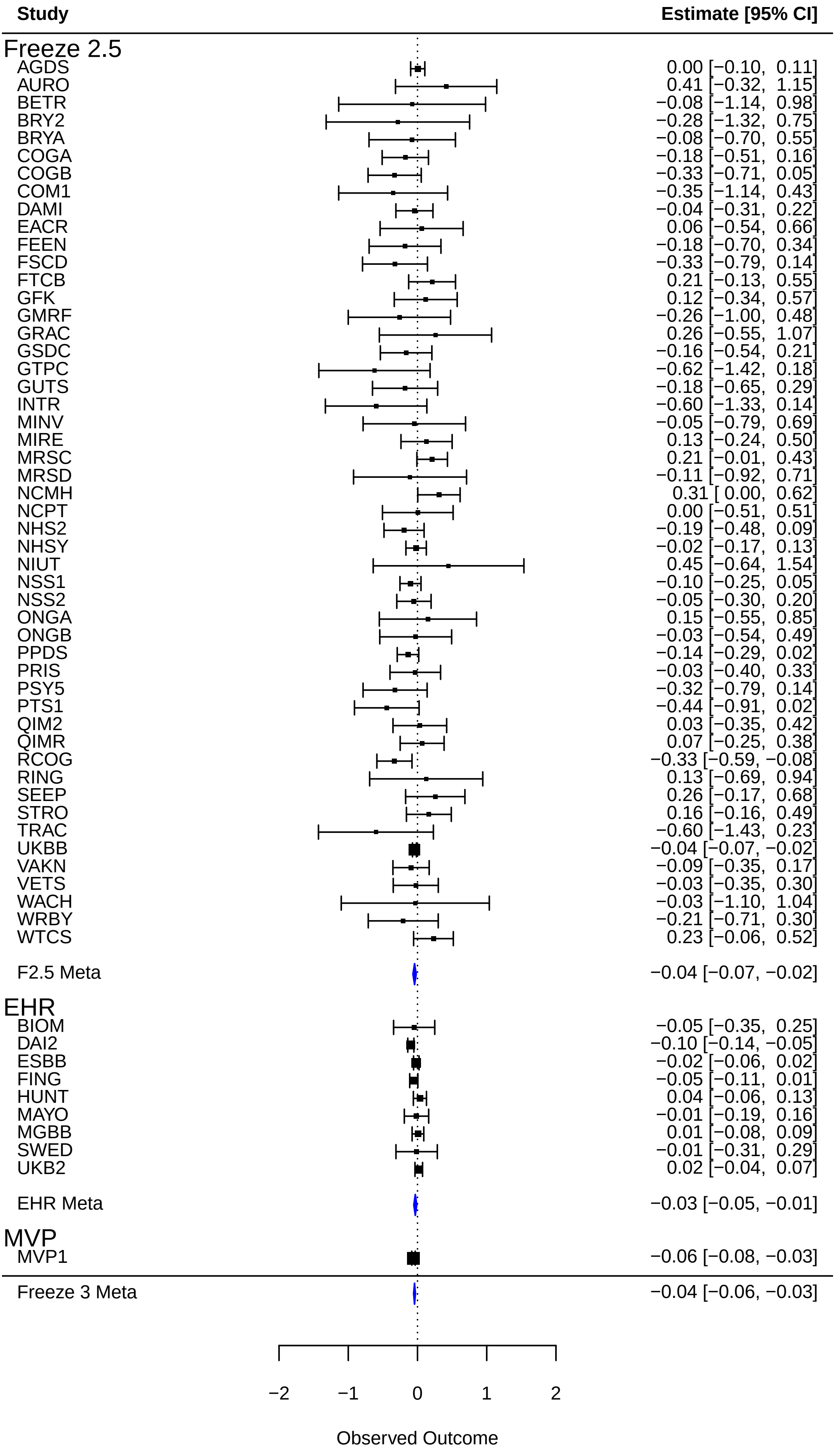

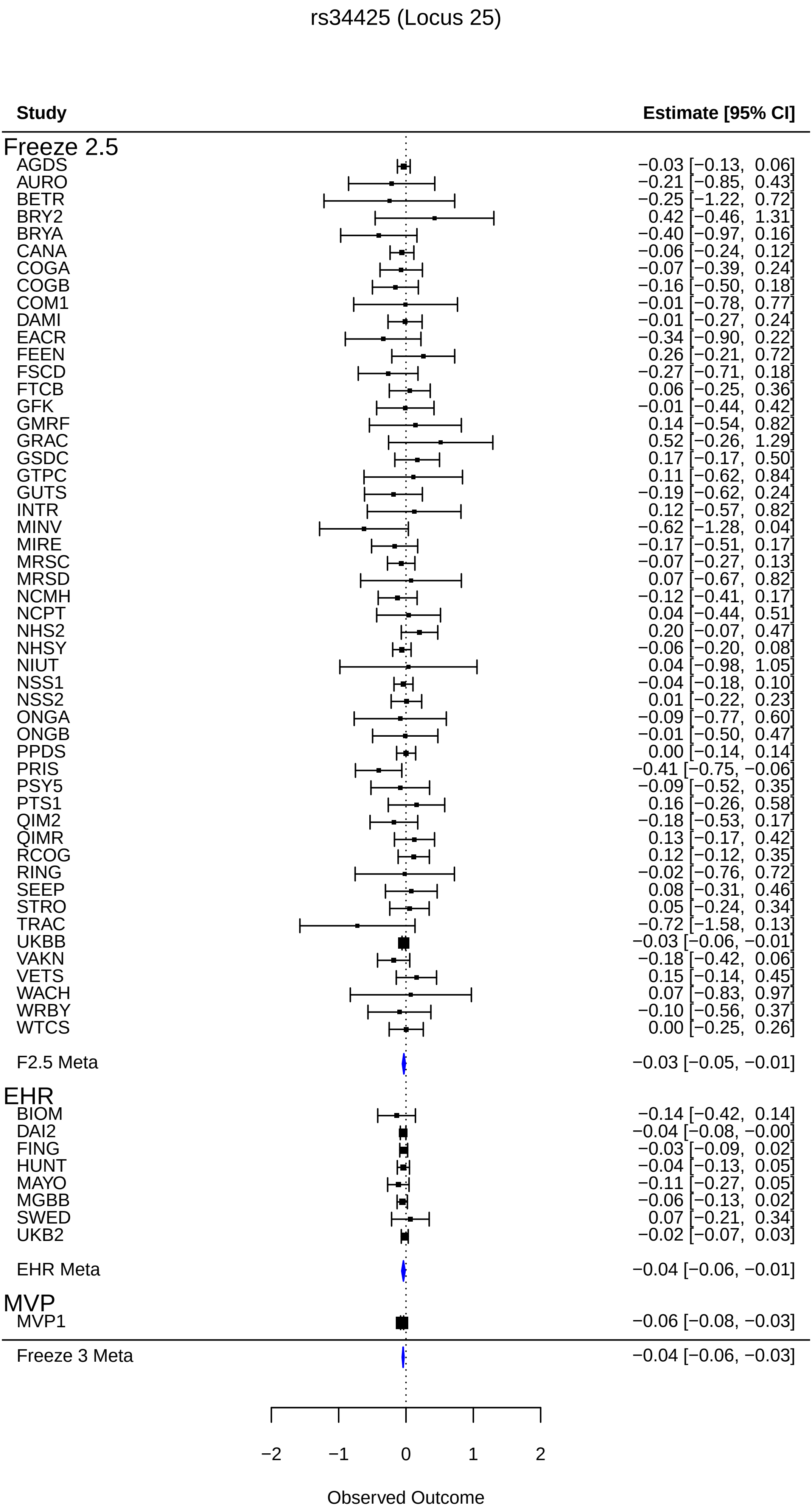
rs34425 (Locus 25)
Study
Estimate [95% CI]
Freeze 2.5
AGDS −0.03 [−0.13, 0.06]
AURO −0.21 [−0.85, 0.43]
BETR −0.25 [−1.22, 0.72]
BRY2 0.42 [−0.46, 1.31]
BRYA −0.40 [−0.97, 0.16]
CANA −0.06 [−0.24, 0.12]
COGA −0.07 [−0.39, 0.24]
COGB −0.16 [−0.50, 0.18]
COM1 −0.01 [−0.78, 0.77]
DAMI −0.01 [−0.27, 0.24]
EACR −0.34 [−0.90, 0.22]
FEEN 0.26 [−0.21, 0.72]
FSCD −0.27 [−0.71, 0.18]
FTCB 0.06 [−0.25, 0.36]
GFK −0.01 [−0.44, 0.42]
GMRF 0.14 [−0.54, 0.82]
GRAC 0.52 [−0.26, 1.29]
GSDC 0.17 [−0.17, 0.50]
GTPC 0.11 [−0.62, 0.84]
GUTS −0.19 [−0.62, 0.24]
INTR 0.12 [−0.57, 0.82]
MINV −0.62 [−1.28, 0.04]
MIRE −0.17 [−0.51, 0.17]
MRSC −0.07 [−0.27, 0.13]
MRSD 0.07 [−0.67, 0.82]
NCMH −0.12 [−0.41, 0.17]
NCPT 0.04 [−0.44, 0.51]
NHS2 0.20 [−0.07, 0.47]
NHSY −0.06 [−0.20, 0.08]
NIUT 0.04 [−0.98, 1.05]
NSS1 −0.04 [−0.18, 0.10]
NSS2 0.01 [−0.22, 0.23]
ONGA −0.09 [−0.77, 0.60]
ONGB −0.01 [−0.50, 0.47]
PPDS 0.00 [−0.14, 0.14]
PRIS −0.41 [−0.75, −0.06]
PSY5 −0.09 [−0.52, 0.35]
PTS1 0.16 [−0.26, 0.58]
QIM2 −0.18 [−0.53, 0.17]
QIMR 0.13 [−0.17, 0.42]
RCOG 0.12 [−0.12, 0.35]
RING −0.02 [−0.76, 0.72]
SEEP 0.08 [−0.31, 0.46]
STRO 0.05 [−0.24, 0.34]
TRAC −0.72 [−1.58, 0.13]
UKBB −0.03 [−0.06, −0.01]
VAKN −0.18 [−0.42, 0.06]
VETS 0.15 [−0.14, 0.45]
WACH 0.07 [−0.83, 0.97]
WRBY −0.10 [−0.56, 0.37]
WTCS 0.00 [−0.25, 0.26]
F2.5 Meta −0.03 [−0.05, −0.01]
EHR
BIOM −0.14 [−0.42, 0.14]
DAI2 −0.04 [−0.08, −0.00]
FING −0.03 [−0.09, 0.02]
HUNT −0.04 [−0.13, 0.05]
MAYO −0.11 [−0.27, 0.05]
MGBB −0.06 [−0.13, 0.02]
SWED 0.07 [−0.21, 0.34]
UKB2 −0.02 [−0.07, 0.03]
EHR Meta −0.04 [−0.06, −0.01]
MVP
MVP1 −0.06 [−0.08, −0.03]
Freeze 3 Meta −0.04 [−0.06, −0.03]
−2
−1
0
1
2
Observed Outcome

rs175086 (Locus 26)

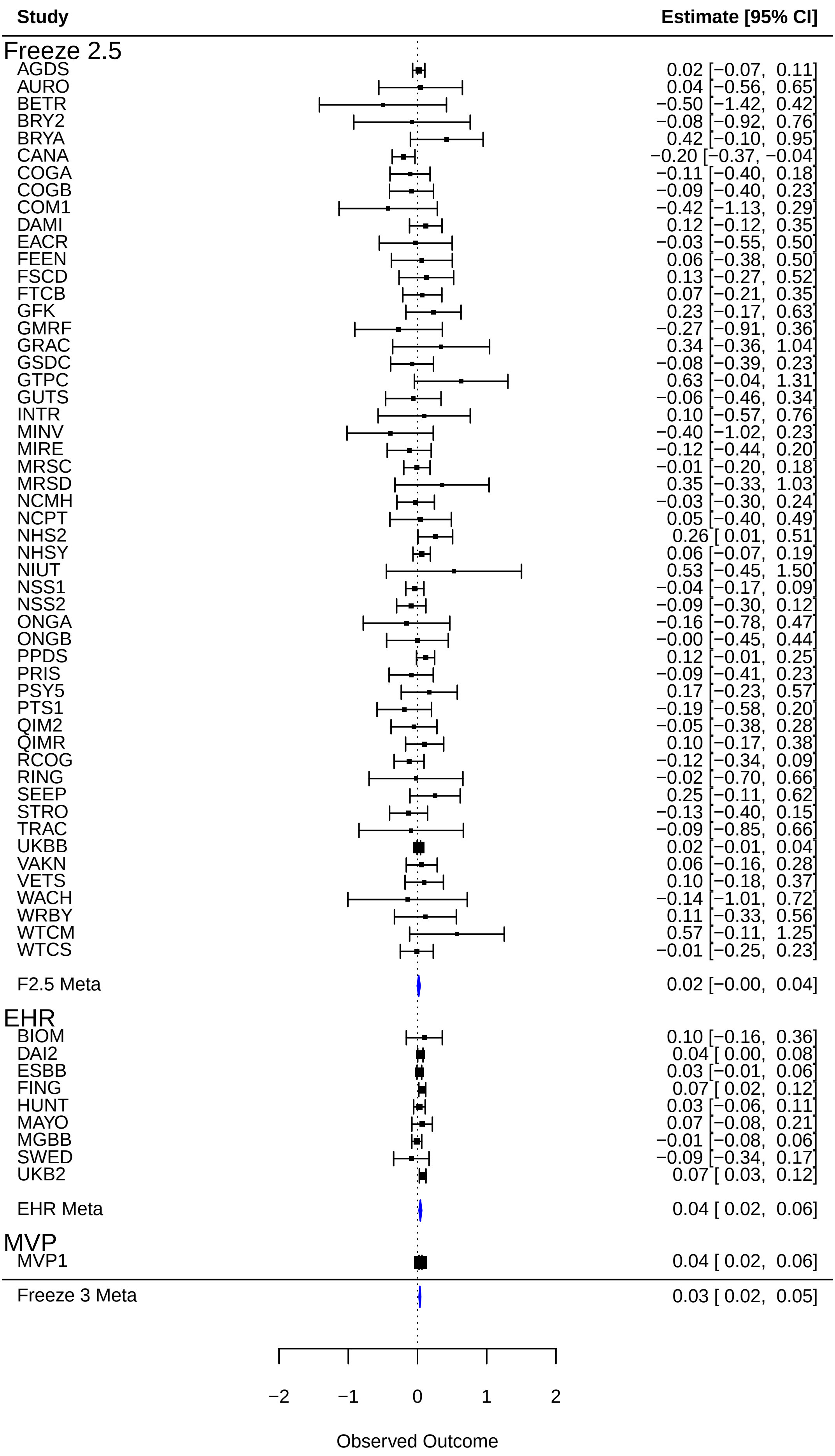

rs4257818 (Locus 27)

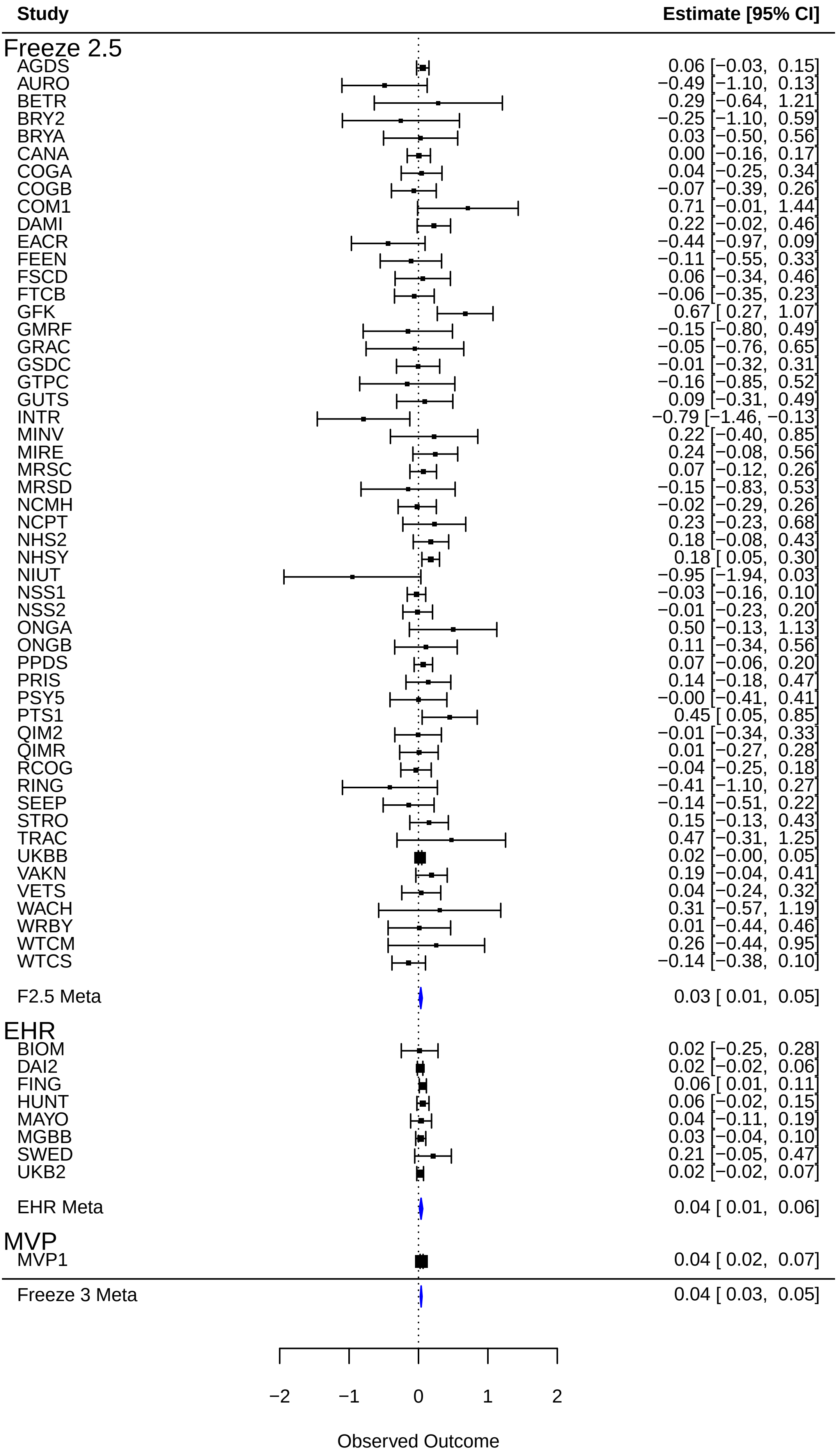

rs13168358 (Locus 28)

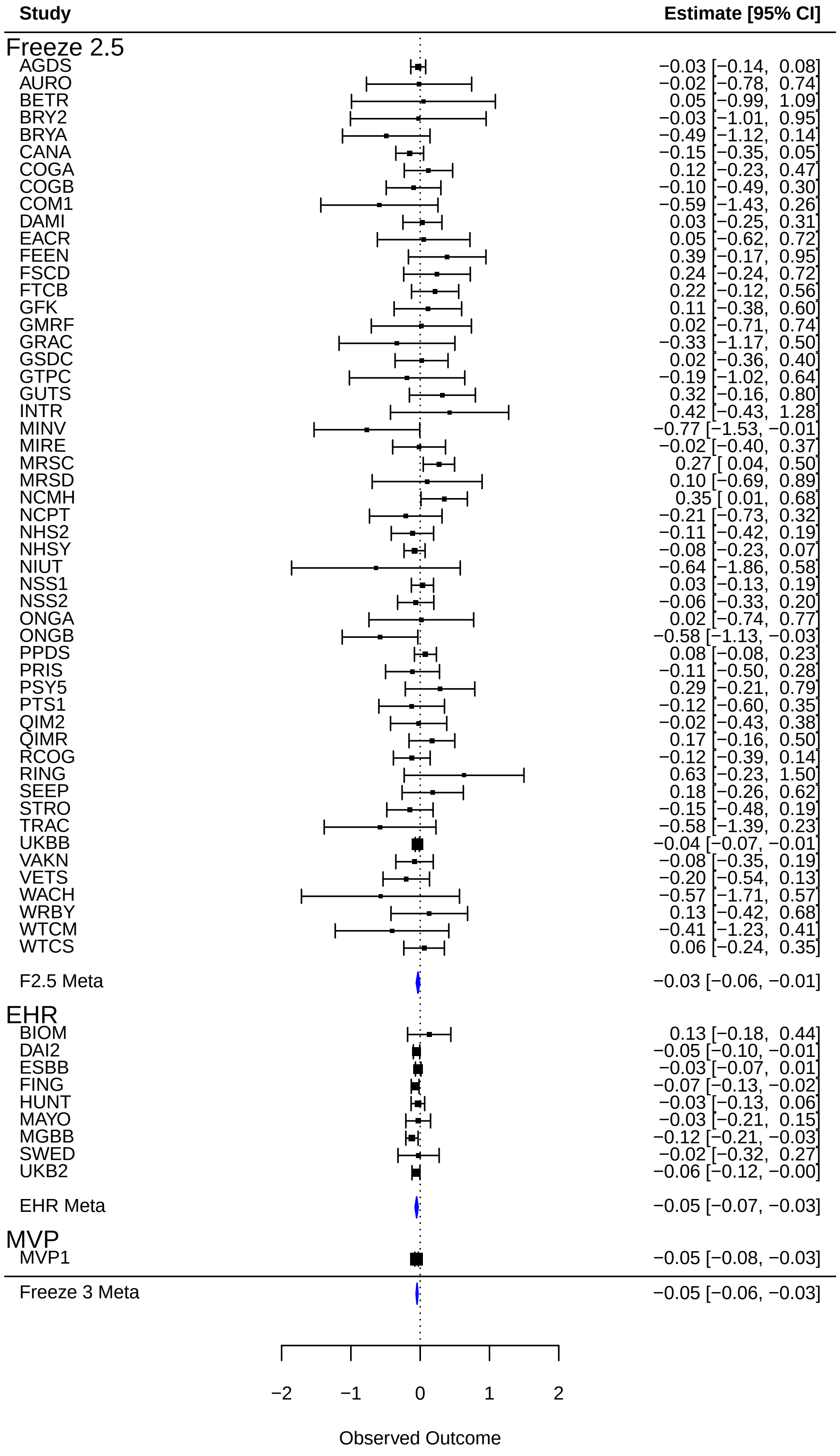

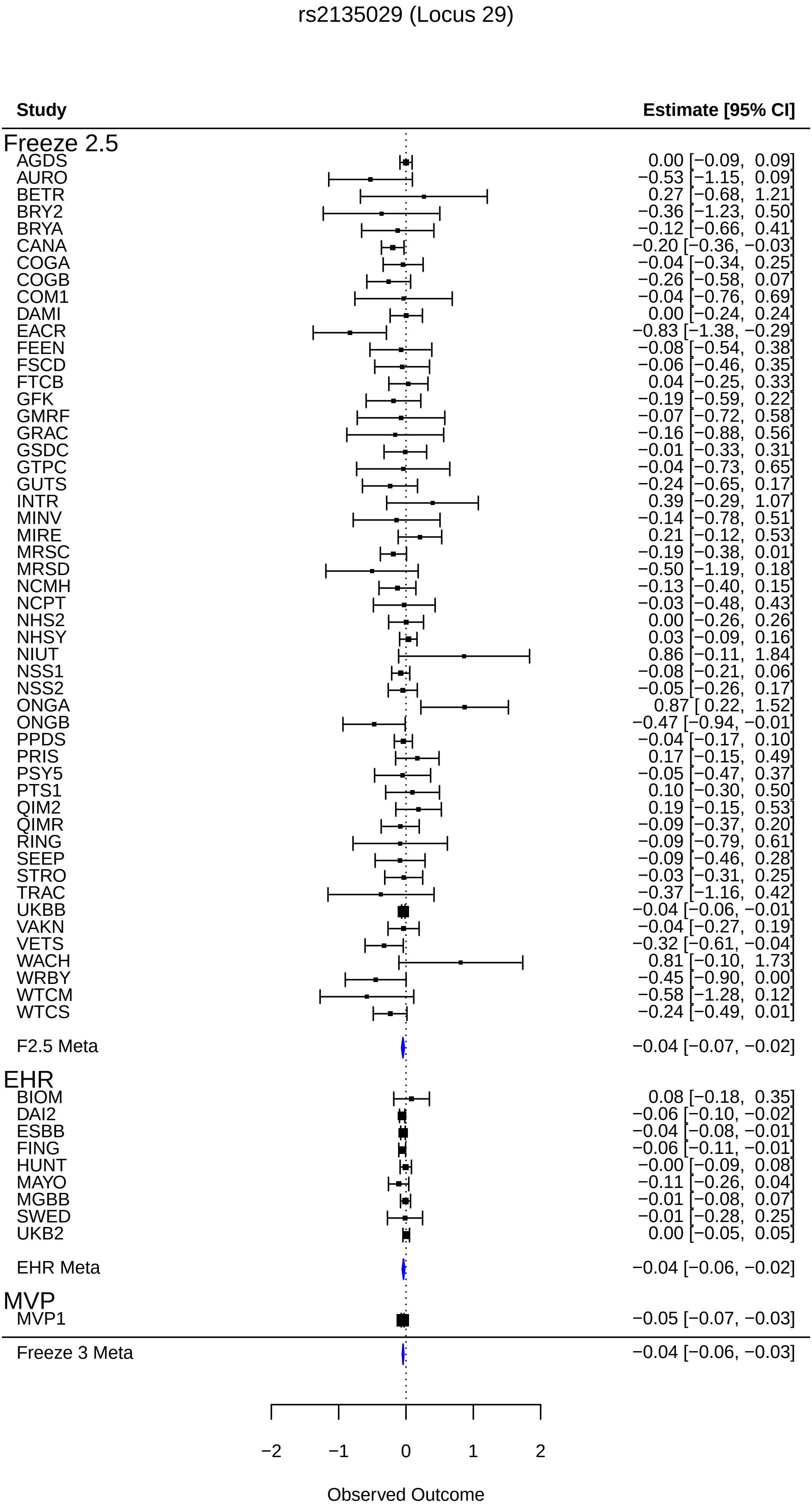
rs2135029 (Locus 29)
Study
Estimate [95% CI]
Freeze 2.5
AGDS 0.00 [−0.09, 0.09]
AURO −0.53 [−1.15, 0.09]
BETR 0.27 [−0.68, 1.21]
BRY2 −0.36 [−1.23, 0.50]
BRYA −0.12 [−0.66, 0.41]
CANA −0.20 [−0.36, −0.03]
COGA −0.04 [−0.34, 0.25]
COGB −0.26 [−0.58, 0.07]
COM1 −0.04 [−0.76, 0.69]
DAMI 0.00 [−0.24, 0.24]
EACR −0.83 [−1.38, −0.29]
FEEN −0.08 [−0.54, 0.38]
FSCD −0.06 [−0.46, 0.35]
FTCB 0.04 [−0.25, 0.33]
GFK −0.19 [−0.59, 0.22]
GMRF −0.07 [−0.72, 0.58]
GRAC −0.16 [−0.88, 0.56]
GSDC −0.01 [−0.33, 0.31]
GTPC −0.04 [−0.73, 0.65]
GUTS −0.24 [−0.65, 0.17]
INTR 0.39 [−0.29, 1.07]
MINV −0.14 [−0.78, 0.51]
MIRE 0.21 [−0.12, 0.53]
MRSC −0.19 [−0.38, 0.01]
MRSD −0.50 [−1.19, 0.18]
NCMH −0.13 [−0.40, 0.15]
NCPT −0.03 [−0.48, 0.43]
NHS2 0.00 [−0.26, 0.26]
NHSY 0.03 [−0.09, 0.16]
NIUT 0.86 [−0.11, 1.84]
NSS1 −0.08 [−0.21, 0.06]
NSS2 −0.05 [−0.26, 0.17]
ONGA 0.87 [ 0.22, 1.52]
ONGB −0.47 [−0.94, −0.01]
PPDS −0.04 [−0.17, 0.10]
PRIS 0.17 [−0.15, 0.49]
PSY5 −0.05 [−0.47, 0.37]
PTS1 0.10 [−0.30, 0.50]
QIM2 0.19 [−0.15, 0.53]
QIMR −0.09 [−0.37, 0.20]
RING −0.09 [−0.79, 0.61]
SEEP −0.09 [−0.46, 0.28]
STRO −0.03 [−0.31, 0.25]
TRAC −0.37 [−1.16, 0.42]
UKBB −0.04 [−0.06, −0.01]
VAKN −0.04 [−0.27, 0.19]
VETS −0.32 [−0.61, −0.04]
WACH 0.81 [−0.10, 1.73]
WRBY −0.45 [−0.90, 0.00]
WTCM −0.58 [−1.28, 0.12]
WTCS −0.24 [−0.49, 0.01]
F2.5 Meta −0.04 [−0.07, −0.02]
EHR
BIOM 0.08 [−0.18, 0.35]
DAI2 −0.06 [−0.10, −0.02]
ESBB −0.04 [−0.08, −0.01]
FING −0.06 [−0.11, −0.01]
HUNT −0.00 [−0.09, 0.08]
MAYO −0.11 [−0.26, 0.04]
MGBB −0.01 [−0.08, 0.07]
SWED −0.01 [−0.28, 0.25]
UKB2 0.00 [−0.05, 0.05]
EHR Meta −0.04 [−0.06, −0.02]
MVP
MVP1 −0.05 [−0.07, −0.03]
Freeze 3 Meta −0.04 [−0.06, −0.03]
−2
−1
0
1
2
Observed Outcome

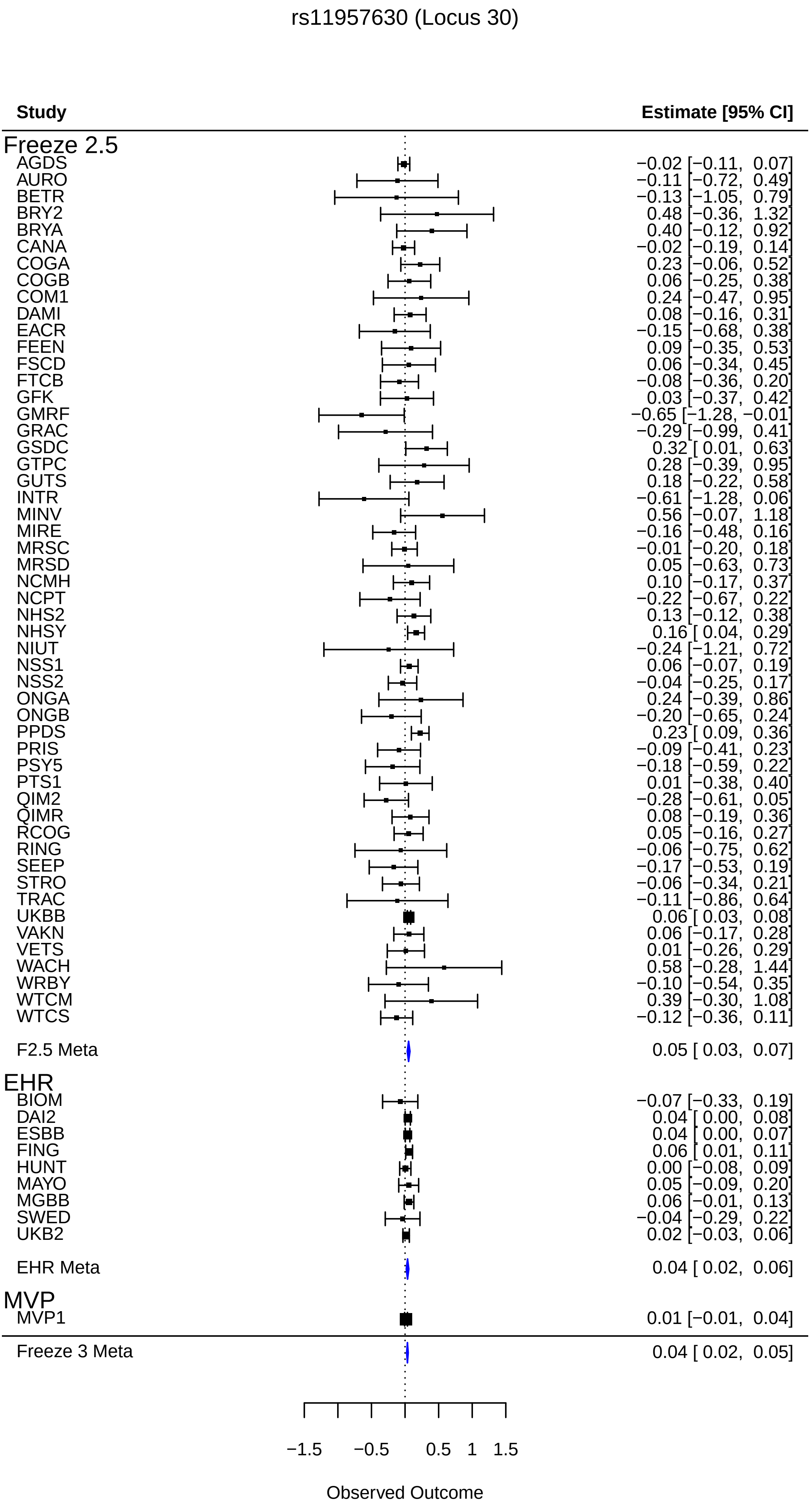
rs11957630 (Locus 30)
Study
Estimate [95% CI]
Freeze 2.5
AGDS −0.02 [−0.11, 0.07]
AURO −0.11 [−0.72, 0.49]
BETR −0.13 [−1.05, 0.79]
BRY2 0.48 [−0.36, 1.32]
BRYA 0.40 [−0.12, 0.92]
CANA −0.02 [−0.19, 0.14]
COGA 0.23 [−0.06, 0.52]
COGB 0.06 [−0.25, 0.38]
COM1 0.24 [−0.47, 0.95]
DAMI 0.08 [−0.16, 0.31]
EACR −0.15 [−0.68, 0.38]
FEEN 0.09 [−0.35, 0.53]
FSCD 0.06 [−0.34, 0.45]
FTCB −0.08 [−0.36, 0.20]
GFK 0.03 [−0.37, 0.42]
GMRF −0.65 [−1.28, −0.01]
GRAC −0.29 [−0.99, 0.41]
GSDC 0.32 [ 0.01, 0.63]
GTPC 0.28 [−0.39, 0.95]
GUTS 0.18 [−0.22, 0.58]
INTR −0.61 [−1.28, 0.06]
MINV 0.56 [−0.07, 1.18]
MIRE −0.16 [−0.48, 0.16]
MRSC −0.01 [−0.20, 0.18]
MRSD 0.05 [−0.63, 0.73]
NCMH 0.10 [−0.17, 0.37]
NCPT −0.22 [−0.67, 0.22]
NHS2 0.13 [−0.12, 0.38]
NHSY 0.16 [ 0.04, 0.29]
NIUT −0.24 [−1.21, 0.72]
NSS1 0.06 [−0.07, 0.19]
NSS2 −0.04 [−0.25, 0.17]
ONGA 0.24 [−0.39, 0.86]
ONGB −0.20 [−0.65, 0.24]
PPDS 0.23 [ 0.09, 0.36]
PRIS −0.09 [−0.41, 0.23]
PSY5 −0.18 [−0.59, 0.22]
PTS1 0.01 [−0.38, 0.40]
QIM2 −0.28 [−0.61, 0.05]
QIMR 0.08 [−0.19, 0.36]
RCOG 0.05 [−0.16, 0.27]
RING −0.06 [−0.75, 0.62]
SEEP −0.17 [−0.53, 0.19]
STRO −0.06 [−0.34, 0.21]
TRAC −0.11 [−0.86, 0.64]
UKBB 0.06 [ 0.03, 0.08]
VAKN 0.06 [−0.17, 0.28]
VETS 0.01 [−0.26, 0.29]
WACH 0.58 [−0.28, 1.44]
WRBY −0.10 [−0.54, 0.35]
WTCM 0.39 [−0.30, 1.08]
WTCS −0.12 [−0.36, 0.11]
F2.5 Meta 0.05 [ 0.03, 0.07]
EHR
BIOM −0.07 [−0.33, 0.19]
DAI2 0.04 [ 0.00, 0.08]
ESBB 0.04 [ 0.00, 0.07]
FING 0.06 [ 0.01, 0.11]
HUNT 0.00 [−0.08, 0.09]
MAYO 0.05 [−0.09, 0.20]
MGBB 0.06 [−0.01, 0.13]
SWED −0.04 [−0.29, 0.22]
UKB2 0.02 [−0.03, 0.06]
EHR Meta 0.04 [ 0.02, 0.06]
MVP
MVP1 0.01 [−0.01, 0.04]
Freeze 3 Meta 0.04 [ 0.02, 0.05]
−1.5
−0.5
0.5
1
1.5
Observed Outcome

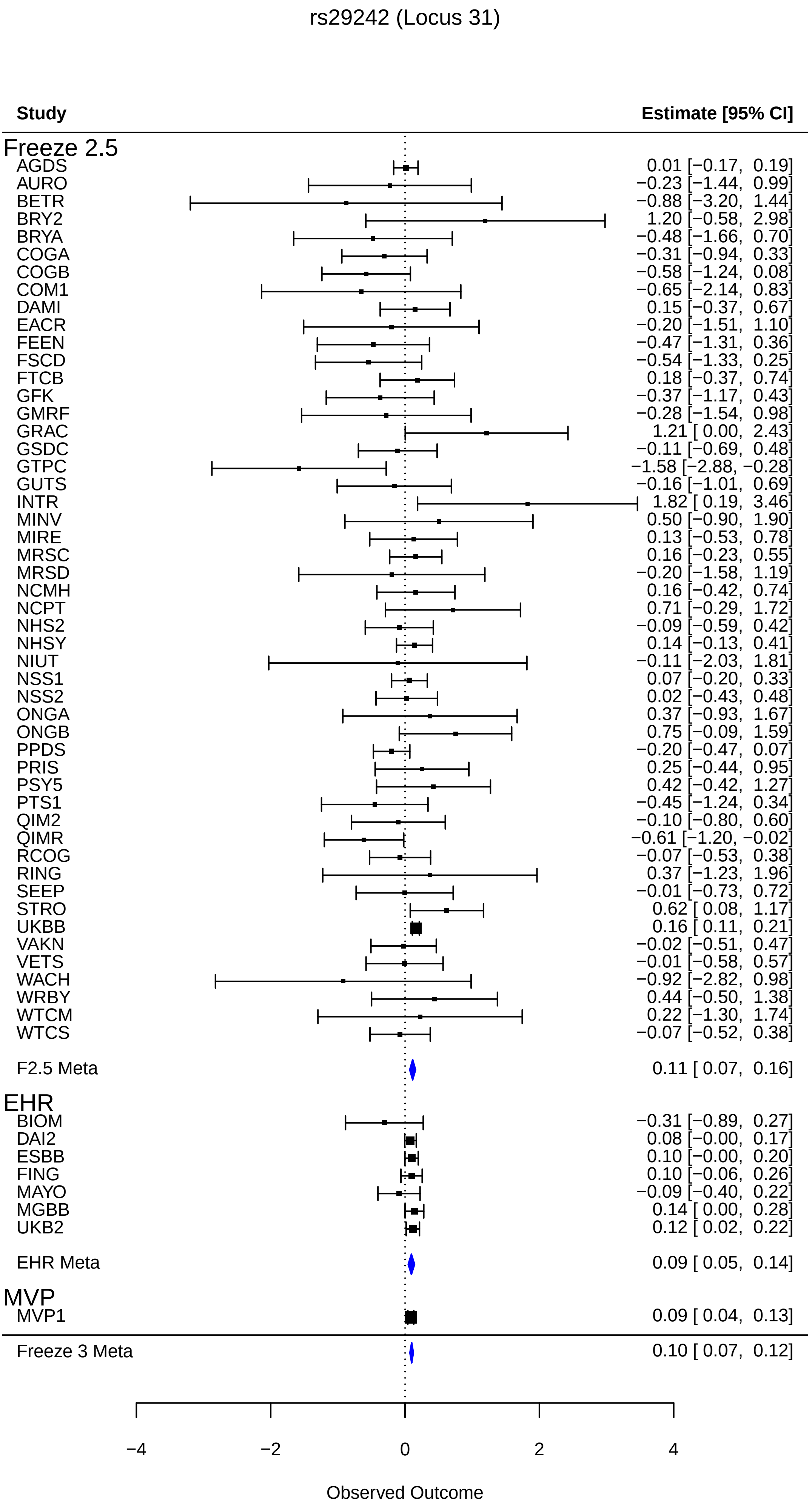
rs29242 (Locus 31)
Study
Estimate [95% CI]
Freeze 2.5
AGDS 0.01 [−0.17, 0.19]
AURO −0.23 [−1.44, 0.99]
BETR −0.88 [−3.20, 1.44]
BRY2 1.20 [−0.58, 2.98]
BRYA −0.48 [−1.66, 0.70]
COGA −0.31 [−0.94, 0.33]
COGB −0.58 [−1.24, 0.08]
COM1 −0.65 [−2.14, 0.83]
DAMI 0.15 [−0.37, 0.67]
EACR −0.20 [−1.51, 1.10]
FEEN −0.47 [−1.31, 0.36]
FSCD −0.54 [−1.33, 0.25]
FTCB 0.18 [−0.37, 0.74]
GFK −0.37 [−1.17, 0.43]
GMRF −0.28 [−1.54, 0.98]
GRAC 1.21 [ 0.00, 2.43]
GSDC −0.11 [−0.69, 0.48]
GTPC −1.58 [−2.88, −0.28]
GUTS −0.16 [−1.01, 0.69]
INTR 1.82 [ 0.19, 3.46]
MINV 0.50 [−0.90, 1.90]
MIRE 0.13 [−0.53, 0.78]
MRSC 0.16 [−0.23, 0.55]
MRSD −0.20 [−1.58, 1.19]
NCMH 0.16 [−0.42, 0.74]
NCPT 0.71 [−0.29, 1.72]
NHS2 −0.09 [−0.59, 0.42]
NHSY 0.14 [−0.13, 0.41]
NIUT −0.11 [−2.03, 1.81]
NSS1 0.07 [−0.20, 0.33]
NSS2 0.02 [−0.43, 0.48]
ONGA 0.37 [−0.93, 1.67]
ONGB 0.75 [−0.09, 1.59]
PPDS −0.20 [−0.47, 0.07]
PRIS 0.25 [−0.44, 0.95]
PSY5 0.42 [−0.42, 1.27]
PTS1 −0.45 [−1.24, 0.34]
QIM2 −0.10 [−0.80, 0.60]
QIMR −0.61 [−1.20, −0.02]
RCOG −0.07 [−0.53, 0.38]
RING 0.37 [−1.23, 1.96]
SEEP −0.01 [−0.73, 0.72]
STRO 0.62 [ 0.08, 1.17]
UKBB 0.16 [ 0.11, 0.21]
VAKN −0.02 [−0.51, 0.47]
VETS −0.01 [−0.58, 0.57]
WACH −0.92 [−2.82, 0.98]
WRBY 0.44 [−0.50, 1.38]
WTCM 0.22 [−1.30, 1.74]
WTCS −0.07 [−0.52, 0.38]
F2.5 Meta 0.11 [ 0.07, 0.16]
EHR
BIOM −0.31 [−0.89, 0.27]
DAI2 0.08 [−0.00, 0.17]
ESBB 0.10 [−0.00, 0.20]
FING 0.10 [−0.06, 0.26]
MAYO −0.09 [−0.40, 0.22]
MGBB 0.14 [ 0.00, 0.28]
UKB2 0.12 [ 0.02, 0.22]
EHR Meta 0.09 [ 0.05, 0.14]
MVP
MVP1 0.09 [ 0.04, 0.13]
Freeze 3 Meta 0.10 [ 0.07, 0.12]
−4
−2
0
2
4
Observed Outcome

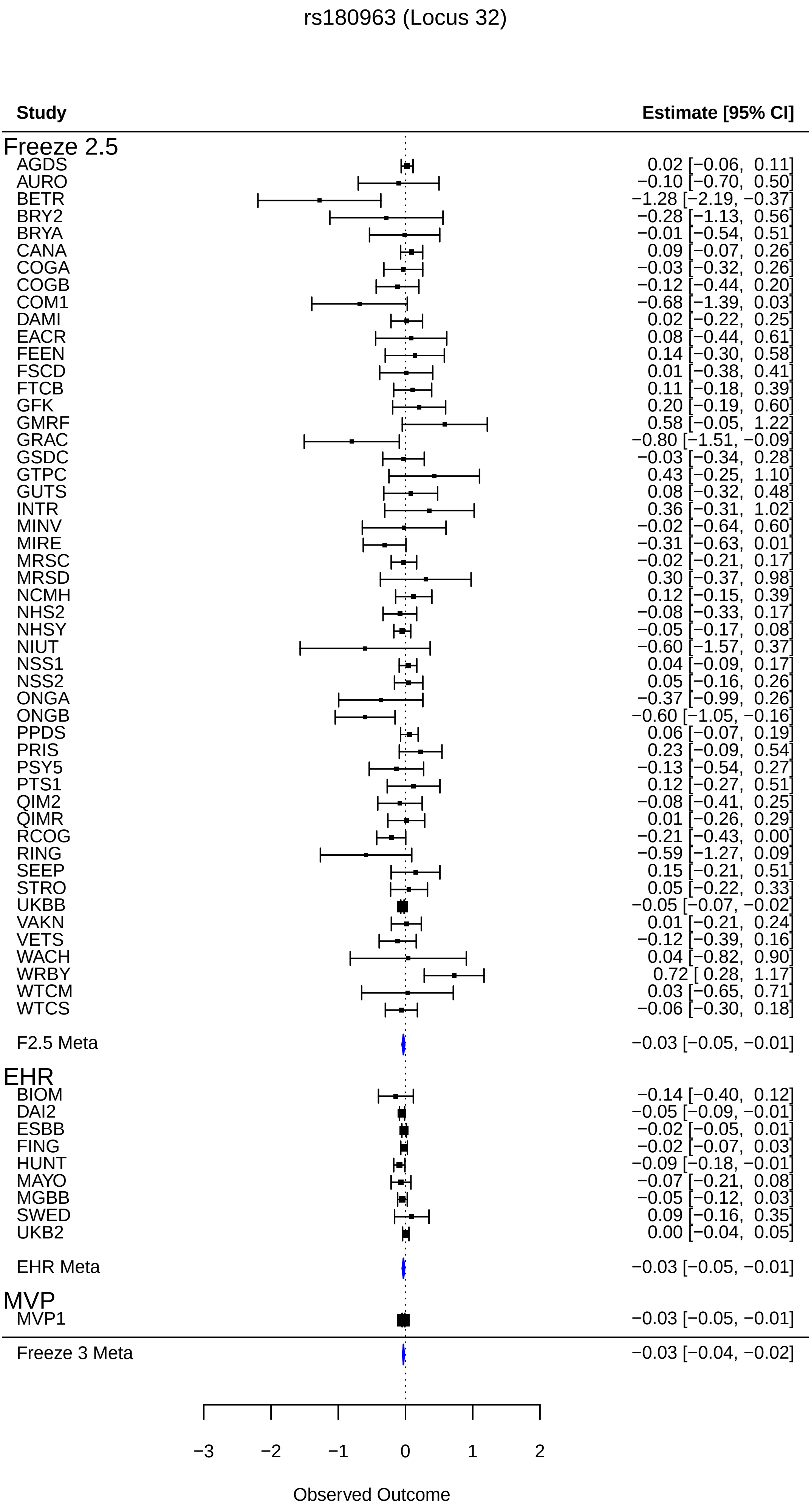
rs180963 (Locus 32)
Study
Estimate [95% CI]
Freeze 2.5
AGDS 0.02 [−0.06, 0.11]
AURO −0.10 [−0.70, 0.50]
BETR −1.28 [−2.19, −0.37]
BRY2 −0.28 [−1.13, 0.56]
BRYA −0.01 [−0.54, 0.51]
CANA 0.09 [−0.07, 0.26]
COGA −0.03 [−0.32, 0.26]
COGB −0.12 [−0.44, 0.20]
COM1 −0.68 [−1.39, 0.03]
DAMI 0.02 [−0.22, 0.25]
EACR 0.08 [−0.44, 0.61]
FEEN 0.14 [−0.30, 0.58]
FSCD 0.01 [−0.38, 0.41]
FTCB 0.11 [−0.18, 0.39]
GFK 0.20 [−0.19, 0.60]
GMRF 0.58 [−0.05, 1.22]
GRAC −0.80 [−1.51, −0.09]
GSDC −0.03 [−0.34, 0.28]
GTPC 0.43 [−0.25, 1.10]
GUTS 0.08 [−0.32, 0.48]
INTR 0.36 [−0.31, 1.02]
MINV −0.02 [−0.64, 0.60]
MIRE −0.31 [−0.63, 0.01]
MRSC −0.02 [−0.21, 0.17]
MRSD 0.30 [−0.37, 0.98]
NCMH 0.12 [−0.15, 0.39]
NHS2 −0.08 [−0.33, 0.17]
NHSY −0.05 [−0.17, 0.08]
NIUT −0.60 [−1.57, 0.37]
NSS1 0.04 [−0.09, 0.17]
NSS2 0.05 [−0.16, 0.26]
ONGA −0.37 [−0.99, 0.26]
ONGB −0.60 [−1.05, −0.16]
PPDS 0.06 [−0.07, 0.19]
PRIS 0.23 [−0.09, 0.54]
PSY5 −0.13 [−0.54, 0.27]
PTS1 0.12 [−0.27, 0.51]
QIM2 −0.08 [−0.41, 0.25]
QIMR 0.01 [−0.26, 0.29]
RCOG −0.21 [−0.43, 0.00]
RING −0.59 [−1.27, 0.09]
SEEP 0.15 [−0.21, 0.51]
STRO 0.05 [−0.22, 0.33]
UKBB −0.05 [−0.07, −0.02]
VAKN 0.01 [−0.21, 0.24]
VETS −0.12 [−0.39, 0.16]
WACH 0.04 [−0.82, 0.90]
WRBY 0.72 [ 0.28, 1.17]
WTCM 0.03 [−0.65, 0.71]
WTCS −0.06 [−0.30, 0.18]
F2.5 Meta −0.03 [−0.05, −0.01]
EHR
BIOM −0.14 [−0.40, 0.12]
DAI2 −0.05 [−0.09, −0.01]
ESBB −0.02 [−0.05, 0.01]
FING −0.02 [−0.07, 0.03]
HUNT −0.09 [−0.18, −0.01]
MAYO −0.07 [−0.21, 0.08]
MGBB −0.05 [−0.12, 0.03]
SWED 0.09 [−0.16, 0.35]
UKB2 0.00 [−0.04, 0.05]
EHR Meta −0.03 [−0.05, −0.01]
MVP
MVP1 −0.03 [−0.05, −0.01]
Freeze 3 Meta −0.03 [−0.04, −0.02]
−3
−2
−1
0
1
2
Observed Outcome

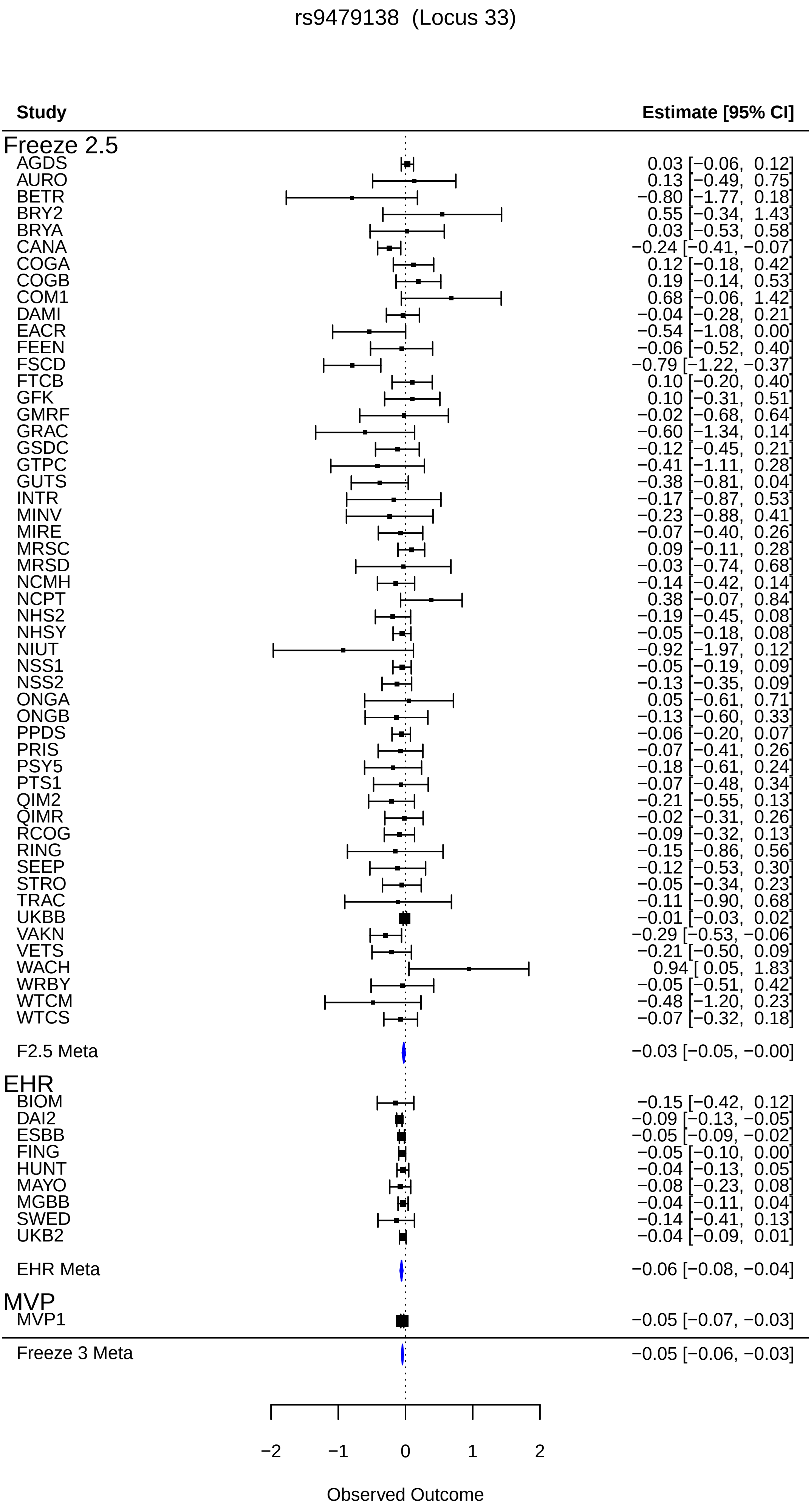
rs9479138 (Locus 33)
Study
Estimate [95% CI]
Freeze 2.5
AGDS 0.03 [−0.06, 0.12]
AURO 0.13 [−0.49, 0.75]
BETR −0.80 [−1.77, 0.18]
BRY2 0.55 [−0.34, 1.43]
BRYA 0.03 [−0.53, 0.58]
CANA −0.24 [−0.41, −0.07]
COGA 0.12 [−0.18, 0.42]
COGB 0.19 [−0.14, 0.53]
COM1 0.68 [−0.06, 1.42]
DAMI −0.04 [−0.28, 0.21]
EACR −0.54 [−1.08, 0.00]
FEEN −0.06 [−0.52, 0.40]
FSCD −0.79 [−1.22, −0.37]
FTCB 0.10 [−0.20, 0.40]
GFK 0.10 [−0.31, 0.51]
GMRF −0.02 [−0.68, 0.64]
GRAC −0.60 [−1.34, 0.14]
GSDC −0.12 [−0.45, 0.21]
GTPC −0.41 [−1.11, 0.28]
GUTS −0.38 [−0.81, 0.04]
INTR −0.17 [−0.87, 0.53]
MINV −0.23 [−0.88, 0.41]
MIRE −0.07 [−0.40, 0.26]
MRSC 0.09 [−0.11, 0.28]
MRSD −0.03 [−0.74, 0.68]
NCMH −0.14 [−0.42, 0.14]
NCPT 0.38 [−0.07, 0.84]
NHS2 −0.19 [−0.45, 0.08]
NHSY −0.05 [−0.18, 0.08]
NIUT −0.92 [−1.97, 0.12]
NSS1 −0.05 [−0.19, 0.09]
NSS2 −0.13 [−0.35, 0.09]
ONGA 0.05 [−0.61, 0.71]
ONGB −0.13 [−0.60, 0.33]
PPDS −0.06 [−0.20, 0.07]
PRIS −0.07 [−0.41, 0.26]
PSY5 −0.18 [−0.61, 0.24]
PTS1 −0.07 [−0.48, 0.34]
QIM2 −0.21 [−0.55, 0.13]
QIMR −0.02 [−0.31, 0.26]
RCOG −0.09 [−0.32, 0.13]
RING −0.15 [−0.86, 0.56]
SEEP −0.12 [−0.53, 0.30]
STRO −0.05 [−0.34, 0.23]
TRAC −0.11 [−0.90, 0.68]
UKBB −0.01 [−0.03, 0.02]
VAKN −0.29 [−0.53, −0.06]
VETS −0.21 [−0.50, 0.09]
WACH 0.94 [ 0.05, 1.83]
WRBY −0.05 [−0.51, 0.42]
WTCM −0.48 [−1.20, 0.23]
WTCS −0.07 [−0.32, 0.18]
F2.5 Meta −0.03 [−0.05, −0.00]
EHR
BIOM −0.15 [−0.42, 0.12]
DAI2 −0.09 [−0.13, −0.05]
ESBB −0.05 [−0.09, −0.02]
FING −0.05 [−0.10, 0.00]
HUNT −0.04 [−0.13, 0.05]
MAYO −0.08 [−0.23, 0.08]
MGBB −0.04 [−0.11, 0.04]
SWED −0.14 [−0.41, 0.13]
UKB2 −0.04 [−0.09, 0.01]
EHR Meta −0.06 [−0.08, −0.04]
MVP
MVP1 −0.05 [−0.07, −0.03]
Freeze 3 Meta −0.05 [−0.06, −0.03]
−2
−1
0
1
2
Observed Outcome

rs34809719 (Locus 34)

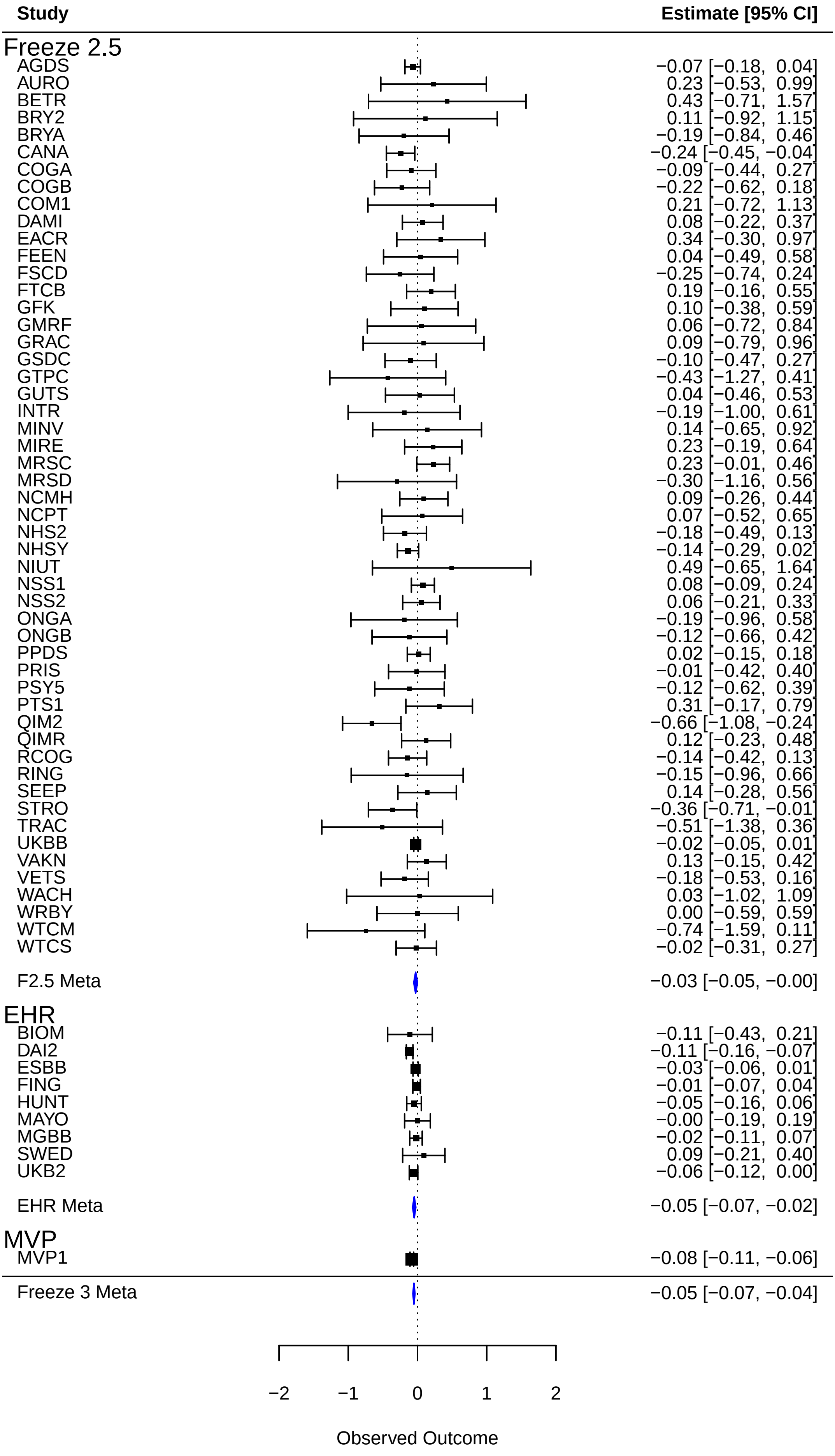

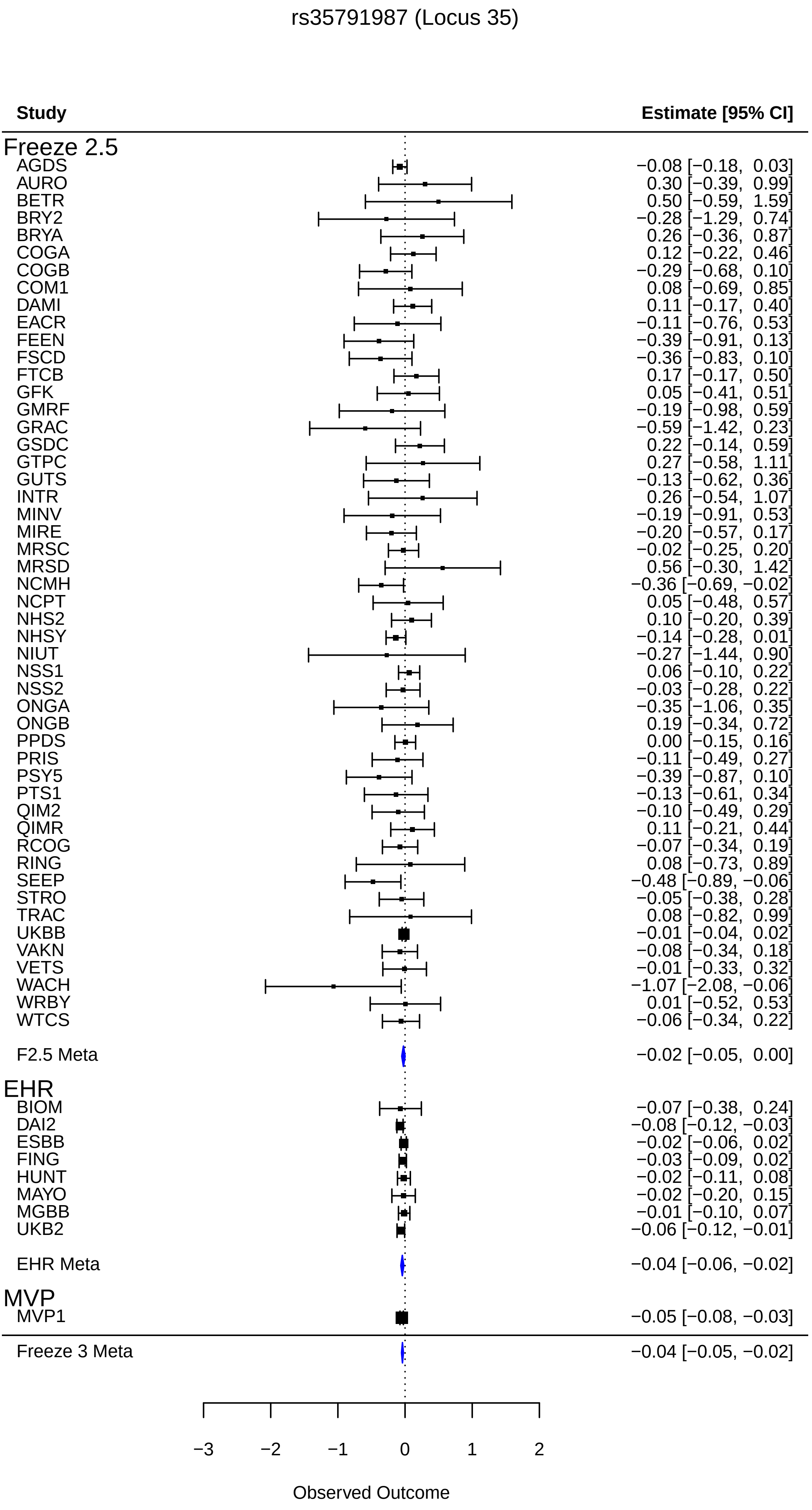
rs35791987 (Locus 35)
Study
Estimate [95% CI]
Freeze 2.5
AGDS −0.08 [−0.18, 0.03]
AURO 0.30 [−0.39, 0.99]
BETR 0.50 [−0.59, 1.59]
BRY2 −0.28 [−1.29, 0.74]
BRYA 0.26 [−0.36, 0.87]
COGA 0.12 [−0.22, 0.46]
COGB −0.29 [−0.68, 0.10]
COM1 0.08 [−0.69, 0.85]
DAMI 0.11 [−0.17, 0.40]
EACR −0.11 [−0.76, 0.53]
FEEN −0.39 [−0.91, 0.13]
FSCD −0.36 [−0.83, 0.10]
FTCB 0.17 [−0.17, 0.50]
GFK 0.05 [−0.41, 0.51]
GMRF −0.19 [−0.98, 0.59]
GRAC −0.59 [−1.42, 0.23]
GSDC 0.22 [−0.14, 0.59]
GTPC 0.27 [−0.58, 1.11]
GUTS −0.13 [−0.62, 0.36]
INTR 0.26 [−0.54, 1.07]
MINV −0.19 [−0.91, 0.53]
MIRE −0.20 [−0.57, 0.17]
MRSC −0.02 [−0.25, 0.20]
MRSD 0.56 [−0.30, 1.42]
NCMH −0.36 [−0.69, −0.02]
NCPT 0.05 [−0.48, 0.57]
NHS2 0.10 [−0.20, 0.39]
NHSY −0.14 [−0.28, 0.01]
NIUT −0.27 [−1.44, 0.90]
NSS1 0.06 [−0.10, 0.22]
NSS2 −0.03 [−0.28, 0.22]
ONGA −0.35 [−1.06, 0.35]
ONGB 0.19 [−0.34, 0.72]
PPDS 0.00 [−0.15, 0.16]
PRIS −0.11 [−0.49, 0.27]
PSY5 −0.39 [−0.87, 0.10]
PTS1 −0.13 [−0.61, 0.34]
QIM2 −0.10 [−0.49, 0.29]
QIMR 0.11 [−0.21, 0.44]
RCOG −0.07 [−0.34, 0.19]
RING 0.08 [−0.73, 0.89]
SEEP −0.48 [−0.89, −0.06]
STRO −0.05 [−0.38, 0.28]
TRAC 0.08 [−0.82, 0.99]
UKBB −0.01 [−0.04, 0.02]
VAKN −0.08 [−0.34, 0.18]
VETS −0.01 [−0.33, 0.32]
WACH −1.07 [−2.08, −0.06]
WRBY 0.01 [−0.52, 0.53]
WTCS −0.06 [−0.34, 0.22]
F2.5 Meta −0.02 [−0.05, 0.00]
EHR
BIOM −0.07 [−0.38, 0.24]
DAI2 −0.08 [−0.12, −0.03]
ESBB −0.02 [−0.06, 0.02]
FING −0.03 [−0.09, 0.02]
HUNT −0.02 [−0.11, 0.08]
MAYO −0.02 [−0.20, 0.15]
MGBB −0.01 [−0.10, 0.07]
UKB2 −0.06 [−0.12, −0.01]
EHR Meta −0.04 [−0.06, −0.02]
MVP
MVP1 −0.05 [−0.08, −0.03]
Freeze 3 Meta −0.04 [−0.05, −0.02]
−3
−2
−1
0
1
2
Observed Outcome

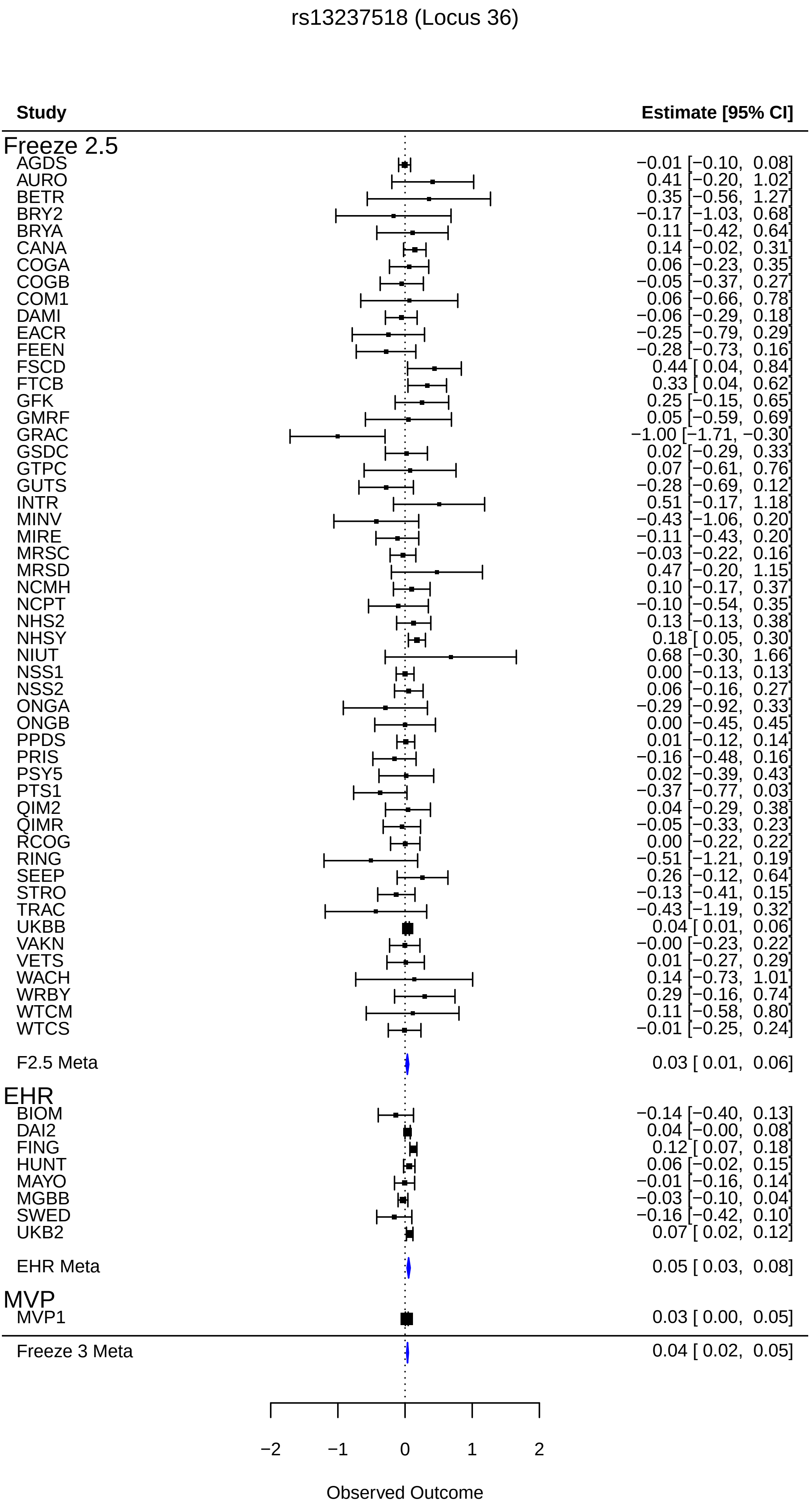

rs13237518 (Locus 36)
Study
Estimate [95% CI]
Freeze 2.5
AGDS −0.01 [−0.10, 0.08]
AURO 0.41 [−0.20, 1.02]
BETR 0.35 [−0.56, 1.27]
BRY2 −0.17 [−1.03, 0.68]
BRYA 0.11 [−0.42, 0.64]
CANA 0.14 [−0.02, 0.31]
COGA 0.06 [−0.23, 0.35]
COGB −0.05 [−0.37, 0.27]
COM1 0.06 [−0.66, 0.78]
DAMI −0.06 [−0.29, 0.18]
EACR −0.25 [−0.79, 0.29]
FEEN −0.28 [−0.73, 0.16]
FSCD 0.44 [ 0.04, 0.84]
FTCB 0.33 [ 0.04, 0.62]
GFK 0.25 [−0.15, 0.65]
GMRF 0.05 [−0.59, 0.69]
GRAC −1.00 [−1.71, −0.30]
GSDC 0.02 [−0.29, 0.33]
GTPC 0.07 [−0.61, 0.76]
GUTS −0.28 [−0.69, 0.12]
INTR 0.51 [−0.17, 1.18]
MINV −0.43 [−1.06, 0.20]
MIRE −0.11 [−0.43, 0.20]
MRSC −0.03 [−0.22, 0.16]
MRSD 0.47 [−0.20, 1.15]
NCMH 0.10 [−0.17, 0.37]
NCPT −0.10 [−0.54, 0.35]
NHS2 0.13 [−0.13, 0.38]
NHSY 0.18 [ 0.05, 0.30]
NIUT 0.68 [−0.30, 1.66]
NSS1 0.00 [−0.13, 0.13]
NSS2 0.06 [−0.16, 0.27]
ONGA −0.29 [−0.92, 0.33]
ONGB 0.00 [−0.45, 0.45]
PPDS 0.01 [−0.12, 0.14]
PRIS −0.16 [−0.48, 0.16]
PSY5 0.02 [−0.39, 0.43]
PTS1 −0.37 [−0.77, 0.03]
QIM2 0.04 [−0.29, 0.38]
QIMR −0.05 [−0.33, 0.23]
RCOG 0.00 [−0.22, 0.22]
RING −0.51 [−1.21, 0.19]
SEEP 0.26 [−0.12, 0.64]
STRO −0.13 [−0.41, 0.15]
TRAC −0.43 [−1.19, 0.32]
UKBB 0.04 [ 0.01, 0.06]
VAKN −0.00 [−0.23, 0.22]
VETS 0.01 [−0.27, 0.29]
WACH 0.14 [−0.73, 1.01]
WRBY 0.29 [−0.16, 0.74]
WTCM 0.11 [−0.58, 0.80]
WTCS −0.01 [−0.25, 0.24]
F2.5 Meta 0.03 [ 0.01, 0.06]
EHR
BIOM −0.14 [−0.40, 0.13]
DAI2 0.04 [−0.00, 0.08]
FING 0.12 [ 0.07, 0.18]
HUNT 0.06 [−0.02, 0.15]
MAYO −0.01 [−0.16, 0.14]
MGBB −0.03 [−0.10, 0.04]
SWED −0.16 [−0.42, 0.10]
UKB2 0.07 [ 0.02, 0.12]
EHR Meta 0.05 [ 0.03, 0.08]
MVP
MVP1 0.03 [ 0.00, 0.05]
Freeze 3 Meta 0.04 [ 0.02, 0.05]
−2
−1
0
1
2
Observed Outcome

rs2107448 (Locus 37)

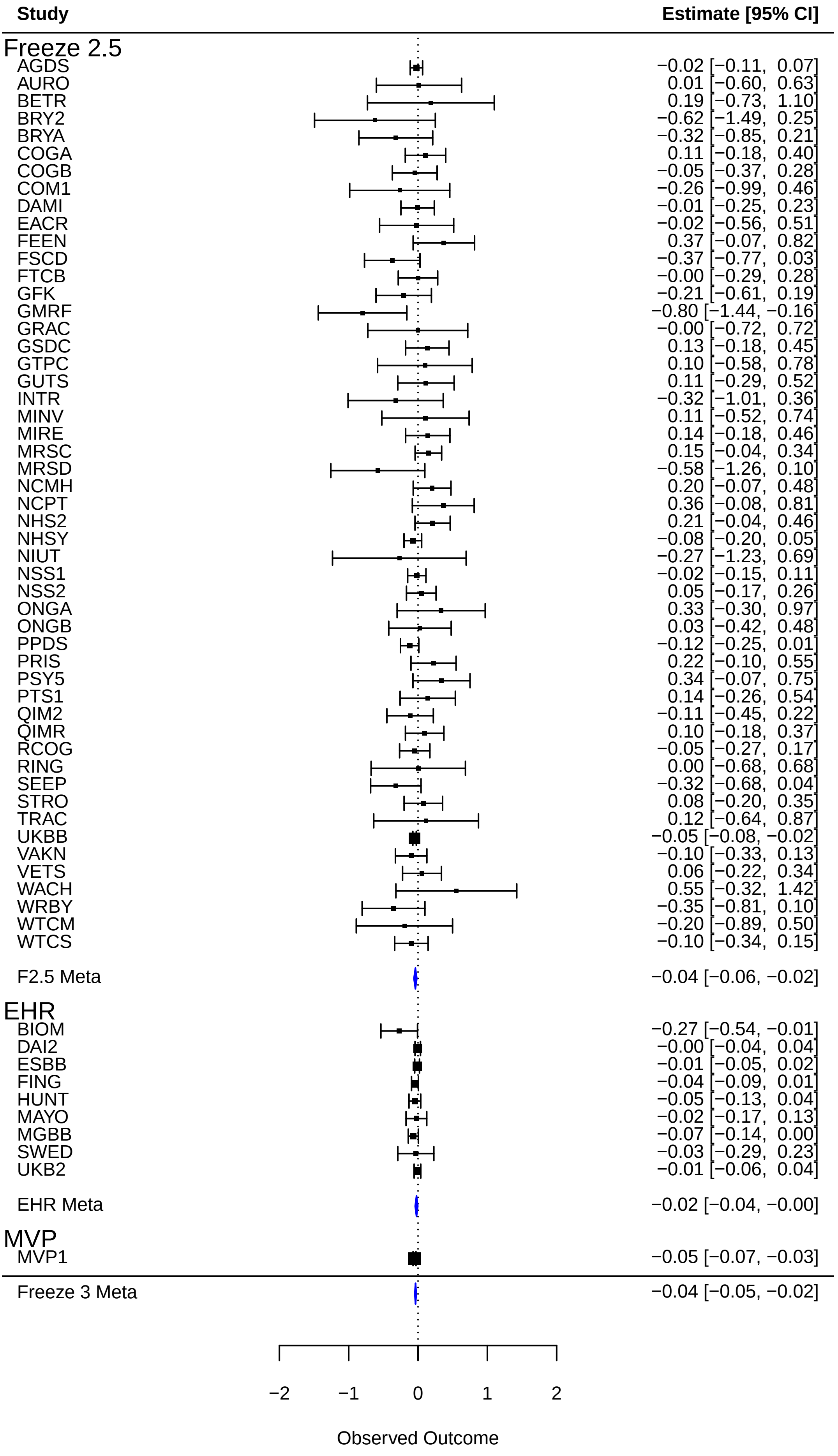

rs58043442 (Locus 38)

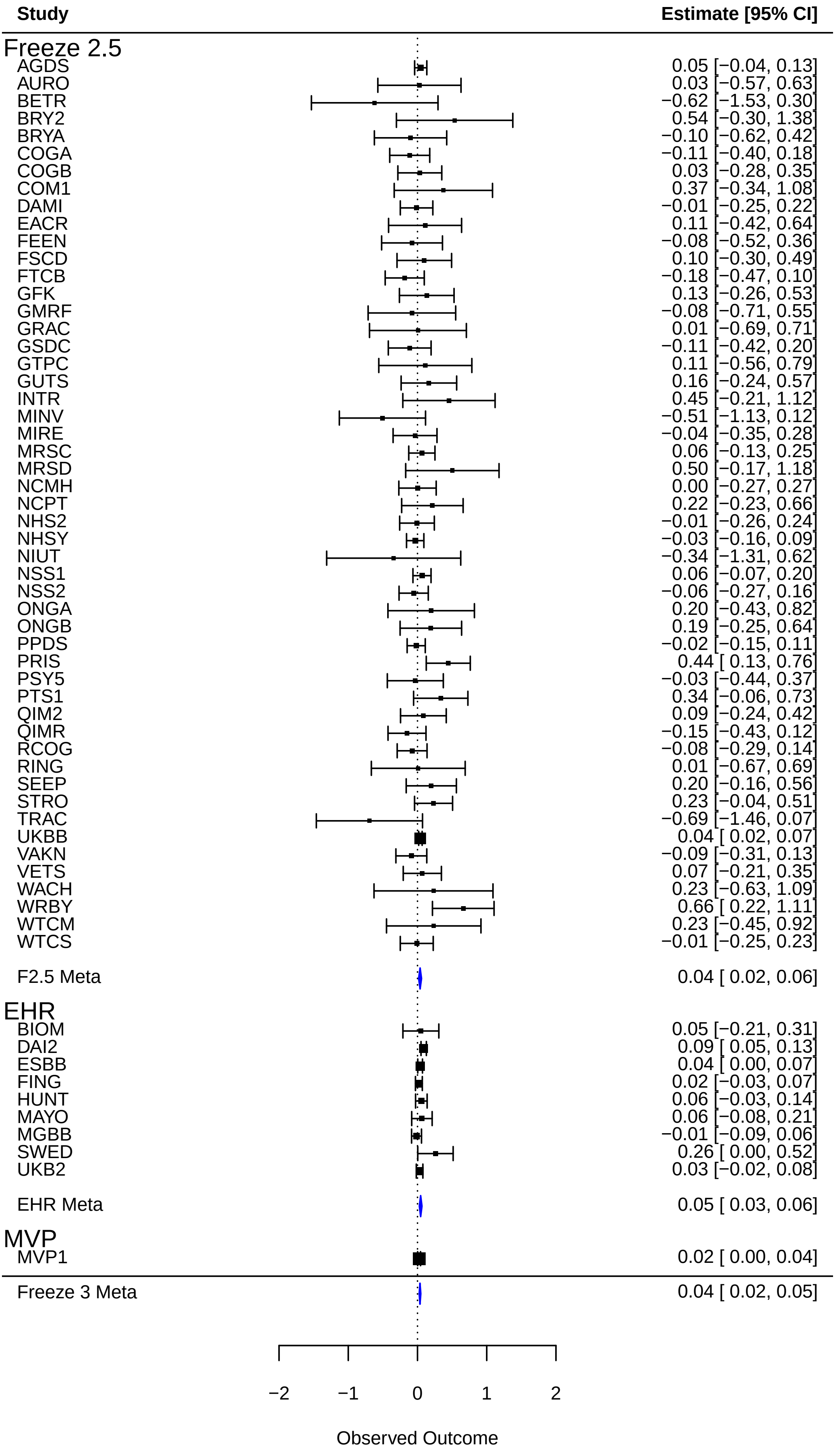

rs2470937 (Locus 39)

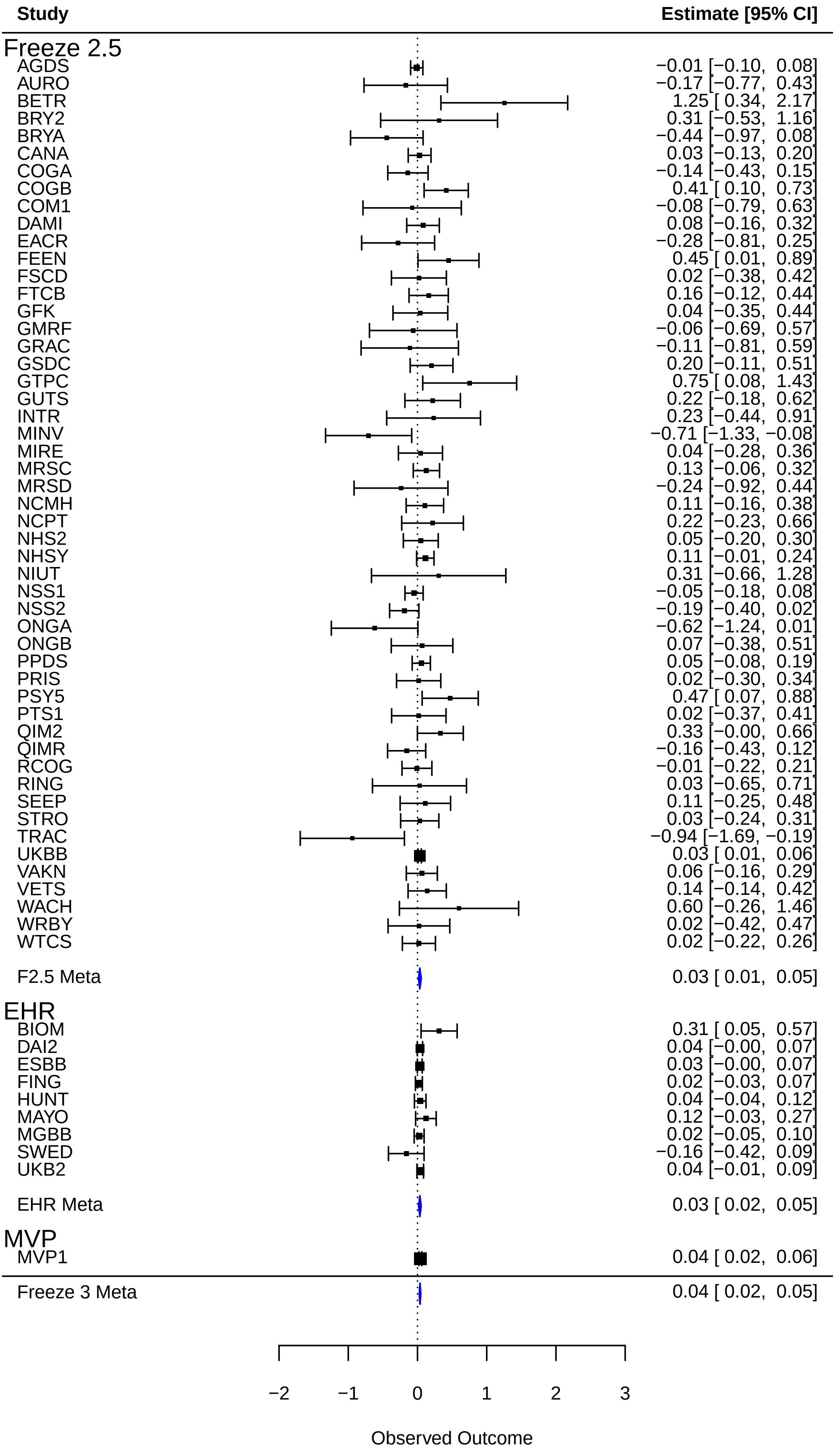

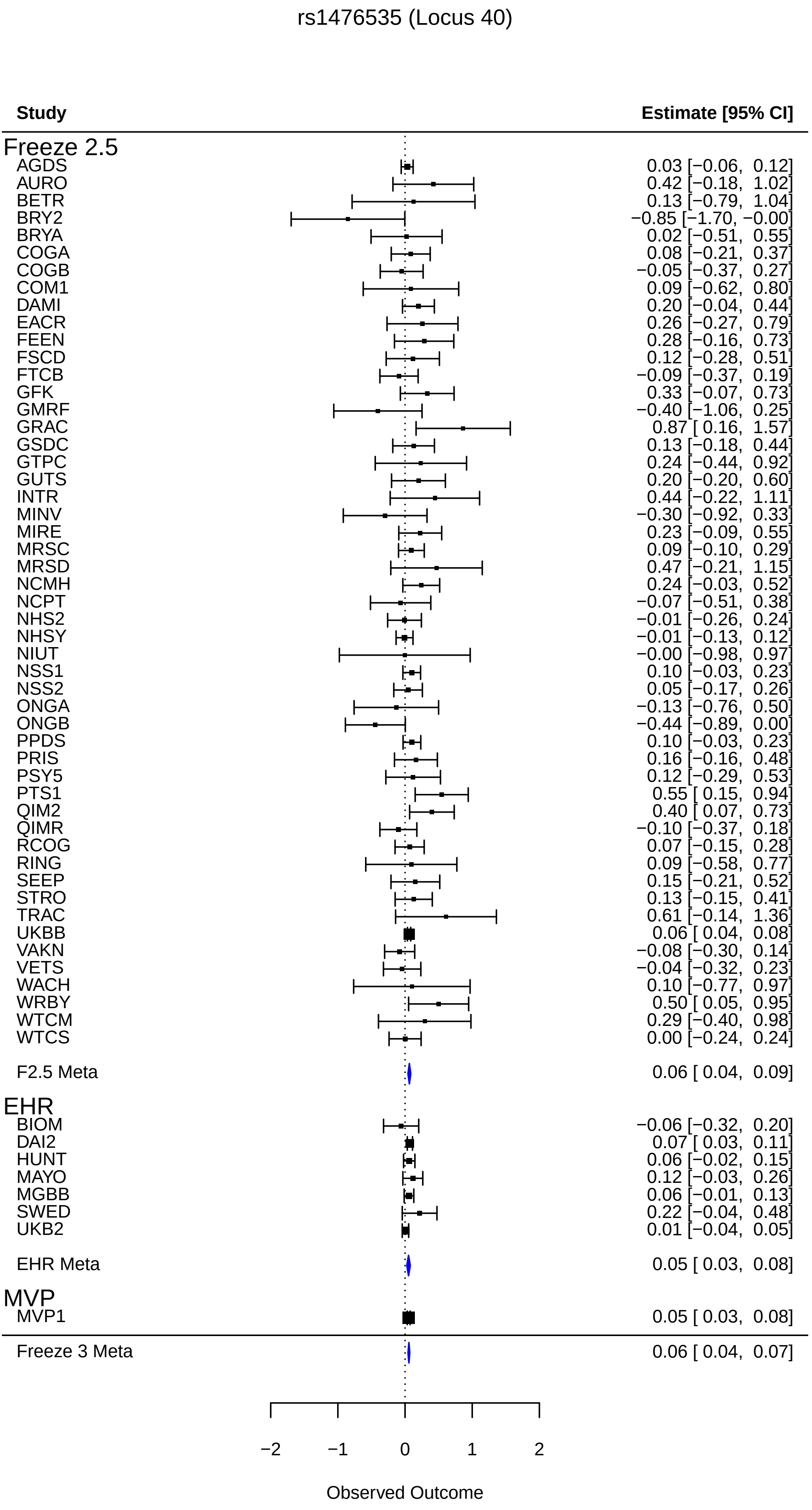
rs1476535 (Locus 40)
Study
Estimate [95% CI]
Freeze 2.5
AGDS 0.03 [−0.06, 0.12]
AURO 0.42 [−0.18, 1.02]
BETR 0.13 [−0.79, 1.04]
BRY2 −0.85 [−1.70, −0.00]
BRYA 0.02 [−0.51, 0.55]
COGA 0.08 [−0.21, 0.37]
COGB −0.05 [−0.37, 0.27]
COM1 0.09 [−0.62, 0.80]
DAMI 0.20 [−0.04, 0.44]
EACR 0.26 [−0.27, 0.79]
FEEN 0.28 [−0.16, 0.73]
FSCD 0.12 [−0.28, 0.51]
FTCB −0.09 [−0.37, 0.19]
GFK 0.33 [−0.07, 0.73]
GMRF −0.40 [−1.06, 0.25]
GRAC 0.87 [ 0.16, 1.57]
GSDC 0.13 [−0.18, 0.44]
GTPC 0.24 [−0.44, 0.92]
GUTS 0.20 [−0.20, 0.60]
INTR 0.44 [−0.22, 1.11]
MINV −0.30 [−0.92, 0.33]
MIRE 0.23 [−0.09, 0.55]
MRSC 0.09 [−0.10, 0.29]
MRSD 0.47 [−0.21, 1.15]
NCMH 0.24 [−0.03, 0.52]
NCPT −0.07 [−0.51, 0.38]
NHS2 −0.01 [−0.26, 0.24]
NHSY −0.01 [−0.13, 0.12]
NIUT −0.00 [−0.98, 0.97]
NSS1 0.10 [−0.03, 0.23]
NSS2 0.05 [−0.17, 0.26]
ONGA −0.13 [−0.76, 0.50]
ONGB −0.44 [−0.89, 0.00]
PPDS 0.10 [−0.03, 0.23]
PRIS 0.16 [−0.16, 0.48]
PSY5 0.12 [−0.29, 0.53]
PTS1 0.55 [ 0.15, 0.94]
QIM2 0.40 [ 0.07, 0.73]
QIMR −0.10 [−0.37, 0.18]
RCOG 0.07 [−0.15, 0.28]
RING 0.09 [−0.58, 0.77]
SEEP 0.15 [−0.21, 0.52]
STRO 0.13 [−0.15, 0.41]
TRAC 0.61 [−0.14, 1.36]
UKBB 0.06 [ 0.04, 0.08]
VAKN −0.08 [−0.30, 0.14]
VETS −0.04 [−0.32, 0.23]
WACH 0.10 [−0.77, 0.97]
WRBY 0.50 [ 0.05, 0.95]
WTCM 0.29 [−0.40, 0.98]
WTCS 0.00 [−0.24, 0.24]
F2.5 Meta 0.06 [ 0.04, 0.09]
EHR
BIOM −0.06 [−0.32, 0.20]
DAI2 0.07 [ 0.03, 0.11]
HUNT 0.06 [−0.02, 0.15]
MAYO 0.12 [−0.03, 0.26]
MGBB 0.06 [−0.01, 0.13]
SWED 0.22 [−0.04, 0.48]
UKB2 0.01 [−0.04, 0.05]
EHR Meta 0.05 [ 0.03, 0.08]
MVP
MVP1 0.05 [ 0.03, 0.08]
Freeze 3 Meta 0.06 [ 0.04, 0.07]
−2
−1
0
1
2
Observed Outcome

rs7806900 (Locus 41)

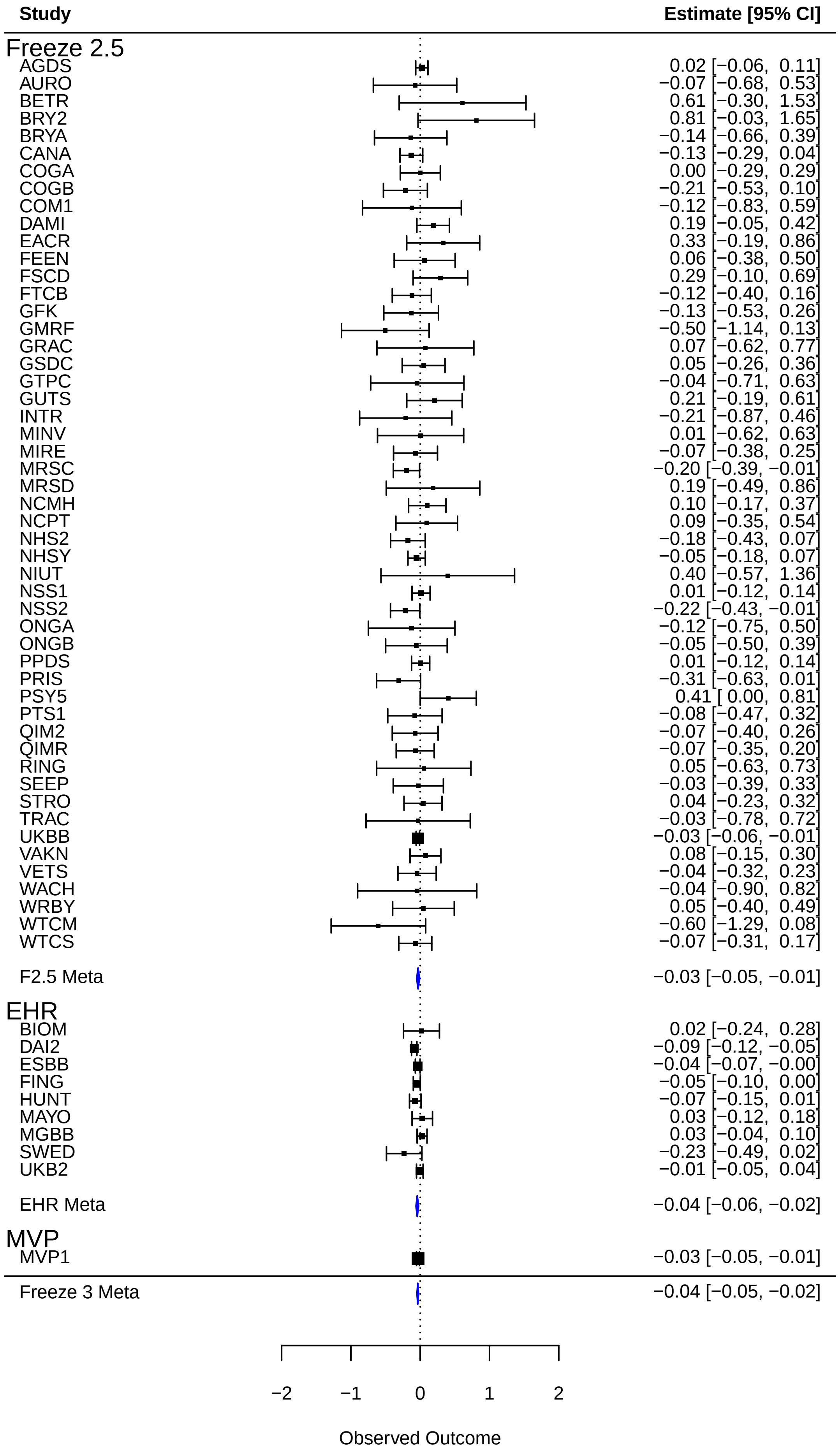

rs61702433 (Locus 42)

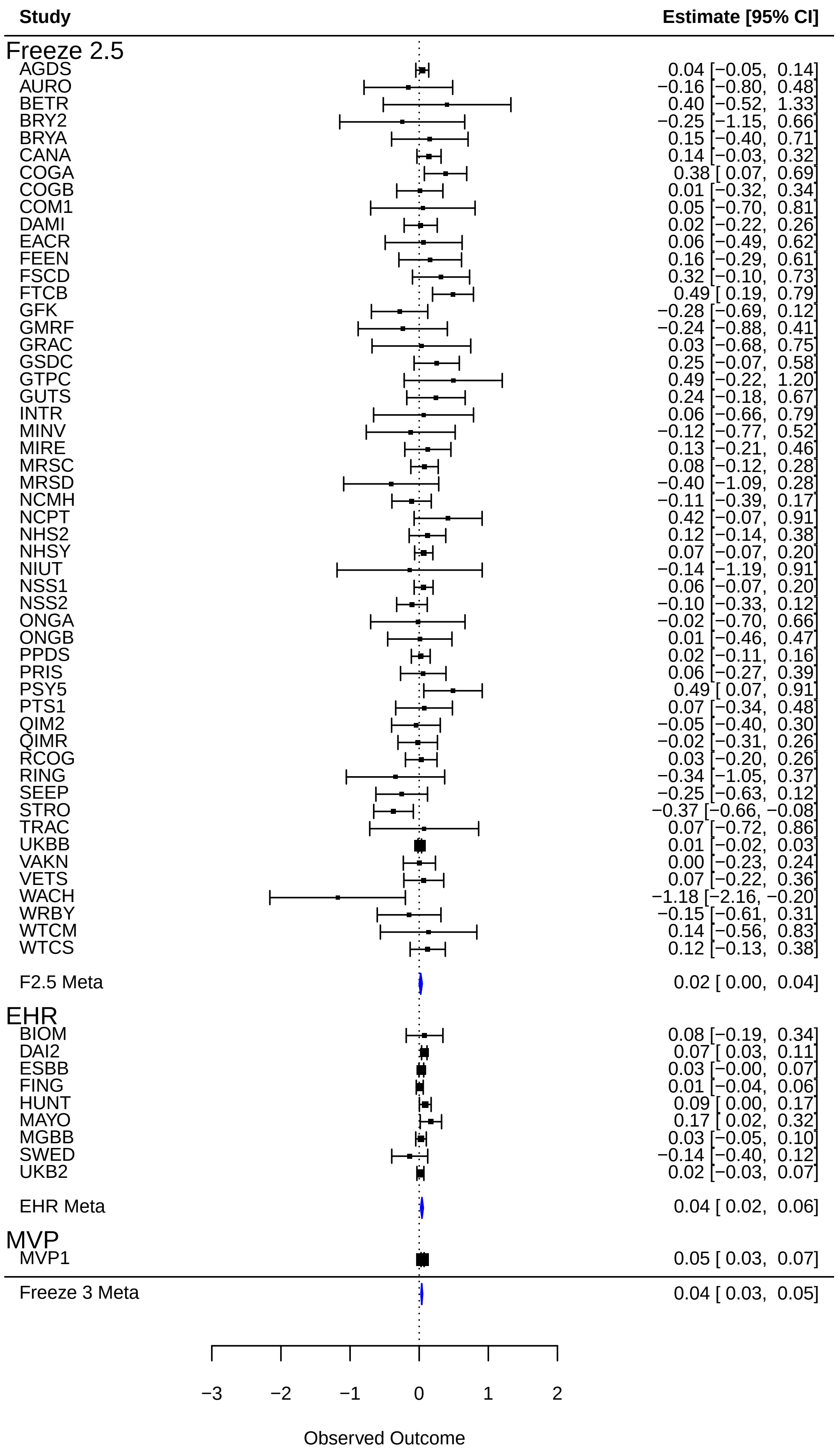

rs10487459 (Locus 43)

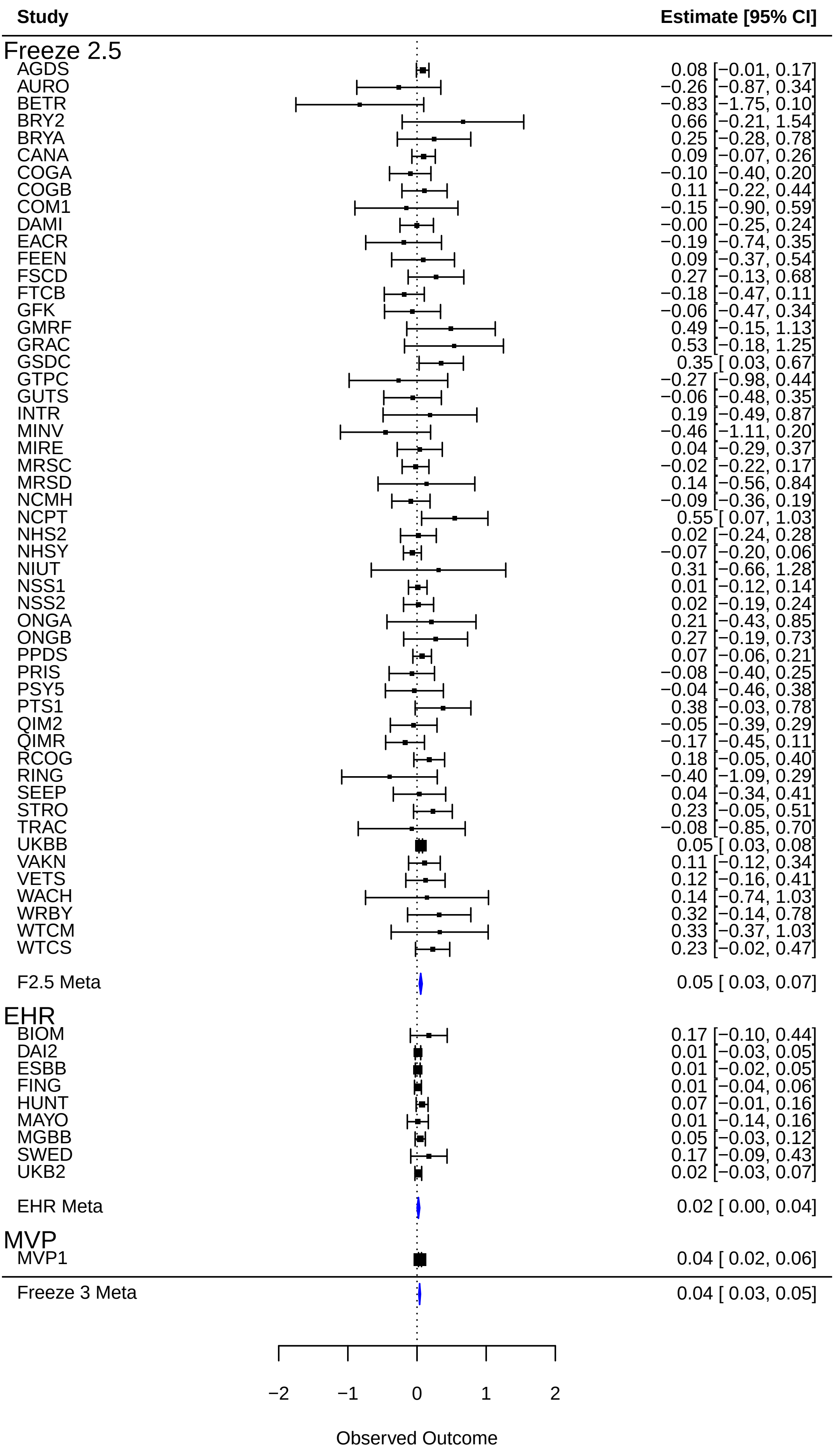

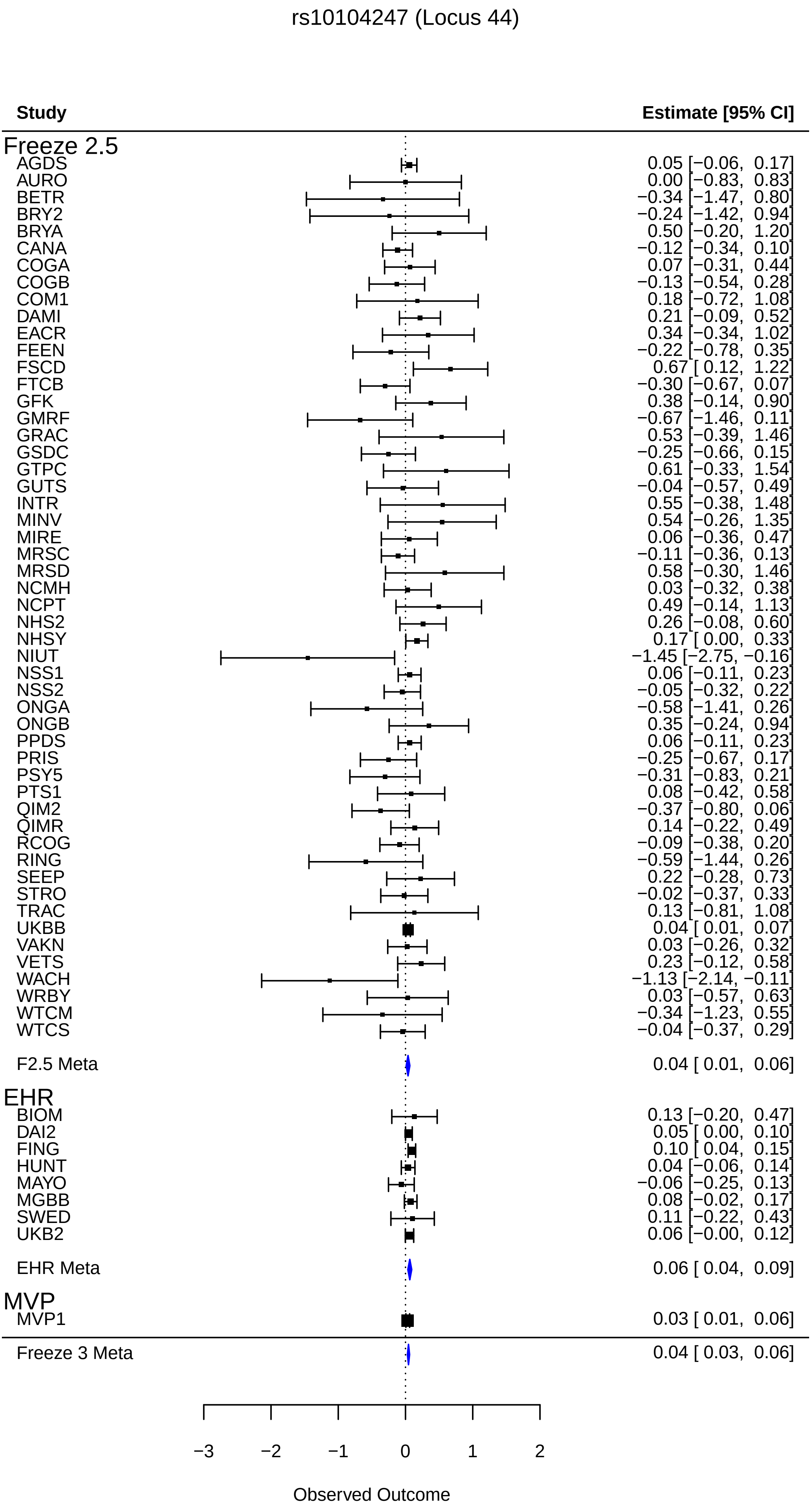
rs10104247 (Locus 44)
Study
Estimate [95% CI]
Freeze 2.5
AGDS 0.05 [−0.06, 0.17]
AURO 0.00 [−0.83, 0.83]
BETR −0.34 [−1.47, 0.80]
BRY2 −0.24 [−1.42, 0.94]
BRYA 0.50 [−0.20, 1.20]
CANA −0.12 [−0.34, 0.10]
COGA 0.07 [−0.31, 0.44]
COGB −0.13 [−0.54, 0.28]
COM1 0.18 [−0.72, 1.08]
DAMI 0.21 [−0.09, 0.52]
EACR 0.34 [−0.34, 1.02]
FEEN −0.22 [−0.78, 0.35]
FSCD 0.67 [ 0.12, 1.22]
FTCB −0.30 [−0.67, 0.07]
GFK 0.38 [−0.14, 0.90]
GMRF −0.67 [−1.46, 0.11]
GRAC 0.53 [−0.39, 1.46]
GSDC −0.25 [−0.66, 0.15]
GTPC 0.61 [−0.33, 1.54]
GUTS −0.04 [−0.57, 0.49]
INTR 0.55 [−0.38, 1.48]
MINV 0.54 [−0.26, 1.35]
MIRE 0.06 [−0.36, 0.47]
MRSC −0.11 [−0.36, 0.13]
MRSD 0.58 [−0.30, 1.46]
NCMH 0.03 [−0.32, 0.38]
NCPT 0.49 [−0.14, 1.13]
NHS2 0.26 [−0.08, 0.60]
NHSY 0.17 [ 0.00, 0.33]
NIUT −1.45 [−2.75, −0.16]
NSS1 0.06 [−0.11, 0.23]
NSS2 −0.05 [−0.32, 0.22]
ONGA −0.58 [−1.41, 0.26]
ONGB 0.35 [−0.24, 0.94]
PPDS 0.06 [−0.11, 0.23]
PRIS −0.25 [−0.67, 0.17]
PSY5 −0.31 [−0.83, 0.21]
PTS1 0.08 [−0.42, 0.58]
QIM2 −0.37 [−0.80, 0.06]
QIMR 0.14 [−0.22, 0.49]
RCOG −0.09 [−0.38, 0.20]
RING −0.59 [−1.44, 0.26]
SEEP 0.22 [−0.28, 0.73]
STRO −0.02 [−0.37, 0.33]
TRAC 0.13 [−0.81, 1.08]
UKBB 0.04 [ 0.01, 0.07]
VAKN 0.03 [−0.26, 0.32]
VETS 0.23 [−0.12, 0.58]
WACH −1.13 [−2.14, −0.11]
WRBY 0.03 [−0.57, 0.63]
WTCM −0.34 [−1.23, 0.55]
WTCS −0.04 [−0.37, 0.29]
F2.5 Meta 0.04 [ 0.01, 0.06]
EHR
BIOM 0.13 [−0.20, 0.47]
DAI2 0.05 [ 0.00, 0.10]
FING 0.10 [ 0.04, 0.15]
HUNT 0.04 [−0.06, 0.14]
MAYO −0.06 [−0.25, 0.13]
MGBB 0.08 [−0.02, 0.17]
SWED 0.11 [−0.22, 0.43]
UKB2 0.06 [−0.00, 0.12]
EHR Meta 0.06 [ 0.04, 0.09]
MVP
MVP1 0.03 [ 0.01, 0.06]
Freeze 3 Meta 0.04 [ 0.03, 0.06]
−3
−2
−1
0
1
2
Observed Outcome

rs4129585 (Locus 45)

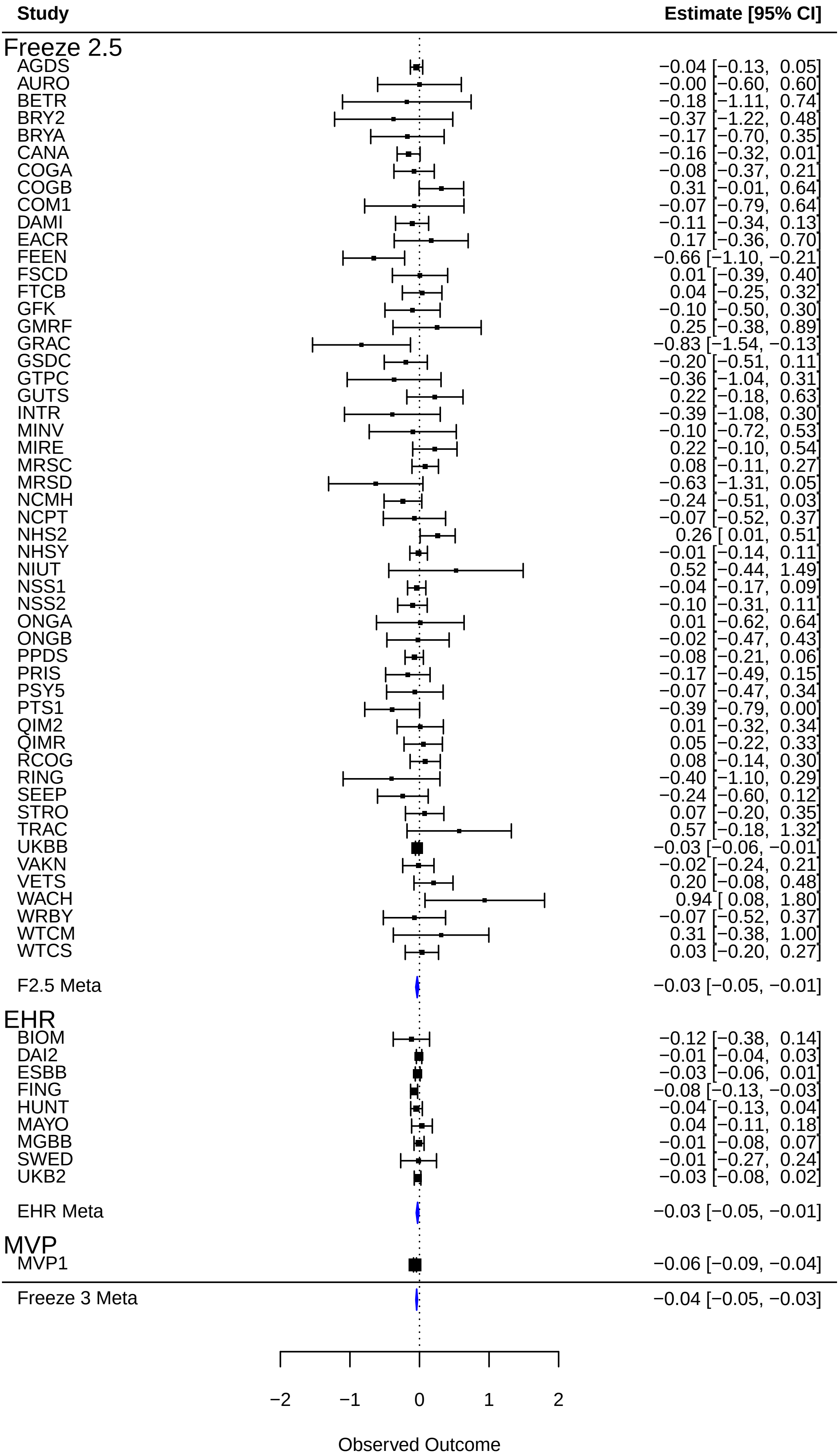

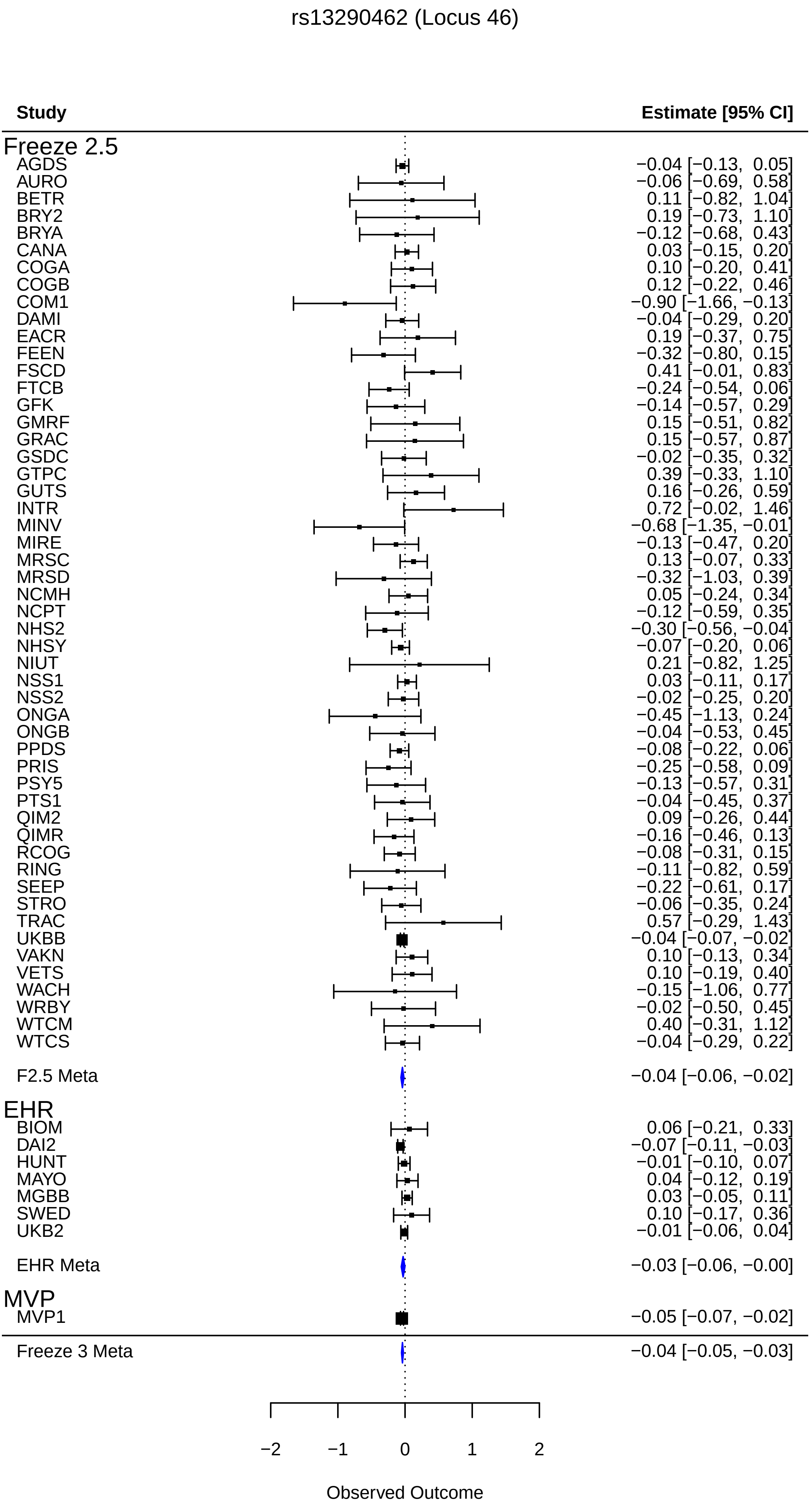

rs13290462 (Locus 46)
Study
Estimate [95% CI]
Freeze 2.5
AGDS −0.04 [−0.13, 0.05]
AURO −0.06 [−0.69, 0.58]
BETR 0.11 [−0.82, 1.04]
BRY2 0.19 [−0.73, 1.10]
BRYA −0.12 [−0.68, 0.43]
CANA 0.03 [−0.15, 0.20]
COGA 0.10 [−0.20, 0.41]
COGB 0.12 [−0.22, 0.46]
COM1 −0.90 [−1.66, −0.13]
DAMI −0.04 [−0.29, 0.20]
EACR 0.19 [−0.37, 0.75]
FEEN −0.32 [−0.80, 0.15]
FSCD 0.41 [−0.01, 0.83]
FTCB −0.24 [−0.54, 0.06]
GFK −0.14 [−0.57, 0.29]
GMRF 0.15 [−0.51, 0.82]
GRAC 0.15 [−0.57, 0.87]
GSDC −0.02 [−0.35, 0.32]
GTPC 0.39 [−0.33, 1.10]
GUTS 0.16 [−0.26, 0.59]
INTR 0.72 [−0.02, 1.46]
MINV −0.68 [−1.35, −0.01]
MIRE −0.13 [−0.47, 0.20]
MRSC 0.13 [−0.07, 0.33]
MRSD −0.32 [−1.03, 0.39]
NCMH 0.05 [−0.24, 0.34]
NCPT −0.12 [−0.59, 0.35]
NHS2 −0.30 [−0.56, −0.04]
NHSY −0.07 [−0.20, 0.06]
NIUT 0.21 [−0.82, 1.25]
NSS1 0.03 [−0.11, 0.17]
NSS2 −0.02 [−0.25, 0.20]
ONGA −0.45 [−1.13, 0.24]
ONGB −0.04 [−0.53, 0.45]
PPDS −0.08 [−0.22, 0.06]
PRIS −0.25 [−0.58, 0.09]
PSY5 −0.13 [−0.57, 0.31]
PTS1 −0.04 [−0.45, 0.37]
QIM2 0.09 [−0.26, 0.44]
QIMR −0.16 [−0.46, 0.13]
RCOG −0.08 [−0.31, 0.15]
RING −0.11 [−0.82, 0.59]
SEEP −0.22 [−0.61, 0.17]
STRO −0.06 [−0.35, 0.24]
TRAC 0.57 [−0.29, 1.43]
UKBB −0.04 [−0.07, −0.02]
VAKN 0.10 [−0.13, 0.34]
VETS 0.10 [−0.19, 0.40]
WACH −0.15 [−1.06, 0.77]
WRBY −0.02 [−0.50, 0.45]
WTCM 0.40 [−0.31, 1.12]
WTCS −0.04 [−0.29, 0.22]
F2.5 Meta −0.04 [−0.06, −0.02]
EHR
BIOM 0.06 [−0.21, 0.33]
DAI2 −0.07 [−0.11, −0.03]
HUNT −0.01 [−0.10, 0.07]
MAYO 0.04 [−0.12, 0.19]
MGBB 0.03 [−0.05, 0.11]
SWED 0.10 [−0.17, 0.36]
UKB2 −0.01 [−0.06, 0.04]
EHR Meta −0.03 [−0.06, −0.00]
MVP
MVP1 −0.05 [−0.07, −0.02]
Freeze 3 Meta −0.04 [−0.05, −0.03]
−2
−1
0
1
2
Observed Outcome

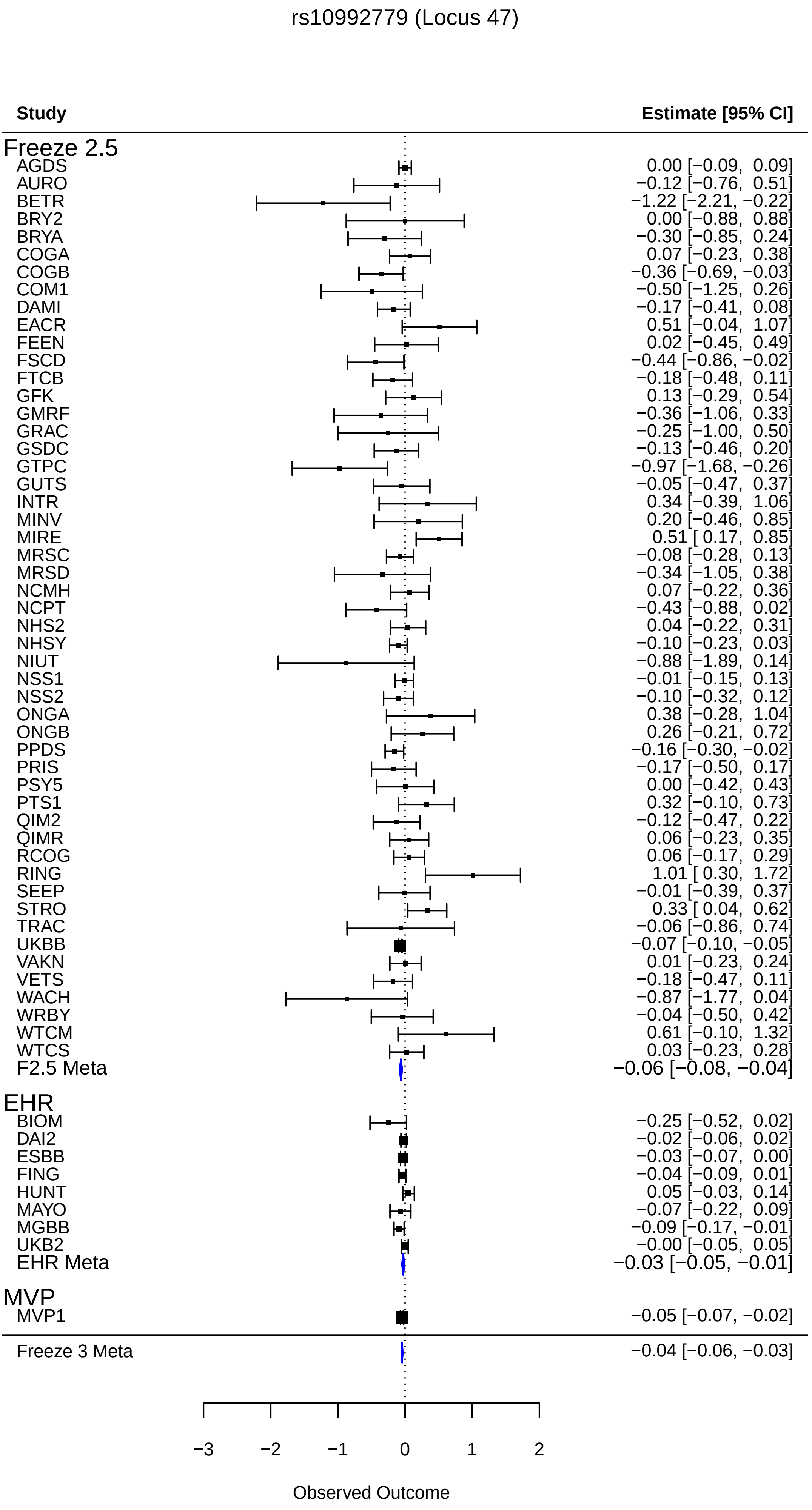
rs10992779 (Locus 47)
Study
Estimate [95% CI]
Freeze 2.5
AGDS 0.00 [−0.09, 0.09]
AURO −0.12 [−0.76, 0.51]
BETR −1.22 [−2.21, −0.22]
BRY2 0.00 [−0.88, 0.88]
BRYA −0.30 [−0.85, 0.24]
COGA 0.07 [−0.23, 0.38]
COGB −0.36 [−0.69, −0.03]
COM1 −0.50 [−1.25, 0.26]
DAMI −0.17 [−0.41, 0.08]
EACR 0.51 [−0.04, 1.07]
FEEN 0.02 [−0.45, 0.49]
FSCD −0.44 [−0.86, −0.02]
FTCB −0.18 [−0.48, 0.11]
GFK 0.13 [−0.29, 0.54]
GMRF −0.36 [−1.06, 0.33]
GRAC −0.25 [−1.00, 0.50]
GSDC −0.13 [−0.46, 0.20]
GTPC −0.97 [−1.68, −0.26]
GUTS −0.05 [−0.47, 0.37]
INTR 0.34 [−0.39, 1.06]
MINV 0.20 [−0.46, 0.85]
MIRE 0.51 [ 0.17, 0.85]
MRSC −0.08 [−0.28, 0.13]
MRSD −0.34 [−1.05, 0.38]
NCMH 0.07 [−0.22, 0.36]
NCPT −0.43 [−0.88, 0.02]
NHS2 0.04 [−0.22, 0.31]
NHSY −0.10 [−0.23, 0.03]
NIUT −0.88 [−1.89, 0.14]
NSS1 −0.01 [−0.15, 0.13]
NSS2 −0.10 [−0.32, 0.12]
ONGA 0.38 [−0.28, 1.04]
ONGB 0.26 [−0.21, 0.72]
PPDS −0.16 [−0.30, −0.02]
PRIS −0.17 [−0.50, 0.17]
PSY5 0.00 [−0.42, 0.43]
PTS1 0.32 [−0.10, 0.73]
QIM2 −0.12 [−0.47, 0.22]
QIMR 0.06 [−0.23, 0.35]
RCOG 0.06 [−0.17, 0.29]
RING 1.01 [ 0.30, 1.72]
SEEP −0.01 [−0.39, 0.37]
STRO 0.33 [ 0.04, 0.62]
TRAC −0.06 [−0.86, 0.74]
UKBB −0.07 [−0.10, −0.05]
VAKN 0.01 [−0.23, 0.24]
VETS −0.18 [−0.47, 0.11]
WACH −0.87 [−1.77, 0.04]
WRBY −0.04 [−0.50, 0.42]
WTCM 0.61 [−0.10, 1.32]
WTCS 0.03 [−0.23, 0.28]
F2.5 Meta −0.06 [−0.08, −0.04]
EHR
BIOM −0.25 [−0.52, 0.02]
DAI2 −0.02 [−0.06, 0.02]
ESBB −0.03 [−0.07, 0.00]
FING −0.04 [−0.09, 0.01]
HUNT 0.05 [−0.03, 0.14]
MAYO −0.07 [−0.22, 0.09]
MGBB −0.09 [−0.17, −0.01]
UKB2 −0.00 [−0.05, 0.05]
EHR Meta −0.03 [−0.05, −0.01]
MVP
MVP1 −0.05 [−0.07, −0.02]
Freeze 3 Meta −0.04 [−0.06, −0.03]
−3
−2
−1
0
1
2
Observed Outcome

rs67736073 (Locus 48)

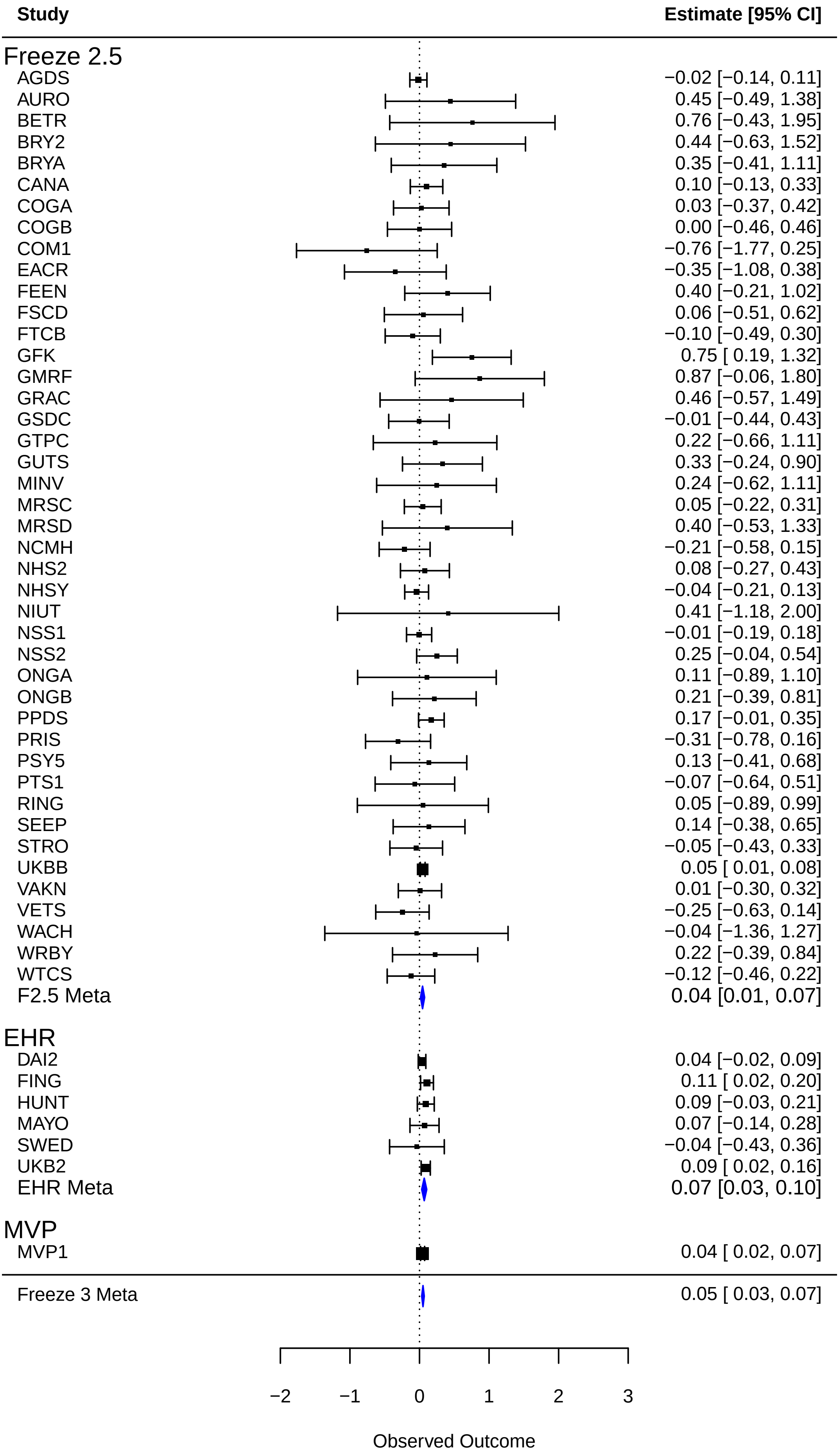

rs1124372 (Locus 49)

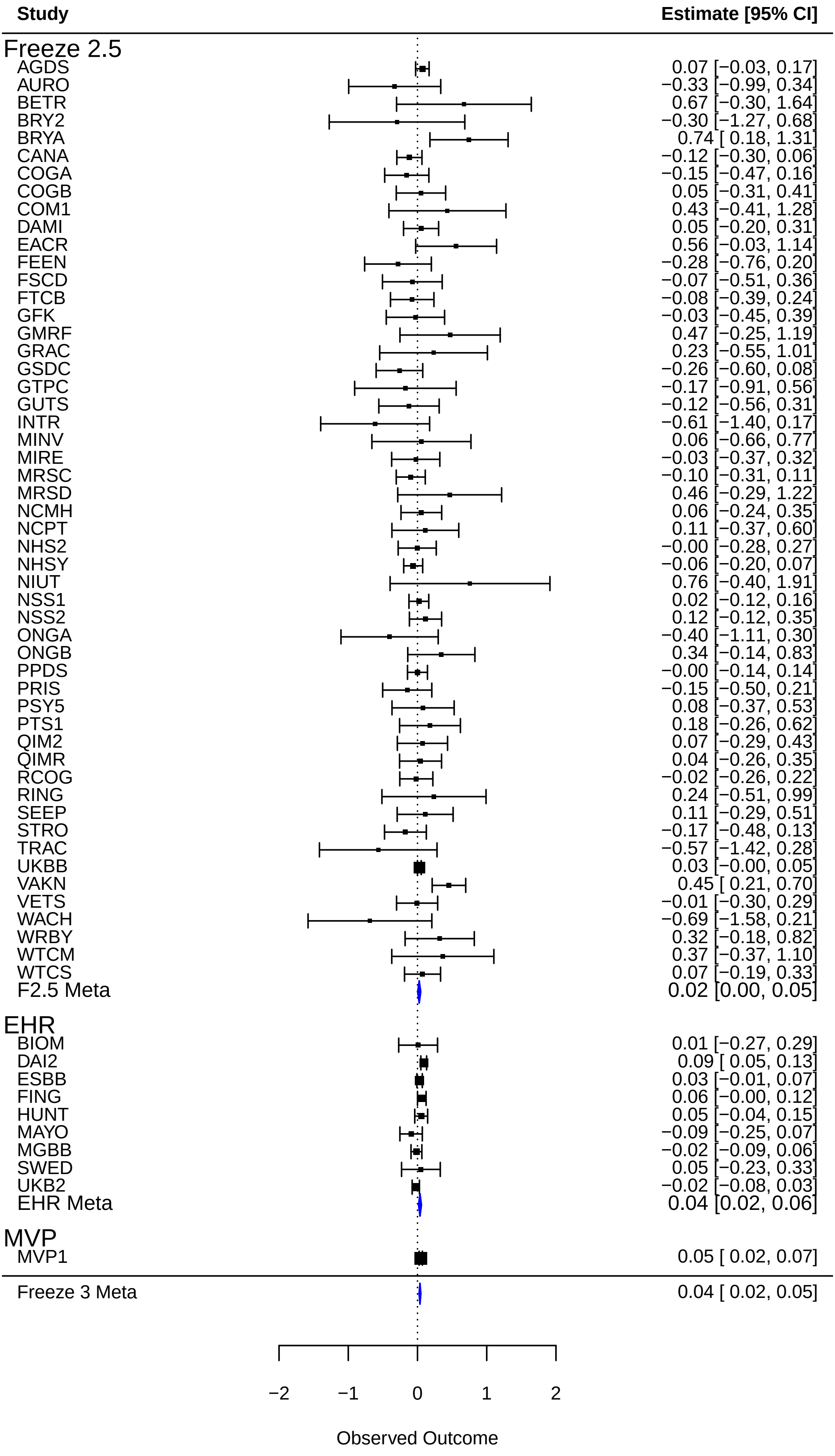

### rs7914674 (Locus 50)

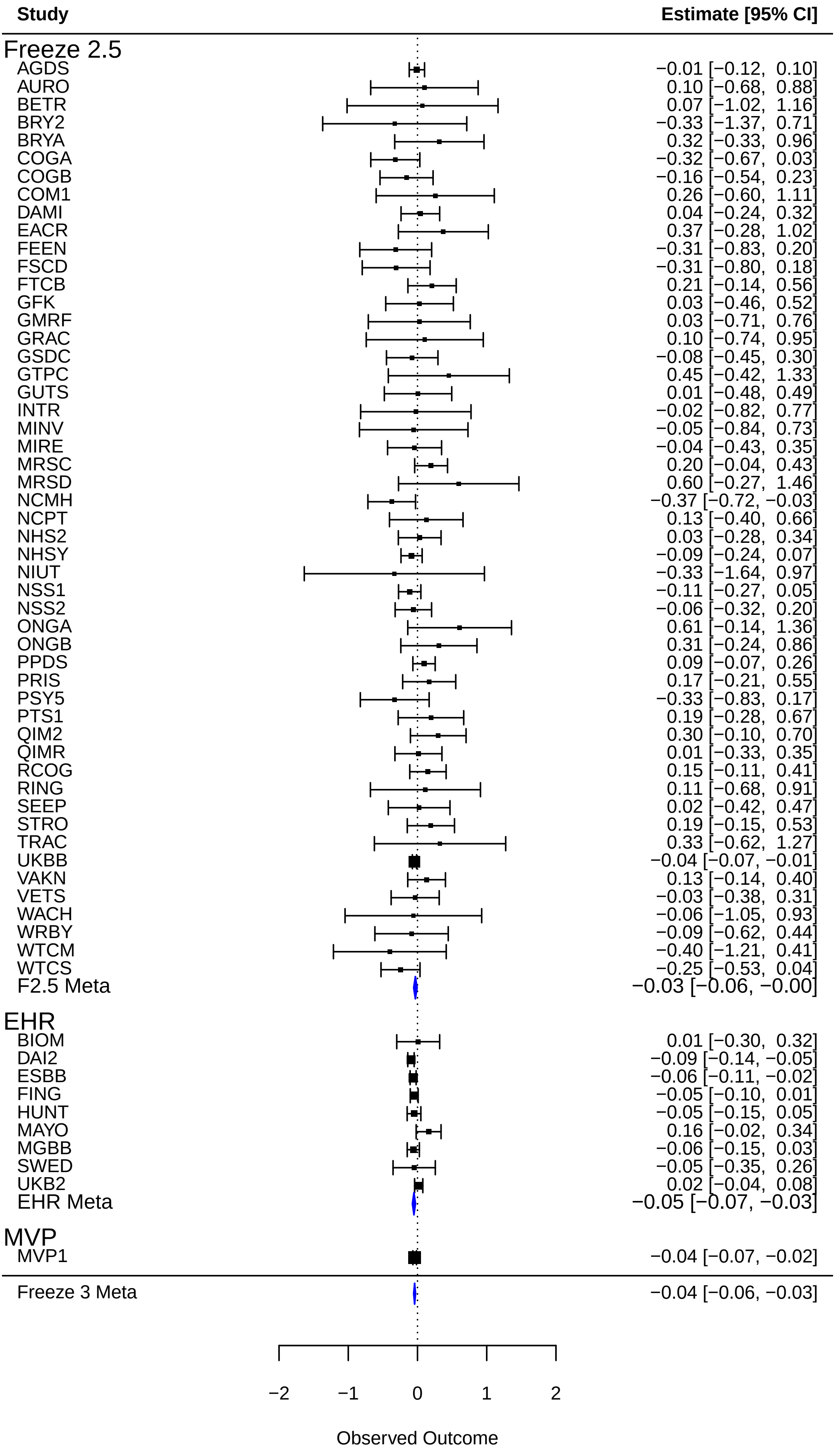

rs11529859 (Locus 51)

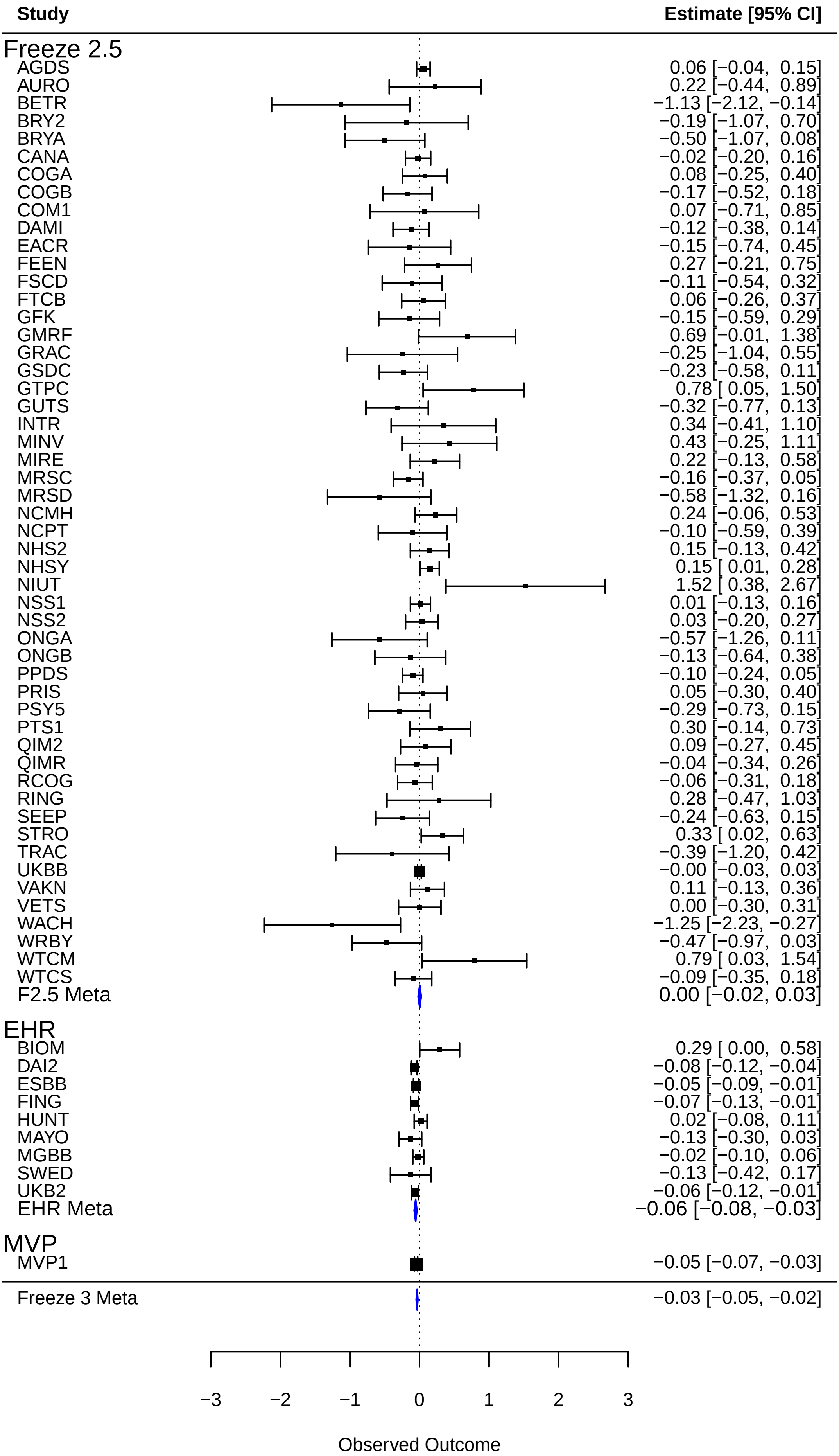

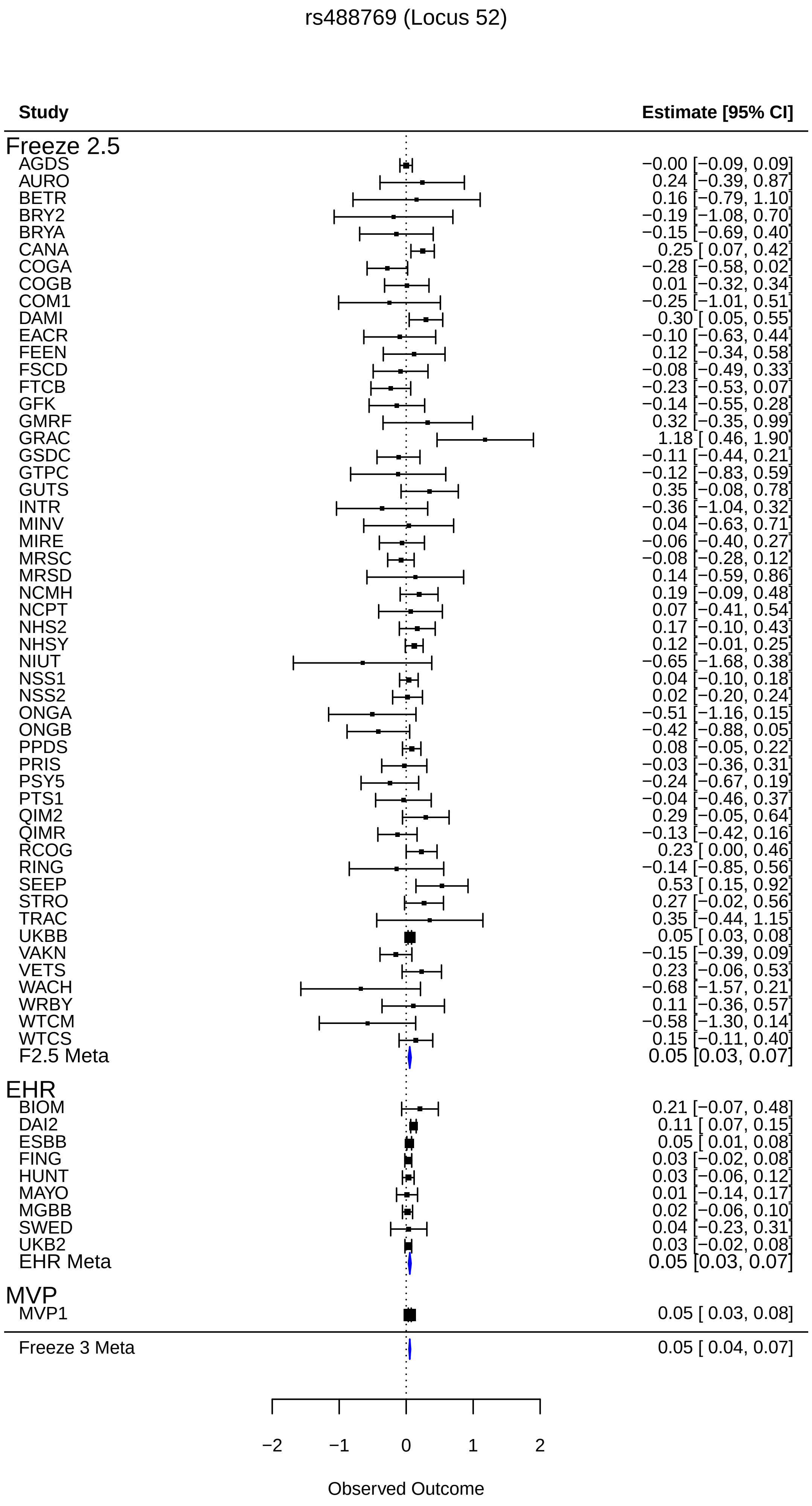
rs488769 (Locus 52)
Study
Estimate [95% CI]
Freeze 2.5
AGDS −0.00 [−0.09, 0.09]
AURO 0.24 [−0.39, 0.87]
BETR 0.16 [−0.79, 1.10]
BRY2 −0.19 [−1.08, 0.70]
BRYA −0.15 [−0.69, 0.40]
CANA 0.25 [ 0.07, 0.42]
COGA −0.28 [−0.58, 0.02]
COGB 0.01 [−0.32, 0.34]
COM1 −0.25 [−1.01, 0.51]
DAMI 0.30 [ 0.05, 0.55]
EACR −0.10 [−0.63, 0.44]
FEEN 0.12 [−0.34, 0.58]
FSCD −0.08 [−0.49, 0.33]
FTCB −0.23 [−0.53, 0.07]
GFK −0.14 [−0.55, 0.28]
GMRF 0.32 [−0.35, 0.99]
GRAC 1.18 [ 0.46, 1.90]
GSDC −0.11 [−0.44, 0.21]
GTPC −0.12 [−0.83, 0.59]
GUTS 0.35 [−0.08, 0.78]
INTR −0.36 [−1.04, 0.32]
MINV 0.04 [−0.63, 0.71]
MIRE −0.06 [−0.40, 0.27]
MRSC −0.08 [−0.28, 0.12]
MRSD 0.14 [−0.59, 0.86]
NCMH 0.19 [−0.09, 0.48]
NCPT 0.07 [−0.41, 0.54]
NHS2 0.17 [−0.10, 0.43]
NHSY 0.12 [−0.01, 0.25]
NIUT −0.65 [−1.68, 0.38]
NSS1 0.04 [−0.10, 0.18]
NSS2 0.02 [−0.20, 0.24]
ONGA −0.51 [−1.16, 0.15]
ONGB −0.42 [−0.88, 0.05]
PPDS 0.08 [−0.05, 0.22]
PRIS −0.03 [−0.36, 0.31]
PSY5 −0.24 [−0.67, 0.19]
PTS1 −0.04 [−0.46, 0.37]
QIM2 0.29 [−0.05, 0.64]
QIMR −0.13 [−0.42, 0.16]
RCOG 0.23 [ 0.00, 0.46]
RING −0.14 [−0.85, 0.56]
SEEP 0.53 [ 0.15, 0.92]
STRO 0.27 [−0.02, 0.56]
TRAC 0.35 [−0.44, 1.15]
UKBB 0.05 [ 0.03, 0.08]
VAKN −0.15 [−0.39, 0.09]
VETS 0.23 [−0.06, 0.53]
WACH −0.68 [−1.57, 0.21]
WRBY 0.11 [−0.36, 0.57]
WTCM −0.58 [−1.30, 0.14]
WTCS 0.15 [−0.11, 0.40]
F2.5 Meta 0.05 [0.03, 0.07]
EHR
BIOM 0.21 [−0.07, 0.48]
DAI2 0.11 [ 0.07, 0.15]
ESBB 0.05 [ 0.01, 0.08]
FING 0.03 [−0.02, 0.08]
HUNT 0.03 [−0.06, 0.12]
MAYO 0.01 [−0.14, 0.17]
MGBB 0.02 [−0.06, 0.10]
SWED 0.04 [−0.23, 0.31]
UKB2 0.03 [−0.02, 0.08]
EHR Meta 0.05 [0.03, 0.07]
MVP
MVP1 0.05 [ 0.03, 0.08]
Freeze 3 Meta 0.05 [ 0.04, 0.07]
−2
−1
0
1
2
Observed Outcome

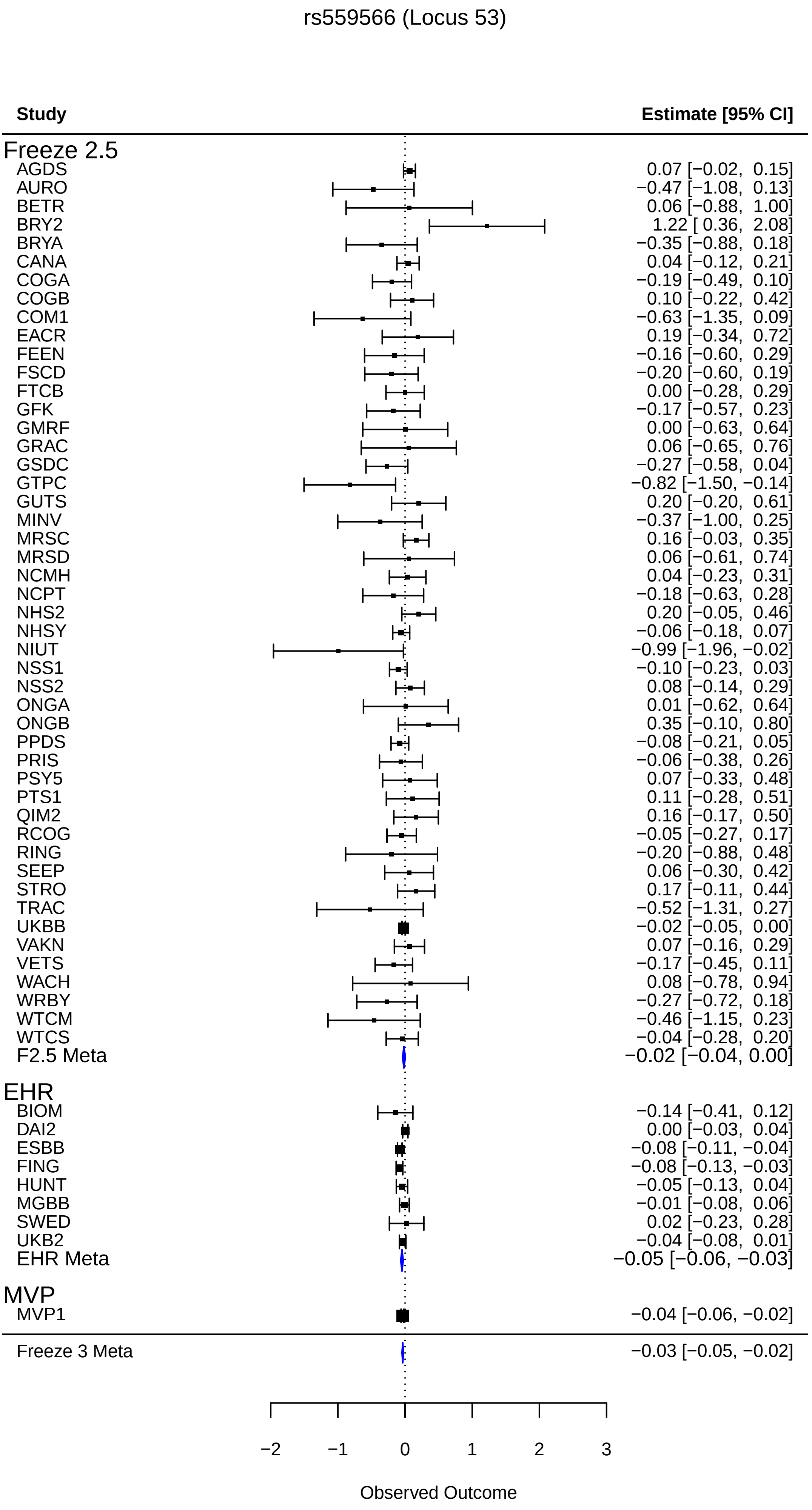
rs559566 (Locus 53)
Study
Estimate [95% CI]
Freeze 2.5
AGDS 0.07 [−0.02, 0.15]
AURO −0.47 [−1.08, 0.13]
BETR 0.06 [−0.88, 1.00]
BRY2 1.22 [ 0.36, 2.08]
BRYA −0.35 [−0.88, 0.18]
CANA 0.04 [−0.12, 0.21]
COGA −0.19 [−0.49, 0.10]
COGB 0.10 [−0.22, 0.42]
COM1 −0.63 [−1.35, 0.09]
EACR 0.19 [−0.34, 0.72]
FEEN −0.16 [−0.60, 0.29]
FSCD −0.20 [−0.60, 0.19]
FTCB 0.00 [−0.28, 0.29]
GFK −0.17 [−0.57, 0.23]
GMRF 0.00 [−0.63, 0.64]
GRAC 0.06 [−0.65, 0.76]
GSDC −0.27 [−0.58, 0.04]
GTPC −0.82 [−1.50, −0.14]
GUTS 0.20 [−0.20, 0.61]
MINV −0.37 [−1.00, 0.25]
MRSC 0.16 [−0.03, 0.35]
MRSD 0.06 [−0.61, 0.74]
NCMH 0.04 [−0.23, 0.31]
NCPT −0.18 [−0.63, 0.28]
NHS2 0.20 [−0.05, 0.46]
NHSY −0.06 [−0.18, 0.07]
NIUT −0.99 [−1.96, −0.02]
NSS1 −0.10 [−0.23, 0.03]
NSS2 0.08 [−0.14, 0.29]
ONGA 0.01 [−0.62, 0.64]
ONGB 0.35 [−0.10, 0.80]
PPDS −0.08 [−0.21, 0.05]
PRIS −0.06 [−0.38, 0.26]
PSY5 0.07 [−0.33, 0.48]
PTS1 0.11 [−0.28, 0.51]
QIM2 0.16 [−0.17, 0.50]
RCOG −0.05 [−0.27, 0.17]
RING −0.20 [−0.88, 0.48]
SEEP 0.06 [−0.30, 0.42]
STRO 0.17 [−0.11, 0.44]
TRAC −0.52 [−1.31, 0.27]
UKBB −0.02 [−0.05, 0.00]
VAKN 0.07 [−0.16, 0.29]
VETS −0.17 [−0.45, 0.11]
WACH 0.08 [−0.78, 0.94]
WRBY −0.27 [−0.72, 0.18]
WTCM −0.46 [−1.15, 0.23]
WTCS −0.04 [−0.28, 0.20]
F2.5 Meta −0.02 [−0.04, 0.00]
EHR
BIOM −0.14 [−0.41, 0.12]
DAI2 0.00 [−0.03, 0.04]
ESBB −0.08 [−0.11, −0.04]
FING −0.08 [−0.13, −0.03]
HUNT −0.05 [−0.13, 0.04]
MGBB −0.01 [−0.08, 0.06]
SWED 0.02 [−0.23, 0.28]
UKB2 −0.04 [−0.08, 0.01]
EHR Meta −0.05 [−0.06, −0.03]
MVP
MVP1 −0.04 [−0.06, −0.02]
Freeze 3 Meta −0.03 [−0.05, −0.02]
−2
−1
0
1
2
3
Observed Outcome

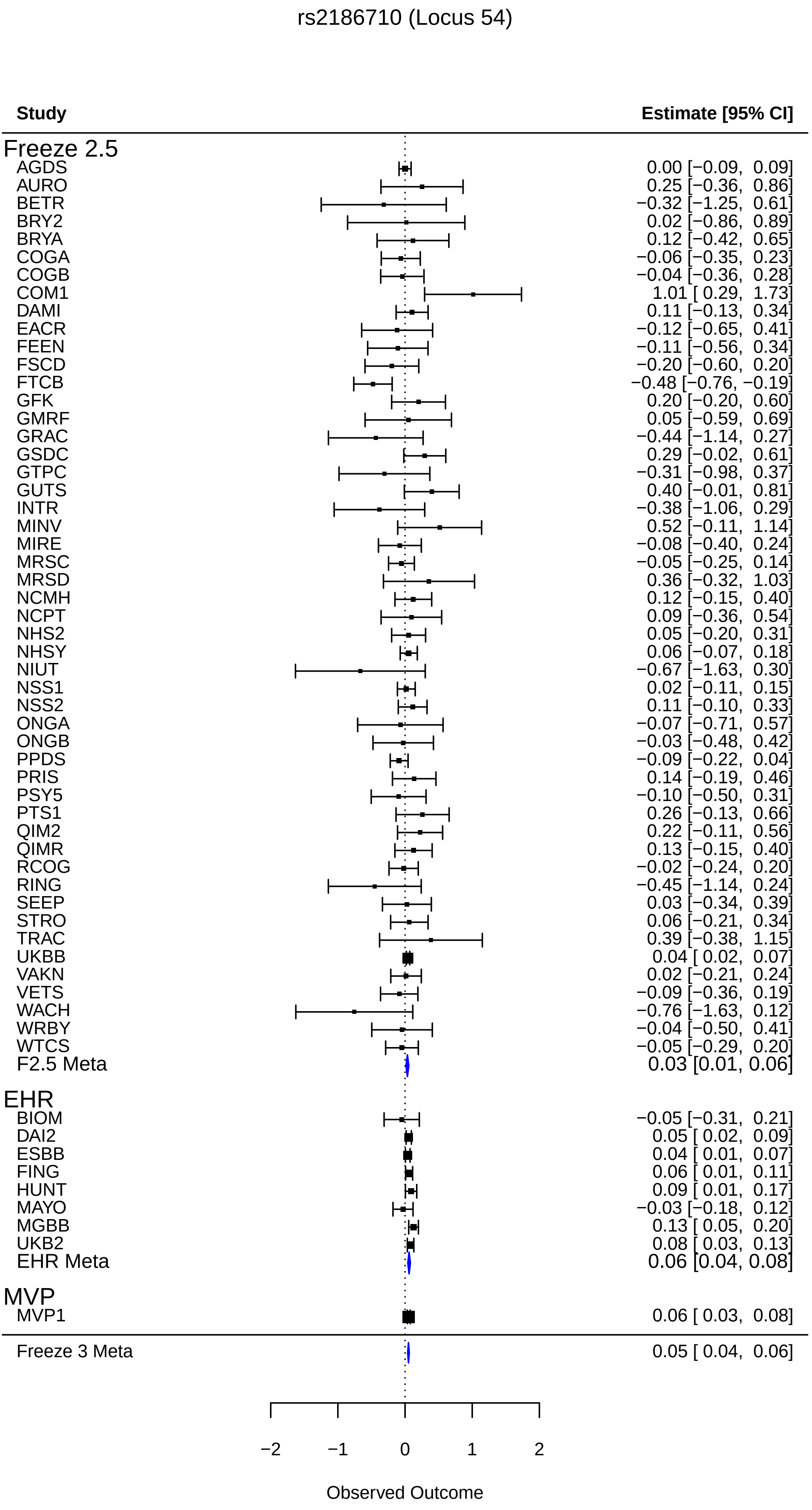
rs2186710 (Locus 54)
Study
Estimate [95% CI]
Freeze 2.5
AGDS 0.00 [−0.09, 0.09]
AURO 0.25 [−0.36, 0.86]
BETR −0.32 [−1.25, 0.61]
BRY2 0.02 [−0.86, 0.89]
BRYA 0.12 [−0.42, 0.65]
COGA −0.06 [−0.35, 0.23]
COGB −0.04 [−0.36, 0.28]
COM1 1.01 [ 0.29, 1.73]
DAMI 0.11 [−0.13, 0.34]
EACR −0.12 [−0.65, 0.41]
FEEN −0.11 [−0.56, 0.34]
FSCD −0.20 [−0.60, 0.20]
FTCB −0.48 [−0.76, −0.19]
GFK 0.20 [−0.20, 0.60]
GMRF 0.05 [−0.59, 0.69]
GRAC −0.44 [−1.14, 0.27]
GSDC 0.29 [−0.02, 0.61]
GTPC −0.31 [−0.98, 0.37]
GUTS 0.40 [−0.01, 0.81]
INTR −0.38 [−1.06, 0.29]
MINV 0.52 [−0.11, 1.14]
MIRE −0.08 [−0.40, 0.24]
MRSC −0.05 [−0.25, 0.14]
MRSD 0.36 [−0.32, 1.03]
NCMH 0.12 [−0.15, 0.40]
NCPT 0.09 [−0.36, 0.54]
NHS2 0.05 [−0.20, 0.31]
NHSY 0.06 [−0.07, 0.18]
NIUT −0.67 [−1.63, 0.30]
NSS1 0.02 [−0.11, 0.15]
NSS2 0.11 [−0.10, 0.33]
ONGA −0.07 [−0.71, 0.57]
ONGB −0.03 [−0.48, 0.42]
PPDS −0.09 [−0.22, 0.04]
PRIS 0.14 [−0.19, 0.46]
PSY5 −0.10 [−0.50, 0.31]
PTS1 0.26 [−0.13, 0.66]
QIM2 0.22 [−0.11, 0.56]
QIMR 0.13 [−0.15, 0.40]
RCOG −0.02 [−0.24, 0.20]
RING −0.45 [−1.14, 0.24]
SEEP 0.03 [−0.34, 0.39]
STRO 0.06 [−0.21, 0.34]
TRAC 0.39 [−0.38, 1.15]
UKBB 0.04 [ 0.02, 0.07]
VAKN 0.02 [−0.21, 0.24]
VETS −0.09 [−0.36, 0.19]
WACH −0.76 [−1.63, 0.12]
WRBY −0.04 [−0.50, 0.41]
WTCS −0.05 [−0.29, 0.20]
F2.5 Meta 0.03 [0.01, 0.06]
EHR
BIOM −0.05 [−0.31, 0.21]
DAI2 0.05 [ 0.02, 0.09]
ESBB 0.04 [ 0.01, 0.07]
FING 0.06 [ 0.01, 0.11]
HUNT 0.09 [ 0.01, 0.17]
MAYO −0.03 [−0.18, 0.12]
MGBB 0.13 [ 0.05, 0.20]
UKB2 0.08 [ 0.03, 0.13]
EHR Meta 0.06 [0.04, 0.08]
MVP
MVP1 0.06 [ 0.03, 0.08]
Freeze 3 Meta 0.05 [ 0.04, 0.06]
−2
−1
0
1
2
Observed Outcome

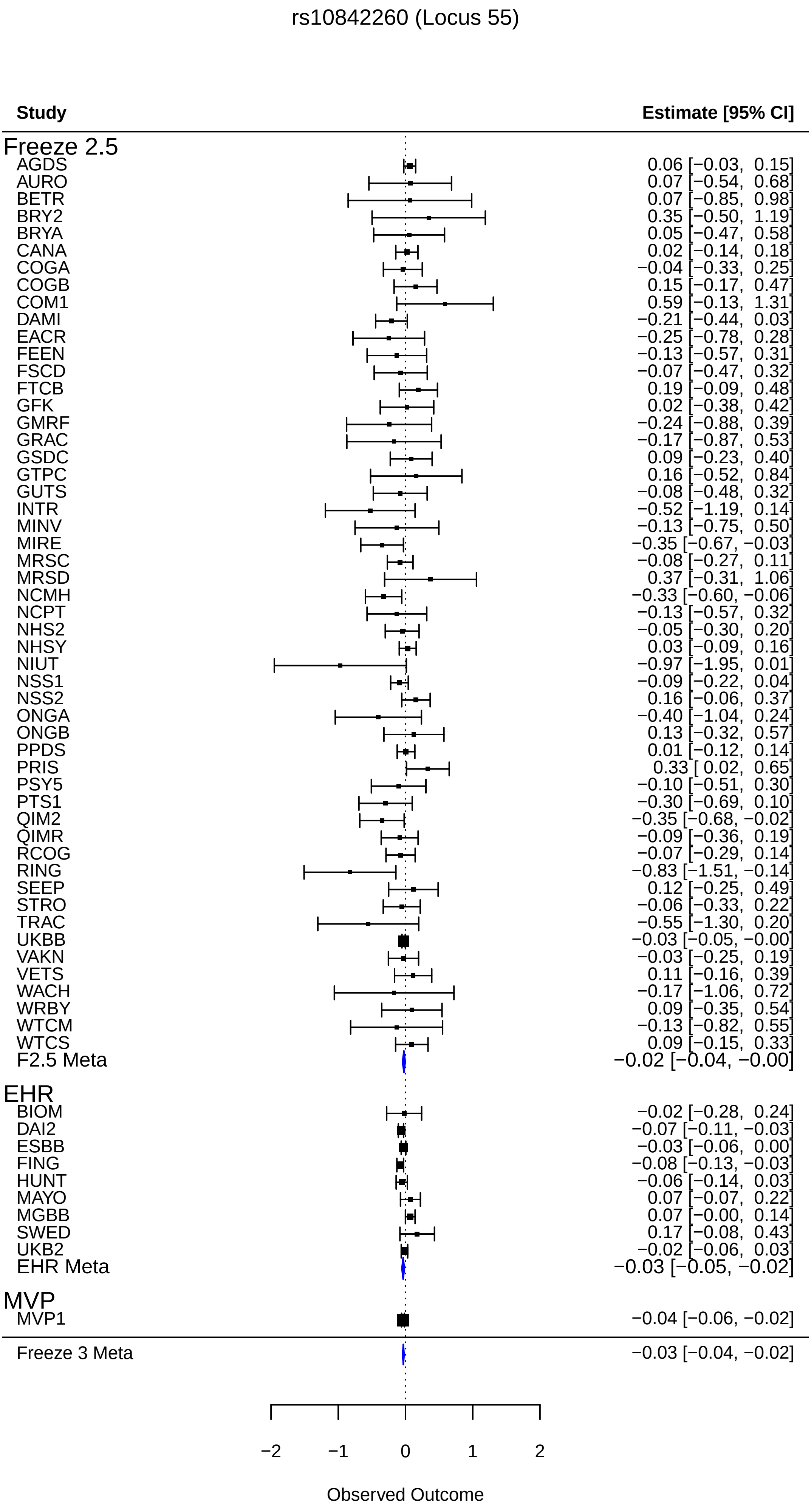
rs10842260 (Locus 55)
Study
Estimate [95% CI]
Freeze 2.5
AGDS 0.06 [−0.03, 0.15]
AURO 0.07 [−0.54, 0.68]
BETR 0.07 [−0.85, 0.98]
BRY2 0.35 [−0.50, 1.19]
BRYA 0.05 [−0.47, 0.58]
CANA 0.02 [−0.14, 0.18]
COGA −0.04 [−0.33, 0.25]
COGB 0.15 [−0.17, 0.47]
COM1 0.59 [−0.13, 1.31]
DAMI −0.21 [−0.44, 0.03]
EACR −0.25 [−0.78, 0.28]
FEEN −0.13 [−0.57, 0.31]
FSCD −0.07 [−0.47, 0.32]
FTCB 0.19 [−0.09, 0.48]
GFK 0.02 [−0.38, 0.42]
GMRF −0.24 [−0.88, 0.39]
GRAC −0.17 [−0.87, 0.53]
GSDC 0.09 [−0.23, 0.40]
GTPC 0.16 [−0.52, 0.84]
GUTS −0.08 [−0.48, 0.32]
INTR −0.52 [−1.19, 0.14]
MINV −0.13 [−0.75, 0.50]
MIRE −0.35 [−0.67, −0.03]
MRSC −0.08 [−0.27, 0.11]
MRSD 0.37 [−0.31, 1.06]
NCMH −0.33 [−0.60, −0.06]
NCPT −0.13 [−0.57, 0.32]
NHS2 −0.05 [−0.30, 0.20]
NHSY 0.03 [−0.09, 0.16]
NIUT −0.97 [−1.95, 0.01]
NSS1 −0.09 [−0.22, 0.04]
NSS2 0.16 [−0.06, 0.37]
ONGA −0.40 [−1.04, 0.24]
ONGB 0.13 [−0.32, 0.57]
PPDS 0.01 [−0.12, 0.14]
PRIS 0.33 [ 0.02, 0.65]
PSY5 −0.10 [−0.51, 0.30]
PTS1 −0.30 [−0.69, 0.10]
QIM2 −0.35 [−0.68, −0.02]
QIMR −0.09 [−0.36, 0.19]
RCOG −0.07 [−0.29, 0.14]
RING −0.83 [−1.51, −0.14]
SEEP 0.12 [−0.25, 0.49]
STRO −0.06 [−0.33, 0.22]
TRAC −0.55 [−1.30, 0.20]
UKBB −0.03 [−0.05, −0.00]
VAKN −0.03 [−0.25, 0.19]
VETS 0.11 [−0.16, 0.39]
WACH −0.17 [−1.06, 0.72]
WRBY 0.09 [−0.35, 0.54]
WTCM −0.13 [−0.82, 0.55]
WTCS 0.09 [−0.15, 0.33]
F2.5 Meta −0.02 [−0.04, −0.00]
EHR
BIOM −0.02 [−0.28, 0.24]
DAI2 −0.07 [−0.11, −0.03]
ESBB −0.03 [−0.06, 0.00]
FING −0.08 [−0.13, −0.03]
HUNT −0.06 [−0.14, 0.03]
MAYO 0.07 [−0.07, 0.22]
MGBB 0.07 [−0.00, 0.14]
SWED 0.17 [−0.08, 0.43]
UKB2 −0.02 [−0.06, 0.03]
EHR Meta −0.03 [−0.05, −0.02]
MVP
MVP1 −0.04 [−0.06, −0.02]
Freeze 3 Meta −0.03 [−0.04, −0.02]
−2
−1
0
1
2
Observed Outcome

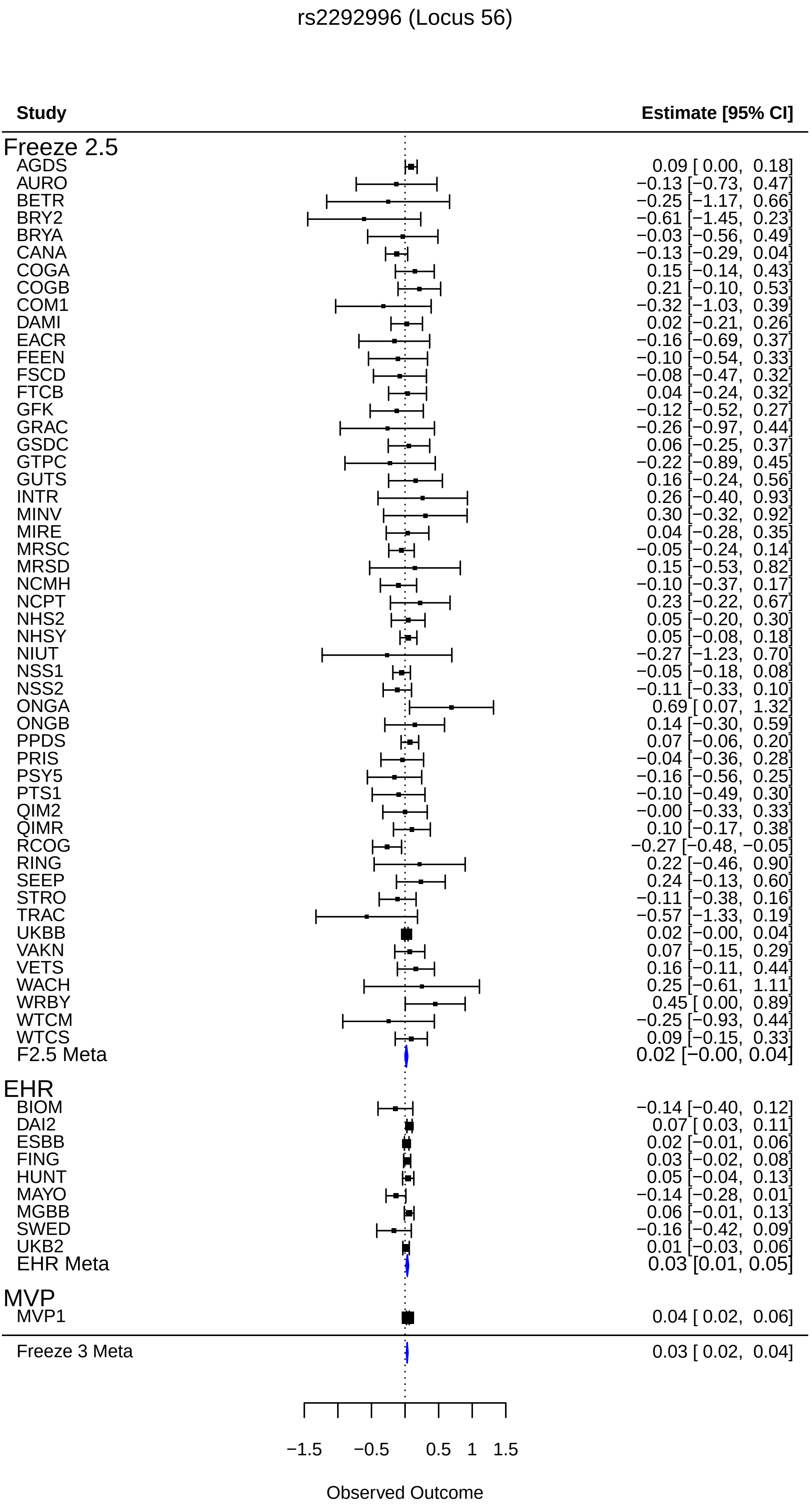
rs2292996 (Locus 56)
Study
Estimate [95% CI]
Freeze 2.5
AGDS 0.09 [ 0.00, 0.18]
AURO −0.13 [−0.73, 0.47]
BETR −0.25 [−1.17, 0.66]
BRY2 −0.61 [−1.45, 0.23]
BRYA −0.03 [−0.56, 0.49]
CANA −0.13 [−0.29, 0.04]
COGA 0.15 [−0.14, 0.43]
COGB 0.21 [−0.10, 0.53]
COM1 −0.32 [−1.03, 0.39]
DAMI 0.02 [−0.21, 0.26]
EACR −0.16 [−0.69, 0.37]
FEEN −0.10 [−0.54, 0.33]
FSCD −0.08 [−0.47, 0.32]
FTCB 0.04 [−0.24, 0.32]
GFK −0.12 [−0.52, 0.27]
GRAC −0.26 [−0.97, 0.44]
GSDC 0.06 [−0.25, 0.37]
GTPC −0.22 [−0.89, 0.45]
GUTS 0.16 [−0.24, 0.56]
INTR 0.26 [−0.40, 0.93]
MINV 0.30 [−0.32, 0.92]
MIRE 0.04 [−0.28, 0.35]
MRSC −0.05 [−0.24, 0.14]
MRSD 0.15 [−0.53, 0.82]
NCMH −0.10 [−0.37, 0.17]
NCPT 0.23 [−0.22, 0.67]
NHS2 0.05 [−0.20, 0.30]
NHSY 0.05 [−0.08, 0.18]
NIUT −0.27 [−1.23, 0.70]
NSS1 −0.05 [−0.18, 0.08]
NSS2 −0.11 [−0.33, 0.10]
ONGA 0.69 [ 0.07, 1.32]
ONGB 0.14 [−0.30, 0.59]
PPDS 0.07 [−0.06, 0.20]
PRIS −0.04 [−0.36, 0.28]
PSY5 −0.16 [−0.56, 0.25]
PTS1 −0.10 [−0.49, 0.30]
QIM2 −0.00 [−0.33, 0.33]
QIMR 0.10 [−0.17, 0.38]
RCOG −0.27 [−0.48, −0.05]
RING 0.22 [−0.46, 0.90]
SEEP 0.24 [−0.13, 0.60]
STRO −0.11 [−0.38, 0.16]
TRAC −0.57 [−1.33, 0.19]
UKBB 0.02 [−0.00, 0.04]
VAKN 0.07 [−0.15, 0.29]
VETS 0.16 [−0.11, 0.44]
WACH 0.25 [−0.61, 1.11]
WRBY 0.45 [ 0.00, 0.89]
WTCM −0.25 [−0.93, 0.44]
WTCS 0.09 [−0.15, 0.33]
F2.5 Meta 0.02 [−0.00, 0.04]
EHR
BIOM −0.14 [−0.40, 0.12]
DAI2 0.07 [ 0.03, 0.11]
ESBB 0.02 [−0.01, 0.06]
FING 0.03 [−0.02, 0.08]
HUNT 0.05 [−0.04, 0.13]
MAYO −0.14 [−0.28, 0.01]
MGBB 0.06 [−0.01, 0.13]
SWED −0.16 [−0.42, 0.09]
UKB2 0.01 [−0.03, 0.06]
EHR Meta 0.03 [0.01, 0.05]
MVP
MVP1 0.04 [ 0.02, 0.06]
Freeze 3 Meta 0.03 [ 0.02, 0.04]
−1.5
−0.5
0.5
1
1.5
Observed Outcome

rs816363 (Locus 57)

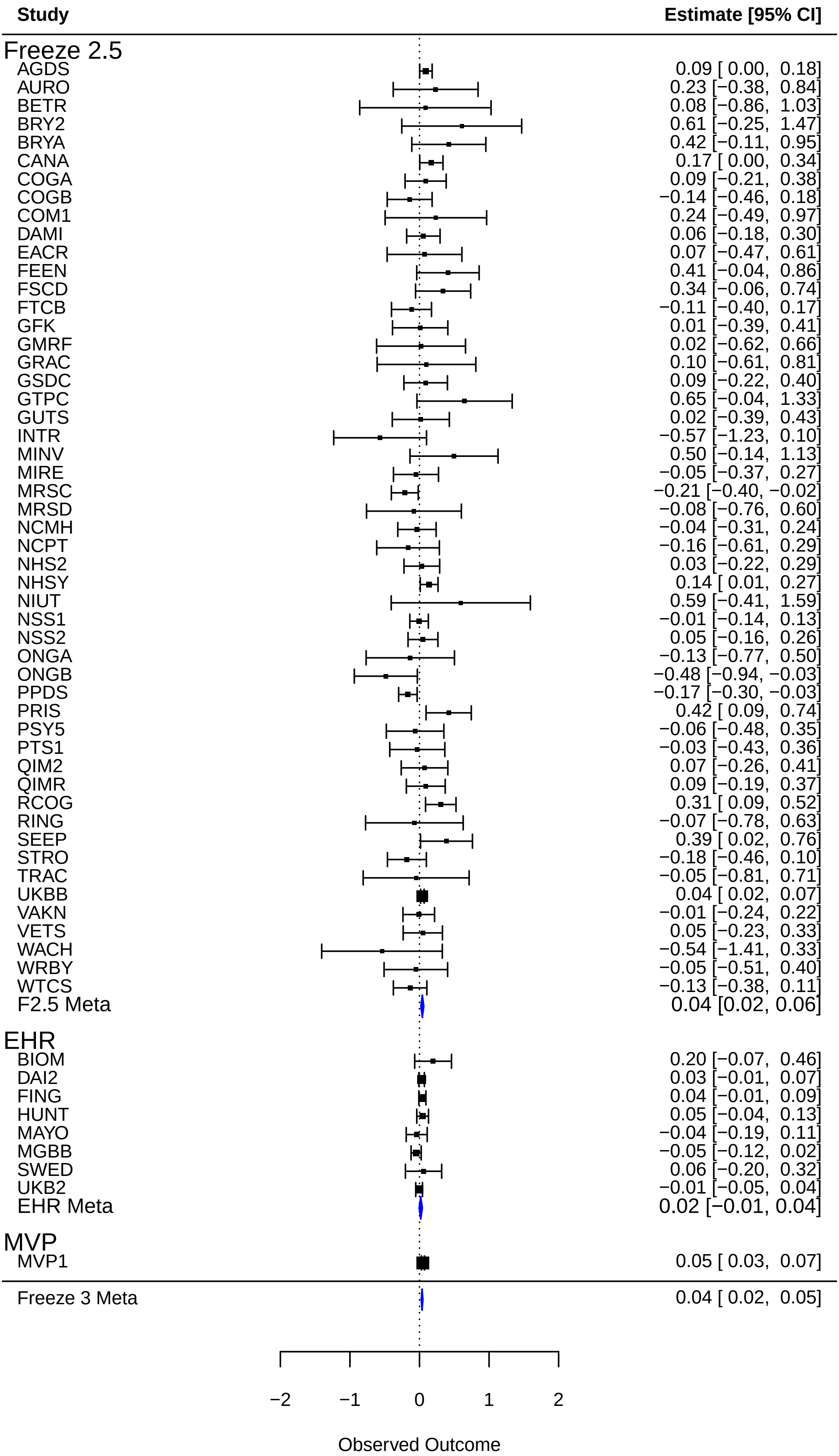

rs61946067 (Locus 58)

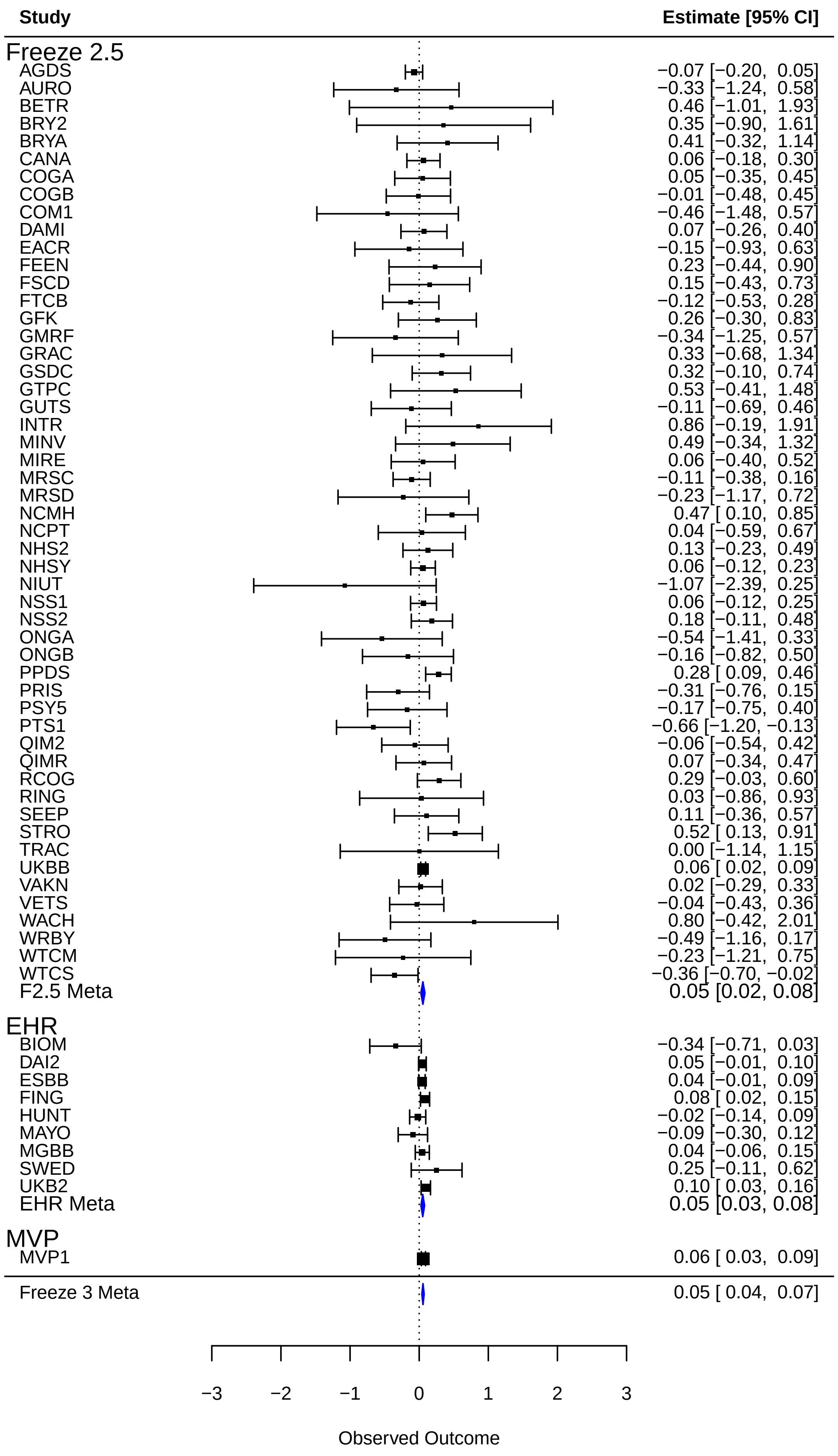

rs1373273 (Locus 59)

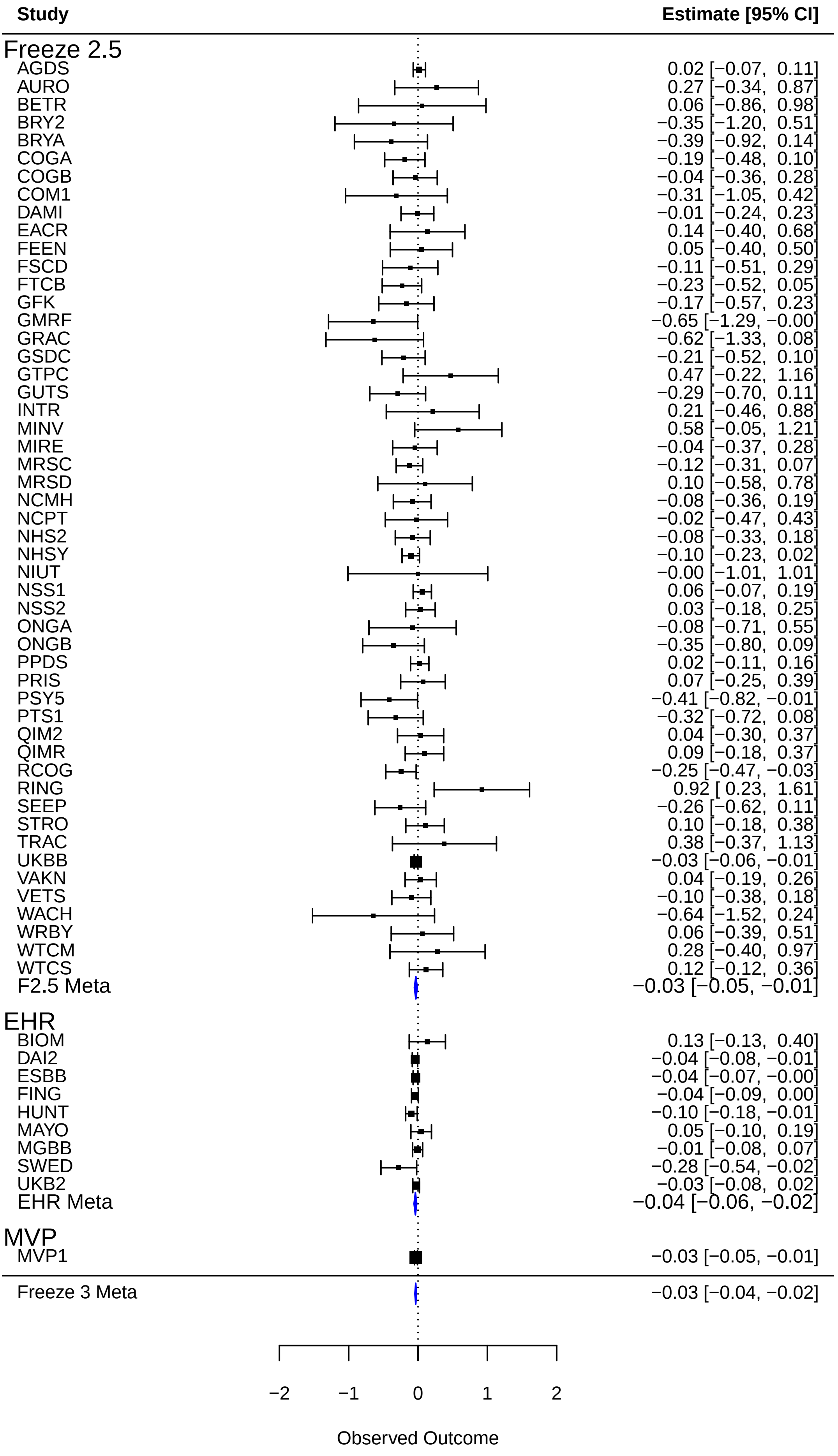

rs7333625 (Locus 60)

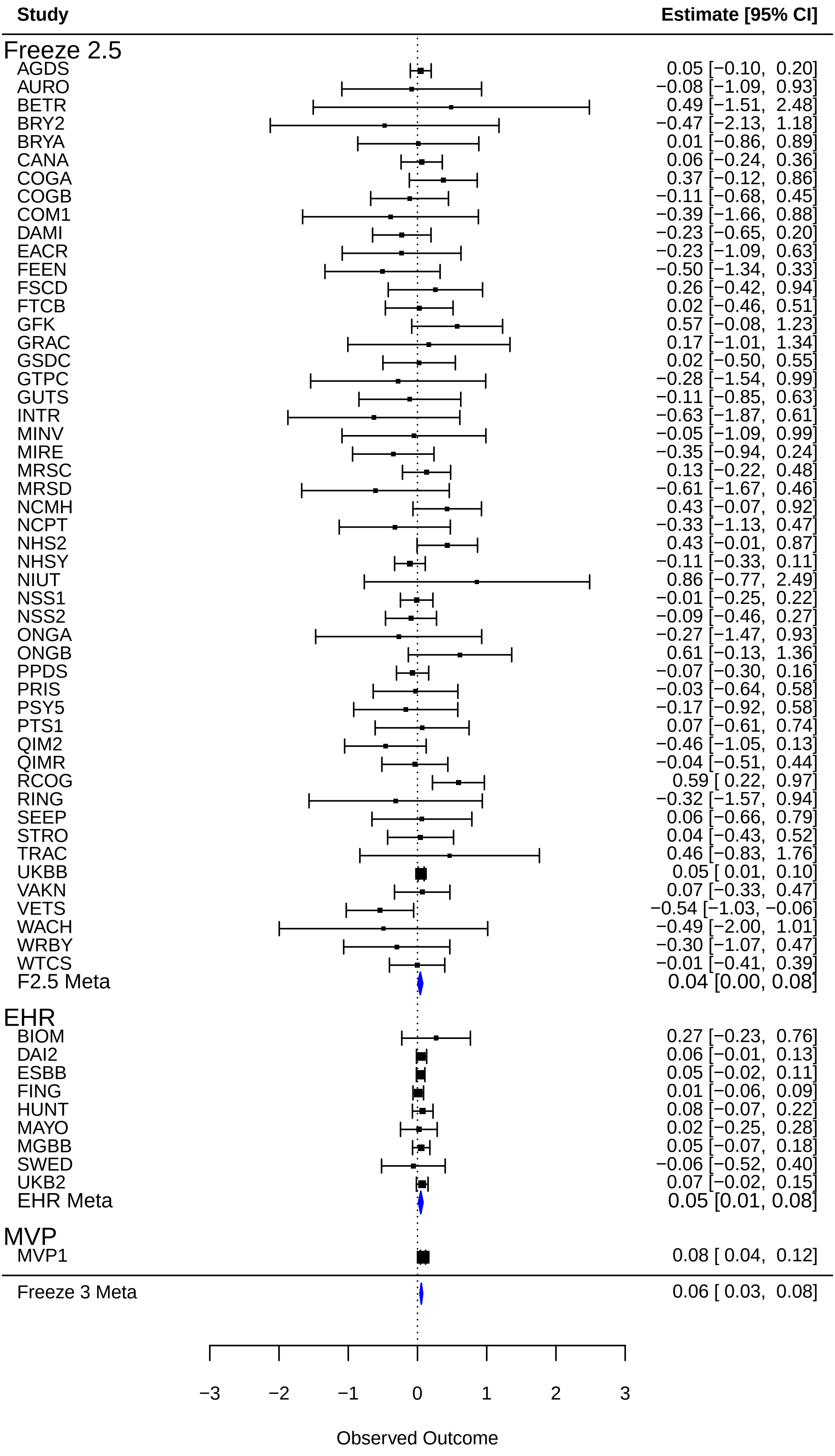

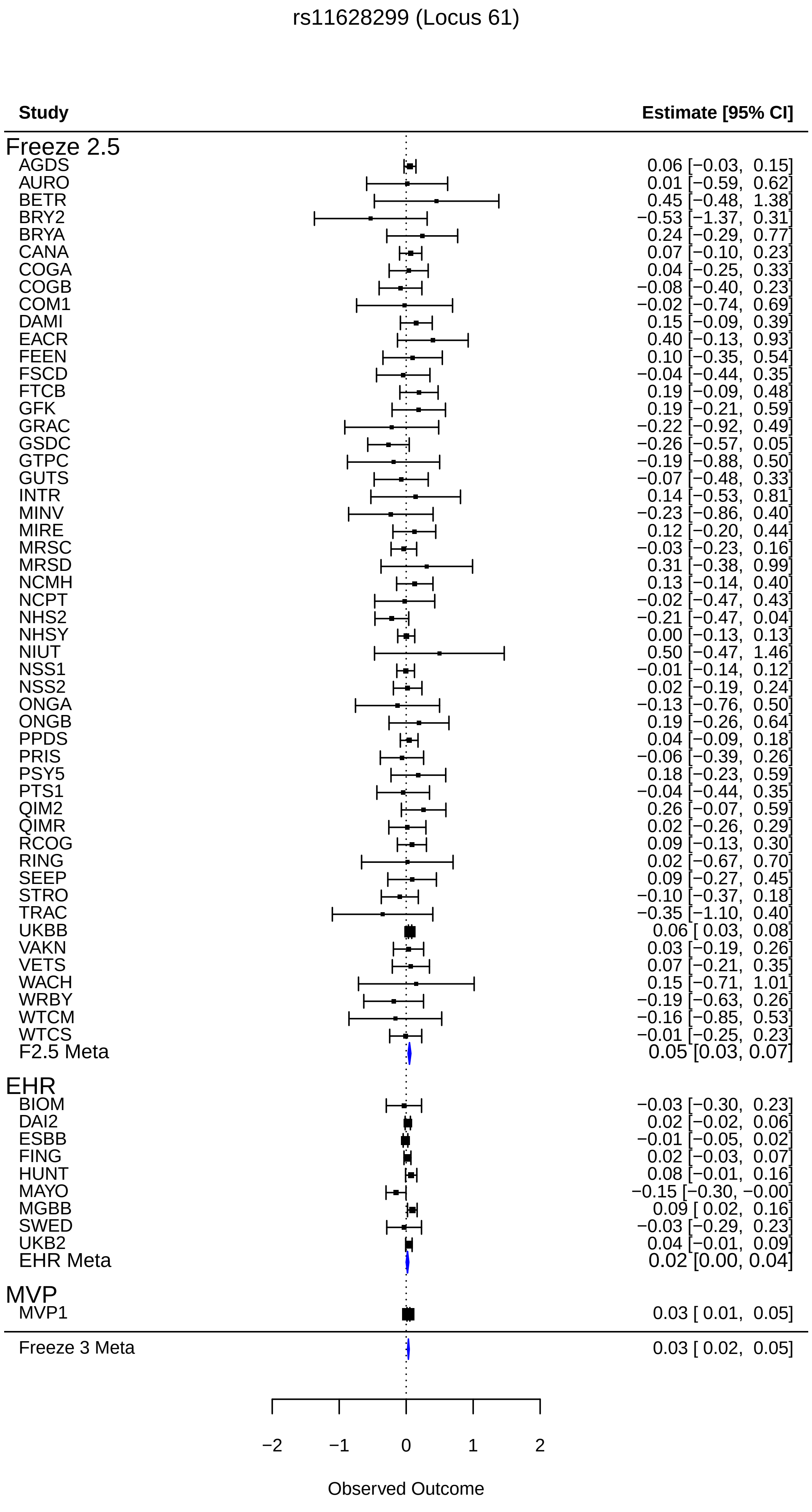
rs11628299 (Locus 61)
Study
Estimate [95% CI]
Freeze 2.5
AGDS 0.06 [−0.03, 0.15]
AURO 0.01 [−0.59, 0.62]
BETR 0.45 [−0.48, 1.38]
BRY2 −0.53 [−1.37, 0.31]
BRYA 0.24 [−0.29, 0.77]
CANA 0.07 [−0.10, 0.23]
COGA 0.04 [−0.25, 0.33]
COGB −0.08 [−0.40, 0.23]
COM1 −0.02 [−0.74, 0.69]
DAMI 0.15 [−0.09, 0.39]
EACR 0.40 [−0.13, 0.93]
FEEN 0.10 [−0.35, 0.54]
FSCD −0.04 [−0.44, 0.35]
FTCB 0.19 [−0.09, 0.48]
GFK 0.19 [−0.21, 0.59]
GRAC −0.22 [−0.92, 0.49]
GSDC −0.26 [−0.57, 0.05]
GTPC −0.19 [−0.88, 0.50]
GUTS −0.07 [−0.48, 0.33]
INTR 0.14 [−0.53, 0.81]
MINV −0.23 [−0.86, 0.40]
MIRE 0.12 [−0.20, 0.44]
MRSC −0.03 [−0.23, 0.16]
MRSD 0.31 [−0.38, 0.99]
NCMH 0.13 [−0.14, 0.40]
NCPT −0.02 [−0.47, 0.43]
NHS2 −0.21 [−0.47, 0.04]
NHSY 0.00 [−0.13, 0.13]
NIUT 0.50 [−0.47, 1.46]
NSS1 −0.01 [−0.14, 0.12]
NSS2 0.02 [−0.19, 0.24]
ONGA −0.13 [−0.76, 0.50]
ONGB 0.19 [−0.26, 0.64]
PPDS 0.04 [−0.09, 0.18]
PRIS −0.06 [−0.39, 0.26]
PSY5 0.18 [−0.23, 0.59]
PTS1 −0.04 [−0.44, 0.35]
QIM2 0.26 [−0.07, 0.59]
QIMR 0.02 [−0.26, 0.29]
RCOG 0.09 [−0.13, 0.30]
RING 0.02 [−0.67, 0.70]
SEEP 0.09 [−0.27, 0.45]
STRO −0.10 [−0.37, 0.18]
TRAC −0.35 [−1.10, 0.40]
UKBB 0.06 [ 0.03, 0.08]
VAKN 0.03 [−0.19, 0.26]
VETS 0.07 [−0.21, 0.35]
WACH 0.15 [−0.71, 1.01]
WRBY −0.19 [−0.63, 0.26]
WTCM −0.16 [−0.85, 0.53]
WTCS −0.01 [−0.25, 0.23]
F2.5 Meta 0.05 [0.03, 0.07]
EHR
BIOM −0.03 [−0.30, 0.23]
DAI2 0.02 [−0.02, 0.06]
ESBB −0.01 [−0.05, 0.02]
FING 0.02 [−0.03, 0.07]
HUNT 0.08 [−0.01, 0.16]
MAYO −0.15 [−0.30, −0.00]
MGBB 0.09 [ 0.02, 0.16]
SWED −0.03 [−0.29, 0.23]
UKB2 0.04 [−0.01, 0.09]
EHR Meta 0.02 [0.00, 0.04]
MVP
MVP1 0.03 [ 0.01, 0.05]
Freeze 3 Meta 0.03 [ 0.02, 0.05]
−2
−1
0
1
2
Observed Outcome

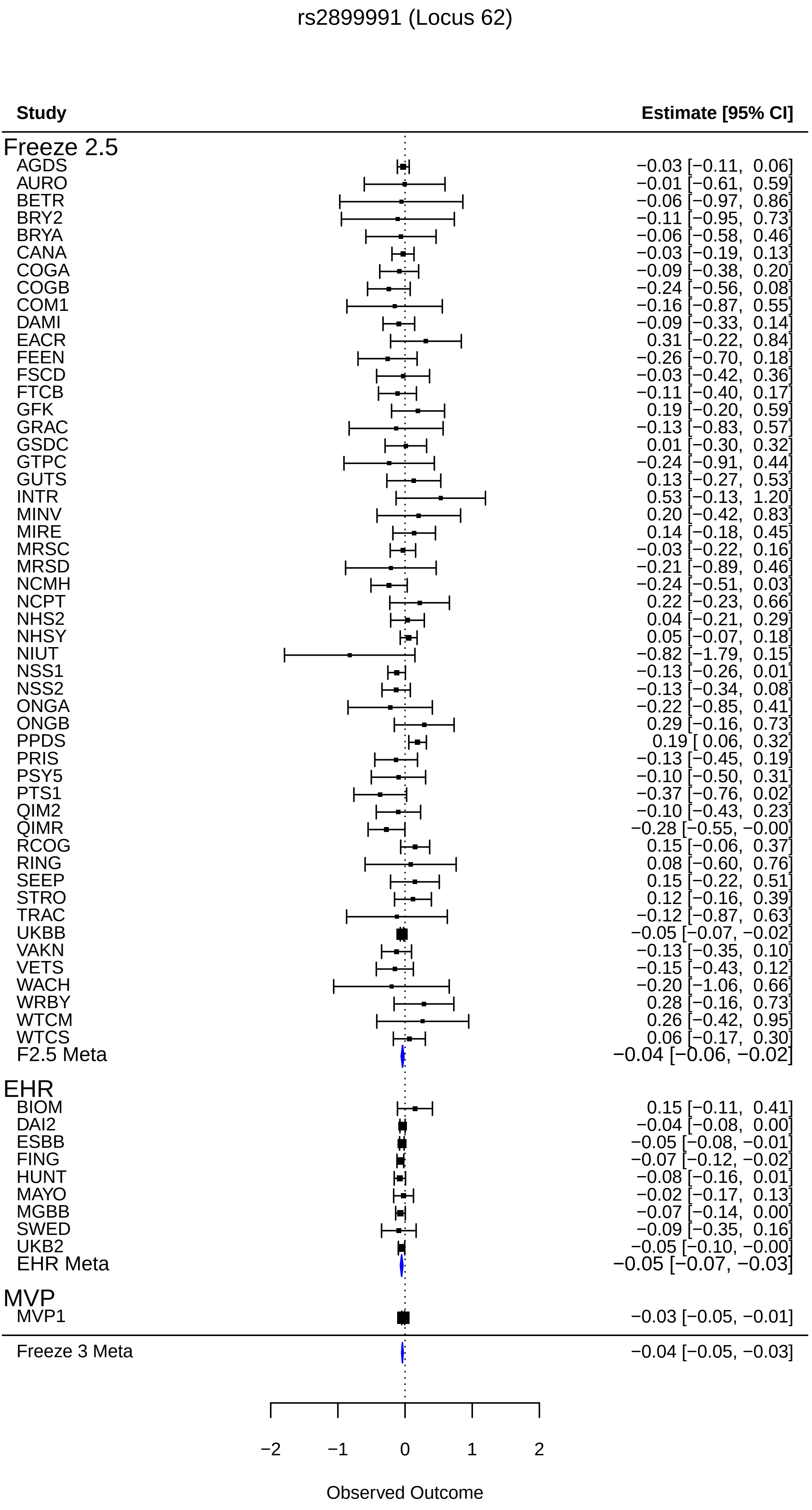
rs2899991 (Locus 62)
Study
Estimate [95% CI]
Freeze 2.5
AGDS −0.03 [−0.11, 0.06]
AURO −0.01 [−0.61, 0.59]
BETR −0.06 [−0.97, 0.86]
BRY2 −0.11 [−0.95, 0.73]
BRYA −0.06 [−0.58, 0.46]
CANA −0.03 [−0.19, 0.13]
COGA −0.09 [−0.38, 0.20]
COGB −0.24 [−0.56, 0.08]
COM1 −0.16 [−0.87, 0.55]
DAMI −0.09 [−0.33, 0.14]
EACR 0.31 [−0.22, 0.84]
FEEN −0.26 [−0.70, 0.18]
FSCD −0.03 [−0.42, 0.36]
FTCB −0.11 [−0.40, 0.17]
GFK 0.19 [−0.20, 0.59]
GRAC −0.13 [−0.83, 0.57]
GSDC 0.01 [−0.30, 0.32]
GTPC −0.24 [−0.91, 0.44]
GUTS 0.13 [−0.27, 0.53]
INTR 0.53 [−0.13, 1.20]
MINV 0.20 [−0.42, 0.83]
MIRE 0.14 [−0.18, 0.45]
MRSC −0.03 [−0.22, 0.16]
MRSD −0.21 [−0.89, 0.46]
NCMH −0.24 [−0.51, 0.03]
NCPT 0.22 [−0.23, 0.66]
NHS2 0.04 [−0.21, 0.29]
NHSY 0.05 [−0.07, 0.18]
NIUT −0.82 [−1.79, 0.15]
NSS1 −0.13 [−0.26, 0.01]
NSS2 −0.13 [−0.34, 0.08]
ONGA −0.22 [−0.85, 0.41]
ONGB 0.29 [−0.16, 0.73]
PPDS 0.19 [ 0.06, 0.32]
PRIS −0.13 [−0.45, 0.19]
PSY5 −0.10 [−0.50, 0.31]
PTS1 −0.37 [−0.76, 0.02]
QIM2 −0.10 [−0.43, 0.23]
QIMR −0.28 [−0.55, −0.00]
RCOG 0.15 [−0.06, 0.37]
RING 0.08 [−0.60, 0.76]
SEEP 0.15 [−0.22, 0.51]
STRO 0.12 [−0.16, 0.39]
TRAC −0.12 [−0.87, 0.63]
UKBB −0.05 [−0.07, −0.02]
VAKN −0.13 [−0.35, 0.10]
VETS −0.15 [−0.43, 0.12]
WACH −0.20 [−1.06, 0.66]
WRBY 0.28 [−0.16, 0.73]
WTCM 0.26 [−0.42, 0.95]
WTCS 0.06 [−0.17, 0.30]
F2.5 Meta −0.04 [−0.06, −0.02]
EHR
BIOM 0.15 [−0.11, 0.41]
DAI2 −0.04 [−0.08, 0.00]
ESBB −0.05 [−0.08, −0.01]
FING −0.07 [−0.12, −0.02]
HUNT −0.08 [−0.16, 0.01]
MAYO −0.02 [−0.17, 0.13]
MGBB −0.07 [−0.14, 0.00]
SWED −0.09 [−0.35, 0.16]
UKB2 −0.05 [−0.10, −0.00]
EHR Meta −0.05 [−0.07, −0.03]
MVP
MVP1 −0.03 [−0.05, −0.01]
Freeze 3 Meta −0.04 [−0.05, −0.03]
−2
−1
0
1
2
Observed Outcome

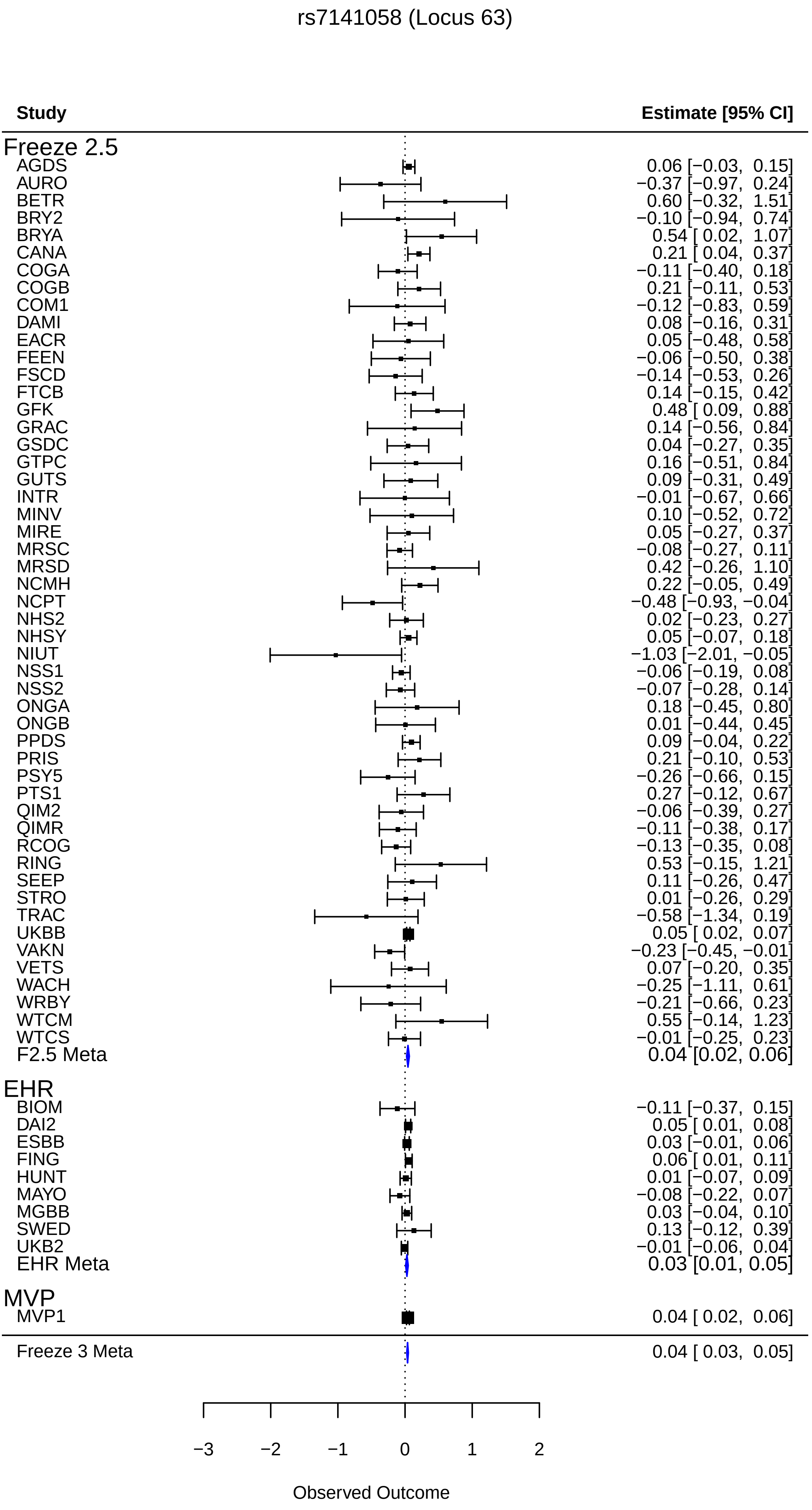
rs7141058 (Locus 63)
Study
Estimate [95% CI]
Freeze 2.5
AGDS 0.06 [−0.03, 0.15]
AURO −0.37 [−0.97, 0.24]
BETR 0.60 [−0.32, 1.51]
BRY2 −0.10 [−0.94, 0.74]
BRYA 0.54 [ 0.02, 1.07]
CANA 0.21 [ 0.04, 0.37]
COGA −0.11 [−0.40, 0.18]
COGB 0.21 [−0.11, 0.53]
COM1 −0.12 [−0.83, 0.59]
DAMI 0.08 [−0.16, 0.31]
EACR 0.05 [−0.48, 0.58]
FEEN −0.06 [−0.50, 0.38]
FSCD −0.14 [−0.53, 0.26]
FTCB 0.14 [−0.15, 0.42]
GFK 0.48 [ 0.09, 0.88]
GRAC 0.14 [−0.56, 0.84]
GSDC 0.04 [−0.27, 0.35]
GTPC 0.16 [−0.51, 0.84]
GUTS 0.09 [−0.31, 0.49]
INTR −0.01 [−0.67, 0.66]
MINV 0.10 [−0.52, 0.72]
MIRE 0.05 [−0.27, 0.37]
MRSC −0.08 [−0.27, 0.11]
MRSD 0.42 [−0.26, 1.10]
NCMH 0.22 [−0.05, 0.49]
NCPT −0.48 [−0.93, −0.04]
NHS2 0.02 [−0.23, 0.27]
NHSY 0.05 [−0.07, 0.18]
NIUT −1.03 [−2.01, −0.05]
NSS1 −0.06 [−0.19, 0.08]
NSS2 −0.07 [−0.28, 0.14]
ONGA 0.18 [−0.45, 0.80]
ONGB 0.01 [−0.44, 0.45]
PPDS 0.09 [−0.04, 0.22]
PRIS 0.21 [−0.10, 0.53]
PSY5 −0.26 [−0.66, 0.15]
PTS1 0.27 [−0.12, 0.67]
QIM2 −0.06 [−0.39, 0.27]
QIMR −0.11 [−0.38, 0.17]
RCOG −0.13 [−0.35, 0.08]
RING 0.53 [−0.15, 1.21]
SEEP 0.11 [−0.26, 0.47]
STRO 0.01 [−0.26, 0.29]
TRAC −0.58 [−1.34, 0.19]
UKBB 0.05 [ 0.02, 0.07]
VAKN −0.23 [−0.45, −0.01]
VETS 0.07 [−0.20, 0.35]
WACH −0.25 [−1.11, 0.61]
WRBY −0.21 [−0.66, 0.23]
WTCM 0.55 [−0.14, 1.23]
WTCS −0.01 [−0.25, 0.23]
F2.5 Meta 0.04 [0.02, 0.06]
EHR
BIOM −0.11 [−0.37, 0.15]
DAI2 0.05 [ 0.01, 0.08]
ESBB 0.03 [−0.01, 0.06]
FING 0.06 [ 0.01, 0.11]
HUNT 0.01 [−0.07, 0.09]
MAYO −0.08 [−0.22, 0.07]
MGBB 0.03 [−0.04, 0.10]
SWED 0.13 [−0.12, 0.39]
UKB2 −0.01 [−0.06, 0.04]
EHR Meta 0.03 [0.01, 0.05]
MVP
MVP1 0.04 [ 0.02, 0.06]
Freeze 3 Meta 0.04 [ 0.03, 0.05]
−3
−2
−1
0
1
2
Observed Outcome

rs10132977 (Locus 64)

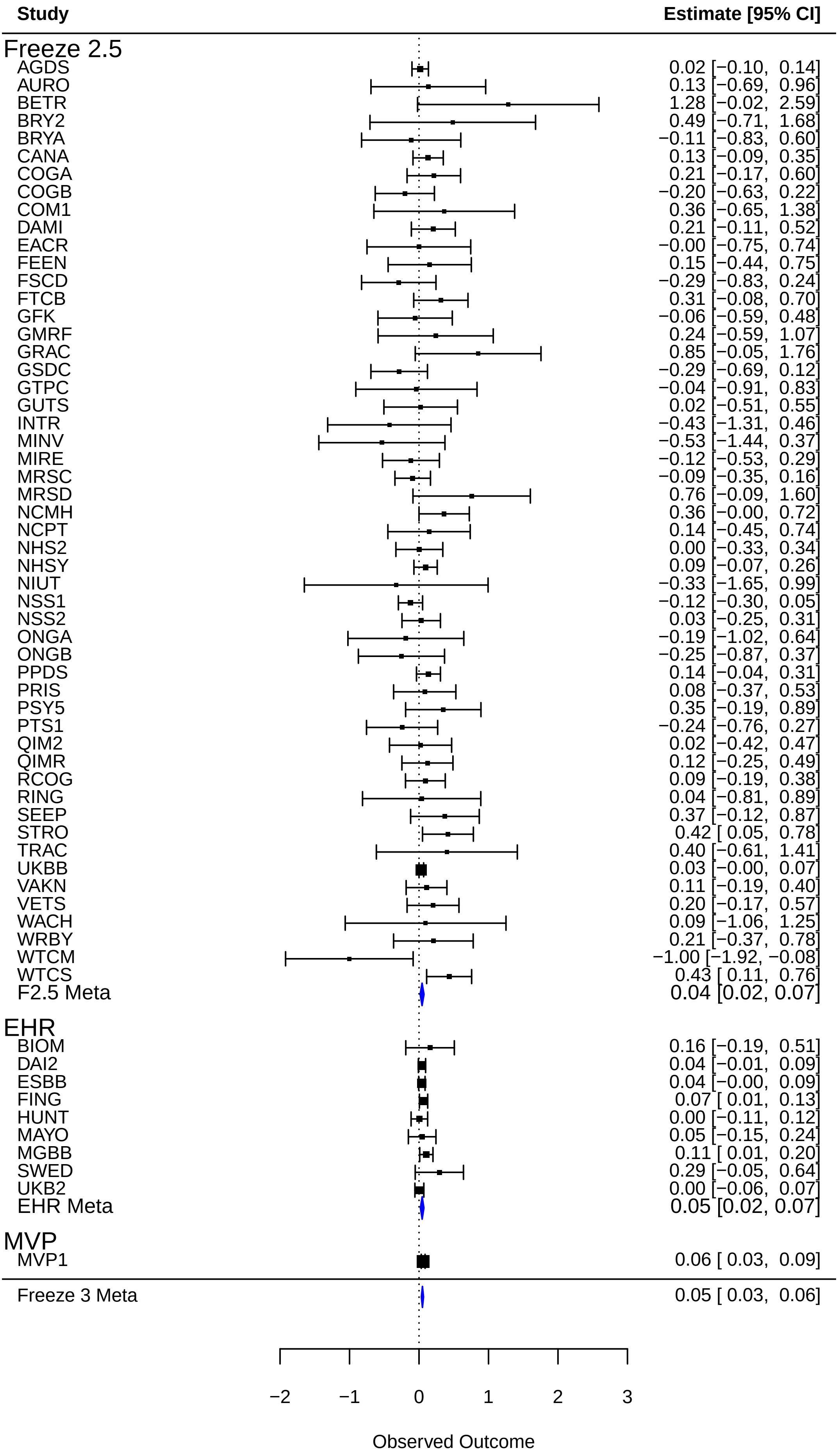

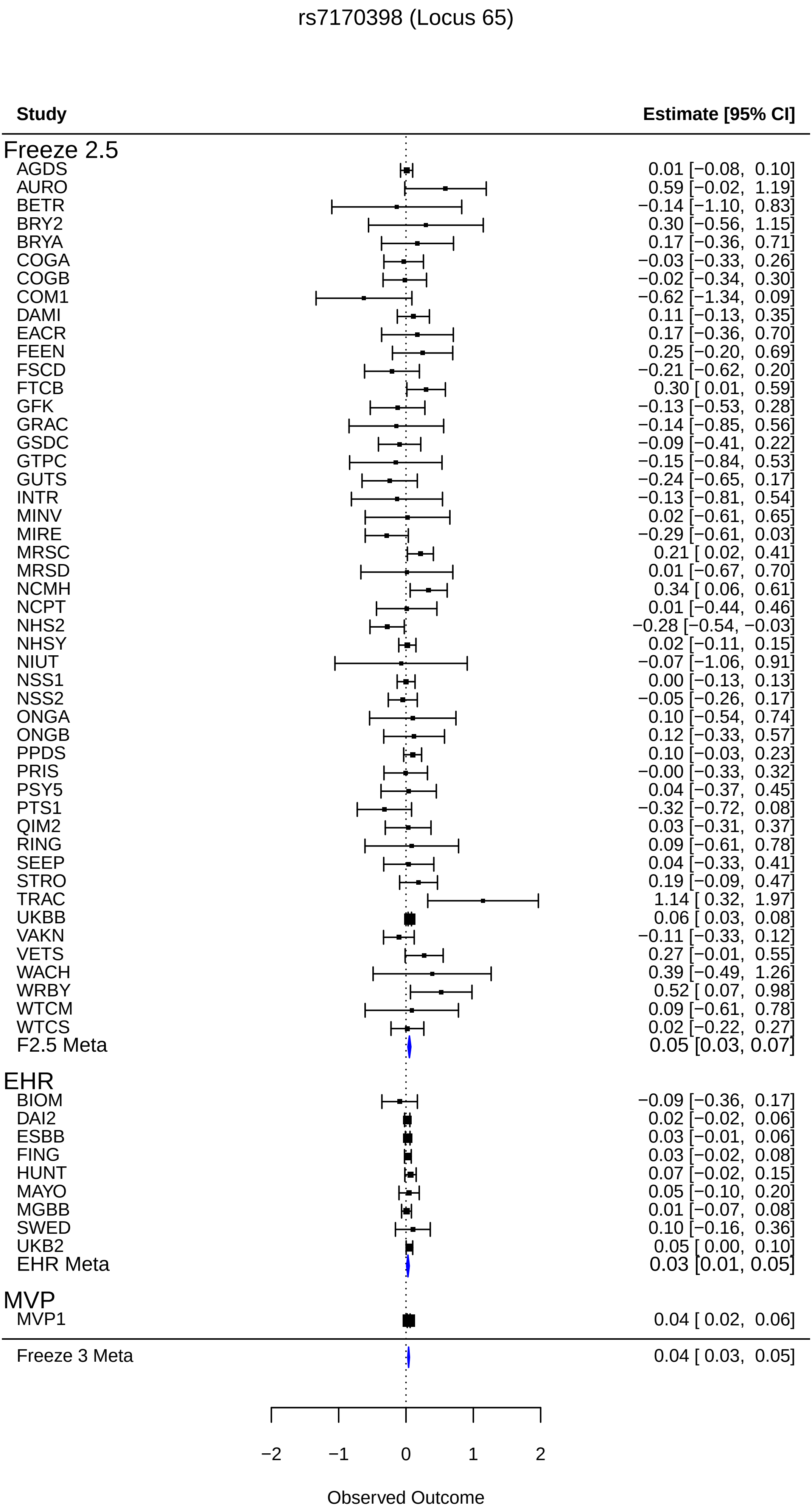
rs7170398 (Locus 65)
Study
Estimate [95% CI]
Freeze 2.5
AGDS 0.01 [−0.08, 0.10]
AURO 0.59 [−0.02, 1.19]
BETR −0.14 [−1.10, 0.83]
BRY2 0.30 [−0.56, 1.15]
BRYA 0.17 [−0.36, 0.71]
COGA −0.03 [−0.33, 0.26]
COGB −0.02 [−0.34, 0.30]
COM1 −0.62 [−1.34, 0.09]
DAMI 0.11 [−0.13, 0.35]
EACR 0.17 [−0.36, 0.70]
FEEN 0.25 [−0.20, 0.69]
FSCD −0.21 [−0.62, 0.20]
FTCB 0.30 [ 0.01, 0.59]
GFK −0.13 [−0.53, 0.28]
GRAC −0.14 [−0.85, 0.56]
GSDC −0.09 [−0.41, 0.22]
GTPC −0.15 [−0.84, 0.53]
GUTS −0.24 [−0.65, 0.17]
INTR −0.13 [−0.81, 0.54]
MINV 0.02 [−0.61, 0.65]
MIRE −0.29 [−0.61, 0.03]
MRSC 0.21 [ 0.02, 0.41]
MRSD 0.01 [−0.67, 0.70]
NCMH 0.34 [ 0.06, 0.61]
NCPT 0.01 [−0.44, 0.46]
NHS2 −0.28 [−0.54, −0.03]
NHSY 0.02 [−0.11, 0.15]
NIUT −0.07 [−1.06, 0.91]
NSS1 0.00 [−0.13, 0.13]
NSS2 −0.05 [−0.26, 0.17]
ONGA 0.10 [−0.54, 0.74]
ONGB 0.12 [−0.33, 0.57]
PPDS 0.10 [−0.03, 0.23]
PRIS −0.00 [−0.33, 0.32]
PSY5 0.04 [−0.37, 0.45]
PTS1 −0.32 [−0.72, 0.08]
QIM2 0.03 [−0.31, 0.37]
RING 0.09 [−0.61, 0.78]
SEEP 0.04 [−0.33, 0.41]
STRO 0.19 [−0.09, 0.47]
TRAC 1.14 [ 0.32, 1.97]
UKBB 0.06 [ 0.03, 0.08]
VAKN −0.11 [−0.33, 0.12]
VETS 0.27 [−0.01, 0.55]
WACH 0.39 [−0.49, 1.26]
WRBY 0.52 [ 0.07, 0.98]
WTCM 0.09 [−0.61, 0.78]
WTCS 0.02 [−0.22, 0.27]
F2.5 Meta 0.05 [0.03, 0.07]
EHR
BIOM −0.09 [−0.36, 0.17]
DAI2 0.02 [−0.02, 0.06]
ESBB 0.03 [−0.01, 0.06]
FING 0.03 [−0.02, 0.08]
HUNT 0.07 [−0.02, 0.15]
MAYO 0.05 [−0.10, 0.20]
MGBB 0.01 [−0.07, 0.08]
SWED 0.10 [−0.16, 0.36]
UKB2 0.05 [ 0.00, 0.10]
EHR Meta 0.03 [0.01, 0.05]
MVP
MVP1 0.04 [ 0.02, 0.06]
Freeze 3 Meta 0.04 [ 0.03, 0.05]
−2
−1
0
1
2
Observed Outcome

rs17514846 (Locus 66)

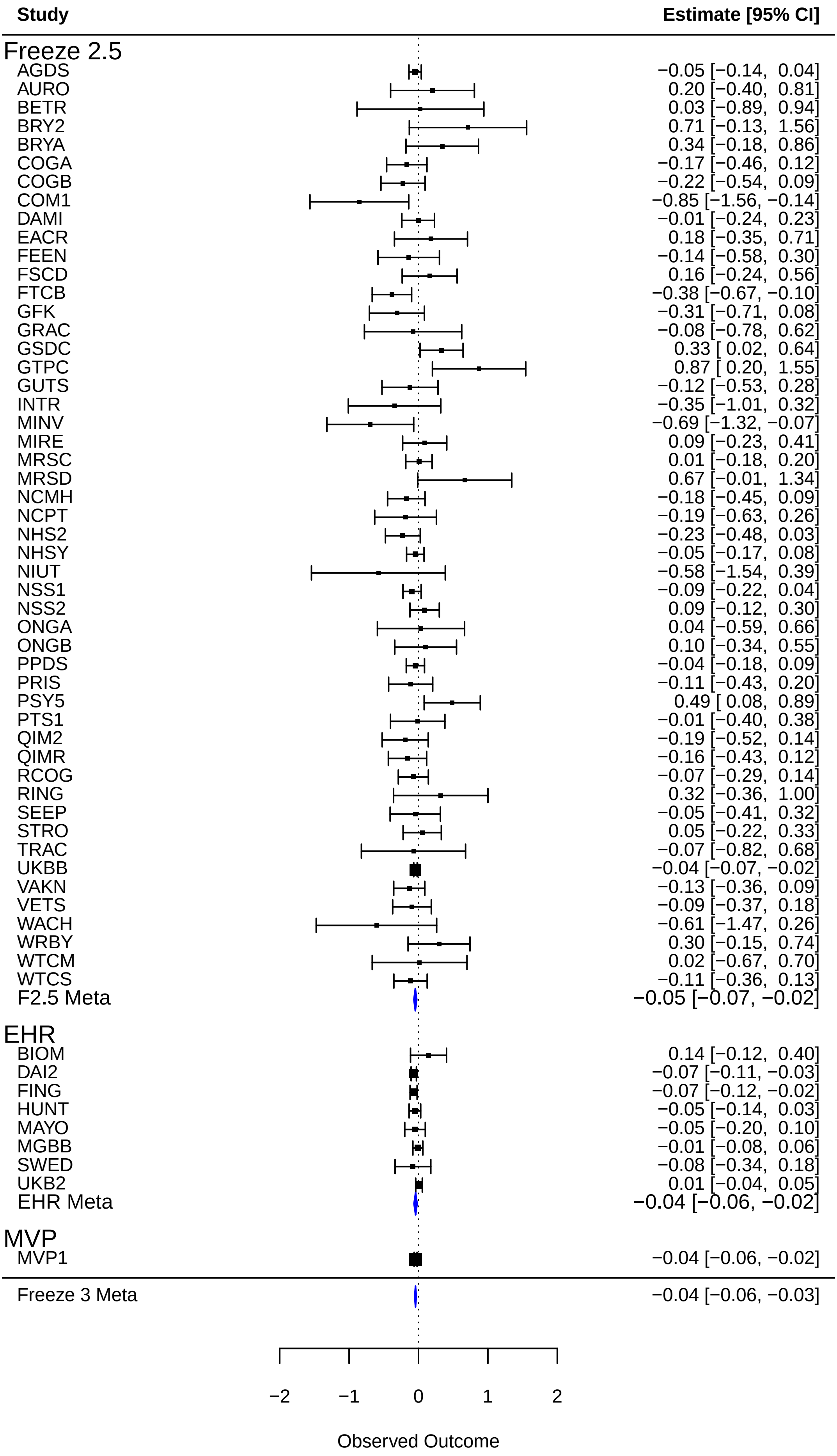

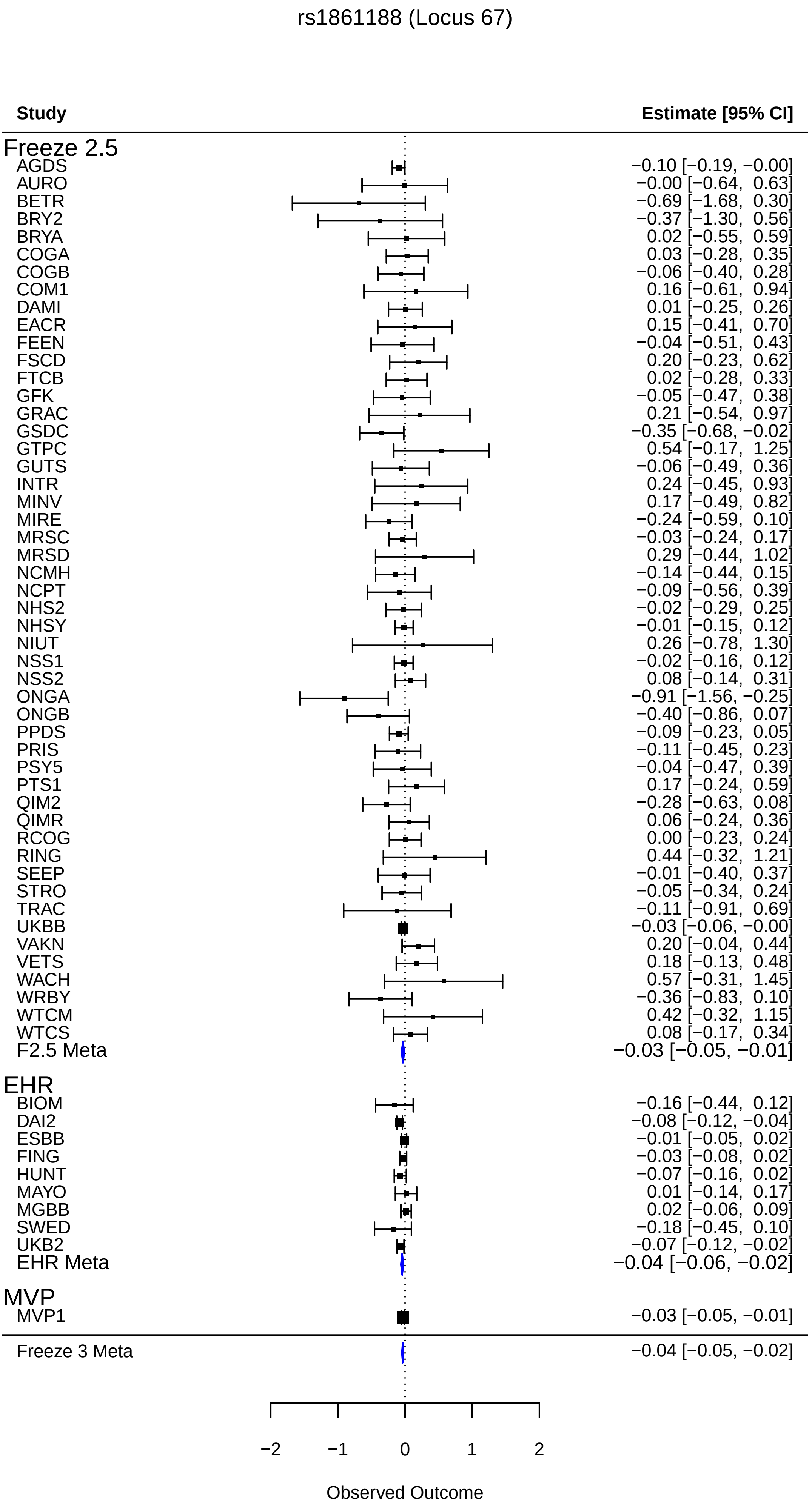
rs1861188 (Locus 67)
Study
Estimate [95% CI]
Freeze 2.5
AGDS −0.10 [−0.19, −0.00]
AURO −0.00 [−0.64, 0.63]
BETR −0.69 [−1.68, 0.30]
BRY2 −0.37 [−1.30, 0.56]
BRYA 0.02 [−0.55, 0.59]
COGA 0.03 [−0.28, 0.35]
COGB −0.06 [−0.40, 0.28]
COM1 0.16 [−0.61, 0.94]
DAMI 0.01 [−0.25, 0.26]
EACR 0.15 [−0.41, 0.70]
FEEN −0.04 [−0.51, 0.43]
FSCD 0.20 [−0.23, 0.62]
FTCB 0.02 [−0.28, 0.33]
GFK −0.05 [−0.47, 0.38]
GRAC 0.21 [−0.54, 0.97]
GSDC −0.35 [−0.68, −0.02]
GTPC 0.54 [−0.17, 1.25]
GUTS −0.06 [−0.49, 0.36]
INTR 0.24 [−0.45, 0.93]
MINV 0.17 [−0.49, 0.82]
MIRE −0.24 [−0.59, 0.10]
MRSC −0.03 [−0.24, 0.17]
MRSD 0.29 [−0.44, 1.02]
NCMH −0.14 [−0.44, 0.15]
NCPT −0.09 [−0.56, 0.39]
NHS2 −0.02 [−0.29, 0.25]
NHSY −0.01 [−0.15, 0.12]
NIUT 0.26 [−0.78, 1.30]
NSS1 −0.02 [−0.16, 0.12]
NSS2 0.08 [−0.14, 0.31]
ONGA −0.91 [−1.56, −0.25]
ONGB −0.40 [−0.86, 0.07]
PPDS −0.09 [−0.23, 0.05]
PRIS −0.11 [−0.45, 0.23]
PSY5 −0.04 [−0.47, 0.39]
PTS1 0.17 [−0.24, 0.59]
QIM2 −0.28 [−0.63, 0.08]
QIMR 0.06 [−0.24, 0.36]
RCOG 0.00 [−0.23, 0.24]
RING 0.44 [−0.32, 1.21]
SEEP −0.01 [−0.40, 0.37]
STRO −0.05 [−0.34, 0.24]
TRAC −0.11 [−0.91, 0.69]
UKBB −0.03 [−0.06, −0.00]
VAKN 0.20 [−0.04, 0.44]
VETS 0.18 [−0.13, 0.48]
WACH 0.57 [−0.31, 1.45]
WRBY −0.36 [−0.83, 0.10]
WTCM 0.42 [−0.32, 1.15]
WTCS 0.08 [−0.17, 0.34]
F2.5 Meta −0.03 [−0.05, −0.01]
EHR
BIOM −0.16 [−0.44, 0.12]
DAI2 −0.08 [−0.12, −0.04]
ESBB −0.01 [−0.05, 0.02]
FING −0.03 [−0.08, 0.02]
HUNT −0.07 [−0.16, 0.02]
MAYO 0.01 [−0.14, 0.17]
MGBB 0.02 [−0.06, 0.09]
SWED −0.18 [−0.45, 0.10]
UKB2 −0.07 [−0.12, −0.02]
EHR Meta −0.04 [−0.06, −0.02]
MVP
MVP1 −0.03 [−0.05, −0.01]
Freeze 3 Meta −0.04 [−0.05, −0.02]
−2
−1
0
1
2
Observed Outcome

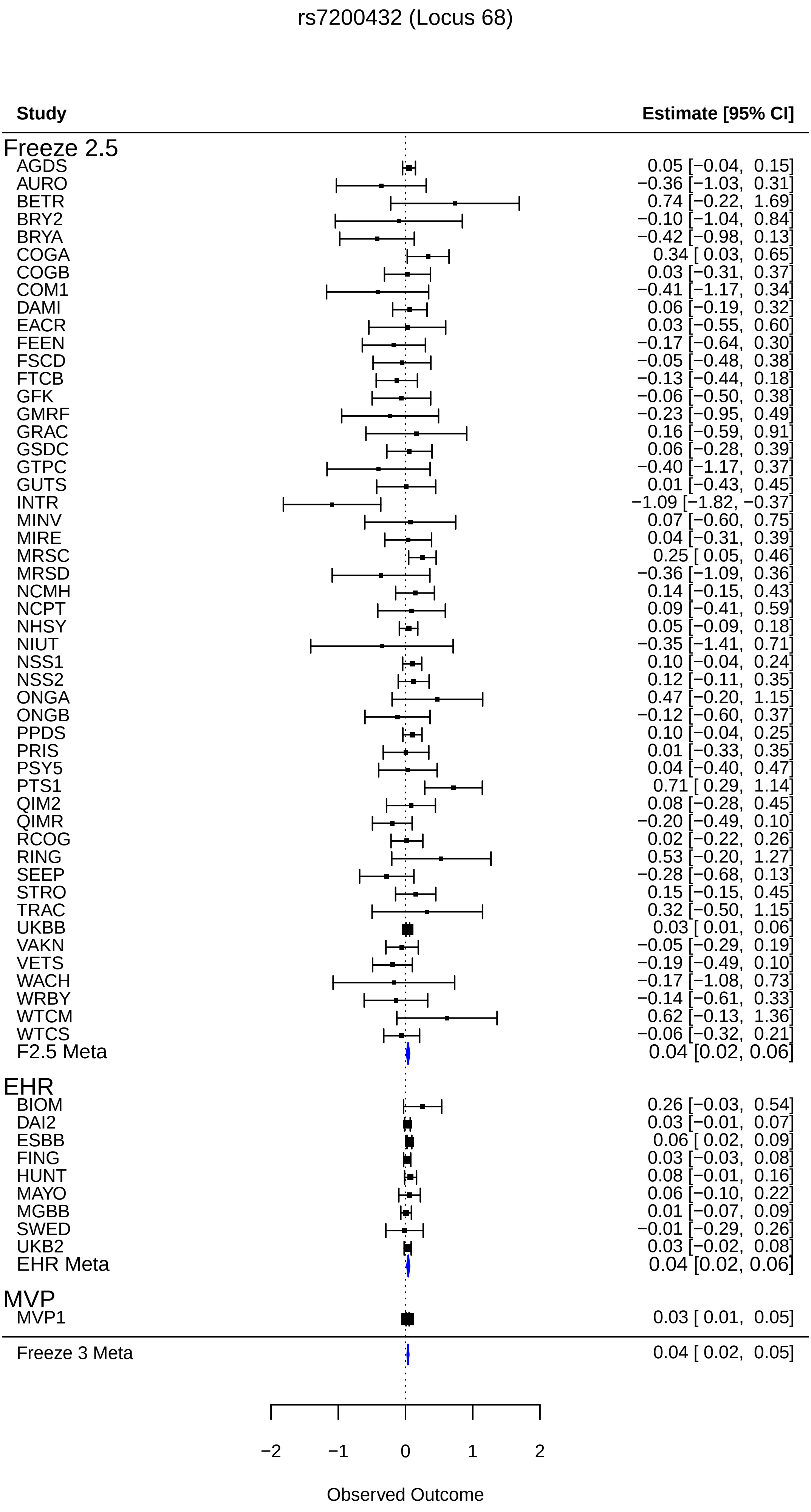
rs7200432 (Locus 68)
Study
Estimate [95% CI]
Freeze 2.5
AGDS 0.05 [−0.04, 0.15]
AURO −0.36 [−1.03, 0.31]
BETR 0.74 [−0.22, 1.69]
BRY2 −0.10 [−1.04, 0.84]
BRYA −0.42 [−0.98, 0.13]
COGA 0.34 [ 0.03, 0.65]
COGB 0.03 [−0.31, 0.37]
COM1 −0.41 [−1.17, 0.34]
DAMI 0.06 [−0.19, 0.32]
EACR 0.03 [−0.55, 0.60]
FEEN −0.17 [−0.64, 0.30]
FSCD −0.05 [−0.48, 0.38]
FTCB −0.13 [−0.44, 0.18]
GFK −0.06 [−0.50, 0.38]
GMRF −0.23 [−0.95, 0.49]
GRAC 0.16 [−0.59, 0.91]
GSDC 0.06 [−0.28, 0.39]
GTPC −0.40 [−1.17, 0.37]
GUTS 0.01 [−0.43, 0.45]
INTR −1.09 [−1.82, −0.37]
MINV 0.07 [−0.60, 0.75]
MIRE 0.04 [−0.31, 0.39]
MRSC 0.25 [ 0.05, 0.46]
MRSD −0.36 [−1.09, 0.36]
NCMH 0.14 [−0.15, 0.43]
NCPT 0.09 [−0.41, 0.59]
NHSY 0.05 [−0.09, 0.18]
NIUT −0.35 [−1.41, 0.71]
NSS1 0.10 [−0.04, 0.24]
NSS2 0.12 [−0.11, 0.35]
ONGA 0.47 [−0.20, 1.15]
ONGB −0.12 [−0.60, 0.37]
PPDS 0.10 [−0.04, 0.25]
PRIS 0.01 [−0.33, 0.35]
PSY5 0.04 [−0.40, 0.47]
PTS1 0.71 [ 0.29, 1.14]
QIM2 0.08 [−0.28, 0.45]
QIMR −0.20 [−0.49, 0.10]
RCOG 0.02 [−0.22, 0.26]
RING 0.53 [−0.20, 1.27]
SEEP −0.28 [−0.68, 0.13]
STRO 0.15 [−0.15, 0.45]
TRAC 0.32 [−0.50, 1.15]
UKBB 0.03 [ 0.01, 0.06]
VAKN −0.05 [−0.29, 0.19]
VETS −0.19 [−0.49, 0.10]
WACH −0.17 [−1.08, 0.73]
WRBY −0.14 [−0.61, 0.33]
WTCM 0.62 [−0.13, 1.36]
WTCS −0.06 [−0.32, 0.21]
F2.5 Meta 0.04 [0.02, 0.06]
EHR
BIOM 0.26 [−0.03, 0.54]
DAI2 0.03 [−0.01, 0.07]
ESBB 0.06 [ 0.02, 0.09]
FING 0.03 [−0.03, 0.08]
HUNT 0.08 [−0.01, 0.16]
MAYO 0.06 [−0.10, 0.22]
MGBB 0.01 [−0.07, 0.09]
SWED −0.01 [−0.29, 0.26]
UKB2 0.03 [−0.02, 0.08]
EHR Meta 0.04 [0.02, 0.06]
MVP
MVP1 0.03 [ 0.01, 0.05]
Freeze 3 Meta 0.04 [ 0.02, 0.05]
−2
−1
0
1
2
Observed Outcome

rs143133717 (Locus 69)

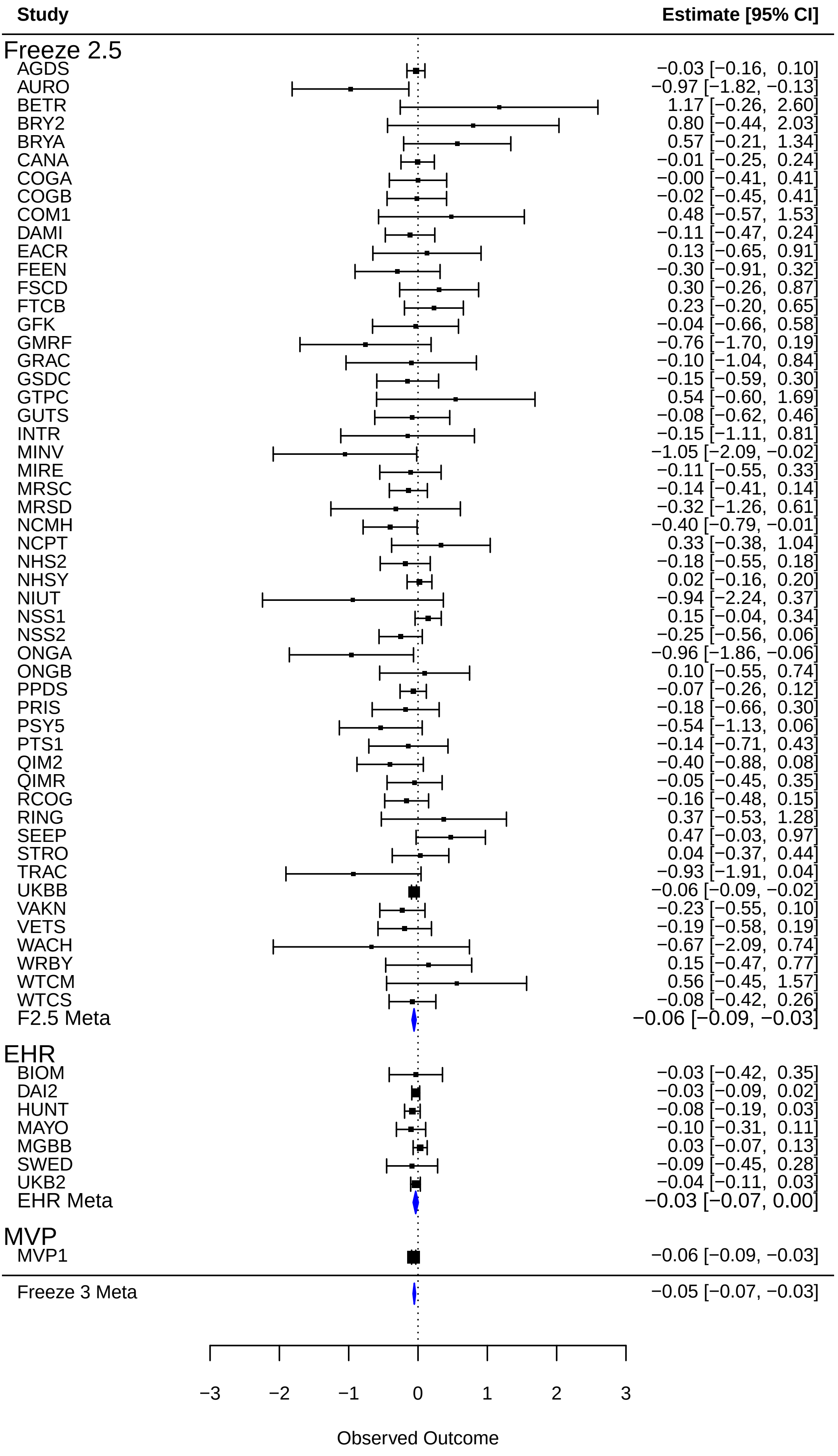

rs2684641 (Locus 70)

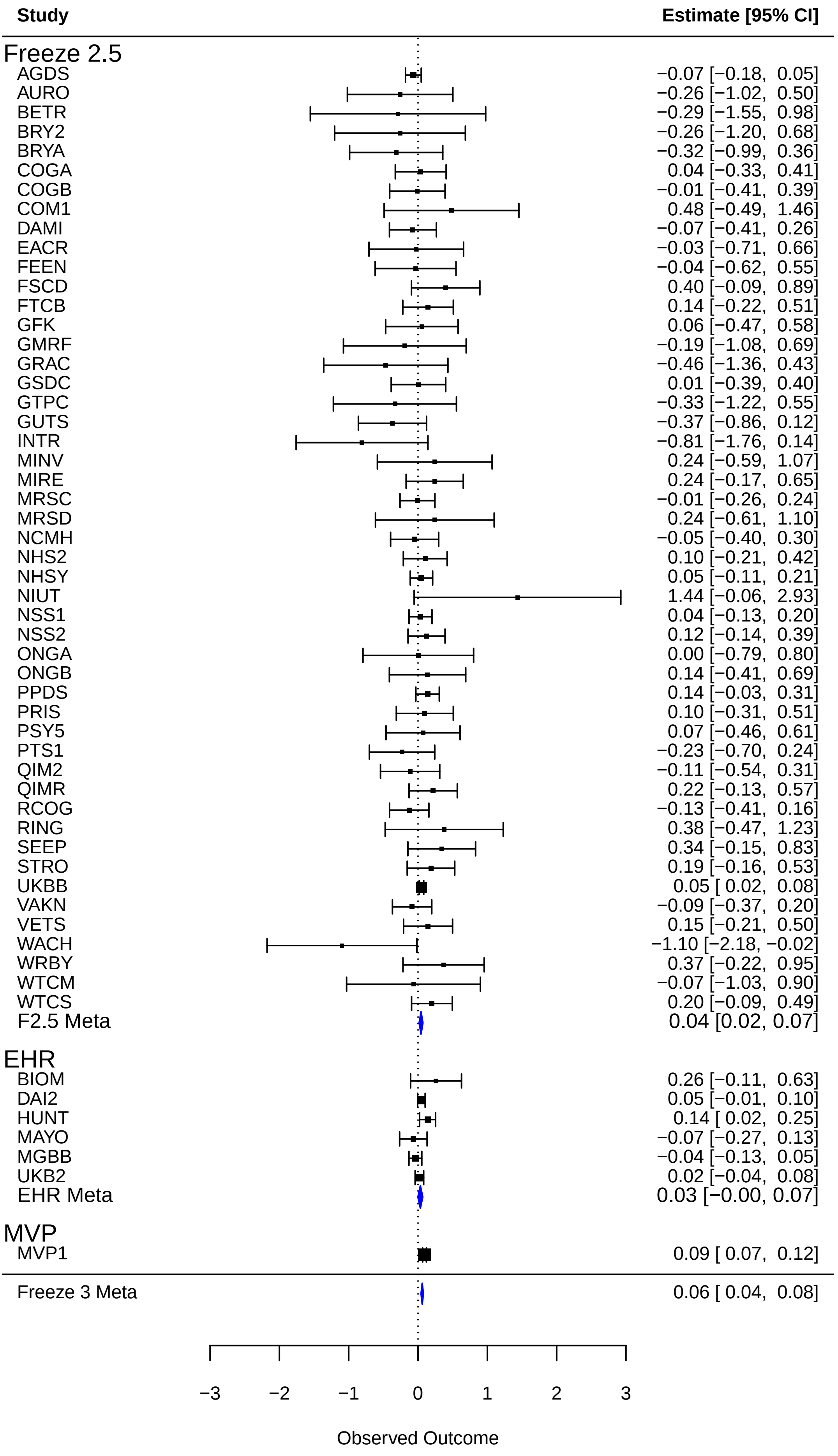

rs73338706 (Locus 71)

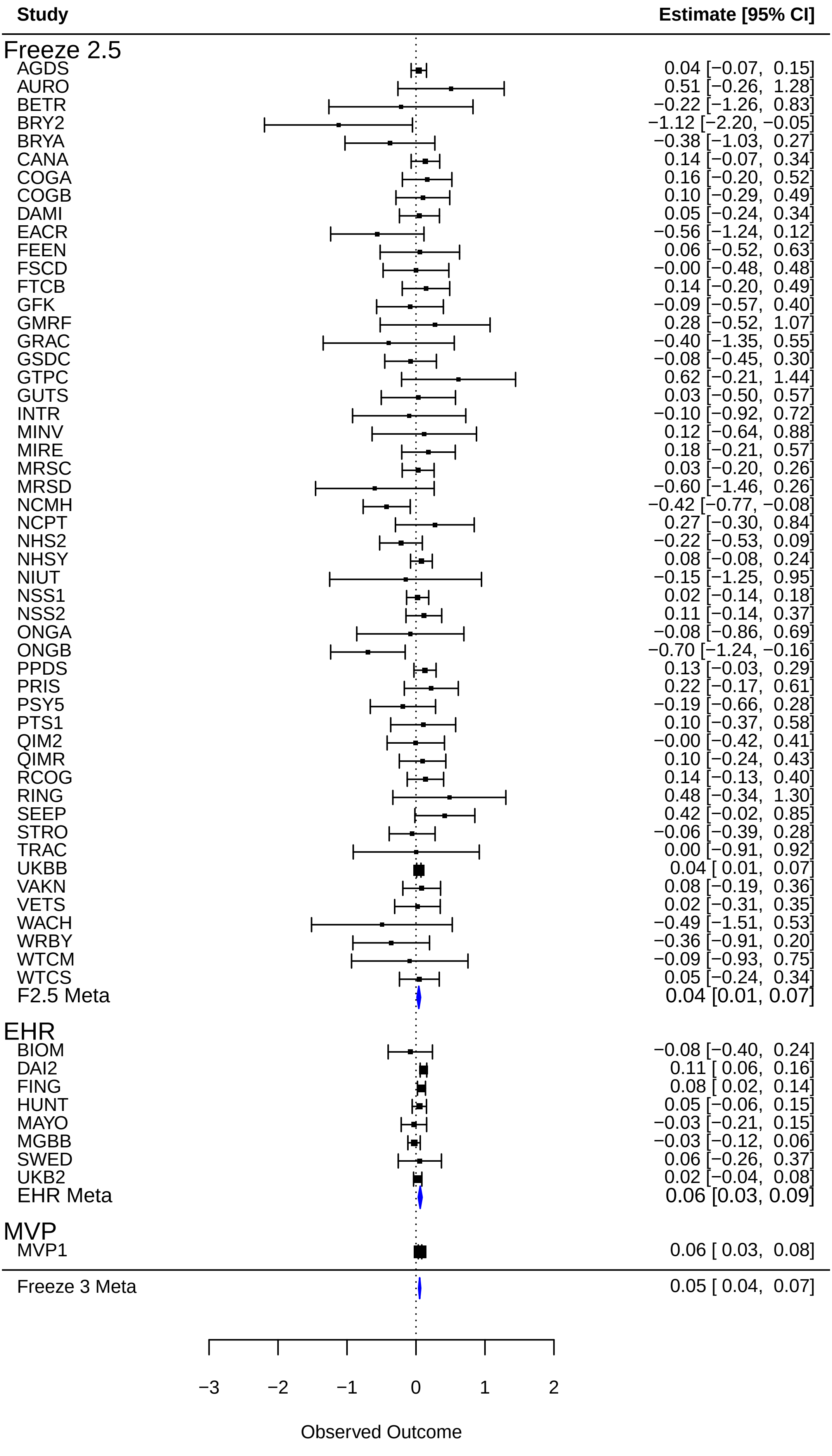

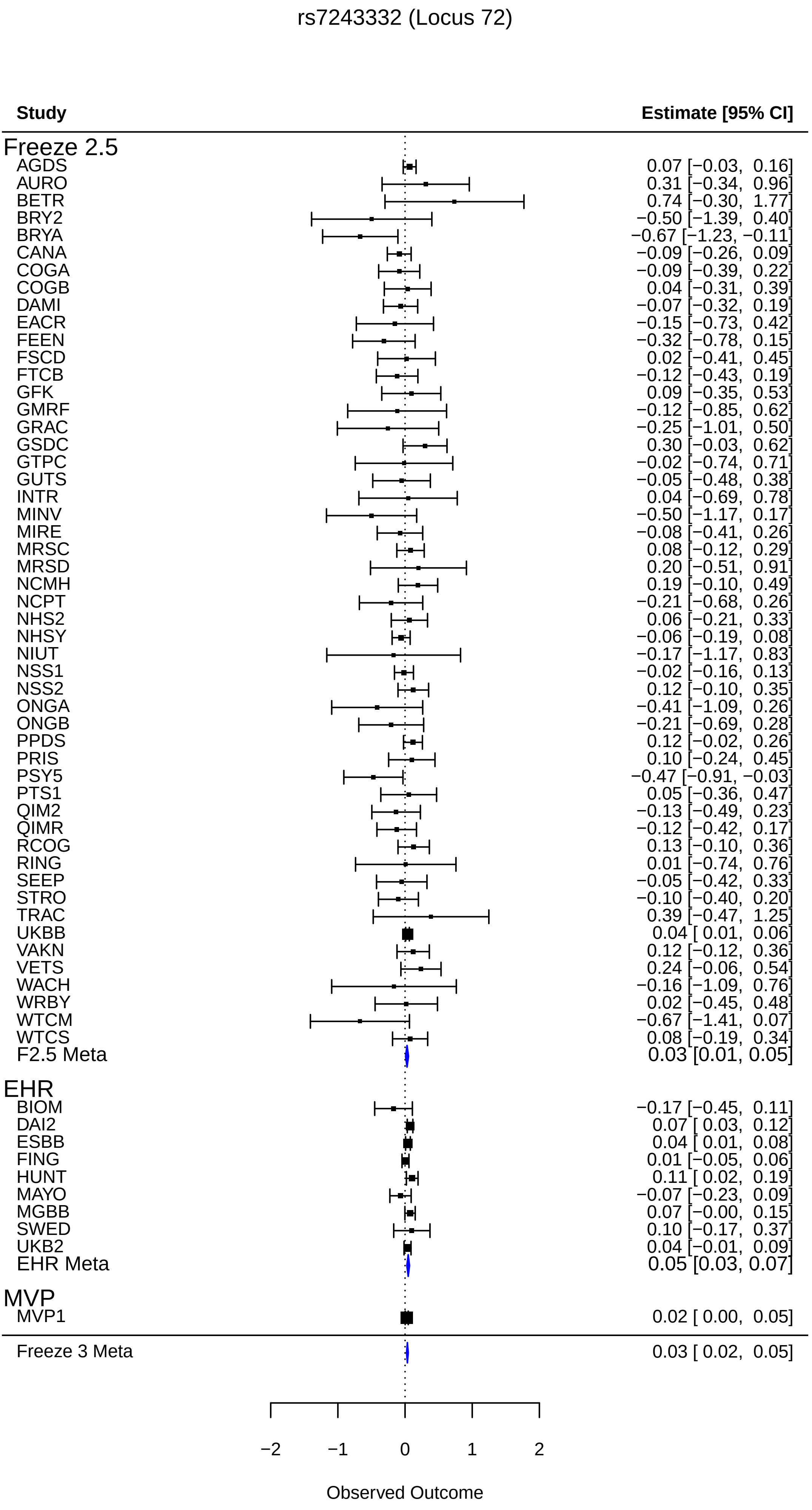
rs7243332 (Locus 72)
Study
Estimate [95% CI]
Freeze 2.5
AGDS 0.07 [−0.03, 0.16]
AURO 0.31 [−0.34, 0.96]
BETR 0.74 [−0.30, 1.77]
BRY2 −0.50 [−1.39, 0.40]
BRYA −0.67 [−1.23, −0.11]
CANA −0.09 [−0.26, 0.09]
COGA −0.09 [−0.39, 0.22]
COGB 0.04 [−0.31, 0.39]
DAMI −0.07 [−0.32, 0.19]
EACR −0.15 [−0.73, 0.42]
FEEN −0.32 [−0.78, 0.15]
FSCD 0.02 [−0.41, 0.45]
FTCB −0.12 [−0.43, 0.19]
GFK 0.09 [−0.35, 0.53]
GMRF −0.12 [−0.85, 0.62]
GRAC −0.25 [−1.01, 0.50]
GSDC 0.30 [−0.03, 0.62]
GTPC −0.02 [−0.74, 0.71]
GUTS −0.05 [−0.48, 0.38]
INTR 0.04 [−0.69, 0.78]
MINV −0.50 [−1.17, 0.17]
MIRE −0.08 [−0.41, 0.26]
MRSC 0.08 [−0.12, 0.29]
MRSD 0.20 [−0.51, 0.91]
NCMH 0.19 [−0.10, 0.49]
NCPT −0.21 [−0.68, 0.26]
NHS2 0.06 [−0.21, 0.33]
NHSY −0.06 [−0.19, 0.08]
NIUT −0.17 [−1.17, 0.83]
NSS1 −0.02 [−0.16, 0.13]
NSS2 0.12 [−0.10, 0.35]
ONGA −0.41 [−1.09, 0.26]
ONGB −0.21 [−0.69, 0.28]
PPDS 0.12 [−0.02, 0.26]
PRIS 0.10 [−0.24, 0.45]
PSY5 −0.47 [−0.91, −0.03]
PTS1 0.05 [−0.36, 0.47]
QIM2 −0.13 [−0.49, 0.23]
QIMR −0.12 [−0.42, 0.17]
RCOG 0.13 [−0.10, 0.36]
RING 0.01 [−0.74, 0.76]
SEEP −0.05 [−0.42, 0.33]
STRO −0.10 [−0.40, 0.20]
TRAC 0.39 [−0.47, 1.25]
UKBB 0.04 [ 0.01, 0.06]
VAKN 0.12 [−0.12, 0.36]
VETS 0.24 [−0.06, 0.54]
WACH −0.16 [−1.09, 0.76]
WRBY 0.02 [−0.45, 0.48]
WTCM −0.67 [−1.41, 0.07]
WTCS 0.08 [−0.19, 0.34]
F2.5 Meta 0.03 [0.01, 0.05]
EHR
BIOM −0.17 [−0.45, 0.11]
DAI2 0.07 [ 0.03, 0.12]
ESBB 0.04 [ 0.01, 0.08]
FING 0.01 [−0.05, 0.06]
HUNT 0.11 [ 0.02, 0.19]
MAYO −0.07 [−0.23, 0.09]
MGBB 0.07 [−0.00, 0.15]
SWED 0.10 [−0.17, 0.37]
UKB2 0.04 [−0.01, 0.09]
EHR Meta 0.05 [0.03, 0.07]
MVP
MVP1 0.02 [ 0.00, 0.05]
Freeze 3 Meta 0.03 [ 0.02, 0.05]
−2
−1
0
1
2
Observed Outcome

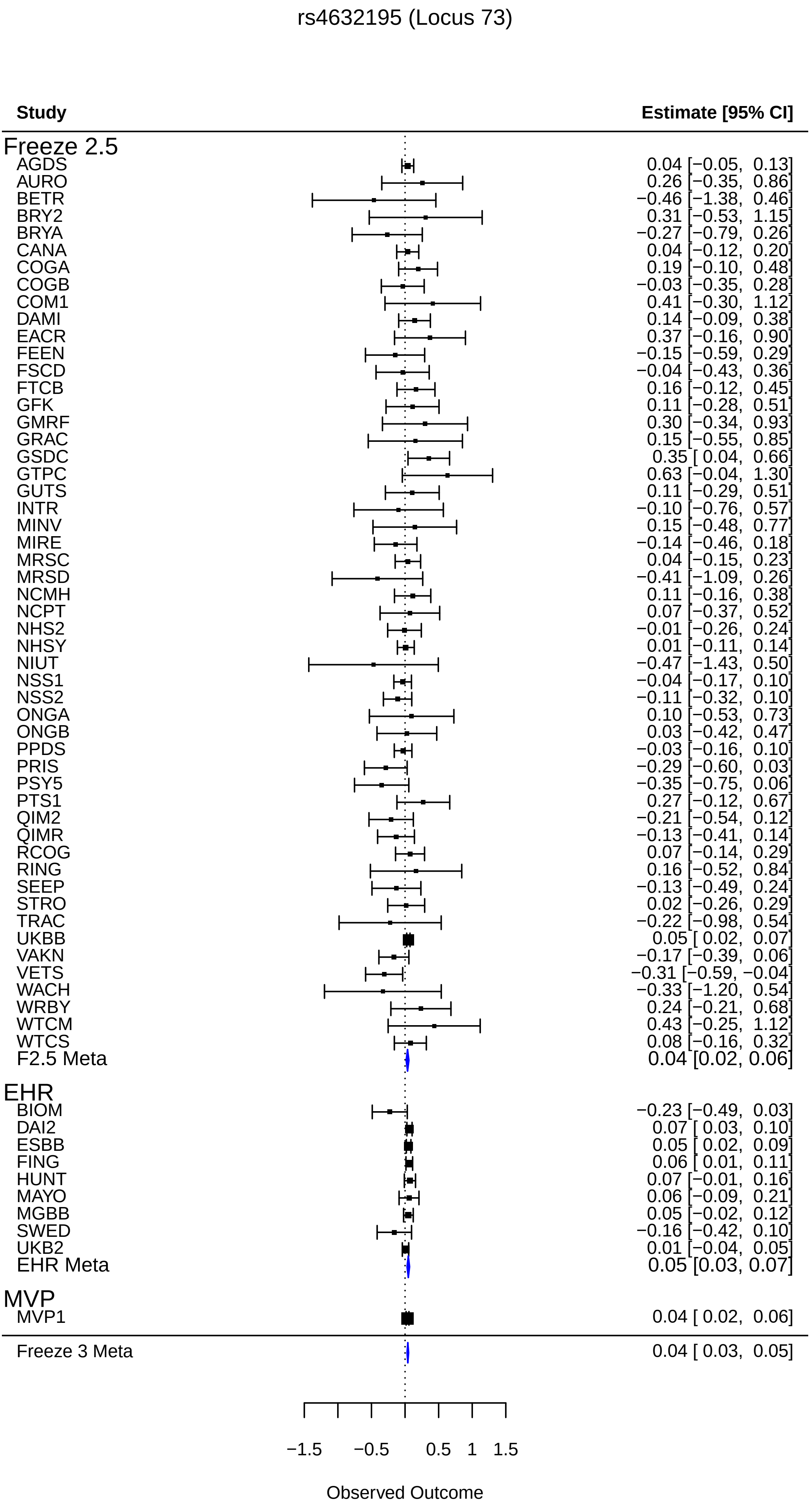
rs4632195 (Locus 73)
Study
Estimate [95% CI]
Freeze 2.5
AGDS 0.04 [−0.05, 0.13]
AURO 0.26 [−0.35, 0.86]
BETR −0.46 [−1.38, 0.46]
BRY2 0.31 [−0.53, 1.15]
BRYA −0.27 [−0.79, 0.26]
CANA 0.04 [−0.12, 0.20]
COGA 0.19 [−0.10, 0.48]
COGB −0.03 [−0.35, 0.28]
COM1 0.41 [−0.30, 1.12]
DAMI 0.14 [−0.09, 0.38]
EACR 0.37 [−0.16, 0.90]
FEEN −0.15 [−0.59, 0.29]
FSCD −0.04 [−0.43, 0.36]
FTCB 0.16 [−0.12, 0.45]
GFK 0.11 [−0.28, 0.51]
GMRF 0.30 [−0.34, 0.93]
GRAC 0.15 [−0.55, 0.85]
GSDC 0.35 [ 0.04, 0.66]
GTPC 0.63 [−0.04, 1.30]
GUTS 0.11 [−0.29, 0.51]
INTR −0.10 [−0.76, 0.57]
MINV 0.15 [−0.48, 0.77]
MIRE −0.14 [−0.46, 0.18]
MRSC 0.04 [−0.15, 0.23]
MRSD −0.41 [−1.09, 0.26]
NCMH 0.11 [−0.16, 0.38]
NCPT 0.07 [−0.37, 0.52]
NHS2 −0.01 [−0.26, 0.24]
NHSY 0.01 [−0.11, 0.14]
NIUT −0.47 [−1.43, 0.50]
NSS1 −0.04 [−0.17, 0.10]
NSS2 −0.11 [−0.32, 0.10]
ONGA 0.10 [−0.53, 0.73]
ONGB 0.03 [−0.42, 0.47]
PPDS −0.03 [−0.16, 0.10]
PRIS −0.29 [−0.60, 0.03]
PSY5 −0.35 [−0.75, 0.06]
PTS1 0.27 [−0.12, 0.67]
QIM2 −0.21 [−0.54, 0.12]
QIMR −0.13 [−0.41, 0.14]
RCOG 0.07 [−0.14, 0.29]
RING 0.16 [−0.52, 0.84]
SEEP −0.13 [−0.49, 0.24]
STRO 0.02 [−0.26, 0.29]
TRAC −0.22 [−0.98, 0.54]
UKBB 0.05 [ 0.02, 0.07]
VAKN −0.17 [−0.39, 0.06]
VETS −0.31 [−0.59, −0.04]
WACH −0.33 [−1.20, 0.54]
WRBY 0.24 [−0.21, 0.68]
WTCM 0.43 [−0.25, 1.12]
WTCS 0.08 [−0.16, 0.32]
F2.5 Meta 0.04 [0.02, 0.06]
EHR
BIOM −0.23 [−0.49, 0.03]
DAI2 0.07 [ 0.03, 0.10]
ESBB 0.05 [ 0.02, 0.09]
FING 0.06 [ 0.01, 0.11]
HUNT 0.07 [−0.01, 0.16]
MAYO 0.06 [−0.09, 0.21]
MGBB 0.05 [−0.02, 0.12]
SWED −0.16 [−0.42, 0.10]
UKB2 0.01 [−0.04, 0.05]
EHR Meta 0.05 [0.03, 0.07]
MVP
MVP1 0.04 [ 0.02, 0.06]
Freeze 3 Meta 0.04 [ 0.03, 0.05]
−1.5
−0.5
0.5
1
1.5
Observed Outcome

rs896686 (Locus 74)

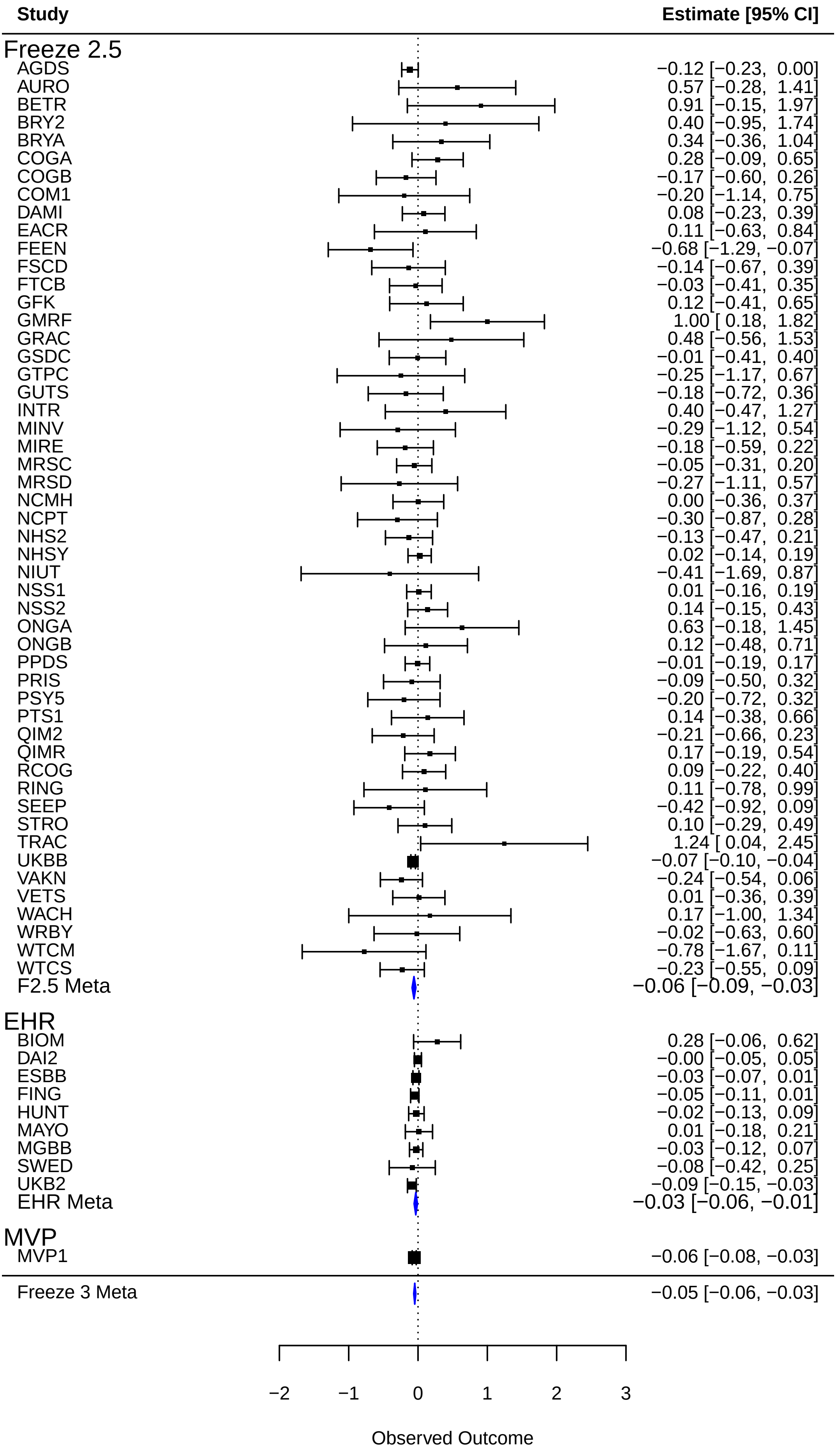

rs7408312 (Locus 75)

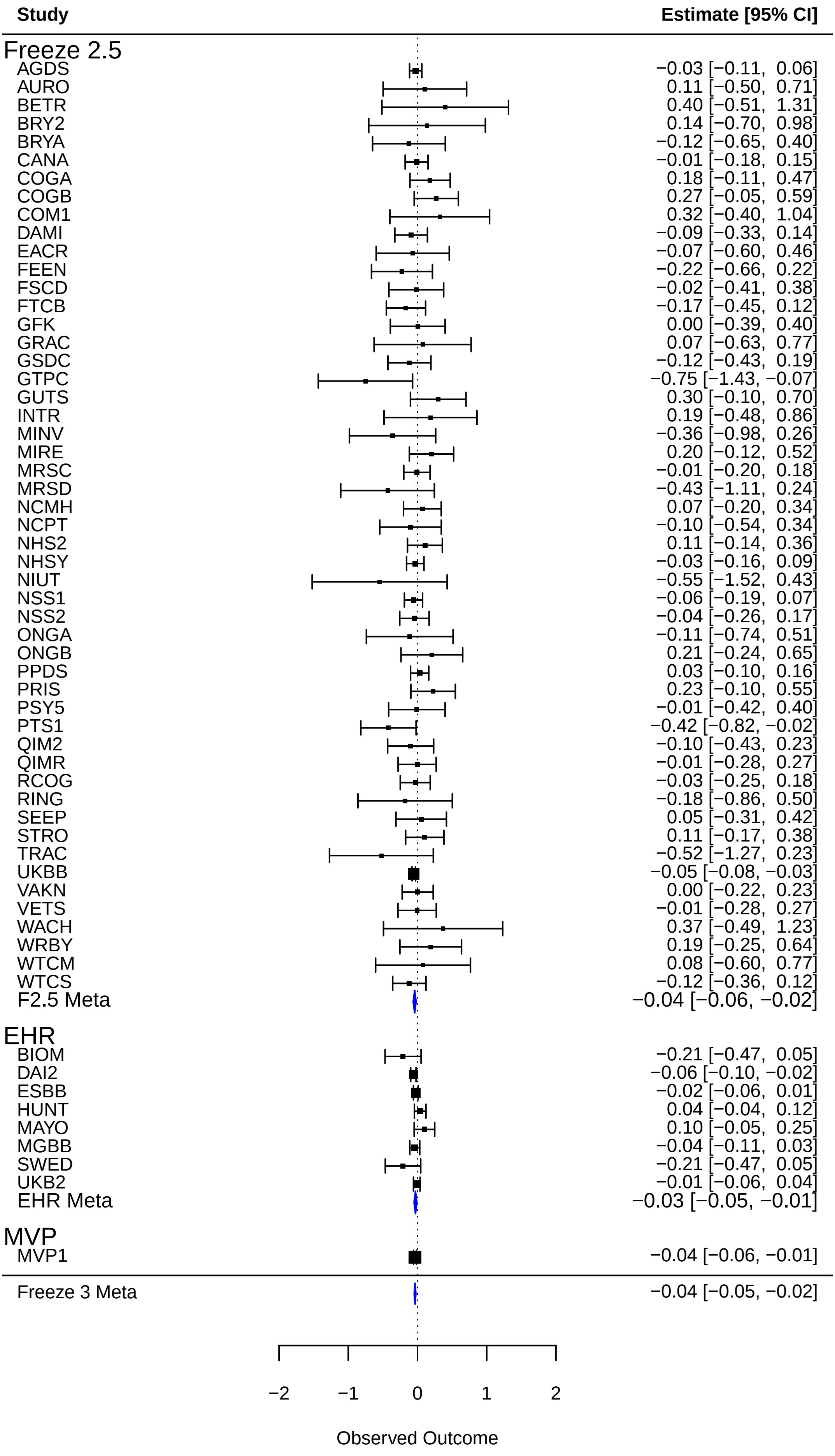

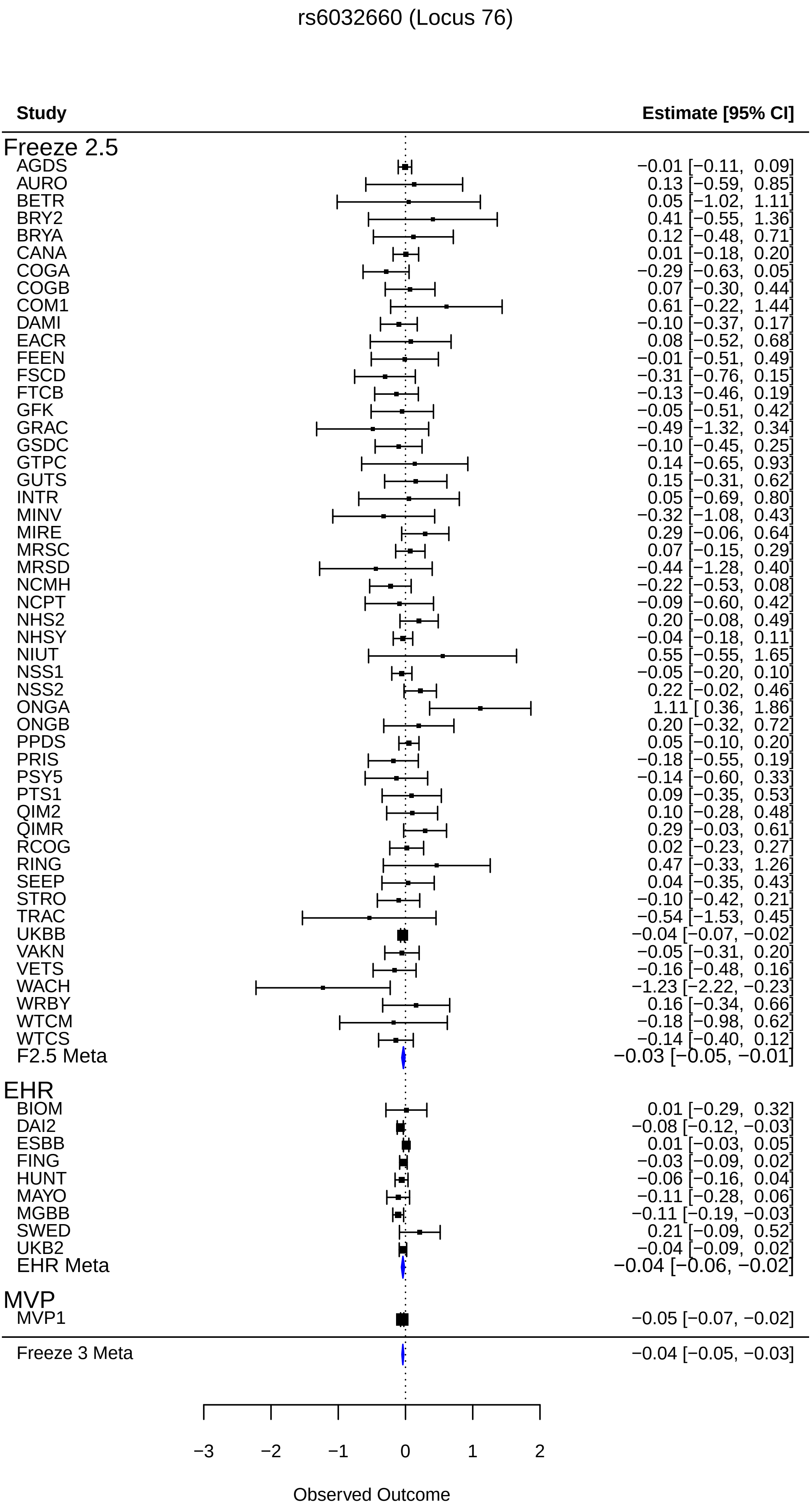
rs6032660 (Locus 76)
Study
Estimate [95% CI]
Freeze 2.5
AGDS −0.01 [−0.11, 0.09]
AURO 0.13 [−0.59, 0.85]
BETR 0.05 [−1.02, 1.11]
BRY2 0.41 [−0.55, 1.36]
BRYA 0.12 [−0.48, 0.71]
CANA 0.01 [−0.18, 0.20]
COGA −0.29 [−0.63, 0.05]
COGB 0.07 [−0.30, 0.44]
COM1 0.61 [−0.22, 1.44]
DAMI −0.10 [−0.37, 0.17]
EACR 0.08 [−0.52, 0.68]
FEEN −0.01 [−0.51, 0.49]
FSCD −0.31 [−0.76, 0.15]
FTCB −0.13 [−0.46, 0.19]
GFK −0.05 [−0.51, 0.42]
GRAC −0.49 [−1.32, 0.34]
GSDC −0.10 [−0.45, 0.25]
GTPC 0.14 [−0.65, 0.93]
GUTS 0.15 [−0.31, 0.62]
INTR 0.05 [−0.69, 0.80]
MINV −0.32 [−1.08, 0.43]
MIRE 0.29 [−0.06, 0.64]
MRSC 0.07 [−0.15, 0.29]
MRSD −0.44 [−1.28, 0.40]
NCMH −0.22 [−0.53, 0.08]
NCPT −0.09 [−0.60, 0.42]
NHS2 0.20 [−0.08, 0.49]
NHSY −0.04 [−0.18, 0.11]
NIUT 0.55 [−0.55, 1.65]
NSS1 −0.05 [−0.20, 0.10]
NSS2 0.22 [−0.02, 0.46]
ONGA 1.11 [ 0.36, 1.86]
ONGB 0.20 [−0.32, 0.72]
PPDS 0.05 [−0.10, 0.20]
PRIS −0.18 [−0.55, 0.19]
PSY5 −0.14 [−0.60, 0.33]
PTS1 0.09 [−0.35, 0.53]
QIM2 0.10 [−0.28, 0.48]
QIMR 0.29 [−0.03, 0.61]
RCOG 0.02 [−0.23, 0.27]
RING 0.47 [−0.33, 1.26]
SEEP 0.04 [−0.35, 0.43]
STRO −0.10 [−0.42, 0.21]
TRAC −0.54 [−1.53, 0.45]
UKBB −0.04 [−0.07, −0.02]
VAKN −0.05 [−0.31, 0.20]
VETS −0.16 [−0.48, 0.16]
WACH −1.23 [−2.22, −0.23]
WRBY 0.16 [−0.34, 0.66]
WTCM −0.18 [−0.98, 0.62]
WTCS −0.14 [−0.40, 0.12]
F2.5 Meta −0.03 [−0.05, −0.01]
EHR
BIOM 0.01 [−0.29, 0.32]
DAI2 −0.08 [−0.12, −0.03]
ESBB 0.01 [−0.03, 0.05]
FING −0.03 [−0.09, 0.02]
HUNT −0.06 [−0.16, 0.04]
MAYO −0.11 [−0.28, 0.06]
MGBB −0.11 [−0.19, −0.03]
SWED 0.21 [−0.09, 0.52]
UKB2 −0.04 [−0.09, 0.02]
EHR Meta −0.04 [−0.06, −0.02]
MVP
MVP1 −0.05 [−0.07, −0.02]
Freeze 3 Meta −0.04 [−0.05, −0.03]
−3
−2
−1
0
1
2
Observed Outcome

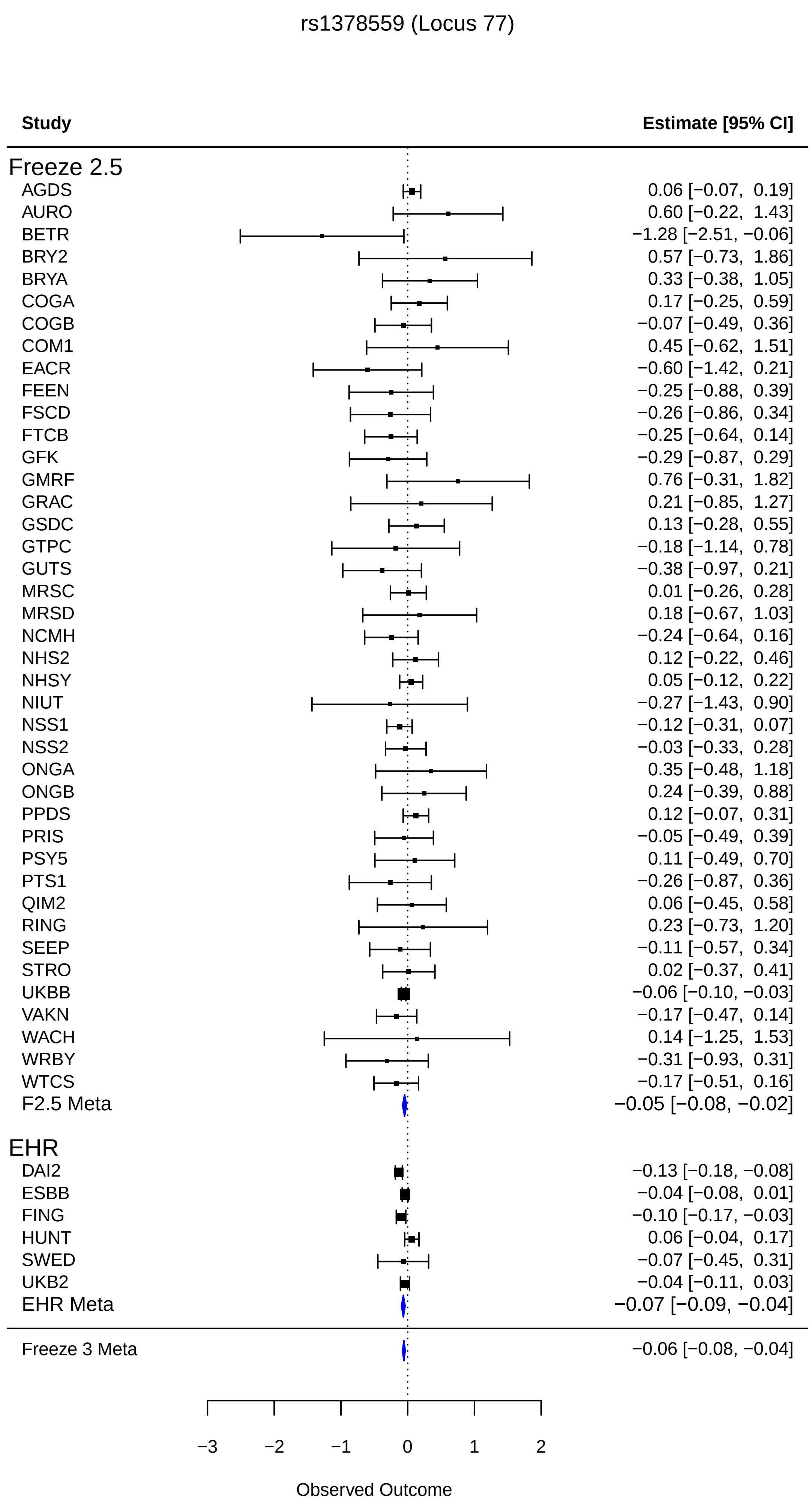
rs1378559 (Locus 77)
Study
Estimate [95% CI]
Freeze 2.5
AGDS 0.06 [−0.07, 0.19]
AURO 0.60 [−0.22, 1.43]
BETR −1.28 [−2.51, −0.06]
BRY2 0.57 [−0.73, 1.86]
BRYA 0.33 [−0.38, 1.05]
COGA 0.17 [−0.25, 0.59]
COGB −0.07 [−0.49, 0.36]
COM1 0.45 [−0.62, 1.51]
EACR −0.60 [−1.42, 0.21]
FEEN −0.25 [−0.88, 0.39]
FSCD −0.26 [−0.86, 0.34]
FTCB −0.25 [−0.64, 0.14]
GFK −0.29 [−0.87, 0.29]
GMRF 0.76 [−0.31, 1.82]
GRAC 0.21 [−0.85, 1.27]
GSDC 0.13 [−0.28, 0.55]
GTPC −0.18 [−1.14, 0.78]
GUTS −0.38 [−0.97, 0.21]
MRSC 0.01 [−0.26, 0.28]
MRSD 0.18 [−0.67, 1.03]
NCMH −0.24 [−0.64, 0.16]
NHS2 0.12 [−0.22, 0.46]
NHSY 0.05 [−0.12, 0.22]
NIUT −0.27 [−1.43, 0.90]
NSS1 −0.12 [−0.31, 0.07]
NSS2 −0.03 [−0.33, 0.28]
ONGA 0.35 [−0.48, 1.18]
ONGB 0.24 [−0.39, 0.88]
PPDS 0.12 [−0.07, 0.31]
PRIS −0.05 [−0.49, 0.39]
PSY5 0.11 [−0.49, 0.70]
PTS1 −0.26 [−0.87, 0.36]
QIM2 0.06 [−0.45, 0.58]
RING 0.23 [−0.73, 1.20]
SEEP −0.11 [−0.57, 0.34]
STRO 0.02 [−0.37, 0.41]
UKBB −0.06 [−0.10, −0.03]
VAKN −0.17 [−0.47, 0.14]
WACH 0.14 [−1.25, 1.53]
WRBY −0.31 [−0.93, 0.31]
WTCS −0.17 [−0.51, 0.16]
F2.5 Meta −0.05 [−0.08, −0.02]
EHR
DAI2 −0.13 [−0.18, −0.08]
ESBB −0.04 [−0.08, 0.01]
FING −0.10 [−0.17, −0.03]
HUNT 0.06 [−0.04, 0.17]
SWED −0.07 [−0.45, 0.31]
UKB2 −0.04 [−0.11, 0.03]
EHR Meta −0.07 [−0.09, −0.04]
Freeze 3 Meta −0.06 [−0.08, −0.04]
−3
−2
−1
0
1
2
Observed Outcome

rs10284205 (Locus 78)

| Study | Estimate [95% CI] |
|---|---|
| **Freeze 2.5** | |
| AGDS | 0.18 [ 0.09, 0.27] |
| AURO | 0.03 [−0.60, 0.65] |
| BETR | −0.29 [−1.22, 0.64] |
| BRY2 | −0.02 [−0.89, 0.85] |
| BRYA | 0.25 [−0.29, 0.78] |
| COGA | −0.10 [−0.40, 0.19] |
| COGB | −0.10 [−0.43, 0.22] |
| COM1 | 0.17 [−0.55, 0.89] |
| EACR | −0.05 [−0.59, 0.48] |
| FEEN | −0.57 [−1.02, −0.13] |
| FSCD | −0.12 [−0.53, 0.28] |
| FTCB | 0.04 [−0.25, 0.33] |
| GFK | 0.26 [−0.15, 0.66] |
| GMRF | 0.10 [−0.56, 0.77] |
| GRAC | 0.47 [−0.26, 1.19] |
| GSDC | −0.16 [−0.48, 0.15] |
| GTPC | 0.19 [−0.49, 0.88] |
| GUTS | 0.07 [−0.34, 0.49] |
| MINV | 0.30 [−0.33, 0.94] |
| MRSC | 0.01 [−0.18, 0.21] |
| MRSD | 0.45 [−0.23, 1.13] |
| NCMH | 0.03 [−0.24, 0.31] |
| NHS2 | 0.24 [−0.02, 0.49] |
| NHSY | 0.13 [−0.00, 0.26] |
| NIUT | −0.02 [−1.01, 0.97] |
| NSS1 | −0.12 [−0.25, 0.02] |
| NSS2 | −0.04 [−0.25, 0.18] |
| ONGA | 0.05 [−0.59, 0.69] |
| ONGB | 0.31 [−0.15, 0.77] |
| PPDS | 0.03 [−0.10, 0.17] |
| PRIS | −0.24 [−0.56, 0.08] |
| PSY5 | 0.27 [−0.15, 0.68] |
| PTS1 | 0.15 [−0.25, 0.56] |
| QIM2 | 0.12 [−0.23, 0.48] |
| RING | −0.24 [−0.94, 0.47] |
| SEEP | −0.14 [−0.50, 0.23] |
| STRO | 0.16 [−0.12, 0.44] |
| UKBB | 0.04 [ 0.02, 0.07] |
| VAKN | 0.15 [−0.08, 0.38] |
| WACH | 0.24 [−0.65, 1.13] |
| WRBY | −0.16 [−0.62, 0.31] |
| WTCS | −0.02 [−0.27, 0.22] |
| F2.5 Meta | 0.05 [0.02, 0.07] |
| **EHR** | |
| BIOV | 0.05 [−0.01, 0.11] |
| DAI2 | 0.03 [−0.01, 0.06] |
| ESBB | 0.02 [−0.02, 0.05] |
| FING | 0.07 [ 0.02, 0.12] |
| HUNT | −0.01 [−0.09, 0.08] |
| SWED | 0.35 [ 0.07, 0.62] |
| UKB2 | 0.04 [−0.01, 0.09] |
| EHR Meta | 0.03 [0.02, 0.05] |
| Freeze 3 Meta | 0.04 [ 0.03, 0.05] |

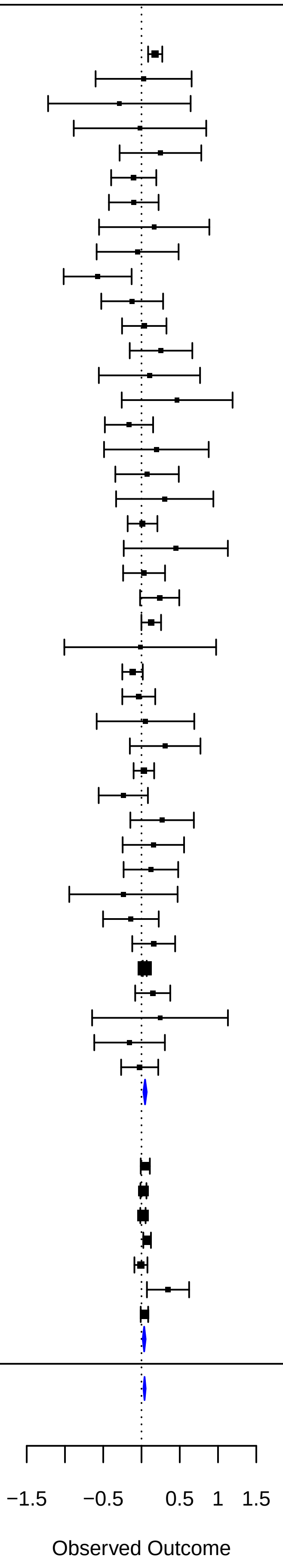

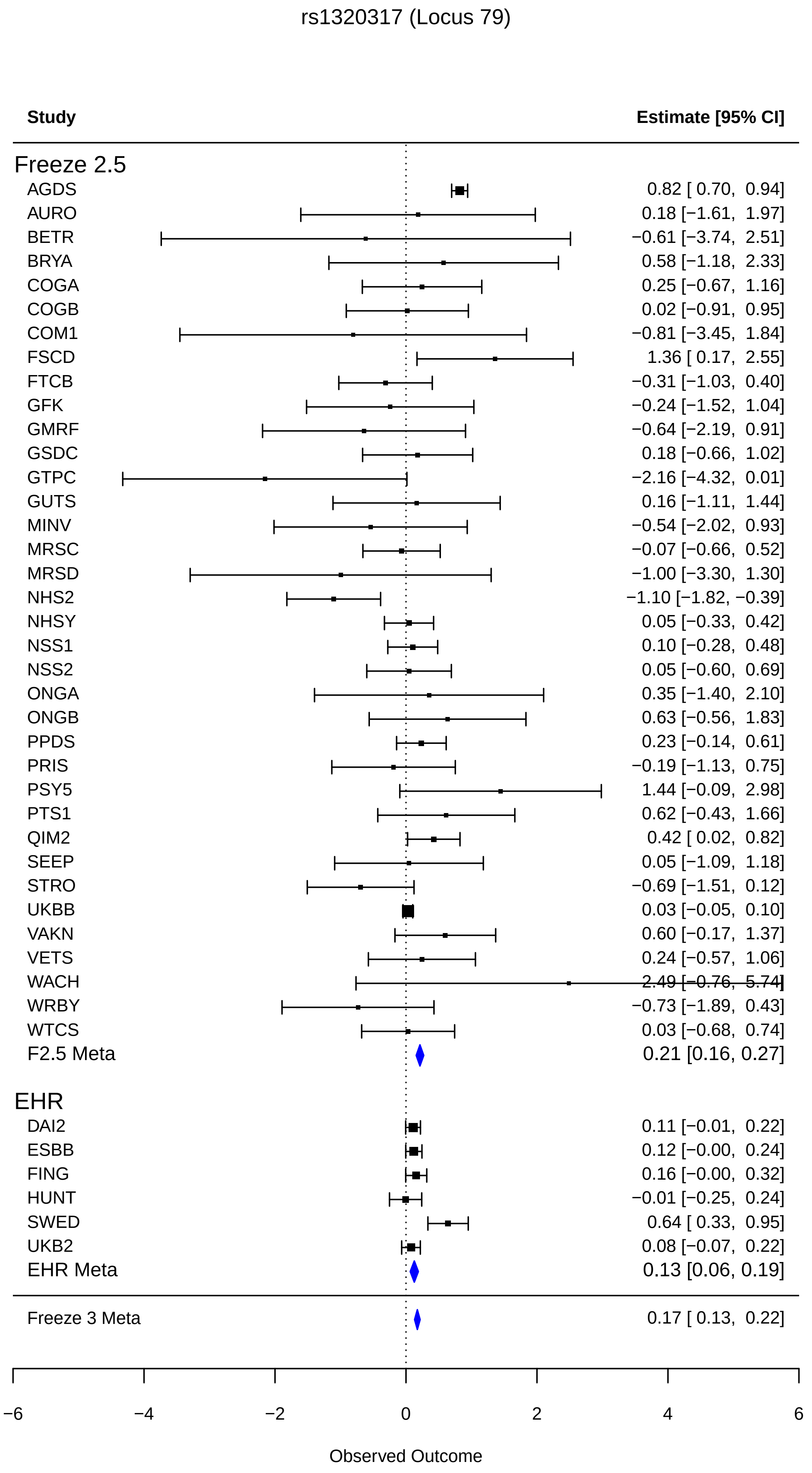
rs1320317 (Locus 79)
Study
Estimate [95% CI]
Freeze 2.5
AGDS 0.82 [ 0.70, 0.94]
AURO 0.18 [−1.61, 1.97]
BETR −0.61 [−3.74, 2.51]
BRYA 0.58 [−1.18, 2.33]
COGA 0.25 [−0.67, 1.16]
COGB 0.02 [−0.91, 0.95]
COM1 −0.81 [−3.45, 1.84]
FSCD 1.36 [ 0.17, 2.55]
FTCB −0.31 [−1.03, 0.40]
GFK −0.24 [−1.52, 1.04]
GMRF −0.64 [−2.19, 0.91]
GSDC 0.18 [−0.66, 1.02]
GTPC −2.16 [−4.32, 0.01]
GUTS 0.16 [−1.11, 1.44]
MINV −0.54 [−2.02, 0.93]
MRSC −0.07 [−0.66, 0.52]
MRSD −1.00 [−3.30, 1.30]
NHS2 −1.10 [−1.82, −0.39]
NHSY 0.05 [−0.33, 0.42]
NSS1 0.10 [−0.28, 0.48]
NSS2 0.05 [−0.60, 0.69]
ONGA 0.35 [−1.40, 2.10]
ONGB 0.63 [−0.56, 1.83]
PPDS 0.23 [−0.14, 0.61]
PRIS −0.19 [−1.13, 0.75]
PSY5 1.44 [−0.09, 2.98]
PTS1 0.62 [−0.43, 1.66]
QIM2 0.42 [ 0.02, 0.82]
SEEP 0.05 [−1.09, 1.18]
STRO −0.69 [−1.51, 0.12]
UKBB 0.03 [−0.05, 0.10]
VAKN 0.60 [−0.17, 1.37]
VETS 0.24 [−0.57, 1.06]
WACH 2.49 [−0.76, 5.74]
WRBY −0.73 [−1.89, 0.43]
WTCS 0.03 [−0.68, 0.74]
F2.5 Meta 0.21 [0.16, 0.27]
EHR
DAI2 0.11 [−0.01, 0.22]
ESBB 0.12 [−0.00, 0.24]
FING 0.16 [−0.00, 0.32]
HUNT −0.01 [−0.25, 0.24]
SWED 0.64 [ 0.33, 0.95]
UKB2 0.08 [−0.07, 0.22]
EHR Meta 0.13 [0.06, 0.19]
Freeze 3 Meta 0.17 [ 0.13, 0.22]
−6
−4
−2
0
2
4
6
Observed Outcome

rs112052534 (Locus 80)

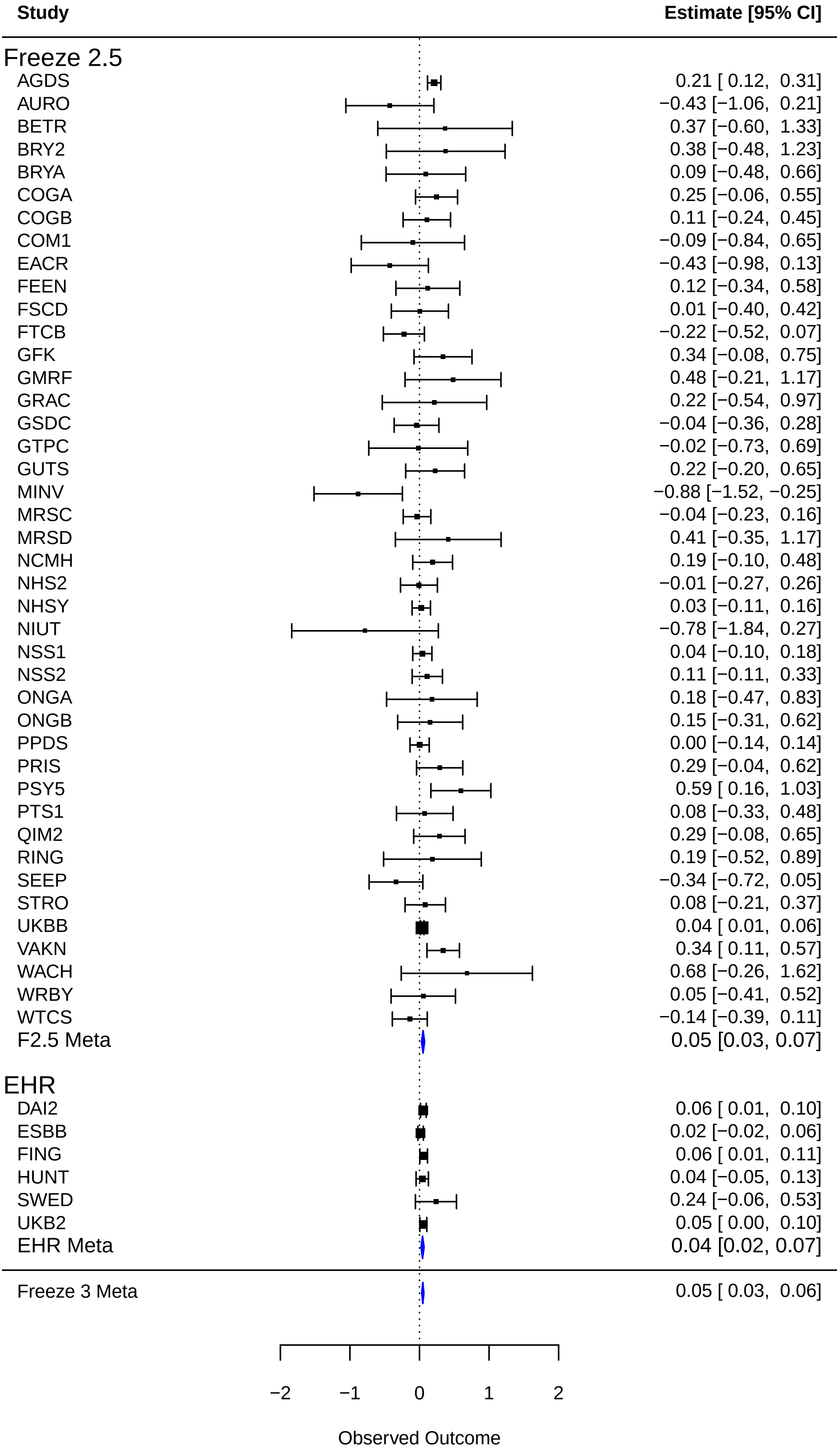

### rs2266850 (Locus 81)

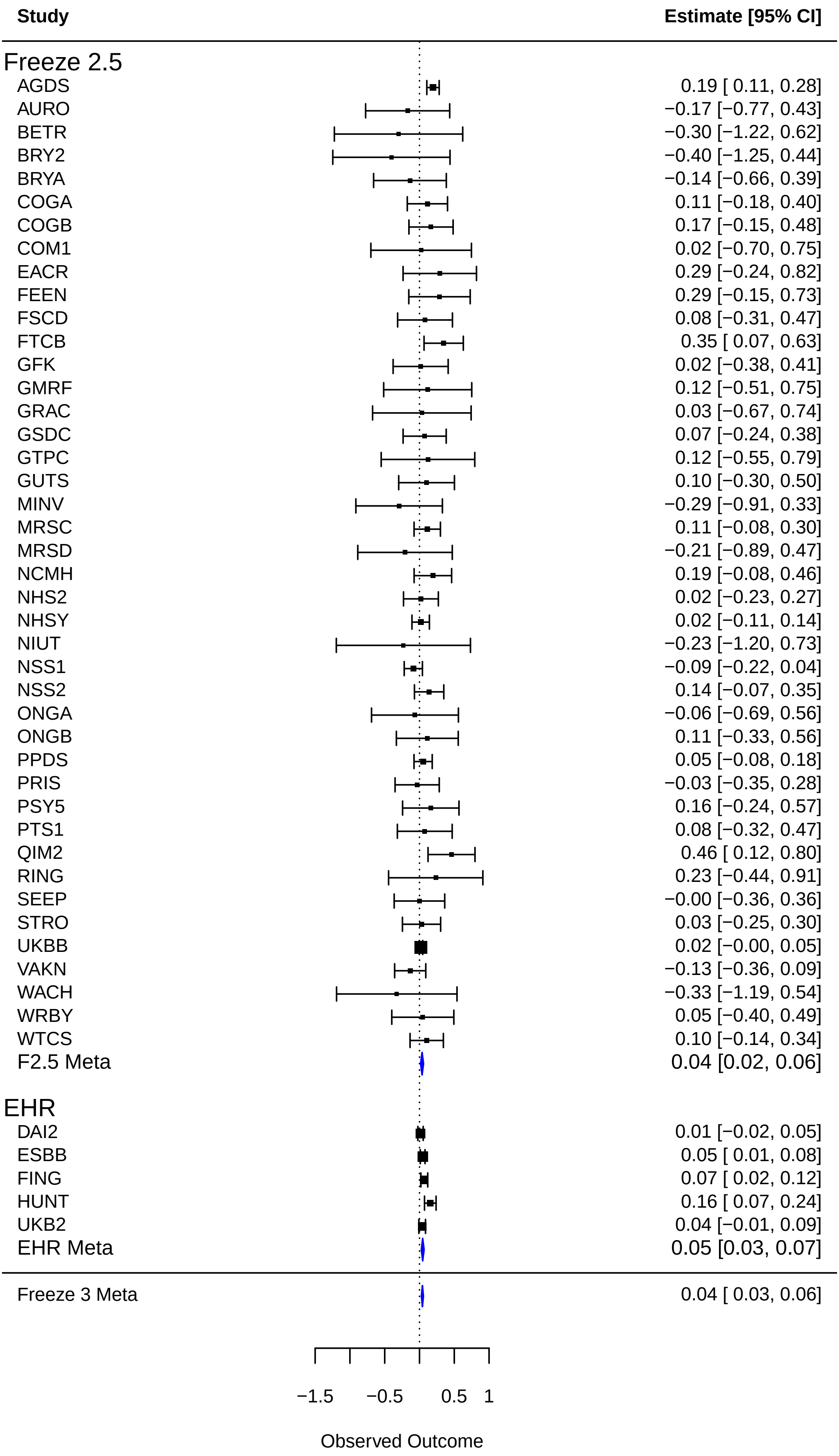
