## Supplemental Data 3 for "Discovery of 95 PTSD loci provides insight into genetic architecture and neurobiology of trauma and stress-related disorders"

#### Supplementary Data 3

Circos Plot of Chromatin Interactions and eQTLs for each chromosome harboring PTSD risk loci based on the EA Meta-Analysis. Outer-most layer: Manhattan plot displaying lead SNPs, where additional independent significant SNPs in loci are colored according to linkage disequilibrium ($r^2$; red [$r^2 > 0.8$], orange [$r^2 > 0.6$], green [$r^2 > 0.4$], blue [$r^2 > 0.2$], grey [$r^2 \leq 0.2$]); -log10 *P*-values for each SNP is indicated on the y-axis. Second layer: chromosome ring with coordinates and genomic risk loci highlighted in blue. Third layer: chromosome ring showing genes mapped by chromatin interactions (orange), eQTL (green), or both (red). Links (orange for chromatin interactions and green for eQTL) are based on all brain tissues available in FUMA v 1.4.1.

#### Chromosome 1

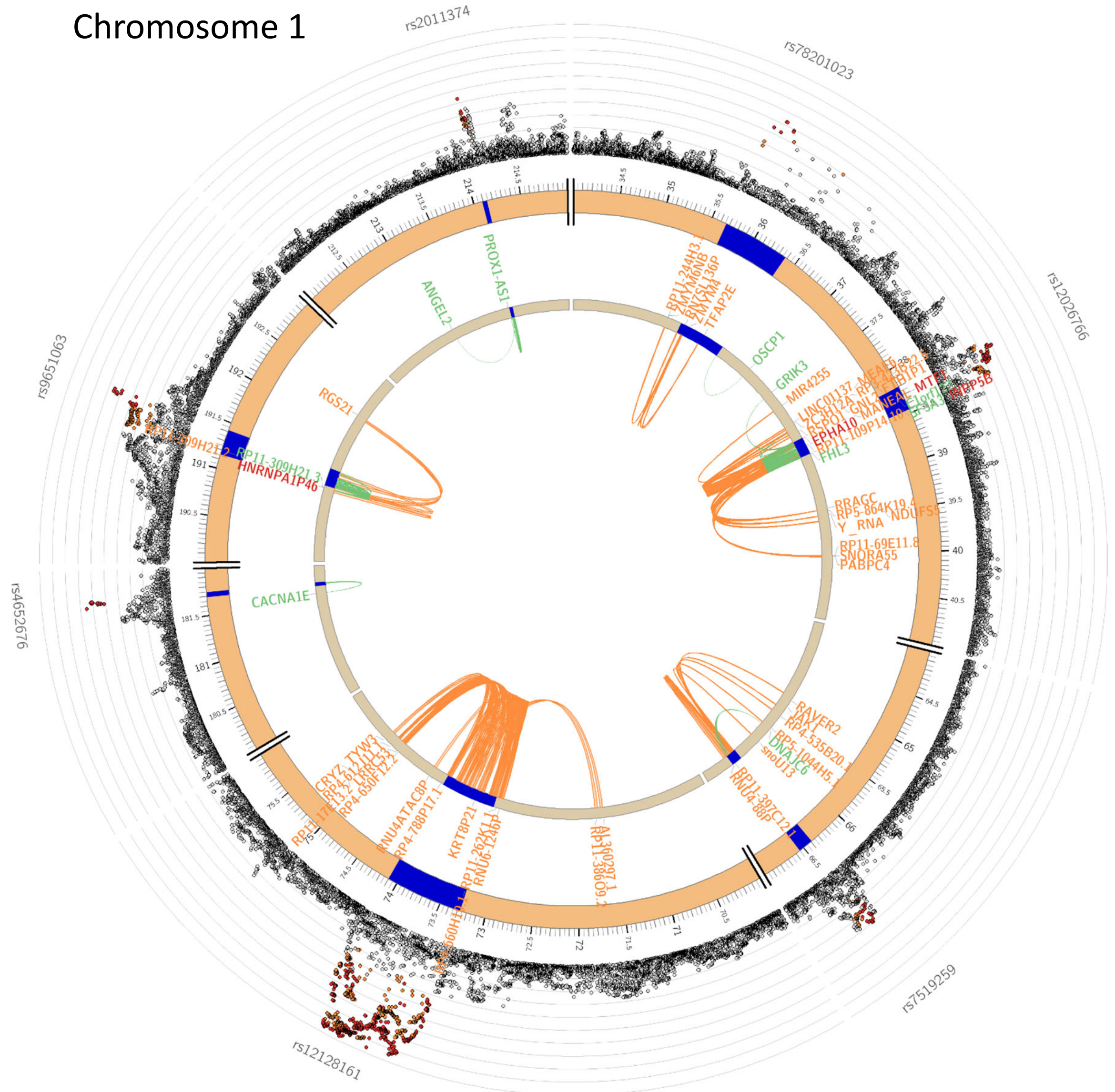

Chromosome 2

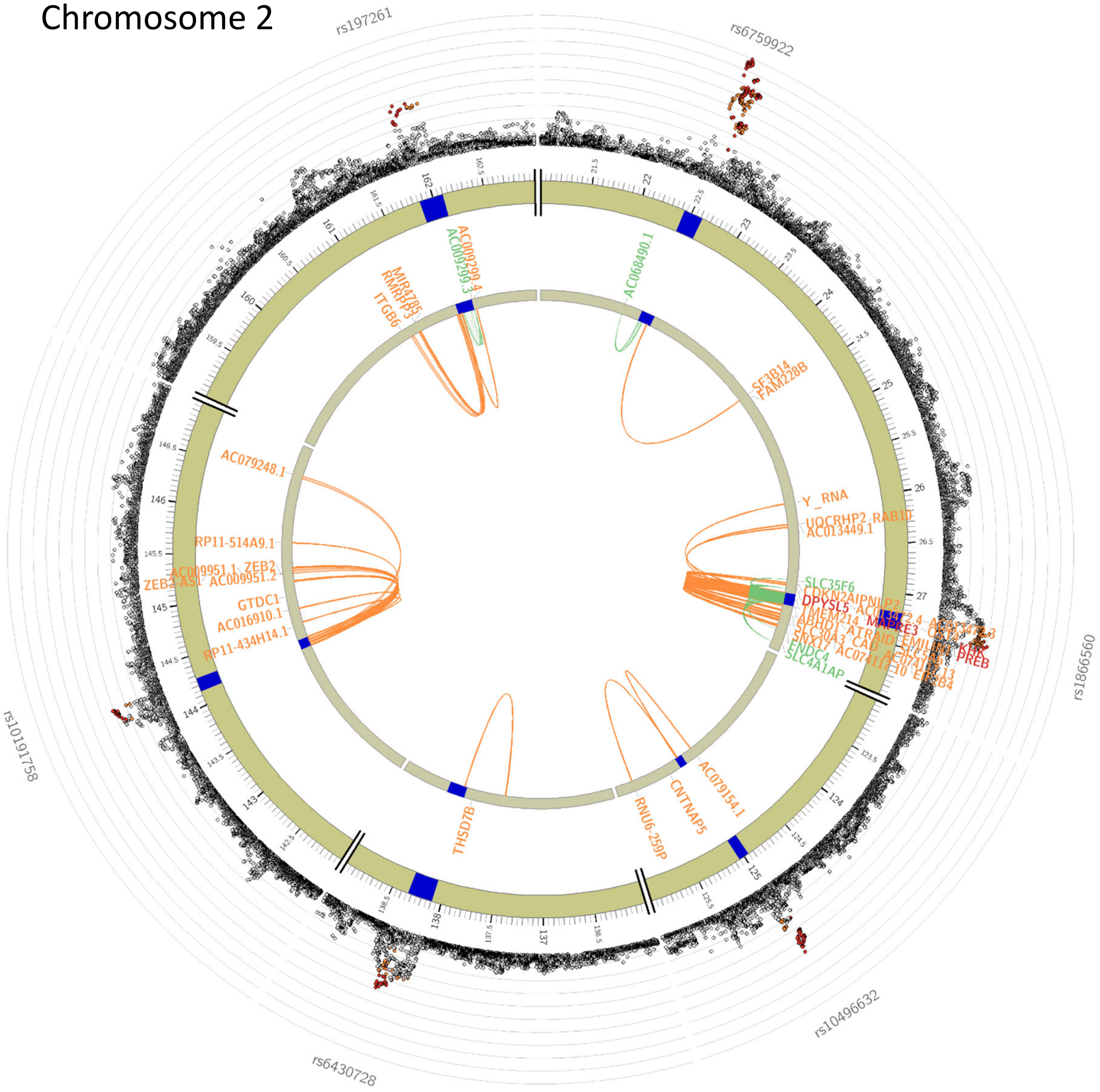

Chromosome 3

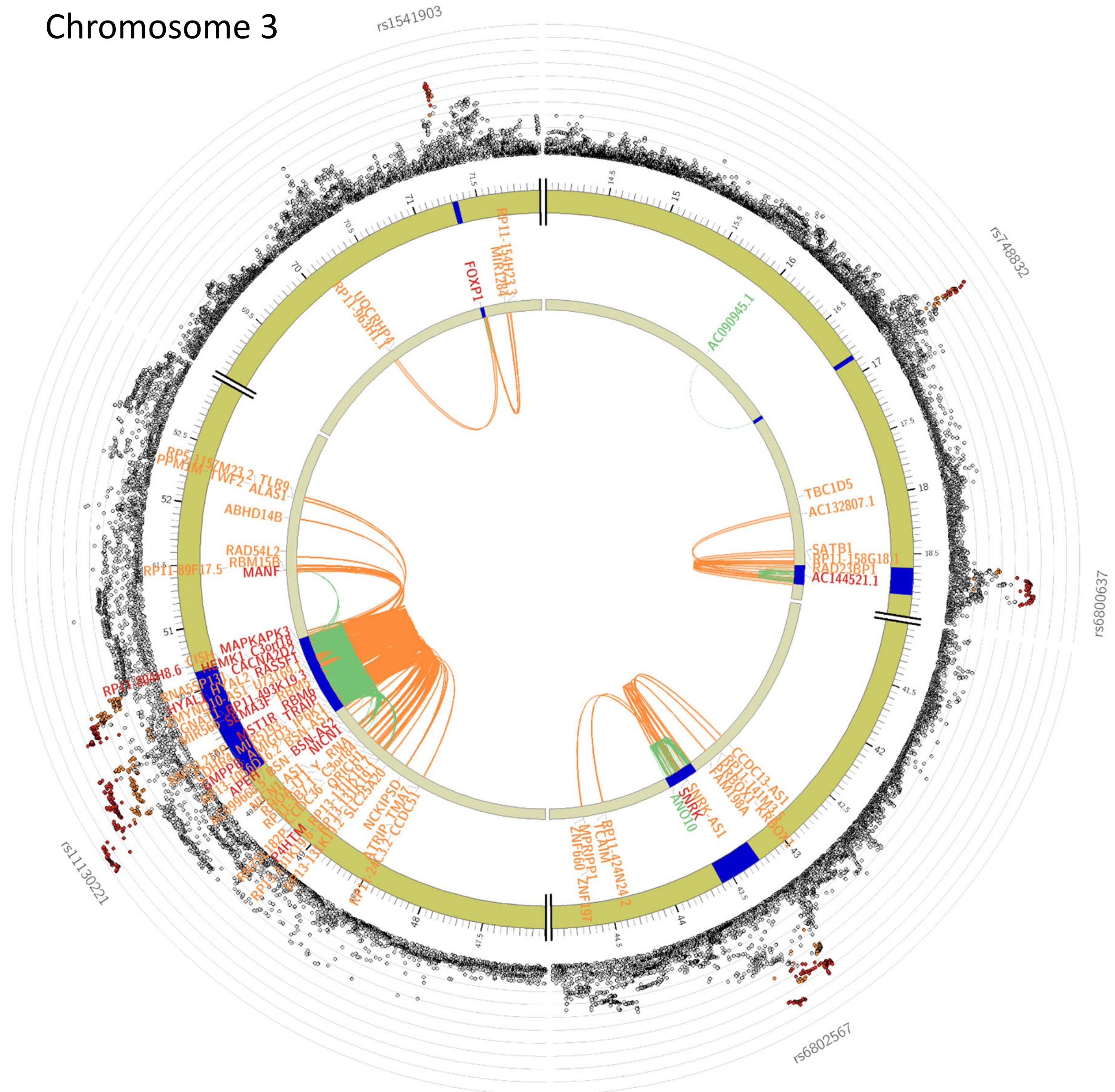

### Chromosome 4

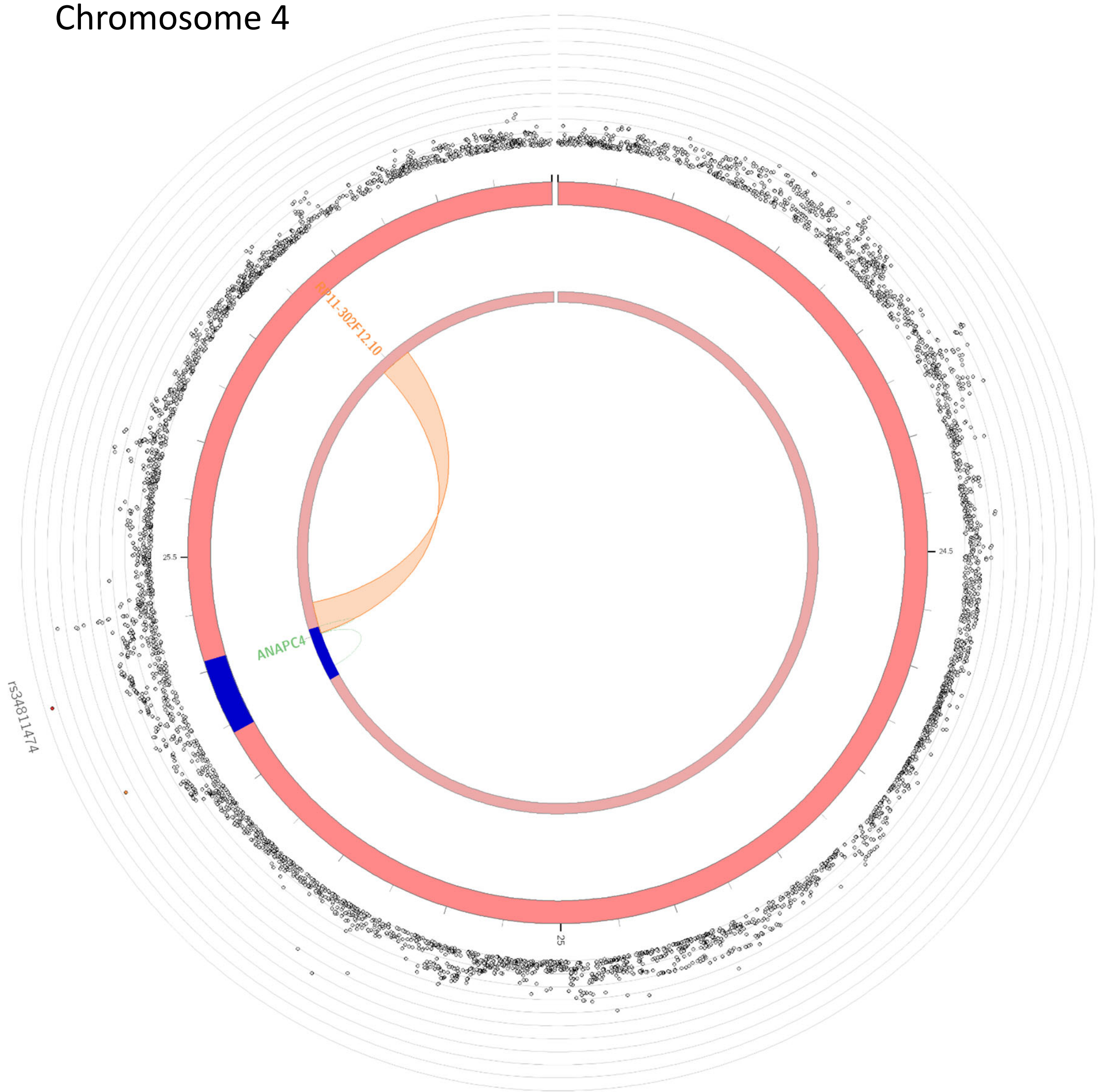

### Chromosome 5

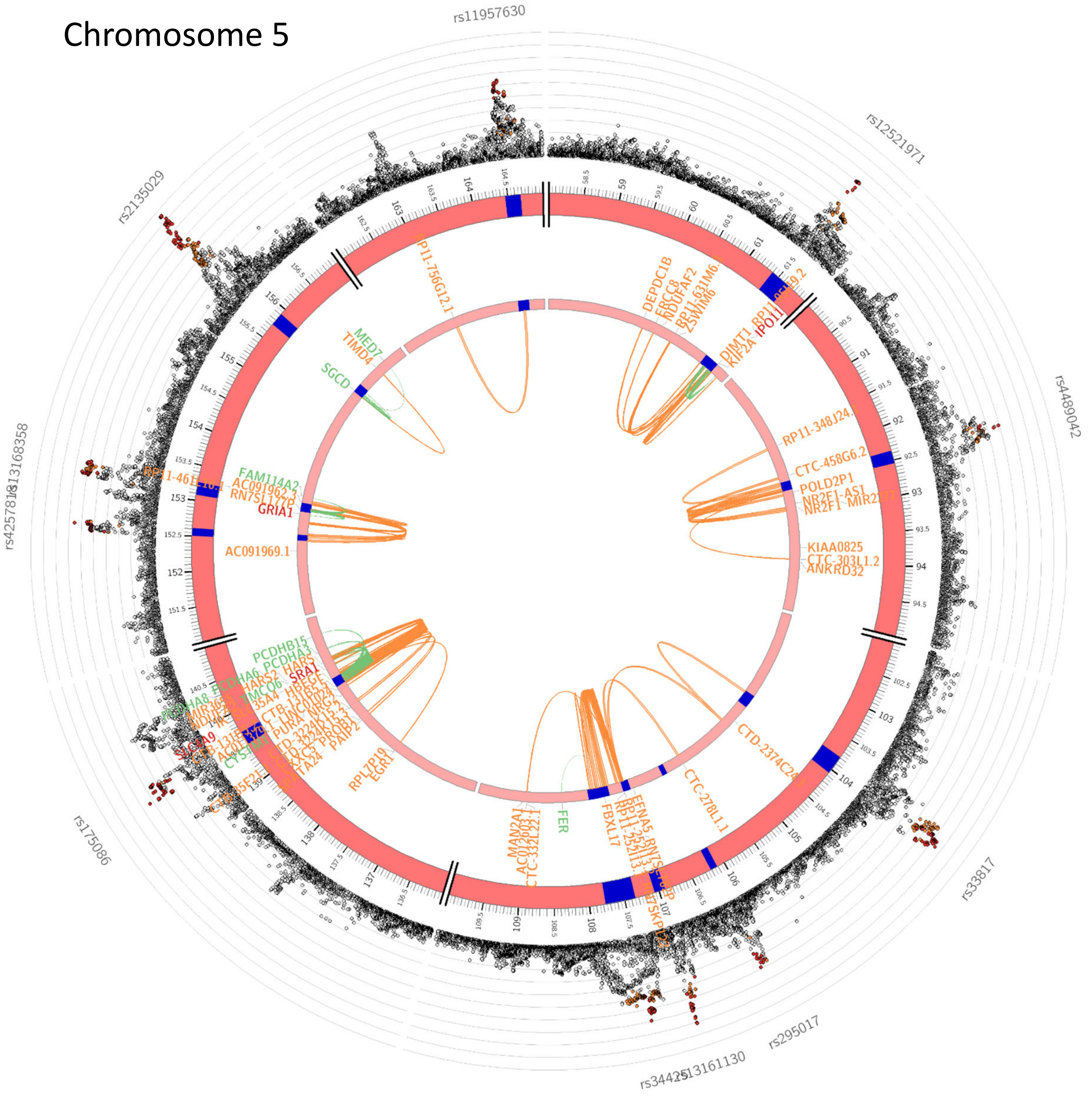

### Chromosome 6

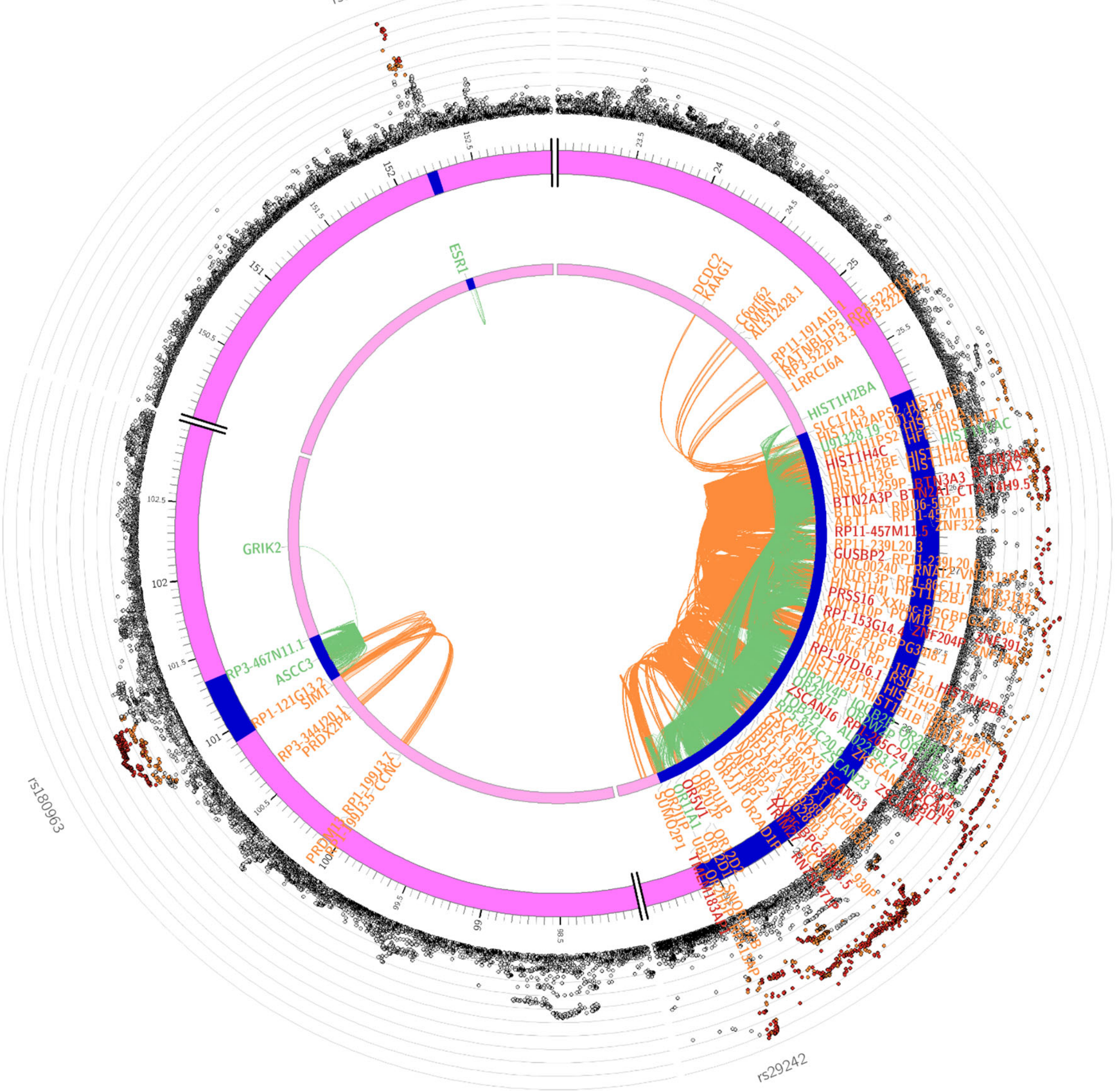

### Chromosome 7

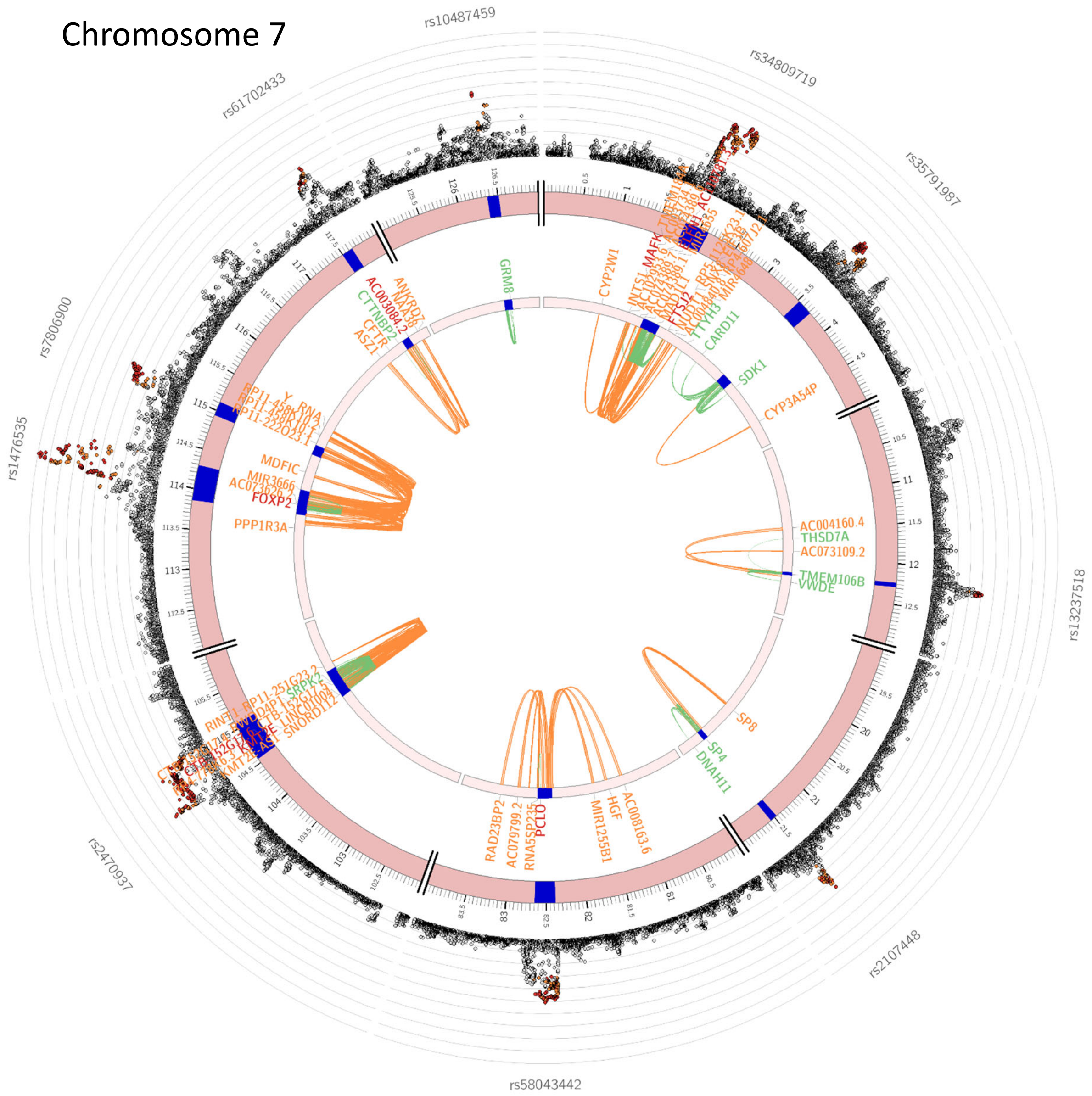

Chromosome 8

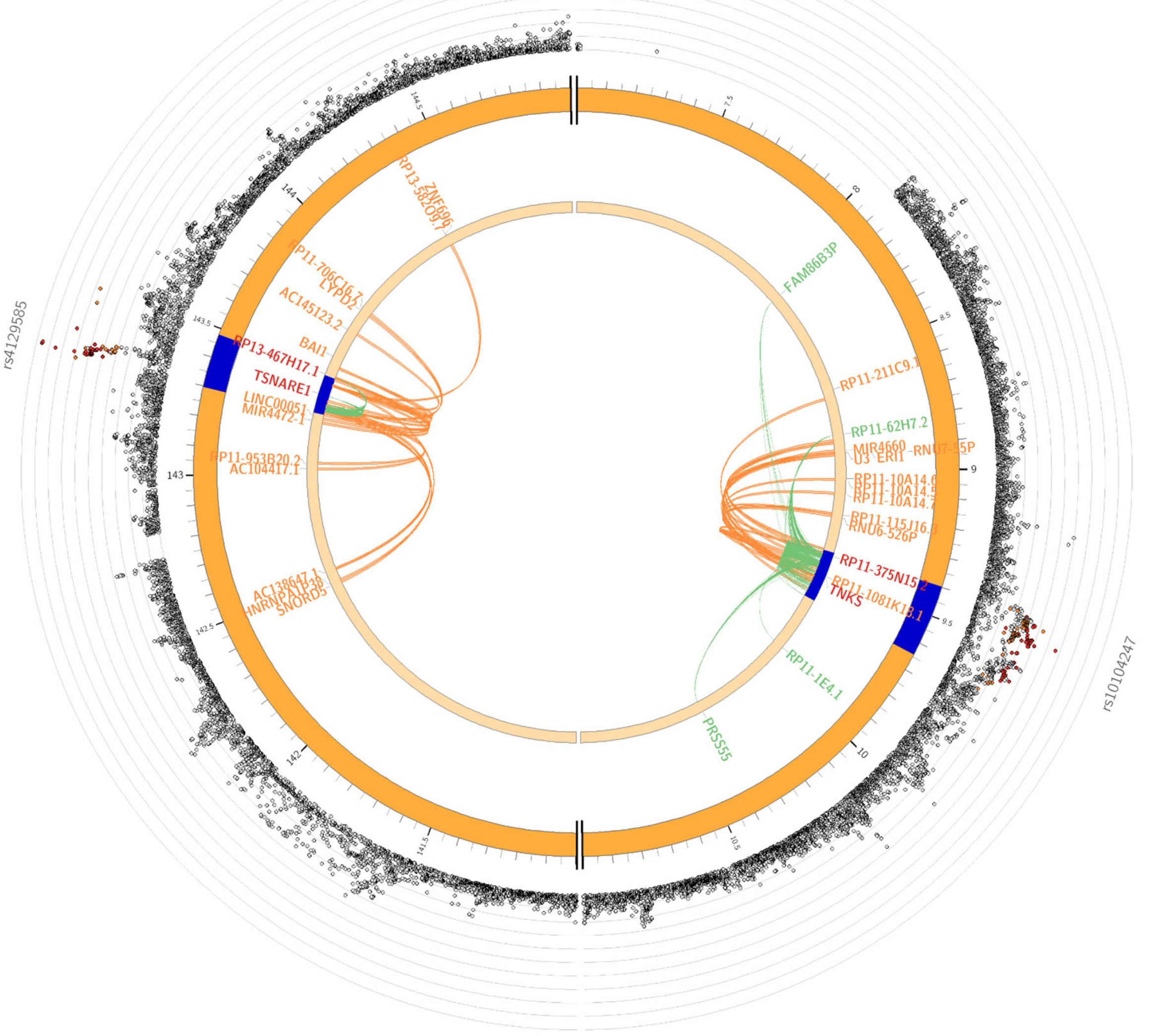

Chromosome 9

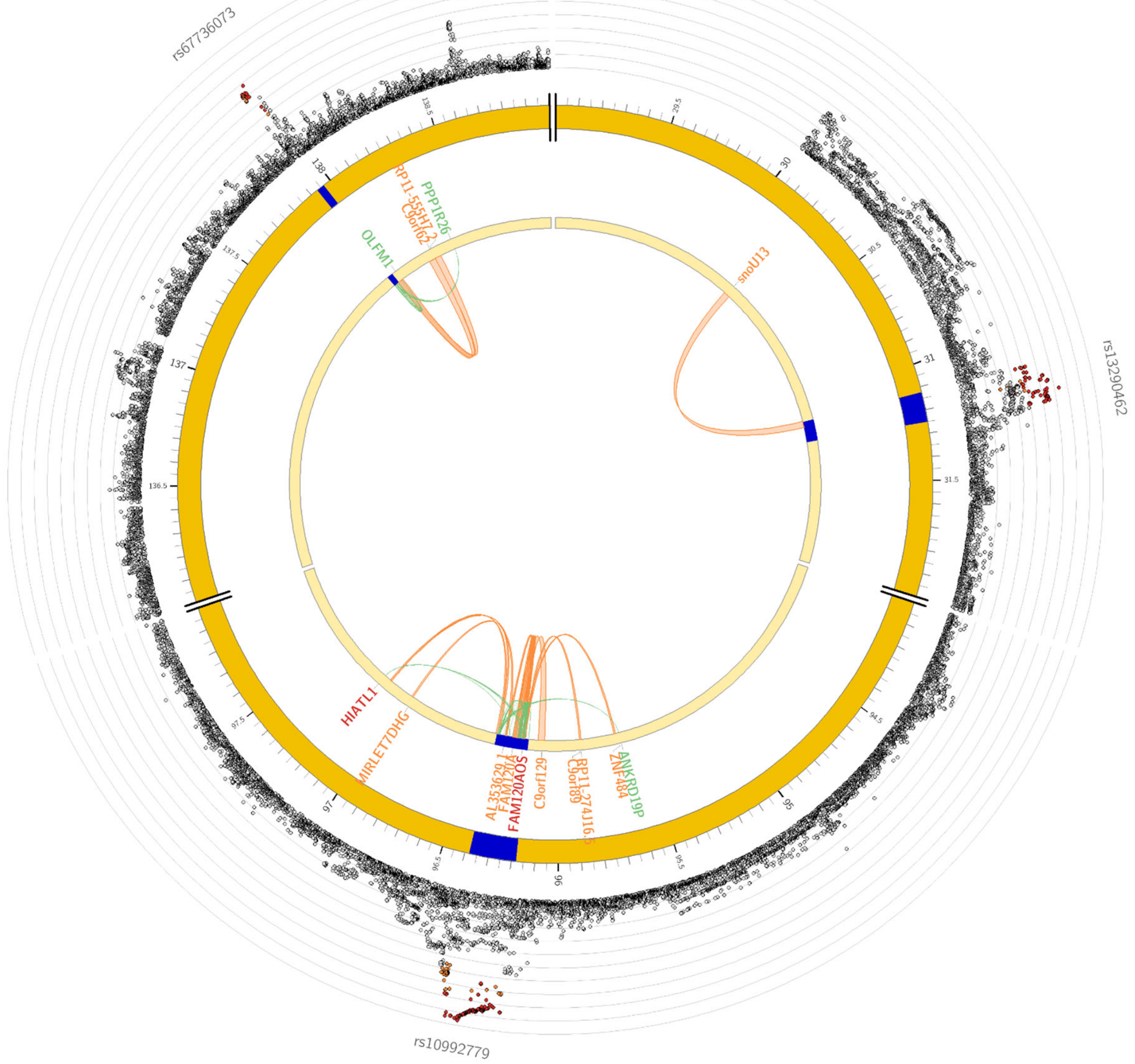

Chromosome 10

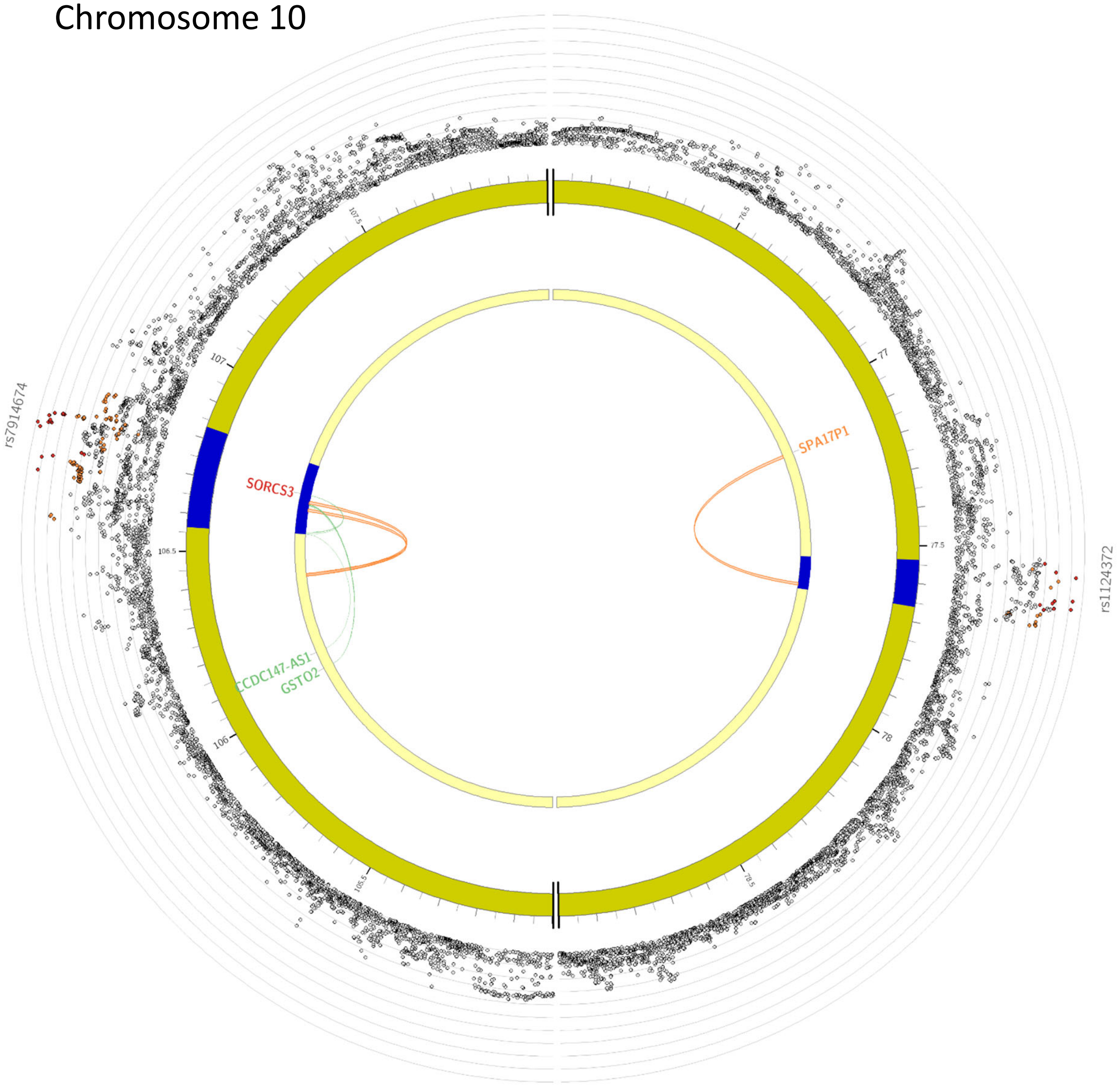

### Chromosome 11

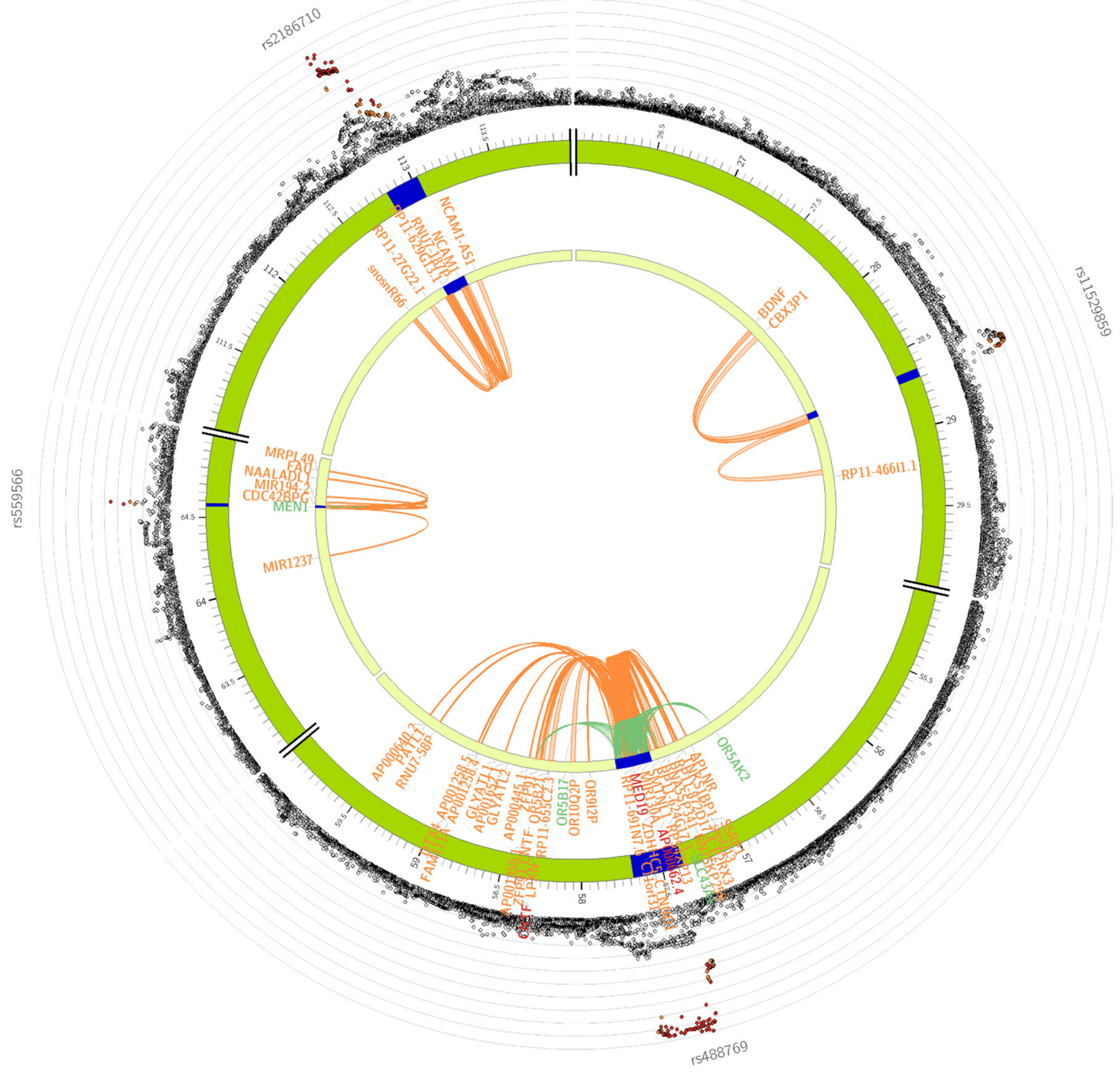

### Chromosome 12

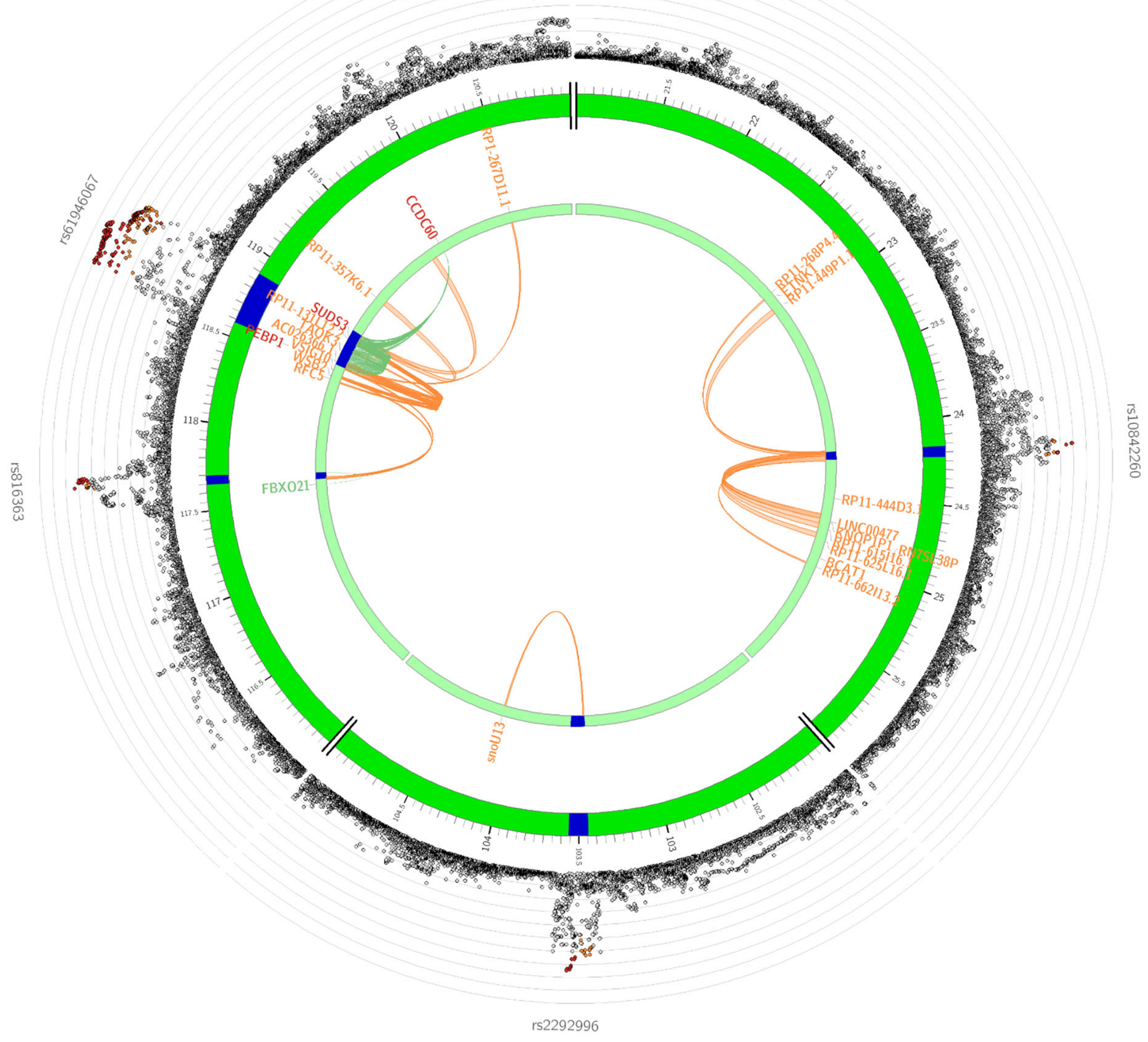

Chromosome 13

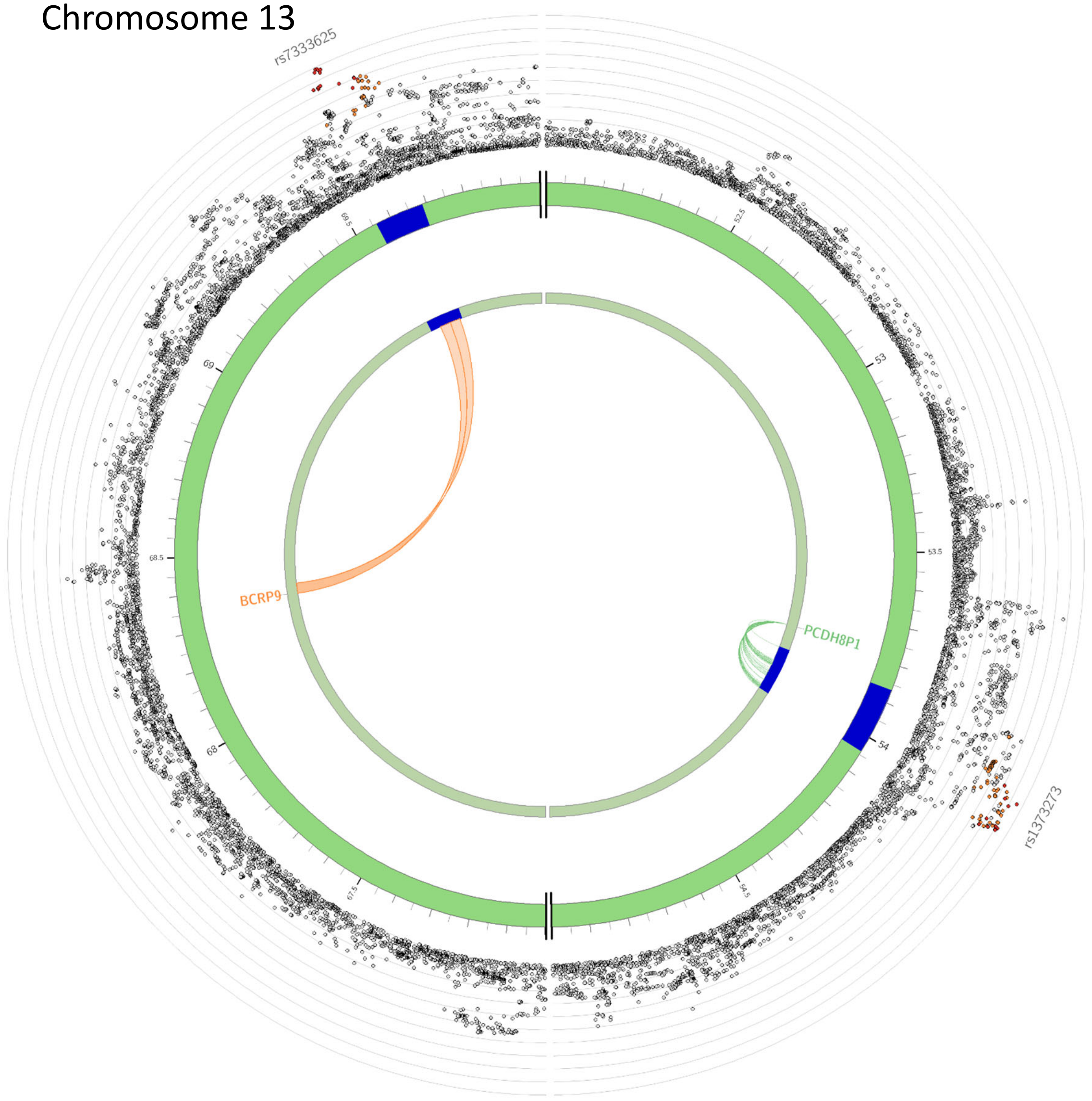

Chromosome 14

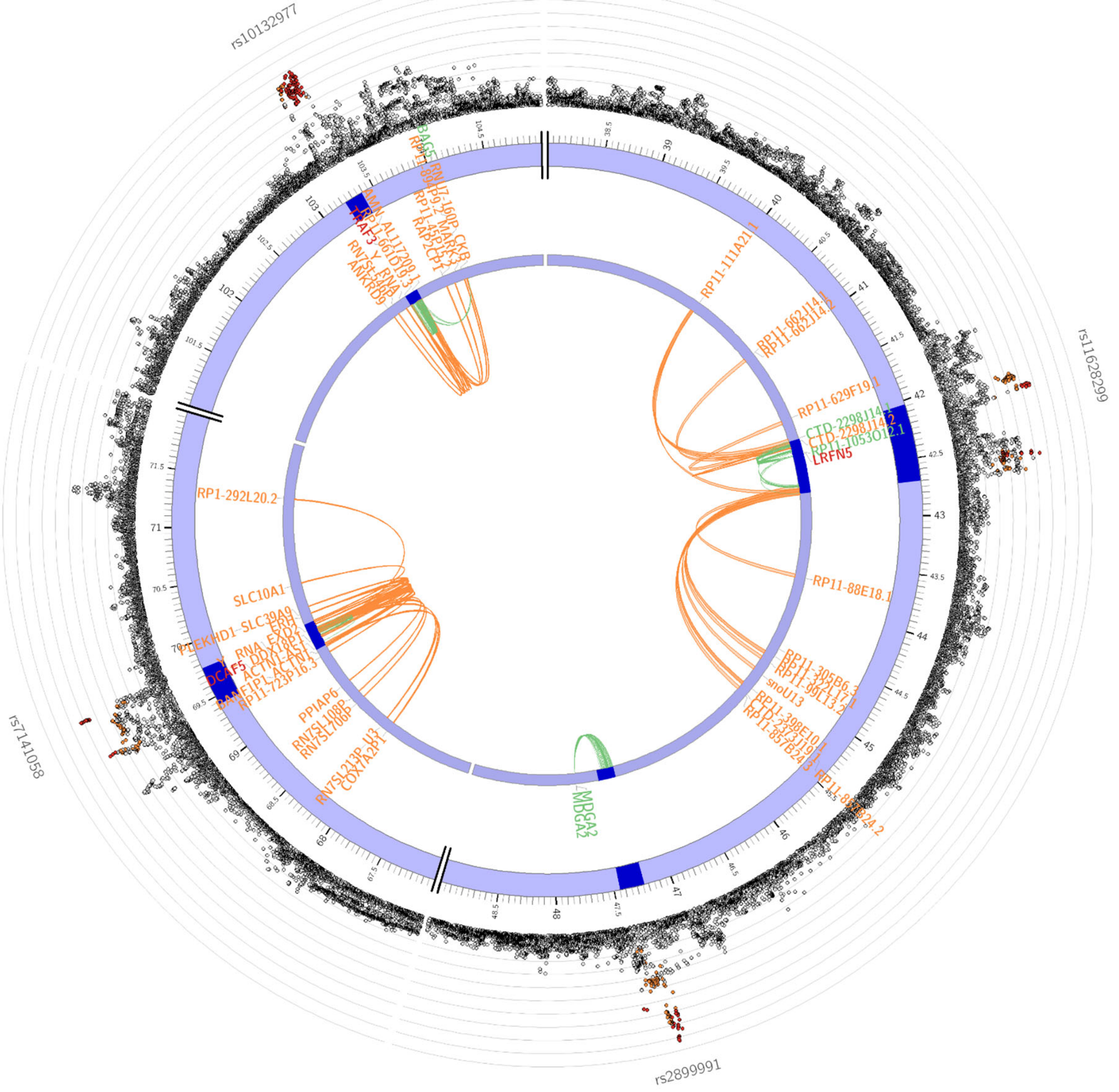

Chromosome 15

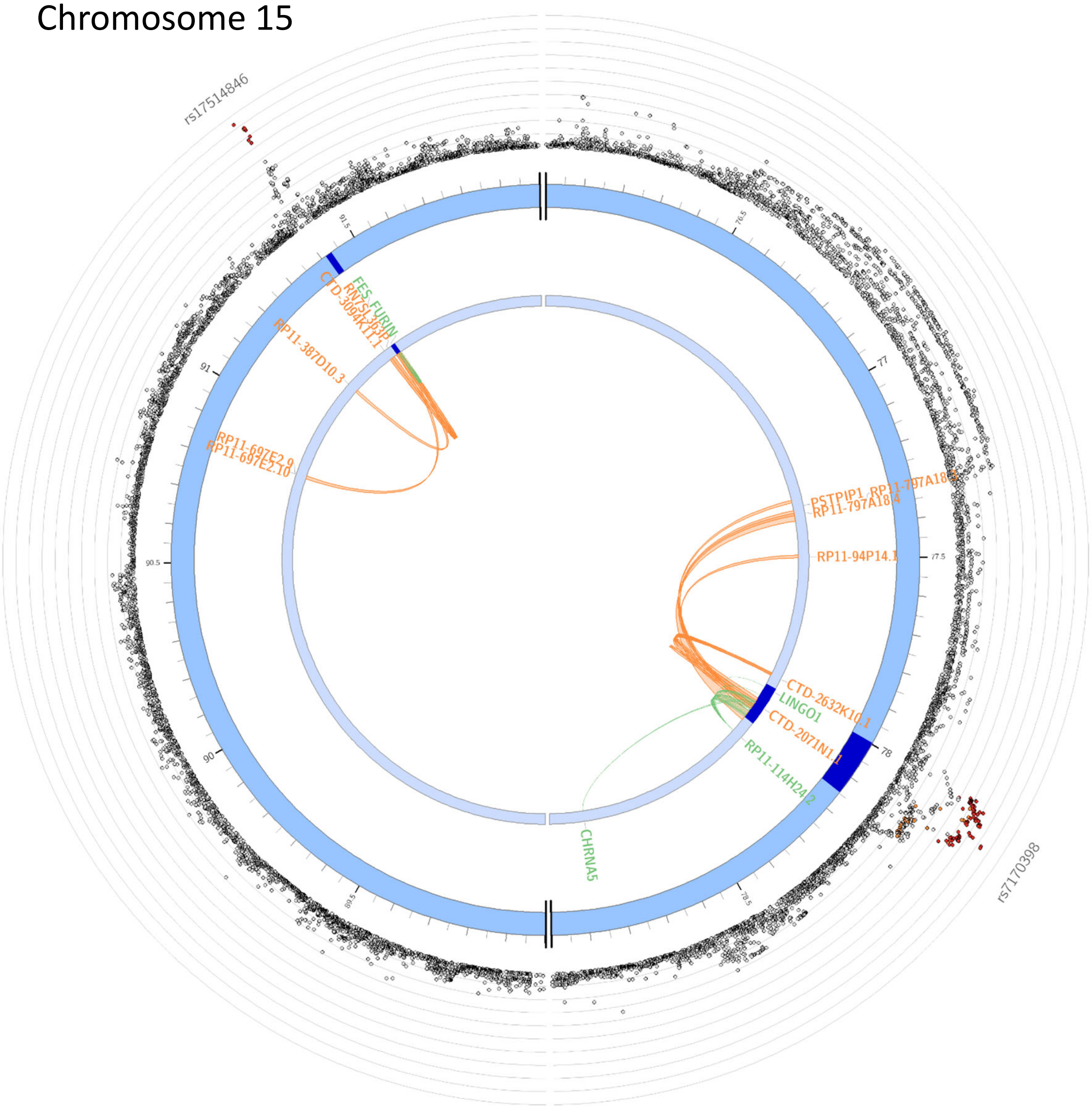

#### Chromosome 16

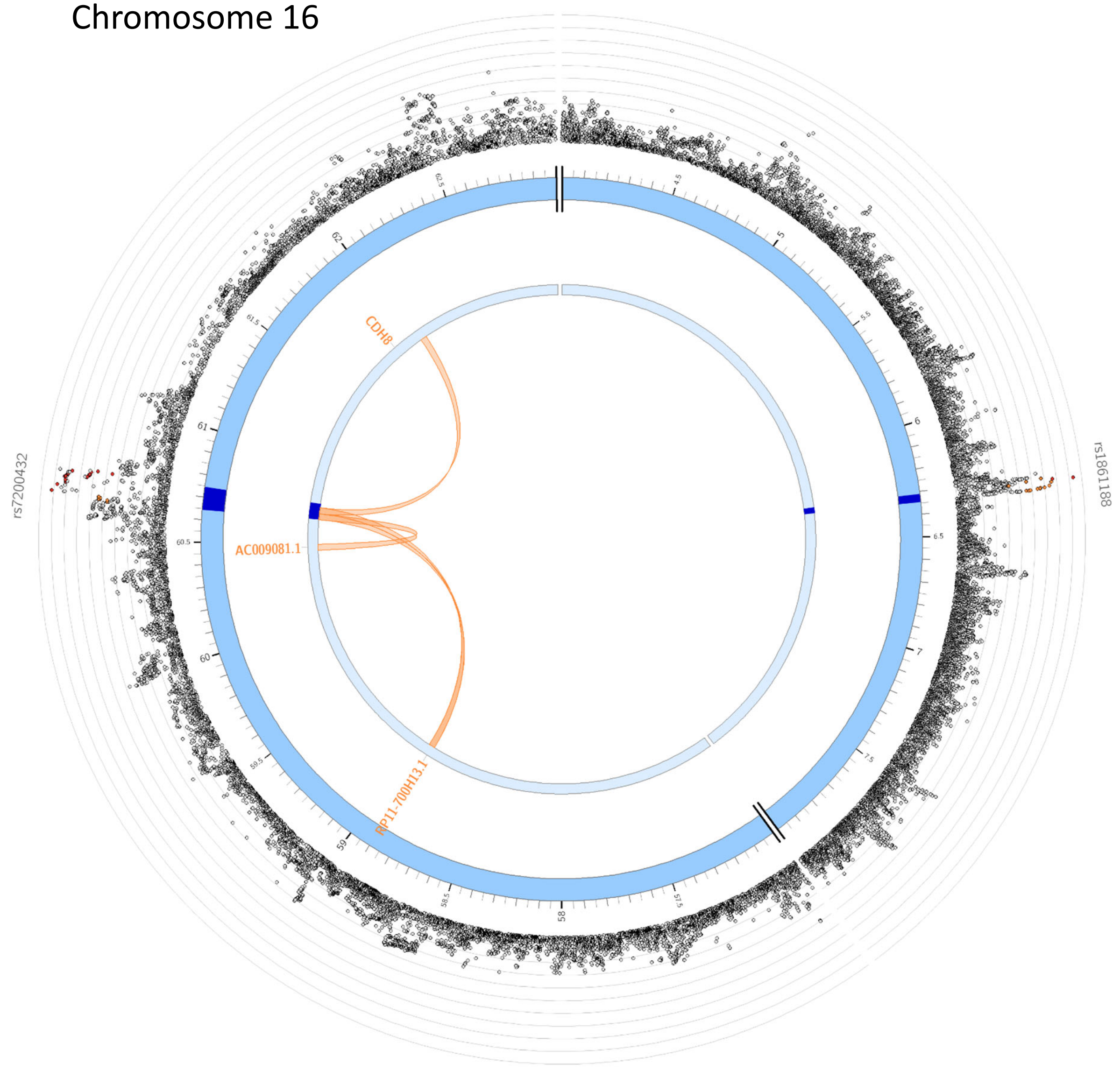

Chromosome 17

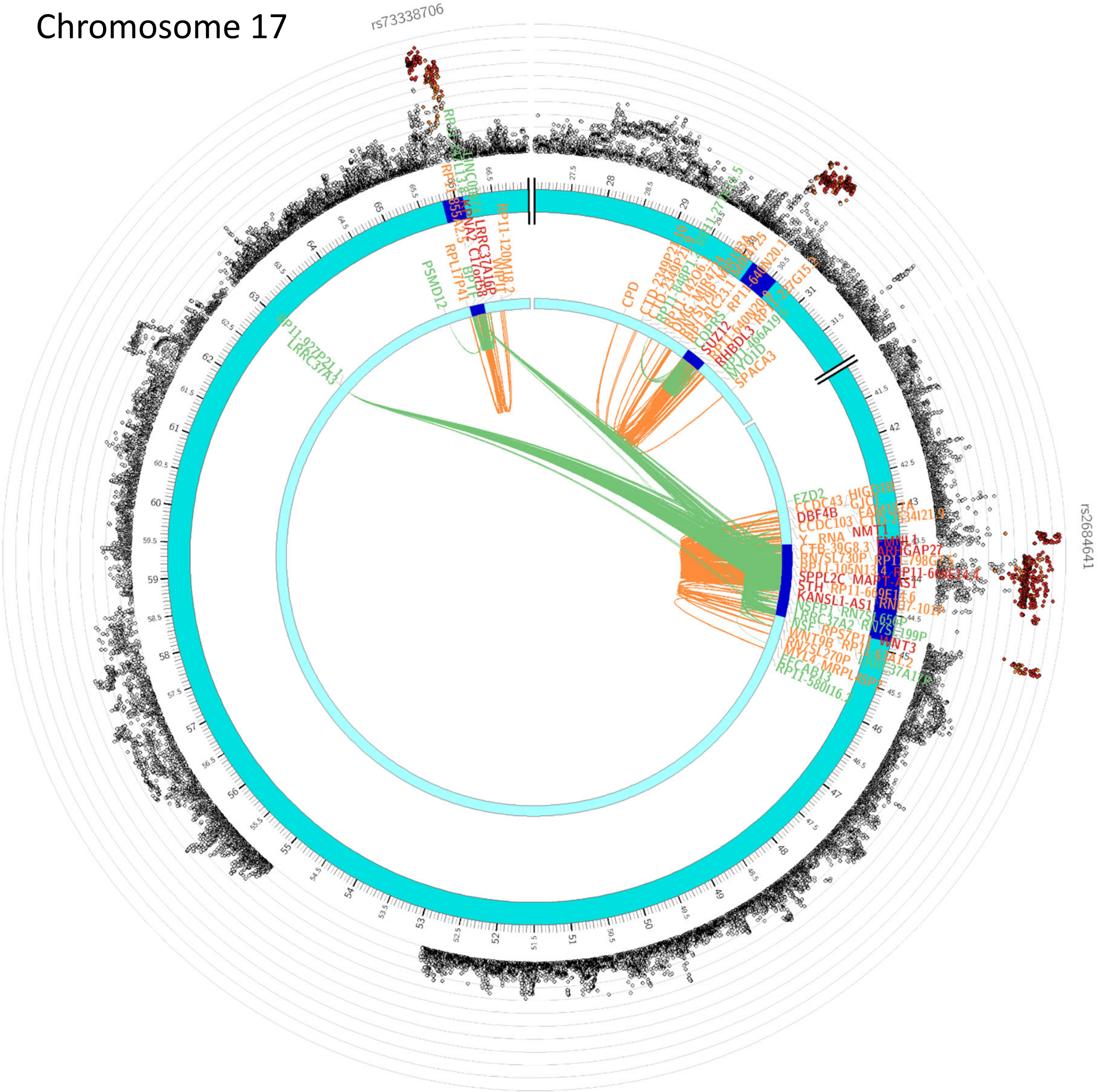

Chromosome 18

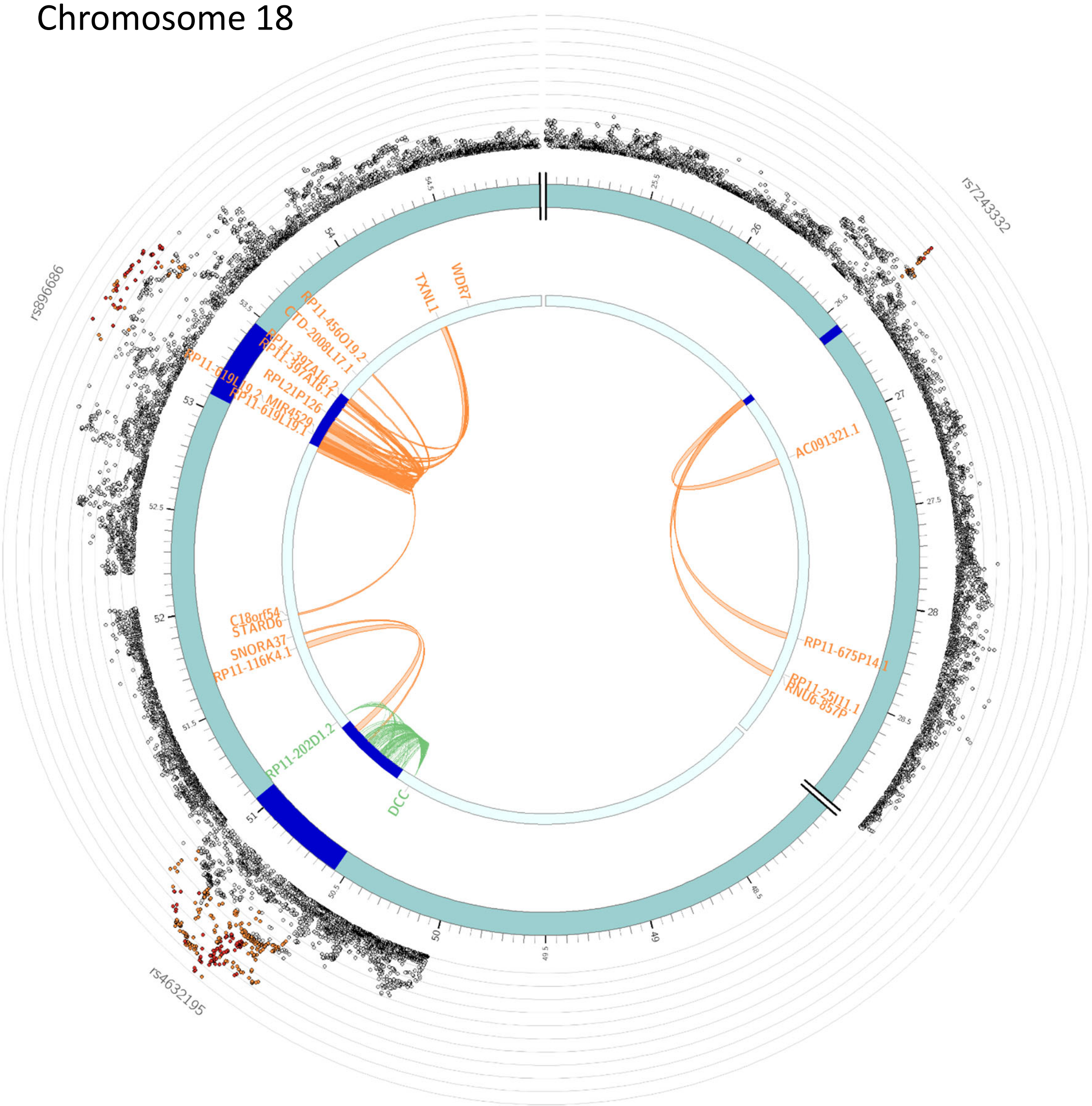

Chromosome 19

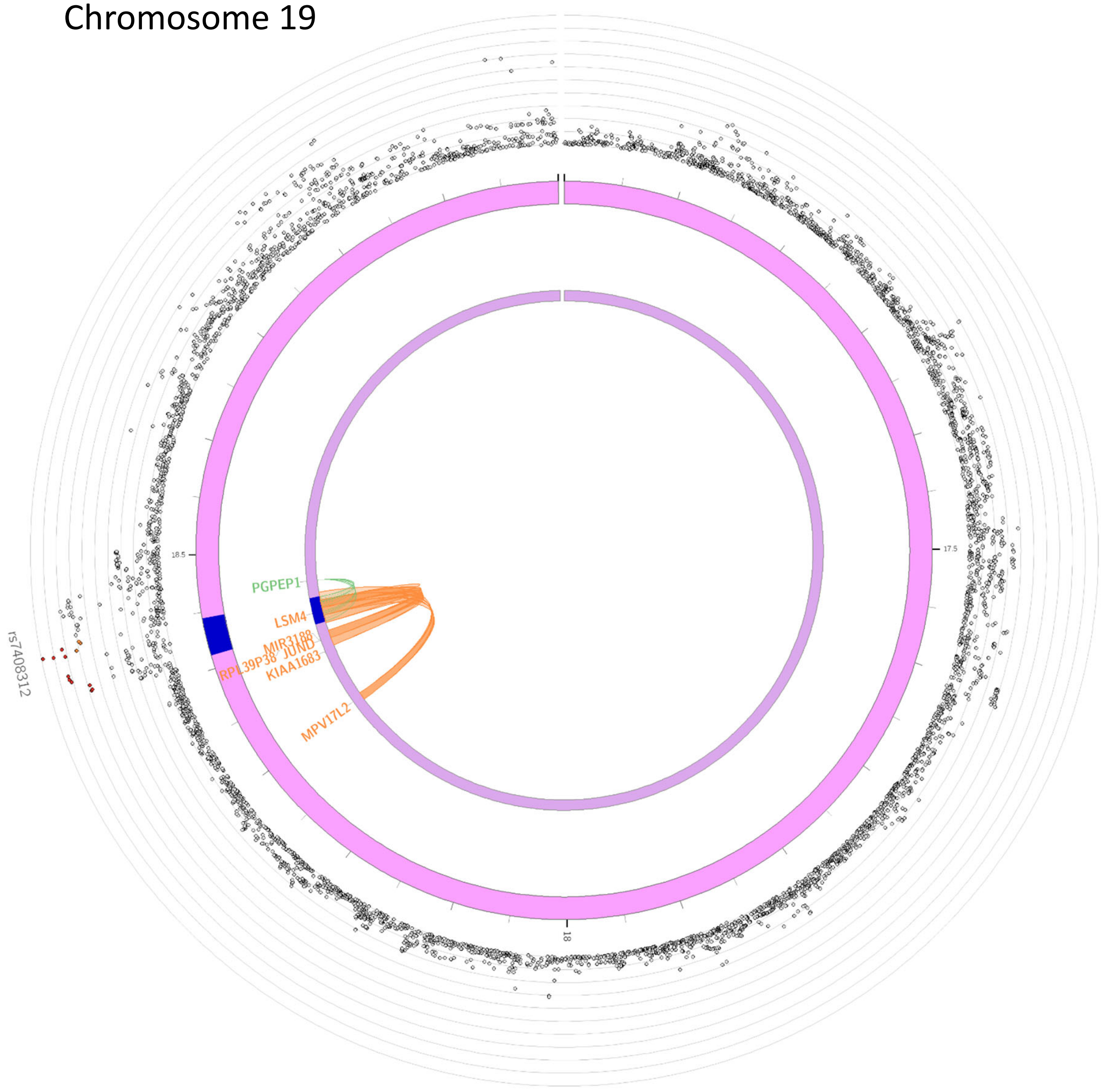

Chromosome 20

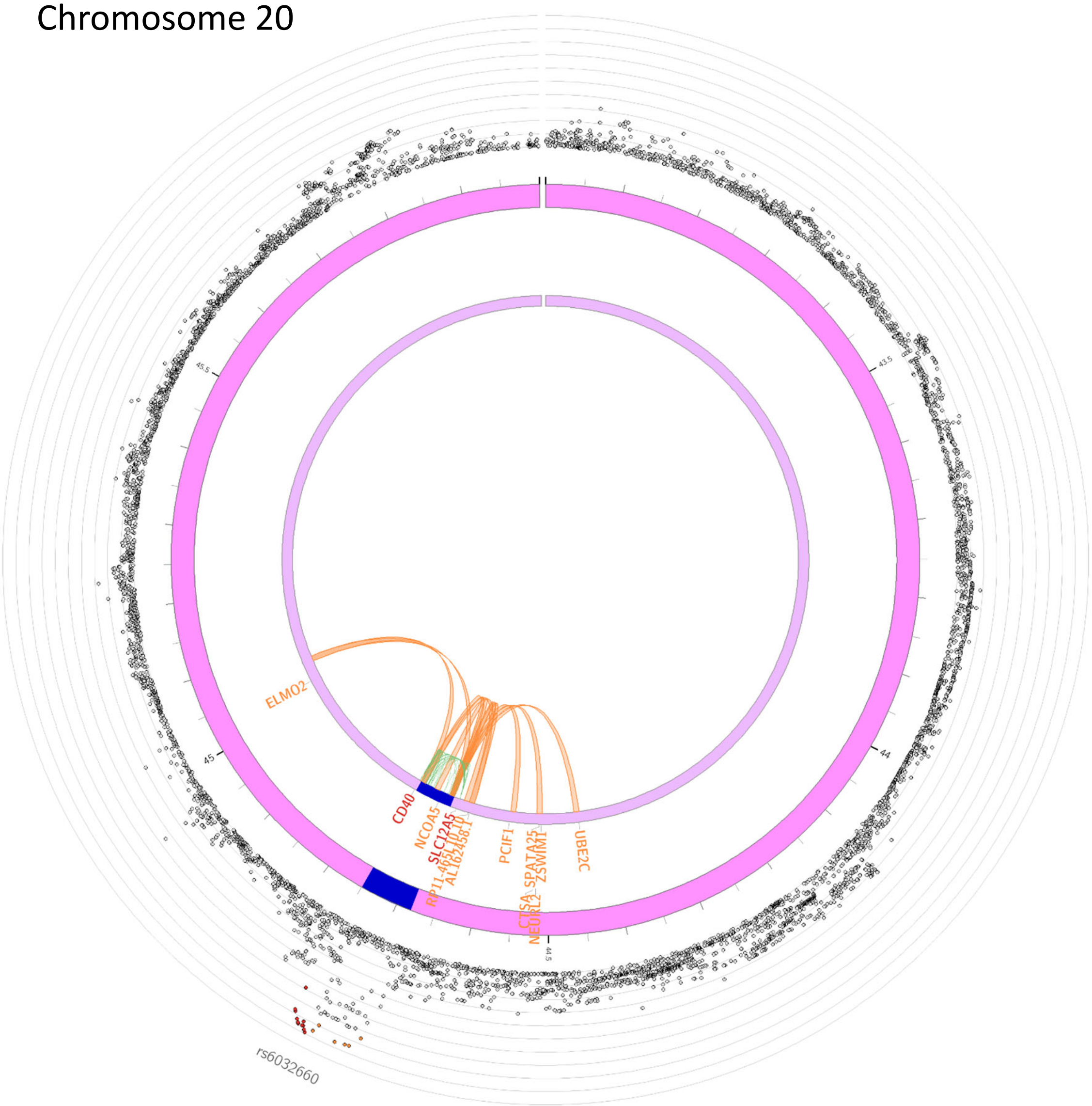

Chromosome X

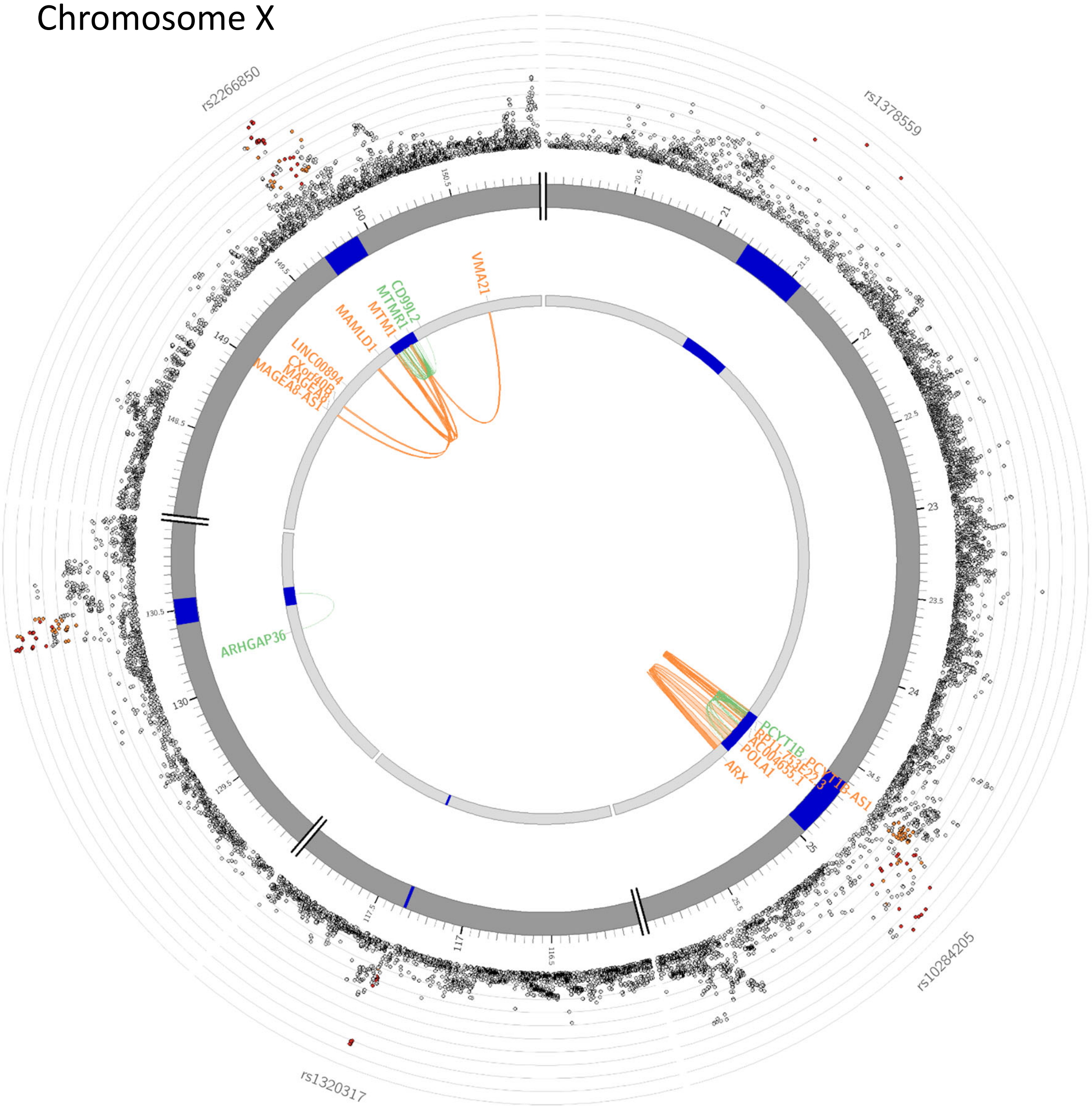
