## Supplemental Methods for "Discovery of 95 PTSD loci provides insight into genetic architecture and neurobiology of trauma and stress-related disorders"

**Supplementary Methods for *Discovery of 95 PTSD loci provides insight into genetic architecture and neurobiology***

**Participating studies**

The following studies were included, listed with the official name of each study followed by a 4-letter abbreviation corresponding to genotyping files and a study number corresponding to **Supplementary Table 1**:

**Army Study to Assess Risk and Resilience in Servicemembers (NSS1, NSS2, PPDS; Supplementary Table 1 #14-16)**

See reference for details.^1^ Potentially traumatic events in childhood and adult civilian trauma were assessed in all participants, as well as military traumatic experience for participants who had been deployed, using a self-administered questionnaire. The extent of traumatic experiences was summarized, separately, in a non-deployment trauma variable and a deployment trauma variable. Both continuous variables are summaries of frequencies of responses to each of the 11 questions regarding non-deployment or deployment trauma. Responses range from Never (0), Once (1), 2-4 Times (2), 5-9 Times (3) or More than 10 Times (4). A combination of a computer-administered version of the Composite International Diagnostic Interview (CIDI) and the Posttraumatic Stress Disorder Checklist (PCL for DSM-IV)^2^ were used to assign diagnoses of lifetime Posttraumatic Stress Disorder (PTSD) according to DSM-IV.^3^ DNA for GWAS analysis was isolated from blood. The Institutional Review Boards of all participating institutions approved this study.

**Ash Wednesday (BRYA; Supplementary Table 1 #10)**

See reference for details.^4^ Potentially traumatic events were identified using the Recent Life Events questionnaire.^5^ The PTSD Checklist (PCL) was used to assess PTSD over the prior 4 weeks by interviews.^2^ Respondents were considered to have a diagnosis if DSM-IV criteria were met. The PCL calculates PTSD symptom severity, which ranged from 0 to 68, by clinical interview. For this cohort (187 Cases, 261 Controls), the mean severity was 3.28 and the standard deviation 11.56. DNA for GWAS analysis was isolated from saliva. The Institutional Review Board of Western Sydney Area Health Service approved this study.

**AURORA (AURO; Supplementary Table 1 #73)**

See reference for details.^6^ Potentially traumatic events were identified by the enrolling Research Assistant at participating Emergency Department sites. Traumatic stressors such as motor vehicle collision, physical assault, sexual assault, fall from >10 feet, and mass casualty events constituted automatic eligibility for enrollment. Individuals were also eligible if they reported actual or threatened serious injury, sexual violence, or death, either by direct exposure, witnessing, or learning about it. The PCL-5 checklist was used to assess PTSD symptoms over the prior thirty days via self-report using web-based surveys three months following the traumatic stress exposure.^7^ Respondents were considered to have substantial PTSD symptoms if they had a score of 31 or more on the PCL-5. The PCL-5 calculates PTSD symptom severity, which ranged from 0-80. For this cohort (348 Cases, 461 Controls). DNA for GWAS analysis was isolated from blood collected in DNA PAXgene tubes. The Institutional Review Board of the University of North Carolina at Chapel Hill approved this study.

**Biomarkers and Experiences in Adolescent Relationships (BRLS; Supplementary Table 1 #64)**

Participants were recruited from an adolescent inpatient psychiatric setting following hospitalization for suicidal thoughts or behaviors. Participants provided blood samples for GWAS, DNA methylation, and gene expression at baseline while inpatient. Potentially traumatic events were assessed using the Childhood Trauma Questionnaire and the Life Events Checklist.^8,9^ The DSM-5 Clinician Administered PTSD Scale for Children and Adolescents was administered by trained and supervised clinical interviewers to assess current PTSD at baseline, three weeks, and six months.^10^

Respondents were considered to have a current diagnosis if criteria were met. The DSM-5 Clinician Administered PTSD Scale for Children and Adolescents calculates total CAPS PTSD symptom severity, which ranged from 0 to 80, by clinical interview.

The DSM-5 Version of the K-SADS was also administered by clinical interviewers to assess lifetime and current mood and anxiety disorders at each timepoint. The study was approved by the Rhode Island Hospital IRB and funded by National Institutes of Mental Health (R01MH105379; PI Nugent).

**Biological Effects of Traumatic Experiences, Treatment and Recovery (BETR; Supplementary Table 1 #48)**

See reference for details.^11^ In total, 57 PTSD patients, 29 veteran controls (combat controls) and 32 civilian controls (healthy controls) were included. Patients were recruited from one of four outpatient clinics of the Military Mental Healthcare Organization, The Netherlands. Patients were included after a psychologist or psychiatrist diagnosed PTSD. PTSD diagnosis was confirmed using the Clinician Administered PTSD scale (CAPS ≥45^12^). The Structural Clinical interview for DSM-IV (SCID-I^13^) was applied to diagnose comorbid disorders. Control participants were recruited via advertisements, and the interviews (SCID and CAPS) were also applied to investigate PTSD symptoms and psychiatric disorders. Inclusion criteria for controls were no current psychiatric or neurological disorder, and no presence of current PTSD symptoms (CAPS ≤15). After receiving a complete written and verbal description of the study all participants gave written informed consent. The Medical Ethical Committee of the UMC Utrecht approved the study, and the study was performed in accordance with the Declaration of Helsinki.

Potentially traumatic events were identified using the Life Events Checklist for DSM-IV (LEC-IV).^9^ The CAPS for DSM-IV was used to assess PTSD over the prior month by trained researchers.^12^ The CAPS calculates PTSD symptom severity, which ranged from 0 to 107, by calculating the total CAPS score. For this cohort (57 Cases, 61 Controls), the mean severity was 50.05 and the standard deviation 32.86. Respondents were considered to have a lifetime diagnosis if DSM-IV criteria were met. Respondents were considered to have a current diagnosis if DSM-IV were met in the previous month. The Institutional Review Board of Utrecht University Medical Center approved this study.

**BioVU: Biomarkers Vanderbilt University (BIOV; Supplementary Table 1 #86)**

PTSD cases and controls were ascertained using EHR data from [n=72,824] European Ancestry patients from BioVU in Vanderbilt University Medical Center. A major goal of BioVU is to generate datasets that incorporate de-identified information derived from medical records (See Synthetic Derivative (SD) described below) and genotype information to identify factors that affect disease susceptibility, disease progression, and/or drug response. The scale of the biobank is large (>280K samples as of April 2020); rapid accrual of samples is a result of Vanderbilt’s unique ability to de-identify the samples, which in part leads to a non-human subject designation with the Institutional Review Board, thus allowing the use of blood samples collected for clinical care and scheduled to be discarded. See reference for details.^14,15^ Once a sample passes all inclusion and exclusion criteria required to participate, it is accepted in the BioVU program. Acceptance triggers the encryption program to assign a unique research ID number. The Secure Hash Algorithm (SHA) is a published and verified hash function employed for BioVU that has the property of generating a unique output for every unique input, and the property that the input cannot be inferred by cryptanalysis of the output. Using the Vanderbilt medical record number as an input, the system generates a unique, 512-bit (128 character) code that serves as a Research Unique Identifier (RUI) and is used to relate samples with de-identified computer data. Patients included in this sample were not filtered based on minimum data floor criteria. Samples accrued to date have a wide range of chronic and acute conditions, based on associated ICD-9 and 10 codes (which are extracted from the corresponding de-identified medical records). Of those currently collected samples, 44% are male and 56% female. The samples reflect the surrounding community and are 63.63% from Caucasians and 8.32% African American. Asian and Hispanic populations comprise 1% each. Pediatric samples (individuals 0 to 18 years of age) comprise 10% of all BioVU samples. PTSD case and control definitions were based on ICD-9/10 codes per the Freeze 3 established definitions (see Supplementary Methods). Due to limited case numbers using the narrow definition (n cases=1,302), only the broad definition was examined (n cases=6,679). DNA for GWAS analysis was isolated from blood. Genetic data derived from both investigator-driven projects and institutional/program initiatives result in a nested in silico resource within the BioVU Databank. By signing the data use agreement, approved investigators are required to redeposit any genetic data derived on BioVU DNA samples back into the BioVU Databank. To date, 94,474 BioVU subjects have associated genotypes measured on the MEGA-Ex platform. The Synthetic Derivative database is a research tool developed to enable studies with de-identified clinical data. The SD collection includes information extracted from the EMR systems described above and indexed by the same one-way RUI used to track samples. Content is changed by deletion or permutation of all identifiers contained within each record, as described in the proposal. The SD contains >2.3 million total records, with highly detailed longitudinal clinical data for approximately one million subjects, with an average clinical record size of 106,727 bytes (about 30 pages of text) and an average of 13 distinct diagnostic codes (ICD 9) per record. The database incorporates data from multiple sources and includes diagnostic and procedure codes (ICD 9 and CPT), basic demographics (age, gender, race), text from clinical care including discharge summaries, nursing notes, progress notes, history and physical, problem lists and multi-disciplinary assessments, laboratory values, ECG diagnoses, clinical text and electronically derived trace values, and inpatient medication orders. All clinical data are updated regularly to include patients new to VUMC, and therefore the SD, and to append new data to clinical records of existing patients as they continue to access care at VUMC. The Vanderbilt Institutional Review Board approved this study (IRB# 201609).

**Bounce Back Now (BOBA; Supplementary Table 1 #18)**

See reference for details.^16^ Potentially traumatic events were identified using National Survey (NSA) on Adolescents PTSD module which was administered by trained interviewers using computer assisted telephone interview technology. The NSA PTSD Module assessed for exposure to five types of potentially traumatic events, in addition to five specific questions about the impact of the tornado (e.g., did the tornado cause damage to your house or property?).^16,17^ The NSA PTSD Module that was administered was used to assess PTSD since the tornado, as well as during any two week period in their lifetimes, and lifetime PTSD was used in the present analyses.^17^ Respondents were considered to have a lifetime diagnosis if DSM-IV PTSD criteria (i.e., at least one re-experiencing symptom, three avoidance symptoms, and two or more arousal symptoms) were met for at least a two week period. For this cohort, 127 individuals with lifetime PTSD were matched by sex and race/ethnicity with 127 controls who did not meet criteria for lifetime PTSD. DNA for GWAS analysis was isolated from saliva collected via Oragene kits. The Institutional Review Board of the Medical University of South Carolina approved this study.

**Brazilian High Risk Cohort Study for Psychiatric Disorders (BRCO; Supplementary Table 1 #72)**

See reference for details.^18^ Potentially traumatic events were identified using The Development and Well-being Assessment (DAWBA).^19^ The DAWBA was used to assess PTSD over the prior 4 weeks by interviewers.^19^ Respondents were considered to have a diagnosis if DSM-IV PTSD criteria were met. Respondents were considered to have a current diagnosis if DSM-IV PTSD criteria were met. For this cohort, we genotyped 74 cases with PTSD, 1170 controls without any other psychiatric disorder and 946 with another psychiatric diagnosis. Besides, parents’ genotypes are also available (N=2007 controls without any psychiatric disorder diagnosed). DNA was isolated from saliva or blood (N=201, probands only). The Institutional Review Board of the Hospital de Clinicas from Porto Alegre, University of São Paulo, and Federal University of São Paulo approved this study.

**Canadian Longitudinal Study on Aging (CANA; Supplementary Table 1 #81)**

See reference for details.^20,21^ The Primary Care Post-Traumatic Stress Disorder (PC-PTSD) screen scale was used to assess PTSD in the last month by interviewers.^22^ Respondents were considered to have a diagnosis if they endorse at least three items on the PC-PTSD screen scale. This cohort has 796 Cases and 15,565 Controls; all subjects are of self-reported European ancestry confirmed by PCA. DNA for GWAS (Affymetrix Axiom arrays) analysis was isolated from blood. The Institutional Review Board of the Centre for Addiction and Mental Health# approved this study.

**Childhood Trauma Study (QIMR; Supplementary Table 1 #30)**

See references for details.^23,24^ Potentially traumatic events were identified using a semi-structured psychiatric diagnostic telephone assessment that included questions on childhood maltreatment.^25^ Lifetime DSM-IV PTSD (binary measure) was assessed using a modified version of the measure from the National Comorbidity Survey by telephone interviewers trained by an experienced clinical psychologist.^26^ For respondents who had experienced more than one potentially traumatic event, assessment of lifetime PTSD focused on the event identified by each respondent as most disturbing. DNA for GWAS analysis was isolated from blood. The Queensland Institute of Medical Research Ethics Committee and the Washington University School of Medicine Human Research Protection Office approved this study.

**Child Trauma and Neural Systems Underlying Emotion Regulation (KMCT, KMC2; Supplementary Table 1 #19, #61)**

Potentially traumatic events were identified using The UCLA PTSD Reaction Index,^27^ the Childhood Experiences of Care and Abuse Interview,^28^ and the Childhood Trauma Questionnaire.^29^ The Clinician Administered PTSD Scale for Children^10^ was used to assess both lifetime and current PTSD by trained clinical interviewers.^28^ Children and a parent or guardian completed the interview, and an or rule was used to assign diagnoses. Respondents were considered to have a diagnosis if DSM-5 criteria were met. Respondents were considered to have a current diagnosis if DSM-5 criteria were met. The UCLA PTSD Reaction Index^27^ calculates PTSD symptom severity. Children and a parent or guardian each completed this measure, and we used the highest score from either the child or parent, which ranged from 0 to 67 in our sample. For this cohort (KMCT: 133 Cases, 122 Controls; KMC2: 59 Cases, 89 Controls), the mean severity was 17.57 and the standard deviation 17.94. DNA for GWAS analysis was isolated from saliva. The Institutional Review Board of the University of Washington approved this study.

**Chitwan Valley Family Study (CVHS; Supplementary Table 1 #67)**

See reference for details.^30^ Potentially traumatic events were identified using a version of the Composite International Diagnostic Interview (CIDI) 3.0, modified for use in Nepal (translation and cultural adaptation, after extensive expert review, pilot testing, and clinical validation).^31^ The PTSD module of the Nepal CIDI was used to assess PTSD over the lifetime (also generating 12-month and 30-day diagnoses), and was administered by professional lay interviewers who were trained and certified through the University of Michigan’s WHO CIDI Training and Reference Centre. Respondents were considered to have a diagnosis if DSM-IV or ICD-10 criteria were met. PTSD symptom severity and interference were also measured. DNA for GWAS analysis was isolated from saliva samples collected using the Genotek Oragene protocol. The Institutional Review Board of the University of Michigan, as well as the Nepal Health Research Council, approved and provided on-going regulation of this study.

**CHOICE (FEEN; Supplementary Table 1 #37)**

See reference for details.^32-41^ Potentially traumatic events were identified using the standard trauma interview.^42^ The PTSD Symptom Scale – Interview (PSS-I) was used to assess PTSD over the prior two weeks for the trauma of interest by postdoctoral or graduate level assessors trained to reliability^13^. The Structured Clinical Interview (SCID-IV) was used to assess lifetime PTSD (not current) for a trauma not the focus of treatment by postdoctoral or graduate level assessors trained to reliability.^43^ Respondents were considered to have a current diagnosis if on the PSS-I they met symptom-level DSM-IV diagnostic criteria. The PSS-I also provides PTSD symptom severity, with a range from 0 to 51. For this cohort (104 Cases), the mean severity was 32.63 and the standard deviation 4.87. DNA for GWAS analysis was isolated from blood. The Institutional Review Board of University Hospitals approved this study.

**Collaborative Study on the Genetics of Alcoholism (RCOG; Supplementary Table 1 #83)**

The Collaborative Study on the Genetics of Alcoholism (COGA) is a large, multi-site study of US families densely affected with alcohol use disorders designed to identify genetic and environmental factors involved in substance use disorders and related psychiatric disorders. See reference for details.^44-46^ Potentially traumatic events were identified using the Semi-Structured Assessment for the Genetics of Alcoholism (SSAGA).^45^ The SSAGA was used to assess PTSD over the lifetime via structured diagnostic interviews conducted by trained research associates.^47^ Respondents were considered to have a lifetime diagnosis if DSM-IV criteria were met. Among those who endorsed having been exposed to a traumatic event in their lifetime, the SSAGA calculates DSM-IV PTSD symptom severity. For this cohort of 4,989 genotyped individuals (599 Cases, 4390 Controls), the mean symptom severity was 2.7 and the standard deviation 4.6. DNA for GWAS analysis was isolated from blood. The Institutional Review Board of SUNY Downstate Medical Center approved this study.

**Cohen Veterans Center Study (COM1; Supplementary Table 1 #50)**

See reference for details.^48-50^ This is a multi-site study that ran through NYUMC and Stanford University and Palo Alto VAMC. The VA Palo Alto Health Care System has an affiliation with Stanford University School of Medicine. Dr. Charles Marmar is the overall PI for this study.

Potentially traumatic events were identified using clinical interview that was administered by a licensed psychologist.^51^ The CAPS was administered by clinicians to assess PTSD for two time periods: the preceding 30 days and a one-month period in the past when symptoms were the worst, by respondent’s subjective account.^12^ The CAPS *calculates PTSD symptom severity by summing the scores for all items, which ranged from 0 to 80 in the full range* and ranged from 0 to 56 in this dataset (in the Cohen Veterans Center study). For this cohort (232 Cases, 802 Controls), the mean severity was 12.14 and the standard deviation 11.85. Respondents were considered to have a current diagnosis if DSM-5 criteria were met in the preceding 30 days. Respondents were considered to have a lifetime diagnosis if DSM-5 criteria were met in the month when symptoms were the worst. The Institutional Review Board of NYUMC and Stanford University approved this study.

**Cortical Excitability: Biomarker and Endophenotype in Combat Related PTSD (WANG; Supplementary Table 1 #59)**

Potentially traumatic events were identified using CAPS Life Event Checklist. The CAPS was used to assess PTSD over the prior 4 weeks, by research coordinators/interviewers.^12^ Respondents were considered to have a diagnosis if CAPS>45. Respondents were considered to have a current diagnosis if CAPS>45. The CAPS calculates PTSD symptom severity, which ranged from 0 to 123 out of a possible 136, by interview.^12^ For this cohort (208 Cases, 87 Controls), the mean severity of cases was 76.22 and the standard deviation was 16.85. DNA for GWAS analysis was isolated from whole blood. The Institutional Review Board of Medical University of South Carolina approved this study.

**Danish military study** **(DAMI; Supplementary Table 1 #28)**

Potentially traumatic events were identified with 11 single items listing potentially traumatic events occurring during deployment. A scale developed for the Danish military, the PRIM-PTSD, was used to assess PTSD-symptoms over the previous 3 months^52^. The PRIM-PTSD calculates PTSD symptom severity, with a possible range of 12-48. For this cohort (462 Cases, 2019 Controls after quality control), the mean severity was 17.97 and the standard deviation 5.92. PTSD cases were defined as having a PRIM-PTSD score at or above 25, equaling a score of 44 on the PTSD Checklist. DNA for GWAS analysis was isolated from neonatal blood spots. The Regional Committee on Health Research Ethics, Region Zealand, approved this study.

**Danish iPSYCH PTSD samples (DAI2; Supplementary Table 1 #85)**

See reference for details.^53-55^ The Danish iPSYCH PTSD samples were identified for analysis using infrastructure provided by the iPSYCH project.^56^ The iPSYCH project is a case cohort study, drawing individuals born in Denmark between 1981 and 2005, and obtaining all cases diagnosed with six disorders plus 30,000 random individuals from the same population cohort as controls. PTSD was not one of the original six diagnoses within iPSYCH, but cases were identified in linked records (i.e. PTSD diagnosis comorbid with one of the six ascertained disorders or PTSD diagnosis in one of the 30,000 iPSYCH “controls”). PTSD was assessed via clinician diagnosis according to ICD-10 (F43.1), as obtained from either of two registers in Denmark: the Danish Psychiatric Central Research Register and/or the Danish National Patient Register. Diagnoses in the registers are for current disorders (i.e. not lifetime). PTSD severity was not available. DNA for GWAS analysis was isolated from bloodspots from the Danish Neonatal Screening Biobank hosted by the Statens Serum Institut, as described previously.^54,55^ The study was approved by the Regional Danish Ethics Committee and the Danish Data Protection Agency.

**DCS Rothbaum Study (DCSR; Supplementary Table 1 #38)**

See reference for details.^57,58^ The authors examined the effectiveness of virtual reality exposure augmented with D-cycloserine or alprazolam, compared with placebo, in reducing PTSD due to military trauma.

After an introductory session, five sessions of virtual reality exposure were augmented with D-cycloserine (50 mg) or alprazolam (0.25 mg) in a double-blind, placebo-controlled randomized clinical trial for 156 Iraq and Afghanistan war veterans with PTSD.^59^

PTSD symptoms significantly improved from pre- to posttreatment across all conditions and were maintained at 3, 6, and 12 months. There were no overall differences in symptoms between D-cycloserine and placebo at any time. Alprazolam and placebo differed significantly on the Clinician-Administered PTSD Scale (CAPS) score at posttreatment and PTSD diagnosis at 3 months posttreatment; the alprazolam group showed a higher rate of PTSD (82.8%) than the placebo group (47.8%).^12^ Between-session extinction learning was a treatment-specific enhancer of outcome for the D-cycloserine group only. At posttreatment, the D-cycloserine group had the lowest cortisol reactivity and smallest startle response during virtual reality scenes.

A six-session virtual reality treatment was associated with reduction in PTSD diagnoses and symptoms in Iraq and Afghanistan veterans, although there was no control condition for the virtual reality exposure. There was no advantage of D-cycloserine for PTSD symptoms in primary analyses. In secondary analyses, alprazolam impaired recovery and D-cycloserine enhanced virtual reality outcome in patients who demonstrated within-session learning. D-cycloserine augmentation reduced cortisol and startle reactivity more than did alprazolam or placebo, findings that are consistent with those in the animal literature.

**Defining Essential Features of Neural Damage (DEFE; Supplementary Table 1 #12)**

See reference for details.^60,61^ Potentially traumatic events were identified using the Clinician-Administered PTSD Scale for DSM-IV (CAPS-IV), the Combat Exposure Scale (CES), and items from the Deployment Risk and Resilience Inventory (DRRI).^12,51,62^ The CAPS-IV was also used to assess PTSD currently (over the prior 2 weeks) and over the lifetime by trained interviewers.^12^ The CAPS-IV calculates PTSD symptom severity, which ranged from 0 to 120. For this cohort (88 Cases, 62 Controls), the mean severity was 31.43 and the standard deviation 25.01. Respondents were considered to have a lifetime diagnosis if they met DSM-IV criteria for PTSD (measured using CAPS-IV) either current or past. Respondents were considered to have a current diagnosis if they met DSM-IV criteria for PTSD (measured using CAPS-IV) at the time of the assessment. The Institutional Review Board of the Minneapolis VA Health Care System approved this study.

**Detroit Neighborhood Health Study (DNHS, ADNH; Supplementary Table 1 #4 and #45)**

See reference for details.^63^ Potentially traumatic events (PTEs) were identified using a list of 19 PTEs.^64^ The PTSD Checklist-Civilian (PCL-C) was used to assess PTSD over the lifetime by self-report during structured telephone interviews by referencing two traumatic events; one the respondent regarded as the worst and a second randomly selected event from the list of remaining PTEs (if the respondent experienced more than one traumatic event).^65^ Respondents were considered to have a diagnosis if all six DSM-IV criteria were met for either the worst or random event. Additional questions assessed the timing, duration, severity or illness, and disability resulting from symptoms. PTSD symptom severity, which ranged from 17 to 85, was assessed by summing the respondents’ ratings of the 17 post-traumatic symptoms on a scale indicating the degree to which the respondent was bothered by a particular symptom as a result of the worst trauma, ranging from 1 (not at all) to 5 (extremely). All DNHS participants, include *N* = 2081, of which cases *N* = 408 and controls *N*=1532. DNA for GWAS analysis was isolated from peripheral blood or saliva. The Institutional Review Board at the University of Michigan and University of North Carolina Chapel Hill approved this study.

**Drakenstein Child Health Study - South African sample (SAFR, SAF2; Supplementary Table 1 #3, #77)**

See reference for details.^66 67 68^ The modified PTSD Symptom Scale (PSS) was used to assess PTSD.^43^ Specifically, the re-experiencing symptom cluster was considered met if the sum of reported symptoms totalled greater than or equal to 1; the avoidance/emotional numbing reported symptoms were greater than or equal to 3; and increased arousal cluster reported symptoms were greater than or equal to 2. Participants who scored above threshold in each of the clusters and had symptom duration for at least 1 month were classified as PTSD cases. For cohort SAF2, second round, (117 cases, 318 controls), controls consisted of 150 participants who were trauma-exposed only, and 168 participants who were trauma unexposed. There were only 87 participants with viable mean severity data, and thus, was not included. The Faculty of Health Sciences human research ethics committee of the University of Cape Town (UCT) approved this study.

**EA CRASH (EACR; Supplementary Table 1 #42)**

See reference for details.^69^ European American individuals were enrolled in the Emergency Department within 24 hours following motor vehicle collision (MVC) trauma/stress.^70^ The Impact of Events Scale-Revised (IES-R) was used to assess PTS symptom severity over the past week by research assistants 1 year following MVC.^70^ Respondents were considered to have a diagnosis if they scored 33 or higher on the IES-R questionnaire. The IES-R inventory calculates PTS symptom severity, which ranged from 0 to 88, by scoring a participant’s answers to 22 questions on a scale of 0-4 about symptoms of avoidance, intrusions, and hyperarousal. For this cohort (88 Cases, 276 Controls), the mean PTS symptom severity was 19.2 and the standard deviation was 18.4, measured 1-year after the MVC. DNA for GWAS analysis was isolated from blood collected in DNA PAXgene tubes. The Institutional Review Board of The University of North Carolina at Chapel Hill approved this study.

**Emory Healthcare Veteran’s Program (EHVP; Supplementary Table 1 #74)**

See reference for details.^71^ Potentially traumatic events were identified using the Life Events Checklist.^9^ The CAPS was used to assess PTSD over the prior month by masters and doctoral level clinicians.^72^ Respondents were considered to have a diagnosis if CAPS PTSD diagnostic criteria were met. The PTSD Symptom Checklist calculates PTSD symptom severity, which ranged from 0 to 80, by self-report.^73^ For this cohort (280 cases, 68 controls), the mean severity was 50.52 and the standard deviation 14.31. DNA for GWAS analysis was isolated from saliva. The Institutional Review Board of Emory University approved this study.

**Estonian Biobank (ESBB; Supplementary Table 1 #93)**

PTSD cases and controls were ascertained using EHR data from 200,000 participants from the Estonian Biobank (EstBB) based on their Health Insurance Fund Treatment Bills. See reference for details.^74^ Individuals included in this sample had no minimum data floor criteria. PTSD case and control definitions were based on ICD-10 codes per the Freeze 3 established definitions (see Supplementary Methods). For definition of post-concussive syndrome ICD-10 code F07.2 was used. We determined 623 cases of the narrow PTSD definition (n= 524 women; n=99 men) and 19,723 cases of broad PTSD (n= 15,251women; n=4,472 men). Controls were defined as undiagnosed participants (n=177,970; n= 114,297 women; n=63,673 men) and for the analysis of narrow PTSD we further excluded among controls all individuals with only 1 diagnosis of PTSD, post-concussive syndrome, and those with other stress disorders (n=177,210; n=113,986 women; n=63,224 men).

DNA for GWAS analysis was isolated from blood. Genotyping of DNA samples from the Estonian Biobank was done at the Core Genotyping Lab of the Institute of Genomics, University of Tartu using the Illumina Global Screening Arrays. Altogether 200,000 samples were genotyped and then PLINK format files were created using Illumina GenomeStudio v2.0.4. During the quality control all individuals with call-rate < 95% or mismatching sex that was defined based on the heterozygosity of X chromosome and sex in the phenotype data, were excluded from the analysis. Variants were filtered by call-rate < 95% and HWE p-value < 1e-4 (autosomal variants only). Variant positions were updated to Genome Reference Consortium Human Build 37 and all variants were changed to be from TOP strand using reference information provided by Dr. Will Rayner from the University of Oxford (<https://www.well.ox.ac.uk/~wrayner/strand/>). Before imputation variants with MAF<1% and Indels were removed. Prephasing was done using the Eagle v2.3 software^75^ (number of conditioning haplotypes Eagle2 uses when phasing each sample was set to: --Kpbwt=20000) and imputation was carried out using Beagle v.28Sep18.793^76,77^ with an effective population size n_e_=20,000. As a reference, Estonian population specific imputation reference of 2,297 WGS samples was used.^78^ We conducted the GWASes using the SAIGE software (version 0.43.1)^79^ adjusting for the first ten principal components of the genotype matrix, as well as for birth year and sex. The Institutional Review Board of Estonian Committee on Bioethics and Human Research approved this study.

**Family Study of Cocaine Dependence and Collaborative Genetic Study of Nicotine Dependence (FSCD, COGA, COGB; Supplementary Table 1 #7-9)**

See references for details.^80,81^ A module from the Diagnostic Interview Schedule for DSM-IV (DIS-IV),^82^ a structured assessment that evaluated the presence or absence of psychiatric disorders according to the DSM-IV^83^ criteria was used to evaluate PTSD in a sample of 471 cases and 3,568 controls. A history of fifteen specific traumatic events were queried including rape or sexual assault, assaultive violence (e.g., shot, stabbed), witnessing trauma to others, and non-violent trauma (e.g., serious accident, sudden death of a loved one). The traumatic events were assessed using closed-ended questions (e.g., Have you ever been raped or sexually assaulted?) with nominal response options (i.e., Yes or No). Participants were asked to select the most distressing event and were subsequently evaluated for symptoms of PTSD. A diagnosis of PTSD was dependent on Criterion A, which required intense fear, helplessness, or horror in association with the most distressing event. Interview data were checked for consistency by a senior editor and entered into a computerized data file. Lifetime psychiatric diagnoses were made by a computer algorithm that analyzed responses to the interview using DSM-IV criteria. The Washington University School of Medicine IRB approved the studies.

**FinnGen (FING; Supplementary Table 1 #90)**

PTSD cases and controls were ascertained using EHR data from 269,078 patients from FinnGen. We did not impose a minimum data floor criterion for patient inclusion for FinnGen. See reference for details.^84,85^ PTSD case and control definitions were based on ICD- 9/10 codes per the Freeze 3 established definitions (see Supplementary Methods). Due to the non-availability of ICD code for post-concussive syndrome (F07.81), patients with post-concussive syndrome were not excluded from controls group for PTSD narrow definition. There is a total of 9,801 cases and 249,993 controls for PTSD broad definition (female cases/controls: 6,793/140,020; male cases/controls: 3,008/109,973) and 863 cases and 249,993 controls for PTSD narrow definition (female cases/controls: 704 /140,020; male cases/controls: 159/109,973).

For the Finnish Institute of Health and Welfare (THL) driven FinnGen preparatory project (FinnGen), all patients and control subjects had provided informed consent for biobank research, based on the Finnish Biobank Act. Alternatively, older cohorts were based on study specific consents and later transferred to the THL Biobank after approval by Valvira, the National Supervisory Authority for Welfare and Health. Recruitment protocols followed the biobank protocols approved by Valvira. The Ethical Review Board of the Hospital District of Helsinki and Uusimaa approved the FinnGen study protocol Nr HUS/990/2017. The FinnGen preparatory project is approved by THL, approval numbers THL/2031/6.02.00/2017, amendments THL/341/6.02.00/2018, THL/2222/6.02.00/2018 and THL/283/6.02.00/2019. All DNA samples and data in this study were pseudonymized.

**Fort Campbell study (FTCB; Supplementary Table 1 #52)**

Fort Campbell is a United States Army installation located astride the Kentucky-Tennessee border between Hopkinsville, Kentucky, and Clarksville, Tennessee. Fort Campbell is home to the 101st Airborne Division and the 160th Special Operations Aviation Regiment. One thousand, seven hundred and ninety-three (N=1793) active duty members of the Army’s 101st Airborne Division who deployed to Afghanistan participated in the study. Each participant was evaluated one-three times at the Fort Campbell U.S. Army military installation. The first evaluation took place prior to deployment in January-February, 2014, the second evaluation took place 3 days upon return from deployment and the third evaluation occurred 90-180 days upon return from deployment.

Potentially traumatic events were identified using self-report questionnaire that included PCL 5.^73^ The PTSD Checklist for DSM 5 (PCL-5) was used to assess PTSD during each phase of the study (pre-deployment, 3 days post deployment and 90-180 days post deployment).^73^ The PCL-5 score calculates PTSD symptom severity by summing the scores for all items, which ranged from 0 to 80 in the full range and ranged from 0 to 75 in this dataset. For this cohort (114 Cases, 1624 Controls), the mean severity was 6.81 and the standard deviation 11.24. Respondents were considered to have a current diagnosis if PCL5 score is at least 33. The Institutional Review Board of NYUMC and HARPO (DoD IRB) approved this study.

**Genetic and Environmental Predictors of Combat-Related PTSD (STRO; Supplementary Table 1 #35)**

See reference for details.^86^ Subjects in this study were included from a larger STRONG STAR pre-/post-deployment study of deploying soldiers from Fort Hood in Killeen, Texas. The data included in this analysis are from the pre-deployment assessment. Potentially traumatic events were identified using the Life Events Checklist (LEC).^9^ The PTSD Checklist–Military Version (PCL-M) was used to assess PTSD over the prior month by self-repor.^2^ Subjects were considered to have a current diagnosis if the total score on the PCL was ≥ 50 and classified as a control if their PCL total score was < 50. In addition, both cases and controls were required to report having directly experienced or witnessing a traumatic event on the LEC. Based on these criteria, N=607 subjects were classified as having current PTSD and N=3,390 were classified as controls. DNA for GWAS analysis was isolated from blood. The Institutional Review Board of University of Texas Health Science Center at San Antonio approved this study.

**Genetic Risk for PTSD (YEHU; Supplementary Table 1 #56)**

See reference for details.^87,88^ Potentially traumatic events were identified using the CAPS, SCID, MINI, and the Life Events Checklist.^9,12,89,90^ The CAPS, SCID, and MINI were used to assess PTSD during the past month or over the prior lifetime by a PhD level clinical psychologist.^12,89,90^ Respondents were considered to have a diagnosis if at least one Criterion B symptom, at least three Criterion C symptoms, at least two Criterion D symptoms, and Criterion A, E, and F (CAPS), J1, J2, and J3 are coded “yes”, at least three or more J4 questions are coded “yes”, at least two J5 answers are coded “yes”, and J6 is coded “yes” (MINI), and/or subject experienced a traumatic event and adverse consequences were experienced, both A criteria were coded 3, at least one B criteria was coded 3, at least three C criteria were coded 3, and at least two D criteria were coded 3 (SCID). Respondents were considered to have a current diagnosis if criteria for PTSD diagnosis were met within the past month. The CAPS calculates PTSD symptom severity, which ranged from 0 to 136. For this cohort (123 cases, 43 controls), the mean severity was 80.19 and the standard deviation 2.23. DNA for GWAS analysis was isolated from whole blood. The Institutional Review Board of James J. Peters VA Medical Center and Icahn School of Medicine at Mount Sinai approved this study.

**Genetics of Posttraumatic Stress Disorder/Substance Use Disorder Comorbidity (KSUD; Supplementary Table 1 #17)**

See reference for details.^91^ Potentially traumatic events were identified using the Life Event Checklist for DSM-5 (LEC-5).^92^ The self-report PTSD Checklist for DSM-5 (PCL-5)^73^ was used to assess PTSD symptoms over the prior month. The PCL-5 contains 20 items which summed provide a measure of PTSD symptom severity (scores range from 0 to 80). For this cohort (137 Cases, 106 Controls), the mean severity was 40.57 and the standard deviation 20.96. Respondents were considered to have a current diagnosis if total symptom severity was at or above 38. The Institutional Review Board of Kent State University approved this study.

**GMRF-QUT (GMFR; Supplementary Table 1 #55)**

See reference for details.^93^ Potentially traumatic events were identified using Criterion A event. The Clinician Administered PTSD Scale for DSM 5 (CAPS-5) was used to assess PTSD over the prior 2 weeks and lifetime by clinical psychologists.^94^

Respondents were considered to have a current diagnosis if CAPS-5 criteria were met. The CAPS-5 calculates PTSD symptom severity, which ranged from 0 to 56. For this cohort (100 Cases, 124 Controls), the mean severity was 9.63 and the standard deviation 10.05. DNA for GWAS analysis was isolated from peripheral blood. Ethics approval for the project was obtained from the Department of Veterans’ Affairs, Greenslopes Private Hospital, and Queensland University of Technology Human Research Ethics Committees. This study was carried out in accordance with The Code of Ethics of the World Medical Association (Declaration of Helsinki).

**Genetics Research and the Childbearing Year (GRAC; Supplementary Table 1 #54)**

See reference for details.^95^ A total of 29 potentially traumatic events were identified using the Life Stressor Checklist^96^. PTSD symptoms were assessed using the National Women’s Study PTSD Module (NWS-PTSD), a widely used scale designed for use by lay interviewers, that consists of 20 items that assess DSM-IIl-R PTSD Criteria B, C, and D symptoms. The NWS-PTSD was performed by trained lay interviewers using computer-aided telephone interviewing and epidemiological methods (forced choice yes or no).^97,98^ Respondents were considered to have a diagnosis if lifetime DSM-IV PTSD criteria were met. Respondents were considered to have a current diagnosis if past month DSM-IV criteria were met. The NWS-PTSD assesses the number of PTSD symptoms endorsed, which ranged from 0 to 17. For this cohort (140 Cases, 138 Controls), the mean PTSD symptom count (out of 17 possible symptoms) was 6.4 and the standard deviation 5.5. DNA for GWAS analysis was isolated from saliva (Oragene tube). The Institutional Review Board of the University of Michigan approved this study.

**Grady Trauma Project (EGHS, GTPC; Supplementary Table 1 #44 and #47)**

See reference for details.^99^ The modified PTSD Symptom Scale (PSS), a psychometrically valid 17-item self-report scale assessing PTSD symptomatology over the prior 2 weeks, was used to assess PTSD.^100^ Consistent with prior literature, the PSS frequency items (0 indicates not at all to 3 indicates ≥5 times a week) to obtain a continuous measure of PTSD symptom severity ranging from 0 to 51. For this sample, the PSS frequency items had standardized α=.90 (mean [SD], 13.81 [11.96]). No clearly established PSS cutoff score for PTSD diagnosis has been established; however, DSM-IV criteria for PTSD can be applied to PSS frequency items to create a proxy variable for PTSD diagnostic status. The Institutional Review Boards of Emory University School of Medicine and Grady Memorial Hospital approved this study.

**Grady Trauma Project – Neuroimaging (DGTP; Supplementary Table 1 #68)**

See reference for details.^101^ Potentially traumatic events were identified using the Life Events Checklist (LEC).^9^ The CAPS-IV was used to assess PTSD over the prior month and lifetime by interviewers. Respondents were considered to have a diagnosis if DSM-IV-TR criteria were met.^12^ The CAPS-IV calculates PTSD symptom severity, which ranged from 1 to 115, by adding the intensity and frequency of each symptom. For this cohort (18 Cases, 9 Controls), the mean severity was 30.13 and the standard deviation 22.6 for current PTSD. DNA for GWAS analysis was isolated from saliva. The Institutional Review Board of Emory University approved this study.

**Growing Up Today Study (GUTS; Supplementary Table 1 #21)**

See reference for details.^102^ Potentially traumatic events were identified using the Brief Trauma Questionnaire,^103^ plus questions on stalking and intimate partner violence (specific events queried included: witness an attack, get attacked, disaster, serious accident, attack on family member, stalked, family member killed in violence, served in war zone/saw war casualties, serious injury to self, physical intimate partner violence, sexual intimate partner violence). Breslau’s Short Screening Scale for DSM-IV PTSD was used to assess PTSD over the lifetime by self-report of symptoms.^104^ Respondents were considered to have a lifetime diagnosis if they reported experiencing 4 or more symptoms. Current diagnosis was not assessed. Breslau’s Short Screening Scale for DSM-IV PTSD calculates PTSD symptom severity, which ranged from 0 to 7, by counting the number of symptoms. For this cohort (312 Cases, 312 Controls), the severity was mean=2.63, SD=2.31 (cases: mean=4.93, SD=0.94; controls: mean=0.34, SD=0.47). DNA for GWAS analysis was isolated from saliva. The Institutional Review Board of Brigham and Women’s Hospital approved this study.

**Impact of PTSD and Distress Tolerance on Treatment Readiness in the Detoxification Setting (DELB; Supplementary Table 1 #71)**

Potentially traumatic events were identified using a modified version of the Traumatic Stress Scale.^105^ The self-report PTSD Checklist for DSM-5 (PCL-5) was used to assess PTSD symptoms over the prior month.^73^ The PCL-5 contains 20 items which summed provide a measure of PTSD symptom severity (scores range from 0 to 80). For this cohort (56 Cases, 21 Controls), the mean severity was 45.92 and the standard deviation 17.26. Respondents were considered to have a current diagnosis if total symptom severity was at or above 38. DNA for GWAS analysis was isolated from saliva. The Institutional Review Board of Kent State University approved this study.

**Injury and Traumatic Stress Consortium (INTR; Supplementary Table 1 #27)**

Subjects were participants in studies of the INTRuST Consortium, some of which are cited.^106-108^ Potentially traumatic events were identified using the Life Events Checklist for DSM-IV (LEC)^2^ and/or the Deployment Risk and Resilience inventory (DRRI).^109^ The PTSD Checklist for DSM-IV (PCL)^2^ was used to indicate a likely diagnosis of PTSD (or healthy control status); in some cases, this diagnosis was corroborated by the Clinician-Administered PTSD Scale (CAPS).^12,110^ The PCL was used as an indicator of PTSD symptom severity, which ranged from 17-85. DNA for GWAS analysis was isolated from whole blood. The Institutional Review Boards of UCSD (the Coordinating Center) and all the participating institutions approved this research.

**IVS (BRYA; Supplementary Table 1 #10)**

See reference for details.^111^ Potentially traumatic events were identified using the Recent Life Events questionnaire.^5^ The Clinician Administered PTSD Scale (CAPS) was used to assess PTSD over the prior 4 weeks by psychologist interviewers.^12^ Respondents were considered to have a current diagnosis if DSM-IV were met. The CAPS calculates PTSD symptom severity, which ranged from 0 to 136, by clinical assessment. For this cohort (90 Cases, 312 Controls), the mean severity was 25.10 and the standard deviation 23.98. DNA for GWAS analysis was isolated from saliva. The Institutional Review Board of Western Sydney Area Health Service approved this study.

**Marine Resiliency Study (MRSC, BAKE; Supplementary Table 1 #1 and #57)**

See reference for details.^112,113^ Participants were recruited from two studies including military personnel: (1) the Marine Resiliency Study, a prospective PTSD study with longitudinal follow-up (pre- and post-exposure to combat stress) of U.S. Marines bound for deployment to Iraq or Afghanistan, and (2) a cross-sectional study involving a cohort of combat-exposed active duty or previously deployed service members (CAVC), including PTSD cases and controls with comparable psychosocial and clinical phenotypes. PTSD was diagnosed up to 3 times, once before deployment and 3 and/or 6 month post deployment. Post-traumatic stress (PTS) symptoms were assessed using a structured diagnostic interview, the Clinician Administered PTSD Scale (CAPS), and PTSD diagnosis followed the DSM-IV criteria.^12^ All participants included in this study met the *DSM-IV* criteria A1 event. For participants assessed at multiple timepoints, the timepoint with the highest CAPS score was used. Genomic DNA was prepared from blood leukocytes and genotyping was carried out by Illumina (http://www.illumina.com/) using the HumanOm-niExpressExome (HOEE) array with 951,117 loci and by RUCDR (http://www.rucdr.org) using the HOEE array with 967,537 loci. The study was approved by the University of California San Diego Institutional Review Board, and all participants provided written informed consent to participate.

**Mass General Brigham Biobank (MGBB; Supplementary Table 1 #87)**

PTSD cases and controls were ascertained using EHR data from n=24,824 patients in the Mass General Brigham (MGB; formerly Partners Healthcare) Biobank in the MGB health system. See reference for details about the Biobank.^114^ Patients included in this sample met a minimum data floor criteria of at least 3 visits after 2005 that were each more than 30 days apart with at least one clinical note. PTSD case and control definitions were based on ICD-9/10 codes per the Freeze 3 established definitions (see Supplementary Methods). DNA for GWAS analysis was isolated from blood and quality control procedures were performed on the genomic data, with participants of European ancestry retained for these analyses. The Institutional Review Board at Mass General Brigham (formerly Partners HealthCare) approved this study

**MayoGC (MAYO; Supplementary Table 1 #94)**

PTSD cases and controls were ascertained using EHR data from 7257 patients from the MayoGC in Mayo Clinic Health System (Table 1). See reference for details.^115^ Patients included in this sample met a minimum data floor criterion of at least three visits 90 days apart to the Mayo Clinic Health System. PTSD case and control definitions were based on ICD-9/10 codes per the Freeze 3 established definitions (see Supplementary Methods). DNA for GWAS analysis was isolated from whole blood. The Institutional Review Board of Mayo Clinic approved this study.

MayoGC is comprised of 10 different genetic studies recruiting patients from the Mayo Clinic Health System.^115^ Each of the genetic datasets were genotyped using different genotyping platforms and each were run through the Mayo Clinic Genotype QC pipeline (Table 2). After QC, each study was imputed using the Michigan Imputation server (Minimac4 1.2.1) with the HRC reference panel (Reference Panel: [hg19]). We note that some of the below studies did not have genotype information for sex chromosomes.

*Table 1: Distribution of each PTSD phenotype in MayoGC dataset that met minimum data floor criterion*

| **Definition** | **Level** | **All**  N = 7257 | **Male**  N = 3632 | **Female**  N = 3625 |
| --- | --- | --- | --- | --- |
| Broad PTSD (1+ code) | Missing | 0 | 0 | 0 |
|  | Control | 6136 | 3240 | 2896 |
|  | Case | 1121 | 392 | 729 |
| Narrow PTSD (2+ code) | Missing | 1077 | 381 | 696 |
|  | Control | 6136 | 3240 | 2896 |
|  | Case | 44 | 11 | 33 |

*Table 2: Study and genotyping chip included in MayoGC analysis*

```
|               |   1|   2|   3|   4|    5|   6|   7|   8|
|:--------------|---:|---:|---:|---:|----:|---:|---:|---:|
|MayoGC-BRAIN   |   0|   0|   0| 142|    0|   0|   0|   0|
|MayoGC-CERHAN3 |   0|   0|   0|   0|    0| 321|   0|   0|
|MayoGC-CLL     | 199|   0|   0|   0|    0|   0|   0|   0|
|MayoGC-EM      |   0|   0|   0|   0| 2910|   0|   0|   0|
|MayoGC-LUNG    |   0|   0| 138| 127|    0|   0|   0|   0|
|MayoGC-NHL     |   0|   0|   0|   0|  169|   0|   0|   0|
|MayoGC-OV1     |   0|   0|   0| 432|    0|   0|   0|   0|
|MayoGC-OV2     |   0| 409|   0|   0|    0|   0|   0|   0|
|MayoGC-PA      |   0|   0|   0|   0|    0|   0| 287| 232|
|MayoGC-VT      |   0|   0|   0|   0| 1891|   0|   0|   0|

Genotyping chip
1: Affymetrix 6.0
2: Human Omni 2.5-4v1
3: Illumina 370K
4: Illumina 610K
5: Illumina 660K
6: Ilumina Omni Express
7: PanScan I-550K
8: PanScan II-610K
```

**McLean Trauma Sample (TEIC; Supplementary Table 1 #39)**

The McLean Trauma Sample consists of three separate studies lead by Drs. Milissa Kaufman and Martin Teicher. See references for details.^116-118^ The first study was conducted at McLean Hospital’s Developmental Biopsychiatry Research Program (PI: Martin Teicher, MD, PhD) entitled “Sensitive Periods, Brain Development and Depression Study”. The group aimed to test the hypothesis that there are discrete sensitive periods when exposure to abuse or loss is maximally associated with risk for developing psychiatric disorders, specifically major depression and that risk for developing depression coincided with exposure to abuse during sensitive periods of hippocampal and prefrontal cortex vulnerability. These hypotheses were tested in a sample of 517 individuals (20-25 years of age) recruited from the community. Degree and timing of developmental exposure to abuse and loss across each childhood stage was quantified retrospectively using the Maltreatment and Abuse Chronology of Exposure (MACE) scale,^119^ as well as Traumatic Antecedent Interview,^120^ Childhood trauma Questionnaire^8^ and Adverse Childhood Experiences scale.^121^ Lifetime and current psychopathology including PTSD was assessed by trained psychologists, psychiatrists and clinical nurse specialists, using the Structured Clinical Interview for DSM-IV-TR.^122^ Respondents were considered to have a current diagnosis if DSM-IV-TR criteria were met in the preceding 30 days.

The second study was also conducted at McLean Hospital’s Developmental Biopsychiatry Research Program (PI: Martin Teicher). The key aims of the project were to test in a prospective study whether neurobiological correlates such as T2-relaxation time in dorsolateral prefrontal and anterior cingulate cortex and a large cerebellar lingual size can predict degree of drug and alcohol use in individuals with histories of childhood abuse and neglect, with an emphasis on sensitive periods of maximal exposure. These hypotheses were tested in a sample of 157 individuals (18-19 years of age) recruited from the community. Structured Clinical Interviews for DSM-IV (SCID-IV) Axis I and II psychiatric disorders were used for diagnoses.^122^ Mental health professionals (psychiatrists, Ph.D. psychologists, clinical nurse specialists) performed all the interviews and psychological assessments. In addition to the Maltreatment and Abuse Chronology of Exposure (MACE) scale,^119^ the 100-item semi-structured Traumatic Antecedents Interview^120^ was also used to assess maltreatment history, as well as the Childhood Trauma Questionnaire^8^ and the Adverse Childhood Experience score.^121^ PTSD was diagnosed using the SCID-IV-TR and the CAPS.^12^

The third study was conducted at McLean’s Dissociative Disorders and Trauma Research Program (PI: Milissa Kaufman, MD, PhD) entitled “Evaluating the Neurobiological Basis of Traumatic Dissociation in a Cross-Diagnostic Sample of Women with Histories of Childhood Abuse and Neglect”. Patients were recruited from inpatient and partial/residential treatment programs at McLean Hospital as part of a larger study on trauma-related dissociation comprised of diagnostic interviews, self-reports, neuropsychological testing, and neuroimaging protocols. All individuals endorsed a history of childhood trauma exposure, as assessed by the Traumatic Events Interview and Childhood Trauma Questionnaire. All participants also met criteria for DSM-5 PTSD as assessed by the Clinician-Administered PTSD Scale for DSM-5.

The Institutional review Board of McLean Hospital approved all studies. All saliva samples were collected using Oragene DNA collection kits (DNA Genotek) according to the instructions of the manufacturer.

**Mexican Adolescent Mental Health Study (MAMH; Supplementary Table 1 #65)**

See reference for details.^123,124^ Potentially traumatic events were identified using the PTSD section of the WHO-WMH-CIDI-A.^125^ The WHO-WMH-CIDI-A was used to assess PTSD over the prior lifetime and 12-months by trained lay interviewers.^126,127^ Respondents were considered to have a lifetime diagnosis if DSM-IV criteria were met during lifetime. Respondents were considered to have a current diagnosis if DSM-IV criteria were met in the prior 12 months. The WHO-WMH-CIDI-A calculates PTSD symptom severity, which ranged from 0 to 17, by counting number of symptoms endorsed. For this cohort (70 Cases, 1079 Controls), the mean severity was 12.86 and the standard deviation 2.53. DNA for GWAS analysis was isolated from saliva. The Institutional Review Board of the National Institute of Psychiatry Ramón de la Fuente Muñiz approved this study.

**Mid-Atlantic Mental Illness Research Education and Clinical Center the study of Post-Deployment Mental Health Study (MIRE; Supplementary Table 1 #26)**

See reference for details of the study of Post-Deployment Mental Health (1,308 cases, 1,914 controls).^128,129^ PTSD was diagnosed using the Structured Clinical Interview for DSM-IV Disorders (SCID) administered by trained interviewers.^13^ In accordance with the DSM-IV, PTSD consisted of three symptom clusters. These included re-experiencing symptoms (B symptoms), avoidance and numbing symptoms (C symptoms) and hyperarousal symptoms (D symptoms). Total PTSD symptoms and symptom clusters (B, C, or D) were measured using the Davidson Trauma Scale for all veterans, including individuals with current PTSD diagnosis and controls. The research was reviewed and approved by the Institutional Review Boards at the Salisbury, NC VA, Hampton, VA VA, Richmond, VAVA, Durham, NC VA and Duke University Medical Centers.

**The Million Veteran Program (MVP1; Supplementary Table 1 #79)**

See reference for details.^130^ Likely PTSD diagnoses were identified using Lasso regression with cross-validation based on manual chart review.^131^ The PCL17 for DSM-IV was used to assess PTSD symptoms over the past month by self-report. Respondents were considered to have a diagnosis if passed the Lasso regression with cross-validation for Electronic Health Records (EHR). The PCL-IV calculates PTSD symptom severity, which ranged from 17 to 85, by sum of scores on a five-point severity scale. For this cohort, (38,346 Cases, 173,661 Controls). DNA for GWAS (MVP 1.0 custom Axiom array) analysis was isolated from blood. The VA Central Institutional Review Board (IRB), as well as VA IRBs in Boston, San Diego, and West Haven, approved this study.

**Mount Sinai BioMe PTSD (BIOM; Supplementary Table 1 #92)**

PTSD cases and controls were ascertained using EHR data from n=52,641 patients from the Mount Sinai BioMe^TM^.^132^ Patients included in this sample were not screened for any floor criteria. PTSD case and control definitions were based on ICD-10 codes per the Freeze 3 established definitions (see Supplementary Methods). Due to limited case numbers using the narrow definition (n=393), only the broad definition was examined. DNA for GWAS analysis was isolated from blood.

**National Centre for Mental Health (NCMH; Supplementary Table 1 #41)**

See www.ncmh.info for details. Potentially traumatic events were identified by participant self-report. The Trauma Screening Questionnaire (TSQ) was used to assess PTSD over the prior 2 weeks by self-report.^133^ Respondents were considered to have a current diagnosis if a score of 6 or more was obtained. The TSQ calculates PTSD symptom severity, which ranged from 0 to 10, by self-report. For this cohort (631 Cases, 653 Controls), the mean severity was 5.32 and the standard deviation 3.47. DNA for GWAS analysis was isolated from blood or saliva. The study was given a favorable ethical opinion by Wales Research Ethics Committee (REC) 2.

**National Health and Resilience in Veterans Study (NHRV; Supplementary Table 1 #13)**

See reference for details.^134^ Potentially traumatic events were identified using the Trauma History Screen.^135^ The PTSD Checklist-Specific (PCL-S) was used to assess both lifetime and past-month PTSD symptoms related to respondent’s ‘worst’ traumatic event over their lifetimes by survey.^2^ Respondents were considered to have screen positive for PTSD if their PCL-S score was ≥50. The PCL-S calculates PTSD symptom severity, which ranged from 17 to 85, by self-report. For this cohort (95 Cases, 1490 Controls), the mean PCL-S severity score was 26.9 and the standard deviation 11.3. DNA for GWAS analysis was isolated from saliva. The Human Subjects Subcommittee of VA Connecticut Healthcare System approved this study.

**NIU Trauma Study (NIUT; Supplementary Table 1 #40)**

See reference for details.^136,137^ Potentially traumatic events were identified using the Traumatic Life Events Questionnaire and 12 questions regarding level of exposure to the campus shooting.^138^ The Distressing Events Questionnaire was used to assess PTSD immediately following the mass shooting (average 3.2 weeks) by self-report.^139^ Respondents were considered to have a diagnosis if their DEQ score was ≥ 18. The DEQ calculates PTSD symptom severity, which ranged from 0 to 66, by self-report. For this cohort (280 Cases, 411 Controls), the mean severity was 16.49 and the standard deviation 12.35. DNA for GWAS analysis was obtained from 204 of the PTSD cases and was isolated from saliva. The Institutional Review Board of Northern Illinois University approved this study.

**The Nord-Trøndelag Health Study (HUNT; Supplementary Table 1 #88)**

See reference for details.^140^ Cases with PTSD were defined by participants meeting at least one of the following two criteria: 1) In the time period 1987 through 2017 at least one hospital contact due to ICD-9 diagnosis *Acute reaction to stress from extreme external events* (308) or *Adjustment reaction* (309), or due to ICD-10 diagnosis *Reaction to severe stress and adjustment disorders* (F43) or *Enduring personality change after catastrophic experience* (F62.0). 2) At the time of participation in the HUNT study past year exposure to imminent mortal danger, and scoring >8 for the Hospital Anxiety and Depression Scale (HADS) for either anxiety (HADS-A) or depression (HADS-D).^141^ About one third (27%) of those exposed to imminent danger during the last year reported HADS-A or HADS-D > 8. The study was approved by the Regional Committee for Medical and Health Research Ethics (ref. 2015/575).

**Nurses Health Study II (NHS2; Supplementary Table 1 #5)**

See reference for details.^142^ Participants identified stressful events they had experienced from a list of 25 events used in diagnostic interviews, ^143, 40,144-146^ and PTSD was assessed in relation to participants’ self-selected worst stressful event. Participants were cued to think of the period following the event during which symptoms were most frequent and intense. They were asked whether they had ever been bothered by each of 17 symptoms and rated each symptom on a Likert-style scale (1: “not at all” to “5: extremely”).^147^ Additional questions assessed the other three DSM-IV criteria: intense fear, horror, or helplessness in response to the event (Criterion A2), symptom duration of at least one month (Criterion E), and clinically significant impairment in functioning due to symptoms (Criterion F)^143^. Based on the diagnostic interview, we created two lifetime PTSD phenotypes as follows.

To meet criteria for lifetime PTSD diagnosis, respondents must have endorsed experiencing one or more of the 5 re-experiencing symptoms, 3 or more of the 7 avoidance/numbing symptoms, 2 or more of the 5 arousal symptoms, and criteria A2, E and F as defined above. In addition to the diagnostic phenotype, we analyzed lifetime PTSD symptom severity which was defined as the sum of the symptom ratings across the 17 questions.

The reliability of the PTSD diagnosis was assessed using a blind review of audiotapes from 50 interviews and the Cohen’s kappa statistic was 1.0 (perfect reliability).^148^ We assessed the validity of our identification of PTSD in a separate cohort, the Detroit Neighborhood Health Study, via clinical interviews among a random subsample of 51 participants and found excellent concordance.^63^ The Partners Human Research Committee approved this study.

**Nurses Health Study II (NHSY; Supplementary Table 1 #22)**

See reference for details.^149^ The Nurses’ Health Study II is an ongoing cohort of 116,430 female nurses initially enrolled in 1989 and followed with biennial questionnaires. The present study included follow-up through 2013. This study included women who returned a supplementary 2008 questionnaire on trauma exposure and PTSD symptoms (*N*=54,763). This questionnaire was sent to a subsample of participants (*N*=60,804, response rate=90.1%). To retain participation in the ongoing longitudinal cohort, only women who have already returned their biennial questionnaire are sent supplementary questionnaires. Women missing data on trauma or PTSD symptoms (*N*=3,930) were excluded. This study was approved by the Institutional Review Board of Brigham and Women’s Hospital. Return of the questionnaire via US mail constitutes implied consent.

Trauma exposure and PTSD symptoms were assessed on a supplementary 2008 questionnaire. The 16-item Brief Trauma Questionnaire queried lifetime exposure to 15 types of traumatic events (e.g., serious car accident, sexual assault) and an additional item queried any traumatic event not covered in the other questions. ^103^ Respondents were asked to identify which trauma was their worst or most distressing; they were then asked their age at this worst trauma as well as their age at their first trauma. PTSD symptoms were assessed in relation to their worst trauma with the 7-item Short Screening Scale for DSM-IV PTSD.^104^ Four or more symptoms on this scale have been associated with PTSD diagnosis (sensitivity=80%, specificity=97%, positive predictive value=71%, negative predictive value=98%).^104^ For cases women with ≥4 PTSD symptoms were selected, for controls women with <4 PTSD symptoms (most had 0) were selected.

**Ohio National Guard (ONGA; Supplementary Table 1 #2)**

See reference for details.^150^ A total of 37 potentially traumatic events were identified using the Clinician-Administered PTSD Scale (CAPS-IV)^12^ and the 1996 Detroit Area Survey of Trauma.^64^ PTSD symptoms were assessed using a 17-item structured interview scale derived from the PTSD Checklist (PCL) for DSM-IV^2^ performed by trained lay telephone interviewers using epidemiological methods (forced choice symptom severity range, 1-5). Reliability of the telephone interview was validated against the criterion standard (in-person CAPS interview by mental health professional) in a clinical subsample (N = 500), demonstrating high specificity (0.92).^151^ Respondents were considered to have a diagnosis if lifetime DSM-IV PTSD criteria were met. Respondents were considered to have a current diagnosis if past month DSM-IV criteria were met. The PCL calculates PTSD symptom severity, which ranged from 17 to 85, by sum of scores of items endorsed. For this cohort (125 Cases, 125 Controls), the mean severity was 38.4 and the standard deviation 17.6. DNA for GWAS analysis was isolated from saliva (Oragene tube). The Institutional Review Board of VA Ann Arbor Health System approved this study.

**Ohio National Guard 2 – New Samples (ONGB; Supplementary Table 1 #76)**

See reference for details.^150^ A total of 37 potentially traumatic events were identified using events from the 1996 Detroit Area Survey of Trauma,^64^ the Life Events Checklist-Civilian Version,^9^ and the Deployment, Risk and Resilience Survey (DRRI).^152^ PTSD symptoms were assessed using a 17-item structured interview scale derived from the PTSD Checklist (PCL) for DSM-IV performed by trained lay telephone interviewers using epidemiological methods (symptom severity range, 1-5).^65^ Reliability of the telephone interview was validated against the criterion standard (in-person CAPS interview by mental health professional) in a clinical subsample (n = 500), demonstrating high specificity (0.92).^151^ Respondents were considered to have a diagnosis if lifetime DSM-IV PTSD criteria were met. Respondents were considered to have a current diagnosis if past month DSM-IV criteria were met. The PCL calculates PTSD symptom severity, which ranged from 17 to 85, by sum of scores of items endorsed. DNA for GWAS analysis was isolated from saliva (Oragene tube). The Institutional Review Board of VA Ann Arbor Health System approved this study.

**OPT (FEEN; Supplementary Table 1 #37)**

Potentially traumatic events were identified using the standard trauma interview.^42^ The PSS-I was used to assess PTSD over the prior two weeks for the trauma of interest by postdoctoral and graduate level assessors trained to reliability.^43^ The SCID-IV was used to assess lifetime PTSD (not current) for a single trauma not the focus of treatment.^13^ Respondents were considered to have a current PTSD diagnosis if they met DSM-IV diagnostic criteria and had a PSS-I score of 25 or greater. The PSS-I also provides PTSD symptom severity, with a range from 0 to 51. For this cohort (118 cases), the mean severity was 30.36 and the standard deviation 6.91. DNA for GWAS analysis was isolated from blood. The Institutional Review Board of University Hospitals approved this study.

**Portugal (PORT; Supplementary Table 1 #20)**

See reference for details.^153^ Potentially traumatic events were identified using CIDI, which included a module on DSM-IV PTSD that inquired about lifetime exposure to each of 27 different traumatic events (criterion A1). Respondents who reported ever experiencing any of the traumatic events were then asked about the number of exposures (NOE) and age at first exposure (AOE) for each.^143^ The CIDI was used to assess PTSD over the prior Lifetime DSM-IV PTSD and other common DSM-IV disorders.^143^ DNA isolated from saliva. The Institutional Review Board of the Nova Medical School, Portugal, approved this study.

**PRISMO (PRIS; Supplementary Table 1 #25)**

See reference for details.^154^ Potentially traumatic events were identified using a 19-item checklist.^154^ The Dutch Self-Rating Inventory for PTSD (SRIP) was used to assess PTSD one month prior to deployment and up to 2 years after deployment.^155^ Respondents were considered to have a current diagnosis if a SRIP total score of ≥ 38 at any measurement was met. The SRIP calculates PTSD symptom severity, which ranged from 22 to 88, by self-report. For this cohort (144 Cases, 815 Controls), the mean severity was 27.30 and the standard deviation 6.38. DNA for GWAS analysis was isolated from whole blood. The Institutional Review Board of Utrecht University Medical Center approved this study.

**Pregnancy Outcomes, Maternal and Infant Study Cohort (PROM; Supplementary Table 1 #23)**

See reference for details.^156^ Potentially traumatic events were experiences of intimate partner violence [identified using Demographic Health Survey Questionnaires and Modules: Domestic Violence Module^157^ and the World Health Organization Multi-Country Study on Violence Against Women]^158^ and history of childhood abuse [identified using the Childhood Physical and Sexual Abuse Questionnaire adopted from the Center for Disease Control and Prevention Adverse Childhood Experiences Study].^159^ The Posttraumatic Stress Disorder Checklist – Civilian Version (PCL-C) was used to assess PTSD over the prior month.^160^ The PCL-C is a self-report measure with 17 items reflecting DSM-IV symptoms of PTSD and closely follows the Diagnostic and Statistical Manual of Mental Disorders (DSM-IV) criteria. For each item, participants were asked how bothered they were by a symptom over the past month on a 5-point Likert scale ranging from 1: “not at all” to 5: “extremely” in regards to their most significant life event stressor. The total score on the PCL-C ranges from 17 to 85. Recent data from our team support that a PCL-C score of 26 or higher on the Spanish-language version, is associated with an 86% sensitivity and 63% specificity in diagnosing PTSD in a Peruvian population using the Diagnostic and Statistical Manual of Mental Disorders (DSM-IV) criteria.^160^

All study procedures were approved by the Instituto Nacional Materno Perinatal in Lima, Peru, and the Office of Human Research Administration at the Harvard T.H. Chan School of Public Health, Boston, MA.

**QIMR Berghofer Medical Research Institute (QIMRB) (AGDS, QIM2; Supplementary Table 1 #80, #82)**

The QIMRB cohort contributed with four studies: Australian Genetics of Depression Study (AGDS)^161^, Prospective Imaging Study of Ageing (PISA)^162^, Genetics of Human Agency (GHA)^163^, and Twenty-Five and Up Study (25Up).^164^ All studies were based on self-report.

The four studies included a screening question for PTSD (e.g. “Have you ever been diagnosed or treated for PTSD by a health care professional?”), leading to 2,218 cases and 18,449 controls.

Additionally, AGDS and PISA included a section in the questionnaire on PTSD. Potentially traumatic events were identified using the Life Events Checklist for DSM-5 (LEC-5; Weathers et al, 2013).^92^ Exposure to trauma was considered when for any given event, participants responded “Happened to me personally”, “Witnessed it happen to someone else”, “Learned about it happening to a close family member or close friend”, or “Part of my job”. Three questions adapted from the DSM-5 were used to identify whether participants had PTSD symptoms in 13,046 participants who reported exposure to trauma. A sum score of the presence of these symptoms was computed (mean=1.74, SD=1.2, ranging from 0 to 3).

DNA for GWAS analysis was isolated from blood or saliva. The Human Research Committee of QIMR Berghofer Medical Research Institute approved this study. There are no duplicate individuals within phenotypes. However, there is overlap between the two phenotypes. There are 20,828 participants are unique across phenotypes. Participants are of European ancestry and they exclude the QIMR contribution to Nievergelt et al.

**Readiness and Resilience in National Guard Soldiers (RING; Supplementary Table 1 #33)**

See reference for details.^165^ Potentially traumatic events were identified using the Clinician-Administered PTSD Scale for DSM-IV (CAPS-IV) and items from the Deployment Risk and Resilience Inventory (DRRI)^12,62^ The CAPS-IV and the PTSD Checklist (PCL) were also used to assess PTSD currently (over the prior 2 weeks) and over the lifetime by trained interviewers and through self-report.^12,166^ The PCL calculates PTSD symptom severity, which ranged from 17 to 73 currently and 17 to 85 lifetime, by self-report. For this cohort (41 Cases, 162 Controls), the mean current severity was 34.03 and the standard deviation was 13.48. The mean lifetime severity was 33.89 and the standard deviation was 12.98. Respondents were considered to have a lifetime diagnosis if they had a PCL score ≥ 50 at the one-year post-deployment time point. Respondents were considered to have a current diagnosis if the same criteria were met at the two-year post-deployment time point. The Institutional Review Board of the Minneapolis VA Health Care System approved this study.

**Risbrough/Norman randomized controlled psychotherapy trial (VRIS; Supplementary Table 1 #58)**

See reference for details.^167^ Potentially traumatic events were identified using the life event checklist. The CAPS was used to assess PTSD over the prior over the past month by interviewers.^94^ Respondents were considered to have a diagnosis if DSM-5 diagnostic criteria using the CAPS-5 (a criterion A trauma and 2 or higher on 1 re-experiencing symptom (criterion b), 1 avoidance symptom (criterion c), 2 negative alterations in cognitions or mood (criterion d), and 2 hyperarousal symptoms (criterion e)) were met. The CAPS calculates PTSD symptom severity, which ranged from 0 to 80 by clinician rater of symptoms and severity. For this cohort (73 Cases, 5 sub-clinical), the mean severity was 42 and the standard deviation 9.6. DNA for GWAS analysis was isolated from saliva using Oragene. The Institutional Review Board of The San Diego VA approved this study.

**SAS (DSAS; Supplementary Table 1 #69)**

See reference for details.^168^ Potentially traumatic events were identified using the Childhood Trauma Questionnaire.^29^ The Diagnostic Interview Schedule for Children (DISC-IV) was used to assess PTSD over the lifetime and past year by interviewers.^169^ Respondents were considered to have a diagnosis if DSM-IV criteria were met. The DISC-IV calculates PTSD symptom severity, which ranged from 0 to 13, by summing intensity of PTSD symptoms. For this cohort (25 Cases, 146 Controls), the mean severity was 1.62 and the standard deviation 3.14. DNA for GWAS analysis was isolated from saliva. The Institutional Review Board of University of Washington approved this study.

**Shared Roots (SHRS; Supplementary Table 1 #46)**

Potentially traumatic events were identified using The Life Events Checklist for DSM-5 (LEC-5).^92^ The Clinician-Administered PTSD Scale for DSM-5 (CAPS-5)^94^ was administered by clinicians to assess PTSD over the prior month. The CAPS-5 and the PTSD Checklist for DSM-5 (PCL-5)^73^ calculates PTSD symptom severity, with a score range of 0 to 80, by adding scores ranging from 0 to 4 for all twenty items. For this cohort (164 Cases, 164 Controls), the mean severity on the PCL-5 was 33.0 and the standard deviation 23.9. A lifetime diagnosis of PTSD was not assessed for in this study. Respondents were considered to have a current diagnosis if DSM-5^170^ criteria based on the CAPS-5 were met. The Institutional Review Board of Stellenbosch University approved this study.

**Shared Roots 2 (SHR2; Supplementary Table 1 #75)**

Potentially traumatic events were identified using The Life Events Checklist for DSM-5 (LEC-5).^92^ The clinician-administered PTSD scale for DSM-5 (CAPS-5)^94^ was used to assess PTSD over the prior month by clinicians. The CAPS-5 and the PTSD Checklist for DSM-5 (PCL-5)^73^ calculates PTSD symptom severity, which ranged from 0 to 80, by adding scores ranging from 0 to 4 for all twenty items. For this cohort (107 Cases, 231 Controls), the mean severity on the PCL-5 was 26.6 and the standard deviation 22.0. A lifetime diagnosis of PTSD was not assessed for in this study. Respondents were considered to have a current diagnosis if DSM-5^170^ criteria based on the CAPS-5 were met. The Institutional Review Board of Stellenbosch University approved this study.

**Southeastern Europe PTSD (SEEP; Supplementary Table 1 #49)**

See reference for details.^171^ Potentially traumatic events were identified using Life Stressor List and List of traumatic events including frequency and severity of traumatic events.^172^ The CAPS was used to assess lifetime PTSD by medical personnel (psychiatrists, psychologists or psychiatric residents).^12^ Respondents were considered to have a diagnosis if DSM-IV criteria were met over lifetime. The CAPS calculates PTSD symptom severity, which ranged from 27 to 141. For this cohort (347 Cases, 339 Controls), the mean severity was 74.3 and the standard deviation 20.2. DNA for GWAS analysis was isolated from EDTA blood. The Institutional Review Board of the universities of Sarajevo, Zagreb, Tuzla, Mostar, Prishtina and Würzburg approved this study.

**Stony Brook University World Trade Center Health Program (WTCS; Supplementary Table 1 #78)**

See reference for details.^173^ Trauma - 9/11 exposure severity - was assessed by a clinician-administered interview at the first monitoring visit, and operationalized dimensionally using an established index composed of the total time spent working at the WTC site, exposure to the cloud of debris from the collapse of the WTC buildings, and work on the pile of debris.^174^ The PTSD Checklist (PCL) - Specific Version^2^ was used to assess WTC-related DSM-IV PTSD symptoms over the prior 1 month by self-report questionnaires. The PCL calculates PTSD symptom severity, which ranged from 17 to 85. For this cohort, the mean severity was 28.67 and the standard deviation 12.61. The Structured Clinical Interview for DSM-IV (SCID)^175^ was administered to 59.8% of the total sample. A lifetime prevalence rate of WTC-related PTSD was 23.8%. DNA for GWAS analysis was isolated from blood samples. The Institutional Review Board of Stony Brook University approved this study.

**Stress Risk and Resilience in Syrian and Iraqi Refugees and Survivors of Torture (SYIR; Supplementary Table 1 #63)**

See reference for details.^176^ Potentially traumatic events were identified using LEC was used a year later, data is available but not included in this report.^5^ The PTSD Checklist for Civilians, DSM IV was used to assess PTSD over the prior lifetime by bilingual clinicians and research assistants.^2^ Respondents were considered to have a diagnosis if Criteria B, C, and D as defined by the scoring of the PTSD Checklist for Civilians, DSM-IV were met. Determining whether an individual meets DSM-IV symptom criteria is defined by at least 1 B item (questions 1-5), 3 C items (questions 6-12), and 2 D items (questions 13-17) being present—that is, the response is 3-5. The PCL calculates PTSD symptom severity, which ranged from 17 to 85, by summing the scores from each of the 17 items that have response options ranging from 1 “Not at all” to 5 “Extremely”. For this cohort (59 cases; 135 controls) the mean severity was 55.875 for cases; 31.41964 for controls; 38.75625 for the total cohort and the standard deviation was 12.62659 for cases; 10.36421 for controls; 15.76388 for the total cohort. DNA for GWAS analysis was isolated from saliva. The Institutional Review Board of Wayne State University approved this study.

**Study of Aftereffects of Trauma: Understanding Response in National Guard (SATU; Supplementary Table 1 #11)**

See reference for details.^177^ Potentially traumatic events were identified using Clinician-Administered PTSD Scale for DSM-IV (CAPS-IV).^12^ The CAPS-IV was also used to assess PTSD currently (over the prior 2 weeks) and over the lifetime by trained interviewers^12^. The CAPS-IV calculates PTSD symptom severity, which ranged from 0 to 105. For this cohort (88 Cases, 62 Controls), the mean severity was 50.33 and the standard deviation 24.55. Respondents were considered to have a lifetime diagnosis if they met DSM-IV criteria for PTSD (measured using CAPS-IV) either current or past. Respondents were considered to have a current diagnosis if they met DSM-IV criteria for PTSD (measured using CAPS-IV) at the time of the assessment. The Institutional Review Board of the Minneapolis VA Health Care System approved this study.

**The Study of Twin Adults: Genes and Environment (STR-STAGE), from the Swedish Twin Registry (SWED; Supplementary Table 1 #89)**

See reference for details.^178^ We identified individuals with PTSD and stress or adjustment disorders from the national outpatient or inpatient register, using corresponding diagnosis codes in the International Classification of Diseases (ICD) version 9 or 10.^179^ Broad PTSD definition considered those with at least one specialist inpatient or outpatient treatment contacts for PTSD (ICD-9: 309.81; ICD-10: F34.1) and/or one contact for stress or adjustment disorder (ICD-9: 309; ICD-10: F43.0, F43.2, F43.8, F43.9) as cases, and included all other participants as controls. Narrow definition considered those with at least two specialist treatment contacts for PTSD as cases, while excluding those with only one diagnosis of PTSD, post-concussive syndrome (ICD-10: F07.81), and those with other stress disorders from controls. In this study, the number of cases in the narrow definition is too few (N<100) to warrant a separate GWAS. We, therefore, conducted a GWAS using the broad PTSD definition (330 cases and 9,347 controls). DNA for GWAS analysis was isolated from saliva samples. The Regional Ethics Review Board in Stockholm approved this study (Dnr 2011/570-31/5, 2012/1107-32, 2018/960-31/ 2).

**Study on Trauma & Resilience (STAR Study) (RCSS; Supplementary Table 1 #70)**

See reference for details.^180^ Potentially traumatic events were identified using the Life Events Checklist at baseline (during index hospitalization) and then again at the 6-month follow-up to assess for new traumatic events since the participant’s index hospitalization for traumatic injury.^92^

The PCL-5 was used to assess PTSD symptoms at study baseline for the participant’s traumatic injury, and at study follow-up 6 months post-date of trauma.^170,181^ It was administered at each study timepoint by trained study interviewers. Study interviewers were at a minimum at the undergraduate level supervised by clinical psychologists. However, most personnel were clinical psychology graduates and fellows.

The CAPS-5 assessed PTSD diagnosis and symptom severity at the 6-month follow-up by similarly trained interviewers.^182^ All respondents met Criterion A due to their traumatic injury requiring hospitalization at a Level 1 Trauma Center. Respondents were considered to have a current PTSD diagnosis if they scored positive via the CAPS-5. Both the PCL-5 and CAPS-5 calculate PTSD symptom severity, which ranged from 0 to 80, by summative calculation.

For this cohort (50 Cases, 222 Controls), at baseline the mean severity via the PCL-5 was 19.57 and the standard deviation 17.37. For this cohort (48 Cases, 102 Controls) at 6-month follow-up, the mean severity via CAPS-5 was 13.55 and the standard deviation was 15.21. Also at 6-month follow-up, the mean severity via the PCL-5 was 20.33 and the standard deviation was 20.24. DNA for GWAS analysis was isolated from blood serum. The Institutional Review Board of the Medical College of Wisconsin approved this study.

**Sydney Neuroimaging (BRY2; Supplementary Table 1 #36)**

Potentially traumatic events were identified using clinical interview to identify history of traumatic events. The CAPS was used to assess PTSD over the prior 4 weeks by Masters level clinical interviewers.^12^ Respondents were considered to have a diagnosis if DSM-IV criteria were met. The CAPS calculates PTSD symptom severity, which ranged from 0 to 136, by clinical interview. For this cohort (82 Cases, 86 Controls), the mean severity was 39.67 and the standard deviation 31.75. DNA for GWAS analysis was isolated from saliva. The Institutional Review Board of Western Sydney Area Health Service approved this study.

**UK Biobank Cohort Description for PGC-PTSD (UKBB; Supplementary Table 1 #60)**

The UK Biobank is an epidemiological resource assessing a range of health-related phenotypes in approximately 500,000 British individuals who were recruited between the ages of 40 and 70.^183^ Genome-wide genotype data is available on all participants, as well as a broad range of health phenotypes assessed at varying intensity. Data from an online follow-up questionnaire assessing common mental health traits, including questions designed to screen for PTSD, was available on 157,366 individuals.^184^

*Phenotype:* PTSD phenotypes were derived from the mental health online follow-up of the UK Biobank (Resource 22 on http://biobank.ctsu.ox.ac.uk). Participants were asked six questions derived from the brief civilian version of the PTSD Checklist Screener (PCL-S;^110^) assessing PTSD symptoms in the prior month. Questions comprised three initial questions related to avoidance of activities, disturbing thoughts, and feeling upset, and three further questions related to feeling distant, feeling irritable and having trouble concentrating (UK Biobank fields 20494-20498, 20508). Each item was scored on a five-point scale according to the amount of concern caused by that item in the past month (1="Not at all" to 5="Extremely"). The final item concerning trouble concentrating was drawn from an equivalent item from the Patient Health Questionnaire-9 (PHQ9) depression questionnaire and was scored on a four-point scale according to frequency of difficulties (1="Not at all" and 4="Nearly every day"). All items were summed for each individual to yield a total score ranging 3-29. Cases were defined as all individuals with a PCL-S score ≥ 13, controls as all individuals who responded to all of the initial three questions and had PCL-S score ≤ 12.

Two sensitivity analyses were performed to assess the stability of the UK Biobank PTSD phenotype. Firstly, controls were limited only to those who answered all initial questions with "Not at all" and so had a PCL-S score = 3. Secondly, PTSD cases and controls were limited to those reporting a lifetime trauma exposure. To assess the impact of reported trauma exposure on the PTSD phenotype, a trauma exposure measure was derived from questions in the mental health online follow-up that related to common triggers of post-traumatic stress-disorder.^184^ These questions asked if participants had ever: experienced combat; had a life-threatening accident; been diagnosed with a life-threatening illness; been a victim of a physically violent crime; been a victim of sexual assault; or witnessed a sudden violent death. Responses were combined to a single variable capturing any report of traumatic experience versus no report.

*Genetic quality control:* Genetic data for analyses was obtained from the full release of the UK Biobank data (N=487,410).^185^ Individuals were removed if this was recommended by the UK Biobank for unusual levels of missingness or heterozygosity; if call rate < 98% on genotyped SNPs; if they were related to another individual in the dataset (KING r < 0.044, equivalent to removing up to third-degree relatives inclusive); and if the phenotypic and genotypic gender information was discordant (X-chromosome homozygosity (FX) < 0.9 for phenotypic males, FX > 0.5 for phenotypic females). Removal of relatives was performed using a greedy algorithm, which minimise exclusions (for example, by excluding the child in a mother-father-child trio). All analyses were limited to individuals of White Western European ancestry, as defined by 4-means clustering on the first two genetic principal components provided by the UK Biobank.^186^ Principal components analysis was also performed on the European-only subset of the data using the software package flashpca2^187^. After quality control, individuals were excluded from analysis if they did not complete the mental health online questionnaire (N=126,522).

Genetic analyses used imputed variants provided by the UK Biobank.^185^ Autosomal genotype data from two highly-overlapping custom genotyping arrays (covering ~800,000 markers) underwent centralised quality control to remove genotyping errors before being imputed in a two-stage imputation to the Haplotype Reference Consortium (HRC) and UK10K (for rarer variants not present in the HRC) reference panels ^(188;189;185)^. In addition, this central quality control, variants for analysis were limited to common variants (minor allele frequency > 0.01) imputed with higher confidence (IMPUTE INFO metric > 0.4). In addition, only variants that were directly genotyped or that were imputed from the HRC were included.^188^

Genome-wide association analyses: Prior to analysis, PTSD status was residualised on six prinicipal components from the genetic data and factors capturing genotyping batch and recruitment centre, using logistic regression. GWAS were then performed on the resulting deviance residuals using linear regressions on imputed genotype dosages in BGenie v1.2, software written for genetic analyses of UK Biobank.^185^

**UK Biobank – EHR Cohort Description (UKB2; Supplementary Table 1 #91)**

PTSD cases and controls were ascertained using primary healthcare data from (n=130 744) patients from the UK Biobank. See reference for details.^190^ Patients included in this sample were not subject to a minimum data floor criterion, requiring only a single reported diagnosis of PTSD or of another stress-related disorder. Case and control definitions were based on Reed codes matched to ICD-10 codes, per the Freeze 3 established "broad" definition (see Supplementary Methods). Due to limited case numbers using the narrow definition (n=81), only the broad definition was examined. Details of the genotyping of this cohort can be found in reference.^191^ Research on the UK Biobank is approved under a generic Research Tissue Bank Approval from the UK North West Multi-centre Research Ethics Committee. The analyses in this paper were conducted as part of project 18177.

**VA Boston-National Center for PTSD Study (NCPT, TRAC; Supplementary Table 1 #31, Supplementary Table 1 #32)**

A total of 437 white non-Hispanic cases and 215 trauma-exposed controls is the composite of two datasets. The first from a cohort of white non-Hispanic subjects as described in a previous GWAS^192^ that passed ancestry filters as performed using SNPweights^193^ according to PGC-PTSD protocols (300 cases and 165 controls; 305 males and 160 females). The majority of this sample consisted of US veterans, but also included the intimate partners of a subset of the veterans. The second cohort was made up of subjects from the Translational Research Center for TBI and Stress Disorders, a VA RR&D Traumatic Brain Injury Center of Excellence at VA Boston Healthcare System (TRACTS) study of US veterans. From TRACTS, 137 white non-Hispanic cases and 50 controls passed ancestry filters based on SNPweights and were included in the analysis. The TRACTS sample is largely male (170 men and 17 women). The genotyping, quality control, filtering and imputation for these cohorts has been described in detail elsewhere.^192,194^ Briefly, genotyping was performed using the Illumina HumanOmni2.5-8 microarrays (Illumina, San Diego, CA). Imputation of non-genotyped SNPs was performed using IMPUTE2^195-198^ and 1000 genomes phase 1 reference data.^199^ Principal components were generated by the program EIGENSTRAT^200^ based on 100,000 SNPs. For both cohorts, participants were administered the Clinician-administered PTSD scale for DSM-IV (CAPS-IV),^12^ a 30-item structured diagnostic interview that assesses the frequency and severity of the 17 DSM-IV PTSD symptoms, 5 associated features and functional impairment, and both current and lifetime PTSD symptoms. These studies were performed under the oversight of the appropriate VA health care facilities institutional review boards.

**Vietnam Era Twin Study of Aging (VETS; Supplementary Table 1 #24)**

See references for details.^201,202^ Potentially traumatic events were identified using the Combat Exposure Index^203^ and the Diagnostic Interview Schedule Version III-Revised (DIS-III-R).^DIS-III-R; 204^ The Vietnam Era Twin Registry PTSD scale^205^ (administered at average age 38) was used to assess PTSD symptoms over the past 6 months and the PCL-civilian version for DSM-IV^206^ (administered at average age 62) was used to assess PTSD over the prior month. These two instruments correlate 0.90 when administered at the same time.^207^ The 17-item PCL calculated PTSD symptom severity, which ranged from 17 to 84. Each response was rated on a 1-5 scale (from “not at all” to “extremely”). For this cohort (60 Cases, 841 Controls), the mean severity was 26.2 and the standard deviation was10.5. For this analysis, respondents were considered to have a diagnosis of PTSD if they met DSM-III-R criteria based on the DIS-II-R interview. DNA for GWAS analysis was isolated from blood. Genotyping was performed by deCODE Genetics, Reykjavik, Iceland. The Institutional Review Boards of the University of California, Sand Diego, Boston University, and the Puget Sound VA Healthcare System approved this study.

**The Women and Children’s Health Study (WACH; Supplementary Table 1 #43)**

See reference for details.^208^ Potentially traumatic events were identified using the Life Events Checklist (LEC) for DSM-5.^92^ The PTSD Checklist for DSM-5 (PCL-5) was used to assess PTSD over the lifetime by interviewers.^181^ Respondents were considered to have a diagnosis of PTSD if the PCL-5 score was >=38. Respondents were considered to have a current diagnosis based only upon self-report during the interview. The PCL-5 calculates PTSD symptom severity, which ranged from 0 to 79. For this cohort (151 Cases, 150 Controls), the mean severity was 52.4 (SD: 11.2) for cases and 31.0 (SD: 23.5) for controls DNA for GWAS analysis was isolated from blood. The Institutional Review Board of the Louisiana State University Health Sciences Center-New Orleans approved this study.

**World Trade Center: Mount Sinai (WTCM; Supplementary Table 1 #84)**

Potentially traumatic events experienced during the 9/11 attacks on the World Trade Center (WTC) and/or during rescue, recovery, and clean-up work following the WTC attacks (e.g., exposure to human remains, received treatment for an illness or injury during WTC recovery work) were identified during interviews and on self-report questionnaires; these were completed by participants on their first health-monitoring visit to the WTC Health Program, an average of 4.3 (SD=2.7) years following 9/11/2001. The Clinician-Administered PTSD Scale (CAPS) was used to assess lifetime highest WTC-related PTSD., administered by trained clinicians during in-person clinical assessments conducted an average of 13.5 (SD=1.3) years following 9/11/2001. Respondents were considered to have a lifetime PTSD diagnosis if they met DSM-IV criteria for lifetime WTC-related PTSD and a total score ≥ 40 on the lifetime CAPS. The total lifetime CAPS score, used to calculate PTSD symptom severity, ranged from 0-119 in the full cohort. This analysis includes three ancestry-specific cohorts; case/control numbers, and mean, range of CAPS are shown in the table below.

DNA for GWAS analysis was isolated from whole blood. The Institutional Review Board of the Icahn School of Medicine at Mount Sinai approved this study.

|  | **CAPS Lifetime** | | | **Sample Numbers** | |
| --- | --- | --- | --- | --- | --- |
| **IDS** | **Range** | **Mean** | **SD** | **Cases** | **Controls** |
| W | 0-119 | 34.726 | 34.31 | 62 | 156 |
| HA | 0-117 | 28.69 | 33.66 | 23 | 49 |
| AA | 0-117 | 35.3 | 33.5 | 15 | 47 |

**Yale-Penn Study (GSDC; Supplementary Table 1 #6)**

See reference for details.^209^ Sample collection and diagnostic interviews were performed by trained interviewers using the Semi-Structured Assessment for Drug Dependence and Alcoholism (SSADDA; available at <https://zork.wustl.edu/nida/study_descriptions/study_1/ssaddav11_2_ns.pdf>) to derive diagnoses for lifetime psychiatric and substance use disorders based on DSM-IV criteria. Twelve types of traumatic events were assessed: experienced direct combat in a war; seriously physically attacked or assaulted; physically abused as a child; seriously neglected as a child; raped; sexually molested or assaulted; threatened with a weapon; held captive or kidnapped; witnessed someone being badly injured or killed; involved in a flood, fire, or other natural disaster; involved in a life-threatening accident; suffered a great shock because one of the above events happened to someone close to you; and other. Participants were asked to list up to three traumatic events and describe the trauma in detail. Those reporting traumatic experiences were then interviewed for potential PTSD symptoms. After the data were scored, PTSD diagnoses were generated based on DSM-IV criteria. The institutional review boards at Yale University School of Medicine, the University of Connecticut Health Center, the University of Pennsylvania School of Medicine, the Medical University of South Carolina, and McLean Hospital approved the study.
