## Supplemental Text for "Discovery of 95 PTSD loci provides insight into genetic architecture and neurobiology of trauma and stress-related disorders"

### European GWAS: Sex chromosomes analysis

#### X chromosome

For the first time in a large GWAS meta-analysis of PTSD, we analyzed the X-chromosome.  Five loci exceeded genome-wide significance, four of which were intragenic.  As can be seen in Supplementary Fig. 1, the top X-chromosome locus is in the *IGSF1* gene (rs112052534, *p*=2.4x10^-9^).  *IGSF1* is highly expressed in the pituitary and hypothalamus, and mutations in *IGSF1* have been linked to central hypothyroidism,^1^ a condition that causes symptoms shared with some psychiatric disorders (e.g. fatigue and concentration difficulties).  The second, third, and fifth most significant loci on the X-chromosome were located in the *MTM1*, *ENSG00000283380*, and *POLA1* genes respectively, which are widely expressed throughout the body, including in the brain.^2^ *MTM1* encodes myotubularin, and mutations in this gene have been linked to myopathies.^3^ *ENSG00000283380* is an RNA gene of the lncRNA class and *POLA1* encodes a catalytic subunit of DNA polymerase. The fourth locus was intergenic. Notably, the *ENSG00000283380* locus was previously reported as being associated with schizophrenia,^4^ with the same top SNP (rs1378559, PTSD: *p*=2.0x10^-8^, schizophrenia: *p*=2.0x10^-17^) and the same direction of effect (T allele associated with higher risk). To our knowledge, none of the other loci have been previously linked to psychiatric disorders, though only a minority of large psychiatric GWAS have included the X chromosome.

#### Y chromosome

Analysis of the Y chromosome yielded no significant loci (Supplementary Fig. 2).  However, it must be noted that, in addition to being the smallest chromosome, coverage of the Y chromosome was much lower than for other chromosomes.  Indeed, imputation methods for the Y chromosome are still not widely used,^5^ and so we took the conservative approach of meta-analyzing only genotyped SNP results across cohorts (after cleaning steps and GWAS were applied to each cohort individually, as was done for the autosomes and the X chromosome).  This meant that, for most Y chromosome variants, the large majority of cohorts *did not* have data available.  Indeed, the largest sample size for any Y chromosome variant was 69,321, whereas the maximum sample size for an autosomal variant was 1,307,247. In other words, the maximum Y chromosome sample size was just 5% of the maximum autosomal sample size.  Of the analyzed variants, the top Y chromosome SNP was rs3902 (*p*=2.5x10^-4^*)*.

### Gene-based findings unique from GWAS

52 genes in gene-based analysis were not already named by FUMA gene-mapping strategies applied to significant SNPs from GWAS. It is possible that these associations arose in gene-based analysis due to carrying independent functional SNPs that were not genome-wide significant. We performed GCTA COJO analysis to examine conditional independence of SNPs within these 52 genes. For 11/52 (21%) of genes (*ASTN2, CACNA2D3, CNTN5, CTNNA2, DRD2, EXOC4, GRM7, KCNMB2, LSAMP*, *SEMA6D,* and *TENM2*) there was evidence (P < 0.05/52) of at least 2 conditionally independent significant SNPs in the gene region. The association with *LSAMP,* which has been reported in other recent GWAS,^6^ as well as *KCNMB2*, may be overstated due to inaccurate gene boundary definitions in FUMA.

#### *LSAMP* association under gene-based tests is inflated

*LSAMP* was reported as a significant gene under gene-based associations. We note that this may be a false positive due to the gene region defined in FUMA annotations. FUMA annotations have *LSAMP* as spanning over an approximately 2.2 megabase region (chr3:115,521,235-117,716,095, GRCh37 coordinates). This is the entire range of all transcripts spanned by *LSAMP* as annotated by Ensembl genes under genome build hg37. Regional plots of our results indicate multiple separate LD blocks spanning beyond *LSAMP*, that are being counted in the gene-based LSAMP association test. This seems to be due to the broad boundary of the gene transcripts under hg37: in updated hg38/GRCh38 coordinates, *LSAMP* transcripts range over a much smaller 1.3 MB region (chr3:115,802,363-117,139,389).

### Review of genes in top 20% of prioritized loci

Results are presented in order of significance of leading variants.

##### ZDHHC5

*ZDHHC5* (Zinc Finger DHHC-Type Palmitoyltransferase 5) encodes an enzyme that belongs to the DHHC family of palmitoyl transferases which catalyzes the transfer of the fatty acid palmitate onto protein substrates.^7^ This modification can regulate protein localization, stability, and function. ZDHHC5 may be involved in the regulation of protein palmitoylation in the brain, which is known to be important for synaptic plasticity and neuronal function, and may be disrupted in schizophrenia.^8,9^ ZDHHC5 has also been linked in association studies to schizophrenia^10,11,12^ as well as Alzheimer's disease.^13^

##### FOXP2

The *FOXP2* (Forkhead Box Protein P2) gene encodes a transcription factor protein that is involved in the development and function of the nervous system, particularly in the areas related to language and speech.^14^ Mutations in the FOXP2 gene have been linked to developmental speech and language disorders, as well as social behavior, cognitive function, and motor control.^15,16^ FOXP2 has also been associated with multiple psychiatric conditions including schizophrenia,^17^ major depression,^18^ and substance use disorders.^19^

##### HLA region

The *HLA* (Human Leukocyte Antigen) region is a cluster of genes located on the short arm of chromosome 6 that encode the major histocompatibility complex (MHC) proteins in humans. MHC proteins play a critical role in the immune response by presenting antigens to T cells, which triggers an immune reaction against the foreign invader. The HLA region is highly polymorphic. This genetic diversity allows for a wide range of antigens to be presented to the immune system, which is important for mounting an effective immune response against diverse pathogens. The HLA region has been associated with the development of autoimmune diseases,^20,21^ the rejection of transplanted tissues and organs,^22^ alongside a vast array of other complex disorders.^23^

##### Chromosome 3 gene-rich region in 3p21, including:

The chromosome 3p21 top hit in our GWAS is a region on the short arm of chromosome 3 that contains a high density of genes. Some of the key genes in the 3p21 region include:

*RNF123 -* The gene *RNF123* (Ring Finger Protein 123) encodes a protein that belongs to the RING finger family of E3 ubiquitin ligases. E3 ubiquitin ligases are enzymes that facilitate the transfer of ubiquitin molecules to target proteins, leading to their degradation by the proteasome or altering their cellular localization and activity.^24^ *RNF123* has been implicated in the development and progression of various cancers and may function as a tumor suppressor.^25^ Of note, RFN123 expression has been linked to depressive disorder and psychosis.^26,27^

*CAMKV -* The gene *CAMKV* (calmodulin-dependent protein kinase 5) codes for a protein called calcium/calmodulin-dependent protein kinase V. This protein is primarily found in the brain and is involved in the regulation of neuronal activity.^28^ Research suggests that CAMKV may play a role in learning and memory processes in the brain via maintenance of dendritic spines.^29^ Prior studies have shown that mutations in the *CAMKV* gene may be associated with schizophrenia^30^ and autism spectrum disorder.^31,32^

*MST1R –* The *MST1R* gene encodes the macrophage-stimulating 1 receptor protein, which is a receptor tyrosine kinase. This protein is involved in several cellular processes, including cell proliferation, differentiation, migration, and survival.^33^ It has been associated with human cancers,^34^ as well as with psychiatric conditions including anorexia nervosa.^35,36^

*RBM6 - RBM6* encodes RNA binding motif protein 6. This protein is involved in the regulation of RNA splicing. *RBM6* has been linked to major depressive disorder in multiple prior studies.^37,38^ Other work have also suggested that RBM6 may be involved in the regulation of synaptic plasticity, which is a key process underlying learning and memory and is also implicated in various psychiatric disorders.^39^

*SEMA3F -* The SEMA3F gene provides instructions for producing a protein called semaphorin 3F, which is a member of the semaphorin family of proteins. Semaphorins are involved in cell growth, differentiation, migration, and axon guidance during development.^40,41^ SEMA3F regulates the activity of the glutamate receptor NMDAR, which is involved in the pathophysiology of several psychiatric conditions, including schizophrenia and depression.^42,43^

### 11q12.1

The 11q12.1 region on the long arm of chromosome 11 houses a number of genes that were significant hits in our study. Studies have identified genetic variants in this region to be associated with multiple psychiatric and neurological conditions, including schizophrenia, bipolar disorder, major depressive disorder, and Alzheimer's disease. Specifically, the genes pinpointed in our work include *CLP1, CTNND1, RP11-691N7.6, SERPING1, TMX2-CTNND1, ZDHHC5*. *CLP1* is a gene that encodes for a protein which is involved in RNA processing and splicing.

*CLP1* (cleavage and polyadenylation factor 1) has been previously seen to be associated with major depression^44^ as well as having differential expression in bipolar disorder.^45^

*CTNND1* (also known as p120-catenin) is a gene that encodes a cytoplasmic protein that plays a role in cell adhesion and signaling. It has been previously associated with PTSD and migraine in humans,^46^ as well as with stress-induced neuroinflammation in mice.^47^

*RP11-691N7.6* is a long non-coding RNA (lncRNA) gene. RP11-691N7.6 has been implicated in the regulation of gene expression, and may interact with other proteins or RNA molecules to modulate cellular processes. It has been found to be dysregulated in cancer, however, further studies are needed to fully understand the function of RP11-691N7.6 and its potential involvement in PTSD.

*SERPING1* (also known as C1 inhibitor) is a gene that encodes a protein involved in the regulation of the complement system of the immune system. While *SERPING1* has not been directly connected to PTSD previously, it is a primary driver of hereditary angioedema, an immune disorder which can be triggered by trauma or emotional stress.^48,49^ Dysregulation of the complement system has been previously shown to impact other neurodevelopmental disorders including schizophrenia, bipolar disorder, major depressive disorder, and rodent systems have indicated a strong role of the complement system in anxiety behaviors.^50^

The *TMX2-CTNND1* fusion gene, and has been identified in a large meta-analysis of OCD.^51^ Individually, *TMX2* and *CTNND1* (discussed above) have also both individually been linked to psychiatric disorders, including opioid use disorder.^52^

The *ZDHHC5* gene codes for a protein called palmitoyltransferase ZDHHC5, which belongs to the zinc finger DHHC-type palmitoyltransferase family of enzymes. Palmitoylation is a post-translational modification that involves the addition of a fatty acid molecule (palmitate) to cysteine residues in proteins, and it has been shown to be involved in a variety of cellular processes, including protein trafficking, signaling, neuronal development, and synaptic plasticity. ZDHHC5 is primarily expressed in the brain and has been implicated in several neurological and psychiatric conditions, including schizophrenia and substance use disorders.^53,8,7^

##### NCAM1

*NCAM1* (Neural Cell Adhesion Molecule 1) is a gene that encodes a cell adhesion molecule involved in cell-cell communication and signal transduction. The NCAM1 gene has been implicated in various psychiatric disorders, including schizophrenia, bipolar disorder, and major depressive disorder.^54,55^ More specifically, it has been connected to multiple brain-related biological processes, including neuronal migration, axonal branching, fasciculation, and synaptogenesis, with a pivotal role in synaptic plasticity. Notably, it has previously been shown to affect memory, with epigenetic modifications to *NCAM1* affecting PTSD symptomology.^56^

##### ESR1

The *ESR1* gene codes for the estrogen receptor alpha, which is a nuclear receptor that binds to the hormone estrogen. In psychiatry, *ESR1* has been implicated in a number of disorders and conditions, including depression, anxiety, suicidal ideation, and has previously been connected with PTSD.^57,58,59^ Research suggests that *ESR1* may influence the development and function of the brain, as well as the regulation of mood, cognition, and behavior.

##### SGCD

The *SGCD* gene (Sarcoglycan Delta) encodes a protein that is involved in the formation of a complex of proteins that make up the dystrophin-associated glycoprotein complex (DGC), which is critical for maintaining the integrity and stability of muscle fibers. While the direct mechanistic connection of *SGCD* to psychiatric conditions is not clear, this gene has been previously associated with lifetime trauma exposure in biobanks.^60^

##### DCC

The *DCC* (Deleted in Colorectal Cancer) gene encodes for a transmembrane receptor protein that is involved in the guidance of developing neurons in the central nervous system. *DCC* is considered a pleiotropic risk factor for a variety of psychiatric disorders, and has been implicated in schizophrenia, bipolar disorder, major depression, suicidal ideation, as well as in PTSD.^61,62,63^

##### FAM120A

The *FAM120A* (Family with sequence similarity 120A) protein product is widely expressed across tissues. While the precise function of this protein is not well understood, recent studies have suggested that FAM120A may play a role in the regulation of immune system function and inflammation. *FAM120A* has been previously associated to PTSD, among other psychiatric phenotypes, in large scale GWAS and GWAS meta-analyses.^60,64,65^

### 15q26.1

The 15q26.1 locus on chromosome 15 has been associated with several different conditions and traits, including psychiatric disorders such as bipolar disorder, schizophrenia, and autism spectrum disorder. One of the most well-studied genes in this region is *CHRNA7*, which codes for a subunit of a type of nicotinic acetylcholine receptor (nAChR) that is involved in various physiological processes, including neural signaling and inflammation. Alterations in CHRNA7 expression or function have been implicated in schizophrenia, bipolar disorder, and epilepsy. Other genes in the 15q26.1 region have also been implicated in psychiatric and neurological disorders, although their precise roles in these conditions are less clear. For example, the gene *KLF13* has been associated with bipolar disorder and has been shown to play a role in neural development and function, while the gene *MTMR10* has been implicated in autism spectrum disorder and may be involved in the regulation of neuronal signaling and synapse function.

The two genes specifically fine-mapped to in our study, *FES* and *FURIN* are close to each other and in moderate to strong LD. Expression of FES and FURIN in the brain was found to result in increased risk of schizophrenia and major depressive disorder.^66^ A variant in *FURIN* was additionally seen to have the highest evidence for gene-by-environment interaction in a study of cumulative lifetime trauma burden.^67^ Similarly, *FES* has been associated with childhood maltreatment in multi-omics analyses of expression QTL and chromatin interaction.^68^

### 7p22.3

The 7p22.3 locus has been implicated in several psychiatric disorders, including schizophrenia and bipolar disorder. One of the most well-studied genes in this region is *CNTNAP2*, which codes for a protein that is involved in neural cell adhesion and communication. Alterations in *CNTNAP2* expression or function have been associated with a range of neuropsychiatric disorders, including autism spectrum disorder, language disorders, and epilepsy. Other genes in the 7p22.3 region have also been implicated in psychiatric and neurological disorders, although their precise roles in these conditions are less clear. For example, the gene *GABBR1* has been associated with schizophrenia and has been shown to play a role in the regulation of inhibitory neurotransmission, while the gene *DLG2* has been implicated in bipolar disorder and may be involved in the regulation of synaptic plasticity.

The two top genes fine-mapped within the region in our study were *AC110781.3* and *MAD1L1*. AC110781.3 is a long non-coding RNA that has been found to be associated with mood instability and schizophrenia, bipolar disorder, depression, and ADHD.^69,70^ *MAD1L1* encodes a protein called mitotic spindle assembly checkpoint protein MAD1, which is involved in the regulation of cell division and the maintenance of genomic stability. The MVP previously found *MAD1L1* to be associated with PTSD in the EUR and cross-ancestry meta-analyses.^71^ The gene has also been replicated in other studies as a marker for resilience, with differences in *MAD1L1* methylation impacting longitudinal PTSD outcomes.^72,73,74^

##### MDGA2

*MDGA2* (MAM domain-containing glycosylphosphatidylinositol anchor 2) is involved in the development and function of the nervous system, and has been implicated in several psychiatric and neurodevelopmental disorders. MDGA2 acts as a cell adhesion molecule and plays an important role in the formation and maintenance of synapses.^75^ The gene has been associated with MDD^76^ and autism^75^ in prior work.

### 3p21.33 / 3p22.1

Two genes are present in this locus: *ANO10* and *SNRK. ANO10* is a gene that encodes for a calcium-activated chloride channel and is involved in several cellular processes, including ion transport and lipid metabolism. It has been linked to ataxia, epilepsy, and learning difficulties in human studies.^77,78,79^ The *SNRK* (SNF1-related kinase) gene encodes for a protein kinase, which is an enzyme that regulates the activity of other proteins through the transfer of phosphate groups. SNRK has been associated with schizophrenia in prior EUR GWAS,^80^ as well as with autism.^81^

##### CNTNAP5

*CNTNAP5* (Contactin Associated Protein-Like 5) is a gene that encodes a protein that is widely expressed in the brain and plays a role in neural development and function. It has been associated with self-reported childhood maltreatment and bipolar disorder in prior studies,^82,83^ as well as with antipsychotic response.^84^ Notably, *CNTNAP5* was found as an overlapping top prioritized gene between PTSD and ASD.^85^

##### TSNARE1

*TSNARE1* (also known as SNAP23) is a gene that encodes a protein involved in vesicle fusion, a process that plays a crucial role in neurotransmitter release in the brain. *TSNARE1* has been specifically associated PTSD directly,^86,87^ as well as with a number of PTSD symptoms, including reexperiencing, avoidance, negative emotional symptoms, and hyperarousal,^88^ in addition to risk-taking.^89^

##### EFNA5

The gene *EFNA5* (ephrin-A5) encodes a protein called ephrin-A5, which is a member of the ephrin family of proteins. In the nervous system, ephrin-A5 is involved in axon guidance and synapse formation during development, and it also plays a role in synaptic plasticity. *EFNA5* was found to be associated with modifying expression in acute stress^90^ and has been seen to be associated in anxiety disorders^91^ as well as traumatic brain injury and recovery.^92^ Importantly, it has been shown to be regulated by nutraceutical omega-3 fatty acids, making it of potential pharmacogenomics utility.^91^

### Phenotype definitions for EHR

#### Case/Control definitions

Cases: Those with at least 1) one diagnosis of PTSD and/or 2) one diagnosis of other stress disorder

Controls: Include all other participants

#### ICD codes utilized

Diagnoses can be ICD-9 or 10. PTSD Dx are bolded, adjustment disorders are italicized.

**Possible ICD-9 codes:**

[*309*](https://urldefense.proofpoint.com/v2/url?u=http-3A__www.icd9data.com_2015_Volume1_290-2D319_300-2D316_309_309.htm&d=DwMFAw&c=G2MiLlal7SXE3PeSnG8W6_JBU6FcdVjSsBSbw6gcR0U&r=QGiOGwneOYKdMI_b-4g5Cita4ycB2h5Um83YWCmxPRE&m=lNvl5Zl-grDHBVMNAPJuB6TCOd8gxK_qWxqLvzNOeP0&s=7JbZn_rpD_ir3585dpJoMJDtQ6N0QQK2BJbeyzsVG5M&e=) *Adjustment reaction*

[*309.0*](https://urldefense.proofpoint.com/v2/url?u=http-3A__www.icd9data.com_2015_Volume1_290-2D319_300-2D316_309_309.0.htm&d=DwMFAw&c=G2MiLlal7SXE3PeSnG8W6_JBU6FcdVjSsBSbw6gcR0U&r=QGiOGwneOYKdMI_b-4g5Cita4ycB2h5Um83YWCmxPRE&m=lNvl5Zl-grDHBVMNAPJuB6TCOd8gxK_qWxqLvzNOeP0&s=yHd66qvFqCqUCKJ5V5CVtEDCUSKJOvW9wtqhKGdfwBo&e=) *Adjustment disorder with depressed mood* [*convert 309.0 to ICD-10-CM*](https://urldefense.proofpoint.com/v2/url?u=http-3A__www.icd10data.com_Convert_309.0&d=DwMFAw&c=G2MiLlal7SXE3PeSnG8W6_JBU6FcdVjSsBSbw6gcR0U&r=QGiOGwneOYKdMI_b-4g5Cita4ycB2h5Um83YWCmxPRE&m=lNvl5Zl-grDHBVMNAPJuB6TCOd8gxK_qWxqLvzNOeP0&s=y_cOzRWWjanbyM0cq2RSpvVDIfF2Vb7nEUiv_zZEct0&e=)

[*309.1*](https://urldefense.proofpoint.com/v2/url?u=http-3A__www.icd9data.com_2015_Volume1_290-2D319_300-2D316_309_309.1.htm&d=DwMFAw&c=G2MiLlal7SXE3PeSnG8W6_JBU6FcdVjSsBSbw6gcR0U&r=QGiOGwneOYKdMI_b-4g5Cita4ycB2h5Um83YWCmxPRE&m=lNvl5Zl-grDHBVMNAPJuB6TCOd8gxK_qWxqLvzNOeP0&s=GSatAh5E5mSS_VXR3I9SbTuHdx1vajshfYPZAn_PCiI&e=) *Prolonged depressive reaction* [*convert 309.1 to ICD-10-CM*](https://urldefense.proofpoint.com/v2/url?u=http-3A__www.icd10data.com_Convert_309.1&d=DwMFAw&c=G2MiLlal7SXE3PeSnG8W6_JBU6FcdVjSsBSbw6gcR0U&r=QGiOGwneOYKdMI_b-4g5Cita4ycB2h5Um83YWCmxPRE&m=lNvl5Zl-grDHBVMNAPJuB6TCOd8gxK_qWxqLvzNOeP0&s=V-TOp_GIZW_Vb4hlat05Ui090QU2JWomx__zYgRvrx8&e=)

[*309.2*](https://urldefense.proofpoint.com/v2/url?u=http-3A__www.icd9data.com_2015_Volume1_290-2D319_300-2D316_309_309.2.htm&d=DwMFAw&c=G2MiLlal7SXE3PeSnG8W6_JBU6FcdVjSsBSbw6gcR0U&r=QGiOGwneOYKdMI_b-4g5Cita4ycB2h5Um83YWCmxPRE&m=lNvl5Zl-grDHBVMNAPJuB6TCOd8gxK_qWxqLvzNOeP0&s=zqrss9RtEklF1SQTZYWdQzer8IWMoOF6UgY5PdNsYZ4&e=) *Adjustment reaction with predominant disturbance of other emotions*

[*309.21*](https://urldefense.proofpoint.com/v2/url?u=http-3A__www.icd9data.com_2015_Volume1_290-2D319_300-2D316_309_309.21.htm&d=DwMFAw&c=G2MiLlal7SXE3PeSnG8W6_JBU6FcdVjSsBSbw6gcR0U&r=QGiOGwneOYKdMI_b-4g5Cita4ycB2h5Um83YWCmxPRE&m=lNvl5Zl-grDHBVMNAPJuB6TCOd8gxK_qWxqLvzNOeP0&s=gaKH-VYo_k1BcqGkguvI0_4TuR8LTRMAmsHJmgQ_cck&e=) *Separation anxiety disorder* [*convert 309.21 to ICD-10-CM*](https://urldefense.proofpoint.com/v2/url?u=http-3A__www.icd10data.com_Convert_309.21&d=DwMFAw&c=G2MiLlal7SXE3PeSnG8W6_JBU6FcdVjSsBSbw6gcR0U&r=QGiOGwneOYKdMI_b-4g5Cita4ycB2h5Um83YWCmxPRE&m=lNvl5Zl-grDHBVMNAPJuB6TCOd8gxK_qWxqLvzNOeP0&s=FmIKXs4tp0Gk5e3wkjgJD-11n0UbtdRDMpsssng6eQc&e=)

[*309.22*](https://urldefense.proofpoint.com/v2/url?u=http-3A__www.icd9data.com_2015_Volume1_290-2D319_300-2D316_309_309.22.htm&d=DwMFAw&c=G2MiLlal7SXE3PeSnG8W6_JBU6FcdVjSsBSbw6gcR0U&r=QGiOGwneOYKdMI_b-4g5Cita4ycB2h5Um83YWCmxPRE&m=lNvl5Zl-grDHBVMNAPJuB6TCOd8gxK_qWxqLvzNOeP0&s=wzISu-wlv6nLBFsNAN9S0_Nt2v9l1mrWrAiF6LiHz8M&e=) *Emancipation disorder of adolescence and early adult life* [*convert 309.22 to ICD-10-CM*](https://urldefense.proofpoint.com/v2/url?u=http-3A__www.icd10data.com_Convert_309.22&d=DwMFAw&c=G2MiLlal7SXE3PeSnG8W6_JBU6FcdVjSsBSbw6gcR0U&r=QGiOGwneOYKdMI_b-4g5Cita4ycB2h5Um83YWCmxPRE&m=lNvl5Zl-grDHBVMNAPJuB6TCOd8gxK_qWxqLvzNOeP0&s=9xyJXAdMcd4v1MhjRs46sfXKvYv6dYEdFXgwXt_DeUA&e=)

[*309.23*](https://urldefense.proofpoint.com/v2/url?u=http-3A__www.icd9data.com_2015_Volume1_290-2D319_300-2D316_309_309.23.htm&d=DwMFAw&c=G2MiLlal7SXE3PeSnG8W6_JBU6FcdVjSsBSbw6gcR0U&r=QGiOGwneOYKdMI_b-4g5Cita4ycB2h5Um83YWCmxPRE&m=lNvl5Zl-grDHBVMNAPJuB6TCOd8gxK_qWxqLvzNOeP0&s=IXGmTnEI2lOg5_0VJsOD-eQ8zLWyCvC9DRytf4CGtCg&e=) *Specific academic or work inhibition* [*convert 309.23 to ICD-10-CM*](https://urldefense.proofpoint.com/v2/url?u=http-3A__www.icd10data.com_Convert_309.23&d=DwMFAw&c=G2MiLlal7SXE3PeSnG8W6_JBU6FcdVjSsBSbw6gcR0U&r=QGiOGwneOYKdMI_b-4g5Cita4ycB2h5Um83YWCmxPRE&m=lNvl5Zl-grDHBVMNAPJuB6TCOd8gxK_qWxqLvzNOeP0&s=WGVMkdBoPNKN-tmTMyU0oSNnIrnRkgSpJENR5vYyrcA&e=)

[*309.24*](https://urldefense.proofpoint.com/v2/url?u=http-3A__www.icd9data.com_2015_Volume1_290-2D319_300-2D316_309_309.24.htm&d=DwMFAw&c=G2MiLlal7SXE3PeSnG8W6_JBU6FcdVjSsBSbw6gcR0U&r=QGiOGwneOYKdMI_b-4g5Cita4ycB2h5Um83YWCmxPRE&m=lNvl5Zl-grDHBVMNAPJuB6TCOd8gxK_qWxqLvzNOeP0&s=fXmO6AA-A-N7ZGgTg5rqh2ndCAA6p9erig_8px8smkM&e=) *Adjustment disorder with anxiety* [*convert 309.24 to ICD-10-CM*](https://urldefense.proofpoint.com/v2/url?u=http-3A__www.icd10data.com_Convert_309.24&d=DwMFAw&c=G2MiLlal7SXE3PeSnG8W6_JBU6FcdVjSsBSbw6gcR0U&r=QGiOGwneOYKdMI_b-4g5Cita4ycB2h5Um83YWCmxPRE&m=lNvl5Zl-grDHBVMNAPJuB6TCOd8gxK_qWxqLvzNOeP0&s=UBSqNHysT5a9680udOo3eE9fGBY53sy-8265vl2y-C8&e=)

[*309.28*](https://urldefense.proofpoint.com/v2/url?u=http-3A__www.icd9data.com_2015_Volume1_290-2D319_300-2D316_309_309.28.htm&d=DwMFAw&c=G2MiLlal7SXE3PeSnG8W6_JBU6FcdVjSsBSbw6gcR0U&r=QGiOGwneOYKdMI_b-4g5Cita4ycB2h5Um83YWCmxPRE&m=lNvl5Zl-grDHBVMNAPJuB6TCOd8gxK_qWxqLvzNOeP0&s=IsZr3tZWMOYyjMwjYPL51RoxSNQonK-zijb8WxUI11U&e=) *Adjustment disorder with mixed anxiety and depressed mood* [*convert 309.28 to ICD-10-CM*](https://urldefense.proofpoint.com/v2/url?u=http-3A__www.icd10data.com_Convert_309.28&d=DwMFAw&c=G2MiLlal7SXE3PeSnG8W6_JBU6FcdVjSsBSbw6gcR0U&r=QGiOGwneOYKdMI_b-4g5Cita4ycB2h5Um83YWCmxPRE&m=lNvl5Zl-grDHBVMNAPJuB6TCOd8gxK_qWxqLvzNOeP0&s=p0WWftG45njEQqZC3kMx3PbwzsDbYyCV9fl1xRB7v_c&e=)

[*309.29*](https://urldefense.proofpoint.com/v2/url?u=http-3A__www.icd9data.com_2015_Volume1_290-2D319_300-2D316_309_309.29.htm&d=DwMFAw&c=G2MiLlal7SXE3PeSnG8W6_JBU6FcdVjSsBSbw6gcR0U&r=QGiOGwneOYKdMI_b-4g5Cita4ycB2h5Um83YWCmxPRE&m=lNvl5Zl-grDHBVMNAPJuB6TCOd8gxK_qWxqLvzNOeP0&s=xR2CKCq5pruwIQXkwClnxU1Fic6ymibJ0bLEzmUNGPE&e=) *Other adjustment reactions with predominant disturbance of other emotions* [*convert 309.29 to ICD-10-CM*](https://urldefense.proofpoint.com/v2/url?u=http-3A__www.icd10data.com_Convert_309.29&d=DwMFAw&c=G2MiLlal7SXE3PeSnG8W6_JBU6FcdVjSsBSbw6gcR0U&r=QGiOGwneOYKdMI_b-4g5Cita4ycB2h5Um83YWCmxPRE&m=lNvl5Zl-grDHBVMNAPJuB6TCOd8gxK_qWxqLvzNOeP0&s=_DIHAeuoNSpTfQ1OS064YgI00Wm2BdHbRGi_sYQErxE&e=)

[*309.3*](https://urldefense.proofpoint.com/v2/url?u=http-3A__www.icd9data.com_2015_Volume1_290-2D319_300-2D316_309_309.3.htm&d=DwMFAw&c=G2MiLlal7SXE3PeSnG8W6_JBU6FcdVjSsBSbw6gcR0U&r=QGiOGwneOYKdMI_b-4g5Cita4ycB2h5Um83YWCmxPRE&m=lNvl5Zl-grDHBVMNAPJuB6TCOd8gxK_qWxqLvzNOeP0&s=RKcPzLZCiLBeXorcsge9xvNbquo6UAKgd8eRN_svj9E&e=) *Adjustment disorder with disturbance of conduct* [*convert 309.3 to ICD-10-CM*](https://urldefense.proofpoint.com/v2/url?u=http-3A__www.icd10data.com_Convert_309.3&d=DwMFAw&c=G2MiLlal7SXE3PeSnG8W6_JBU6FcdVjSsBSbw6gcR0U&r=QGiOGwneOYKdMI_b-4g5Cita4ycB2h5Um83YWCmxPRE&m=lNvl5Zl-grDHBVMNAPJuB6TCOd8gxK_qWxqLvzNOeP0&s=DYimWnr6myLgz31etd5WdGieiYobrKw3k1-bFnIAfnw&e=)

[*309.4*](https://urldefense.proofpoint.com/v2/url?u=http-3A__www.icd9data.com_2015_Volume1_290-2D319_300-2D316_309_309.4.htm&d=DwMFAw&c=G2MiLlal7SXE3PeSnG8W6_JBU6FcdVjSsBSbw6gcR0U&r=QGiOGwneOYKdMI_b-4g5Cita4ycB2h5Um83YWCmxPRE&m=lNvl5Zl-grDHBVMNAPJuB6TCOd8gxK_qWxqLvzNOeP0&s=S9JbyRBFzzNDnzxeEdmFJmnK0P1720qlwjzuz26EW2Q&e=) *Adjustment disorder with mixed disturbance of emotions and conduct* [*convert 309.4 to ICD-10-CM*](https://urldefense.proofpoint.com/v2/url?u=http-3A__www.icd10data.com_Convert_309.4&d=DwMFAw&c=G2MiLlal7SXE3PeSnG8W6_JBU6FcdVjSsBSbw6gcR0U&r=QGiOGwneOYKdMI_b-4g5Cita4ycB2h5Um83YWCmxPRE&m=lNvl5Zl-grDHBVMNAPJuB6TCOd8gxK_qWxqLvzNOeP0&s=M3fXOCu1RTdnpa-N1tHm67uRSxO_NbnW6-REnaIAoac&e=)

[*309.8*](https://urldefense.proofpoint.com/v2/url?u=http-3A__www.icd9data.com_2015_Volume1_290-2D319_300-2D316_309_309.8.htm&d=DwMFAw&c=G2MiLlal7SXE3PeSnG8W6_JBU6FcdVjSsBSbw6gcR0U&r=QGiOGwneOYKdMI_b-4g5Cita4ycB2h5Um83YWCmxPRE&m=lNvl5Zl-grDHBVMNAPJuB6TCOd8gxK_qWxqLvzNOeP0&s=BY30FWc7tEz98OxyqWTtvaxSHVDAV6Ormxjoj4qLAhg&e=) *Other specified adjustment reactions*

[**309.81**](https://urldefense.proofpoint.com/v2/url?u=http-3A__www.icd9data.com_2015_Volume1_290-2D319_300-2D316_309_309.81.htm&d=DwMFAw&c=G2MiLlal7SXE3PeSnG8W6_JBU6FcdVjSsBSbw6gcR0U&r=QGiOGwneOYKdMI_b-4g5Cita4ycB2h5Um83YWCmxPRE&m=lNvl5Zl-grDHBVMNAPJuB6TCOd8gxK_qWxqLvzNOeP0&s=5iCw-0SfxDyyXTy72xpDYc7M405mS47-Uk4r7Uw-ASg&e=) **Posttraumatic stress disorder** [**convert 309.81 to ICD-10-CM**](https://urldefense.proofpoint.com/v2/url?u=http-3A__www.icd10data.com_Convert_309.81&d=DwMFAw&c=G2MiLlal7SXE3PeSnG8W6_JBU6FcdVjSsBSbw6gcR0U&r=QGiOGwneOYKdMI_b-4g5Cita4ycB2h5Um83YWCmxPRE&m=lNvl5Zl-grDHBVMNAPJuB6TCOd8gxK_qWxqLvzNOeP0&s=8Ut-qCueFXlxUD7KbNxNHhZPc6MpT6foY9M3Krj2kXY&e=)

[*309.82*](https://urldefense.proofpoint.com/v2/url?u=http-3A__www.icd9data.com_2015_Volume1_290-2D319_300-2D316_309_309.82.htm&d=DwMFAw&c=G2MiLlal7SXE3PeSnG8W6_JBU6FcdVjSsBSbw6gcR0U&r=QGiOGwneOYKdMI_b-4g5Cita4ycB2h5Um83YWCmxPRE&m=lNvl5Zl-grDHBVMNAPJuB6TCOd8gxK_qWxqLvzNOeP0&s=xUHIiXtmCK1AZOgB-oU8hJ9CVAgLA_xcTOm9dZCHcFY&e=) *Adjustment reaction with physical symptoms* [*convert 309.82 to ICD-10-CM*](https://urldefense.proofpoint.com/v2/url?u=http-3A__www.icd10data.com_Convert_309.82&d=DwMFAw&c=G2MiLlal7SXE3PeSnG8W6_JBU6FcdVjSsBSbw6gcR0U&r=QGiOGwneOYKdMI_b-4g5Cita4ycB2h5Um83YWCmxPRE&m=lNvl5Zl-grDHBVMNAPJuB6TCOd8gxK_qWxqLvzNOeP0&s=XapoVAVYO10mnvGuXGw6F33AuDvUQs07wjThLp2-32U&e=)

[*309.83*](https://urldefense.proofpoint.com/v2/url?u=http-3A__www.icd9data.com_2015_Volume1_290-2D319_300-2D316_309_309.83.htm&d=DwMFAw&c=G2MiLlal7SXE3PeSnG8W6_JBU6FcdVjSsBSbw6gcR0U&r=QGiOGwneOYKdMI_b-4g5Cita4ycB2h5Um83YWCmxPRE&m=lNvl5Zl-grDHBVMNAPJuB6TCOd8gxK_qWxqLvzNOeP0&s=yI8LKyV-TOQUlY7ORyLs8Z8FR_44ASuAxFQFRmbs6gs&e=) *Adjustment reaction with withdrawal* [*convert 309.83 to ICD-10-CM*](https://urldefense.proofpoint.com/v2/url?u=http-3A__www.icd10data.com_Convert_309.83&d=DwMFAw&c=G2MiLlal7SXE3PeSnG8W6_JBU6FcdVjSsBSbw6gcR0U&r=QGiOGwneOYKdMI_b-4g5Cita4ycB2h5Um83YWCmxPRE&m=lNvl5Zl-grDHBVMNAPJuB6TCOd8gxK_qWxqLvzNOeP0&s=TQ_OYfTlor5XLEf3hiqse-RACPO-oFtegPcGQd_RvII&e=)

[*309.89*](https://urldefense.proofpoint.com/v2/url?u=http-3A__www.icd9data.com_2015_Volume1_290-2D319_300-2D316_309_309.89.htm&d=DwMFAw&c=G2MiLlal7SXE3PeSnG8W6_JBU6FcdVjSsBSbw6gcR0U&r=QGiOGwneOYKdMI_b-4g5Cita4ycB2h5Um83YWCmxPRE&m=lNvl5Zl-grDHBVMNAPJuB6TCOd8gxK_qWxqLvzNOeP0&s=4lHUlBuRPC-RvCTSGr8bVBEhNAJvqw9d2WDLiiaFGlY&e=) *Other specified adjustment reactions* [*convert 309.89 to ICD-10-CM*](https://urldefense.proofpoint.com/v2/url?u=http-3A__www.icd10data.com_Convert_309.89&d=DwMFAw&c=G2MiLlal7SXE3PeSnG8W6_JBU6FcdVjSsBSbw6gcR0U&r=QGiOGwneOYKdMI_b-4g5Cita4ycB2h5Um83YWCmxPRE&m=lNvl5Zl-grDHBVMNAPJuB6TCOd8gxK_qWxqLvzNOeP0&s=CDDUwKBnRbqIqrqzX-FCOyhRmQe1XTKXWX50kSSZIWk&e=)

[*309.9*](https://urldefense.proofpoint.com/v2/url?u=http-3A__www.icd9data.com_2015_Volume1_290-2D319_300-2D316_309_309.9.htm&d=DwMFAw&c=G2MiLlal7SXE3PeSnG8W6_JBU6FcdVjSsBSbw6gcR0U&r=QGiOGwneOYKdMI_b-4g5Cita4ycB2h5Um83YWCmxPRE&m=lNvl5Zl-grDHBVMNAPJuB6TCOd8gxK_qWxqLvzNOeP0&s=MWjXlGRjw811BV-q8L8TNB9z6Iekv-a6N_Q5xivLKMs&e=) *Unspecified adjustment reaction* [*convert 309.9 to ICD-10-CM*](https://urldefense.proofpoint.com/v2/url?u=http-3A__www.icd10data.com_Convert_309.9&d=DwMFAw&c=G2MiLlal7SXE3PeSnG8W6_JBU6FcdVjSsBSbw6gcR0U&r=QGiOGwneOYKdMI_b-4g5Cita4ycB2h5Um83YWCmxPRE&m=lNvl5Zl-grDHBVMNAPJuB6TCOd8gxK_qWxqLvzNOeP0&s=6z58al9CqEq8MLzpFZdFRQnMSdetzOmDXFEsKm9BLmk&e=)

**Possible ICD-10 codes:**

[*F43*](https://urldefense.proofpoint.com/v2/url?u=https-3A__www.icd10data.com_ICD10CM_Codes_F01-2DF99_F40-2DF48_F43-2D_F43&d=DwMFAw&c=G2MiLlal7SXE3PeSnG8W6_JBU6FcdVjSsBSbw6gcR0U&r=QGiOGwneOYKdMI_b-4g5Cita4ycB2h5Um83YWCmxPRE&m=lNvl5Zl-grDHBVMNAPJuB6TCOd8gxK_qWxqLvzNOeP0&s=0UUo4JX_CdpnmaCVNo-xAlSDEWf6r1D6L7ibSmsAqRs&e=) *Reaction to severe stress, and adjustment disorders*

*o*	[*F43.0*](https://urldefense.proofpoint.com/v2/url?u=https-3A__www.icd10data.com_ICD10CM_Codes_F01-2DF99_F40-2DF48_F43-2D_F43.0&d=DwMFAw&c=G2MiLlal7SXE3PeSnG8W6_JBU6FcdVjSsBSbw6gcR0U&r=QGiOGwneOYKdMI_b-4g5Cita4ycB2h5Um83YWCmxPRE&m=lNvl5Zl-grDHBVMNAPJuB6TCOd8gxK_qWxqLvzNOeP0&s=zUTcSUi5935mwZLbIkVMxKVHyn3zG7zvTUWETnQk52E&e=) *Acute stress reaction*

**o**	[**F43.1**](https://urldefense.proofpoint.com/v2/url?u=https-3A__www.icd10data.com_ICD10CM_Codes_F01-2DF99_F40-2DF48_F43-2D_F43.1&d=DwMFAw&c=G2MiLlal7SXE3PeSnG8W6_JBU6FcdVjSsBSbw6gcR0U&r=QGiOGwneOYKdMI_b-4g5Cita4ycB2h5Um83YWCmxPRE&m=lNvl5Zl-grDHBVMNAPJuB6TCOd8gxK_qWxqLvzNOeP0&s=0jD7YfNK9PxbCTXztC5D9mRAKberugOBg6c_21xmUN4&e=) **Post-traumatic stress disorder (PTSD)**

**§**	[**F43.10**](https://urldefense.proofpoint.com/v2/url?u=https-3A__www.icd10data.com_ICD10CM_Codes_F01-2DF99_F40-2DF48_F43-2D_F43.10&d=DwMFAw&c=G2MiLlal7SXE3PeSnG8W6_JBU6FcdVjSsBSbw6gcR0U&r=QGiOGwneOYKdMI_b-4g5Cita4ycB2h5Um83YWCmxPRE&m=lNvl5Zl-grDHBVMNAPJuB6TCOd8gxK_qWxqLvzNOeP0&s=tRP4j9p51aqrJjpsg16SEq0A-XTe6vqHpFNZn2k5Sj8&e=) **Post-traumatic stress disorder, unspecified**

**§**	[**F43.11**](https://urldefense.proofpoint.com/v2/url?u=https-3A__www.icd10data.com_ICD10CM_Codes_F01-2DF99_F40-2DF48_F43-2D_F43.11&d=DwMFAw&c=G2MiLlal7SXE3PeSnG8W6_JBU6FcdVjSsBSbw6gcR0U&r=QGiOGwneOYKdMI_b-4g5Cita4ycB2h5Um83YWCmxPRE&m=lNvl5Zl-grDHBVMNAPJuB6TCOd8gxK_qWxqLvzNOeP0&s=OlT4d0gepSX5llc36_WbE1CNclL_z1GlkLd5VELy2ow&e=) **Post-traumatic stress disorder, acute**

**§**	[**F43.12**](https://urldefense.proofpoint.com/v2/url?u=https-3A__www.icd10data.com_ICD10CM_Codes_F01-2DF99_F40-2DF48_F43-2D_F43.12&d=DwMFAw&c=G2MiLlal7SXE3PeSnG8W6_JBU6FcdVjSsBSbw6gcR0U&r=QGiOGwneOYKdMI_b-4g5Cita4ycB2h5Um83YWCmxPRE&m=lNvl5Zl-grDHBVMNAPJuB6TCOd8gxK_qWxqLvzNOeP0&s=1KKzUcNSogbrEUwJwZTjBtHAPze1Xm7dBpvyot09700&e=) **Post-traumatic stress disorder, chronic**

*o*	[*F43.2*](https://urldefense.proofpoint.com/v2/url?u=https-3A__www.icd10data.com_ICD10CM_Codes_F01-2DF99_F40-2DF48_F43-2D_F43.2&d=DwMFAw&c=G2MiLlal7SXE3PeSnG8W6_JBU6FcdVjSsBSbw6gcR0U&r=QGiOGwneOYKdMI_b-4g5Cita4ycB2h5Um83YWCmxPRE&m=lNvl5Zl-grDHBVMNAPJuB6TCOd8gxK_qWxqLvzNOeP0&s=QgIXypBri8MqdoOPKhJKJBIm98sUx6cT6ywxQteJT7g&e=) *Adjustment disorders*

*§*	[*F43.20*](https://urldefense.proofpoint.com/v2/url?u=https-3A__www.icd10data.com_ICD10CM_Codes_F01-2DF99_F40-2DF48_F43-2D_F43.20&d=DwMFAw&c=G2MiLlal7SXE3PeSnG8W6_JBU6FcdVjSsBSbw6gcR0U&r=QGiOGwneOYKdMI_b-4g5Cita4ycB2h5Um83YWCmxPRE&m=lNvl5Zl-grDHBVMNAPJuB6TCOd8gxK_qWxqLvzNOeP0&s=RFFVNjKc6I1C2_jejsR_R2UxKyATr3s5gQ3d7IzeSnc&e=) *Adjustment disorder, unspecified*

*§*	[*F43.21*](https://urldefense.proofpoint.com/v2/url?u=https-3A__www.icd10data.com_ICD10CM_Codes_F01-2DF99_F40-2DF48_F43-2D_F43.21&d=DwMFAw&c=G2MiLlal7SXE3PeSnG8W6_JBU6FcdVjSsBSbw6gcR0U&r=QGiOGwneOYKdMI_b-4g5Cita4ycB2h5Um83YWCmxPRE&m=lNvl5Zl-grDHBVMNAPJuB6TCOd8gxK_qWxqLvzNOeP0&s=lb4_3vy0JV9wDca5P9nBmN3neIH74qyqrveBLgiJHS0&e=) *Adjustment disorder with depressed mood*

*§*	[*F43.22*](https://urldefense.proofpoint.com/v2/url?u=https-3A__www.icd10data.com_ICD10CM_Codes_F01-2DF99_F40-2DF48_F43-2D_F43.22&d=DwMFAw&c=G2MiLlal7SXE3PeSnG8W6_JBU6FcdVjSsBSbw6gcR0U&r=QGiOGwneOYKdMI_b-4g5Cita4ycB2h5Um83YWCmxPRE&m=lNvl5Zl-grDHBVMNAPJuB6TCOd8gxK_qWxqLvzNOeP0&s=DG99zeoEPVhX3UmVibh_X4WfDNodZo4E_r-CtgV3wOc&e=) *Adjustment disorder with anxiety*

*§*	[*F43.23*](https://urldefense.proofpoint.com/v2/url?u=https-3A__www.icd10data.com_ICD10CM_Codes_F01-2DF99_F40-2DF48_F43-2D_F43.23&d=DwMFAw&c=G2MiLlal7SXE3PeSnG8W6_JBU6FcdVjSsBSbw6gcR0U&r=QGiOGwneOYKdMI_b-4g5Cita4ycB2h5Um83YWCmxPRE&m=lNvl5Zl-grDHBVMNAPJuB6TCOd8gxK_qWxqLvzNOeP0&s=PLW-3rGecgESPhszmmgQZzTbWMw017y5t7YxK5J2210&e=) *Adjustment disorder with mixed anxiety and depressed mood*

*§*	[*F43.24*](https://urldefense.proofpoint.com/v2/url?u=https-3A__www.icd10data.com_ICD10CM_Codes_F01-2DF99_F40-2DF48_F43-2D_F43.24&d=DwMFAw&c=G2MiLlal7SXE3PeSnG8W6_JBU6FcdVjSsBSbw6gcR0U&r=QGiOGwneOYKdMI_b-4g5Cita4ycB2h5Um83YWCmxPRE&m=lNvl5Zl-grDHBVMNAPJuB6TCOd8gxK_qWxqLvzNOeP0&s=q7jtpkr6efkzsKrOqNcqW1BCFhhSh5HWnP07-5wq4IU&e=) *Adjustment disorder with disturbance of conduct*

*§*	[*F43.25*](https://urldefense.proofpoint.com/v2/url?u=https-3A__www.icd10data.com_ICD10CM_Codes_F01-2DF99_F40-2DF48_F43-2D_F43.25&d=DwMFAw&c=G2MiLlal7SXE3PeSnG8W6_JBU6FcdVjSsBSbw6gcR0U&r=QGiOGwneOYKdMI_b-4g5Cita4ycB2h5Um83YWCmxPRE&m=lNvl5Zl-grDHBVMNAPJuB6TCOd8gxK_qWxqLvzNOeP0&s=cEDfy0TizCH8ZJ2WwI2n2v2Goe4ltp2fundyzz_UuEU&e=) *Adjustment disorder with mixed disturbance of emotions and conduct*

*§*	[*F43.29*](https://urldefense.proofpoint.com/v2/url?u=https-3A__www.icd10data.com_ICD10CM_Codes_F01-2DF99_F40-2DF48_F43-2D_F43.29&d=DwMFAw&c=G2MiLlal7SXE3PeSnG8W6_JBU6FcdVjSsBSbw6gcR0U&r=QGiOGwneOYKdMI_b-4g5Cita4ycB2h5Um83YWCmxPRE&m=lNvl5Zl-grDHBVMNAPJuB6TCOd8gxK_qWxqLvzNOeP0&s=FmLAMTkqdeRYxiaa5O8TUVYARkCpY9B8xC02Ti0peDA&e=) *Adjustment disorder with other symptoms*

*o*	[*F43.8*](https://urldefense.proofpoint.com/v2/url?u=https-3A__www.icd10data.com_ICD10CM_Codes_F01-2DF99_F40-2DF48_F43-2D_F43.8&d=DwMFAw&c=G2MiLlal7SXE3PeSnG8W6_JBU6FcdVjSsBSbw6gcR0U&r=QGiOGwneOYKdMI_b-4g5Cita4ycB2h5Um83YWCmxPRE&m=lNvl5Zl-grDHBVMNAPJuB6TCOd8gxK_qWxqLvzNOeP0&s=mjYiPat6n3hYFvTW1sjJ05I0A2M18gC4kfyZrG69V8o&e=) *Other reactions to severe stress*

*o*	[*F43.9*](https://urldefense.proofpoint.com/v2/url?u=https-3A__www.icd10data.com_ICD10CM_Codes_F01-2DF99_F40-2DF48_F43-2D_F43.9&d=DwMFAw&c=G2MiLlal7SXE3PeSnG8W6_JBU6FcdVjSsBSbw6gcR0U&r=QGiOGwneOYKdMI_b-4g5Cita4ycB2h5Um83YWCmxPRE&m=lNvl5Zl-grDHBVMNAPJuB6TCOd8gxK_qWxqLvzNOeP0&s=HRhKWGB6Vwm_aHFPrqUOGlRKGk3dry-Pug3p61MwSaE&e=) *Reaction to severe stress, unspecified*

### FUMA Parameters

[jobinfo]

created_at = 2022-10-11 19:13:33

title = eur_ptsd_pcs_v4_aug3_2021.allchr.fuma.gz_Fix_commonmind

[version]

FUMA = v1.4.1

MAGMA = v1.08

GWAScatalog = e104_r2021-09-15

ANNOVAR = 2017-07-17

[inputfiles]

gwasfile = eur_ptsd_pcs_v4_aug3_2021.allchr.fuma.gz

chrcol = Chromosome

poscol = Position

rsIDcol = MarkerName

pcol = P-value

eacol = Allele1

neacol = Allele2

orcol = NA

becol = Zscore

secol = NA

leadSNPsfile = NA

addleadSNPs = 1

regionsfile = NA

[params]

N = NA

Ncol = Weight

exMHC = 1

MHCopt = annot

extMHC = NA

ensembl = v92

genetype = all

leadP = 5e-8

gwasP = 0.05

r2 = 0.6

r2_2 = 0.1

refpanel = 1KG/Phase3

pop = EUR

MAF = 0

refSNPs = 1

mergeDist = 250

[magma]

magma = 1

magma_window = 0

magma_exp = GTEx/v8/gtex_v8_ts_avg_log2TPM:GTEx/v8/gtex_v8_ts_general_avg_log2TPM

[posMap]

posMap = 1

posMapWindowSize = 10

posMapAnnot = NA

posMapCADDth = 0

posMapRDBth = NA

posMapChr15 = NA

posMapChr15Max = NA

posMapChr15Meth = NA

posMapAnnoDs = NA

posMapAnnoMeth = NA

[eqtlMap]

eqtlMap = 1

eqtlMaptss = eQTLcatalogue/BrainSeq_ge_brain.txt.gz:PsychENCODE/PsychENCODE_eQTLs.txt.gz:CMC/CMC_SVA_cis.txt.gz:CMC/CMC_SVA_trans.txt.gz:CMC/CMC_NoSVA_cis.txt.gz:CMC/CMC_NoSVA_trans.txt.gz:BRAINEAC/CRBL.txt.gz:BRAINEAC/FCTX.txt.gz:BRAINEAC/HIPP.txt.gz:BRAINEAC/MEDU.txt.gz:BRAINEAC/OCTX.txt.gz:BRAINEAC/PUTM.txt.gz:BRAINEAC/SNIG.txt.gz:BRAINEAC/TCTX.txt.gz:BRAINEAC/THAL.txt.gz:BRAINEAC/WHMT.txt.gz:BRAINEAC/aveALL.txt.gz:GTEx/v8/Brain_Amygdala.txt.gz:GTEx/v8/Brain_Anterior_cingulate_cortex_BA24.txt.gz:GTEx/v8/Brain_Caudate_basal_ganglia.txt.gz:GTEx/v8/Brain_Cerebellar_Hemisphere.txt.gz:GTEx/v8/Brain_Cerebellum.txt.gz:GTEx/v8/Brain_Cortex.txt.gz:GTEx/v8/Brain_Frontal_Cortex_BA9.txt.gz:GTEx/v8/Brain_Hippocampus.txt.gz:GTEx/v8/Brain_Hypothalamus.txt.gz:GTEx/v8/Brain_Nucleus_accumbens_basal_ganglia.txt.gz:GTEx/v8/Brain_Putamen_basal_ganglia.txt.gz:GTEx/v8/Brain_Spinal_cord_cervical_c-1.txt.gz:GTEx/v8/Brain_Substantia_nigra.txt.gz

eqtlMapSig = 1

eqtlMapP = 1

eqtlMapCADDth = 0

eqtlMapRDBth = NA

eqtlMapChr15 = NA

eqtlMapChr15Max = NA

eqtlMapChr15Meth = NA

eqtlMapAnnoDs = NA

eqtlMapAnnoMeth = NA

[ciMap]

ciMap = 1

ciMapBuiltin = EP/PsychENCODE/EP_links_oneway.txt.gz:HiC/PsychENCODE/Promoter_anchored_loops.txt.gz:HiC/Giusti-Rodriguez_et_al_2019/Adult_Cortex.txt.gz:HiC/Giusti-Rodriguez_et_al_2019/Fetal_Cortex.txt.gz:HiC/GSE87112/Dorsolateral_Prefrontal_Cortex.txt.gz:HiC/GSE87112/Hippocampus.txt.gz:HiC/GSE87112/Neural_Progenitor_Cell.txt.gz

ciMapFileN = 0

ciMapFiles = NA

ciMapFDR = 1e-6

ciMapPromWindow = 250-500

ciMapRoadmap = NA

ciMapEnhFilt = 0

ciMapPromFilt = 0

ciMapCADDth = 0

ciMapRDBth = NA

ciMapChr15 = NA

ciMapChr15Max = NA

ciMapChr15Meth = NA

ciMapAnnoDs = NA

ciMapAnnoMeth = NA

### FUMA cell type analysis datasets

The following datasets were used in the cell type analysis:

Allen_Human_LGN_level2

Allen_Human_MTG_level2

DroNc_Human_Hippocampus

GSE104276_Human_Prefrontal_cortex_per_ages

GSE104276_Human_Prefrontal_cortex_all_ages

Allen_Human_MTG_level1

Allen_Human_LGN_level1

GSE67835_Human_Cortex

GSE67835_Human_Cortex_woFetal

Linnarsson_GSE101601_Human_Temporal_cortex

Linnarsson_GSE76381_Human_Midbrain

PsychENCODE_Adult:PsychENCODE_Developmental

### Acknowledgements

**Army Study to Assess Risk and Resilience in Servicemembers (NSS1, NSS2, PPDS)( Supplementary Table 1 #14, Supplementary Table 1 #15, Supplementary Table 1 #16)**

Funding:

Army STARRS was sponsored by the Department of the Army and funded under cooperative agreement number U01MH087981 (2009-2015) with the National Institutes of Health, National Institute of Mental Health (NIH/NIMH). Subsequently, STARRS-LS was sponsored and funded by the Department of Defense (USUHS grant number HU0001-15-2-0004). The contents are solely the responsibility of the authors and do not necessarily represent the views of the Department of Health and Human Services, NIMH, the Department of the Army, or the Department of Defense.

The Army STARRS Team consists of:

Co-Principal Investigators: Robert J. Ursano, MD (Uniformed Services University of the Health Sciences) and Murray B. Stein, MD, MPH (University of California San Diego and VA San Diego Healthcare System)

Site Principal Investigators: Steven Heeringa, PhD (University of Michigan), James Wagner, PhD (University of Michigan) and Ronald C. Kessler, PhD (Harvard Medical School)

Army liaison/consultant: Kenneth Cox, MD, MPH (US Army Public Health Center)

Other team members: Pablo A. Aliaga, MS (Uniformed Services University of the Health Sciences); COL David M. Benedek, MD (Uniformed Services University of the Health Sciences); Susan Borja, PhD (NIMH); Tianxi Cai, ScD (Harvard School of Public Health); Laura Campbell-Sills, PhD (University of California San Diego); Chia-Yen Chen, ScD (Harvard Medical School); Carol S. Fullerton, PhD (Uniformed Services University of the Health Sciences); Nancy Gebler, MA (University of Michigan); Joel Gelernter, MD (Yale University); Robert K. Gifford, PhD (Uniformed Services University of the Health Sciences); Feng He, MS (University of California San Diego); Paul E. Hurwitz, MPH (Uniformed Services University of the Health Sciences); Sonia Jain, PhD (University of California San Diego); Kevin Jensen, PhD (Yale University); Kristen Jepsen, PhD (University of California San Diego); Tzu-Cheg Kao, PhD (Uniformed Services University of the Health Sciences); Lisa Lewandowski-Romps, PhD (University of Michigan); Holly Herberman Mash, PhD (Uniformed Services University of the Health Sciences); James E. McCarroll, PhD, MPH (Uniformed Services University of the Health Sciences); Adam X. Maihofer (University of California San Diego); Colter Mitchell, PhD (University of Michigan); James A. Naifeh, PhD (Uniformed Services University of the Health Sciences); Tsz Hin Hinz Ng, MPH (Uniformed Services University of the Health Sciences); Caroline M. Nievergelt, PhD (University of California San Diego); Matthew K. Nock, PhD (Harvard University); Stephan Ripke, MD (Harvard Medical School); Nancy A. Sampson, BA (Harvard Medical School); CDR Patcho Santiago, MD, MPH (Uniformed Services University of the Health Sciences); Ronen Segman, MD (Hadassah University Hospital, Israel); Jordan W. Smoller, MD, ScD (Harvard Medical School); Xiaoying Sun, MS (University of California San Diego); Erin Ware PhD (University of Michigan); LTC Gary H. Wynn, MD (Uniformed Services University of the Health Sciences); Alan M. Zaslavsky, PhD (Harvard Medical School); and Lei Zhang, MD (Uniformed Services University of the Health Sciences).

**Ash Wednesday and IVS (BRYA)(Supplementary Table 1 #10)**

This project was funded by a National Health and Medical Research Council Grant (1073041).

**AURORA (AURO; Supplementary Table 1 #73)**

This work was supported by National Institute of Diabetes and Digestive and Kidney Diseases (K01AR071504 to S.D.L.) and National Institute of Mental Health (R01AR060852 to S.A.M.).

**Biomarkers and Experiences in Adolescent Relationships (BRLS; Supplementary Table 1 #64)**

This project was supported by National Institute of Mental Health (R01MH105379 to N.R.N.).

**Bounce Back Now (BOBA)(Supplementary Table 1 #18)**

This work was supported by 1R01MH081056 (PI: Ruggiero), 1R01MH081056-S1 (PI: Amstadter), as well as K02 AA023239 (PI: Amstadter) and K01 AA025692 (PI: Sheerin).

**Canadian Longitudinal Study on Aging (CANA; Supplementary Table 1 #81)**

This study was supported by the Brain and Behavior Research Foundation, Larry and Judy Tanenbaum Family Foundation, and CAMH Foundation.

**Childhood Trauma Study (QIMR) (Supplementary Table 1 #30)**

This work was primarily supported by National Institute of Health grants to ECN (AA13446; AA011998_5978). Additional support includes grants to ACH (AA10249, AA07728, AA11998, AA13321), NGM (AA13326), PAFM (DA12854; DA027995).

**Child Trauma and Neural Systems Underlying Emotion Regulation (KMCT, KMC2; Supplementary Table 1 #19, #61)**

This work was funded by the R01-MH103291 (PI: McLaughlin), R01-MH103291-S1 (PI: McLaughlin), and R01-MH103291-S2 (PI: McLaughlin).

**Cohen Veterans Center Study and Fort Campbell study (COM1, FTCB)(Supplementary Table 1 #50, Supplementary Table 1 #52)**

These studies were supported by the Steve and Alexandra Cohen Foundation and the Department of Defense (DoD: W81XWH-09-2-0044 to C.R.M. and W911NF-09-1-0298 to R.Y.).

**Collaborative Study on the Genetics of Alcoholism (RCOG; Supplementary Table 1 #83)**

This study was supported by the National Institute on Alcohol Abuse and Alcoholism (U10AA008401 to H.E., V.H., J.M., B.P.).

**Cortical Excitability: Biomarker and Endophenotype in Combat Related PTSD (WANG)( Supplementary Table 1 #59)**

This work was supported by VA Merit Review awards 1I21RX001725-01, 1I01CX000487-01A1, and 1R34MH078854-01.

**Danish military study** **(DAMI)(Supplementary Table 1 #28)**

This research has been conducted using the Danish National Biobank resource, supported by the Novo Nordisk Foundation. The study was supported by the Research and Knowledge Centre, The Danish Veteran Centre and funded by the Danish Ministry of Defence as part of the 3rd September 2014 agreement on strengthened initiatives for Danish veterans.

**Danish iPSYCH PTSD samples (DAIP)(Supplementary Table 1 #29)**

The iPSYCH team acknowledges funding by the Lundbeck Foundation (grant numbers R102-A9118 and R155-2014- 1724) and the universities and university hospitals of Aarhus and Copenhagen. The Danish National Biobank resource was supported by the Novo Nordisk Foundation. Data handling and analysis on the GenomeDK HPC facility was supported by NIMH (1U01MH109514-01 to Michael O’Donovan and ADB). High-performance computer capacity for handling and statistical analysis of iPSYCH data on the GenomeDK HPC facility was provided by the Centre for Integrative Sequencing, iSEQ, Aarhus University, Denmark (grant to ADB).

**Danish iPSYCH PTSD samples (DAI2; Supplementary Table 1 #85)**

The iPSYCH team was supported by grants from the Lundbeck Foundation (R102-A9118, R155-2014-1724, and R248-2017-2003), NIH/NIMH (1U01MH109514-01 and 1R01MH124851-01 to A.D.B.) and the Universities and University Hospitals of Aarhus and Copenhagen. The Danish National Biobank resource was supported by the Novo Nordisk Foundation. High-performance computer capacity for handling and statistical analysis of iPSYCH data on the GenomeDK HPC facility was provided by the Center for Genomics and Personalized Medicine and the Centre for Integrative Sequencing, iSEQ, Aarhus University, Denmark (grant to A.D.B.).

**DCS Rothbaum Study (DCSR)(Supplementary Table 1 #38)**

For support, this study was funded by by NIMH grant R01 MH-70880 to Dr. Rothbaum, Clinicaltrials.gov identiﬁer: NCT00356278**.**

**Defining Essential Features of Neural Damage (DEFE)(Supplementary Table 1 #12)**

Congressionally Directed Medical Research Programs, W81XWH-08–2–0038

Department of Veteran Affairs Rehabilitation Research and Development Service, 1IK1RX002325, I01RX000622

**Detroit Neighborhood Health Study (DNHS, ADNH)(Supplementary Table 1 #4, Supplementary Table 1 #45)**

DNHS was funded by NIH Awards R01DA022720, R01DA022720-S1, and RC1MH088283 to Allison E. Aiello; and R01MD011728 to Monica Uddin. We are grateful to all of the participants and staff for their contributions to the DNHS.

**Drakenstein Child Health Study - South African sample (SAFR, SAF2; Supplementary Table 1 #3, #77)**

Research reported in this publication was supported by the South African Medical Research Council (SAMRC) Unit on Risk & Resilience in Mental Disorders and a Self-Initiated Research Grant (NK).  The views and opinions expressed are those of the authors and do not necessarily represent the official views of the SAMRC.

**EA CRASH (EACR)(Supplementary Table 1 #42)**

EA CRASH was supported by the National Institute of Arthritis and Musculoskeletal and Skin Diseases of the National Institutes of Health under Award Number R01-AR056328. The content is solely the responsibility of the authors and does not necessarily represent the views of this funding agency.

**Emory Healthcare Veteran’s Program (EHVP; Supplementary Table 1 #74)**

This work is supported by the Wounded Warrior Project and Multidisciplinary Association of Psychedelics Studies.

**Estonian Biobank (ESBB; Supplementary Table 1 #93)**

The study is supported by the Estonian Research Council (PSG615 to K.L.).

**Family Study of Cocaine Dependence and Collaborative Genetic Study of Nicotine Dependence (FSCD, COGA, COGB)(Supplementary Table 1 #7, Supplementary Table 1 #8, Supplementary Table 1 #9)**

The Collaborative Genetic Study of Nicotine Dependence (COGEND) was supported by National Cancer Institute grant P01CA089392 to Laura Bierut. The Family Study of Cocaine Dependence (FSCD) was supported by National Institute on Drug Abuse grants R01DA013423 and R01DA019963 to Laura Bierut. Funding support for genotyping was provided by the NIH GEI (U01HG004438), the National Institute on Alcohol Abuse and Alcoholism, the National Institute on Drug Abuse, and the NIH contract "High throughput genotyping for studying the genetic contributions to human disease" (HHSN268200782096C).

**FinnGen (FING; Supplementary Table 1 #90)**

We want to acknowledge the participants and investigators of FinnGen study. The FinnGen project is funded by two grants from Business Finland (HUS 4685/31/2016 and UH 4386/31/2016) and the following industry partners: AbbVie Inc., AstraZeneca UK Ltd, Biogen MA Inc., Bristol Myers Squibb (and Celgene Corporation & Celgene International II Sàrl), Genentech Inc., Merck Sharp & Dohme LCC, Pfizer Inc., GlaxoSmithKline Intellectual Property Development Ltd., Sanofi US Services Inc., Maze Therapeutics Inc., Janssen Biotech Inc, Novartis AG, and Boehringer Ingelheim International GmbH. Following biobanks are acknowledged for delivering biobank samples to FinnGen: Auria Biobank (www.auria.fi/biopankki), THL Biobank (www.thl.fi/biobank), Helsinki Biobank (www.helsinginbiopankki.fi), Biobank Borealis of Northern Finland (https://www.ppshp.fi/Tutkimus-ja-opetus/Biopankki/Pages/Biobank-Borealis-briefly-in-English.aspx), Finnish Clinical Biobank Tampere (www.tays.fi/en-US/Research_and_development/Finnish_Clinical_Biobank_Tampere), Biobank of Eastern Finland (www.ita-suomenbiopankki.fi/en), Central Finland Biobank (www.ksshp.fi/fi-FI/Potilaalle/Biopankki), Finnish Red Cross Blood Service Biobank (www.veripalvelu.fi/verenluovutus/biopankkitoiminta), Terveystalo Biobank (www.terveystalo.com/fi/Yritystietoa/Terveystalo-Biopankki/Biopankki/) and Arctic Biobank (https://www.oulu.fi/en/university/faculties-and-units/faculty-medicine/northern-finland-birth-cohorts-and-arctic-biobank). All Finnish Biobanks are members of BBMRI.fi infrastructure (www.bbmri.fi). Finnish Biobank Cooperative -FINBB (https://finbb.fi/) is the coordinator of BBMRI-ERIC operations in Finland. The Finnish biobank data can be accessed through the Fingenious® services (https://site.fingenious.fi/en/) managed by FINBB.

Patients and control subjects in FinnGen provided informed consent for biobank research, based on the Finnish Biobank Act. Alternatively, separate research cohorts, collected prior the Finnish Biobank Act came into effect (in September 2013) and start of FinnGen (August 2017), were collected based on study-specific consents and later transferred to the Finnish biobanks after approval by Fimea, the National Supervisory Authority for Welfare and Health. Recruitment protocols followed the biobank protocols approved by Fimea. The Coordinating Ethics Committee of the Hospital District of Helsinki and Uusimaa (HUS) approved the FinnGen study protocol Nr HUS/990/2017.

The FinnGen study is approved by Finnish Institute for Health and Welfare (permit numbers: THL/2031/6.02.00/2017, THL/1101/5.05.00/2017, THL/341/6.02.00/2018, THL/2222/6.02.00/2018, THL/283/6.02.00/2019, THL/1721/5.05.00/2019, THL/1524/5.05.00/2020, and THL/2364/14.02/2020), Digital and population data service agency (permit numbers: VRK43431/2017-3, VRK/6909/2018-3, VRK/4415/2019-3), the Social Insurance Institution (permit numbers: KELA 58/522/2017, KELA 131/522/2018, KELA 70/522/2019, KELA 98/522/2019, KELA 138/522/2019, KELA 2/522/2020, KELA 16/522/2020 and Statistics Finland (permit numbers: TK-53-1041-17 and TK-53-90-20).

The Biobank Access Decisions for FinnGen samples and data utilized in FinnGen Data Freeze 6 include: THL Biobank BB2017_55, BB2017_111, BB2018_19, BB_2018_34, BB_2018_67, BB2018_71, BB2019_7, BB2019_8, BB2019_26, BB2020_1, Finnish Red Cross Blood Service Biobank 7.12.2017, Helsinki Biobank HUS/359/2017, Auria Biobank AB17-5154, Biobank Borealis of Northern Finland_2017_1013, Biobank of Eastern Finland 1186/2018, Finnish Clinical Biobank Tampere MH0004, Central Finland Biobank 1-2017, and Terveystalo Biobank STB 2018001.

**GMRF-QUT (GMFR)(Supplementary Table 1 #55)**

The Queensland Branch of the Returned and Services League of Australia (RSL) funded the PTSD Initiative at the Gallipoli Medical Research Institute. The Australian Government Department of Veterans’ Affairs provided transport for eligible participants. We gratefully acknowledge the dedicated efforts of the participants and their families, and the clinical and support staff involved in data collection. Specifically, we would like to thank the members of the PTSD Initiative team: Sarah McLeay, PhD, Wendy Harvey, MPH, Madeline Romaniuk, DPsych(Clin), Darrell Crawford, MD, David Colquhoun, MBBS, Ross McD Young, PhD, Miriam Dwyer, BSc, John Gibson, MBBS, Robyn O’Sullivan, MBBS, Graham Cooksley, MBBS, Christopher Strakosch, MD, Rachel Thomson, PhD, Joanne Voisey, PhD, Bruce Lawford, MBBS. We would also like to thank Emile Touma and Vikram Goel for performing psychiatric assessments, Terence Harvey for developing the study database and QUT for financial support.

**Grady Trauma Project (EGHS, GTPC)(Supplementary Table 1 #44, Supplementary Table 1 #47)**

This study was supported by the National Institutes of Health, MH071537 and MH096764.

**Grady Trauma Project – Neuroimaging (DGTP; Supplementary Table 1 #68)**

This work is supported by National Institute of Health (AT011267 to N.F.) and National institute of Mental Health (MH120299 to N.F.; MH111671-01A1 to N.F.; R01MH100122 to T.J.; R01MH111682 to T.J.).

**Injury and Traumatic Stress Consortium (INTR)(Supplementary Table 1 #27)**

The PTSD and TBI INjury and TRaUmatic STress Clinical Consortium (INTRuST) was funded by a grant from the United States Department of Defense (PI: Stein, MB): W81XWH08-2-0159. Members of the INTRuST Consortium Biorepository Working Group who contributed to this work include: Gerald A. Grant MD, Christine E. Marx MD, Mark S. George MD, Thomas W. McAllister MD, Norberto Andaluz MD, Lori Shutter MD, Raul Comibra MD, Ross D. Zafonte DO, Sonia Jain PhD, Xue-Jun Qin, and Michael Hauser PhD.

**Marine Resiliency Study (MRSC, BAKE)(Supplementary Table 1 #1, Supplementary Table 1 #57)**

The Marine Corps, Navy Bureau of Medicine and Surgery (BUMED) and VA Health Research and Development (HSR&D) provided funding for MRS data collection and analysis (PI DGB) and NIH R01MH093500 funded the GWAS assays and analysis (PI CMN). The study was supported by the Veterans Administration Center of Excellence for Stress and Mental Health (CESAMH). Acknowledged are Victoria B. Risbrough Ph.D (VA San Diego Healthcare System & UCSD), Mark A. Geyer (UCSD), Daniel T. O’Connor (UCSD), all MRS investigators, as well as the MRS administrative core and data collection staff. The authors also thank the Marine and Navy Corpsmen volunteers for their military service and participation in MRS.

**MayoGC (MAYO; Supplementary Table 1 #94)**

The study was supported by the National Institute of Mental Health (R01 MH121924 to J.M.B.).

**McLean Trauma Sample (TEIC)(Supplementary Table 1 #39)**

The Kaufman lab would like to thank the participants for making this research possible; the staff of the Hill Center for Women and Proctor House II, McLean Hospital; The work was supported by National Institute of Mental Health (NIMH) grant R21MH112956 to MLK, and NIMH fellowship grant F32MH109274 to LAML, the Anonymous Women’s Health Fund to MLK, the O’Keefe Family Foundation to MLK, the Trauma Scholars Fund to MLK, and the Frazier Foundation Grant for Mood and Anxiety Research to KJR.

The Teicher Lab would like to thank all the participants for being a part of these studies and the staff of Developmental Biopsychiatry Research Program, McLean Hospital. The studies were supported by National Institute of Mental Health (NIMH) grant RO1 MH91391 and National Institute on Drug Abuse R01 DA17846 to MHT.

**Mexican Adolescent Mental Health Study (MAMH; Supplementary Table 1 #65)**

This study was supported by CONACYT-SEP-SSEDF-2003 CO1-22 to C. Benjet and SEP-2004-CO1-46594/A-1 to C.S.C.

**Mid-Atlantic Mental Illness Research Education and Clinical Center the study of Post-Deployment Mental Health Study (MIRE)(Supplementary Table 1 #26)**

Preparation of this manuscript was supported by a Clinical Sciences Research and Development (CSR&D) Research Career Scientist Award (#11S-RCS-009) to Dr. Beckham, a CSR&D Career Development Award (#IK2 CX000525) to Dr. Kimbrel, and Biomedical and Laboratory Research and Development (BLR&D) Merit Award to Dr. Beckham from the U.S. Department of Veterans Affairs (VA). This work was also supported by the VA Mid-Atlantic Mental Illness Research, Education and Clinical Center (MIRECC), the Durham Veterans Affairs Medical Center, the VA Office of Mental Health Services, and the VA Office of Research and Development. The Mid-Atlantic MIRECC Workgroup contributors for this paper include: Mira Brancu, PhD, Patrick S. Calhoun, PhD, Eric Dedert, PhD, Eric B. Elbogen, PhD, John A. Fairbank, PhD, Robin A. Hurley, MD, Jason D. Kilts, PhD, Angela Kirby, MS, Christine E. Marx, MD, MS, Scott D. McDonald, PhD, Scott D. Moore, MD, PhD, Rajendra A. Morey, MD, MS, Jennifer C. Naylor, PhD, Treven C. Pickett, PsyD, Jared Rowland, PhD, Cindy Swinkels, PhD, Steven T. Szabo, MD, PhD, Katherine H. Taber, PhD., Larry A. Tupler, PhD, Elizabeth E. Van Voorhees, PhD, H. Ryan Wagner, Ph.D., Ruth E. Yoash-Gantz, PsyD. Dedert is funded by a Department of Veterans Affairs Clinical Science Research and Development Career Development Award (IK2CX000718). Naylor is funded by a Department of Veterans Affairs Rehabilitation Research and Development Career Development Award (1lK2RX000908). Van Voorhees is funded by a Department of Veterans Affairs Rehabilitation Research and Development Career Development Award (1K2RX001298). The views expressed in this article are those of the authors and do not necessarily reflect the position or policy of the Department of Veterans Affairs or the United States government.

**The Million Veteran Program (MVP1; Supplementary Table 1 #79)**

This work was also supported by funding from the Department of Veterans Affairs Office of Research and Development grants I01CX001849, MVP025 to M.B.S. and J.G. D.F.L. was supported by a Career Development Award CDA-2 from the Veterans Affairs Office of Research and Development (1IK2BX005058-01A2).

**Mount Sinai BioMe PTSD (BIOM; Supplementary Table 1 #92)**

This study was supported by the National Institute of Mental Health (R01 MH124839 to L.M.H.; R01 MH118278 to L.M.H.) and National Institute of Environmental Health Sciences (R01 ES033630 to L.M.H.).

**National Centre for Mental Health (NCMH)(Supplementary Table 1 #41)**

This project was supported by the National Centre for Mental Health (NCMH). NCMH is funded by Welsh Government through Health and Care Research Wales.

**National Health and Resilience in Veterans Study (NHRV; Supplementary Data 1 #13)**

The National Health and Resilience in Veterans Study is supported by the U.S. Department of Veterans Affairs National Center for Posttraumatic Stress Disorder.

**NIU Trauma Orcutt (NIUT)(Supplementary Table 1 #40)**

This study was supported by the Joyce Foundation (Dr Orcutt) and NIH, HD049907and MH085436.

**Nurses Health Study II (NHS2, NHSY)(Supplementary Table 1 #5)(Supplementary Table 1 #22)**

NHSII PTSD Sub-Study was funded by National Institute on Mental Health awards RO1 MH093612, MH078928 to Karestan C Koenen. The NHSII cohort is funded in part by UM1 CA176726.

**Ohio National Guard (ONGA)(Supplementary Table 1 #2)**

The funding information for the Ohio Army National Guard cohort is:

Dept. of Army, Telecommunication and Advanced Technology Research Center (TATRC) Award #s W81XWH-15-1-0080 and W81XWH-10-1-0579 “Ohio Army National Guard Mental Health Initiative: Genetics of Risk and Resilience for Deployment-Related Stress Disorders”.

**Ohio National Guard 2 – New Samples (ONGB; Supplementary Table 1 #76)**

This work is supported by the National Heart, Lung, and Blood Institute (T32HL098048 to L.S.).

**OPT and CHOICE (FEEN)(Supplementary Table 1 #37)**

This research is funded by the National Institute of Mental Health (NIMH; R01MH066347, R01MH066348) and the William T. Dahms, MD, Clinical Research Unit, funded under the Cleveland Clinical and Translational Science Award (UL1 RR024989).

Pfizer Inc. supplied the medication at no cost but had no input in the trial development, conduct, analysis, or interpretation. Drs. Zoellner, Roy-Byrne, Mavissakalian, and Feeny have no competing interests to disclose.

We would like to thank all participants, therapists, and psychiatrists involved in the study and acknowledge the vital contributions of study researchers and administrators in Seattle, Washington and Cleveland, Ohio. Specifically, we would like to thank the investigative team on the grants: Jason Doctor, Ph.D., Joshua McDavid, MD, Alice S. Friedman, MSN, ARNP, and Nora McNamara, MD. Afsoon Eftekhari, Ph.D., and Lisa Stines Doane, Ph.D. were integral in the implementation of this study. Edna Foa, Ph.D. and her team provided PE integrity ratings. We would like to acknowledge Susan Silva, Ph.D., Eric Youngstrom, Ph.D., Kevin King, Ph.D., and Andrew A. Cooper, Ph.D. for their statistical consultation and analyses.

**Pregnancy Outcomes, Maternal and Infant Cohort Study (PROM)(Supplementary Table 1 #23)**

This research was supported by awards from the Eunice Kennedy Shriver Institute of Child Health and Human Development (R01-HD-059835 and R01 HD059827).

**QIMR Berghofer Medical Research Institute (QIMRB) (AGDS, QIM2; Supplementary Table 1 #80, Supplementary Table 1 #82)**

Support for these studies came from National Health and Medical Research Council (APP2016346 to I.B.H.; APP1172917 to S.E.M.). PISA (Prospective Imaging Study of Aging: Genes, Brain and Behaviour) is funded by the National Health and Medical Research Council (NHMRC Grant ID: APP1095227).

**Readiness and Resilience in National Guard Soldiers (RING)(Supplementary Table 1 #33)**

Support for this study came from the Center for Veterans Research and Education.

**Risbrough/Norman randomized controlled psychotherapy trial (VRIS)(Supplementary Table 1 #58)**

Samples and data collection were funded by VA Office of Clinical Science Research and Development (5IO1CX000756 to SBN) and the VA Office of Basic Science Research and Development (1I01BX002558 to VR).  Additional salary support was funded by the VA National Center for PTSD (Norman) and VA Center of Excellence for Stress and Mental Health (Risbrough).

**Shared Roots (SHRS)(Supplementary Table 1 #46)**

The Shared Roots project is supported by the South African Medical Research Council for the Shared Roots Flagship Project, Grant no. MRC-RFA-IFSP-01-2013/SHARED ROOTS through funding received from the South African National Treasury under its Economic Competitiveness and Support Package. Its contents are solely the responsibility of the authors and do not necessarily represent the official views of the South African Medical Research Council. Additional funding was received from the South African Research Chairs Initiative of the South African Department of Science and Technology and National Research Foundation.

**Southeastern Europe PTSD (SEEP)(Supplementary Table 1 #49)**

Recruitment of the SEE-PTSD cohort was funded by the DAAD.

**Stony Brook University World Trade Center Health Program (WTCS; Supplementary Table 1 #78)**

This work is supported by National Institute of Health (U01OH011864 to M.W., R.K., and B.L.) and National Institute of Mental Health (R01MH123619 to A.D.; R01AG049953 to S.A.P.C.).

**Stress Risk and Resilience in Syrian and Iraqi Refugees and Survivors of Torture (SYIR; Supplementary Table 1 #63)**

This study was supported by National Institute of Mental Health (F31MH120927; PI: L.G.) and the State of Michigan Lycaki/Young Foundation (PI: A.J.).

**STRONG STAR Genetic and Environmental Predictors of Combat-Related PTSD (STRO) (Supplementary Table 1 #35)**

Funding for the STRONG STAR sample collection was made possible by the U.S. Department of Defense through the U.S. Army Medical Research and Materiel Command, Congressionally Directed Medical Research Programs, Psychological Health and Traumatic Brain Injury Research Program awards W81XWH-08-02-109 (Alan Peterson) and W81XWH-08-02-0110 (Douglas Williamson), and W81XWH-08-02-0114 (Brett Litz).

**Study of Aftereffects of Trauma: Understanding Response in National Guard (SATU)(Supplementary Table 1 #11)**

Congressionally Directed Medical Research Programs, W81XWH-08–2–0038

Department of Veteran Affairs Rehabilitation Research and Development Service, 1IK1RX002325, I01RX000622

**Study on Trauma & Resilience (STAR Study) (RCSS; Supplementary Table 1 #70)**

This study was supported by National Institute of Health (R21 MH 102838-01 A1 to T.d-C.).

**The Study of Twin Adults: Genes and Environment (STR-STAGE), from the Swedish Twin Registry (SWED; Supplementary Table 1 #89)**

The Swedish Twin Registry is managed by Karolinska Institutet as a core facility and receives funding through the Swedish Research Council under the grant 2021-00180. This study is supported by the European Research Council (Stress Gene, Grant Agreement ID 726413; Grant Agreement ID 101042183) and the National Institute of Mental Health (R01 MH123724).

**Sydney Neuroimaging (BRY2)(Supplementary Table 1 #36)**

This project was funded by a National Health and Medical Research Council Grant (1073041).

**Trøndelag Health Study (HUNT)(Supplementary Table 1 #88)**

The Trøndelag Health Study (HUNT) is a collaboration between HUNT Research Centre (Faculty of Medicine and Health Sciences, Norwegian University of Science and Technology NTNU), Trøndelag County Council, Central Norway Regional Health Authority, and the Norwegian Institute of Public Health. The genotyping was financed by the National Institute of health (NIH), University of Michigan, The Norwegian Research council, and Central Norway Regional Health Authority and the Faculty of Medicine and Health Sciences, Norwegian University of Science and Technology (NTNU). The genotype quality control and imputation has been conducted by the K.G. Jebsen center for genetic epidemiology, Department of public health and nursing, Faculty of medicine and health sciences, Norwegian University of Science and Technology (NTNU).

**UK Biobank (UKBB)(Supplementary Table 1 #60)**

This research has been conducted using the UK Biobank resource (Application No. 41209). This study represents independent research part funded by the National Institute for Health Research (NIHR) Biomedical Research Centre at South London and Maudsley NHS Foundation Trust and King’s College London. The views expressed are those of the authors and not necessarily those of the NHS, the NIHR or the Department of Health and Social Care. High performance computing facilities were funded with capital equipment grants from the GSTT Charity (TR130505) and Maudsley Charity (980). GB and JRIC acknowledge funding from Cohen Veterans Bioscience.

**UK Biobank – EHR Cohort Description (UKB2; Supplementary Table 1 #91)**

This paper represents independent research part funded by the NIHR Maudsley Biomedical Research Centre at South London and Maudsley NHS Foundation Trust and King’s College London. The views expressed are those of the authors and not necessarily those of the NIHR or the UK Department of Health and Social Care.

**VA Boston-National Center for PTSD Study (NCPT, TRACT)(Supplementary Table 1 #31, Supplementary Table 1 #32)**

This research was supported in part by National Institute of Mental Health Award RO1MH079806 (MWM), Department of Veterans Affairs, Clinical Science Research & Development Program Award 5I01CX000431-02 (MWM), Department of Veterans Affairs, Biomedical Laboratory Research & Development Program Award 1I01BX002150-01 (MWM), The Translational Research Center for TBI and Stress Disorders (TRACTS), A VA Traumatic Brain Injury National Network Rehabilitation Research and Development Center award B9254-C (R.E.M. and W.P.M.), Department of Veterans Affairs. This research is the result of work supported with resources and the use of facilities at the Pharmacogenomics Analysis Laboratory, Research and Development Service, Central Arkansas Veterans Healthcare System, Little Rock, Arkansas. This work was also supported by a Career Development Award to E. J. Wolf from the Department of Veterans Affairs, Clinical Sciences Research, and Development Program.

**Vietnam Era Twin Study of Aging (VETS) (Supplementary Table 1 #24)**

This research was supported by National Institute on Aging R01 AG018386, AG022982, AG050595 (W.S.K.), R01 AG018384 (M.J.L.), R03 AG046413 (C.E.F), and K08 AG047903 (M.S.P), and the VA San Diego Center of Excellence for Stress and Mental Health Healthcare System. The content is the responsibility of the authors and does not necessarily represent official views of the NIA, NIH, or VA. The Cooperative Studies Program of the U.S. Department of Veterans Affairs provided financial support for development and maintenance of the Vietnam Era Twin Registry. We would also like to acknowledge the continued cooperation and participation of the members of the VET Registry and their families.

**The Women and Children’s Health Study (WACH)(Supplementary Table 1 #43)**

Funding: This study was primarily supported by the National Institute of Environmental Health Sciences (grant 1U01ES021497)

**World Trade Center: Mount Sinai (WTCM; Supplementary Table 1 #84)**

This study was supported by the National Institute for Occupational Safety and Health (U01 OH010407 to A.F. and R.H.P.; U01 OH010986 to A.F.).

**Yale-Penn Study (GSDC)(Supplementary Table 1 #6)**

This study was supported by National Institutes of Health Grants RC2 DA028909, R01 DA12690, R01 DA12849, R01 DA18432, R01 AA11330, and R01 AA017535 and the Veterans Affairs VISN 1 and VISN 4 Mental Illness Research, Educational, and Clinical Centers; and the VA National Center for PTSD Research.

Genotyping services for a part of our genome-wide association study were provided by the Center for Inherited Disease Research and the Yale Center for Genome Analysis. Center for Inherited Disease Research is fully funded through a Federal contract from the National Institutes of Health to The Johns Hopkins University (contract number N01-HG-65403).

**Million Veteran Program (MVP) (replication cohort)**

This research includes data from the Million Veteran Program (MVP), Office of Research and Development, Veterans Health Administration, and was supported by MVP and the VA Cooperative Studies Program (CSP) study #575B.

### Supplementary Figures

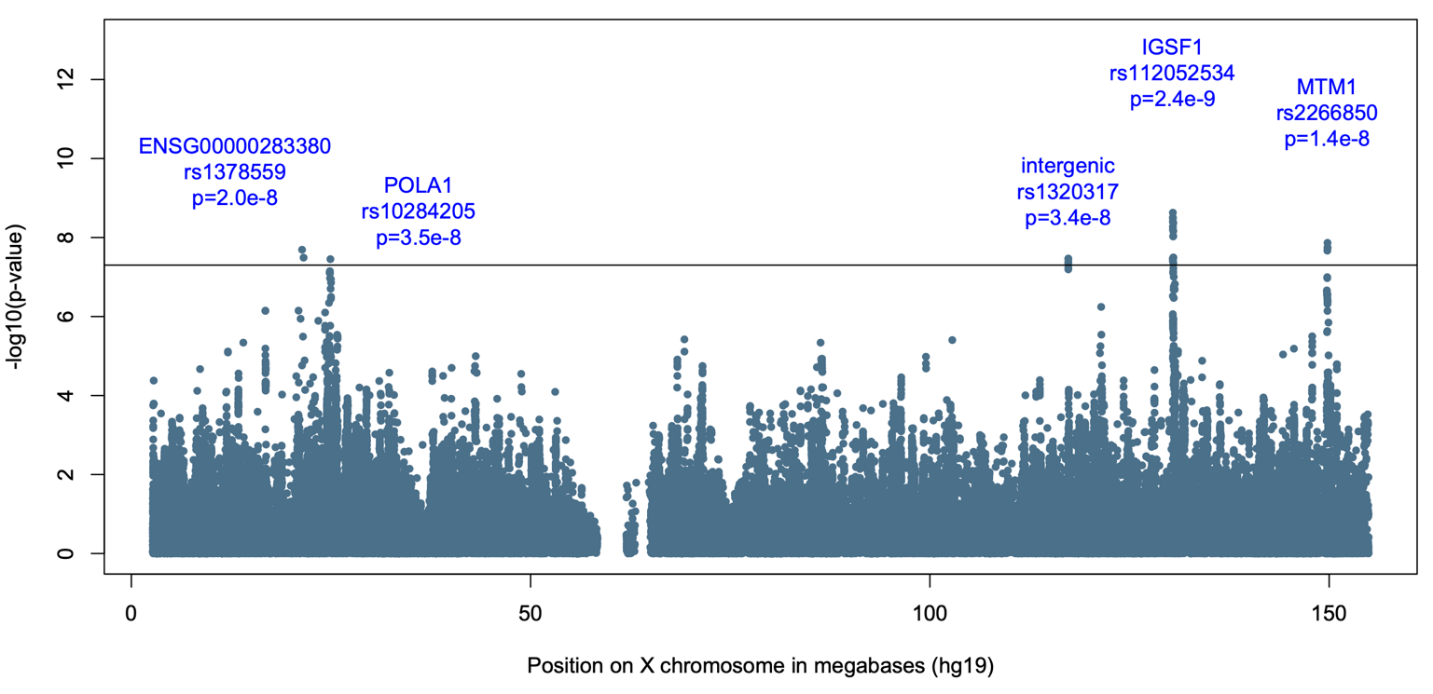

**Supplementary Figure 1: Manhattan plots of EA GWAS results on the X chromosome.**

The *x axis* refers to the position on the genome, ordered by chromosome and base-pair position. The *y axis* refers to the −log10 p-value of association from GWAS. Each circle represents the association between a given single nucleotide polymorphism and PTSD. Circle colors alternate between chromosomes: even chromosomes colored blue and odd chromosomes colored black. The horizontal red bar indicates genome-wide significance (p < 5x10^-8^). Names of genes where significant SNPs reside are labeled.

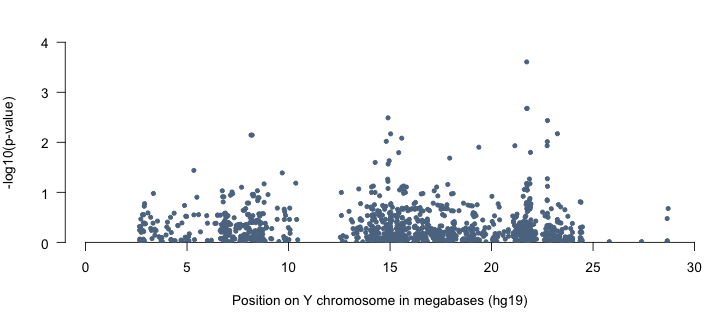

**Supplementary Figure 2: Manhattan plots of EA GWAS results on the Y chromosome.**

The *x axis* refers to the position on the genome, ordered by chromosome and base-pair position. The *y axis* refers to the −log10 p-value of association from GWAS. Each circle represents the association between a given single nucleotide polymorphism and PTSD. Circle colors alternate between chromosomes: even chromosomes colored blue and odd chromosomes colored black. The horizontal red bar indicates genome-wide significance (p < 5x10^-8^).

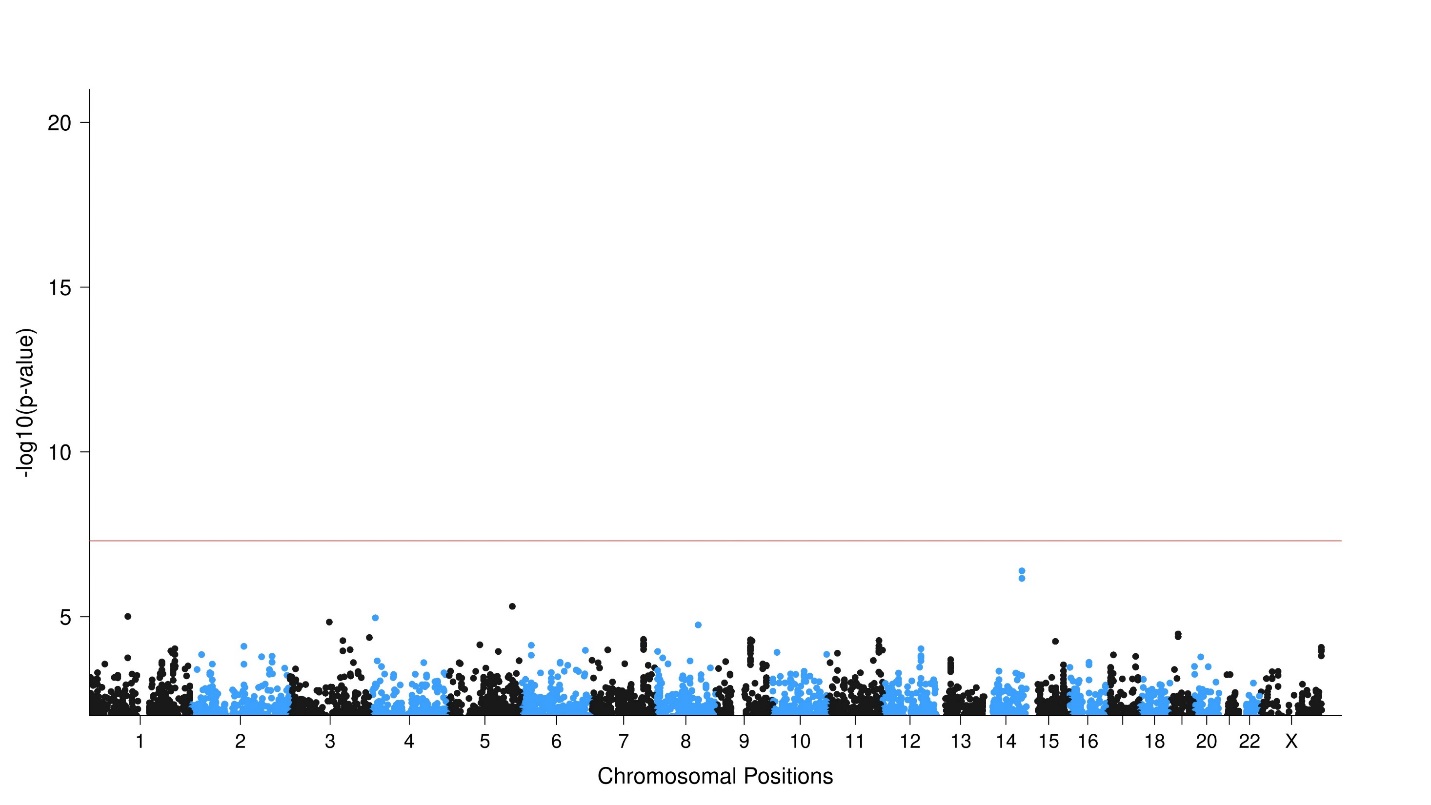

**Supplementary Figure 3: Manhattan plots of EA GWAS heterogeneity test results.**

The *x axis* refers to the position on the genome, ordered by chromosome and base-pair position. The *y axis* refers to the −log10 p-value of the heterogeneity test association from meta-analysis. Each circle represents the association between a given single nucleotide polymorphism and PTSD. Circle colors alternate between chromosomes: even chromosomes colored blue and odd chromosomes colored black. The horizontal red bar indicates genome-wide significance (p < 5x10^-8^).

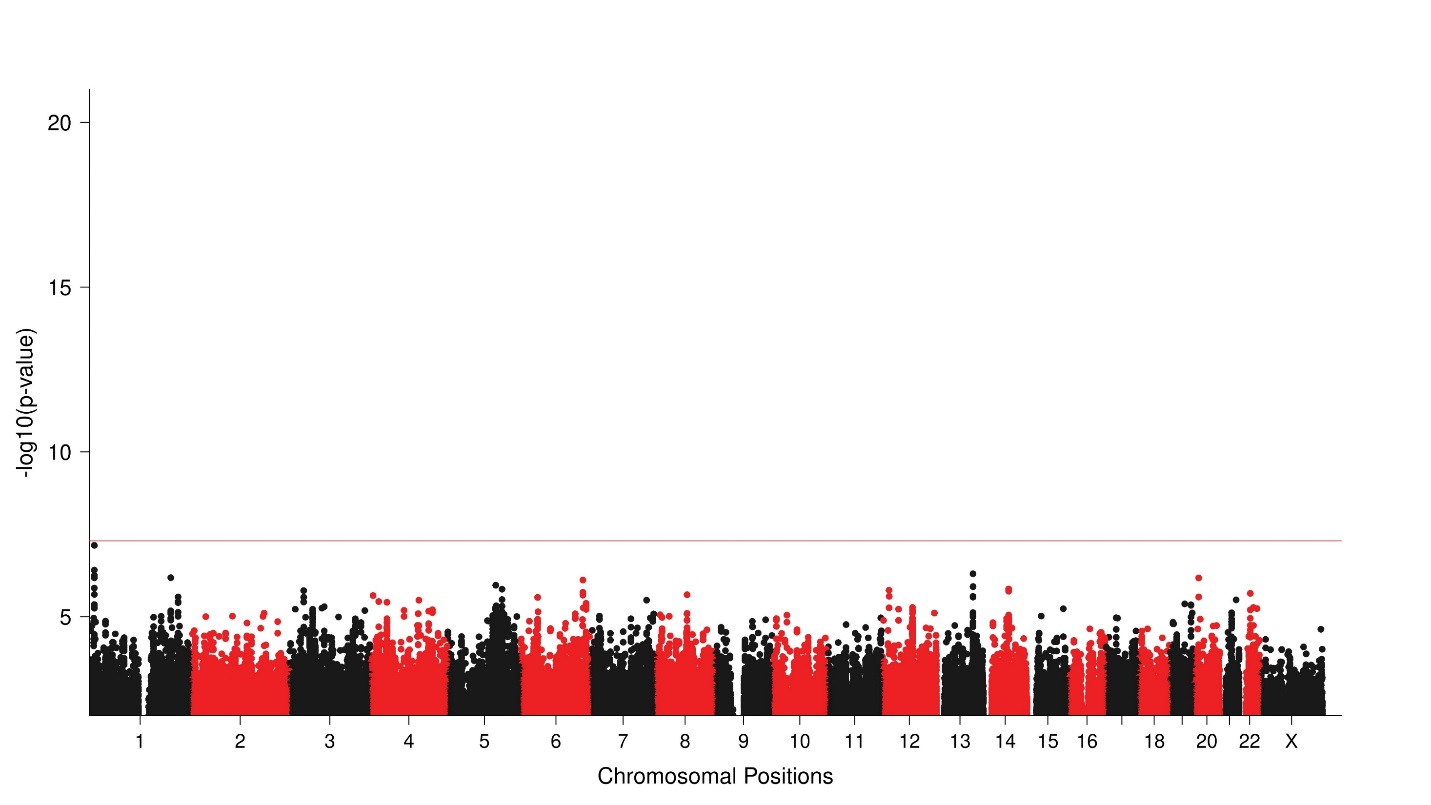

**Supplementary Figure 4. Manhattan plots of AA GWAS**

The *x axis* refers to the position on the genome, ordered by chromosome and base-pair position. The *y axis* refers to the −log10 p-value of association from GWAS. Each circle represents the association between a given single nucleotide polymorphism and PTSD. Circle colors alternate between chromosomes: even chromosomes colored red and odd chromosomes colored black. The horizontal red bar indicates genome-wide significance (p < 5x10^-8^).

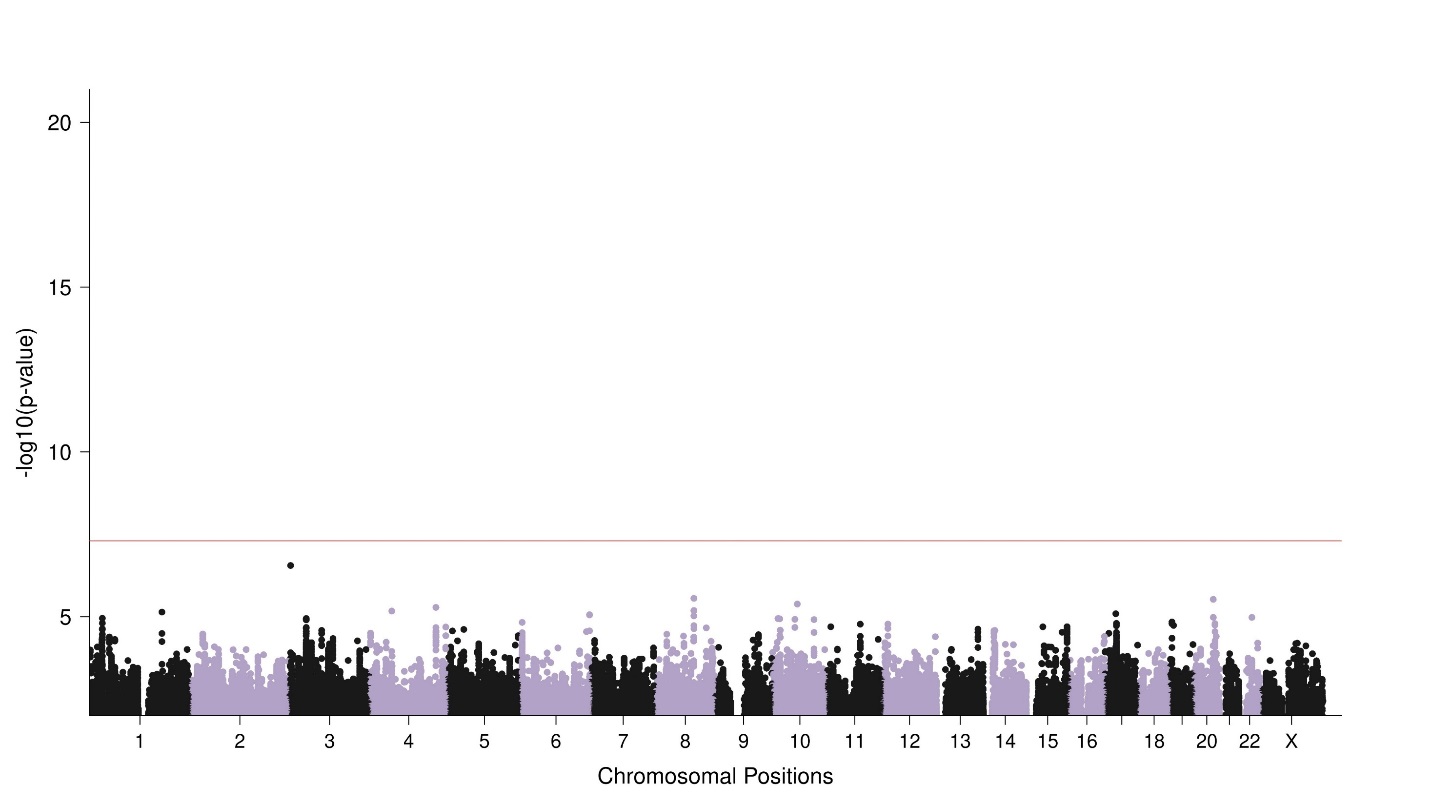

**Supplementary Figure 5: Manhattan plots of LAT GWAS**

The *x axis* refers to the position on the genome, ordered by chromosome and base-pair position. The *y axis* refers to the −log10 p-value of association from GWAS. Each circle represents the association between a given single nucleotide polymorphism and PTSD. Circle colors alternate between chromosomes: even chromosomes colored blue and odd chromosomes colored black. The horizontal red bar indicates genome-wide significance (p < 5x10^-8^).

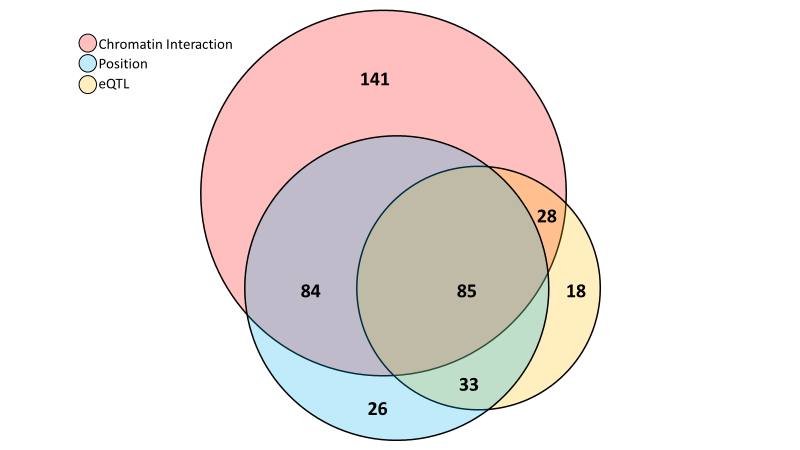

**Supplementary Figure 6: FUMA SNP2Gene gene-mapping counts.**

Genome-wide significant SNPS were mapped to genes using FUMA, based on positional, eQTL and chromatin interaction information of SNPs. Euler plot depicts the number of genes mapped by each gene-mapping method (chromatin interaction mapping, red; positional mapping, blue; eQTL mapping, yellow). Circles are sized in proportion to the number of genes mapped by a given strategy. Numbers within circles indicate the number of genes mapped by a given intersection of methods.

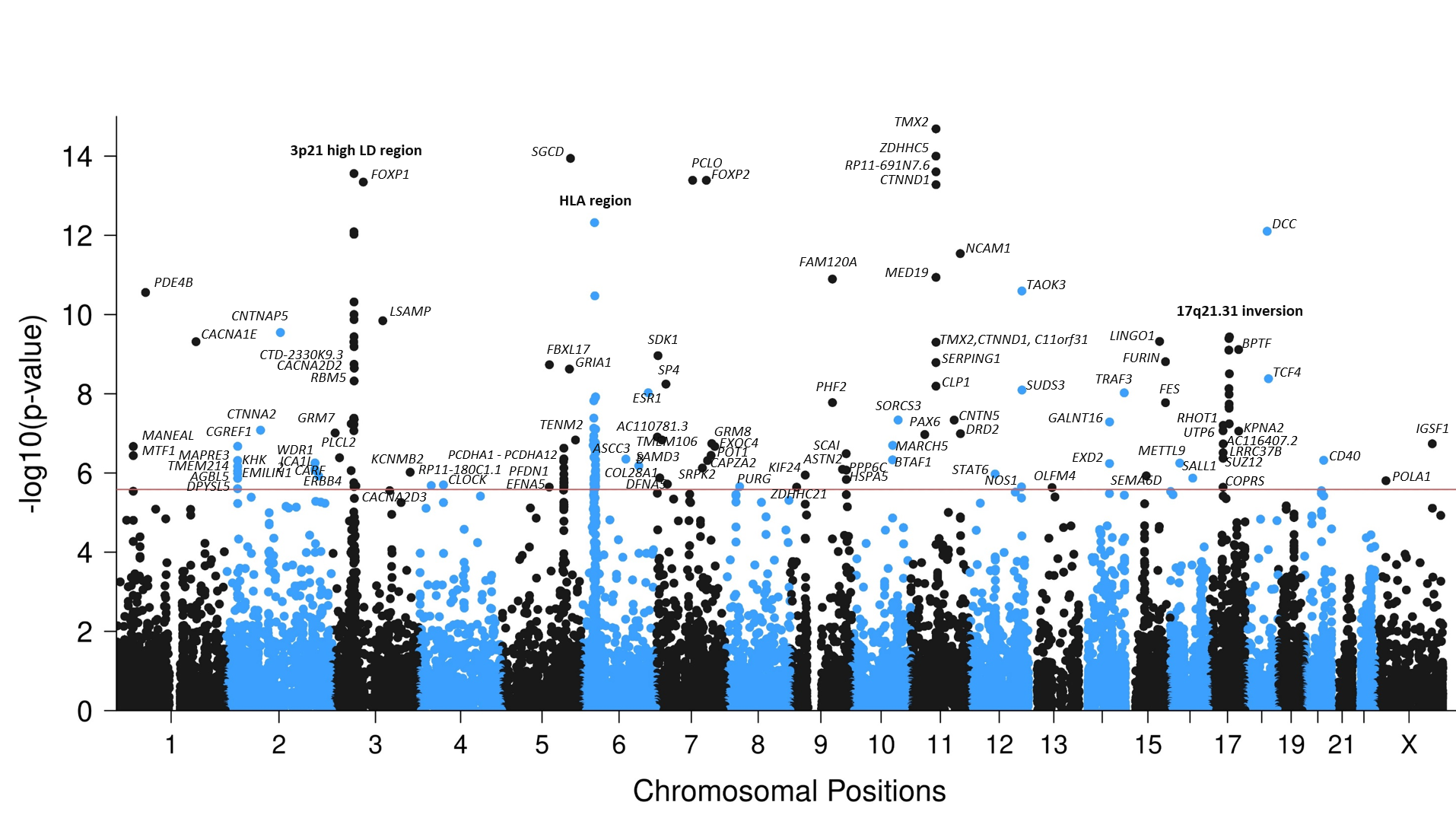

**Supplementary figure 7: Manhattan plots of gene-based associations from EA GWAS.**

The *x axis* refers to the position on the genome, ordered by chromosome and base-pair position. The *y axis* refers to the −log10 p value of association from gene-based analysis in MAGMA. Each circle represents the association between a given gene and the PTSD. Circle colors alternate between chromosomes, with even chromosomes colored blue and odd chromosomes colored black. The horizontal red bar indicates gene-wide significance (p < 0.05/19,106). All gene-wide significant gene names are labeled to the right of the corresponding dot. Three peaks are labeled by a descriptive name, gene names are provided here in order of p-value: 3p21 high LD region: *GRM7, PLCL2, SNRK, ANO10, LAMB2, TCTA, NICN1, BSN, MST1, RNF123, AMIGO3, GMPPB, IP6K1, CDHR4, UBA7, TRAIP, CAMKV, MST1R, CTD-2330K9.3, MON1A, RBM6, RBM5, SEMA3F, CACNA2D2, CACNA2D3.* HLA region: *BTN3A2, PRSS16, HIST1H2AJ, HIST1H2BN, HIST1H3I, HIST1H4L, HIST1H3J, OR2B2, ZKSCAN8, ZSCAN9, ZKSCAN4, PGBD1, OR12D3, GABBR1, TRIM31, TRIM26, PSORS1C1, HLA-C, HLA-B, LST1, PRRC2A, BAG6, CLIC1, MSH5, MSH5-SAPCD1, VWA7, VARS, LSM2, C6orf48, SKIV2L, TNXB, ATF6B*. 17q21.31 inversion: *ARHGAP27, PLEKHM1, CRHR1, SPPL2C, MAPT, STH, KANSL1, ARL17B, NSF, WNT3*

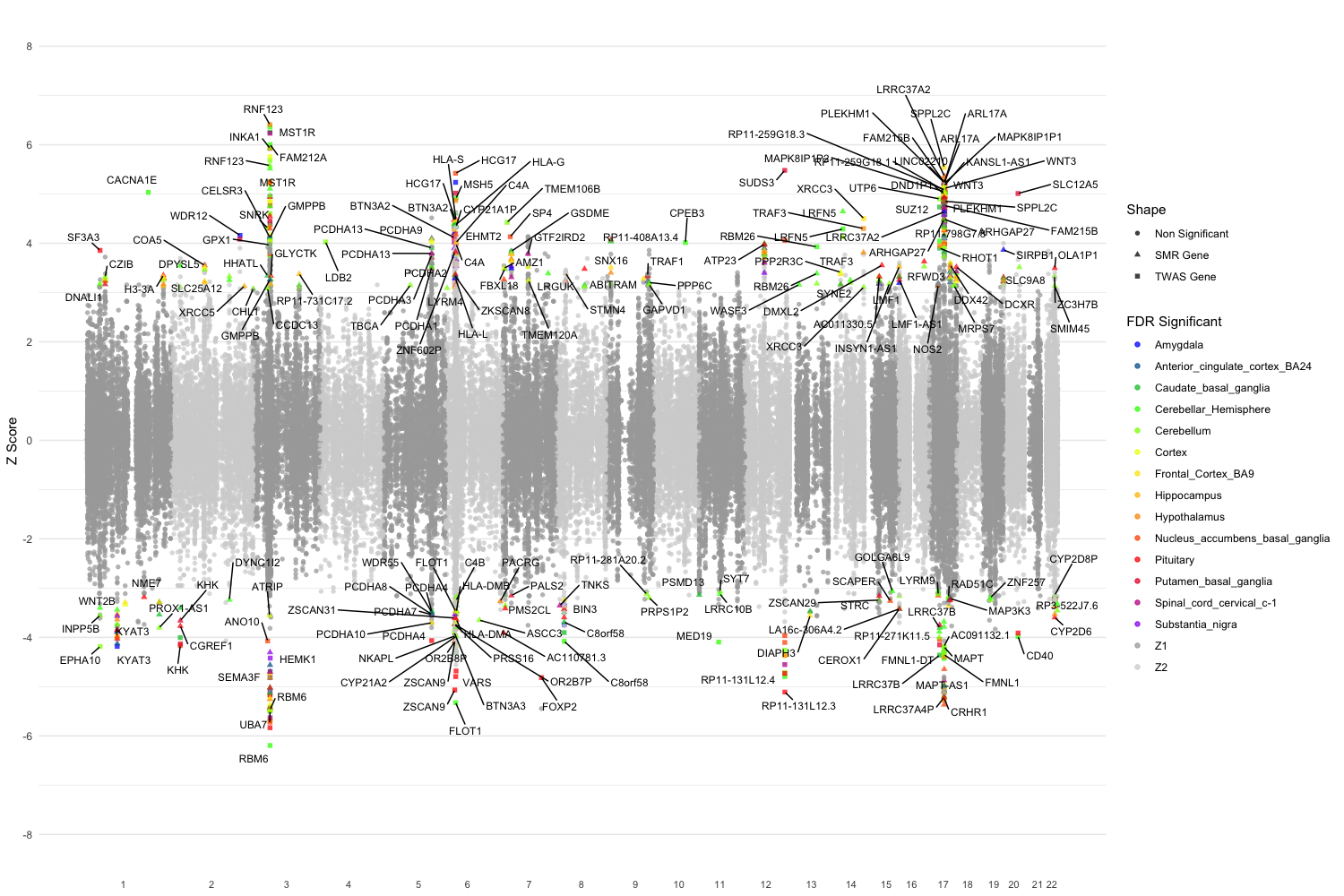

**Supplementary Figure 8: FDR significant eQTL SMR and TWAS results.**

The plot depicts genome-wide significant eQTL SMR (colored triangles) and TWAS (colored squares) results for each brain tissue/pituitary. FDR significant genes are labeled and colored according to the tissue type examined (refer to legend). Non-significant genes are depicted as grey circles. Values above the line indicate overexpression in cases relative to control, values below the line indicate under expression in cases relative to controls.

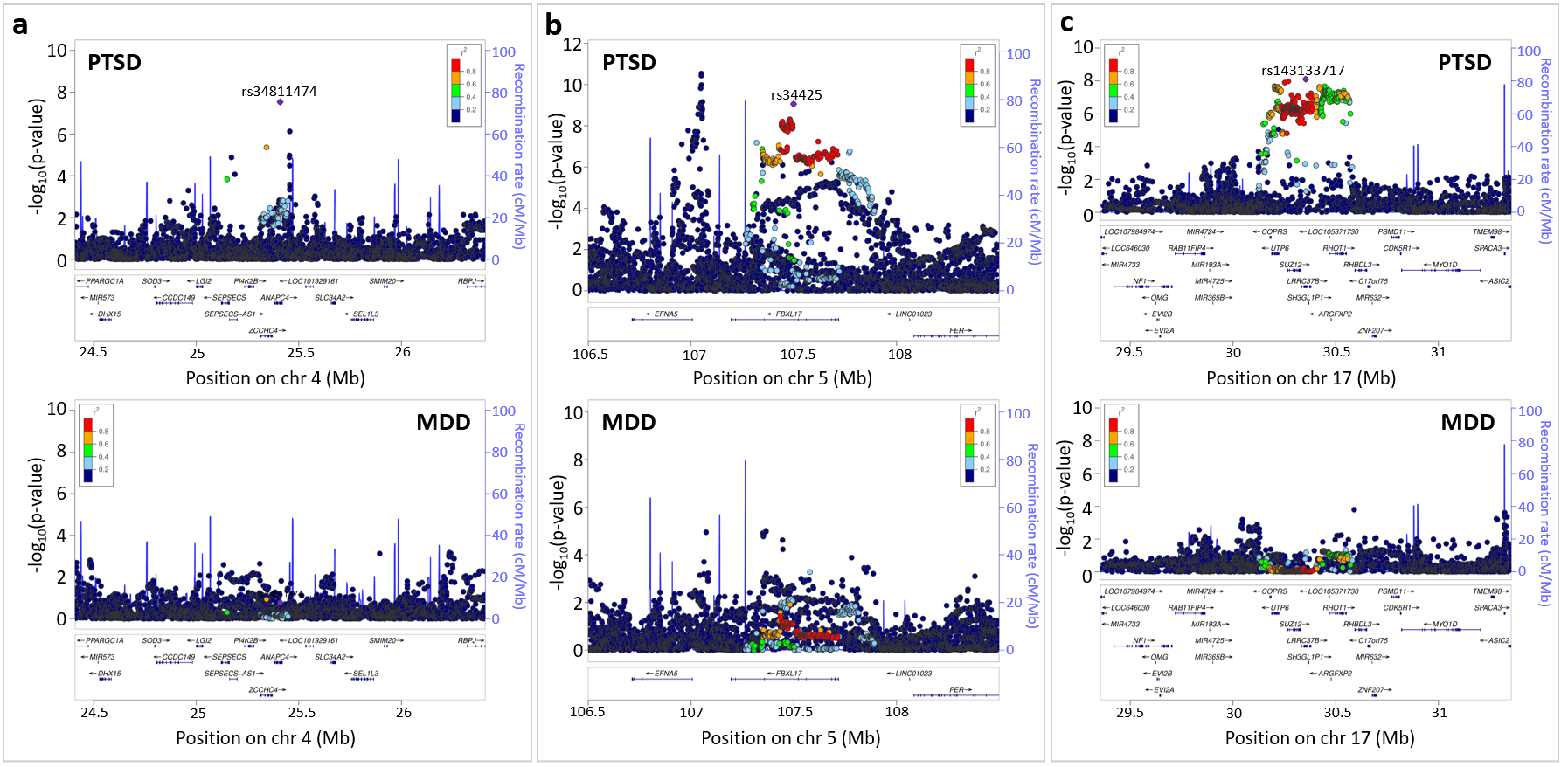

**Supplementary Figure 9: Regional comparisons between PTSD and MDD for three loci with low local genetic correlations between disorders.**

Local genetic correlations (derived from LAVA) between PTSD and major depression (MDD) were estimated within 81 significant loci from our EA PTSD GWAS. The figure showcases 3 example loci with non-Bonferroni significant genetic correlations between PTSD and MDD (p > 0.05/81), where we used regional association plots to visually contrast the local patterns of association seen in EA PTSD GWAS (top of panel) against those seen in MDD GWAS (bottom of panel). **a**, Locus #21. The local genetic correlation between PTSD and MDD was rho = -0.19 (95%CI=[-1.00,0.98], p = 0.71). **b**, Locus #28. The local genetic correlation between PTSD and MDD was rho = 0.86 (95%CI=[0.30,1.00], p = 0.01). **c**, Locus #81. The local genetic correlation between PTSD and MDD was rho = 0.21 (95%CI=[-0.54,0.94], P = 0.48). For each regional association plot, the x-axis represents the base pair (hg19) genetic position of SNPs. The y-axis represents –log p-values of SNP association with PTSD. LD estimates of surrounding SNPs with the labeled index SNP (LD r^2^ values estimated based on 1KGP3 Europeans) is indicated by color (color bar on side of plot indicates color coding of r^2^ values). Local estimates of recombination rate are indicated in light blue (legend on vertical axis at right). Gene names, strands, and boundaries are shown in the box below the regional plot.

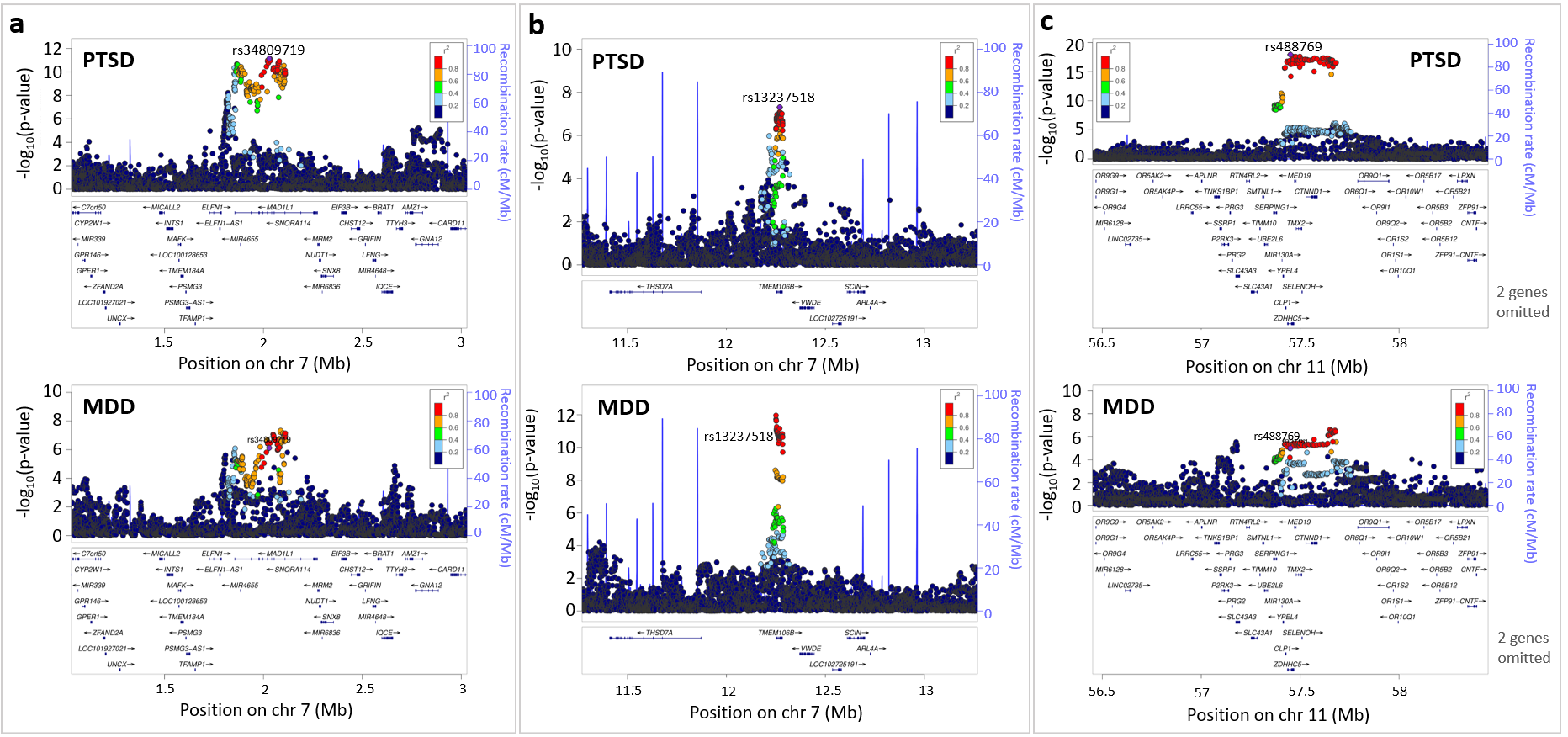

**Supplementary Figure 10: Regional comparisons between PTSD and MDD for three loci with high local genetic correlations between disorders.**

Local genetic correlations (derived from LAVA) between PTSD and major depression (MDD) were estimated within 81 significant loci from our EA PTSD GWAS. The figure showcases 3 arbitrarily chosen example loci with significant genetic correlations between PTSD and MDD (p < 0.05/81), we used regional association plots to visually contrast the local patterns of association seen in EA PTSD GWAS (top of panel) against those seen in MDD GWAS (bottom of panel). **a**, Locus #38. The local genetic correlation between PTSD and MDD was rho = 0.88 (95%CI=[0.52,1.00], p = 0.00041). **b**, Locus #40. The local genetic correlation between PTSD and MDD was rho = 1.00 (95%CI=[0.71,1.00], p = 0.00036). **c**, Locus #62. The local genetic correlation between PTSD and MDD was rho = 0.95 (95%CI=[0.65,1.00], p = 0.00004). For each regional association plot, the x-axis represents the base pair (hg19) genetic position of SNPs. The y-axis represents –log p-values of SNP association with PTSD. LD estimates of surrounding SNPs with the labeled index SNP (LD r^2^ values estimated based on 1KGP3 Europeans) is indicated by color (color bar on side of plot indicates color coding of r^2^ values). Local estimates of recombination rate are indicated in light blue (legend on vertical axis at right). Gene names, strands, and boundaries are shown in the box below the regional plot.
